# The methodologies to assess the effects of non-pharmaceutical interventions during COVID-19: a systematic review

**DOI:** 10.1101/2022.04.14.22273858

**Authors:** Nicolas Banholzer, Adrian Lison, Dennis Özcelik, Tanja Stadler, Stefan Feuerriegel, Werner Vach

## Abstract

Non-pharmaceutical interventions, such as school closures and stay-at-home orders, have been implemented around the world to control the spread of SARS-CoV-2. Their effects on health-related outcomes have been the subject of numerous empirical studies. However, these studies show fairly large variation among methodologies in use, reflecting the absence of an established methodological framework. On the one hand, variation in methodologies may be desirable to assess the robustness of results; on the other hand, a lack of common standards can impede comparability among studies. To establish a comprehensive overview over the methodologies in use, we conducted a systematic review of studies assessing the effects of non-pharmaceutical interventions on health-related outcomes between January 1, 2020 and January 12, 2021 (n=248). We identified substantial variation in methodologies with respect to study setting, outcome, intervention, methodological approach, and effect assessment. On this basis, we point to shortcomings of existing studies and make recommendations for the design of future studies.

## 1 Introduction

In response to the COVID-19 pandemic, countries around the world have implemented non-pharmaceutical interventions. These include a variety of public health measures implemented by governments with the intention of controlling, preventing, and mitigating transmission, e. g. school closures, stay-at-home orders, and mandates for compulsory wearing of masks in public places^1–4^. The widespread use of these interventions has raised interest in empirically studying their effects on health-related outcomes reflecting disease dynamics, e. g. the number of new cases or infection rates^5–10^. Such studies can play an important role in informing the discussion about the effectiveness of interventions. In particular, insights from the COVID-19 pandemic may contribute to an evidence-based public-health response in subsequent COVID-19 waves or future pandemics.

Accordingly, a plethora of studies assessing the effects of non-pharmaceutical interventions during the COVID-19 pandemic have been published. Their findings have been summarized by several meta-analyses^11–15^; nonetheless, each meta-analysis considered a different subset of studies. We argue that the latter is due to substantial variation in the methodologies used to conduct empirical studies on the effects of non-pharmaceutical interventions. The resulting lack of similarity constrains meta-analyses to a comparably small and specific subset of the overall evidence.

There are different reasons to expect variation in methodologies in the studies on the effectiveness of non-pharmaceutical interventions for controlling a pandemic. One possibility is the lack of empirical data before the COVID-19 pandemic, so that early studies have been largely theoretical^16^. Empirically assessing the effects of non-pharmaceutical interventions is therefore a relatively new subject, and corresponding studies do not build on an established scientific framework. Another possibility is that empirical assessments have been approached with different methods and domain knowledge by researchers from various fields, e. g. computational biology, infectious disease epidemiology, public health, economics, and statistical modeling.

Variation in methodologies can be manifold. Different study settings, outcomes, interventions, methodological approaches, and ways to assess effects may be used. On the one hand, such variation may be desired as it allows to assess the robustness of results against individual assumptions and methodologies. On the other hand, variation in methodologies can impede comparability among studies, which is necessary to arrive at conclusive evidence regarding the effects of non-pharmaceutical interventions.

Here, we systematically review the methodologies for studying the effects of non-pharmaceutical interventions on health-related outcomes published between January 1, 2020 and January 12, 2021 (n=248). Thereby, we aim to inform about different methodologies that were used by previous studies and promote common standards so that future studies can align with existing ones. In particular, we explore shortcomings of current studies and provide seven recommendations for subsequent studies on the effects of non-pharmaceutical interventions.

## 2 Results

Our review follows general guidelines for systematic literature reviews^17^ and is reported according the PRISMA 2020 statement^18^. The methodology was preregistered in a review protocol at PROSPERO^19^. Figure 1 shows the PRISMA flow diagram of our identification process. We conducted a systematic database search for peer-reviewed research articles from January 1, 2020 up to January 12, 2021 (see Materials and methods, Section 5), yielding 2,929 unique records of studies for screening. Through title and abstract screening, we identified 411 studies as potentially relevant and evaluated their full texts. Of these, we excluded 163 studies that did not meet the eligibility criteria. The most frequent reasons for exclusion were that (i) studies primarily simulated the effects of interventions in hypothetical scenarios rather than making inferences from observational data; (ii) studies had a different objective than assessing intervention effects, and (iii) studies only assessed the effects of population behavior (most often mobility) on health-related outcomes, but not the effects of interventions. The remaining n=248 studies met our eligibility criteria and were included for subsequent data extraction. Importantly, 35 studies in our review sample contained multiple (i. e. up to three) analyses, e. g. with different methodological approaches, leading to 285 different analyses included. If not indicated otherwise, our results are presented at the level of individual analyses (and *not* at the level of studies).

**Fig 1.**
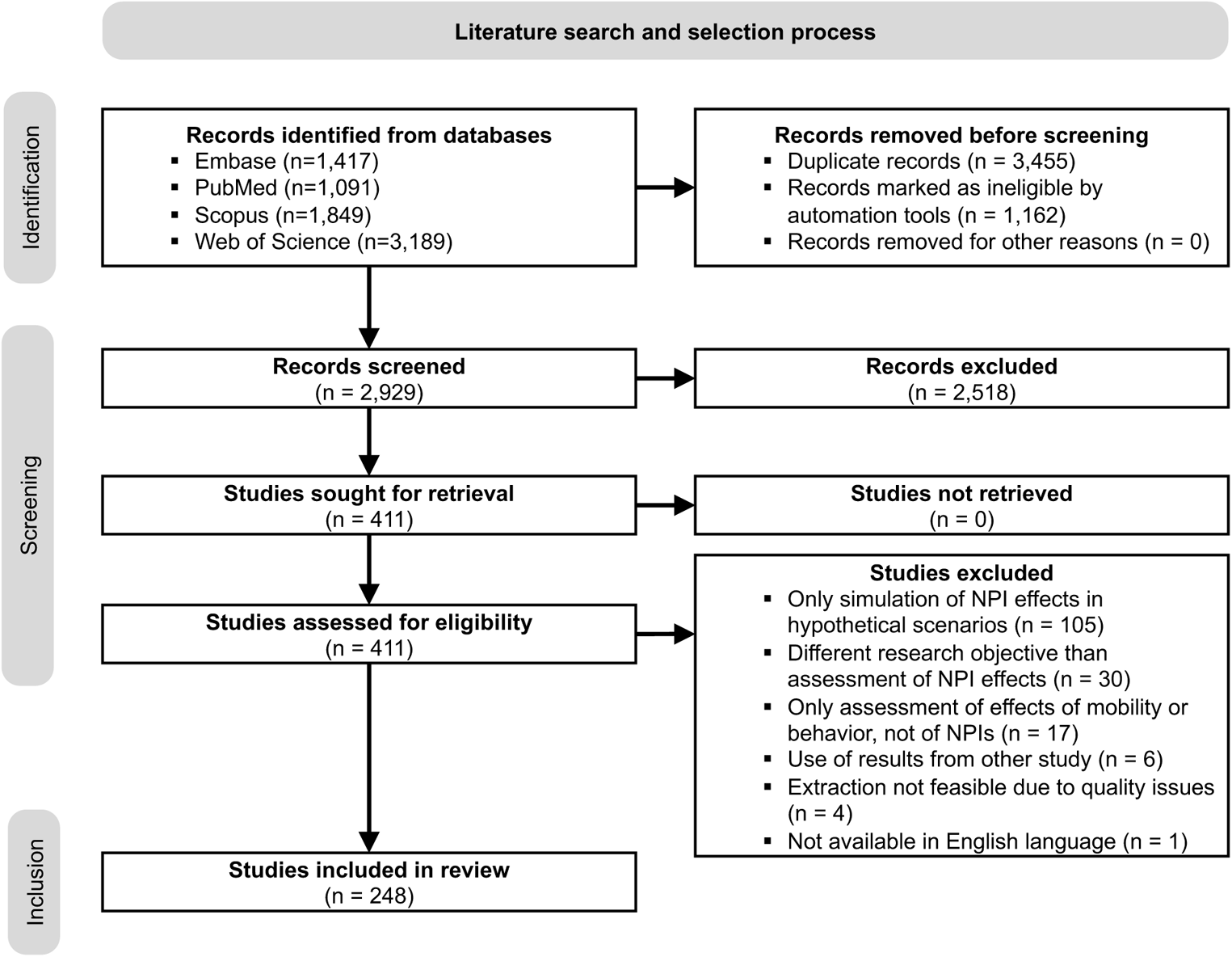
PRISMA flow diagram. Overall, n=248 studies were included. Some studies contain multiple analyses, such that the number of analyses included in the review is 285.

We characterized the analyses along five dimensions (SI Appendix D): study setting (D.1), outcome (D.2), intervention (D.3), methodological approach (D.4), and effect assessment (D.5). In the Results section, if not stated otherwise, we use the term *interventions* to refer to non-pharmaceutical interventions. Where appropriate, we also point to exemplary studies of specific characteristics. Due to the large size of our review sample, however, we refrain from referencing all studies in the main manuscript and instead refer to our complete data extraction report in SI Appendix E.

### 2.1 Study setting

The analyses vary in their scope across populations, geographic areas, and study period. A systematic classification of the study setting is shown in Table 1.

**Table 1.** Systematic classification and frequency of the study setting (D.1).

| **D.1.1: Number of populations included** | Frequency |
| --- | --- |
| Single (country, state, city, etc.) | 118 (41 %) |
| Multiple (countries, states, cities, etc.) | 167 (59 %) |
| **D.1.2: Level of populations included** |  |
| National (country-level) | 117 (41 %) |
| Subnational (e. g. state-level) | 71 (25 %) |
| Both national and subnational (country- and e. g. state-level) | 97 (34 %) |
| **D.1.3: Geographic areas covered<sup>‡</sup>** |  |
| Asia | 144 (51 %) |
| Europe | 109 (38 %) |
| North America | 91 (32 %) |
| Middle East and Africa | 49 (17 %) |
| Central and South America | 46 (16 %) |
| Oceania | 42 (15 %) |
| **D.1.4: Number of countries covered** |  |
| Multiple countries | 66 (23 %) |
| Single country (including multiple populations from a single country)<sup>‡</sup> | 219 (77 %) |
| ↳ China | ↳ 54 (25 %) |
| ↳ United States | ↳ 43 (20 %) |
| ↳ India | ↳ 11 ( 5 %) |
| ↳ Italy | ↳ 11 ( 5 %) |
| ↳ Other | ↳ 100 (46 %) |
| **D.1.5: Study period** |  |
| Start and end date span first epidemic wave | 161 (56 %) |
| One or more exceptions<sup>‡</sup> | 124 (44 %) |
| ↳ End date in growth phase of wave | ↳ 44 (35 %) |
| ↳ Same end date for several populations with diverse epidemic trajectories | ↳ 38 (31 %) |
| ↳ End date at peak of wave | ↳ 16 (13 %) |
| ↳ End date could not be evaluated | ↳ 14 (11 %) |
| ↳ Other | ↳ 14 (11 %) |

<sup>‡</sup> Multiple categories per analysis are possible. Frequencies refer to number of analyses to which category applies, proportions thus do not sum to 100 %.

#### Population

More than half of the analyses studied multiple populations (n=167; 59 %), i. e. multiple countries or subnational regions (e. g. states or cities). The remainder focused on a single population (n=118; 41 %), i. e. a single country or subnational region. The analyses were performed at the national level (n=117; 41 %), the subnational level (n=97; 34 %), or both (n=71; 25 %). If both levels were studied, the country and all its subnational regions were oftentimes considered, e. g. all states of the United States. Geographically, Asia (n=144; 51 %), Europe (n=109; 38 %), and North America (n=91; 32 %) were much more frequently analyzed than the rest of the world. Regional disparities may be explained by the fact that analyses frequently focused on some specific countries. For instance, in analyses of a single country, the most frequent country was China (n=54; 25 %), from where the pandemic originated. Other frequently studied countries, such as the United States (n=43; 20 %), India (n=11; 5 %), and Italy (n=11; 5 %), may have received more attention due to particularly high incidence and mortality during the first epidemic wave.

#### Study period

Typically, the study period covered both a rise and decline in new cases of the first epidemic wave in the analyzed population, and started before and ended after the analyzed interventions were implemented (n=161; 56 %). However, many analyses also deviated from this pattern in one or several aspects. Most often, study periods had a comparatively early end date, i. e. the study period ended already at the peak (n=16; 13 %) or still in the growth phase (n=44; 35 %) of the wave. There was also a considerable number of analyses that used the same study period for populations which were in different epidemic phases (n=38; 31 %). In such cases, the end date of the study period was still within the epidemic growth phase for some populations but already in the control phase for other populations.

### 2.2 Outcome

The studies in our review sample used different types of health-related outcomes or surrogates. For every analysis, we identified the “raw outcome”, i. e. the outcome data which were self-collected or obtained from external sources and used as input for the analysis. In around half of the analyses, the raw outcome was analyzed directly to assess the effects of interventions. The other half of analyses, however, involved an intermediate step, in which another outcome was computed from the raw outcome. This “computed outcome” was then analyzed instead of the raw outcome, or sometimes in addition to it. A systematic classification of the outcomes are shown in Table 2.

**Table 2.**
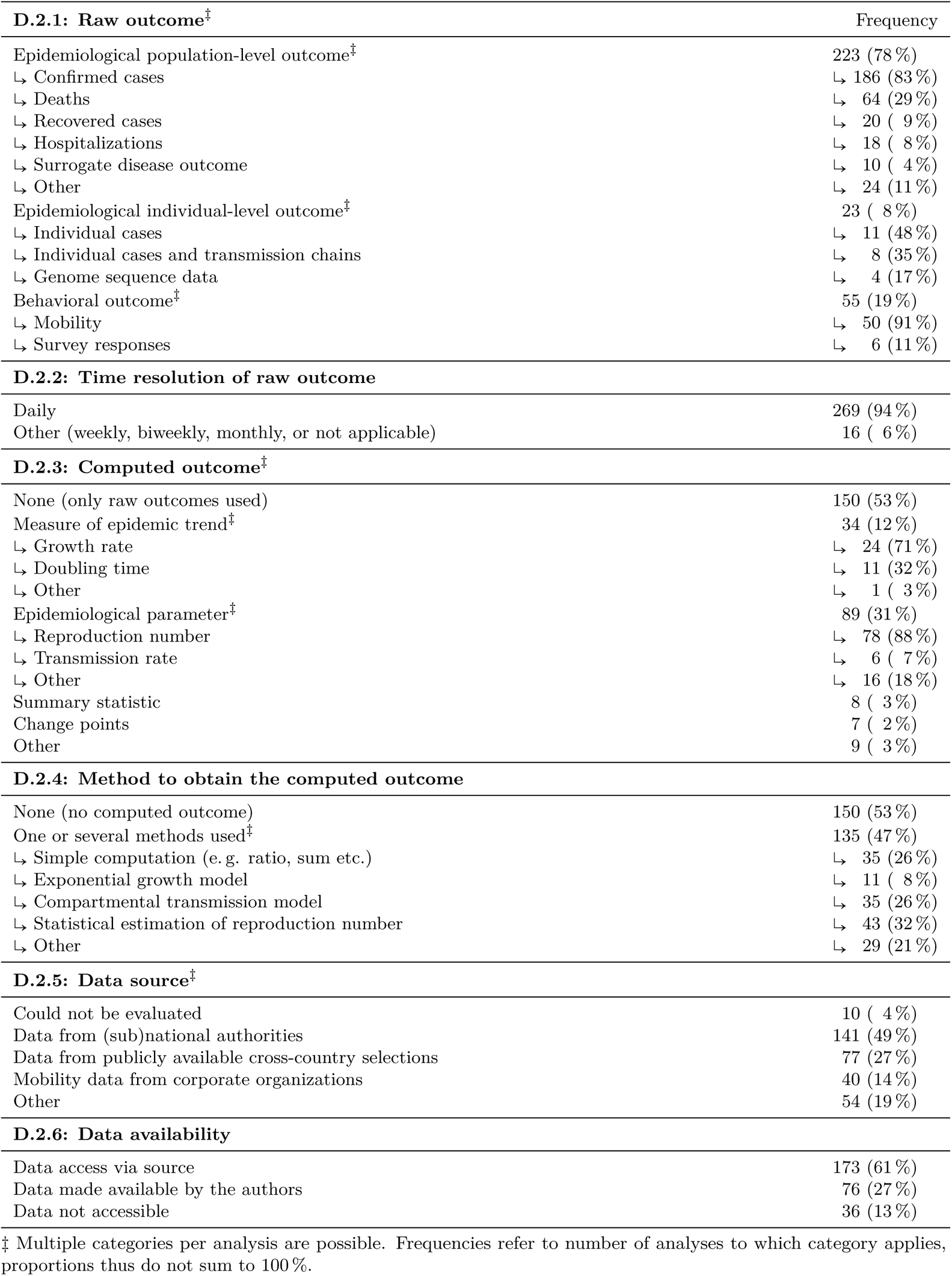
Systematic classification and frequency of the outcome (D.2).

#### Raw outcome

We identified three main types of raw outcome data used, namely (i) epidemiological population-level data, (ii) epidemiological individual-level data, and (iii) behavioral data.

##### (i) Epidemiological population-level data

The majority of analyses used population-level data on epidemiological outcomes (n=223; 78 %). The most frequent types were surveillance data, mainly the number of confirmed cases, but also deaths, hospitalizations, recovered cases, and, less frequently, intensive care unit (ICU) admissions. Importantly, some of these outcomes, such as recovered cases, were predominantly used to fit transmission models, in which case the effect of interventions was rather measured in terms of a different latent outcome (see D.5 Methodological approach, Section 2.4). Frequently, authors also included several types of data (e. g. both cases and deaths), either to perform a separate analysis for each (e. g. as a robustness check) or to combine them in a joint model (e. g. a transmission model). Some analyses used surveillance data on other diseases than COVID-19, with influenza being the most popular choice. Such surrogate diseases have often been monitored over an extended period of time, which allows comparing their spread during the COVID-19 pandemic to earlier years. Notably, we found only three analyses that used external data on latent epidemiological population-level outcomes (e. g. the reproduction number). All other analyses using a latent outcome self-computed it from raw data in an intermediate step (see D.2.2 Computed outcome, Section 2.2).

##### (ii) Epidemiological individual-level data

Instead of population-level data, some analyses also used individual-level epidemiological data (n=23; 8 %). These were in particular data about individual cases with case ID, demographics, and epidemiological characteristics (e. g. the date of symptom onset or travel history). In some instances, this included contact tracing data with links between index and secondary cases, allowing the reconstruction of transmission chains. Two analyses also used genome sequence data of clinical SARS-CoV-2 samples^20,21^.

##### (iii) Behavioral data

In addition to epidemiological data, a relevant share of analyses employed data on population behavior (n=55; 19 %), mainly mobility data. These data were usually obtained through tracking of mobile phone movements and provided as aggregates at the population level, based on summary statistics such as the daily number of trips made, distance travelled, time spent at certain locations, or population flow between regions. Another, less frequently used source of information on human behavior were surveys regarding social distancing practices, such as adherence to interventions, face mask usage, daily face-to-face contacts, or recent traveling.

#### Time resolution of raw data

Almost all raw data were obtained at daily resolution (n=269; 94 %). Exceptions were data on surrogate diseases or from surveys, where reporting was usually only broken down in weekly, biweekly, or monthly intervals (n=16; 6 %).

#### Computed outcome

Around half of the analyses involved an intermediate step, in which a latent outcome was computed from the raw data before assessing the effects of interventions. We identified four main types of computed outcomes:

1) Measures of epidemic trend (n=34; 12 %) were computed to describe the overall trend of the epidemic, e. g. through the growth rate or doubling time of confirmed cases or hospitalizations.
2) Epidemiological parameters (n=89; 31 %) were computed to measure specific infection dynamics, oftentimes in terms of the reproduction number. A few analyses also used individual-level epidemiological data to compute epidemiologically relevant time spans such as the serial interval or the time from symptom onset to isolation.
3) Summary statistics (n=8; 3 %) were computed to describe the progression of an epidemic in a certain population, e. g. the time until a certain number of documented cumulative cases was reached, or the time until the reproduction number first fell below one.
4) Change points in the outcome (n=7; 2 %) were computed with the aim to find time points of presumably structural changes in epidemic dynamics and compare them with implementation dates of interventions in the subsequent analysis^10,22,23^. Typically, change points were computed for the time series of confirmed cases or mobility.

Of note, the raw outcome was not always used only for obtaining the computed outcome, e. g. changes both in the number of new confirmed cases (raw outcome) and in the reproduction number (computed outcome) were sometimes analyzed.

#### Method to obtain the computed outcome

1. Measures of epidemic trend were often obtained through simple computation (e. g. growth rate as percentage change in confirmed cases). Other analyses used simple modeling approaches, e. g. fitting an exponential growth model to the time series and extracting the exponential growth rate or doubling time from the estimated parameters.
2. Epidemiological parameters were mostly estimated from confirmed cases or deaths. Some approaches fitted a compartmental transmission model to the raw epidemiological outcome. For this, the parameter of interest was either allowed to vary over time, or the model was fitted independently on different time periods. Other approaches employed a statistical method to directly estimate reproduction numbers from the observed outcome. Here, the method by Cori et al.^24^ as implemented in the popular software package “EpiEstim”^25^ for estimation of the instantaneous effective reproduction number was used in a large number of analyses. However, we found that statistical methods were not always applied correctly, which could have led to bias in the inferred transmission dynamics (see SI Appendix A). Sometimes, authors also used methods to estimate reproduction numbers from contact matrices^26^ (derived from surveys on personal contacts) or from transmission chains^27,28^ (derived from contact tracing data).
3. Summary statistics were typically obtained through simple computation.
4. Change points in the outcome were obtained by fitting a compartmental transmission model with special parameters representing points in time when the transmission rate changes. Other analyses used special change point detection algorithms.

#### Data source

The majority of authors directly accessed surveillance data from national health authorities or other governmental bodies (n=141; 49 %). In the case of individual-level data, which may be subject to privacy regulations, authors were often themselves affiliated to the relevant health authority. To obtain population-level data, a considerable share of analyses also used publicly available data from cross-country selections (n=77; 27 %), e. g. the European Centre for Disease Prevention and Control (ECDC)^29^, the Johns Hopkins University (JHU)^30^, or Worldometer^31^, which offer aggregated surveillance data internationally from various sources for the pandemic. Mobile phone tracking data were usually provided by corporate organizations (n=40; 14 %) such as Google^32^, Apple^33^, or Baidu^34^. A few analyses were also based on data collected by the authors, e. g. survey data on behavioral outcomes, seroprevalence studies, or data collected at a local facility such as a hospital.

#### Data availability

Data for the raw outcome was usually publicly available, in particular for epidemiological population-level outcomes such as cases and deaths because such data could oftentimes be accessed via the source that is documented in the manuscript (n=173; 61 %). In several cases, the data was made publicly available by the study authors (n=76; 27 %), e. g. by depositing the analyzed data in a public repository. For a small, yet considerable number of analyses, data was not accessible (n=36; 13 %) as the data was neither made publicly available nor the source of the data could be identified. Of note, data on epidemiological individual-level data was typically not available due to privacy concerns. Furthermore, corporate mobility data was widely available in the past, but access has recently been restricted by many providers.

### 2.3 Intervention

The analyses vary in the types of exposures and non-pharmaceutical interventions. A systematic classification is shown in Table 3.

**Table 3.** Systematic classification and frequency of the interventions (D.3).

| **D.3.1: Terminology for interventions<sup>†‡</sup>** | Frequency |
| --- | --- |
| Not applicable (only specific term for intervention type) | 22 ( 9 %) |
| Measures | 135 (54 %) |
| Interventions | 65 (26 %) |
| Policies | 16 ( 6 %) |
| Other | 14 ( 6 %) |
| **D.3.2: Terminology for the specific type of non-pharmaceutical interventions<sup>†‡</sup>** | Frequency |
| Not applicable (only general term for interventions) | 3 ( 1 %) |
| Non-pharmaceutical | 49 (16 %) |
| Control | 48 (16 %) |
| Social distancing | 45 (15 %) |
| Other | 159 (52 %) |
| **D.3.3: Exposure types** |  |
| One single intervention | 43 (15 %) |
| Multiple separate interventions | 31 (11 %) |
| One combination of interventions | 84 (29 %) |
| Multiple combinations of interventions | 20 ( 7 %) |
| All interventions together | 70 (25 %) |
| Other | 37 (13 %) |
| **D.3.4: Types of single interventions** |  |
| Not applicable (no single interventions analyzed) | 211 (74 %) |
| One or multiple single interventions analyzed (as defined in D.3.4 of the Documentation manual)<sup>‡</sup> | 74 (26 %) |
| ↳ Stay-at-home order | ↳ 44 (59 %) |
| ↳ Other | ↳ 27 (36 %) |
| ↳ School closure | ↳ 25 (34 %) |
| ↳ Workplace closure | ↳ 20 (27 %) |
| ↳ International travel restrictions | ↳ 17 (23 %) |
| ↳ Declaration of a state of emergency | ↳ 13 (18 %) |
| ↳ Bans of large gatherings | ↳ 13 (18 %) |
| ↳ Venue closure | ↳ 12 (16 %) |
| ↳ Bans of small gatherings | ↳ 10 (14 %) |
| **D.3.5: Coding of interventions** |  |
| **↳ D.3.6: Source of intervention data** |  |
| Not applicable (no specific interventions analyzed) | 74 (26 %) |
| Not necessary (no joint analysis of interventions across multiple populations) | 137 (48 %) |
| ↳ Could not be evaluated | ↳ 98 (72 %) |
| ↳ Government or news websites | ↳ 30 (22 %) |
| ↳ Other | ↳ 9 ( 7 %) |
| Necessary (joint analysis of interventions across multiple populations) | 74 (26 %) |
| ↳ Could not be evaluated | ↳ 9 (12 %) |
| ↳ Coding done by authors | ↳ 20 (27 %) |
| ↳ Use of externally coded data | ↳ 45 (61 %) |
| **D.3.7: Availability of data on exposure** |  |
| Not applicable (no specific interventions analyzed) | 73 (26 %) |
| Raw data documented in the manuscript | 136 (48 %) |
| Access to externally coded data via source | 32 (11 %) |
| Coded data made available by the authors | 34 (12 %) |
| Coded data not available | 10 ( 4 %) |

<sup>†</sup> Results for this subdimension are reported at the study-level, and not the level of analysis (i.e. one study can contain multiple analyses). If a study uses more than one term predominantly, then both are counted and added to the total count.
<sup>‡</sup> Multiple categories per analysis are possible. Frequencies refer to number of analyses to which category applies, proportions thus do not sum to 100 %.

#### Terminology for non-pharmaceutical interventions

Varying terminology was used by the literature to refer to non-pharmaceutical interventions. This is reflected in our search string, where we used a large set of terms in order to capture a broad range of relevant studies. One part of our search string considers the different terminology for interventions in general, while the other part considers the terminology for the specific type of non-pharmaceutical interventions. In our review, the most frequent terms for interventions were “measures” (n=135; 54 %), followed by “interventions” (n=65; 26 %) and “policies” (n=16; 6 %). The most frequent terms for the specific type of non-pharmaceutical interventions were “non-pharmaceutical” (n=49; 16 %), “control” (n=48; 16 %), and “social distancing” (n=45; 15 %). While terminology sometimes reflected the specific types of non-pharmaceutical interventions that were analyzed, differences in terminology may also be the result of different research backgrounds of the study authors.

#### Exposure types and types of single interventions

A considerable number of analyses examined one single (n=43; 15 %)^5,35^ or multiple interventions separately (n=31; 11 %)^2,7^. Among these analyses, the effects of school closures (n=25; 34 %) and stay-at-home orders (n=44; 59 %) were assessed most frequently, which may be due to these interventions being particularly controversial in the public discourse^36,37^. The majority of analyses, however, did not analyze the separate effects of multiple interventions but rather analyzed the joint effect for a combination of multiple interventions (n=84; 29 %)^38–40^, which is often the case when multiple interventions were implemented on the same day and when thus the separate effects could not be disentangled. A considerable number of analyses were even less specific by only analyzing whether interventions were altogether effective but without attributing effects to specific interventions (n=70; 25 %)^41,42^. Other ways to assess the effectiveness of interventions were: examining the start time of intervention^23,43^, e. g. to assess the effect of different delays with which governments responded to the pandemic^43^; dividing the public health response into different periods^44,45^; dividing interventions into different categories^46,47^; or summarizing the stringency of interventions to a numerical index at a specific time point^48,49^.

Of note, analyses that assessed the effects of a combination of interventions often referred to this combination as “lockdown”. In the underlying analysis, such lockdowns typically included multiple interventions implemented on the same day^39,50^. However, the specific interventions included in lockdowns varied considerably between populations. We therefore considered “lockdown” as an umbrella term for different combinations of interventions rather than as a specific type of intervention. Furthermore, some studies did not only assess the effects of non-pharmaceutical interventions on mobility, but also the effects of changes in mobility on population-level epidemiological outcomes. In these analyses, human mobility was typically defined as a continuous exposure. We extracted information on such complementary analyses of mobility as an addendum to the main review (see SI Appendix E).

#### Coding of interventions

When multiple populations were jointly analyzed, coding of interventions may have been necessary in order to reconcile differences in the definitions of interventions between populations. For instance, the term “school closures” could refer to the closure of primary or secondary schools or universities.

Differences across populations are thus reconciled during coding by deciding upon the type of intervention and providing a common name and definition that is then applied to all populations. Such coding of interventions was necessary in around a quarter of analyses (n=74; 26 %).

#### Source of intervention data

If coding of interventions was *not* necessary, authors often obtained intervention data (i. e. the date of interventions) from a government or news website (n=30; 22 %). Unfortunately however, the data source was often not provided by the authors and could thus not be evaluated (n=98; 72 %). If coding of interventions was necessary, then study authors either coded the data themselves (n=20; 27,%), i. e. collected the data from government or news websites and systematically categorized them, or used externally coded data instead (n=45; 61 %). The most popular choices for externally coded data were the Oxford Government Response Tracker^1^ and, for the United States, the New York Times^51^.

#### Availability of data on exposure

Authors using exposure data (type of exposure, interventions, and implementation dates) where coding of interventions was not necessary usually documented the data in the manuscript. Some authors using externally coded data chose to make the data available themselves (n=34; 12 %), e. g. by depositing it in a public repository, although many only referenced external data (n=32; 11 %).

### 2.4 Methodological approach

A variety of methodological approaches were used to assess the effects of interventions. The methodological approaches extracted here describe the actual stage of estimating the intervention effect. A systematic classification of the methodological approaches is shown in Table 4.

**Table 4.** Systematic classification and frequency of the methodological approach (D.4).

| **D.4.1: Empirical approach** |  |  |  | Total freq. |
| --- | --- | --- | --- | --- |
| **D:** Descriptive |  |  |  | 151 (53 %) |
| **P:** Parametric |  |  |  | 94 (33 %) |
| **C:** Counterfactual |  |  |  | 40 (14 %) |
| **D.4.2: Use of exposure variation** | (D) | (P) | (C) |  |
| Only variation over time for a single population | 78 | 23 | 24 | 125 (44 %) |
| Only variation over time for multiple populations | 63 | 22 | 10 | 95 (33 %) |
| Only variation between populations | 4 | 14 | 0 | 18 ( 6 %) |
| Both variation over time and between populations | 6 | 35 | 6 | 47 (16 %) |
| **D.4.3: Method** |  |  |  |  |
| Description of change over time | 136 | — | — | 136 (48 %) |
| ↳ Description of time course |  |  |  | ↳ 49 (36 %) |
| ↳ Comparison of time periods |  |  |  | ↳ 87 (64 %) |
| Comparison of populations | 8 | — | — | 8 ( 3 %) |
| Comparison of change points with intervention dates | 7 | — | — | 7 ( 2 %) |
| Non-mechanistic model | — | 61 | 17 | 78 (27 %) |
| ↳ Generalized linear model |  |  |  | ↳ 51 (65 %) |
| ↳ Exponential growth model |  |  |  | ↳ 11 (14 %) |
| ↳ Other |  |  |  | ↳ 16 (21 %) |
| Mechanistic model | — | 30 | 13 | 43 (15 %) |
| ↳ Compartmental single-population transmission model |  |  |  | ↳ 29 (67 %) |
| ↳ Compartmental meta-population transmission model |  |  |  | ↳ 4 ( 9 %) |
| ↳ Semi-mechanistic Bayesian transmission model |  |  |  | ↳ 5 (12 %) |
| ↳ Other |  |  |  | ↳ 5 (12 %) |
| Synthetic controls | — | — | 6 | 6 ( 2 %) |
| Other | 0 | 3 | 4 | 7 ( 2 %) |
| **D.4.4: Code availability** |  |  |  |  |
| None (not available) | 121 | 66 | 33 | 220 (77 %) |
| Publicly available | 30 | 28 | 7 | 65 (23 %) |

*Empirical approach:* (D) descriptive, (P) parametric, and (C) counterfactual
‡ Multiple categories per analysis are possible. Frequencies refer to number of analyses to which category applies, proportions thus do not sum to 100 %.

#### Empirical approach

We distinguished three general empirical approaches for assessing the effects of interventions, namely (**D**) descriptive, (**P**) parametric, and (**C**) counterfactual approaches.

- **(D)** Descriptive approaches (n=151; 53 %): These approaches provided descriptive summaries of the outcome over time or between populations, and related variation in these summaries to the presence or absence of different interventions. For example, some analyses compared changes in the growth rate of observed cases before and after interventions were implemented^52,53^. Of note, descriptive approaches could involve modeling as part of an intermediate step, where a latent outcome was computed from the raw outcome (see Computed outcome), while, afterward, a descriptive approach was used to assess the effect of interventions on the latent outcome. For example, some analyses used a single-population compartmental transmission model to estimate the time-varying reproduction number and then compared the reproduction number before and after interventions were implemented^54–56^.
- **(P)** Parametric approaches (n=94; 33 %): These approaches formulated an explicit link between the intervention and the outcome, where the effects of interventions were quantified via a parameter in a model. Most frequently these were regression-like links to estimate the effects of interventions on the reproduction number^2,8^.
- **(C)** Counterfactual approaches (n=40; 14 %): These approaches assessed the effect of interventions by comparing the observed outcome with a counterfactual outcome based on an explicit scenario in which the interventions were not implemented. For example, the observed number of cases was compared with the number of cases that would have been observed if the exponential growth in cases had continued as before the implementation of interventions^57,58^.

#### Use of exposure variation

Effects of interventions were assessed by exploiting variation in the exposure to the intervention over time, between populations, or both. Assessments exploiting exposure variation over time contrasted the outcome in time periods when specific measures were in place with the outcome in time periods when they were not in place. In contrast, assessments exploiting exposure variation between populations were based on a comparison of the outcome between populations that were subject to specific measures with populations that were not. In the majority of analyses, a single population was studied and thus only variation over time could be exploited in order to assess the effects of interventions (n=125; 44 %). In around one third of analyses, multiple populations were studied but only variation over time was exploited (n=95; 33 %), i. e. the effects of interventions were assessed within each population separately. A small number of analyses exploited only variation between populations (n=18; 6 %). Less than one in five analyses exploited both variation over time and between populations (n=47; 16 %).

#### Method

We grouped the different methods used into (i) description of change over time, (ii) comparison of populations, (iii) comparison of change points with intervention dates, (iv) non-mechanistic model, (v) mechanistic model, and (vi) synthetic controls. We review these in the following.

##### (i) Description of change over time

The large majority of analyses following a descriptive approach examined the change of the outcome over time to assess the intervention effect (n=136; 48 %). In some of these analyses, the focus was on the course of the outcome over time, typically by attributing the observed change (e. g. a reduction in new cases over time) to the analyzed interventions (n=49; 36 %). For example, the outcome was assessed at regular or irregular intervals, which were not necessarily aligned with the implementation dates of interventions^41,42,59^. The majority of analyses, however, followed the logic of an interrupted time series analysis, i. e. the outcome was explicitly compared between time periods before and after interventions (n=87; 64 %)^60–63^.

##### (ii) Comparison of populations

A few descriptive analyses compared outcomes via summary statistics only between populations (i. e. without considering variation over time) to assess intervention effects (n=8; 3 %). In such analyses, the outcomes were compared between populations that were stratified by different exposure to interventions (e. g. populations that implemented a certain intervention and populations that did not)^64–66^.

##### (iii) Comparison of change points with intervention dates

Some descriptive analyses checked whether the dates of estimated change points in outcomes and the implementation dates of interventions coincide (n=7; 2 %)^10,22,67^. If both dates were more or less in agreement, this was taken as evidence confirming the effectiveness of the intervention. However, change point detection methods could also yield change points prior to the implementation of interventions, which was sometimes interpreted as a sign of additional factors influencing the outcome (e. g. proactive social distancing)^22^.

##### (iv) Non-mechanistic model

Non-mechanistic models are statistical models that typically make *no* explicit assumptions about the mechanisms that drive infection dynamics. Such models were used in both parametric and counterfactual approaches (n=78; 27 %).

In parametric approaches, non-mechanistic models – almost always (generalized) linear regression models – were used to model a direct link between interventions and outcome. Typically, dummy variables were used to indicate when (variation over time)^9,68,69^ or where (variation between populations)^70–72^ interventions were implemented. Analyses exploiting both variation over time and between populations typically used panel regression methods^5,73,74^.

In counterfactual approaches, the non-mechanistic models used were mostly exponential growth models, and sometimes time series models (e. g. AR(I)MA or exponential smoothing)^38,44,58^. These models were fitted using data prior to when an intervention was implemented and then extrapolated the outcome afterwards.

##### (v) Mechanistic model

Mechanistic models have a structure that makes, to some extent, explicit assumptions about the mechanisms that drive infection dynamics. They were used in both parametric and counterfactual approaches (n=43; 15 %).

In parametric approaches, the effect of an intervention was represented via a parameter that was functionally linked to the disease dynamics (i. e. via a latent variable) of the model. This was typically achieved by parameterizing the transmission rate or reproduction number as a function of binary variables, indicating whether interventions were implemented or not^2,75–77^. Others linked the effects of interventions to the contact rate, the transmission probability upon contact, or to entries in the contact matrix^78–80^. A few modeling approaches also represented the intervention via an explicit structure or dynamic in the model, e. g. a compartment for quarantined individuals with a quarantine rate^47,81^ or an exponential decay of the susceptible population^46,47,82^.

The most popular mechanistic models used in parametric approaches were compartmental transmission models. These models were fitted to the time series of cases, hospitalizations, recovered cases, deaths, or several simultaneously. With the exception of one meta-population model^83^, all compartmental models used in analyses following a parametric approach were single-population models. If multiple populations were analyzed, each population was modeled separately. A few parametric analyses also used a semi-mechanistic Bayesian transmission model with a time-discrete renewal process, similar to the one in an early influential paper by Flaxman et al.^8^. These analyses fitted a Bayesian hierarchical model with stochastic elements for disease transmission and ascertainment on observed time series for cases, deaths, or both^2,8,84^. The model was usually fitted to data from several populations, modeling separately the time course in each population but estimating the parameters for the intervention effects jointly across populations. Rarely, analyses used highly complex models such as individual-based transmission models simulating the behavior of individual agents, or phylodynamic models inferring both virus phylogenies and transmission dynamics from genome sequence data.

In counterfactual approaches, mechanistic models were, similar to non-mechanistic models, calibrated to data before the implementation of an intervention and then projected the outcome for the time after the intervention, while keeping the model parameters fixed^85–87^. Thus, no relationship between intervention and outcome is explicitly modeled. Regularly, these analyses used meta-population or individual-based models that incorporated migration dynamics through mobility data and a network between individuals or populations^87–89^.

##### (vi) Synthetic controls

Some counterfactual approaches used synthetic control methods (n=6; 2 %). Here, a counterfactual scenario was constructed by computing the counterfactual outcome as a weighted combination of observations from a pool of “control” populations in which the intervention was not implemented^43,90,91^. Weights were fitted so as to give more importance to control populations similar to the intervention population. In these analyses, the course of the outcome before intervention was often used as the primary measure of similarity^6,90,91^. Sometimes, further factors such as geographic proximity or population characteristics were also considered^90,92^.

#### Code availability

For around one in four analyses, a link to a publicly accessible repository containing the computer code implemented for a specific analysis was provided (n=65; 23 %). Overall, the code availability was comparably higher for parametric approaches, where one in three analyses provided a link.

### 2.5 Effect assessment

The analyses in our review sample varied in their form of effect assessment, i. e. how the effect was quantified, whether uncertainty was reported, and whether sensitivity analyses or subgroup assessments were conducted. A systematic classification of the effect assessment is shown in Table 5.

**Table 5.** Systematic classification and frequency of different effect assessments (D.5).

| **D.5.1: Reporting of intervention effect** |  |  |  | Total freq. |
| --- | --- | --- | --- | --- |
| **QS:** Qualitative statement |  |  |  | 53 (19 %) |
| **CO:** Comparison of outcome values |  |  |  | 73 (26 %) |
| **QC:** Quantification of change in outcome values |  |  |  | 159 (56 %) |
| **D.5.2: Measure of effect<sup>‡</sup>** | **(QS)** | **(CO)** | **(QC)** |  |
| Change in reproduction number | 22 | 44 | 29 | 95 (33 %) |
| Change in confirmed cases | 16 | 15 | 38 | 69 (24 %) |
| Change in mobility | 9 | 6 | 28 | 43 (15 %) |
| Other | 18 | 29 | 100 | 147 (52 %) |
| **D.5.3: Reporting of uncertainty** |  |  |  |  |
| Not applicable |  |  |  | 52 (18 %) |
| Yes |  |  |  | 154 (54 %) |
| No |  |  |  | 79 (28 %) |
| **D.5.4: Sensitivity analysis (including computed outcomes)** |  |  |  |  |
| None (no sensitivity analyses w.r.t effect) |  |  |  | 217 (76 %) |
| One or more sensitivity analyses |  |  |  | 68 (24 %) |
| ↳ Model specification varied |  |  |  | ↳ 36 (53 %) |
| ↳ Epidemiological parameters varied |  |  |  | ↳ 29 (43 %) |
| ↳ Different or modified outcome used |  |  |  | ↳ 17 (25 %) |
| ↳ Same analysis with (sub)population excluded |  |  |  | ↳ 16 (24 %) |
| ↳ Different coding of interventions used |  |  |  | ↳ 10 (15 %) |
| ↳ Start or end date of study period varied |  |  |  | ↳ 4 (6 %) |
| **D.5.5: Subgroup assessment** |  |  |  |  |
| None (no subgroups) |  |  |  | 250 (88 %) |
| One or more subgroups<sup>‡</sup> |  |  |  | 35 (12 %) |
| ↳ Based on socioeconomic indicators |  |  |  | ↳ 23 (66 %) |
| ↳ Based on epidemiological indicators |  |  |  | ↳ 16 (46 %) |
| ↳ Based on public health response |  |  |  | ↳ 9 (26 %) |
| ↳ Based on geographic areas |  |  |  | ↳ 6 (17 %) |

*Reporting of intervention effect:* **(QS)** qualitative statement, **(CO)** comparison of outcome values, and **(QC)** quantification of change in outcome values
<sup>‡</sup> Multiple categories per analysis are possible. Frequencies refer to number of analyses to which category applies, proportions thus do not sum to 100 %.

#### Reporting of intervention effect, measure of effect, and reporting of uncertainty

The are difference in how effects of interventions were reported. Around one in five analyses made a qualitative assessment of the intervention effect (n=53; 19 %), e. g. by qualitatively describing the change in the outcome over time following the implementation of interventions. More frequent was the reporting of comparisons of outcome values to assess the intervention effect (n=73; 26 %), i. e. by comparing the outcome values before an intervention with the outcome values after an intervention. Around half of the analyses reported a quantitative change in outcome values (n=159; 56 %), e. g. by computing the difference in the outcome values before and after an intervention, or estimating the difference via a parameter in a statistical model. The intervention effect was oftentimes measured in terms of a change in the reproduction number (n=95; 33 %), in confirmed cases (n=69; 24 %), or in mobility (n=43; 15 %), but many other measures of effect were also common. Notably, a relevant number of analyses in our review sample focused its methodology on estimating the reproduction number as the computed outcome and then conducted only a qualitative assessment of the intervention effects afterwards. Uncertainty was reported in around one half of the analyses (n=154; 54 %), e. g. via standard error, confidence intervals, and credible intervals.

#### Sensitivity analyses

We checked all works for sensitivity analyses that were specifically conducted to examine the robustness of the reported intervention effects. Many studies conducted sensitivity analyses only related to the predicted outcome or model fit, but not to the intervention effect. Overall, the vast majority of analyses did not conduct sensitivity analyses with regard to the intervention effect (n=217; 76 %).

Of those that did, sensitivity analyses focused on model extensions or adjustments in which the model specification was varied (n=36; 53 %), e. g. by changing the structure of a transmission model or by adjusting the estimated effects of interventions for additional variables in a regression model. Others analyzed sensitivity with respect to variations in epidemiological parameters (n=29; 43 %), e. g. by assuming a different basic reproduction number, generation or serial interval, infectious period, or reporting delay distribution. Only few analyses tested sensitivity with regard to data: i. e. using different or modified outcomes^69,93^ (n=17; 25 %); using a different coding of interventions^3,7^ (n=10; 15 %); or repeating the same analysis but excluding (sub)populations^2^ (n=16; 24 %). Of the different methodological approaches, analyses with semi-mechanistic Bayesian transmission models generally conducted more comprehensive sensitivity analyses. For example, one work^2^ performed checks regarding data, model, and epidemiological parameters and presented the results prominently in the main manuscript.

#### Subgroup assessment

The effects of interventions were rarely assessed within subgroups of the population (n=35; 12 %). Two thirds of such assessments were within subgroups created based on socioeconomic indicators (n=23; 66 %), e. g. assessing intervention effects within groups of individuals of different age and gender^41^ or assessing the effect of lockdown within low- and high-income income regions^6^. Less frequent were subgroups based on epidemiological indicators^5^, the public health response^94^, or geographic areas^95^.

## 3 Discussion

Our systematic review covers over 240 studies published between January 2020 and January 2021. Insights from this review can inform different types of future studies: (i) studies using data from the same period that extend our knowledge on aspects that have so far been rarely investigated; (ii) studies using data from subsequent periods that generate new insights or corroborate existing ones; and (iii) studies using data from a future pandemic caused by another virus. Although the preconditions to conduct these studies differ, they share the goals and challenges of the studies in our review sample. Accordingly, the results from our systematic review allow us to make seven recommendations for the design of future studies, which are discussed in the following.

### Recommendation 1: Exploring the value of rarely analyzed outcomes

During the COVID-19 pandemic, both surveillance data on confirmed cases, hospitalizations, or deaths^29,30^, and mobility data from mobile phones^32,33^ have become publicly available at scale. This has enabled a large number of studies assessing the effect of non-pharmaceutical interventions on population-level epidemiological outcomes and on human mobility (Table 2, D.2.1). However, there remains considerable potential to explore the value of other outcomes and data sources that have so far been rarely analyzed.

First, more detailed insights into the effects of non-pharmaceutical interventions could be gained from using individual-level data. This has been demonstrated by studies in our review sample, e. g. by relating the effects of non-pharmaceutical interventions to the serial interval using symptom onset data^41^, to transmission chains using contact tracing data^87^, or to virus migration rates using genome sequence data^20^. We hope to see more such analyses as more individual-level data becomes available. Second, there can be great merit in analyses advancing our understanding of the mechanisms by which non-pharmaceutical interventions work. Our review sample contained many analyses assessing the effects of interventions on human behavior, or of human behavior on epidemiological outcomes. The majority of these analyses used mobility data from mobile phones (Table 2, D.2.1); however, more effort is needed to obtain a complete picture of how population behavior mediates the effects of non-pharmaceutical interventions. For example, interventions may influence behaviour and transmission through further factors not captured by previous studies, and, moreover, the relationship between mobility and disease transmission may change over time^96,97^. Additional insights can be gained from analyses using behavioral data from other sources, e. g. surveys evaluating compliance with mask mandates^62^ or the number of daily contacts^98^. Moreover, we see value in analyzing interventions, behavior and epidemiological outcomes jointly, i. e. in the form of a mediation analysis^99,100^, allowing to differentiate the direct and indirect effect of non-pharmaceutical interventions.

### Recommendation 2: Exploiting variation in the exposure to interventions between populations

Variation in the exposure to interventions (i. e. when, where and which interventions were implemented) is required in order to empirically assess their effects. Variation over time was the most common choice in our review sample (Table 4, D.4.2). Here, the effects were previously assessed within populations based on differences in outcomes before and after non-pharmaceutical interventions were implemented. However, such changes may falsely be attributed to non-pharmaceutical interventions if they are subject to confounding by concurring time trends. We thus recommend to also exploit exposure variation between populations, i. e. with respect to the timing and the types of single interventions that were implemented. This was done by only one in five analyses in our review sample (Table 4, D.4.2), although the types and timing of interventions varied considerably between the populations. This leaves a valuable source of variation largely untapped.

### Recommendation 3: Dissecting the effects of single interventions

Evidenced-based decision making requires empirical estimates for the effects of single non-pharmaceutical interventions (e. g. school closures or stay-at-home orders). However, the majority of analyses assessed the effects of population-specific combinations of interventions such as lockdowns (Table 3, D.3.3). The underlying analyses typically studied only a single population (or multiple populations separately) where multiple interventions were implemented on the same day, and, as a result, the separate effects of interventions cannot be disentangled. For future work, we recommend more effort to conduct analyses across multiple populations, so that the separate effects of single interventions can be dissected.

### Recommendation 4: Careful coding of interventions

Systematic coding of intervention data is necessary to compare the effects of non-pharmaceutical interventions across multiple populations (Tabble 3, D.3.5). However, the process of such systematic coding involves subjective decisions regarding if and when non-pharmaceutical interventions have been implemented^2^. These decisions are sometimes unavoidable, and they are also inherent to the development of public databases^101^. As a result, different coding of interventions could impact the results, and, therefore, coding decisions should be made transparent and documented carefully. In addition, we recommend to examine empirically the impact when using a different coding of interventions, e. g. by comparing the results across different input data.

### Recommendation 5: Promoting comparability across different analyses

During a pandemic, public health policy has a strong focus on the number of confirmed cases, hospitalizations, and deaths, making them obvious outcomes to evaluate the effectiveness of interventions. However, non-pharmaceutical interventions act only indirectly or with a certain delay on these observable outcomes. Typically, non-pharmaceutical interventions should influence the behaviour of the population, which should reduce transmission (e. g. by limiting the contact rate), which in turn should affect the number of new infections and, subsequently, observed outcomes like confirmed cases, hospitalizations, or deaths. The question of how to assess intervention effects along this path has been answered differently by the studies in our review sample. We identified four main types of analyses; see (1)–(4) in Box 1. In the following, we discuss the different types with regard to their ability of enabling a comparison of results between studies.

#### Box 1: Different types of analyses to assess the effects of non-pharmaceutical interventions.

**(1) Observed outcome directly linked to interventions**

A raw, observed outcome is analyzed directly by evaluating differences (i) over time with an interrupted time-series analysis comparing the outcome before vs. after an intervention, (ii) between populations with a cross-sectional analysis comparing populations exposed vs. not exposed to an intervention, or (iii) both over time and between populations with a panel data analysis. Mechanistic modeling is typically not involved in this type of analysis, with one exception, namely counterfactual approaches using a transmission model to project the observed outcome after intervention.

**(2) Computed, unobserved outcome linked to interventions**

In contrast to type (1), the intervention effect is measured in terms of an unobserved outcome. This is computed from the raw outcome and then analyzed in a similar manner as in (1). Mechanistic modeling can be involved in computing the unobserved outcome, for example by using a model to estimate the reproduction number or transmission rate from the number of new cases.

**(3) Observed outcome linked to interventions via unobserved outcome in mechanistic model**

Observed outcomes are used to fit a mechanistic model (e. g. compartmental transmission model) that includes a latent variable representing an unobserved outcome (e. g. the reproduction number), which in turn is parameterized as a function of interventions. For instance, a regression-like link is used within the mechanistic model to estimate the effect of interventions on the transmission rate as a latent variable.

**(4) Change points in outcome related to exposure**

Change points are estimated in the time series of an observed or unobserved outcome. The estimated change points are then related to the implementation dates of interventions. If the estimated change points agree well with the actual implementation dates of interventions, this is interpreted as evidence for the effectiveness of interventions.

Analyses of type (1), where the observed outcome is directly linked to interventions, can avoid mechanistic modeling by directly analyzing an observed outcome such as cases or deaths. Here, a central challenge is to take into account the uncertain delay between the implementation of non-pharmaceutical interventions and their effects on the observable outcome. The fact that infections and subsequent outcomes such as confirmed cases follow exponential dynamics during an epidemic wave makes it difficult to compare intervention effects measured by observable outcomes across different epidemic phases. In contrast to that, analyses following type (2) or (3) employ mechanistic modeling, allowing to assess the effects of interventions on latent, unobservable outcomes such as the transmission rate or the reproduction number. Since these latent outcomes can be inferred from different observed outcomes like cases or deaths, it becomes possible to compare analyses that use different raw data. The difference between type (2) and (3) is that for (2) the estimation of the latent outcome is separated from the effect assessment. Such separation reduces model complexity, however, often at the expense of incomplete uncertainty assessments. The reason is that most analyses based on type (2) only incorporated uncertainty involved in modeling the effect of interventions on the latent outcome, thus leaving out uncertainty involved in the computation of the latent outcome. This also means that differences in uncertainty assessments must be carefully taken into account when synthesizing results from multiple studies.

Finally, analyses following type (4), where change points in the outcome are related to the exposure, take a very different approach that shares few assumptions with the other approaches. A comparison of change points can verify the presence of an effect, yet without quantifying its size. As a result, such findings are best complemented with an analysis of type (1), (2), or (3).

### Recommendation 6: Understanding variation in effectiveness across subgroups

The effects of non-pharmaceutical interventions may vary between and within populations. However, only one in ten analyses in our review sample examined variation across subgroups or populations (Table 4, D.4.2). Estimating and explaining such variation could help understand the conditions under which interventions are more or less effective for a specific subgroup, potentially allowing policy makers to tailor interventions to a specific subgroup or setting. Our review points out two approaches that can contribute towards a better understanding of the varying effectiveness of non-pharmaceutical interventions: (i) comparing the effects of interventions between subgroups of the same population (e. g. between the young and elderly population); and (ii) comparing the effects of interventions between different populations and relating differences to population-specific characteristics (e. g. population density).

### Recommendation 7: Assessing sensitivity within and between studies

While variation in methodologies can complicate the comparability among studies, it may help to identify the influence of certain methodological choices on the results. Here, the public availability of data for outcomes and interventions holds potential for sensitivity analyses within studies as well as comparisons between studies. Specifically, the same analysis could be repeated with different sets of publicly available data as part of the same study. This way, sensitivity of the findings with respect to the choice of outcome and intervention data could be assessed within studies, reducing the risk of bias from specific outcome data (e. g. incomplete case ascertainment due to limited testing capacity etc.) or the specific coding of interventions. For example, the number of new cases, deaths, or both could be used as the raw outcome in mechanistic models with a comparable latent outcome^2^. However, other aspects, in particular the specific setting and methodologies used, are presumably more difficult to vary as part of a sensitivity analysis, and may therefore need to be compared between different studies. Important for such comparisons is giving access to the preprocessed data, yet most study authors accessed these data directly from national authorities or external data providers without making the preprocessed data additionally available (Table 2, D.2.5-D.2.6 and Table 3, D.3.6-D.3.7). To promote reproducibility, study authors should always make the preprocessed data available for two reasons. First, future access to the data via the original source may be restricted (this has already been observed for corporate mobility data). Second, originally analyzed and recently accessed data may be different as publicly available epidemiological data and coded intervention data can be subject to updates, revisions, and corrections. In addition, publication of computed code is crucial to support comparisons of methodological approaches, specific modeling choices, and input data.

## 4 Conclusions

Our review of more than 240 studies on the effects of non-pharmaceutical interventions revealed substantial variation in methodologies. Until specific best practices emerge, further heterogeneity in studies is inevitable and can also be beneficial, e. g. for assessing robustness of the results with respect to method and input data. Nevertheless, some standardization is required in order to synthesize evidence on the effects of non-pharmaceutical interventions from multiple studies. So far, a lack of common standards and substantial variation in the methodologies used have created a challenge for meta-analyses to summarize and compare the reported effects from existing studies^11–15^. Here, our methodology review can serve as a basis for subsequent meta-analyses to factor in the variety of existing methodologies when pooling and comparing the large number of effects that have been reported for non-pharmaceutical interventions on health-related outcomes. More importantly though, our review promotes common standards and reduces barriers to comparability across studies by making recommendations for the design of future studies.

A general limitation of the studies included in our review is that it is difficult to estimate causal effects based on population-level observational data and to rule out unobserved confounding. This is also highlighted by the persistent debate on the role of voluntary behavioral change in curbing transmission^102–105^. Evidence from observational studies on the effects of non-pharmaceutical interventions should thus be evaluated in conjunction with evidence from other fields. For example, evidence on the infectiousness of school children and parental strategies to fill the care gap can produce independent predictions about the effects of school closures. Similarly, laboratory evidence regarding the effectiveness of masks together with evidence on compliance with masks can produce independent predictions of the effectiveness of mask mandates.

During the COVID-19 pandemic, a tremendous amount of publicly available epidemiological data has been generated. The ease of access to this data allowed many researchers to contribute work, using a variety of methodologies to assess the effects of non-pharmaceutical interventions on health-related outcomes. With researchers from diverse fields contributing, there is a unique opportunity to benefit from the various inputs in developing a methodological foundation for timely and robust assessments during future pandemics. This will however require a thorough examination of the present methodologies in order to share lessons learnt and develop best practices. Our systematic review can be viewed as a first such attempt.

## 5 Materials and methods

We tailored our review to the challenge of mapping a potentially diverse set methodologies from a large number of studies. To ensure rigour and consistency, we preregistered the procedures for all stages of the review process, following common guidelines for systematic literature reviews^17^. Certain guidelines were not applicable to a methodology review as ours. In particular, our eligibility criteria and risk of bias assessment reflect the objective of this review, which was not to evaluate the effects of non-pharmaceutical interventions, but to map the variation in methodologies used. The preregistered methodology was documented in a review protocol at PROSPERO^19^.

We report our review according to the PRISMA 2020 statement^18^. A completed PRISMA 2020 checklist is provided in SI Appendix G. We conceptualized the review by drawing on experience from our own primary research in the field^4,97^. The search strategy was developed jointly and executed by an experienced information consultant. Then, two authors (NB and AL) performed study selection, data extraction, and synthesis, while having regular meetings with the complete author team.

### 5.1 Eligibility criteria for studies

In the following, we describe our eligibility criteria, which informed our search strategy and were systematically applied during study selection. Importantly, if a study contained multiple analyses of which only some fulfilled our eligibility criteria, we included the study but extracted only the eligible analyses. This may sometimes not correspond to the main analysis of a paper or may include more than one analysis per study.

#### Study design

In this review, we considered observational studies assessing the effects of non-pharmaceutical interventions on outcomes related to the COVID-19 disease. We focused on retrospective analyses that used real-world observational data to assess the effects of non-pharmaceutical interventions. Specifically, we excluded modeling studies that predominantly worked with synthetic data or projected future transmission dynamics based on hypothetical scenarios without assessing the effects of interventions empirically.

#### Population

We considered studies assessing the effects of non-pharmaceutical interventions on the population in one or several geographic regions. Our review was not limited to a specific geographic region, i. e. all national and subnational regions worldwide were considered. We furthermore included studies analyzing specific subpopulations in a certain region (e. g. certain age groups). We also considered analyses using individual-level data, as long as the intervention effect was assessed on a population level.

#### Outcome

The main outcomes considered were health-related outcomes at population level that are associated with COVID-19 (e. g. confirmed cases, hospitalizations, and deaths), and epidemiological outcomes characterizing infection dynamics such as reproduction numbers or transmission rates. We also considered similar outcomes associated with other infectious diseases (e. g. influenza), if used as a surrogate for COVID-19. Moreover, behavioral outcomes potentially mediating the effect of non-pharmaceutical interventions were included (e. g. human mobility). In contrast, we excluded analyses assessing the effects of non-pharmaceutical interventions solely on other outcomes not directly related to infectious diseases (e. g. psychological well-being or economic activities).

#### Intervention

As non-pharmaceutical interventions, we considered the implementation of health policy measures in the context of the COVID-19 pandemic. Specifically, we included any intervention related to social distancing (e. g. school closures, venue closures, workplace closures), containment (e. g. contact tracing, quarantining), population flow (e. g. border closures), or personal protection (e. g. facial mask mandates). Analyses were considered regardless of whether they assessed the effect of a single intervention, the effects of multiple interventions separately, or the effect of a combination of interventions. For simplicity, we refer to these as non-pharmaceutical interventions throughout the review, while recognizing that also other terms have been used in the literature. Importantly, we accounted for various alternative terms in our literature search (see Search strategy below). We excluded interventions not directly related to disease control (e. g. economic measures like social benefits).

### 5.2 Search strategy

Our search strategy was informed by our personal experience with studies in the field from our own primary research on the impact of non-pharmaceutical interventions. The search was supported by an experienced information consultant.

We searched for peer-reviewed original research articles in English language that were accepted, published, or in press between January 1, 2020 and January 12, 2021. In our review protocol, we specified that we would also include preprints in our search. However, due to their enormous volume, we eventually decided not to consider gray literature or preprints in our review. Our results therefore only cover methodologies used by articles peer-reviewed at the time of search, among which we already found considerable variation. To account for potentially new methodologies in articles published after the time of search, we also considered further recent studies on the effects of non-pharmaceutical interventions in our discussion and put them into the context of our review findings.

We searched the databases Embase, PubMed, Scopus, and Web of Science. These databases include, among others, MEDLINE, Biological Abstracts, CAB Abstracts, and Global Health. We composed our search query of four components to be contained in the publication title or abstract: (1) a synonym for “non-pharmaceutical intervention”, (2) a synonym for “estimation” or “assessment”, (3) a synonym for “effect”, and (4) a synonym for “COVID-19”. Starting from a precompiled list of 18 references based on our primary research on the effects of non-pharmaceutical interventions, we created and repeatedly extended a collection of synonyms for each of the above components, thereby achieving a broad search while keeping the number of selected studies manageable. The strings for our search queries are provided in SI Appendix B. Importantly, we decided not to include search terms for single interventions such as face masks or travel restrictions, as this would have resulted in an unmanageable number of studies that were not concerned with the population-level impact of the non-pharmaceutical intervention. Nevertheless, our search query found studies on single interventions through other terms describing non-pharmaceutical interventions.

### 5.3 Data collection and analysis

#### Study selection

As a first step, we screened the titles of the studies retrieved from the database search for keywords clearly suggesting that the study would not meet our predefined eligibility criteria (e. g. “mental health” or “air quality”). The compiled set of keywords (see SI Appendix B) was used to automatically identify cases for exclusion via the publication title. For all remaining studies, two authors (AL and NB) checked the eligibility criteria and individually decided on inclusion or exclusion. Each of the two authors checked the eligibility for one half of the studies via the following process: First, studies were checked by title and, if in doubt, by abstract. Then, if still in doubt, studies were checked by full text and discussed by both authors. Any disagreements were resolved with involvement of a third author (WV). Generally, we followed an inclusive approach by keeping all studies that could not be excluded with high confidence. At each stage, all decisions were recorded in a spreadsheet.

#### Data extraction

We extracted data from all included studies in a spreadsheet. Our extraction strategy reflected the exploratory nature of our analysis and thus allowed for new data items to be added throughout the process. Therefore, we maintained a detailed manual with all data items and the potential values for each item (see SI Appendix D). Before extraction, a preliminary version of the data extraction form was created based on reporting items from checklists for observational studies (STROBE^106^, RECORD^107^), a template for public health policy interventions (TIDieR-PHP^108^), and our personal experience with studies in the field from our own primary research on the impact of non-pharmaceutical interventions. Aside from bibliographic information, the data to be extracted consisted of information on the study setting, outcome, intervention, methodological approach, and effect assessment.

The extraction process was structured in four rounds. During the first round, two authors (AL and NB) extracted data from an initial set of 20 publications, blinded to each other’s coding. The coding was then compared, and any differences were discussed to resolve ambiguities. Corresponding changes were recorded by updating the extraction form and manual, and applied subsequently. In the second round, the two reviewers each extracted data from one half of the remaining publications and checked the other half coded by their colleague. Color-coding was used to highlight uncertain or ambiguous entries for the other reviewer or to mark such entries for further discussion. Regular meetings were held between the two authors (AL and NB) to discuss these uncertainties and ambiguities. All disagreements were resolved through discussion, if needed by involving a third author (WV). Thereby, the data extraction manual and form were continuously refined and kept up-to-date. In particular, the list of values that could be potentially assigned to each data item was continuously extended and harmonized as new studies were extracted. In the third round, the data extraction form and manual were simplified by merging data items or categories that, retrospectively, were found redundant, or by relabeling items and categories to define them more precisely. This was done with particular attention to enable comparability among the extracted analyses as well as readability of the results. In the fourth round, the final scheme was applied to all studies.

#### Quality assessment

The goal of this systematic review was not to perform a meta-analysis or narrative synthesis of the effects of non-pharmaceutical interventions, but to compare the included studies along methodological dimensions and to analyze the variation in study setting, outcome, intervention, methodological approach, and effect assessment. Therefore, no risk of bias assessment with regard to the study results was conducted. Our minimum requirement for quality was that most information on the aforementioned dimensions could be extracted from the manuscript and/or supplementary material. This minimum requirement was not met by four studies, which were thus excluded. For other studies where only some methodological information was missing, we noted in the data extraction sheet that this information “could not be evaluated”.

#### Data synthesis

The results of the data extraction were synthesized in tabular form by recording the frequency of categories per item. We reported the frequency for each item of the main dimensions (study setting, outcome, intervention, methodological approach, and effect assessment) individually. For some items, we conducted further specialized analyses, for example by computing the frequencies of categories for different methodologies separately, or by qualitatively evaluating the supplementary information added to certain entries during extraction. Insights from these additional analyses were reported textually. Furthermore, we synthesized common analysis types based on patterns identified in the methodological approaches. Lastly, based on our findings, we derived specific recommendations for future studies with regard to scope, robustness, and comparability, and put them into the context of more recent studies that were not part of our review sample.

## Data Availability

The full data extracted from the studies in this review is available in machine-readable format at https://github.com/adrian-lison/methodologies-npi-effects.

## Declarations

### Ethics approval

Ethics approval was not required for this study.

## Funding

NB and SF acknowledge funding from the Swiss National Science Foundation (SNSF) as part of the Eccellenza grant 186932 on “Data-driven health management”. The funding bodies had no control over design, conduct, data, analysis, review, reporting, or interpretation of the research conducted.

### Contributions

NB and AL contributed to conceptualization, literature search, study selection, data extraction, data synthesis, and manuscript writing, review & editing. DO contributed to literature search and manuscript review & and editing. TS contributed to manuscript review & editing. SF contributed to manuscript writing, review & editing. WV contributed to conceptualization, study selection, data extraction, data synthesis, manuscript writing, review & editing.

