## Supplementary material for "The methodologies to assess the effects of non-pharmaceutical interventions during COVID-19: a systematic review": SI appendix

#### Appendices

**A Remarks on analyses inferring transmission dynamics** **2**

**B Literature Search** **3**

**C Detailed breakdown of methods by empirical approach** **8**

**D Data extraction manual** **9**

**E Data extraction report** **25**

**F Excluded studies** **618**

**G PRISMA checklist** **660**

**References** **664**

### A Remarks on analyses inferring transmission dynamics

Almost all of the analyses in our review sample that used an unobserved outcome representing transmission dynamics (e. g. the reproduction number or transmission rate) inferred it from one or several observed outcomes (e. g. cases or deaths). For this, non-parametric methods or transmission models were typically used; however, these methods were not always correctly applied.

First, many analyses partially or completely neglected the stochastic time lags in the process from infection to reporting, including disease progression (time from infection to symptom onset, hospitalization, or death) and delays in ascertainment (testing, forwarding to health authority). That is, to estimate the reproduction number or transmission rate, non-parametric methods were often directly applied to the time series of confirmed cases, hospitalizations, or deaths, without accounting for the delays between infection and reporting (e. g.[1–3]). Similarly, analyses using transmission models sometimes equated confirmed cases with infections (e. g.[4–6]). This can lead to overconfident estimates of the reproduction number or transmission rate that may be both shifted in time (e. g. by the mean incubation period and reporting delay) and blurred (e. g. if reports of cases infected on a given day are spread over successive days)[7].

Second, authors must be aware that there exist different versions of the reproduction number $R$, which have been developed for particular use cases and require different interpretations. Aside from the method by Cori et al.[8], some authors also used other approaches for estimation of the instantaneous reproduction number[9,10], basic reproduction number[11,12], or case reproduction number[13]. However, some analyses did not use methods suitable for the research question at hand or the data analyzed. For example, the methods by White et al.[11] or Wallinga and Lipsitch[12] were designed for estimating the basic reproduction number during the early period of exponential growth in an epidemic, but some analyses applied them also to the time after interventions were implemented[14–16], which was typically a period of subexponential growth. Similarly, the case reproduction number as estimated with the method by Wallinga and Teunis[13] is a forward-looking reproduction number that does not describe transmission at one specific point in time, and is thus not ideal for assessing the population-level effect of an intervention on a specific date.

Overall, differences in the methods used for inferring transmission dynamics could, to some extent, also explain different findings regarding the effectiveness of non-pharmaceutical interventions.

### B Literature Search

#### Database search queries

We composed our search queries of four components to be contained in the publication title or abstract: (1) a synonym for "non-pharmaceutical intervention", (2) a synonym for "estimation" or "assessment", (3) a synonym for "effect", and (4) a synonym for "COVID-19". In the following, we provide the detailed string of our search query for each database searched, i. e. Embase, PubMed, Scopus, and Web of Science. The differences in query strings between databases are mostly of syntactical nature and result from the specific conventions of each search interface.

***Embase***

```
SEARCH QUERY:=
      (‘non pharmac*’ OR ‘non-pharmac*’ OR ‘nonpharmac*’ OR ‘anti contagion’
      OR ‘anti-contagion’ OR ‘anticontagion’ OR ‘distancing’ OR ‘contact
      restriction*’ OR ‘containment strateg*’ OR ‘containment measure*’ OR
      ‘containment polic*’ OR ‘confinement strateg*’ OR ‘confinement measure*’ OR
      ‘confinement polic*’ OR ‘quarantine strateg*’ OR ‘quarantine measure*’ OR
      ‘quarantine polic*’ OR ‘control strateg*’ OR ‘control measure*’ OR ‘control
      polic*’ OR ‘mitigation measure*’ OR ‘mitigation strateg*’ OR ‘mitigation
      polic*’ OR ‘government action*’ OR ‘government measure*’ OR ‘government
      response*’ OR ‘government intervention*’ OR ‘public health action*’ OR
      ‘public health measure*’ OR ‘public health response*’ OR ‘public health
      polic*’ OR ‘public health intervention*’)
   AND
      (estimat* OR evaluat* OR assess* OR quantify* OR calculat* OR infer* OR
      rank* OR explain*)
   AND
      (effect* OR effic* OR impact* OR associ* OR link* OR relat* OR chang*)
   AND
      ’coronavirus infection’/exp (#ncov* already included)
```

***PubMed***

SEARCH QUERY:=

((non pharmac*[Title/Abstract]) OR (nonpharmac*[Title/Abstract]) OR (non-pharmac*[Title/Abstract]) OR (anti contagion[Title/Abstract]) OR (anticontagion[Title/Abstract]) OR (anti-contagion[Title/Abstract]) OR (distancing[Title/Abstract]) OR (contact restriction*[Title/Abstract]) OR (containment strateg*[Title/Abstract]) OR (containment measure*[Title/Abstract]) OR (containment polic*[Title/Abstract]) OR (confinement strateg*[Title/Abstract]) OR (confinement measure*[Title/Abstract]) OR (confinement polic*[Title/Abstract]) OR (quarantine strateg*[Title/Abstract]) OR (quarantine measure*[Title/Abstract]) OR (quarantine polic*[Title/Abstract]) OR (control strateg*[Title/Abstract]) OR (control measure*[Title/Abstract]) OR (control polic*[Title/Abstract]) OR (mitigation measure*[Title/Abstract]) OR (mitigation strateg*[Title/Abstract]) OR (mitigation polic*[Title/Abstract]) OR (government action*[Title/Abstract]) OR (government measure*[Title/Abstract]) OR (government response*[Title/Abstract]) OR (government intervention*[Title/Abstract]) OR (public health action*[Title/Abstract]) OR (public health measure*[Title/Abstract]) OR (public health response*[Title/Abstract]) OR (public health polic*[Title/Abstract]) OR (public health intervention*[Title/Abstract]))

AND

((estimat*[Title/Abstract]) OR (evaluat*[Title/Abstract]) OR (assess*[Title/Abstract]) OR (quantify*[Title/Abstract]) OR (calculat*[Title/Abstract]) OR (infer*[Title/Abstract]) OR (rank*[Title/Abstract]) OR (explain*[Title/Abstract]))

AND

((effect*[Title/Abstract]) OR (effic*[Title/Abstract]) OR (impact*[Title/Abstract]) OR (associ*[Title/Abstract]) OR

```
    (link*[Title/Abstract]) OR (relat*[Title/Abstract]) OR
    (chang*[Title/Abstract]))
  AND
    (("SARS-CoV-2"[Mesh]) OR ("COVID-19"[Mesh]))
```

***Scopus***

```
SEARCH QUERY:=
  TITLE-ABS(
    ((non W/0 pharmac*) OR (non-pharmac*) OR (nonpharmac*) OR (anti W/0
    contagion) OR (anti-contagion) OR (anticontagion) OR (distancing) OR
    (contact W/0 restriction*) OR (containment W/0 strateg*) OR (containment
    W/0 measure*) OR (containment W/0 polic*) OR (confinement W/0 strateg*)
    OR (confinement W/0 measure*) OR (confinement W/0 polic*) OR (quarantine
    W/0 strateg*) OR (quarantine W/0 measure*) OR (quarantine W/0 polic*) OR
    (control W/0 strateg*) OR (control W/0 measure*) OR (control W/0 polic*)
    OR (mitigation W/0 measure*) OR (mitigation W/0 strateg*) OR (mitigation
    W/0 polic*) OR (government W/0 action*) OR (government W/0 measure*) OR
    (government W/0 response*) OR (government W/0 intervention*) OR (public
    W/0 health W/0 action*) OR (public W/0 health W/0 measure*) OR (public
    W/0 health W/0 response*) OR (public W/0 health W/0 polic*) OR (public w/0
    health w/0 intervention*))
  AND
    ((estimat*) OR (evaluat*) OR (assess*) OR (calculat*) OR (infer*) OR (rank*)
    OR (explain*))
  AND
    ((effect*) OR (effic*) OR (impact*) OR (associ*) OR (link*) OR (relat*) OR
    (chang*))
  AND
    ((ncov*) OR (covid*) OR (sars*) OR (coronavir*) OR (corona W/0 vir*))
  )
```

***Web of Science***

We independently searched by TI and AB (title and abstract) for non-pharmaceutical interventions, and then combined both searches using the Boolean operator OR.

```
SEARCH QUERY:=
      AB/TI=(‘non pharmac*’ OR ‘non-pharmac*’ OR ‘nonpharmac*’ OR ‘anti contagion’
      OR ‘anti-contagion’ OR ‘anticontagion’ OR ‘distancing’ OR ‘contact
      restriction*’ OR ‘containment strateg*’ OR ‘containment measure*’ OR
      ‘containment polic*’ OR ‘confinement strateg*’ OR ‘confinement measure*’ OR
      ‘confinement polic*’ OR ‘quarantine strateg*’ OR ‘quarantine measure*’ OR
      ‘quarantine polic*’ OR ‘control strateg*’ OR ‘control measure*’ OR ‘control
      polic*’ OR ‘mitigation measure*’ OR ‘mitigation strateg*’ OR ‘mitigation
      polic*’ OR ‘government action*’ OR ‘government measure*’ OR ‘government
      response*’ OR ‘government intervention*’ OR ‘public health action*’ OR
      ‘public health measure*’ OR ‘public health response*’ OR ‘public health
      polic*’ OR ‘public health intervention*’)
   AND
      ALL=(estimat* OR evaluat* OR assess* OR quantify* OR calculat* OR infer* OR
      rank* OR explain*)
   AND
      ALL=(effect* OR effic* OR impact* OR associ* OR link* OR relat* OR chang*)
   AND
      ALL=(ncov* OR covid* OR sars* OR corona$vir*)
```

#### Initial filtering of studies based on keywords

We screened the titles of the studies retrieved from the database search for keywords clearly suggesting that the study would not meet our predefined eligibility criteria. The following set of keywords was compiled:

`air, alcohol, attitudes, clinical, consumption, depression, distress, eating, education, environmental, experience, food, habits, knowledge, medical, mental, nursing, orthopaedic, perceptions, pollution, psychological, psychosocial, quality, remote, services, sleep, students, surgery, telehealth, telemedicine, training, transplant, trauma, virtual, wellbeing, workers`

The above keywords were thoroughly checked in order to minimize the risk of erroneous exclusion of studies. The compiled set of keywords was then used to identify cases for exclusion via the publication title. That is, studies containing one or more exclusion keywords in the publication title (abstract or full text not considered) were excluded.

### C Detailed breakdown of methods by empirical approach

For better readability of the manuscript, the frequencies of methods at the lower level were not broken down by empirical approach to ensure conciseness. The detailed breakdown of the methods by empirical approach is shown in Table S1.

**Table S1.** Detailed breakdown of method frequencies by empirical approach.

| **Method** | **(D)** | **(P)** | **(C)** | Total freq. |
|---|---|---|---|---|
| Description of change over time | 136 | — | — | 136 (48 %) |
| ↳ Description of time course | ↳ 49 | ↳ — | ↳ — | ↳ 49 (36 %) |
| ↳ Comparison of time periods | ↳ 87 | ↳ — | ↳ — | ↳ 87 (64 %) |
| Comparison of populations | 8 | — | — | 8 ( 3 %) |
| Comparison of change points with intervention dates | 7 | — | — | 7 ( 2 %) |
| Non-mechanistic model | — | 61 | 17 | 78 (27 %) |
| ↳ Generalized linear model | ↳ — | ↳ 49 | ↳ 2 | ↳ 51 (65 %) |
| ↳ Exponential growth model | ↳ — | ↳ 0 | ↳ 11 | ↳ 11 (14 %) |
| ↳ Other | ↳ — | ↳ 12 | ↳ 4 | ↳ 16 (21 %) |
| Mechanistic model | — | 30 | 13 | 43 (15 %) |
| ↳ Compartmental single-population transmission model | ↳ — | ↳ 21 | ↳ 8 | ↳ 29 (67 %) |
| ↳ Compartmental meta-population transmission model | ↳ — | ↳ 1 | ↳ 3 | ↳ 4 ( 9 %) |
| ↳ Semi-mechanistic Bayesian transmission model | ↳ — | ↳ 5 | ↳ 0 | ↳ 5 (12 %) |
| ↳ Other | ↳ — | ↳ 3 | ↳ 2 | ↳ 5 (12 %) |
| Synthetic controls | — | — | 6 | 6 ( 2 %) |
| Other | 0 | 3 | 4 | 7 ( 2 %) |

*Empirical approach*: **(D)** descriptive, **(P)** parametric, and **(C)** counterfactual

### D Data extraction manual

The following data extraction manual describes the items that we extracted for each dimension. The data extraction manual was used throughout the data extraction process.

##### *Summary of the data extraction process*

The data extraction manual was developed along with the data collection and analysis process. We started with a pre-defined list of items. For each item, we built a pre-defined set of categories from the first 20 studies that we extracted. Later, if necessary, more categories were added during extraction. Some items were not categorized (see, for example, Items D.3.1 and D.3.2), and some items were merged or removed if they became redundant (i. e. if the entries from one item could easily be determined from the entries of other items). The final list of items is shown below in the table of contents.

##### *Studies with multiple entries*

Some studies assessed the effects of multiple types of non-pharmaceutical interventions with different methodological approaches. As such, these studies can have multiple entries, that is, one for each analysis. No additional entry was added if only different outcomes were analyzed but with respect to the same interventions and methodological approaches. Different outcomes were separated by comma.

##### *Reporting*

In the data extraction report, there is one row for each item. Multiple values are separated by comma (see for example an entry with values "`cases, deaths, recovered`" for Item D.2.1). Specific information for the entered data are given in parentheses, and further details are written after a colon, e. g. "`behavioral outcome (mobility: population outflow from Wuhan)`".

---

### D.0 Study information

##### D.0.1 Author

The author(s) of the study.

##### D.0.2 Title

The title of the study.

##### D.0.3 Year

The year the study was published.

##### D.0.4 Journal

The journal where the study was published.

##### D.0.5 DOI

The digital object identifier (DOI).

##### D.0.6 Date received

The date the study was received.

##### D.0.7 Date accepted

The date the study was accepted. If not available, then the date when the paper was first published in print or online (depending on what came first), as noted in parentheses.

### D.1 Study setting

##### D.1.1 Number of populations included

Whether a single or multiple populations (countries, states, cities) are analyzed.

- **Single**: A single population, e. g. a single country or subnational region is analyzed.
- **Multiple**: Multiple populations are analyzed, e. g. multiple countries or multiple subnational regions.

##### D.1.2 Level of populations included

Whether populations are studied at the national or a subnational level. Can be both, e. g. if a country as a whole and its subnational regions are studied. The specific population is given in parentheses.

- **National**: Populations are studied at the national level, i. e. countries.
- **Subnational**: Populations are studied at the subnational level, e. g. state or city.

##### D.1.3 Geographic areas covered

The geographic areas of the populations that are studied.

- **Asia**
- **Europe**
- **North America**
- **Middle East & Africa**
- **Central & South America**
- **Oceania**

##### D.1.4 Number of countries covered

Whether the analysis covers populations from multiple countries or from a single country (e. g. China, United States, India, Italy, etc).

- **Multiple countries**: Populations from multiple countries, i. e. multiple countries or subnational regions from multiple countries.
- **Single country**: Populations from a single country, i. e. a single country or subnational regions from a single country. If applicable, then the name of that country.

##### D.1.5 Study period

Any specifics regarding the start or end dates of the study period in relation to the course of the epidemic in which the intervention(s) were assessed. We checked if analyses covered both a rise and decline in cases to assess the effects of interventions. Any observed deviations from this setup are noted. If the raw outcome is behavioral, then only deviations of the study period from the timing of interventions are noted. As the vast majority of studies analyzed the first wave in the respective population, it is also noted when a study included data from a subsequent wave.

- **Nothing specific (Start and end date span first epidemic wave)**: Study period covers rise and decline in new cases (with respect to confirmed cases as reported by "Our World in Data"[17]) during the first epidemic wave in the analyzed population, and starts before and ends after the analyzed interventions were implemented.
- **Late start**: The study period starts comparatively late for one or several of the populations analyzed, either with respect to the wave (e. g. only after the peak) or with respect to the intervention (e. g. only after the intervention was implemented).
- **End date in growth phase of wave**: The study period has a comparatively early end date compared to the epidemic wave in one or several of the populations analyzed, i. e. cases are still clearly rising.
- **End date at peak of wave**: The study period ends (roughly) at the peak of the first wave in one or several of the populations analyzed, i. e. new cases are stabilizing or start declining.

- **Same end date for several populations with diverse epidemic trajectories**: Several populations are included in the analysis, and, during the study period, the course of the epidemic varies significantly across the populations. Typically, this would mean that the end date of the study period was still within the epidemic growth phase for some populations and already in the control phase for other populations.
- **Start date could not be evaluated**: The start of the study period could not be evaluated.
- **End date could not be evaluated**: The end of the study period could not be evaluated.
- **Includes second wave**: Study includes data from a second wave in one or several of the populations analyzed. This can either mean that the study analyses several waves (separately or jointly) or only focuses on the second wave. Details noted in parentheses.
- **Other**: Anything else noteworthy about the study period, details noted in parentheses.

#### D.2 Outcome

During the data extraction process, we observed that some analyses directly used the raw data from the outcome to assess the effect of an intervention, while other analyses involved an intermediate step in which an unobserved outcome was computed from the raw outcome. In the latter case, the computed outcome was then analyzed instead of the raw outcome, or sometimes in addition to it. Therefore, we subsequently distinguish between the raw outcome (Item D.2.1) and the computed outcome (Item D.2.3).

##### *D.2.1 Raw outcome*

The raw outcome from the data that was available to the study.

- **Epidemiological population-level outcome**: Cases (documented number of cumulative or new cases), deaths, recovered cases, hospitalizations, intensive care unit (ICU) hospitalizations, ICU beds occupation, emergency room visits, severe cases (people severely ill from COVID-19), active cases (number of active cases as obtained from external data), basic or effective reproduction number (as obtained from external data), exported cases (cases exported from one population to another, as obtained from external data), imported cases (cases imported from one population to another, as obtained from external data), tests (number of COVID-19 tests conducted, or number of positive tests), test positivity rate (ratio of positive tests to total tests), date of first reported case in a country, or surrogate disease (a similar epidemiological outcome as for COVID-19, but for a different disease, e. g. cases of influenza, with exact outcome and disease noted in parentheses).
- **Epidemiological individual-level outcome**: (1) Individual cases, i. e. cases with case ID, demographics, clinical characteristics, and epidemiological characteristics (e. g. the date of symptom onset, travel history, potentially close contacts), which can used to estimate the serial interval; (2) individual cases and transmission chains, i. e. cases with case ID and the link between index and secondary case that allow inference of transmission chains, in order to compute the analyzed outcome (e. g. estimating the serial interval and/or the reproduction number); or (3) genome sequence data, i. e. genome sequence data of clinical SARS-CoV-2 samples.

- **Behavioral outcome**: (1) Mobility (mobility data, e. g. time spent in various locations or distance travelled) or (2) survey responses (survey response data, e. g. on social distancing or hygiene behavior).

##### D.2.2 Time resolution of raw outcome

Time resolution of data for the raw outcome (Item D.2.1), e. g. daily, weekly, monthly, etc.

##### D.2.3 Computed outcome

Outcomes computed in an intermediate step from a raw outcome and then used in the main effect assessment. Detailed computed outcome in parentheses.

- **None**: No intermediate step, i. e. the raw outcome is used in the main assessment.
- **Measure of epidemic trend**: Doubling time, growth rate, or other. Usually all of these measures are derived from confirmed cases. If otherwise, the respective outcome is noted after a colon in the data extraction report.
- **Epidemiological parameter**: Reproduction number (basic or effective reproduction number, which is usually estimated from the confirmed number of cases; if not, the respective outcome is noted after a colon)*, transmission rate (also known as effective contact rate), contact rate, probability of transmission upon contact, hospitalization rate, recovery rate, death rate, case fatality rate, serial interval, time from symptom onset to testing or to isolation, length of infectiousness period, or other (details noted after a colon in the data extraction report).
- **Summary statistic**: Any statistic summarizing the state, course, or severity of the epidemic in a population. Examples are the time until a certain number of cases are reached, the time until the peak of the epidemic is reached, or the time until the reproduction number fell below 1.
- **Change points**: Points in time where there is a structural change in the outcome, estimated using a change point detection method. The respective outcome used for change point detection is noted after a colon.
- **Other**: Any other computed outcome which does not fit into the above categories. Details noted in parentheses.

##### D.2.4 Method to obtain the computed outcome

If applicable, the method which is used to obtain the computed outcome (Item D.2.3). If different methods are used for different computed outcomes, the corresponding outcome is noted in parentheses.

- **Not applicable**: No computed outcome; instead, the raw outcome is used for analysis.
- **Simple computation**: Simple computation, e. g. computing the growth in cases between two consecutive time periods.
- **Exponential growth model**: Fitting of an exponential growth curve to the raw outcome (e. g. confirmed cases) in order to estimate epidemiological parameters such as the exponential growth rate or doubling time.

*Note that the reproduction number is also often used as a measure of epidemic trend (e. g. a value of $R > 1$ is interpreted as exponential spreading). We have classified it as epidemiological parameter here because of its foundation in infectious disease modeling and its frequent use in mechanistic models.

- **Compartmental transmission model**: (1) Compartmental single-population transmission model (no spatial structure) or (2) compartmental meta-population transmission model (several spatially distinguished meta-populations which interact to some degree, e. g. by infection across meta-populations and/or migration between meta-populations). Meta-population models usually imply a network structure between populations, and mobility data are often used to inform the interaction parameters.
- **Individual-based transmission model**: Model for transmission dynamics at the individual level, oftentimes with a network structure or via simulation of individual behavior and contact patterns.
- **Phylodynamic model**: Phylogenetic model with population dynamics that is fitted using genome sequence data.
- **Semi-mechanistic Bayesian transmission model**: Bayesian hierarchical model with a time-discrete, stochastic renewal process of transmission.
- **Statistical estimation of reproduction number**: Estimation of the reproduction number based on the time series of confirmed cases, deaths, hospitalized, etc. The exact method used to estimate the reproduction number is noted in parentheses: (1) Cori et al.[8] (estimates the instantaneous effective reproduction number); (2) Thompson et al.[10] (extension of the method by Cori et al., so that it integrates data on known pairs of index and secondary cases to directly estimate the serial interval); (3) an der Heiden and Hamouda[9] (simplification of the method by Cori et al., assumes a fixed serial interval); (4) Wallinga and Teunis[13] (estimates the case/cohort effective reproductive number, i. e. the expected number of secondary infections that will be caused by an individual infected at time $t$); (5) White et al.[11] (maximum-likelihood based method for estimating the basic reproduction number); or (6) Wallinga and Lipsitch[12] (uses the exponential growth rate for estimating the basic reproduction number).
- **Statistical estimation of reproduction number from individual data**: Estimation of $R$ based on individual case data with information to infer transmission chains. The exact method used to estimate the reproduction number from individual case data is noted in parentheses: (1) Wallinga and Teunis[13] (same method as above, but using individual data); (2) Lloyd and Smith[18] (estimates the reproduction number as the population-average of individual reproduction numbers); or (3) Diekmann et al.[19] (estimates $R$ from the dominant eigenvalue of a next generation matrix).
- **Back projection**: Backward computation of a time series of cases by one epidemiological event (e. g. infection) from a time series of cases by another event (e. g. confirmation) using an empirical distribution of the delay between the two events.
- **Change point analysis**: Change point detection method applied to the raw outcome.
- **Other**: Other less frequent approaches that are used to estimate or compute the analyzed outcome. Details given in parentheses.

##### D.2.5 Data source

A categorization of the data source for the raw outcome (Item D.2.1). The specific source is given in parentheses. Note that, for (sub)national data, this is usually a governmental health authority, e. g. the department or ministry of health, the center for disease control, the health commission, or a committee. The terminology varies; e. g. sometimes the name of the authority is translated, sometimes the higher- or lower-level authority name is provided, or sometimes only the name of the database hosted by the authority is provided. Therefore, all kinds of governmental health authorities and their databases are summarized under "health authority".

- **Could not be evaluated**: Data source for the outcome not mentioned in the manuscript.
- **Data from (sub)national authorities**: The data are accessed from a (sub)national authority, e. g. the national center for disease control.
- **Data from publicly available cross-country selections**: Examples of publicly available cross-country selections include (1) the John Hopkins University (JHU)[20]; (2) the European Center for Disease Prevention and Control (ECDC)[21]; (3) Our World In Data[17], (4) the World Health Organization (WHO)[22]; and (5) Worldometer[23].
- **Mobility data from corporate organizations**: Private/corporate organization sharing mobility data, e. g. Google COVID-19 Community Mobility Reports[24], Apple COVID-19 Mobility Trends Reports[25], or SafeGraph Places Data and Foot Traffic Insight[26].
- **Data from (sub)national public organizations**: The data are accessed directly from a (sub)national public (or not-for-profit) organization, e. g. the national statistics institute or transport organization.
- **Data from public media outlets**: Data from media outlets, e. g. the New York Times.
- **Data from other research projects**: Data from research projects, e. g. a published study or preprint.
- **Data from publicly available open-source projects**: Open-source projects where volunteers collected data from government and/or news websites and/or other sources, which they share via a publicly available repository.
- **Data collected by authors**: The study authors collected the data themselves, e. g. behavioral outcomes via surveys, seroprevalence studies, or data collected at local facilities, e. g. emergency room visits at hospitals.

##### D.2.6 Data availability

Data availability for the raw outcome.

- **Data not accessible**: Data are not available, i.e. no access is provided by the authors and the source of the data is not documented.
- **Data access via source**: Data accessible from original source. Of note, this does not imply that the data are easily accessible or that access is granted. In particular, we labeled raw data on epidemiological individual-level outcomes as *not accessible*, unless the study authors provided a link to the exact source of their raw data in the manuscript.

- **Data made available by the authors**: Authors make the data available, e.g. by depositing the data they used for analysis into a publicly accessible repository.

#### D.3 Intervention

##### D.3.1 Terminology for interventions

The term that is predominantly used by the authors to refer to "interventions".

- **Not applicable**: Only a term for the specific type of non-pharmaceutical interventions is used.
- **Measures**
- **Interventions**
- **Policies**
- **Strategies**
- **Orders**
- **Actions**

##### D.3.2 Terminology for the specific type of non-pharmaceutical interventions

The term that is predominantly used by the authors to refer to the specific type of non-pharmaceutical interventions.

- **Not applicable**: Only a term for "interventions" is used.
- **Non-pharmaceutical**
- **Social distancing**
- **Control**
- **Public health**
- **Containment**
- **Other**: Other less frequently used terms (e. g. "mitigation", "preventive", "government") or terms describing particular non-pharmaceutical interventions (e. g. "stay-at-home orders" and "travel restrictions").

##### D.3.3 Exposure types

Categorization of exposure types. The specific interventions are given in parentheses, e. g. "school closure" or "confinement of the population by decree of the Spanish Government on March 14". Variations of the "Wuhan City Lockdown" or the "Indian National Lockdown" occurred frequently and are thus consistently labeled.

- **One single intervention**: One single intervention implemented on a specific date. Example: school closures implemented on March 15.
- **Multiple separate interventions**: Multiple separate interventions implemented at different dates. Example: school closures implemented on March 15 and stay-at-home orders implemented on April 1.

- **One combination of interventions**: One combination of multiple interventions implemented at the same date. Example: a lockdown on April 1 which included the closure of schools, the closure of all non-essential businesses, and a stay-at-home order.
- **Multiple combinations of interventions**: Multiple combinations of single interventions implemented at different dates, e. g. school closures and gathering bans implemented on March 15 as well as workplace closures and stay-at-home orders implemented on April 1. Of note, this category applies if there is at least one combination of multiple interventions. Example: school closures on March 15 and lockdown including workplace closures and stay-at-home orders on April 1.
- **All interventions together**: No specific interventions are analyzed but rather whether all interventions together were effective. As a result, one observes whether transmission was reduced over time as interventions were implemented.
- **Phases of interventions**: The public health response is divided into phases of interventions and the effects of interventions are assessed within the phases rather than assessing the effects of specific interventions implemented at a specific date. Example: the public health response is divided into two phases, namely (1) from March 15 and April 1 where school closures, bans of large gatherings, and venue closures were implemented, and (2) from April 2 to April 15 where workplace closures and stay-at-home orders were implemented.
- **Start of interventions**: Usually represented as the first date any intervention was implemented.
- **Categories of interventions**: Categorizing interventions into groups, e. g. quarantine and containment measures.
- **Stringency index**: Summarizing the stringency of interventions into a numerical index, e. g. the stringency index from the Oxford Covid-19 Government Response Tracker[27]

##### D.3.4 Types of single interventions

When single interventions are analyzed, the following interventions are consistently labeled:

- **Stay-at-home order**: Order to stay at home and otherwise confinement of the population, i. e. being allowed to leave the house only with few exceptions, such as going shopping or to the pharmacy.
- **School closure**: Closure of primary or secondary schools, or universities.
- **Workplace closure**: Closure of workplaces, i. e. closure of (most) non-essential businesses.
- **Venue closure**: Closure of venues, e. g. bars, restaurants, theaters, religious places, etc.
- **Declaration of a state of emergency**: Declaration of a state of emergency. Of note, in some cases, it could not be evaluated based on the manuscript if the emergency declaration was just a declaration or if it involved additional interventions.
- **Bans of large gatherings**: Ban of large gatherings of people, often the cancellation of large public events involving hundreds or thousands of people.

- **Bans of small gatherings**: Ban of small gatherings of people, often the cancellation of small private gatherings involving only a dozen of people.
- **Internal movement restrictions**: Restrictions on internal movement between subnational regions.
- **Public transport closure**: Closure of public transport.
- **International travel restrictions**: Restrictions on free international travel, i. e. restricting travel from or to some or all international destinations.
- **Mask mandate**: Compulsory order to wear masks in public indoor settings.

###### D.3.5 Coding of interventions

Whether coding of intervention data is necessary, i. e. the interventions and dates need to be categorized in a systematic way so that the interventions are comparable across multiple populations. For instance, one country has closed secondary schools while another has closed both primary and secondary schools. To jointly assess the effects, the data are reconciled by coding school closures as the closure of primary schools. Coding of data is usually necessary when multiple populations are analyzed jointly, whereas the data are usually not further reconciled when a few populations and specific interventions are analyzed separately.

- **Not applicable**: No specific interventions are analyzed.
- **Not necessary**: Coding of interventions is not necessary because the data are not analyzed jointly, i. e. interventions remain specific to the populations and no attempts are made to compare interventions between multiple populations by analyzing them jointly.
- **Necessary**: Coding of interventions is necessary because the data are analyzed jointly, i. e. interventions need to be coded so that they can be compared across multiple populations.

###### D.3.6 Source of intervention data

Source of intervention data. Specific source is provided in parentheses.

- **Not applicable**: No specific interventions are analyzed.
- **Could not be evaluated**: Source could not be evaluated.
- **Government or news websites**: Date and type of interventions for raw (non-coded) data collected from government or news websites.
- **Another study**: Date and type of interventions for raw (non-coded) data collected from another study.
- **Coding done by the authors**: Coding of interventions done by the authors based on data collected from government or news websites.
- **Use of externally coded data**: Coded intervention data are obtained from an external provider or another published study or preprint.

##### D.3.7 *Availability of data on exposure*

Statement regarding data availability for the exposure data.

- **Not applicable**: No specific interventions are analyzed.
- **Raw data documented in the manuscript**: Coding of data is not necessary, and the raw data are documented in the manuscript (i. e. exposure type and intervention dates).
- **Access to externally coded data via source**: Coding of data is necessary, externally coded data are used, and it is stated in the manuscript where the data can be accessed.
- **Coded data made available by the authors**: Coding of data is necessary, coding is done by the authors or externally coded data are used, and the data are made publicly available by the authors, e. g. by depositing the data into a public repository or providing a structured table of the coded data in the manuscript or supplements.
- **Coded data not available**: Coding of data is necessary, but the data are not available; i. e. the data source could not be evaluated, and the data are not made available by the authors.

#### D.4 Methodological approach

The methodological approach describes the stage of effect assessment. Possible intermediate steps in the analysis, which may also involve substantial methodology, are described in Section D.2.

##### D.4.1 *Empirical approach*

The general empirical approach used to assess the effects of interventions on outcomes.

- **Descriptive**: The effect of an intervention is assessed descriptively without explicit formulation of an outcome-intervention relationship within a mathematical model. Nevertheless, mechanistic or non-mechanistic modeling may be involved in other parts of the analysis, e. g. for computing an unobserved outcome (see Section D.2).
- **Parametric**: The effect of an intervention is represented by a parameter in a mechanistic or non-mechanistic model. By fitting the model to data and estimating the parameter, the effect of the intervention is quantified.
- **Counterfactual**: A counterfactual outcome representing an explicit scenario in which the intervention was not implemented is computed. This is either done by projecting the outcome from the pre-intervention period or by using outcomes in other populations. The effect of the intervention is then assessed by comparing the observed outcome with the counterfactual outcome during the post-intervention period.

##### D.4.2 *Use of exposure variation*

The variation in exposure to interventions that is exploited to assess the effects of interventions.

- **Only variation over time for a single population**: A single population is analyzed. Here, only variation in the exposure over time can be exploited to assess the effects of interventions.
- **Only variation over time for multiple populations**: Multiple populations are analyzed, but only the variation over time is exploited, i. e. the effects of interventions are assessed within each population separately.

- **Only variation between populations**: Multiple populations are analyzed, but only the variation between populations is exploited, not the variation in each population over time.
- **Both variation over time and between populations**: Multiple populations are analyzed and both variation over time and between populations are exploited.

##### D.4.3 Method

A categorization of the method used to assess the effects of the intervention on the outcome.

- **Description of change over time**: (1) Description of time course (reported outcome or changes in the reported outcome are qualitatively described over time, sometimes by providing values for the outcome at representative time points), or (2) comparison of time periods (comparison of the outcome along time periods of different interventions, typically before and after an intervention is implemented).
- **Comparison of populations**: Comparison of populations subject to different interventions, usually comparing a population subject to an intervention with a population not subject to this intervention.
- **Comparison of change points with intervention dates**: Comparison of inferred change points in the outcome with observed dates of the interventions. If both time points are concurrent, this would be seen as evidence for an effect of the intervention.
- **Non-mechanistic model**: Non-mechanistic model with a direct relationship between intervention and outcome. For example, the outcome may be an epidemiological variable such as the number of new cases, and the intervention is represented by a binary variable that equals one if the intervention is implemented and zero otherwise. Can either be (1) exponential growth model (fitting of an exponential growth equation); (2) time series model (time series model such as AR(I)MA, exponential smoothing etc.); (3) generalized linear (regression) model (including ordinary linear regression); (4) correlation (correlation coefficient computed between outcome and intervention, where intervention is usually a continuous variable representing a social distancing index); or (5) other.
- **Mechanistic model**: A model that has a structure that makes, to some extent, explicit assumptions about the mechanisms that drive infection dynamics. Can be either (1) compartmental single-population transmission model (single population, i. e. no spatial structure); (2) compartmental meta-population transmission model (several spatially distinguished meta-populations which interact to some degree, e. g. by infection across meta-populations and/or migration between meta-populations; this typically has a network structure between meta-populations, for which often mobility data are used to inform interaction parameters); (3) semi-mechanistic Bayesian transmission model (Bayesian hierarchical model with time-discrete, stochastic renewal process); (4) individual-based transmission model (model of transmission dynamics at individual level, often with network structure or simulation of individual behavior and contact patterns); or (5) phylodynamic model (phylogenetic model with population dynamics that is fitted using genome sequence data).
- **Synthetic controls**: A method based on synthetic controls is used to compare the outcome of a target population with a synthetic scenario constructed from on a pool of "control" populations.

- **Other**: Any other method that does not fit in one of the above categories.

**Addendum for mechanistic models in parametric approaches** If a parametric approach is used and the method is a mechanistic model, then the parameter representing the intervention effect must be functionally linked to some latent variable representing disease dynamics. This makes explicit how the intervention is assumed to influence disease dynamics in the model. The corresponding latent variable is recorded as follows.

- **Not applicable**: No model with a latent variable that is functionally linked to the intervention is used.
- **Reproduction number**: Intervention changes the reproduction number.
- **Transmission rate**: Intervention changes the transmission rate, i. e. effective contact rate.
- **Contact rate**: Intervention changes the contact rate.
- **Probability of transmission upon contact**: Intervention changes the probability of transmission upon contact.
- **Contact matrix**: Intervention changes individual entries of the contact matrix, representing scaled contact rates between different subpopulations (e. g. age groups).
- **Quarantine rate**: Intervention changes the rate at which individuals are quarantined.
- **Susceptible decay**: Intervention removes susceptible individuals without adding infected individuals.
- **Other**: Other type of latent variable, details noted in parentheses.

##### *D.4.4 Code availability*

Statement regarding the availability of the code. We acknowledge that many analyses, in particular descriptive analyses, would not require code to reproduce the results. Thus, this item should be analyzed within specific categories, e. g. analyses that use a parametric approach.

- **None**: No code is made publicly available.
- **Publicly available**: The code is made publicly available.

#### D.5 Effect assessment

##### *D.5.1 Reporting of intervention effect*

How the effects of interventions on outcomes are reported.

- **Qualitative statement**: Effects are reported without quantification, usually by qualitatively stating changes in the outcome that are likely related to the implemented interventions.
- **Comparison of outcome values**: Effects are reported via a comparison of outcome values, usually by comparing the outcome value before the intervention with the outcome value after the intervention.
- **Quantification of change in outcome values**: Effects are reported by quantifying changes in outcome values, usually by computing or estimating the difference in the outcome values with and without the intervention,

e. g. in the form of a model parameter. Of note, in some cases, the difference between quantification of the change in and comparison of outcome values can be subtle. For instance, let $B$ be the outcome value before an intervention and $A$ be the outcome after an intervention. A comparison would be to note that $A$ is lower than $B$, whereas a quantification would be to compute the effect of the intervention as the difference $A - B$.

###### D.5.2 Measure of effect

On which outcome scale the effects of interventions are measured.

- Change in one of the outcomes of Item D.2.1, Item D.2.3, or Item D.4.3.
- Change in any other outcome scale derived from Item D.2.1, Item D.2.3, or Item D.4.3.

###### D.5.3 Reporting of uncertainty

Whether uncertainty regarding the effects of interventions on outcomes is reported. If several outcomes are analyzed, we report if uncertainty is reported for the effect of interventions on at least one outcome.

- **Not applicable**: Effects are not quantified, thus uncertainty reporting is not applicable.
- **Yes**: Uncertainty is reported by providing information on imprecision, e. g. by reporting the standard deviation, standard error, confidence interval, $p$-value, credible interval, etc. of the estimated effect of interventions. For comparison of outcome values, uncertainty is reported for the outcome values. For quantification of the change in outcome values, uncertainty is reported for the quantified change or estimated parameter.
- **No**: Effects are quantified, but uncertainty is not reported.

###### D.5.4 Sensitivity analysis

Categorization of analyses that are conducted for assessing sensitivity with respect to the effect of interventions on the outcome. Detailed categories in parentheses. Note that we also considered sensitivity analyses for models used in descriptive approaches (to obtain a computed outcome) if the sensitivity was assessed with regard to the intervention effect.

- **None**: No sensitivity analyses are conducted.
- **Different coding of interventions used**: Assessing sensitivity with respect to the use of differently coded intervention data. Details in parentheses.
- **Different or modified outcome used**: Assessing sensitivity with respect to the use of different or modified outcomes. Details in parentheses.
- **Same analysis with (sub)population(s) excluded**: Assessing sensitivity with respect to the exclusion of specific (sub)populations for the same analysis.
- **Epidemiological parameters varied**: Assessing sensitivity with respect to varying assumptions regarding epidemiological parameters, e. g. serial interval or delay from infection to reporting of a new case.
- **Start or end date of study period varied**: Assessing sensitivity with respect to variations of the start and end date of the study period.

- **Model specification varied**: Assessing sensitivity with respect to changes in the model structure or specification, e. g. extending the model or adjusting the effects of NPIs for additional variables.

###### D.5.5 Subgroup assessment

The subgroups within which the effects of interventions on outcomes are assessed. These subgroups are usually created by grouping the populations based on the values of some other variable.

- **None**: No subgroup assessments.
- **Socioeconomic indicators**: Subgroups based on age, gender, income (e. g. household income at the individual level or by national income at the population level), nationality, population density, household size, health index (e. g. score for the global health security index), or occupation (i. e. workers with and without health care occupation at the individual level).
- **Epidemiological indicators**: Subgroups based on infection setting (e. g. workplace, household, school or community), pre-intervention epidemic trend (e. g. similar epidemic trends before the implementation of interventions), source of infection (whether the infection can be traced back to a local case or an imported case), isolation delay (e. g. varying delay until a person was isolated after symptom onset at the individual level), or disease severity (disease severity with COVID-19 at the individual level).
- **Public health response**: Subgroups based on response time (time till governments responded to the pandemic with an intervention) or whether being subject to other measures (e. g. whether being subject to contact tracing at the individual level or regions with/without other interventions implemented at the population level).
- **Geographic areas**: Subgroups based on regions (e. g. Europe, Asia, Middle East).

### E Data extraction report

The following section provides a detailed report of all information extracted from the studies in our review sample. The corresponding items are described in the Data extraction manual. The extracted information is presented for each individual analysis, where some studies contain multiple analyses (referred to as Analysis 1, 2, etc.). Of note, some studies did not only assess the effects of non-pharmaceutical interventions on mobility, but also the effects of changes in mobility on population-level epidemiological outcomes. We also extracted information on such complementary analyses that use "mobility as an exposure" and present them along with their main analyses of intervention effects in the following.

---

**Abdalla et al. 2021** . . . . . **38**

Analysis 1 . . . . . 38

Analysis 2 . . . . . 39

Analysis 3, mobility as exposure . . . . . 41

**Abouk & Heydari 2021** . . . . . **43**

**Abuhasel et al. 2020** . . . . . **45**

**Adekunle et al. 2020** . . . . . **47**

**Aguiar et al. 2020** . . . . . **49**

Analysis 1 . . . . . 49

Analysis 2 . . . . . 50

**Akimkin et al. 2020** . . . . . **52**

**Aldila et al. 2020** . . . . . **54**

**Ali et al. 2020** . . . . . **56**

**Alimohamadi et al. 2020** . . . . . **58**

**Aloi et al. 2020** . . . . . **60**

**Alumran 2020** . . . . . **62**

**Anderson et al. 2020** . . . . . **64**

**Anzai et al. 2020** . . . . . **66**

**Aravindakshan et al. 2020** . . . . . **68**

Analysis 1 . . . . . 68

Analysis 2, mobility as exposure . . . . . 69

**Arshed et al. 2020** . . . . . **71**

**Asongu et al. 2020** . . . . . . . . . . . . . . . . . . . . . . . . . . . . . . . **73**

**Auger et al. 2020** . . . . . . . . . . . . . . . . . . . . . . . . . . . . . . . . **75**

**Badr et al. 2020** . . . . . . . . . . . . . . . . . . . . . . . . . . . . . . . . **77**

Analysis 1 . . . . . . . . . . . . . . . . . . . . . . . . . . . . . . . . . . . 77

Analysis 2, mobility as exposure . . . . . . . . . . . . . . . . . . . . . . . . 79

**Baker et al. 2020** . . . . . . . . . . . . . . . . . . . . . . . . . . . . . . . . **81**

Analysis 1 . . . . . . . . . . . . . . . . . . . . . . . . . . . . . . . . . . . 81

Analysis 2 . . . . . . . . . . . . . . . . . . . . . . . . . . . . . . . . . . . 83

**Banerjee & Nayak 2020** . . . . . . . . . . . . . . . . . . . . . . . . . . . . **85**

**Bemanian et al. 2020** . . . . . . . . . . . . . . . . . . . . . . . . . . . . . **87**

**Bennett 2021** . . . . . . . . . . . . . . . . . . . . . . . . . . . . . . . . . **88**

**Bo et al. 2021** . . . . . . . . . . . . . . . . . . . . . . . . . . . . . . . . . **90**

**Bönisch et al. 2020** . . . . . . . . . . . . . . . . . . . . . . . . . . . . . . **92**

Analysis 1 . . . . . . . . . . . . . . . . . . . . . . . . . . . . . . . . . . . 92

Analysis 2, mobility as exposure . . . . . . . . . . . . . . . . . . . . . . . . 94

**Braithwaite et al. 2020** . . . . . . . . . . . . . . . . . . . . . . . . . . . . **96**

**Brauner et al. 2021** . . . . . . . . . . . . . . . . . . . . . . . . . . . . . . **98**

**Candido et al. 2020** . . . . . . . . . . . . . . . . . . . . . . . . . . . . . . **100**

Analysis 1 . . . . . . . . . . . . . . . . . . . . . . . . . . . . . . . . . . . 100

Analysis 2, mobility as exposure . . . . . . . . . . . . . . . . . . . . . . . . 103

**Cao et al. 2020** . . . . . . . . . . . . . . . . . . . . . . . . . . . . . . . . **105**

**Castaneda & Saygili 2020** . . . . . . . . . . . . . . . . . . . . . . . . . . . **107**

**Cento et al. 2020** . . . . . . . . . . . . . . . . . . . . . . . . . . . . . . . **109**

**Chakraborty & Ghosh 2020** . . . . . . . . . . . . . . . . . . . . . . . . . . **111**

**Chatterjee et al. 2020** . . . . . . . . . . . . . . . . . . . . . . . . . . . . . **112**

**Chaudhry et al. 2020** . . . . . . . . . . . . . . . . . . . . . . . . . . . . . **114**

**Chaves et al. 2020** . . . . . . . . . . . . . . . . . . . . . . . . . . . . . . **116**

**Cheema et al. 2021** . . . . . . . . . . . . . . . . . . . . . . . . . . . . . . **118**

**Chen et al. 2020a** . . . . . . . . . . . . . . . . . . . . . . . . . . . . . . . **120**

**Chen et al. 2020b** . . . . . . . . . . **122**

**Chen et al. 2020c** . . . . . . . . . . **124**

**Cheng et al. 2020a** . . . . . . . . . . **126**

**Cheng et al. 2020b** . . . . . . . . . . **128**

**Cheng et al. 2020c** . . . . . . . . . . **129**

Analysis 1 . . . . . . . . . . 130

Analysis 2 . . . . . . . . . . 131

Analysis 3, mobility as exposure . . . . . . . . . . 133

**Cheng et al. 2020d** . . . . . . . . . . **135**

**Chernozhukov et al. 2021** . . . . . . . . . . **137**

Analysis 1 . . . . . . . . . . 137

Analysis 2, mobility as exposure . . . . . . . . . . 139

**Choi & Ki 2020** . . . . . . . . . . **141**

**Choi et al. 2020** . . . . . . . . . . **143**

**Cobb & Seale 2020** . . . . . . . . . . **145**

**Coletti et al. 2020** . . . . . . . . . . **146**

**Collins & Duffy 2020** . . . . . . . . . . **148**

**Cooper et al. 2020** . . . . . . . . . . **150**

**Courtemanche et al. 2020** . . . . . . . . . . **152**

**Cowling et al. 2020** . . . . . . . . . . **154**

Analysis 1 . . . . . . . . . . 154

Analysis 2 . . . . . . . . . . 156

**Crokidakis 2020** . . . . . . . . . . **158**

**Czerwińska & Szenborn 2020** . . . . . . . . . . **160**

**da Silva & Tsigaris 2020** . . . . . . . . . . **162**

**Dai et al. 2020** . . . . . . . . . . **163**

**Dandekar et al. 2020** . . . . . . . . . . **165**

**de Brito Cruz 2020** . . . . . . . . . . **167**

Analysis 1 . . . . . . . . . . 167

Analysis 2, mobility as exposure . . . 169

**Dehning et al. 2020 . . . 171**

**Dickson et al. 2020 . . . 173**

**Dikid et al. 2020 . . . 175**

**Djurović 2020 . . . 177**

**Domenico et al. 2020 . . . 179**

**Donsimoni et al. 2020 . . . 180**

**Du et al. 2020 . . . 182**

Analysis 1 . . . 182

Analysis 2 . . . 184

**Duhon et al. 2021 . . . 186**

**Džiugys et al. 2020 . . . 188**

**Edjoc et al. 2020 . . . 190**

**Eguchi et al. 2020 . . . 192**

**Fatmi 2020 . . . 194**

**Feng et al. 2020 . . . 195**

**Fernández-Recio 2020 . . . 197**

**Flaxman et al. 2020 . . . 199**

**Frank 2020 . . . 202**

**Friedrich et al. 2020 . . . 204**

**Friedson et al. 2020 . . . 205**

Analysis 1 . . . 205

Analysis 2 . . . 207

**Gallaway et al. 2020 . . . 209**

**Gao et al. 2020 . . . 211**

Analysis 1 . . . 211

Analysis 2 . . . 213

**Gatto et al. 2020 . . . 215**

**Ge et al. 2020 . . . 217**

**Ghosal et al. 2020** . . . . . . . . . . **219**
**Gill et al. 2020** . . . . . . . . . . **220**
Analysis 1 . . . . . . . . . . 220
Analysis 2 . . . . . . . . . . 222
**Gregori et al. 2020** . . . . . . . . . . **224**
**Guirao 2020** . . . . . . . . . . **226**
**Guo & Xiao 2020** . . . . . . . . . . **228**
**Gupta et al. 2021** . . . . . . . . . . **230**
Analysis 1 . . . . . . . . . . 230
Analysis 2 . . . . . . . . . . 232
**Hao et al. 2020** . . . . . . . . . . **234**
**Haug et al. 2020** . . . . . . . . . . **235**
**He et al. 2020a** . . . . . . . . . . **237**
**He et al. 2020b** . . . . . . . . . . **239**
**Henríquez et al. 2020** . . . . . . . . . . **241**
Analysis 1 . . . . . . . . . . 241
Analysis 2 . . . . . . . . . . 243
**Holtz et al. 2020** . . . . . . . . . . **245**
**Hsiang et al. 2020** . . . . . . . . . . **247**
**Huang et al. 2020** . . . . . . . . . . **249**
**Huber & Langen 2020** . . . . . . . . . . **251**
Analysis 1 . . . . . . . . . . 251
Analysis 2 . . . . . . . . . . 253
Analysis 3 . . . . . . . . . . 254
**Hyafil & Moriña 2021** . . . . . . . . . . **256**
**Islam et al. 2020** . . . . . . . . . . **258**
**Jardine et al. 2020** . . . . . . . . . . **260**
**Jarvis et al. 2020** . . . . . . . . . . **262**
**Jefferies et al. 2020** . . . . . . . . . . **264**

**Jeffrey et al. 2020** . . . . . . . . . . **266**
Analysis 1 . . . . . . . . . . 266
Analysis 2, mobility as exposure . . . . . . . . . . 268
**Ji et al. 2020** . . . . . . . . . . **270**
**Jia et al. 2020** . . . . . . . . . . **272**
**Jüni et al. 2020** . . . . . . . . . . **273**
**Kang & Kim 2020** . . . . . . . . . . **275**
**Karnakov et al. 2020** . . . . . . . . . . **277**
Analysis 1 . . . . . . . . . . 277
Analysis 2 . . . . . . . . . . 279
**Kaufman et al. 2020** . . . . . . . . . . **281**
**Kendall et al. 2020** . . . . . . . . . . **283**
Analysis 1 . . . . . . . . . . 283
Analysis 2 . . . . . . . . . . 285
**Khan et al. 2020** . . . . . . . . . . **287**
**Kharroubi & Saleh 2020** . . . . . . . . . . **289**
**Kim et al. 2020a** . . . . . . . . . . **291**
**Kim et al. 2020b** . . . . . . . . . . **292**
**Klimek-Tulwin & Tulwin 2020** . . . . . . . . . . **294**
**Koh et al. 2020** . . . . . . . . . . **296**
Analysis 1 . . . . . . . . . . 296
Analysis 2 . . . . . . . . . . 298
Analysis 3, mobility as exposure . . . . . . . . . . 300
**Kraemer et al. 2020** . . . . . . . . . . **302**
Analysis 1 . . . . . . . . . . 302
Analysis 2 . . . . . . . . . . 304
Analysis 3, mobility as exposure . . . . . . . . . . 306
**Krishna 2020** . . . . . . . . . . **308**
**Kucharski et al. 2020** . . . . . . . . . . **309**

**Kumar et al. 2020** . . . . . . . . . . . . . . . . . . . . . . . . . . . . **312**
**Kuo & Fu 2021** . . . . . . . . . . . . . . . . . . . . . . . . . . . . . **314**
**Lau et al. 2020** . . . . . . . . . . . . . . . . . . . . . . . . . . . . . **315**
Analysis 1 . . . . . . . . . . . . . . . . . . . . . . . . . . . . . . . . 316
Analysis 2 . . . . . . . . . . . . . . . . . . . . . . . . . . . . . . . . 317
**Lee et al. 2020** . . . . . . . . . . . . . . . . . . . . . . . . . . . . . **319**
**Lei et al. 2020** . . . . . . . . . . . . . . . . . . . . . . . . . . . . . **321**
**Lemaitre et al. 2020** . . . . . . . . . . . . . . . . . . . . . . . . . . **323**
Analysis 1 . . . . . . . . . . . . . . . . . . . . . . . . . . . . . . . . 323
Analysis 2 . . . . . . . . . . . . . . . . . . . . . . . . . . . . . . . . 325
Analysis 3, mobility as exposure . . . . . . . . . . . . . . . . . . . . . . 327
**Li et al. 2020a** . . . . . . . . . . . . . . . . . . . . . . . . . . . . . **329**
**Li et al. 2020b** . . . . . . . . . . . . . . . . . . . . . . . . . . . . . **331**
**Li et al. 2020c** . . . . . . . . . . . . . . . . . . . . . . . . . . . . . **332**
Analysis 1 . . . . . . . . . . . . . . . . . . . . . . . . . . . . . . . . 332
Analysis 2 . . . . . . . . . . . . . . . . . . . . . . . . . . . . . . . . 334
**Li et al. 2021** . . . . . . . . . . . . . . . . . . . . . . . . . . . . . **336**
**Lim et al. 2020a** . . . . . . . . . . . . . . . . . . . . . . . . . . . . **338**
Analysis 1 . . . . . . . . . . . . . . . . . . . . . . . . . . . . . . . . 338
Analysis 2, mobility as exposure . . . . . . . . . . . . . . . . . . . . . . 340
**Lim et al. 2020b** . . . . . . . . . . . . . . . . . . . . . . . . . . . . **342**
**Linka et al. 2020** . . . . . . . . . . . . . . . . . . . . . . . . . . . . **344**
Analysis 1 . . . . . . . . . . . . . . . . . . . . . . . . . . . . . . . . 344
Analysis 2, mobility as exposure . . . . . . . . . . . . . . . . . . . . . . 346
**Liu et al. 2020a** . . . . . . . . . . . . . . . . . . . . . . . . . . . . **347**
**Liu et al. 2020b** . . . . . . . . . . . . . . . . . . . . . . . . . . . . **349**
Analysis 1 . . . . . . . . . . . . . . . . . . . . . . . . . . . . . . . . 349
Analysis 2, mobility as exposure . . . . . . . . . . . . . . . . . . . . . . 351
**Loeffler-Wirth et al. 2020** . . . . . . . . . . . . . . . . . . . . . . . **353**

**Lurie et al. 2020** . . . . . . . . . . . . . . . . . . . . . . . . . . . **355**
Analysis 1 . . . . . . . . . . . . . . . . . . . . . . . . . . . . . . . . . . . 355
Analysis 2 . . . . . . . . . . . . . . . . . . . . . . . . . . . . . . . . . . . 357
**Maier & Brockmann 2020** . . . . . . . . . . . . . . . . . . . . . . . **358**
**Malheiro et al. 2020** . . . . . . . . . . . . . . . . . . . . . . . . . **360**
**Manevski et al. 2020** . . . . . . . . . . . . . . . . . . . . . . . . . **362**
**Marschner 2020** . . . . . . . . . . . . . . . . . . . . . . . . . . . . **364**
**McCarthy et al. 2020** . . . . . . . . . . . . . . . . . . . . . . . . **366**
**McGrail et al. 2020** . . . . . . . . . . . . . . . . . . . . . . . . . **368**
**McKee et al. 2020** . . . . . . . . . . . . . . . . . . . . . . . . . . **370**
**McKenzie & Adams 2020** . . . . . . . . . . . . . . . . . . . . . . . **372**
**Medline et al. 2020** . . . . . . . . . . . . . . . . . . . . . . . . . **374**
**Miller et al. 2020** . . . . . . . . . . . . . . . . . . . . . . . . . . **376**
**Mitra et al. 2020** . . . . . . . . . . . . . . . . . . . . . . . . . . **378**
**Moon et al. 2020** . . . . . . . . . . . . . . . . . . . . . . . . . . **380**
**Moreno et al. 2020** . . . . . . . . . . . . . . . . . . . . . . . . . **382**
**Moshammer et al. 2020** . . . . . . . . . . . . . . . . . . . . . . . **384**
**Mukandavire et al. 2020** . . . . . . . . . . . . . . . . . . . . . . **386**
**Munayco et al. 2020** . . . . . . . . . . . . . . . . . . . . . . . . **387**
**Murillo-Zamora et al. 2020** . . . . . . . . . . . . . . . . . . . . . **389**
**Ng et al. 2020** . . . . . . . . . . . . . . . . . . . . . . . . . . . . **391**
**Niazkar et al. 2020** . . . . . . . . . . . . . . . . . . . . . . . . . **393**
Analysis 1 . . . . . . . . . . . . . . . . . . . . . . . . . . . . . . . . . . . 393
Analysis 2 . . . . . . . . . . . . . . . . . . . . . . . . . . . . . . . . . . . 395
**Nie et al. 2020** . . . . . . . . . . . . . . . . . . . . . . . . . . . **397**
**Niwa et al. 2020** . . . . . . . . . . . . . . . . . . . . . . . . . . **399**
**Odagaki 2020** . . . . . . . . . . . . . . . . . . . . . . . . . . . . **401**
**Ouchetto et al. 2020** . . . . . . . . . . . . . . . . . . . . . . . . **402**
**Pan et al. 2020a** . . . . . . . . . . . . . . . . . . . . . . . . . . **404**

**Pan et al. 2020b** . . . . . . . . . . . . . . . . . . . . . . . . . . . . . . . **406**

**Pang et al. 2020** . . . . . . . . . . . . . . . . . . . . . . . . . . . . . . . **408**

**Pei et al. 2020** . . . . . . . . . . . . . . . . . . . . . . . . . . . . . . . . **410**

**Peixoto et al. 2020** . . . . . . . . . . . . . . . . . . . . . . . . . . . . . **412**

**Peng et al. 2020** . . . . . . . . . . . . . . . . . . . . . . . . . . . . . . . **414**

**Pépin et al. 2020** . . . . . . . . . . . . . . . . . . . . . . . . . . . . . . . **416**

**Piovani et al. 2021** . . . . . . . . . . . . . . . . . . . . . . . . . . . . . **417**

**Price et al. 2020** . . . . . . . . . . . . . . . . . . . . . . . . . . . . . . . **419**

**Pullano et al. 2020** . . . . . . . . . . . . . . . . . . . . . . . . . . . . . **421**

Analysis 1 . . . . . . . . . . . . . . . . . . . . . . . . . . . . . . . . . . . 421

Analysis 2 . . . . . . . . . . . . . . . . . . . . . . . . . . . . . . . . . . . 423

Analysis 3 . . . . . . . . . . . . . . . . . . . . . . . . . . . . . . . . . . . 425

**Pung et al. 2020** . . . . . . . . . . . . . . . . . . . . . . . . . . . . . . . **427**

**Qiu et al. 2020** . . . . . . . . . . . . . . . . . . . . . . . . . . . . . . . . **429**

Analysis 1 . . . . . . . . . . . . . . . . . . . . . . . . . . . . . . . . . . . 429

Analysis 2 . . . . . . . . . . . . . . . . . . . . . . . . . . . . . . . . . . . 431

**Quaife et al. 2020** . . . . . . . . . . . . . . . . . . . . . . . . . . . . . . **433**

**Rahman et al. 2020** . . . . . . . . . . . . . . . . . . . . . . . . . . . . . . **435**

Analysis 1 . . . . . . . . . . . . . . . . . . . . . . . . . . . . . . . . . . . 435

Analysis 2, mobility as exposure . . . . . . . . . . . . . . . . . . . . . . . . . 437

**Reis et al. 2020** . . . . . . . . . . . . . . . . . . . . . . . . . . . . . . . . **439**

**Riccardo et al. 2020** . . . . . . . . . . . . . . . . . . . . . . . . . . . . . **441**

**Rieger & Wang 2020** . . . . . . . . . . . . . . . . . . . . . . . . . . . . . **443**

**Ryu et al. 2020** . . . . . . . . . . . . . . . . . . . . . . . . . . . . . . . . **445**

**Saez et al. 2020** . . . . . . . . . . . . . . . . . . . . . . . . . . . . . . . . **447**

**Salje et al. 2020** . . . . . . . . . . . . . . . . . . . . . . . . . . . . . . . **448**

**Salvatore et al. 2020** . . . . . . . . . . . . . . . . . . . . . . . . . . . . . **450**

**Santamaria et al. 2020** . . . . . . . . . . . . . . . . . . . . . . . . . . . . **452**

**Saurabh et al. 2020** . . . . . . . . . . . . . . . . . . . . . . . . . . . . . . **454**

**Scarabel et al. 2020** . . . . . . . . . . . . . . . . . . . . . . . . . . . . 456

**Sebastian et al. 2020** . . . . . . . . . . . . . . . . . . . . . . . . . . . . 458

Analysis 1 . . . . . . . . . . . . . . . . . . . . . . . . . . . . . . . . . . . 458

Analysis 2 . . . . . . . . . . . . . . . . . . . . . . . . . . . . . . . . . . . 460

**Sebastiani et al. 2020** . . . . . . . . . . . . . . . . . . . . . . . . . . . 462

**Seemann et al. 2020** . . . . . . . . . . . . . . . . . . . . . . . . . . . . 464

**Sehra et al. 2020** . . . . . . . . . . . . . . . . . . . . . . . . . . . . . . 466

Analysis 1 . . . . . . . . . . . . . . . . . . . . . . . . . . . . . . . . . . . 466

Analysis 2, mobility as exposure . . . . . . . . . . . . . . . . . . . . . . . 467

**Sekine et al. 2020** . . . . . . . . . . . . . . . . . . . . . . . . . . . . . 469

**Sewell & And 2020** . . . . . . . . . . . . . . . . . . . . . . . . . . . . . 471

Analysis 1 . . . . . . . . . . . . . . . . . . . . . . . . . . . . . . . . . . . 471

Analysis 2 . . . . . . . . . . . . . . . . . . . . . . . . . . . . . . . . . . . 473

**Shanmugam 2020** . . . . . . . . . . . . . . . . . . . . . . . . . . . . . . 475

**Shi et al. 2020a** . . . . . . . . . . . . . . . . . . . . . . . . . . . . . . . 477

Analysis 1 . . . . . . . . . . . . . . . . . . . . . . . . . . . . . . . . . . . 477

Analysis 2 . . . . . . . . . . . . . . . . . . . . . . . . . . . . . . . . . . . 479

**Shi et al. 2020b** . . . . . . . . . . . . . . . . . . . . . . . . . . . . . . . 481

**Shim et al. 2021** . . . . . . . . . . . . . . . . . . . . . . . . . . . . . . . 483

**Siedner et al. 2020** . . . . . . . . . . . . . . . . . . . . . . . . . . . . . 485

**Signorelli et al. 2020** . . . . . . . . . . . . . . . . . . . . . . . . . . . . 487

**Silva et al. 2020** . . . . . . . . . . . . . . . . . . . . . . . . . . . . . . . 488

**Singh & Singh 2020** . . . . . . . . . . . . . . . . . . . . . . . . . . . . . 490

**Singh et al. 2020** . . . . . . . . . . . . . . . . . . . . . . . . . . . . . . 492

**Sinha 2020** . . . . . . . . . . . . . . . . . . . . . . . . . . . . . . . . . . 494

**Spaccaferri et al. 2020** . . . . . . . . . . . . . . . . . . . . . . . . . . . 496

**Sun et al. 2020** . . . . . . . . . . . . . . . . . . . . . . . . . . . . . . . 498

**Supino et al. 2020** . . . . . . . . . . . . . . . . . . . . . . . . . . . . . . 500

**Suraya et al. 2020** . . . . . . . . . . . . . . . . . . . . . . . . . . . . . 502

Analysis 1 . . . . . . . . . . 502
Analysis 2 . . . . . . . . . . 504
**Talmoudi et al. 2020 . . . . . . . . . . 506**
**Tamagusko & Ferreira 2020 . . . . . . . . . . 508**
Analysis 1 . . . . . . . . . . 508
Analysis 2 . . . . . . . . . . 509
**Tchole et al. 2020 . . . . . . . . . . 511**
**Thai et al. 2020 . . . . . . . . . . 513**
**Tian et al. 2020 . . . . . . . . . . 516**
Analysis 1 . . . . . . . . . . 516
Analysis 2 . . . . . . . . . . 517
**Tian et al. 2021 . . . . . . . . . . 519**
**Tiwari et al. 2020 . . . . . . . . . . 521**
**Tsai et al. 2020 . . . . . . . . . . 523**
**Turk et al. 2020 . . . . . . . . . . 525**
**Tutsoy et al. 2020 . . . . . . . . . . 527**
**Utamura et al. 2020 . . . . . . . . . . 529**
**Valcarcel et al. 2020 . . . . . . . . . . 531**
**Valencia et al. 2020 . . . . . . . . . . 533**
Analysis 1 . . . . . . . . . . 533
Analysis 2 . . . . . . . . . . 534
**Van Dyke et al. 2020 . . . . . . . . . . 536**
**Vannoni et al. 2020 . . . . . . . . . . 538**
**Verma et al. 2020 . . . . . . . . . . 540**
**Vicentini et al. 2020 . . . . . . . . . . 542**
**Wagner et al. 2020 . . . . . . . . . . 544**
**Wahaibi et al. 2020 . . . . . . . . . . 546**
**Wan et al. 2020 . . . . . . . . . . 548**
**Wang et al. 2020a . . . . . . . . . . 550**

**Wang et al. 2020b** . . . . . . . . . . . . . . . . . . . . . . . . . . . . **552**

**Wang et al. 2020c** . . . . . . . . . . . . . . . . . . . . . . . . . . . . **554**

Analysis 1 . . . . . . . . . . . . . . . . . . . . . . . . . . . . . . . . . . . . . . . 554

Analysis 2 . . . . . . . . . . . . . . . . . . . . . . . . . . . . . . . . . . . . . . . 556

**Wang et al. 2020d** . . . . . . . . . . . . . . . . . . . . . . . . . . . . **557**

**Wang et al. 2020e** . . . . . . . . . . . . . . . . . . . . . . . . . . . . **559**

**Wells et al. 2020** . . . . . . . . . . . . . . . . . . . . . . . . . . . . . **561**

**Wibbens et al. 2020** . . . . . . . . . . . . . . . . . . . . . . . . . . . **563**

**Wieland 2020** . . . . . . . . . . . . . . . . . . . . . . . . . . . . . . . **565**

Analysis 1 . . . . . . . . . . . . . . . . . . . . . . . . . . . . . . . . . . . . . . . 565

Analysis 2 . . . . . . . . . . . . . . . . . . . . . . . . . . . . . . . . . . . . . . . 567

**Wielechowski et al. 2020** . . . . . . . . . . . . . . . . . . . . . . . . **569**

Analysis 1 . . . . . . . . . . . . . . . . . . . . . . . . . . . . . . . . . . . . . . . 569

Analysis 2, mobility as exposure . . . . . . . . . . . . . . . . . . . . . . . . . . 571

**Wong et al. 2020a** . . . . . . . . . . . . . . . . . . . . . . . . . . . . **573**

**Wong et al. 2020b** . . . . . . . . . . . . . . . . . . . . . . . . . . . . **575**

Analysis 1 . . . . . . . . . . . . . . . . . . . . . . . . . . . . . . . . . . . . . . . 575

Analysis 2, mobility as exposure . . . . . . . . . . . . . . . . . . . . . . . . . . 577

**Wong et al. 2020c** . . . . . . . . . . . . . . . . . . . . . . . . . . . . **578**

Analysis 1 . . . . . . . . . . . . . . . . . . . . . . . . . . . . . . . . . . . . . . . 579

Analysis 2 . . . . . . . . . . . . . . . . . . . . . . . . . . . . . . . . . . . . . . . 580

**Wu et al. 2020** . . . . . . . . . . . . . . . . . . . . . . . . . . . . . . . **582**

Analysis 1 . . . . . . . . . . . . . . . . . . . . . . . . . . . . . . . . . . . . . . . 582

Analysis 2 . . . . . . . . . . . . . . . . . . . . . . . . . . . . . . . . . . . . . . . 584

**Xia et al. 2020** . . . . . . . . . . . . . . . . . . . . . . . . . . . . . . . **586**

**Xiao et al. 2020** . . . . . . . . . . . . . . . . . . . . . . . . . . . . . . **588**

**Xu et al. 2020** . . . . . . . . . . . . . . . . . . . . . . . . . . . . . . . **590**

Analysis 1 . . . . . . . . . . . . . . . . . . . . . . . . . . . . . . . . . . . . . . . 590

Analysis 2 . . . . . . . . . . . . . . . . . . . . . . . . . . . . . . . . . . . . . . . 591

Analysis 3, mobility as exposure . . . . . . . . . . . . . . . . . . . . . . . . . . . 593
**Yang et al. 2020** . . . . . . . . . . . . . . . . . . . . . . . . . . . . . . . . . . . . **595**
**Ye & Hu 2020** . . . . . . . . . . . . . . . . . . . . . . . . . . . . . . . . . . . . . . **597**
**Yehya et al. 2020** . . . . . . . . . . . . . . . . . . . . . . . . . . . . . . . . . . . **599**
**You 2020** . . . . . . . . . . . . . . . . . . . . . . . . . . . . . . . . . . . . . . . . **601**
**You et al. 2020** . . . . . . . . . . . . . . . . . . . . . . . . . . . . . . . . . . . . . **603**
**Yuan et al. 2020** . . . . . . . . . . . . . . . . . . . . . . . . . . . . . . . . . . . . **604**
**Zhang et al. 2020a** . . . . . . . . . . . . . . . . . . . . . . . . . . . . . . . . . . . **606**
**Zhang et al. 2020b** . . . . . . . . . . . . . . . . . . . . . . . . . . . . . . . . . . . **608**
**Zhang et al. 2020c** . . . . . . . . . . . . . . . . . . . . . . . . . . . . . . . . . . . **610**
**Zhang et al. 2020d** . . . . . . . . . . . . . . . . . . . . . . . . . . . . . . . . . . . **612**
**Zhao & Chen 2020** . . . . . . . . . . . . . . . . . . . . . . . . . . . . . . . . . . . . **614**
**Zhao et al. 2020** . . . . . . . . . . . . . . . . . . . . . . . . . . . . . . . . . . . . . **615**

| Abdalla et al. 2021 (Analysis 1) | |
| --- | --- |
| **Study information** | |
| Author | Abdalla M, Abar A, Beiter ER, Saad M |
| Title | Asynchrony between Individual and Government Actions Accounts for Disproportionate Impact of COVID-19 on Vulnerable Communities |
| Year | 2021 |
| Journal | American Journal of Preventive Medicine |
| DOI | 10.1016/j.amepre.2020.10.012 |
| Date received | – could not be evaluated – |
| Date accepted | 2020-11-13 (published) |
| **Study setting** | |
| Number of populations included | multiple |
| Level of populations included | both national and subnational (1124 US counties in 42 US states) |
| Geographic areas covered | North America |
| Number of countries covered | single country (US) |
| Study period | end date could not be evaluated |
| **Outcome** | |
| Raw outcome | epidemiological population-level outcome (cases) |
| Time resolution of raw outcome | daily |
| Computed outcome | none |
| Method to obtain the computed outcome | – not applicable – |
| Data source | data from public media outlets (New York Times) |
| Data availability | data access via source |

– continued on next page –

| | |
|---|---|
| **Intervention** | |
| Terminology for interventions | actions |
| Terminology for the specific type of non-pharmaceutical interventions | government |
| Exposure types | one single intervention (stay-at-home order) |
| Types of single interventions | stay-at-home order |
| Coding of interventions | necessary |
| Source of intervention data | – could not be evaluated – |
| Availability of data on exposure | coded data not available |
| **Methodological approach** | |
| Empirical approach | parametric |
| Use of exposure variation | both variation over time and between populations |
| Method | non-mechanistic model (generalized linear model) |
| Code availability | none |
| **Effect assessment** | |
| Reporting of intervention effect | quantification of change in outcome values |
| Measure of effect | cases |
| Reporting of uncertainty | yes |
| Sensitivity analysis | epidemiological parameters varied |
| Subgroup assessment | none |

**Abdalla et al. 2021 (Analysis 2)**

| | |
|---|---|
| **Study information** | |
| Author | Abdalla M, Abar A, Beiter ER, Saad M |

– continued on next page –

| | |
|---|---|
| Title | Asynchrony between Individual and Government Actions Accounts for Disproportionate Impact of COVID-19 on Vulnerable Communities |
| Year | 2021 |
| Journal | American Journal of Preventive Medicine |
| DOI | 10.1016/j.amepre.2020.10.012 |
| Date received | – could not be evaluated – |
| Date accepted | 2020-11-13 (published) |
| **Study setting** | |
| Number of populations included | multiple |
| Level of populations included | both national and subnational (1124 US counties in 42 US states) |
| Geographic areas covered | North America |
| Number of countries covered | single country (US) |
| Study period | end date could not be evaluated |
| **Outcome** | |
| Raw outcome | behavioral outcome (mobility: time spent at home) |
| Time resolution of raw outcome | daily |
| Computed outcome | change points (change points: mobility) |
| Method to obtain the computed outcome | change point analysis |
| Data source | mobility data from corporate organizations (Google) |
| Data availability | data access via source |
| **Intervention** | |
| Terminology for interventions | actions |
| Terminology for the specific type of non-pharmaceutical interventions | government |
| Exposure types | one single intervention (stay-at-home order) |

– continued on next page –

| | |
|---|---|
| Types of single interventions | stay-at-home order |
| Coding of interventions | necessary |
| Source of intervention data | – could not be evaluated – |
| Availability of data on exposure | coded data not available |
| **Methodological approach** | |
| Empirical approach | descriptive |
| Use of exposure variation | both variation over time and between populations |
| Method | comparison of change points with intervention dates |
| Code availability | none |
| **Effect assessment** | |
| Reporting of intervention effect | quantification of change in outcome values |
| Measure of effect | change points: mobility |
| Reporting of uncertainty | no |
| Sensitivity analysis | none |
| Subgroup assessment | none |

| **Abdalla et al. 2021 (Analysis 3, mobility as exposure)** | |
|---|---|
| **Study information** | |
| Author | Abdalla M, Abar A, Beiter ER, Saad M |
| Title | Asynchrony between Individual and Government Actions Accounts for Disproportionate Impact of COVID-19 on Vulnerable Communities |
| Year | 2021 |
| Journal | American Journal of Preventive Medicine |
| DOI | 10.1016/j.amepre.2020.10.012 |
| Date received | – could not be evaluated – |

– continued on next page –

| | |
|---|---|
| Date accepted | 13.11.2020 (published) |
| **Study setting** | |
| Number of populations included | multiple |
| Level of populations included | both national and subnational (1124 US counties in 42 US states) |
| Geographic areas covered | North America |
| Number of countries covered | single country (US) |
| Study period | end date could not be evaluated |
| **Outcome** | |
| Raw outcome | epidemiological population-level outcome (cases) |
| Time resolution of raw outcome | daily |
| Computed outcome | none |
| Method to obtain the computed outcome | – not applicable – |
| Data source | data from public media outlets (New York Times) |
| Data availability | data access via source |
| **Intervention** | |
| Terminology for interventions | actions |
| Terminology for the specific type of non-pharmaceutical interventions | government |
| Exposure types | change points (mobility: time spent at home) |
| Types of single interventions | – not applicable – |
| Coding of interventions | – not applicable – |
| Source of intervention data | mobility data by corporate organizations (Google) |
| Availability of data on exposure | data access via source |
| **Methodological approach** | |
| Empirical approach | parametric |

– continued on next page –

| | |
|---|---|
| Use of exposure variation | both variation over time and between populations |
| Method | non-mechanistic model (generalized linear model) |
| Code availability | none |
| **Effect assessment** | |
| Reporting of intervention effect | quantification of change in outcome values |
| Measure of effect | cases |
| Reporting of uncertainty | yes |
| Sensitivity analysis | epidemiological parameters varied |
| Subgroup assessment | none |

**Abouk & Heydari 2021**

| | |
|---|---|
| **Study information** | |
| Author | Abouk R, Heydari B |
| Title | The Immediate Effect of COVID-19 Policies on Social-Distancing Behavior in the United States |
| Year | 2021 |
| Journal | Public Health Reports |
| DOI | 10.1177/0033354920976575 |
| Date received | – could not be evaluated – |
| Date accepted | 2020-01-05 (published) |
| **Study setting** | |
| Number of populations included | multiple |
| Level of populations included | both national and subnational (50 US states and the District of Columbia ) |
| Geographic areas covered | North America |

– continued on next page –

| | |
|---|---|
| Number of countries covered | single country (US) |
| Study period | start and end date span first epidemic wave: behavioral |
| **Outcome** | |
| Raw outcome | behavioral outcome (mobility: time spent in various locations) |
| Time resolution of raw outcome | – not applicable – |
| Computed outcome | none |
| Method to obtain the computed outcome | – not applicable – |
| Data source | mobility data from corporate organizations (Google) |
| Data availability | data access via source |
| **Intervention** | |
| Terminology for interventions | policies |
| Terminology for the specific type of non-pharmaceutical interventions | social distancing |
| Exposure types | multiple separate interventions (stay- at-home order, limited stay- at-home order, workplace closure, bans of large gatherings, school closure, venue closure) |
| Types of single interventions | stay-at-home order, workplace closure, bans of large gatherings, school closure, venue closure |
| Coding of interventions | necessary |
| Source of intervention data | use of externally coded data (Kaiser Family Foundation) |
| Availability of data on exposure | access to externally coded data via source |
| **Methodological approach** | |
| Empirical approach | parametric |
| Use of exposure variation | both variation over time and between populations |
| Method | non-mechanistic model (generalized linear model) |

– continued on next page –

| | |
|---|---|
| Code availability | none |
| **Effect assessment** | |
| Reporting of intervention effect | quantification of change in outcome values |
| Measure of effect | mobility: time spent in various locations |
| Reporting of uncertainty | yes |
| Sensitivity analysis | same analysis with (sub)population(s) excluded, model specification varied, different coding of interventions used |
| Subgroup assessment | none |

**Abuhasel et al. 2020**

| | |
|---|---|
| **Study information** | |
| Author | Abuhasel KA, Khadr M, Alquraish MM |
| Title | Analyzing and Forecasting COVID -19 Pandemic in the Kingdom of Saudi Arabia Using ARIMA and SIR Models |
| Year | 2020 |
| Journal | Computational Intelligence |
| DOI | 10.1111/coin.12407 |
| Date received | 2020-06-01 |
| Date accepted | 2020-09-06 |
| **Study setting** | |
| Number of populations included | single |
| Level of populations included | national (Saudi Arabia) |
| Geographic areas covered | Middle East and Africa |
| Number of countries covered | single country (Saudi Ariba) |

– continued on next page –

| | |
|---|---|
| Study period | end date at peak of wave |
| **Outcome** | |
| Raw outcome | epidemiological population-level outcome (cases, deaths, recovered cases) |
| Time resolution of raw outcome | daily |
| Computed outcome | none |
| Method to obtain the computed outcome | – not applicable – |
| Data source | data from (sub)national authorities |
| Data availability | data access via source |
| **Intervention** | |
| Terminology for interventions | strategies |
| Terminology for the specific type of non-pharmaceutical interventions | containment, control |
| Exposure types | all interventions together |
| Types of single interventions | – not applicable – |
| Coding of interventions | – not applicable – |
| Source of intervention data | – not applicable – |
| Availability of data on exposure | – not applicable – |
| **Methodological approach** | |
| Empirical approach | counterfactual |
| Use of exposure variation | only variation over time for a single population |
| Method | mechanistic model (compartmental single-population transmission model) |
| Code availability | none |
| **Effect assessment** | |
| Reporting of intervention effect | comparison of outcome values |
| Measure of effect | infections |

– continued on next page –

| | |
|---|---|
| Reporting of uncertainty | no |
| Sensitivity analysis | none |
| Subgroup assessment | none |

| **Adekunle et al. 2020** | |
|---|---|
| **Study information** | |
| Author | Adekunle A, Meehan M, Rojas-Alvarez D, Trauer J, McBryde E |
| Title | Delaying the COVID-19 Epidemic in Australia: Evaluating the Effectiveness of International Travel Bans |
| Year | 2020 |
| Journal | Australian and New Zealand Journal of Public Health |
| DOI | 10.1111/1753-6405.13016 |
| Date received | 2020-04-01 |
| Date accepted | 2020-05-01 |
| **Study setting** | |
| Number of populations included | single |
| Level of populations included | national (Australia) |
| Geographic areas covered | Oceania |
| Number of countries covered | single country (Australia) |
| Study period | end date at peak of wave |
| **Outcome** | |
| Raw outcome | epidemiological population-level outcome (imported cases) |
| Time resolution of raw outcome | daily |
| Computed outcome | none |

– continued on next page –

| | |
|---|---|
| Method to obtain the computed outcome | – not applicable – |
| Data source | data from other research projects (EpiForecasts project by the London School of Hygiene & Tropical Medicine or study by Abbott et al.) |
| Data availability | data access via source |
| **Intervention** | |
| Terminology for interventions | interventions |
| Terminology for the specific type of non-pharmaceutical interventions | – not applicable – |
| Exposure types | one single intervention (travel restrictions (Australian travel bans on travellers from Wuhan, China, Iran, South Korea, Italy)) |
| Types of single interventions | international travel restrictions |
| Coding of interventions | not necessary |
| Source of intervention data | – could not be evaluated – |
| Availability of data on exposure | raw data documented in the manuscript |
| **Methodological approach** | |
| Empirical approach | counterfactual |
| Use of exposure variation | only variation over time for a single population |
| Method | mechanistic model (compartmental metapopulation transmission model) |
| Code availability | none |
| **Effect assessment** | |
| Reporting of intervention effect | quantification of change in outcome values |
| Measure of effect | imported cases |
| Reporting of uncertainty | no |
| Sensitivity analysis | none |
| Subgroup assessment | none |

| Aguiar et al. 2020 (Analysis 1) | |
|---|---|
| **Study information** | |
| Author | Aguiar M, Ortuondo EM, Van-Dierdonck JB, Mar J, Stollenwerk N |
| Title | Modelling COVID 19 in the Basque Country from Introduction to Control Measure Response |
| Year | 2020 |
| Journal | Scientific Reports |
| DOI | 10.1038/s41598-020-74386-1 |
| Date received | 2020-07-20 |
| Date accepted | 2020-09-28 |
| **Study setting** | |
| Number of populations included | single |
| Level of populations included | subnational (Basque) |
| Geographic areas covered | Europe |
| Number of countries covered | single country (Spain) |
| Study period | start and end date span first epidemic wave |
| **Outcome** | |
| Raw outcome | epidemiological population-level outcome (cases, hospitalizations, ICU hospitalizations, recovered cases, deaths) |
| Time resolution of raw outcome | daily |
| Computed outcome | none |
| Method to obtain the computed outcome | – not applicable – |
| Data source | data from (sub)national authorities |

– continued on next page –

| | |
|---|---|
| Data availability | data access via source |
| **Intervention** | |
| Terminology for interventions | measures |
| Terminology for the specific type of non-pharmaceutical interventions | control |
| Exposure types | all interventions together |
| Types of single interventions | – not applicable – |
| Coding of interventions | – not applicable – |
| Source of intervention data | – not applicable – |
| Availability of data on exposure | – not applicable – |
| **Methodological approach** | |
| Empirical approach | counterfactual |
| Use of exposure variation | only variation over time for a single population |
| Method | mechanistic model (compartmental single-population transmission model) |
| Code availability | none |
| **Effect assessment** | |
| Reporting of intervention effect | qualitative statement |
| Measure of effect | cases, hospitalizations, ICU hospitalizations, recovered, deaths |
| Reporting of uncertainty | – not applicable – |
| Sensitivity analysis | none |
| Subgroup assessment | none |

**Aguiar et al. 2020 (Analysis 2)**

| |
|---|
| **Study information** |

– continued on next page –

| | |
|---|---|
| Author | Aguiar M, Ortuondo EM, Van-Dierdonck JB, Mar J, Stollenwerk N |
| Title | Modelling COVID 19 in the Basque Country from Introduction to Control Measure Response |
| Year | 2020 |
| Journal | Scientific Reports |
| DOI | 10.1038/s41598-020-74386-1 |
| Date received | 2020-07-20 |
| Date accepted | 2020-09-28 |
| **Study setting** | |
| Number of populations included | single |
| Level of populations included | subnational (Basque) |
| Geographic areas covered | Europe |
| Number of countries covered | single country (Spain) |
| Study period | start and end date span first epidemic wave |
| **Outcome** | |
| Raw outcome | epidemiological population-level outcome (cases, hospitalizations, ICU hospitalizations, recovered cases, deaths) |
| Time resolution of raw outcome | daily |
| Computed outcome | measure of epidemic trend (growth rate, growth rate: hospitalized, growth rate: ICU hospitalized, growth rate: recovered, growth rate: deaths), epidemiological parameter (reproduction number) |
| Method to obtain the computed outcome | simple computation (growth rate), compartmental transmission model (compartmental single-population transmission model: reproduction number) |
| Data source | data from (sub)national authorities |
| Data availability | data access via source |
| **Intervention** | |

– continued on next page –

| | |
|---|---|
| Terminology for interventions | measures |
| Terminology for the specific type of non-pharmaceutical interventions | control |
| Exposure types | all interventions together |
| Types of single interventions | – not applicable – |
| Coding of interventions | – not applicable – |
| Source of intervention data | – not applicable – |
| Availability of data on exposure | – not applicable – |
| **Methodological approach** | |
| Empirical approach | descriptive |
| Use of exposure variation | only variation over time for a single population |
| Method | description of change over time (description of time course) |
| Code availability | none |
| **Effect assessment** | |
| Reporting of intervention effect | qualitative statement |
| Measure of effect | reproduction number, growth rate, growth rate: hospitalizations, growth rate: ICU hospitalizations, growth rate: recovered, growth rate: deaths |
| Reporting of uncertainty | – not applicable – |
| Sensitivity analysis | none |
| Subgroup assessment | none |

**Akimkin et al. 2020**

| |
|---|
| **Study information** |

– continued on next page –

| | |
|---|---|
| Author | Akimkin VG, Kuzin SN, Semenenko TA, Shipulina OY, Yatsyshina SB, Tivanova EV, Kalenskaya AV, Solovyova IV, Vershinina MA, Kvasova OA, Ploskireva AA, Mamoshina MV, Elkina MA, Klushkina VV, Andreeva EE, Ivanenko AV |
| Title | Patterns of the SARS-CoV-2 Epidemic Spread in a Megacity |
| Year | 2020 |
| Journal | Problems of Virology |
| DOI | 10.36233/0507-4088-2020-65-4-203-211 |
| Date received | – could not be evaluated – |
| Date accepted | – could not be evaluated – |
| **Study setting** | |
| Number of populations included | single |
| Level of populations included | subnational (Moscow) |
| Geographic areas covered | Asia, Europe |
| Number of countries covered | single country (Russia) |
| Study period | start and end date span first epidemic wave |
| **Outcome** | |
| Raw outcome | epidemiological population-level outcome (cases) |
| Time resolution of raw outcome | daily |
| Computed outcome | measure of epidemic trend (growth rate) |
| Method to obtain the computed outcome | simple computation |
| Data source | data from (sub)national authorities (Government of the Russian Federation) |
| Data availability | data access via source |
| **Intervention** | |
| Terminology for interventions | measures |

– continued on next page –

| | |
|---|---|
| Terminology for the specific type of non-pharmaceutical interventions | prevention |
| Exposure types | all interventions together (timeline of measures) |
| Types of single interventions | – not applicable – |
| Coding of interventions | – not applicable – |
| Source of intervention data | – not applicable – |
| Availability of data on exposure | – not applicable – |
| **Methodological approach** | |
| Empirical approach | descriptive |
| Use of exposure variation | only variation over time for a single population |
| Method | description of change over time (comparison of time periods) |
| Code availability | none |
| **Effect assessment** | |
| Reporting of intervention effect | comparison of outcome values |
| Measure of effect | growth rate, cases |
| Reporting of uncertainty | yes |
| Sensitivity analysis | none |
| Subgroup assessment | none |

**Aldila et al. 2020**

| | |
|---|---|
| **Study information** | |
| Author | Aldila D, Khoshnaw SH, Safitri E, Anwar YR, Bakry AR, Samiadji BM, Anugerah DA, Gh MF, Ayulani ID, Salim SN |
| Title | A Mathematical Study on the Spread of COVID-19 Considering Social Distancing and Rapid Assessment: The Case of Jakarta, Indonesia |

– continued on next page –

| | |
|---|---|
| Year | 2020 |
| Journal | Chaos, Solitons & Fractals |
| DOI | 10.1016/j.chaos.2020.110042 |
| Date received | 2020-05-22 |
| Date accepted | 2020-06-18 |
| **Study setting** | |
| Number of populations included | single |
| Level of populations included | subnational (Jakarta) |
| Geographic areas covered | Asia |
| Number of countries covered | single country (Indonesia) |
| Study period | end date in growth phase of wave |
| **Outcome** | |
| Raw outcome | epidemiological population-level outcome (hospitalizations, cases) |
| Time resolution of raw outcome | daily |
| Computed outcome | epidemiological parameter (reproduction number, transmission rate, hospitalization rate) |
| Method to obtain the computed outcome | compartmental transmission model (single-population) |
| Data source | – could not be evaluated – |
| Data availability | data made available by the authors |
| **Intervention** | |
| Terminology for interventions | – not applicable – |
| Terminology for the specific type of non-pharmaceutical interventions | social distancing |
| Exposure types | one combination of interventions (closure of schools and venues) |
| Types of single interventions | – not applicable – |

– continued on next page –

| | |
|---|---|
| Coding of interventions | not necessary |
| Source of intervention data | government or news websites |
| Availability of data on exposure | raw data documented in the manuscript |
| **Methodological approach** | |
| Empirical approach | descriptive |
| Use of exposure variation | only variation over time for a single population |
| Method | description of change over time (comparison of time periods) |
| Code availability | none |
| **Effect assessment** | |
| Reporting of intervention effect | quantification of change in outcome values |
| Measure of effect | reproduction number, transmission rate, hospitalization rate |
| Reporting of uncertainty | no |
| Sensitivity analysis | none |
| Subgroup assessment | none |

**Ali et al. 2020**

| | |
|---|---|
| **Study information** | |
| Author | Ali ST, Wang L, Lau EH, Xu XK, Du Z, Wu Y, Leung GM, Cowling BJ |
| Title | Serial Interval of SARS-CoV-2 Was Shortened over Time by Nonpharmaceutical Interventions |
| Year | 2020 |
| Journal | Science |
| DOI | 10.1126/science.abc9004 |
| Date received | 2020-05-20 |

– continued on next page –

| | |
|---|---|
| Date accepted | 2020-07-13 |
| **Study setting** | |
| Number of populations included | single |
| Level of populations included | national (China) |
| Geographic areas covered | Asia |
| Number of countries covered | single country (China) |
| Study period | start and end date span first epidemic wave |
| **Outcome** | |
| Raw outcome | epidemiological individual-level outcome (individual cases and transmission chains) |
| Time resolution of raw outcome | daily |
| Computed outcome | epidemiological parameter (serial interval) |
| Method to obtain the computed outcome | simple computation |
| Data source | data from (sub)national authorities |
| Data availability | data made available by the authors |
| **Intervention** | |
| Terminology for interventions | interventions |
| Terminology for the specific type of non-pharmaceutical interventions | non-pharmaceutical |
| Exposure types | all interventions together |
| Types of single interventions | – not applicable – |
| Coding of interventions | – not applicable – |
| Source of intervention data | – not applicable – |
| Availability of data on exposure | – not applicable – |
| **Methodological approach** | |

– continued on next page –

| | |
|---|---|
| Empirical approach | descriptive |
| Use of exposure variation | only variation over time for a single population |
| Method | description of change over time (description of time course) |
| Code availability | publicly available |
| **Effect assessment** | |
| Reporting of intervention effect | quantification of change in outcome values |
| Measure of effect | serial interval |
| Reporting of uncertainty | yes |
| Sensitivity analysis | none |
| Subgroup assessment | socioeconomic indicators (age, gender), epidemiological indicators (infection setting, isolation delay) |

**Alimohamadi et al. 2020**

| | |
|---|---|
| **Study information** | |
| Author | Alimohamadi Y, Holakouie-Naieni K, Sepandi M, Taghdir M |
| Title | Effect of Social Distancing on COVID-19 Incidence and Mortality in Iran Since February 20 to May 13, 2020: An Interrupted Time Series Analysis |
| Year | 2020 |
| Journal | Risk Management and Healthcare Policy |
| DOI | 10.2147/rmhp.s265079 |
| Date received | 2020-06-08 |
| Date accepted | 2020-08-29 |
| **Study setting** | |
| Number of populations included | single |

– continued on next page –

| | |
|---|---|
| Level of populations included | national (Iran) |
| Geographic areas covered | Middle East and Africa |
| Number of countries covered | single country (Iran) |
| Study period | start and end date span first epidemic wave |
| **Outcome** | |
| Raw outcome | epidemiological population-level outcome (cases, deaths) |
| Time resolution of raw outcome | daily |
| Computed outcome | none |
| Method to obtain the computed outcome | – not applicable – |
| Data source | data from publicly available cross-country selections (OurWorldInData) |
| Data availability | data access via source |
| **Intervention** | |
| Terminology for interventions | measures |
| Terminology for the specific type of non-pharmaceutical interventions | social distancing |
| Exposure types | one single intervention (venue closure) |
| Types of single interventions | venue closure |
| Coding of interventions | not necessary |
| Source of intervention data | – could not be evaluated – |
| Availability of data on exposure | raw data documented in the manuscript |
| **Methodological approach** | |
| Empirical approach | parametric |
| Use of exposure variation | only variation over time for a single population |
| Method | non-mechanistic model (generalized linear model) |

– continued on next page –

| | |
|---|---|
| Code availability | none |
| **Effect assessment** | |
| Reporting of intervention effect | quantification of change in outcome values |
| Measure of effect | cases, deaths |
| Reporting of uncertainty | yes |
| Sensitivity analysis | none |
| Subgroup assessment | none |

**Aloi et al. 2020**

| | |
|---|---|
| **Study information** | |
| Author | Aloi A, Alonso B, Benavente J, Cordera R, Echániz E, González F, Ladisa C, Lezama-Romanelli R, López-Parra Á, Mazzei V, Perrucci L, Prieto-Quintana D, Rodríguez A, Sañudo R |
| Title | Effects of the COVID-19 Lockdown on Urban Mobility: Empirical Evidence from the City of Santander (Spain) |
| Year | 2020 |
| Journal | Sustainability |
| DOI | 10.3390/su12093870 |
| Date received | 2020-04-14 |
| Date accepted | 2020-05-15 |
| **Study setting** | |
| Number of populations included | single |
| Level of populations included | subnational (Santander (Spain)) |
| Geographic areas covered | Europe |
| Number of countries covered | single country (Spain) |

– continued on next page –

| | |
|---|---|
| Study period | start and end date span first epidemic wave: behavioral |
| **Outcome** | |
| Raw outcome | behavioral outcome (mobility: public transport data & sensors and surveillance cams) |
| Time resolution of raw outcome | daily |
| Computed outcome | none |
| Method to obtain the computed outcome | – not applicable – |
| Data source | data from (sub)national authorities (Santander City Council) |
| Data availability | data not accessible |
| **Intervention** | |
| Terminology for interventions | measures |
| Terminology for the specific type of non-pharmaceutical interventions | confinement, quarantine |
| Exposure types | one combination of interventions (confinement of the population imposed by the Spanish Government on March 15) |
| Types of single interventions | – not applicable – |
| Coding of interventions | not necessary |
| Source of intervention data | – could not be evaluated – |
| Availability of data on exposure | raw data documented in the manuscript |
| **Methodological approach** | |
| Empirical approach | descriptive |
| Use of exposure variation | only variation over time for a single population |
| Method | description of change over time (comparison of time periods) |
| Code availability | none |
| **Effect assessment** | |

– continued on next page –

| | |
|---|---|
| Reporting of intervention effect | quantification of change in outcome values |
| Measure of effect | mobility: various measures of within-city mobility for motor vehicles & public transport & walking |
| Reporting of uncertainty | no |
| Sensitivity analysis | none |
| Subgroup assessment | none |

**Alumran 2020**

| | |
|---|---|
| **Study information** | |
| Author | Alumran A |
| Title | Role of Precautionary Measures in Containing the Natural Course of Novel Coronavirus Disease |
| Year | 2020 |
| Journal | Journal of Multidisciplinary Healthcare |
| DOI | 10.2147/jmdh.s261643 |
| Date received | 2020-05-12 |
| Date accepted | 2020-07-02 |
| **Study setting** | |
| Number of populations included | single |
| Level of populations included | national (Saudi Arabia) |
| Geographic areas covered | Middle East and Africa |
| Number of countries covered | single country (Saudi Ariba) |
| Study period | end date in growth phase of wave |
| **Outcome** | |

– continued on next page –

| | |
|---|---|
| Raw outcome | epidemiological population-level outcome (cases, deaths) |
| Time resolution of raw outcome | daily |
| Computed outcome | none |
| Method to obtain the computed outcome | – not applicable – |
| Data source | data from (sub)national authorities (cases and deaths), data from publicly available cross-country selections (Worldometer: cases and deaths) |
| Data availability | data access via source |
| **Intervention** | |
| Terminology for interventions | measures |
| Terminology for the specific type of non-pharmaceutical interventions | prevention |
| Exposure types | all interventions together (timeline of measures) |
| Types of single interventions | – not applicable – |
| Coding of interventions | – not applicable – |
| Source of intervention data | – not applicable – |
| Availability of data on exposure | – not applicable – |
| **Methodological approach** | |
| Empirical approach | counterfactual |
| Use of exposure variation | only variation over time for a single population |
| Method | non-mechanistic model (exponential growth model) |
| Code availability | none |
| **Effect assessment** | |
| Reporting of intervention effect | quantification of change in outcome values |
| Measure of effect | cases, deaths |
| Reporting of uncertainty | no |

– continued on next page –

| | |
|---|---|
| Sensitivity analysis | none |
| Subgroup assessment | none |

**Anderson et al. 2020**

| | |
|---|---|
| **Study information** | |
| Author | Anderson SC, Edwards AM, Yerlanov M, Mulberry N, Stockdale JE, Iyaniwura SA, Falcao RC, Otterstatter MC, Irvine MA, Janjua NZ, Coombs D, Colijn C |
| Title | Quantifying the Impact of COVID-19 Control Measures Using a Bayesian Model of Physical Distancing |
| Year | 2020 |
| Journal | PLOS Computational Biology |
| DOI | 10.1371/journal.pcbi.1008274 |
| Date received | 2020-05-21 |
| Date accepted | 2020-08-19 |
| **Study setting** | |
| Number of populations included | multiple |
| Level of populations included | both national and subnational (British Columbia, New York, Washington, Florida, California, New Zealand) |
| Geographic areas covered | North America |
| Number of countries covered | single country (Canada) |
| Study period | same end date for several populations with diverse epidemic trajectories |
| **Outcome** | |
| Raw outcome | epidemiological population-level outcome (cases) |
| Time resolution of raw outcome | daily |

– continued on next page –

| | |
|---|---|
| Computed outcome | none |
| Method to obtain the computed outcome | – not applicable – |
| Data source | data from (sub)national authorities (British Columbia and New Zealand), data from public media outlets (COVID Tracking Project by The Atlantic: US) |
| Data availability | data made available by the authors |
| **Intervention** | |
| Terminology for interventions | measures |
| Terminology for the specific type of non-pharmaceutical interventions | physical distancing |
| Exposure types | all interventions together |
| Types of single interventions | – not applicable – |
| Coding of interventions | – not applicable – |
| Source of intervention data | – not applicable – |
| Availability of data on exposure | – not applicable – |
| **Methodological approach** | |
| Empirical approach | parametric |
| Use of exposure variation | only variation over time for multiple populations |
| Method | mechanistic model (compartmental single-population transmission model) |
| Latent variable functionally linked to intervention | contact rate |
| Code availability | publicly available |
| **Effect assessment** | |
| Reporting of intervention effect | quantification of change in outcome values |
| Measure of effect | contact rate |

– continued on next page –

| | |
|---|---|
| Reporting of uncertainty | yes |
| Sensitivity analysis | epidemiological parameters varied |
| Subgroup assessment | none |

| **Anzai et al. 2020** | |
|---|---|
| **Study information** | |
| Author | Anzai A, Kobayashi T, Linton NM, Kinoshita R, Hayashi K, Suzuki A, Yang Y, Jung SM, Miyama T, Akhmetzhanov AR, Nishiura H |
| Title | Assessing the Impact of Reduced Travel on Exportation Dynamics of Novel Coronavirus Infection (COVID-19) |
| Year | 2020 |
| Journal | Journal of Clinical Medicine |
| DOI | 10.3390/jcm9020601 |
| Date received | 2020-02-13 |
| Date accepted | 2020-02-20 |
| **Study setting** | |
| Number of populations included | multiple |
| Level of populations included | national (Japan (imported cases from China)) |
| Geographic areas covered | Asia |
| Number of countries covered | single country (Japan) |
| Study period | end date in growth phase of wave |
| **Outcome** | |
| Raw outcome | epidemiological population-level outcome (cases) |
| Time resolution of raw outcome | daily |

– continued on next page –

| | |
|---|---|
| Computed outcome | none |
| Method to obtain the computed outcome | – not applicable – |
| Data source | data collected by authors (from data from (sub)national authorities and news websites quoting official outbreak reports) |
| Data availability | data made available by the authors |
| **Intervention** | |
| Terminology for interventions | – not applicable – |
| Terminology for the specific type of non-pharmaceutical interventions | travel restrictions |
| Exposure types | one combination of interventions (Wuhan City Lockdown) |
| Types of single interventions | – not applicable – |
| Coding of interventions | not necessary |
| Source of intervention data | – could not be evaluated – |
| Availability of data on exposure | raw data documented in the manuscript |
| **Methodological approach** | |
| Empirical approach | counterfactual |
| Use of exposure variation | both variation over time and between populations |
| Method | non-mechanistic model (generalized linear model) |
| Code availability | none |
| **Effect assessment** | |
| Reporting of intervention effect | quantification of change in outcome values |
| Measure of effect | cases |
| Reporting of uncertainty | yes |
| Sensitivity analysis | epidemiological parameters varied |
| Subgroup assessment | none |

| Aravindakshan et al. 2020 (Analysis 1) | |
|---|---|
| **Study information** | |
| Author | Aravindakshan A, Boehnke J, Gholami E, Nayak A |
| Title | Preparing for a Future COVID-19 Wave: Insights and Limitations from a Data-Driven Evaluation of Non-Pharmaceutical Interventions in Germany |
| Year | 2020 |
| Journal | Scientific Reports |
| DOI | 10.1038/s41598-020-76244-6 |
| Date received | 2020-07-01 |
| Date accepted | 2020-10-26 |
| **Study setting** | |
| Number of populations included | multiple |
| Level of populations included | both national and subnational (Germanys states) |
| Geographic areas covered | Europe |
| Number of countries covered | single country (Germany) |
| Study period | start and end date span first epidemic wave: behavioral |
| **Outcome** | |
| Raw outcome | behavioral outcome (mobility: time spent in retail and recreation) |
| Time resolution of raw outcome | daily |
| Computed outcome | none |
| Method to obtain the computed outcome | – not applicable – |
| Data source | mobility data from corporate organizations (Google) |
| Data availability | data made available by the authors |

– continued on next page –

| | |
|---|---|
| **Intervention** | |
| Terminology for interventions | interventions |
| Terminology for the specific type of non-pharmaceutical interventions | non-pharmaceutical |
| Exposure types | multiple separate interventions (retail outlets closure (e.g. restaurants), contact restriction (movement in public places restricted to just two people), stay-at-home order, non-essential services closure (e.g. trade shows), educational facilities closure (e.g. school and university closure), initial business closure (e.g. restaurants), international border closure) |
| Types of single interventions | workplace closure, venue closure, bans of small gatherings, stay-at-home order, international travel restrictions |
| Coding of interventions | necessary |
| Source of intervention data | use of externally coded data (Insitute for Health Metrics and Evaluation, ACAPS Government Measures Dataset) |
| Availability of data on exposure | coded data made available by the authors |
| **Methodological approach** | |
| Empirical approach | parametric |
| Use of exposure variation | both variation over time and between populations |
| Method | non-mechanistic model (generalized linear model) |
| Code availability | publicly available |
| **Effect assessment** | |
| Reporting of intervention effect | quantification of change in outcome values |
| Measure of effect | mobility: time spent in retail and recreation |
| Reporting of uncertainty | yes |
| Sensitivity analysis | none |
| Subgroup assessment | none |

| Aravindakshan et al. 2020 (Analysis 2, mobility as exposure) | |
|---|---|
| **Study information** | |
| Author | Aravindakshan A, Boehnke J, Gholami E, Nayak A |
| Title | Preparing for a Future COVID-19 Wave: Insights and Limitations from a Data-Driven Evaluation of Non-Pharmaceutical Interventions in Germany |
| Year | 2020 |
| Journal | Scientific Reports |
| DOI | 10.1038/s41598-020-76244-6 |
| Date received | 01.07.2020 |
| Date accepted | 26.10.2020 |
| **Study setting** | |
| Number of populations included | multiple |
| Level of populations included | both national and subnational (Germanys states) |
| Geographic areas covered | Europe |
| Number of countries covered | single country (Germany) |
| Study period | start and end date span first epidemic wave |
| **Outcome** | |
| Raw outcome | epidemiological population-level outcome (cases) |
| Time resolution of raw outcome | daily |
| Computed outcome | none |
| Method to obtain the computed outcome | – not applicable – |
| Data source | data from (sub)national authorities |
| Data availability | data made available by the authors |
| **Intervention** | |
| Terminology for interventions | interventions |

– continued on next page –

**Aravindakshan et al. 2020 (Analysis 2, mobility as exposure)** – continued from previous page

| | |
|---|---|
| Terminology for the specific type of non-pharmaceutical interventions | non-pharmaceutical |
| Exposure types | mobility (time spent in retail and recreation, predicted from regression model on NPIs) |
| Types of single interventions | – not applicable – |
| Coding of interventions | – not applicable – |
| Source of intervention data | mobility data by corporate organizations (Google) |
| Availability of data on exposure | data access via source |
| **Methodological approach** | |
| Empirical approach | counterfactual |
| Use of exposure variation | only variation over time for multiple populations |
| Method | mechanistic model (compartmental single-population transmission model) |
| Code availability | publicly available |
| **Effect assessment** | |
| Reporting of intervention effect | quantification of change in outcome values |
| Measure of effect | cases |
| Reporting of uncertainty | yes |
| Sensitivity analysis | none |
| Subgroup assessment | none |

**Arshed et al. 2020**

| | |
|---|---|
| **Study information** | |
| Author | Arshed N, Meo MS, Farooq F |
| Title | Empirical Assessment of Government Policies and Flattening of the COVID 19 Curve |

– continued on next page –

| | |
|---|---|
| Year | 2020 |
| Journal | Journal of Public Affairs |
| DOI | 10.1002/pa.2333 |
| Date received | 2020-06-22 |
| Date accepted | 2020-07-26 |
| **Study setting** | |
| Number of populations included | multiple |
| Level of populations included | national (190 countries worldwide) |
| Geographic areas covered | Europe, Middle East and Africa, Asia, Oceania, Central and South America, North America |
| Number of countries covered | multiple countries |
| Study period | same end date for several populations with diverse epidemic trajectories |
| **Outcome** | |
| Raw outcome | epidemiological population-level outcome (cases) |
| Time resolution of raw outcome | daily |
| Computed outcome | summary statistic (number of days until flattening of the curve) |
| Method to obtain the computed outcome | simple computation |
| Data source | data from publicly available cross-country selections (JHU) |
| Data availability | data access via source |
| **Intervention** | |
| Terminology for interventions | policies |
| Terminology for the specific type of non-pharmaceutical interventions | government |

– continued on next page –

| | |
|---|---|
| Exposure types | multiple separate interventions (school closure, bans of large gatherings, workplace closure, stay-at-home order, international travel restrictions, internal movement restrictions, public transport closure) |
| Types of single interventions | school closure, bans of large gatherings, workplace closure, stay-at-home order, international travel restrictions, internal movement restrictions, public transport closure |
| Coding of interventions | necessary |
| Source of intervention data | use of externally coded data (OxCGRT) |
| Availability of data on exposure | access to externally coded data via source |
| **Methodological approach** | |
| Empirical approach | parametric |
| Use of exposure variation | only variation between populations |
| Method | non-mechanistic model (other: generalized additive model) |
| Code availability | none |
| **Effect assessment** | |
| Reporting of intervention effect | quantification of change in outcome values |
| Measure of effect | number of days until flattening of the curve |
| Reporting of uncertainty | yes |
| Sensitivity analysis | none |
| Subgroup assessment | none |

**Asongu et al. 2020**

| | |
|---|---|
| **Study information** | |
| Author | Asongu SA, Diop S, Nnanna J |

– continued on next page –

| | |
|---|---|
| Title | The Geography of the Effectiveness and Consequences of Covid-19 Measures: Global Evidence |
| Year | 2020 |
| Journal | Journal of Public Affairs |
| DOI | 10.1002/pa.2483 |
| Date received | 2020-07-21 |
| Date accepted | 2020-09-11 |
| **Study setting** | |
| Number of populations included | multiple |
| Level of populations included | national (186 countries worldwide) |
| Geographic areas covered | Europe, Middle East and Africa, Asia, Oceania, Central and South America, North America |
| Number of countries covered | multiple countries |
| Study period | same end date for several populations with diverse epidemic trajectories |
| **Outcome** | |
| Raw outcome | epidemiological population-level outcome (cases) |
| Time resolution of raw outcome | daily |
| Computed outcome | summary statistic (peak reached or not) |
| Method to obtain the computed outcome | other (visual inspection) |
| Data source | data from publicly available cross-country selections (ECDC) |
| Data availability | data access via source |
| **Intervention** | |
| Terminology for interventions | measures |
| Terminology for the specific type of non-pharmaceutical interventions | prevention, mitigation |

– continued on next page –

| | |
|---|---|
| Exposure types | categories of interventions (lockdown, movement restrictions, social distancing, governance/economic measures, public health measures (further detailed in manuscript)) |
| Types of single interventions | – not applicable – |
| Coding of interventions | necessary |
| Source of intervention data | use of externally coded data (ACAPS Government Measures Dataset) |
| Availability of data on exposure | access to externally coded data via source |
| **Methodological approach** | |
| Empirical approach | descriptive |
| Use of exposure variation | only variation between populations |
| Method | comparison of populations |
| Code availability | none |
| **Effect assessment** | |
| Reporting of intervention effect | quantification of change in outcome values |
| Measure of effect | peak reached or not |
| Reporting of uncertainty | yes |
| Sensitivity analysis | none |
| Subgroup assessment | geographic areas (regions) |

**Auger et al. 2020**

| | |
|---|---|
| **Study information** | |
| Author | Auger KA, Shah SS, Richardson T, Hartley D, Hall M, Warniment A, Timmons K, Bosse D, Ferris SA, Brady PW, Schondelmeyer AC, Thomson JE |

– continued on next page –

| | |
|---|---|
| Title | Association between Statewide School Closure and COVID-19 Incidence and Mortality in the US |
| Year | 2020 |
| Journal | JAMA |
| DOI | 10.1001/jama.2020.14348 |
| Date received | – could not be evaluated – |
| Date accepted | 2020-07-17 |
| **Study setting** | |
| Number of populations included | multiple |
| Level of populations included | both national and subnational (50 US states) |
| Geographic areas covered | North America |
| Number of countries covered | single country (US) |
| Study period | same end date for several populations with diverse epidemic trajectories |
| **Outcome** | |
| Raw outcome | epidemiological population-level outcome (cases, deaths) |
| Time resolution of raw outcome | daily |
| Computed outcome | none |
| Method to obtain the computed outcome | – not applicable – |
| Data source | data from publicly available cross-country selections (JHU) |
| Data availability | data access via source |
| **Intervention** | |
| Terminology for interventions | interventions |
| Terminology for the specific type of non-pharmaceutical interventions | non-pharmaceutical |

– continued on next page –

| | |
|---|---|
| Exposure types | single individual intervention (school closure (other controls: stay-at-home or shelter-in-place order, nonessential business closure, restaurant and bar closure, and prohibition of gatherings with more than 10 people)) |
| Types of single interventions | school closure |
| Coding of interventions | – not applicable – |
| Source of intervention data | coding done by authors |
| Availability of data on exposure | coded data made available by the authors |
| **Methodological approach** | |
| Empirical approach | parametric |
| Use of exposure variation | both variation over time and between populations |
| Method | non-mechanistic model (generalized linear model) |
| Code availability | none |
| **Effect assessment** | |
| Reporting of intervention effect | quantification of change in outcome values |
| Measure of effect | cases, deaths |
| Reporting of uncertainty | yes |
| Sensitivity analysis | epidemiological parameters varied |
| Subgroup assessment | epidemiological indicators (pre-intervention epidemic trend), socioeconomic indicators (population density, age, health care occupation), public health response (subject to other measures) |

**Badr et al. 2020 (Analysis 1)**

| | |
|---|---|
| **Study information** | |
| Author | Badr HS, Du H, Marshall M, Dong E, Squire MM, Gardner LM |

– continued on next page –

| | |
|---|---|
| Title | Association between Mobility Patterns and COVID-19 Transmission in the USA: A Mathematical Modelling Study |
| Year | 2020 |
| Journal | The Lancet Infectious Diseases |
| DOI | 10.1016/s1473-3099(20)30553-3 |
| Date received | – could not be evaluated – |
| Date accepted | 2020-07-01 (published) |
| **Study setting** | |
| Number of populations included | multiple |
| Level of populations included | subnational (25 US counties) |
| Geographic areas covered | North America |
| Number of countries covered | single country (US) |
| Study period | start and end date span first epidemic wave: behavioral |
| **Outcome** | |
| Raw outcome | behavioral outcome (mobility: number of trips between and within counties, relative to baseline) |
| Time resolution of raw outcome | daily |
| Computed outcome | none |
| Method to obtain the computed outcome | – not applicable – |
| Data source | mobility data from corporate organizations (Teralytics) |
| Data availability | data not accessible |
| **Intervention** | |
| Terminology for interventions | measures |
| Terminology for the specific type of non-pharmaceutical interventions | social distancing |

– continued on next page –

| | |
|---|---|
| Exposure types | one single intervention (stay-at-home order (state- and county-level)) |
| Types of single interventions | stay-at-home order |
| Coding of interventions | necessary |
| Source of intervention data | coding done by authors |
| Availability of data on exposure | coded data made available by the authors |
| **Methodological approach** | |
| Empirical approach | descriptive |
| Use of exposure variation | only variation over time for multiple populations |
| Method | description of change over time (description of time course) |
| Code availability | none |
| **Effect assessment** | |
| Reporting of intervention effect | qualitative statement |
| Measure of effect | mobility: smoothed mobility ratio |
| Reporting of uncertainty | – not applicable – |
| Sensitivity analysis | none |
| Subgroup assessment | none |

**Badr et al. 2020 (Analysis 2, mobility as exposure)**

| | |
|---|---|
| **Study information** | |
| Author | Badr HS, Du H, Marshall M, Dong E, Squire MM, Gardner LM |
| Title | Association between Mobility Patterns and COVID-19 Transmission in the USA: A Mathematical Modelling Study |
| Year | 2020 |
| Journal | The Lancet Infectious Diseases |
| DOI | 10.1016/s1473-3099(20)30553-3 |

– continued on next page –

| | |
|---|---|
| Date received | – could not be evaluated – |
| Date accepted | 01.07.2020 (published) |
| **Study setting** | |
| Number of populations included | multiple |
| Level of populations included | subnational (25 US counties) |
| Geographic areas covered | North America |
| Number of countries covered | single country (US) |
| Study period | same end date for several populations with diverse epidemic trajectories |
| **Outcome** | |
| Raw outcome | epidemiological population-level outcome (cases) |
| Time resolution of raw outcome | daily |
| Computed outcome | measure of epidemic trend (growth rate ratio) |
| Method to obtain the computed outcome | simple computation |
| Data source | data from publicly available cross-country selections (JHU) |
| Data availability | data access via source |
| **Intervention** | |
| Terminology for interventions | measures |
| Terminology for the specific type of non-pharmaceutical interventions | social distancing |
| Exposure types | mobility (number of trips between and within counties, relative to baseline) |
| Types of single interventions | – not applicable – |
| Coding of interventions | – not applicable – |
| Source of intervention data | mobility data by corporate organizations (Teralytics) |
| Availability of data on exposure | data not accessible |
| **Methodological approach** | |

– continued on next page –

| | |
|---|---|
| Empirical approach | parametric |
| Use of exposure variation | both variation over time and between populations |
| Method | non-mechanistic model (generalized linear model, correlation) |
| Code availability | none |
| **Effect assessment** | |
| Reporting of intervention effect | quantification of change in outcome values |
| Measure of effect | growth rate ratio |
| Reporting of uncertainty | yes |
| Sensitivity analysis | different or modified outcome used (mobility aggregated by state) |
| Subgroup assessment | none |

**Baker et al. 2020 (Analysis 1)**

| | |
|---|---|
| **Study information** | |
| Author | Baker RE, Park SW, Yang W, Vecchi GA, Metcalf CJ, Grenfell BT |
| Title | The Impact of COVID-19 Nonpharmaceutical Interventions on the Future Dynamics of Endemic Infections |
| Year | 2020 |
| Journal | Proceedings of the National Academy of Sciences |
| DOI | 10.1073/pnas.2013182117 |
| Date received | 2020-06-24 |
| Date accepted | 2020-10-13 |
| **Study setting** | |
| Number of populations included | multiple |

– continued on next page –

| | |
|---|---|
| Level of populations included | subnational (4 US states: Florinda, Minnesota, Oregan & SW Washingston, Texas) |
| Geographic areas covered | North America |
| Number of countries covered | single country (US) |
| Study period | end date in growth phase of wave |
| **Outcome** | |
| Raw outcome | epidemiological population-level outcome (surrogate disease: test positivity rate for RSV and influenza) |
| Time resolution of raw outcome | weekly |
| Computed outcome | none |
| Method to obtain the computed outcome | – not applicable – |
| Data source | data from (sub)national authorities |
| Data availability | data made available by the authors |
| **Intervention** | |
| Terminology for interventions | interventions |
| Terminology for the specific type of non-pharmaceutical interventions | non-pharmaceutical |
| Exposure types | all interventions together |
| Types of single interventions | – not applicable – |
| Coding of interventions | – not applicable – |
| Source of intervention data | – not applicable – |
| Availability of data on exposure | – not applicable – |
| **Methodological approach** | |
| Empirical approach | descriptive |
| Use of exposure variation | only variation over time for multiple populations |

– continued on next page –

| | |
|---|---|
| Method | description of change over time (comparison of time periods) |
| Code availability | publicly available |
| **Effect assessment** | |
| Reporting of intervention effect | qualitative statement |
| Measure of effect | test positivity rate: RSV & influenza |
| Reporting of uncertainty | – not applicable – |
| Sensitivity analysis | none |
| Subgroup assessment | none |

**Baker et al. 2020 (Analysis 2)**

| | |
|---|---|
| **Study information** | |
| Author | Baker RE, Park SW, Yang W, Vecchi GA, Metcalf CJ, Grenfell BT |
| Title | The Impact of COVID-19 Nonpharmaceutical Interventions on the Future Dynamics of Endemic Infections |
| Year | 2020 |
| Journal | Proceedings of the National Academy of Sciences |
| DOI | 10.1073/pnas.2013182117 |
| Date received | 2020-06-24 |
| Date accepted | 2020-10-13 |
| **Study setting** | |
| Number of populations included | multiple |
| Level of populations included | both national and subnational (majority of US states) |
| Geographic areas covered | North America |
| Number of countries covered | single country (US) |

– continued on next page –

| | |
|---|---|
| Study period | same end date for several populations with diverse epidemic trajectories |
| **Outcome** | |
| Raw outcome | epidemiological population-level outcome (surrogate disease: test positivity rate for influenza) |
| Time resolution of raw outcome | weekly |
| Computed outcome | none |
| Method to obtain the computed outcome | – not applicable – |
| Data source | data from (sub)national authorities |
| Data availability | data made available by the authors |
| **Intervention** | |
| Terminology for interventions | interventions |
| Terminology for the specific type of non-pharmaceutical interventions | non-pharmaceutical |
| Exposure types | all interventions together |
| Types of single interventions | – not applicable – |
| Coding of interventions | – not applicable – |
| Source of intervention data | – not applicable – |
| Availability of data on exposure | – not applicable – |
| **Methodological approach** | |
| Empirical approach | descriptive |
| Use of exposure variation | only variation over time for multiple populations |
| Method | description of change over time (comparison of time periods) |
| Code availability | publicly available |
| **Effect assessment** | |
| Reporting of intervention effect | quantification of change in outcome values |

– continued on next page –

| | |
|---|---|
| Measure of effect | test positivity rate: influenza |
| Reporting of uncertainty | no |
| Sensitivity analysis | none |
| Subgroup assessment | none |

**Banerjee & Nayak 2020**

| | |
|---|---|
| **Study information** | |
| Author | Banerjee T, Nayak A |
| Title | U.S. County Level Analysis to Determine If Social Distancing Slowed the Spread of COVID-19 |
| Year | 2020 |
| Journal | Revista Panamericana de Salud Pública |
| DOI | 10.26633/rpsp.2020.90 |
| Date received | 2020-04-23 |
| Date accepted | 2020-07-03 |
| **Study setting** | |
| Number of populations included | multiple |
| Level of populations included | both national and subnational (US and all counties) |
| Geographic areas covered | North America |
| Number of countries covered | single country (US) |
| Study period | end date in growth phase of wave |
| **Outcome** | |
| Raw outcome | epidemiological population-level outcome (cases) |
| Time resolution of raw outcome | daily |

– continued on next page –

| | |
|---|---|
| Computed outcome | none |
| Method to obtain the computed outcome | – not applicable – |
| Data source | data from (sub)national authorities |
| Data availability | data access via source |
| **Intervention** | |
| Terminology for interventions | interventions |
| Terminology for the specific type of non-pharmaceutical interventions | non-pharmaceutical |
| Exposure types | start of interventions (first NPI out of potentially several NPIs taken in each country) |
| Types of single interventions | – not applicable – |
| Coding of interventions | necessary |
| Source of intervention data | use of externally coded data (New York Times, Keystone) |
| Availability of data on exposure | access to externally coded data via source |
| **Methodological approach** | |
| Empirical approach | parametric |
| Use of exposure variation | both variation over time and between populations |
| Method | non-mechanistic model (generalized linear model) |
| Code availability | none |
| **Effect assessment** | |
| Reporting of intervention effect | quantification of change in outcome values |
| Measure of effect | cases |
| Reporting of uncertainty | yes |
| Sensitivity analysis | model specification varied |
| Subgroup assessment | none |

| Bemanian et al. 2020 | |
|---|---|
| **Study information** | |
| Author | Bemanian A, Ahn KW, O'brien M, Rausch DJ, Weston B, Beyer KMM |
| Title | Investigating the Trajectory of the COVID-19 Outbreak in Milwaukee County and Projected Effects of Relaxed Distancing |
| Year | 2020 |
| Journal | Wisconsin Medical Journal |
| DOI | – not applicable – |
| Date received | – could not be evaluated – |
| Date accepted | 2020-06-01 (published) |
| **Study setting** | |
| Number of populations included | single |
| Level of populations included | subnational (Milwaukee county (Wisconsin, USA)) |
| Geographic areas covered | North America |
| Number of countries covered | single country (US) |
| Study period | end date in growth phase of wave |
| **Outcome** | |
| Raw outcome | epidemiological population-level outcome (cases) |
| Time resolution of raw outcome | daily |
| Computed outcome | epidemiological parameter (reproduction number) |
| Method to obtain the computed outcome | statistical estimation of reproduction number (Cori et al.) |
| Data source | data from (sub)national authorities |
| Data availability | data access via source |
| **Intervention** | |

– continued on next page –

| | |
|---|---|
| Terminology for interventions | measures |
| Terminology for the specific type of non-pharmaceutical interventions | physical distancing |
| Exposure types | multiple combinations of interventions (initial social distancing measures (school closure, gathering bans), stay-at-home order) |
| Types of single interventions | – not applicable – |
| Coding of interventions | not necessary |
| Source of intervention data | – could not be evaluated – |
| Availability of data on exposure | raw data documented in the manuscript |
| **Methodological approach** | |
| Empirical approach | descriptive |
| Use of exposure variation | only variation over time for a single population |
| Method | description of change over time (description of time course) |
| Code availability | none |
| **Effect assessment** | |
| Reporting of intervention effect | qualitative statement |
| Measure of effect | reproduction number |
| Reporting of uncertainty | – not applicable – |
| Sensitivity analysis | none |
| Subgroup assessment | none |

**Bennett 2021**

| | |
|---|---|
| **Study information** | |
| Author | Bennett M |

– continued on next page –

| | |
|---|---|
| Title | All Things Equal? Heterogeneity in Policy Effectiveness against COVID-19 Spread in Chile |
| Year | 2021 |
| Journal | World Development |
| DOI | 10.1016/j.worlddev.2020.105208 |
| Date received | – could not be evaluated – |
| Date accepted | 2020-09-15 |
| **Study setting** | |
| Number of populations included | multiple |
| Level of populations included | both national and subnational (municipalities in Santiago and Chile where COVID was spread, number unclear) |
| Geographic areas covered | Central and South America |
| Number of countries covered | single country (Chile) |
| Study period | end date in growth phase of wave |
| **Outcome** | |
| Raw outcome | epidemiological population-level outcome (cases), behavioral outcome (mobility: transit station mobility index & subway validations) |
| Time resolution of raw outcome | daily |
| Computed outcome | none |
| Method to obtain the computed outcome | – not applicable – |
| Data source | data from (sub)national authorities (health authority: cases, Ministry of Transportation: subway validations), mobility data from corporate organizations (Google: transit station mobility index) |
| Data availability | data access via source |
| **Intervention** | |
| Terminology for interventions | – not applicable – |

– continued on next page –

| | |
|---|---|
| Terminology for the specific type of non-pharmaceutical interventions | lockdown |
| Exposure types | one combination of interventions (lockdown) |
| Types of single interventions | – not applicable – |
| Coding of interventions | necessary |
| Source of intervention data | – could not be evaluated – |
| Availability of data on exposure | coded data made available by the authors |
| **Methodological approach** | |
| Empirical approach | counterfactual |
| Use of exposure variation | both variation over time and between populations |
| Method | synthetic controls |
| Code availability | none |
| **Effect assessment** | |
| Reporting of intervention effect | quantification of change in outcome values |
| Measure of effect | cases, mobility: transit station mobility index, mobility: subway validations |
| Reporting of uncertainty | yes |
| Sensitivity analysis | same analysis with (sub)population(s) excluded |
| Subgroup assessment | socioeconomic indicators (income) |

**Bo et al. 2021**

| | |
|---|---|
| **Study information** | |
| Author | Bo Y, Guo C, Lin C, Zeng Y, Li HB, Zhang Y, Hossain MS, Chan JW, Yeung DW, Kwok KO, Wong SY, Lau AK, Lao XQ |
| Title | Effectiveness of Non-Pharmaceutical Interventions on COVID-19 Transmission in 190 Countries from 23 January to 13 April 2020 |

– continued on next page –

| | |
|---|---|
| Year | 2021 |
| Journal | International Journal of Infectious Diseases |
| DOI | 10.1016/j.ijid.2020.10.066 |
| Date received | 2020-07-27 |
| Date accepted | 2020-10-21 |
| **Study setting** | |
| Number of populations included | multiple |
| Level of populations included | both national and subnational (235 cities from 10 countries + 180 countries worldwide) |
| Geographic areas covered | Europe, Middle East and Africa, Asia, Oceania, Central and South America, North America |
| Number of countries covered | multiple countries |
| Study period | end date could not be evaluated |
| **Outcome** | |
| Raw outcome | epidemiological population-level outcome (cases) |
| Time resolution of raw outcome | daily |
| Computed outcome | epidemiological parameter (reproduction number) |
| Method to obtain the computed outcome | statistical estimation of reproduction number (Cori et al.) |
| Data source | data from publicly available cross-country selections (JHU: cases), mobility data from corporate organizations (Wind financial database: cases) |
| Data availability | data access via source |
| **Intervention** | |
| Terminology for interventions | interventions |
| Terminology for the specific type of non-pharmaceutical interventions | non-pharmaceutical |

– continued on next page –

| | |
|---|---|
| Exposure types | start of interventions (the first day an NPI was implemented within a group (NPI groups: travel restriction, social distancing, mask wearing mandate, isolation or quarantine)) |
| Types of single interventions | – not applicable – |
| Coding of interventions | necessary |
| Source of intervention data | coding done by authors |
| Availability of data on exposure | coded data not available |
| **Methodological approach** | |
| Empirical approach | parametric |
| Use of exposure variation | both variation over time and between populations |
| Method | non-mechanistic model (generalized linear model) |
| Code availability | none |
| **Effect assessment** | |
| Reporting of intervention effect | quantification of change in outcome values |
| Measure of effect | reproduction number |
| Reporting of uncertainty | yes |
| Sensitivity analysis | epidemiological parameters varied, model specification varied, same analysis with (sub)population(s) excluded |
| Subgroup assessment | geographic areas (regions), socioeconomic indicators (population density, age, health index: global health security index) |

**Bönisch et al. 2020 (Analysis 1)**

**Study information**

– continued on next page –

| | |
|---|---|
| Author | Bönisch S, Wegscheider K, Krause L, Sehner S, Wiegel S, Zapf A, Moser S, Becher H |
| Title | Effects of Coronavirus Disease (COVID-19) Related Contact Restrictions in Germany, March to May 2020, on the Mobility and Relation to Infection Patterns |
| Year | 2020 |
| Journal | Frontiers in Public Health |
| DOI | 10.3389/fpubh.2020.568287 |
| Date received | 2020-05-31 |
| Date accepted | 2020-09-18 |
| **Study setting** | |
| Number of populations included | single |
| Level of populations included | national (Germany) |
| Geographic areas covered | Europe |
| Number of countries covered | single country (Germany) |
| Study period | start and end date span first epidemic wave: behavioral |
| **Outcome** | |
| Raw outcome | behavioral outcome (mobility: median daily distance) |
| Time resolution of raw outcome | daily |
| Computed outcome | none |
| Method to obtain the computed outcome | – not applicable – |
| Data source | mobility data from corporate organizations (GapFish) |
| Data availability | data not accessible |
| **Intervention** | |
| Terminology for interventions | – not applicable – |

– continued on next page –

| | |
|---|---|
| Terminology for the specific type of non-pharmaceutical interventions | contact restrictions |
| Exposure types | one combination of interventions (lockdown period from 23.03 to 05.04.2020 following the closure of schools and non-essential public services, TV adress by Angela Merkel, contact restrictions) |
| Types of single interventions | – not applicable – |
| Coding of interventions | not necessary |
| Source of intervention data | government or news websites |
| Availability of data on exposure | raw data documented in the manuscript |
| **Methodological approach** | |
| Empirical approach | counterfactual |
| Use of exposure variation | only variation over time for a single population |
| Method | non-mechanistic model (time series model) |
| Code availability | none |
| **Effect assessment** | |
| Reporting of intervention effect | quantification of change in outcome values |
| Measure of effect | mobility: median daily distance |
| Reporting of uncertainty | yes |
| Sensitivity analysis | none |
| Subgroup assessment | socioeconomic indicators (age, gender) |

**Bönisch et al. 2020 (Analysis 2, mobility as exposure)**

| | |
|---|---|
| **Study information** | |
| Author | Bönisch S, Wegscheider K, Krause L, Sehner S, Wiegel S, Zapf A, Moser S, Becher H |

– continued on next page –

| | |
|---|---|
| Title | Effects of Coronavirus Disease (COVID-19) Related Contact Restrictions in Germany, March to May 2020, on the Mobility and Relation to Infection Patterns |
| Year | 2020 |
| Journal | Frontiers in Public Health |
| DOI | 10.3389/fpubh.2020.568287 |
| Date received | 31.05.2020 |
| Date accepted | 18.09.2020 |
| **Study setting** | |
| Number of populations included | single |
| Level of populations included | national (Germany) |
| Geographic areas covered | Europe |
| Number of countries covered | single country (Germany) |
| Study period | start and end date span first epidemic wave |
| **Outcome** | |
| Raw outcome | epidemiological population-level outcome (cases, reproduction number) |
| Time resolution of raw outcome | daily |
| Computed outcome | none |
| Method to obtain the computed outcome | – not applicable – |
| Data source | data from (sub)national authorities |
| Data availability | data access via source |
| **Intervention** | |
| Terminology for interventions | – not applicable – |
| Terminology for the specific type of non-pharmaceutical interventions | contact restrictions |

– continued on next page –

| | |
|---|---|
| Exposure types | mobility (relative reduction in median daily distance in the study period) |
| Types of single interventions | – not applicable – |
| Coding of interventions | – not applicable – |
| Source of intervention data | mobility data by corporate organizations (GapFish) |
| Availability of data on exposure | data not accessible |
| **Methodological approach** | |
| Empirical approach | descriptive |
| Use of exposure variation | only variation over time for a single population |
| Method | description of change over time (description of time course) |
| Code availability | none |
| **Effect assessment** | |
| Reporting of intervention effect | qualitative statement |
| Measure of effect | cases, reproduction number |
| Reporting of uncertainty | – not applicable – |
| Sensitivity analysis | none |
| Subgroup assessment | none |

**Braithwaite et al. 2020**

| | |
|---|---|
| **Study information** | |
| Author | Braithwaite J, Tran Y, Ellis LA, Westbrook J |
| Title | The 40 Health Systems, COVID-19 (40HS, C-19) Study |
| Year | 2020 |
| Journal | International Journal for Quality in Health Care |
| DOI | 10.1093/intqhc/mzaa113 |

– continued on next page –

| | |
|---|---|
| Date received | 2020-06-24 |
| Date accepted | 2020-09-08 |
| **Study setting** | |
| Number of populations included | multiple |
| Level of populations included | national (40 mostly OECD countries) |
| Geographic areas covered | Europe, Middle East and Africa, Asia, Oceania, Central and South America, North America |
| Number of countries covered | multiple countries |
| Study period | same end date for several populations with diverse epidemic trajectories |
| **Outcome** | |
| Raw outcome | epidemiological population-level outcome (cases, deaths) |
| Time resolution of raw outcome | daily |
| Computed outcome | none |
| Method to obtain the computed outcome | – not applicable – |
| Data source | data from publicly available cross-country selections (OurWorldInData) |
| Data availability | data access via source |
| **Intervention** | |
| Terminology for interventions | measures |
| Terminology for the specific type of non-pharmaceutical interventions | public health |
| Exposure types | stringency index (OxCGRT Stringency Index (at 3 time points)) |
| Types of single interventions | – not applicable – |
| Coding of interventions | necessary |
| Source of intervention data | use of externally coded data (OxCGRT) |
| Availability of data on exposure | access to externally coded data via source |

– continued on next page –

| | |
|---|---|
| **Methodological approach** | |
| Empirical approach | descriptive |
| Use of exposure variation | only variation between populations |
| Method | comparison of populations |
| Code availability | none |
| **Effect assessment** | |
| Reporting of intervention effect | quantification of change in outcome values |
| Measure of effect | cases, deaths |
| Reporting of uncertainty | yes |
| Sensitivity analysis | none |
| Subgroup assessment | none |

**Brauner et al. 2021**

| | |
|---|---|
| **Study information** | |
| Author | Brauner JM, Mindermann S, Sharma M, Johnston D, Salvatier J, Gavenčiak T, Stephenson AB, Leech G, Altman G, Mikulik V, Norman AJ, Monrad JT, Besiroglu T, Ge H, Hartwick MA, Teh YW, Chindelevitch L, Gal Y, Kulveit J |
| Title | Inferring the Effectiveness of Government Interventions against COVID-19 |
| Year | 2021 |
| Journal | Science |
| DOI | 10.1126/science.abd9338 |
| Date received | 2020-07-21 |
| Date accepted | 2020-12-08 |

– continued on next page –

| | |
|---|---|
| **Study setting** | |
| Number of populations included | multiple |
| Level of populations included | national (34 European, 7 non-European countries) |
| Geographic areas covered | Asia, Central and South America, Europe, Middle East and Africa, Oceania |
| Number of countries covered | multiple countries |
| Study period | start and end date span first epidemic wave |
| **Outcome** | |
| Raw outcome | epidemiological population-level outcome (cases, deaths) |
| Time resolution of raw outcome | daily |
| Computed outcome | none |
| Method to obtain the computed outcome | – not applicable – |
| Data source | data from publicly available cross-country selections (JHU) |
| Data availability | data made available by the authors |
| **Intervention** | |
| Terminology for interventions | interventions |
| Terminology for the specific type of non-pharmaceutical interventions | non-pharmaceutical |
| Exposure types | multiple separate interventions (bans of small gatherings, bans of large gatherings, venue closure, workplace closure, stay-at-home order, school closure) |
| Types of single interventions | bans of small gatherings, bans of large gatherings, venue closure, workplace closure, stay-at-home order, school closure |
| Coding of interventions | necessary |
| Source of intervention data | coding done by authors |
| Availability of data on exposure | coded data made available by the authors |

– continued on next page –

| | |
|---|---|
| **Methodological approach** | |
| Empirical approach | parametric |
| Use of exposure variation | both variation over time and between populations |
| Method | mechanistic model (semi-mechanistic Bayesian transmission model) |
| Latent variable functionally linked to intervention | reproduction number |
| Code availability | publicly available |
| **Effect assessment** | |
| Reporting of intervention effect | quantification of change in outcome values |
| Measure of effect | reproduction number |
| Reporting of uncertainty | yes |
| Sensitivity analysis | different or modified outcome used (number of confirmed cases adjusted using time-varying ascertainment rate estimated by study), different coding of interventions used (specific NPIs left out), model specification varied, start or end date of study period varied, epidemiological parameters varied, same analysis with (sub)population(s) excluded |
| Subgroup assessment | none |

**Candido et al. 2020 (Analysis 1)**

| |
|---|
| **Study information** |

– continued on next page –

| | |
|---|---|
| Author | Candido DS, Claro IM, de Jesus JG, Souza WM, Moreira FR, Dellicour S, Mellan TA, du Plessis L, Pereira RH, Sales FC, Manuli ER, Thézé J, Almeida L, Menezes MT, Voloch CM, Fumagalli MJ, Coletti TM, da Silva CA, Ramundo MS, Amorim MR, Hoeltgebaum HH, Mishra S, Gill MS, Carvalho LM, Buss LF, Prete CA, Ashworth J, Nakaya HI, Peixoto PS, Brady OJ, Nicholls SM, Tanuri A, Rossi ÁD, Braga CK, Gerber AL, de C. Guimarães AP, Gaburo N, Alencar CS, Ferreira AC, Lima CX, Levi JE, Granato C, Ferreira GM, Francisco RS, Granja F, Garcia MT, Moretti ML, Perroud MW, Castiñeiras TM, Lazari CS, Hill SC, de Souza Santos AA, Simeoni CL, Forato J, Sposito AC, Schreiber AZ, Santos MN, de Sá CZ, Souza RP, Resende-Moreira LC, Teixeira MM, Hubner J, Leme PA, Moreira RG, Nogueira ML, Ferguson NM, Costa SF, Proenca-Modena JL, Vasconcelos AT, Bhatt S, Lemey P, Wu CH, Rambaut A, Loman NJ, Aguiar RS, Pybus OG, Sabino EC, And NR |
| Title | Evolution and Epidemic Spread of SARS-CoV-2 in Brazil |
| Year | 2020 |
| Journal | Science |
| DOI | 10.1126/science.abd2161 |
| Date received | 2020-06-10 |
| Date accepted | 2020-07-16 |
| **Study setting** | |
| Number of populations included | multiple |
| Level of populations included | both national and subnational (Brazil and its major cities) |
| Geographic areas covered | Central and South America |
| Number of countries covered | single country (Brazil) |
| Study period | end date in growth phase of wave |
| **Outcome** | |
| Raw outcome | epidemiological individual-level outcome (genome sequence data), behavioral outcome (mobility: air travel) |

– continued on next page –

| | |
|---|---|
| Time resolution of raw outcome | daily |
| Computed outcome | other (virus migration) |
| Method to obtain the computed outcome | phylodynamic model (phylogeographic analysis) |
| Data source | data from (sub)national authorities (Civil Aviation Agency: mobility), data collected by authors (viral sequence data) |
| Data availability | data made available by the authors |
| **Intervention** | |
| Terminology for interventions | interventions |
| Terminology for the specific type of non-pharmaceutical interventions | non-pharmaceutical |
| Exposure types | single combination of interventions (school closure, retail and service closure, emergency declaration) |
| Types of single interventions | – not applicable – |
| Coding of interventions | not necessary |
| Source of intervention data | – could not be evaluated – |
| Availability of data on exposure | raw data documented in the manuscript |
| **Methodological approach** | |
| Empirical approach | descriptive |
| Use of exposure variation | only variation over time for multiple populations |
| Method | description of change over time (description of time course) |
| Code availability | publicly available |
| **Effect assessment** | |
| Reporting of intervention effect | qualitative statement |
| Measure of effect | mobility: air travel, virus migration |
| Reporting of uncertainty | – not applicable – |

– continued on next page –

| | |
|---|---|
| Sensitivity analysis | none |
| Subgroup assessment | none |

| **Candido et al. 2020 (Analysis 2, mobility as exposure)** | |
|---|---|
| **Study information** | |
| Author | Candido DS, Claro IM, de Jesus JG, Souza WM, Moreira FR, Dellicour S, Mellan TA, du Plessis L, Pereira RH, Sales FC, Manuli ER, Thézé J, Almeida L, Menezes MT, Voloch CM, Fumagalli MJ, Coletti TM, da Silva CA, Ramundo MS, Amorim MR, Hoeltgebaum HH, Mishra S, Gill MS, Carvalho LM, Buss LF, Prete CA, Ashworth J, Nakaya HI, Peixoto PS, Brady OJ, Nicholls SM, Tanuri A, Rossi ÁD, Braga CK, Gerber AL, de C. Guimarães AP, Gaburo N, Alencar CS, Ferreira AC, Lima CX, Levi JE, Granato C, Ferreira GM, Francisco RS, Granja F, Garcia MT, Moretti ML, Perroud MW, Castiñeiras TM, Lazari CS, Hill SC, de Souza Santos AA, Simeoni CL, Forato J, Sposito AC, Schreiber AZ, Santos MN, de Sá CZ, Souza RP, Resende-Moreira LC, Teixeira MM, Hubner J, Leme PA, Moreira RG, Nogueira ML, Ferguson NM, Costa SF, Proenca-Modena JL, Vasconcelos AT, Bhatt S, Lemey P, Wu CH, Rambaut A, Loman NJ, Aguiar RS, Pybus OG, Sabino EC, And NR |
| Title | Evolution and Epidemic Spread of SARS-CoV-2 in Brazil |
| Year | 2020 |
| Journal | Science |
| DOI | 10.1126/science.abd2161 |
| Date received | 10.06.2020 |
| Date accepted | 16.07.2020 |
| **Study setting** | |
| Number of populations included | multiple |
| Level of populations included | both national and subnational (Brazil and its major cities) |

– continued on next page –

| | |
|---|---|
| Geographic areas covered | Central and South America |
| Number of countries covered | single country (Brazil) |
| Study period | end date in growth phase of wave |
| **Outcome** | |
| Raw outcome | epidemiological population-level outcome (cases, deaths) |
| Time resolution of raw outcome | daily |
| Computed outcome | none |
| Method to obtain the computed outcome | – not applicable – |
| Data source | data from (sub)national authorities |
| Data availability | data made available by the authors |
| **Intervention** | |
| Terminology for interventions | interventions |
| Terminology for the specific type of non-pharmaceutical interventions | non-pharmaceutical |
| Exposure types | mobility (time spent in various locations, social isolation index) |
| Types of single interventions | – not applicable – |
| Coding of interventions | – not applicable – |
| Source of intervention data | mobility data by corporate organizations (Google: time spent in various locations, InLoco: social isolation index) |
| Availability of data on exposure | data access via source |
| **Methodological approach** | |
| Empirical approach | parametric |
| Use of exposure variation | only variation over time for multiple populations |
| Method | mechanistic model (semi-mechanistic Bayesian transmission model) |
| Code availability | publicly available |

– continued on next page –

| | |
|---|---|
| **Effect assessment** | |
| Reporting of intervention effect | qualitative statement |
| Measure of effect | reproduction number |
| Reporting of uncertainty | – not applicable – |
| Sensitivity analysis | none |
| Subgroup assessment | none |

**Cao et al. 2020**

| | |
|---|---|
| **Study information** | |
| Author | Cao Y, Hiyoshi A, Montgomery S |
| Title | COVID-19 Case-Fatality Rate and Demographic and Socioeconomic Influencers: Worldwide Spatial Regression Analysis Based on Country-Level Data |
| Year | 2020 |
| Journal | BMJ Open |
| DOI | 10.1136/bmjopen-2020-043560 |
| Date received | 2020-08-17 |
| Date accepted | 2020-09-16 |
| **Study setting** | |
| Number of populations included | multiple |
| Level of populations included | national (209 countries/territories worldwide) |
| Geographic areas covered | Europe, Middle East and Africa, Asia, Oceania, Central and South America, North America |
| Number of countries covered | multiple countries |

– continued on next page –

| | |
|---|---|
| Study period | start and end date span first epidemic wave |
| **Outcome** | |
| Raw outcome | epidemiological population-level outcome (cases, deaths) |
| Time resolution of raw outcome | daily |
| Computed outcome | summary statistic (case-fatality rate) |
| Method to obtain the computed outcome | simple computation |
| Data source | data from publicly available cross-country selections (OurWorldInData) |
| Data availability | data access via source |
| **Intervention** | |
| Terminology for interventions | measures |
| Terminology for the specific type of non-pharmaceutical interventions | quarantine |
| Exposure types | stringency index (OxCGRT Stringency Index) |
| Types of single interventions | – not applicable – |
| Coding of interventions | necessary |
| Source of intervention data | use of externally coded data (OxCGRT) |
| Availability of data on exposure | access to externally coded data via source |
| **Methodological approach** | |
| Empirical approach | parametric |
| Use of exposure variation | only variation between populations |
| Method | non-mechanistic model (other: spatial regression model) |
| Code availability | none |
| **Effect assessment** | |
| Reporting of intervention effect | quantification of change in outcome values |
| Measure of effect | case-fatality rate |

– continued on next page –

| | |
|---|---|
| Reporting of uncertainty | yes |
| Sensitivity analysis | different or modified outcome used (crude cause-specific death rate instead of CFR) |
| Subgroup assessment | public health response (testing policy), socioeconomic indicators (income) |

| **Castaneda & Saygili 2020** | |
|---|---|
| **Study information** | |
| Author | Castaneda MA, Saygili M |
| Title | The Effect of Shelter-in-Place Orders on Social Distancing and the Spread of the COVID-19 Pandemic: A Study of Texas |
| Year | 2020 |
| Journal | Frontiers in Public Health |
| DOI | 10.3389/fpubh.2020.596607 |
| Date received | 2020-08-27 |
| Date accepted | 2020-11-05 |
| **Study setting** | |
| Number of populations included | multiple |
| Level of populations included | subnational (Counties in Texas) |
| Geographic areas covered | North America |
| Number of countries covered | single country (US) |
| Study period | end date in growth phase of wave |
| **Outcome** | |
| Raw outcome | epidemiological population-level outcome (cases, deaths), behavioral outcome (mobility: share of population sheltering in home) |
| Time resolution of raw outcome | daily |

– continued on next page –

| | |
|---|---|
| Computed outcome | measure of epidemic trend (growth rate, growth rate: deaths) |
| Method to obtain the computed outcome | simple computation |
| Data source | data from public media outlets (New York Times: cases), mobility data from corporate organizations (SafeGraph: mobility) |
| Data availability | data access via source |
| **Intervention** | |
| Terminology for interventions | measures |
| Terminology for the specific type of non-pharmaceutical interventions | social distancing |
| Exposure types | multiple separate interventions (emergency declaration, stay-at-home order (county- and statewide)) |
| Types of single interventions | declaration of a state of emergency, stay-at-home order |
| Coding of interventions | necessary |
| Source of intervention data | use of externally coded data (National Association of Counties) |
| Availability of data on exposure | access to externally coded data via source |
| **Methodological approach** | |
| Empirical approach | parametric |
| Use of exposure variation | both variation over time and between populations |
| Method | non-mechanistic model (generalized linear model) |
| Code availability | none |
| **Effect assessment** | |
| Reporting of intervention effect | quantification of change in outcome values |
| Measure of effect | growth rate, growth rate: deaths, mobility: share of population sheltering in home |
| Reporting of uncertainty | yes |

– continued on next page –

| | |
|---|---|
| Sensitivity analysis | none |
| Subgroup assessment | public health response (subject to other measures) |

| **Cento et al. 2020** | |
|---|---|
| **Study information** | |
| Author | Cento V, Alteri C, Merli M, Ruscio FD, Tartaglione L, Rossotti R, Travi G, Vecchi M, Raimondi A, Nava A, Colagrossi L, Fumagalli R, Ughi N, Epis OM, Fanti D, Beretta A, Galbiati F, Scaglione F, Vismara C, Puoti M, Campisi D, Perno CF |
| Title | Effectiveness of Infection-Containment Measures on SARS-CoV-2 Seroprevalence and Circulation from May to July 2020, in Milan, Italy |
| Year | 2020 |
| Journal | PLOS ONE |
| DOI | 10.1371/journal.pone.0242765 |
| Date received | 2020-08-10 |
| Date accepted | 2020-11-09 |
| **Study setting** | |
| Number of populations included | single |
| Level of populations included | subnational (Milan (Italy)) |
| Geographic areas covered | Europe |
| Number of countries covered | single country (Italy) |
| Study period | start and end date span first epidemic wave |
| **Outcome** | |
| Raw outcome | epidemiological population-level outcome (test positivity rate: anti-N IgG seroprevalence) |

– continued on next page –

| | |
|---|---|
| Time resolution of raw outcome | daily |
| Computed outcome | none |
| Method to obtain the computed outcome | – not applicable – |
| Data source | data collected by authors |
| Data availability | data access via source |
| **Intervention** | |
| Terminology for interventions | measures |
| Terminology for the specific type of non-pharmaceutical interventions | containment |
| Exposure types | one combination of interventions (lockdown of March 9) |
| Types of single interventions | – not applicable – |
| Coding of interventions | not necessary |
| Source of intervention data | – could not be evaluated – |
| Availability of data on exposure | raw data documented in the manuscript |
| **Methodological approach** | |
| Empirical approach | descriptive |
| Use of exposure variation | only variation over time for a single population |
| Method | description of change over time (comparison of time periods) |
| Code availability | none |
| **Effect assessment** | |
| Reporting of intervention effect | comparison of outcome values |
| Measure of effect | test positivity rate: anti-N IgG seroprevalence |
| Reporting of uncertainty | yes |
| Sensitivity analysis | none |

– continued on next page –

| | |
|---|---|
| Subgroup assessment | none |

#### Chakraborty & Ghosh 2020

| | |
|---|---|
| **Study information** | |
| Author | Chakraborty T, Ghosh I |
| Title | Real-Time Forecasts and Risk Assessment of Novel Coronavirus (COVID-19) Cases: A Data-Driven Analysis |
| Year | 2020 |
| Journal | Chaos, Solitons & Fractals |
| DOI | 10.1016/j.chaos.2020.109850 |
| Date received | 2020-04-10 |
| Date accepted | 2020-04-24 |
| **Study setting** | |
| Number of populations included | multiple |
| Level of populations included | national (50 countries around the globe) |
| Geographic areas covered | Europe, Middle East and Africa, Asia, Oceania, Central and South America, North America |
| Number of countries covered | multiple countries |
| Study period | same end date for several populations with diverse epidemic trajectories |
| **Outcome** | |
| Raw outcome | epidemiological population-level outcome (cases, deaths) |
| Time resolution of raw outcome | daily |
| Computed outcome | epidemiological parameter (case-fatality rate) |
| Method to obtain the computed outcome | simple computation |

– continued on next page –

| | |
|---|---|
| Data source | data from publicly available cross-country selections (Worldometer) |
| Data availability | data made available by the authors |
| **Intervention** | |
| Terminology for interventions | – not applicable – |
| Terminology for the specific type of non-pharmaceutical interventions | lockdown |
| Exposure types | one combination of interventions (lockdown with specific date for each country (only days since lockdown are provided)) |
| Types of single interventions | – not applicable – |
| Coding of interventions | necessary |
| Source of intervention data | – could not be evaluated – |
| Availability of data on exposure | coded data made available by the authors |
| **Methodological approach** | |
| Empirical approach | parametric |
| Use of exposure variation | only variation between populations |
| Method | non-mechanistic model (other: feature importance from machine learning model with lockdown period as predictor) |
| Code availability | publicly available |
| **Effect assessment** | |
| Reporting of intervention effect | quantification of change in outcome values |
| Measure of effect | case-fatality rate |
| Reporting of uncertainty | no |
| Sensitivity analysis | none |
| Subgroup assessment | none |

| Chatterjee et al. 2020 | |
|---|---|
| **Study information** | |
| Author | Chatterjee S, Sarkar A, Chatterjee S, Karmakar M, Paul R |
| Title | Studying the Progress of COVID-19 Outbreak in India Using SIRD Model |
| Year | 2020 |
| Journal | Indian Journal of Physics |
| DOI | 10.1007/s12648-020-01766-8 |
| Date received | 2020-05-15 |
| Date accepted | 2020-06-03 |
| **Study setting** | |
| Number of populations included | multiple |
| Level of populations included | both national and subnational (India and 4 states, South Korea, USA, Germany, Spain) |
| Geographic areas covered | Asia |
| Number of countries covered | single country (India) |
| Study period | same end date for several populations with diverse epidemic trajectories |
| **Outcome** | |
| Raw outcome | epidemiological population-level outcome (cases, deaths) |
| Time resolution of raw outcome | daily |
| Computed outcome | none |
| Method to obtain the computed outcome | – not applicable – |
| Data source | data from publicly available open-source projects (covid19india.org: India), data from publicly available cross-country selections (Worldometer: other countries) |
| Data availability | data access via source |
| **Intervention** | |

– continued on next page –

| | |
|---|---|
| Terminology for interventions | measures |
| Terminology for the specific type of non-pharmaceutical interventions | containment |
| Exposure types | one combination of interventions (Indian National Lockdown) |
| Types of single interventions | – not applicable – |
| Coding of interventions | not necessary |
| Source of intervention data | – could not be evaluated – |
| Availability of data on exposure | raw data documented in the manuscript |
| **Methodological approach** | |
| Empirical approach | parametric |
| Use of exposure variation | only variation over time for multiple populations |
| Method | mechanistic model (compartmental single-population transmission model) |
| Latent variable functionally linked to intervention | transmission rate |
| Code availability | none |
| **Effect assessment** | |
| Reporting of intervention effect | quantification of change in outcome values |
| Measure of effect | transmission rate, reproduction number |
| Reporting of uncertainty | no |
| Sensitivity analysis | none |
| Subgroup assessment | none |

**Chaudhry et al. 2020**

| |
|---|
| **Study information** |

– continued on next page –

| | |
|---|---|
| Author | Chaudhry R, Dranitsaris G, Mubashir T, Bartoszko J, Riazi S |
| Title | A Country Level Analysis Measuring the Impact of Government Actions, Country Preparedness and Socioeconomic Factors on COVID-19 Mortality and Related Health Outcomes |
| Year | 2020 |
| Journal | EClinicalMedicine |
| DOI | 10.1016/j.eclinm.2020.100464 |
| Date received | 2020-05-05 |
| Date accepted | 2020-07-02 |
| **Study setting** | |
| Number of populations included | multiple |
| Level of populations included | national (50 countries around the globe) |
| Geographic areas covered | Europe, Middle East and Africa, Asia, Oceania, Central and South America, North America |
| Number of countries covered | multiple countries |
| Study period | same end date for several populations with diverse epidemic trajectories |
| **Outcome** | |
| Raw outcome | epidemiological population-level outcome (cases, recovered cases, severe cases, deaths) |
| Time resolution of raw outcome | daily |
| Computed outcome | none |
| Method to obtain the computed outcome | – not applicable – |
| Data source | data from publicly available cross-country selections (JHU: other countries, WHO: other countries, Worldometer: other countries), data from (sub)national authorities (US) |
| Data availability | data access via source |

– continued on next page –

| | |
|---|---|
| **Intervention** | |
| Terminology for interventions | policies, measures |
| Terminology for the specific type of non-pharmaceutical interventions | health policies, containment |
| Exposure types | multiple separate interventions (travel restrictions (types of travel restrictions, partial border closures, complete border closures) stay-at-home order (partial lockdown, complete lockdown, stay-at-home order)) |
| Types of single interventions | international travel restrictions, stay-at-home order |
| Coding of interventions | necessary |
| Source of intervention data | coding done by authors |
| Availability of data on exposure | coded data not available |
| **Methodological approach** | |
| Empirical approach | parametric |
| Use of exposure variation | only variation between populations |
| Method | non-mechanistic model (generalized linear model) |
| Code availability | none |
| **Effect assessment** | |
| Reporting of intervention effect | quantification of change in outcome values |
| Measure of effect | cases, recovered, severe cases, deaths |
| Reporting of uncertainty | yes |
| Sensitivity analysis | none |
| Subgroup assessment | none |

| Chaves et al. 2020 | |
|---|---|
| **Study information** | |
| Author | Chaves LF, Hurtado LA, Rojas MR, Friberg MD, Rodríguez RM, Avila-Aguero ML |
| Title | COVID-19 Basic Reproduction Number and Assessment of Initial Suppression Policies in Costa Rica |
| Year | 2020 |
| Journal | Mathematical Modelling of Natural Phenomena |
| DOI | 10.1051/mmnp/2020019 |
| Date received | 2020-04-07 |
| Date accepted | 2020-05-07 |
| **Study setting** | |
| Number of populations included | single |
| Level of populations included | national (Costa Rica) |
| Geographic areas covered | Central and South America |
| Number of countries covered | single country (Costa Rica) |
| Study period | end date at peak of wave |
| **Outcome** | |
| Raw outcome | epidemiological population-level outcome (cases) |
| Time resolution of raw outcome | daily |
| Computed outcome | epidemiological parameter (reproduction number) |
| Method to obtain the computed outcome | statistical estimation of reproduction number (Thompson et al.), compartmental transmission model (single-population) |
| Data source | data from (sub)national authorities |
| Data availability | data access via source |
| **Intervention** | |
| Terminology for interventions | policies |

– continued on next page –

| | |
|---|---|
| Terminology for the specific type of non-pharmaceutical interventions | suppression |
| Exposure types | phases of interventions (timeline of measures divided into three phases) |
| Types of single interventions | – not applicable – |
| Coding of interventions | not necessary |
| Source of intervention data | government or news websites |
| Availability of data on exposure | raw data documented in the manuscript |
| **Methodological approach** | |
| Empirical approach | descriptive |
| Use of exposure variation | only variation over time for a single population |
| Method | description of change over time (comparison of time periods) |
| Code availability | none |
| **Effect assessment** | |
| Reporting of intervention effect | comparison of outcome values |
| Measure of effect | reproduction number |
| Reporting of uncertainty | yes |
| Sensitivity analysis | none |
| Subgroup assessment | none |

**Cheema et al. 2021**

| | |
|---|---|
| **Study information** | |
| Author | Cheema SA, Kifayat T, Rahman AR, Khan U, Zaib A, Khan I, Nisar KS |
| Title | Is Social Distancing, and Quarantine Effective in Restricting COVID-19 Outbreak? Statistical Evidences from Wuhan, China |

– continued on next page –

| | |
|---|---|
| Year | 2021 |
| Journal | Computers, Materials & Continua |
| DOI | 10.32604/cmc.2020.012096 |
| Date received | 2020-06-14 |
| Date accepted | 2020-07-07 |
| **Study setting** | |
| Number of populations included | single |
| Level of populations included | subnational (Wuhan) |
| Geographic areas covered | Asia |
| Number of countries covered | single country (China) |
| Study period | late start |
| **Outcome** | |
| Raw outcome | epidemiological population-level outcome (cases, deaths) |
| Time resolution of raw outcome | daily |
| Computed outcome | none |
| Method to obtain the computed outcome | – not applicable – |
| Data source | data from (sub)national authorities |
| Data availability | data access via source |
| **Intervention** | |
| Terminology for interventions | measures, policies |
| Terminology for the specific type of non-pharmaceutical interventions | social distancing |
| Exposure types | one combination of interventions (Wuhan City Lockdown) |
| Types of single interventions | – not applicable – |
| Coding of interventions | not necessary |

– continued on next page –

| | |
|---|---|
| Source of intervention data | – could not be evaluated – |
| Availability of data on exposure | raw data documented in the manuscript |
| **Methodological approach** | |
| Empirical approach | descriptive |
| Use of exposure variation | only variation over time for a single population |
| Method | description of change over time (comparison of time periods) |
| Code availability | none |
| **Effect assessment** | |
| Reporting of intervention effect | quantification of change in outcome values |
| Measure of effect | cases, deaths |
| Reporting of uncertainty | yes |
| Sensitivity analysis | none |
| Subgroup assessment | none |

| **Chen et al. 2020a** | |
|---|---|
| **Study information** | |
| Author | Chen SL, Yen AM, Lai CC, Hsu CY, Chan CC, Chen TH |
| Title | An Index for Lifting Social Distancing during the COVID-19 Pandemic: Algorithm Recommendation for Lifting Social Distancing |
| Year | 2020 |
| Journal | Journal of Medical Internet Research |
| DOI | 10.2196/22469 |
| Date received | 2020-07-13 |
| Date accepted | 2020-09-04 |

– continued on next page –

| | |
|---|---|
| **Study setting** | |
| Number of populations included | multiple |
| Level of populations included | national (186 countries worldwide) |
| Geographic areas covered | Europe, Middle East and Africa, Asia, Oceania, Central and South America, North America |
| Number of countries covered | multiple countries |
| Study period | same end date for several populations with diverse epidemic trajectories |
| **Outcome** | |
| Raw outcome | epidemiological population-level outcome (cases, deaths, recovered cases) |
| Time resolution of raw outcome | daily |
| Computed outcome | other (lifting social distancing index) |
| Method to obtain the computed outcome | simple computation |
| Data source | data from publicly available cross-country selections (JHU) |
| Data availability | data access via source |
| **Intervention** | |
| Terminology for interventions | measures |
| Terminology for the specific type of non-pharmaceutical interventions | social distancing |
| Exposure types | all interventions together |
| Types of single interventions | – not applicable – |
| Coding of interventions | – not applicable – |
| Source of intervention data | – not applicable – |
| Availability of data on exposure | – not applicable – |
| **Methodological approach** | |
| Empirical approach | descriptive |

– continued on next page –

| | |
|---|---|
| Use of exposure variation | only variation over time for multiple populations |
| Method | description of change over time (description of time course) |
| Code availability | none |
| **Effect assessment** | |
| Reporting of intervention effect | qualitative statement |
| Measure of effect | lifting social distancing index |
| Reporting of uncertainty | – not applicable – |
| Sensitivity analysis | none |
| Subgroup assessment | none |

**Chen et al. 2020b**

| | |
|---|---|
| **Study information** | |
| Author | Chen S, Chen Q, Yang W, Xue L, Liu Y, Yang J, Wang C, Bärnighausen T |
| Title | Buying Time for an Effective Epidemic Response: The Impact of a Public Holiday for Outbreak Control on COVID-19 Epidemic Spread |
| Year | 2020 |
| Journal | Engineering |
| DOI | 10.1016/j.eng.2020.07.018 |
| Date received | 2020-05-07 |
| Date accepted | 2020-07-13 |
| **Study setting** | |
| Number of populations included | multiple |
| Level of populations included | both national and subnational (China Hubei province and other provinces) |
| Geographic areas covered | Asia |

– continued on next page –

| | |
|---|---|
| Number of countries covered | single country (China) |
| Study period | end date in growth phase of wave |
| **Outcome** | |
| Raw outcome | epidemiological population-level outcome (cases, deaths, recovered cases) |
| Time resolution of raw outcome | daily |
| Computed outcome | none |
| Method to obtain the computed outcome | – not applicable – |
| Data source | data from (sub)national authorities |
| Data availability | data access via source |
| **Intervention** | |
| Terminology for interventions | measures |
| Terminology for the specific type of non-pharmaceutical interventions | control |
| Exposure types | one combination of interventions (Wuhan City Lockdown) |
| Types of single interventions | – not applicable – |
| Coding of interventions | not necessary |
| Source of intervention data | government or news websites |
| Availability of data on exposure | raw data documented in the manuscript |
| **Methodological approach** | |
| Empirical approach | parametric |
| Use of exposure variation | both variation over time and between populations |
| Method | mechanistic model (compartmental single-population transmission model) |
| Latent variable functionally linked to intervention | transmission rate |
| Code availability | none |

– continued on next page –

| | |
|---|---|
| **Effect assessment** | |
| Reporting of intervention effect | quantification of change in outcome values |
| Measure of effect | transmission rate |
| Reporting of uncertainty | no |
| Sensitivity analysis | none |
| Subgroup assessment | none |

**Chen et al. 2020c**

| | |
|---|---|
| **Study information** | |
| Author | Chen YT, Yen YF, Yu SH, Su EC |
| Title | An Examination on the Transmission of COVID-19 and the Effect of Response Strategies: A Comparative Analysis |
| Year | 2020 |
| Journal | International Journal of Environmental Research and Public Health |
| DOI | 10.3390/ijerph17165687 |
| Date received | 2020-06-24 |
| Date accepted | 2020-07-31 |
| **Study setting** | |
| Number of populations included | multiple |
| Level of populations included | national (six countries: China, Korea, Japan, Italy, the USA, Brazil) |
| Geographic areas covered | Asia, Central and South America, Europe, North America |
| Number of countries covered | multiple countries |
| Study period | start and end date span first epidemic wave |
| **Outcome** | |

– continued on next page –

| | |
|---|---|
| Raw outcome | epidemiological population-level outcome (cases) |
| Time resolution of raw outcome | daily |
| Computed outcome | summary statistic (time to peak, maximum percentage reduction relative to peak after peak) |
| Method to obtain the computed outcome | simple computation |
| Data source | data from publicly available cross-country selections (WHO) |
| Data availability | data access via source |
| **Intervention** | |
| Terminology for interventions | strategies |
| Terminology for the specific type of non-pharmaceutical interventions | response |
| Exposure types | all interventions together |
| Types of single interventions | – not applicable – |
| Coding of interventions | – not applicable – |
| Source of intervention data | – not applicable – |
| Availability of data on exposure | – not applicable – |
| **Methodological approach** | |
| Empirical approach | descriptive |
| Use of exposure variation | both variation over time and between populations |
| Method | comparison of populations |
| Code availability | none |
| **Effect assessment** | |
| Reporting of intervention effect | qualitative statement |
| Measure of effect | time to peak, maximum percentage reduction relative to peak after peak |
| Reporting of uncertainty | – not applicable – |

– continued on next page –

| | |
|---|---|
| Sensitivity analysis | none |
| Subgroup assessment | none |

**Cheng et al. 2020a**

| | |
|---|---|
| **Study information** | |
| Author | Cheng W, Yu Z, Liu S, Sun W, Ling F, Pan J, Chen E |
| Title | Successful Interruption of Seasonal Influenza Transmission under the COVID-19 Rapid Response in Zhejiang Province, China |
| Year | 2020 |
| Journal | Public Health |
| DOI | 10.1016/j.puhe.2020.10.011 |
| Date received | 2020-08-04 |
| Date accepted | 2020-09-13 |
| **Study setting** | |
| Number of populations included | single |
| Level of populations included | subnational (Zhejiang Province (China)) |
| Geographic areas covered | Asia |
| Number of countries covered | single country (China) |
| Study period | start and end date span first epidemic wave |
| **Outcome** | |
| Raw outcome | epidemiological population-level outcome (surrogate disease: cases influenza & hospital visits with influenza-like illnesses & test positivity rate influenza) |
| Time resolution of raw outcome | weekly |
| Computed outcome | none |

– continued on next page –

| | |
|---|---|
| Method to obtain the computed outcome | – not applicable – |
| Data source | data from (sub)national authorities (National Notifiable Diseases Reporting System: influenza, National Sentinel Influenza Surveillance Network: influenza-like illnesses) |
| Data availability | data access via source |
| **Intervention** | |
| Terminology for interventions | interventions |
| Terminology for the specific type of non-pharmaceutical interventions | non-pharmaceutical |
| Exposure types | one combination of interventions (level-1 public health response with many different measures (including travel restrictions, gathering bans, school and venue closure)) |
| Types of single interventions | – not applicable – |
| Coding of interventions | not necessary |
| Source of intervention data | another study (Tian et al.) |
| Availability of data on exposure | raw data documented in the manuscript |
| **Methodological approach** | |
| Empirical approach | descriptive |
| Use of exposure variation | only variation over time for a single population |
| Method | description of change over time (comparison of time periods) |
| Code availability | none |
| **Effect assessment** | |
| Reporting of intervention effect | quantification of change in outcome values |
| Measure of effect | cases: influenza, emergency room visits: influenza-like illness, test positivity rate: influenza |
| Reporting of uncertainty | yes |

– continued on next page –

| | |
|---|---|
| Sensitivity analysis | none |
| Subgroup assessment | none |

| **Cheng et al. 2020b** | |
|---|---|
| **Study information** | |
| Author | Cheng Q, Liu Z, Cheng G, Huang J |
| Title | Heterogeneity and Effectiveness Analysis of COVID-19 Prevention and Control in Major Cities in China through Time-Varying Reproduction Number Estimation |
| Year | 2020 |
| Journal | Scientific Reports |
| DOI | 10.1038/s41598-020-79063-x |
| Date received | 2020-02-14 |
| Date accepted | 2020-12-03 |
| **Study setting** | |
| Number of populations included | multiple |
| Level of populations included | subnational (25 cities in China) |
| Geographic areas covered | Asia |
| Number of countries covered | single country (China) |
| Study period | end date at peak of wave |
| **Outcome** | |
| Raw outcome | epidemiological population-level outcome (cases) |
| Time resolution of raw outcome | daily |
| Computed outcome | epidemiological parameter (reproduction number) |

– continued on next page –

| | |
|---|---|
| Method to obtain the computed outcome | statistical estimation of reproduction number (Cori et al.) |
| Data source | data from (sub)national authorities |
| Data availability | data made available by the authors |
| **Intervention** | |
| Terminology for interventions | measures |
| Terminology for the specific type of non-pharmaceutical interventions | prevention, control |
| Exposure types | all interventions together |
| Types of single interventions | – not applicable – |
| Coding of interventions | – not applicable – |
| Source of intervention data | – not applicable – |
| Availability of data on exposure | – not applicable – |
| **Methodological approach** | |
| Empirical approach | descriptive |
| Use of exposure variation | only variation over time for multiple populations |
| Method | description of change over time (description of time course) |
| Code availability | none |
| **Effect assessment** | |
| Reporting of intervention effect | qualitative statement |
| Measure of effect | reproduction number |
| Reporting of uncertainty | – not applicable – |
| Sensitivity analysis | none |
| Subgroup assessment | none |

| Cheng et al. 2020c (Analysis 1) | |
|---|---|
| **Study information** | |
| Author | Cheng C, Zhang T, Song C, Shen S, Jiang Y, Zhang X |
| Title | The Coupled Impact of Emergency Responses and Population Flows on the COVID-19 Pandemic in China |
| Year | 2020 |
| Journal | GeoHealth |
| DOI | 10.1029/2020gh000332 |
| Date received | 2020-10-08 |
| Date accepted | 2020-12-18 |
| **Study setting** | |
| Number of populations included | multiple |
| Level of populations included | both national and subnational (China and its provinces) |
| Geographic areas covered | Asia |
| Number of countries covered | single country (China) |
| Study period | start and end date span first epidemic wave |
| **Outcome** | |
| Raw outcome | epidemiological population-level outcome (cases) |
| Time resolution of raw outcome | daily |
| Computed outcome | none |
| Method to obtain the computed outcome | – not applicable – |
| Data source | data from publicly available open-source projects (public repository aggregating data from health authorities and other sources) |
| Data availability | data access via source |
| **Intervention** | |

– continued on next page –

| | |
|---|---|
| Terminology for interventions | responses |
| Terminology for the specific type of non-pharmaceutical interventions | emergency |
| Exposure types | stringency index (computation of a index based on level and timing of the public health response in the provinces) |
| Types of single interventions | – not applicable – |
| Coding of interventions | necessary |
| Source of intervention data | coding done by authors |
| Availability of data on exposure | coded data made available by the authors |
| **Methodological approach** | |
| Empirical approach | parametric |
| Use of exposure variation | both variation over time and between populations |
| Method | non-mechanistic model (correlation) |
| Code availability | none |
| **Effect assessment** | |
| Reporting of intervention effect | quantification of change in outcome values |
| Measure of effect | cases |
| Reporting of uncertainty | yes |
| Sensitivity analysis | none |
| Subgroup assessment | none |

**Cheng et al. 2020c (Analysis 2)**

| | |
|---|---|
| **Study information** | |
| Author | Cheng C, Zhang T, Song C, Shen S, Jiang Y, Zhang X |

– continued on next page –

| | |
|---|---|
| Title | The Coupled Impact of Emergency Responses and Population Flows on the COVID-19 Pandemic in China |
| Year | 2020 |
| Journal | GeoHealth |
| DOI | 10.1029/2020gh000332 |
| Date received | 2020-10-08 |
| Date accepted | 2020-12-18 |
| **Study setting** | |
| Number of populations included | multiple |
| Level of populations included | both national and subnational (China and its provinces) |
| Geographic areas covered | Asia |
| Number of countries covered | single country (China) |
| Study period | start and end date span first epidemic wave |
| **Outcome** | |
| Raw outcome | epidemiological population-level outcome (cases) |
| Time resolution of raw outcome | daily |
| Computed outcome | none |
| Method to obtain the computed outcome | – not applicable – |
| Data source | data from publicly available open-source projects (public repository aggregating data from health authorities and other sources) |
| Data availability | data access via source |
| **Intervention** | |
| Terminology for interventions | responses |
| Terminology for the specific type of non-pharmaceutical interventions | emergency |

– continued on next page –

| | |
|---|---|
| Exposure types | stringency index (computation of a index based on level and timing of the public health response in the provinces) |
| Types of single interventions | – not applicable – |
| Coding of interventions | necessary |
| Source of intervention data | coding done by authors |
| Availability of data on exposure | coded data made available by the authors |
| **Methodological approach** | |
| Empirical approach | parametric |
| Use of exposure variation | both variation over time and between populations |
| Method | non-mechanistic model (other: weighted regression) |
| Code availability | none |
| **Effect assessment** | |
| Reporting of intervention effect | quantification of change in outcome values |
| Measure of effect | cases |
| Reporting of uncertainty | yes |
| Sensitivity analysis | none |
| Subgroup assessment | none |

**Cheng et al. 2020c (Analysis 3, mobility as exposure)**

| | |
|---|---|
| **Study information** | |
| Author | Cheng C, Zhang T, Song C, Shen S, Jiang Y, Zhang X |
| Title | The Coupled Impact of Emergency Responses and Population Flows on the COVID-19 Pandemic in China |
| Year | 2020 |
| Journal | GeoHealth |

– continued on next page –

| | |
|---|---|
| DOI | 10.1029/2020gh000332 |
| Date received | 08.10.2020 |
| Date accepted | 18.12.2020 |
| **Study setting** | |
| Number of populations included | multiple |
| Level of populations included | both national and subnational (China and its provinces) |
| Geographic areas covered | Asia |
| Number of countries covered | single country (China) |
| Study period | start and end date span first epidemic wave |
| **Outcome** | |
| Raw outcome | epidemiological population-level outcome (cases) |
| Time resolution of raw outcome | daily |
| Computed outcome | none |
| Method to obtain the computed outcome | – not applicable – |
| Data source | data from publicly available open-source projects (public repository aggregating data from health authorities and other sources) |
| Data availability | data access via source |
| **Intervention** | |
| Terminology for interventions | responses |
| Terminology for the specific type of non-pharmaceutical interventions | emergency |
| Exposure types | mobility (proportion of population moving out of Wuhan that moves to certain province, relative population inflow into province from across country) |
| Types of single interventions | – not applicable – |
| Coding of interventions | – not applicable – |

– continued on next page –

| | |
|---|---|
| Source of intervention data | mobility data by corporate organizations (Baidu) |
| Availability of data on exposure | data access via source |
| **Methodological approach** | |
| Empirical approach | descriptive |
| Use of exposure variation | both variation over time and between populations |
| Method | non-mechanistic model (correlation) |
| Code availability | none |
| **Effect assessment** | |
| Reporting of intervention effect | quantification of change in outcome values |
| Measure of effect | cases |
| Reporting of uncertainty | yes |
| Sensitivity analysis | none |
| Subgroup assessment | none |

**Cheng et al. 2020d**

| | |
|---|---|
| **Study information** | |
| Author | Cheng VC, Wong SC, Chuang VW, So SY, Chen JH, Sridhar S, To KK, Chan JF, Hung IF, Ho PL, Yuen KY |
| Title | The Role of Community-Wide Wearing of Face Mask for Control of Coronavirus Disease 2019 (COVID-19) Epidemic Due to SARS-CoV-2 |
| Year | 2020 |
| Journal | Journal of Infection |
| DOI | 10.1016/j.jinf.2020.04.024 |
| Date received | – could not be evaluated – |

– continued on next page –

| | |
|---|---|
| Date accepted | 2020-04-17 |
| **Study setting** | |
| Number of populations included | multiple |
| Level of populations included | national (Hong Kong Special Administrative Region, Germany, France, Italy, Spain, UK, South Korea, Singapore, US) |
| Geographic areas covered | Asia, Europe, North America |
| Number of countries covered | multiple countries |
| Study period | same end date for several populations with diverse epidemic trajectories |
| **Outcome** | |
| Raw outcome | epidemiological population-level outcome (cases) |
| Time resolution of raw outcome | daily |
| Computed outcome | none |
| Method to obtain the computed outcome | – not applicable – |
| Data source | data from publicly available cross-country selections (WHO) |
| Data availability | data access via source |
| **Intervention** | |
| Terminology for interventions | interventions |
| Terminology for the specific type of non-pharmaceutical interventions | non-pharmaceutical |
| Exposure types | one single intervention (mask mandate) |
| Types of single interventions | mask mandate |
| Coding of interventions | not necessary |
| Source of intervention data | government or news websites |
| Availability of data on exposure | raw data documented in the manuscript |
| **Methodological approach** | |

– continued on next page –

| | |
|---|---|
| Empirical approach | descriptive |
| Use of exposure variation | only variation between populations |
| Method | comparison of populations |
| Code availability | none |
| **Effect assessment** | |
| Reporting of intervention effect | quantification of change in outcome values |
| Measure of effect | cases |
| Reporting of uncertainty | yes |
| Sensitivity analysis | none |
| Subgroup assessment | none |

**Chernozhukov et al. 2021 (Analysis 1)**

| | |
|---|---|
| **Study information** | |
| Author | Chernozhukov V, Kasahara H, Schrimpf P |
| Title | Causal Impact of Masks, Policies, Behavior on Early Covid-19 Pandemic in the U.S |
| Year | 2021 |
| Journal | Journal of Econometrics |
| DOI | 10.1016/j.jeconom.2020.09.003 |
| Date received | 2020-07-03 |
| Date accepted | 2020-09-15 |
| **Study setting** | |
| Number of populations included | multiple |

– continued on next page –

| | |
|---|---|
| Level of populations included | both national and subnational (US and its states) |
| Geographic areas covered | North America |
| Number of countries covered | single country (US) |
| Study period | same end date for several populations with diverse epidemic trajectories |
| **Outcome** | |
| Raw outcome | epidemiological population-level outcome (cases, deaths), behavioral outcome (mobility: time spent in various locations) |
| Time resolution of raw outcome | daily |
| Computed outcome | measure of epidemic trend (growth rate, growth rate: deaths) |
| Method to obtain the computed outcome | simple computation |
| Data source | data from publicly available cross-country selections (New York Times), mobility data from corporate organizations (Google) |
| Data availability | data made available by the authors |
| **Intervention** | |
| Terminology for interventions | policies |
| Terminology for the specific type of non-pharmaceutical interventions | social distancing |
| Exposure types | multiple separate interventions (stay-at-home, closed nonessential businesses, closed K-12 schools, closed restaurants except takeout, closed movie theaters, and face mask mandates for employees in public facing businesses) |
| Types of single interventions | stay-at-home order, workplace closure, school closure, venue closure, mask mandate |
| Coding of interventions | necessary |
| Source of intervention data | use of externally coded data (Boston University COVID-19 US State Policy Database) |
| Availability of data on exposure | coded data made available by the authors |

– continued on next page –

| | |
|---|---|
| **Methodological approach** | |
| Empirical approach | parametric |
| Use of exposure variation | both variation over time and between populations |
| Method | non-mechanistic model (generalized linear model) |
| Code availability | publicly available |
| **Effect assessment** | |
| Reporting of intervention effect | quantification of change in outcome values |
| Measure of effect | growth rate, growth rate:deaths, mobility: time spent in various locations |
| Reporting of uncertainty | yes |
| Sensitivity analysis | different coding of interventions used, same analysis with (sub)population(s) excluded, model specification varied |
| Subgroup assessment | none |

| **Chernozhukov et al. 2021 (Analysis 2, mobility as exposure)** | |
|---|---|
| **Study information** | |
| Author | Chernozhukov V, Kasahara H, Schrimpf P |
| Title | Causal Impact of Masks, Policies, Behavior on Early Covid-19 Pandemic in the U.S |
| Year | 2021 |
| Journal | Journal of Econometrics |
| DOI | 10.1016/j.jeconom.2020.09.003 |
| Date received | 2020-07-03 |
| Date accepted | 2020-09-15 |
| **Study setting** | |
| Number of populations included | multiple |

– continued on next page –

| | |
|---|---|
| Level of populations included | both national and subnational (US and its states) |
| Geographic areas covered | North America |
| Number of countries covered | single country (US) |
| Study period | same end date for several populations with diverse epidemic trajectories |
| **Outcome** | |
| Raw outcome | epidemiological population-level outcome (cases, deaths) |
| Time resolution of raw outcome | daily |
| Computed outcome | measure of epidemic trend (growth rate, growth rate: deaths) |
| Method to obtain the computed outcome | simple computation |
| Data source | data from publicly available cross-country selections (New York Times) |
| Data availability | data made available by the authors |
| **Intervention** | |
| Terminology for interventions | policies |
| Terminology for the specific type of non-pharmaceutical interventions | social distancing |
| Exposure types | mobility (time spent in various locations) |
| Types of single interventions | – not applicable – |
| Coding of interventions | – not applicable – |
| Source of intervention data | mobility data by corporate organizations (Google) |
| Availability of data on exposure | data made available by the authors |
| **Methodological approach** | |
| Empirical approach | parametric |
| Use of exposure variation | both variation over time and between populations |
| Method | non-mechanistic model (generalized linear model) |

– continued on next page –

**Chernozhukov et al. 2021 (Analysis 2, mobility as exposure)** – continued from previous page

| | |
|---|---|
| Code availability | publicly available |
| **Effect assessment** | |
| Reporting of intervention effect | quantification of change in outcome values |
| Measure of effect | growth rate, growth rate: deaths |
| Reporting of uncertainty | yes |
| Sensitivity analysis | model specification varied |
| Subgroup assessment | none |

**Choi & Ki 2020**

| | |
|---|---|
| **Study information** | |
| Author | Choi S, Ki M |
| Title | Analyzing the Effects of Social Distancing on the COVID-19 Pandemic in Korea Using Mathematical Modeling |
| Year | 2020 |
| Journal | Epidemiology and Health |
| DOI | 10.4178/epih.e2020064 |
| Date received | 2020-06-16 |
| Date accepted | 2020-08-19 |
| **Study setting** | |
| Number of populations included | single |
| Level of populations included | national (South Korea) |
| Geographic areas covered | Asia |
| Number of countries covered | single country (South Korea) |
| Study period | start and end date span first epidemic wave |

– continued on next page –

| | |
|---|---|
| **Outcome** | |
| Raw outcome | epidemiological population-level outcome (cases) |
| Time resolution of raw outcome | daily |
| Computed outcome | epidemiological parameter (reproduction number) |
| Method to obtain the computed outcome | compartmental transmission model (single-population) |
| Data source | data from (sub)national authorities |
| Data availability | data access via source |
| **Intervention** | |
| Terminology for interventions | measures |
| Terminology for the specific type of non-pharmaceutical interventions | social distancing |
| Exposure types | phases of interventions (social distancing policies of varying strictness divided into 5 phases but where the phases are manually derived based on observed changes in the outcome (the phases are guided by the NPI dates, but changepoints were defined manually by inspecting the 2nd order difference (second derivative) of the cumulative case time series)) |
| Types of single interventions | – not applicable – |
| Coding of interventions | not necessary |
| Source of intervention data | – could not be evaluated – |
| Availability of data on exposure | raw data documented in the manuscript |
| **Methodological approach** | |
| Empirical approach | descriptive |
| Use of exposure variation | only variation over time for a single population |
| Method | description of change over time (comparison of time periods) |
| Code availability | none |
| **Effect assessment** | |

– continued on next page –

| | |
|---|---|
| Reporting of intervention effect | comparison of outcome values |
| Measure of effect | reproduction number |
| Reporting of uncertainty | no |
| Sensitivity analysis | none |
| Subgroup assessment | none |

| **Choi et al. 2020** | |
|---|---|
| **Study information** | |
| Author | Choi Y, Kim JS, Choi H, Lee H, Lee CH |
| Title | Assessment of Social Distancing for Controlling COVID-19 in Korea: An Age-Structured Modeling Approach |
| Year | 2020 |
| Journal | International Journal of Environmental Research and Public Health |
| DOI | 10.3390/ijerph17207474 |
| Date received | 2020-08-28 |
| Date accepted | 2020-10-12 |
| **Study setting** | |
| Number of populations included | single |
| Level of populations included | subnational (Seougl and Gyenggi provinces in South Korea) |
| Geographic areas covered | Asia |
| Number of countries covered | single country (South Korea) |
| Study period | includes second wave (two waves) |
| **Outcome** | |
| Raw outcome | epidemiological population-level outcome (cases, deaths, recovered cases) |

– continued on next page –

| | |
|---|---|
| Time resolution of raw outcome | daily |
| Computed outcome | epidemiological parameter (reproduction number, transmission probability) |
| Method to obtain the computed outcome | compartmental transmission model (single-population) |
| Data source | data from (sub)national authorities |
| Data availability | data access via source |
| **Intervention** | |
| Terminology for interventions | measures |
| Terminology for the specific type of non-pharmaceutical interventions | social distancing |
| Exposure types | all interventions together (timeline of measures divided into three phases) |
| Types of single interventions | – not applicable – |
| Coding of interventions | – not applicable – |
| Source of intervention data | – not applicable – |
| Availability of data on exposure | – not applicable – |
| **Methodological approach** | |
| Empirical approach | descriptive |
| Use of exposure variation | only variation over time for a single population |
| Method | description of change over time (comparison of time periods) |
| Code availability | none |
| **Effect assessment** | |
| Reporting of intervention effect | comparison of outcome values |
| Measure of effect | reproduction number, probability of transmission upon contact |
| Reporting of uncertainty | no |
| Sensitivity analysis | none |

– continued on next page –

| | |
|---|---|
| Subgroup assessment | socioeconomic indicators (age) |

| **Cobb & Seale 2020** | |
|---|---|
| **Study information** | |
| Author | Cobb JS, Seale MA |
| Title | Examining the Effect of Social Distancing on the Compound Growth Rate of COVID-19 at the County Level (United States) Using Statistical Analyses and a Random Forest Machine Learning Model |
| Year | 2020 |
| Journal | Public Health |
| DOI | 10.1016/j.puhe.2020.04.016 |
| Date received | 2020-04-10 |
| Date accepted | 2020-04-13 |
| **Study setting** | |
| Number of populations included | multiple |
| Level of populations included | both national and subnational (US counties) |
| Geographic areas covered | North America |
| Number of countries covered | single country (US) |
| Study period | end date in growth phase of wave |
| **Outcome** | |
| Raw outcome | epidemiological population-level outcome (cases) |
| Time resolution of raw outcome | daily |
| Computed outcome | measure of epidemic trend (growth rate) |
| Method to obtain the computed outcome | simple computation |

– continued on next page –

| | |
|---|---|
| Data source | data from (sub)national authorities (till March 25), data from public media outlets (New York Times: beyond March 25) |
| Data availability | data access via source |
| **Intervention** | |
| Terminology for interventions | measures |
| Terminology for the specific type of non-pharmaceutical interventions | social distancing |
| Exposure types | multiple separate interventions (presidential recommendation to limit gatherings, shelter-in-place order) |
| Types of single interventions | other, stay-at-home order |
| Coding of interventions | necessary |
| Source of intervention data | – could not be evaluated – |
| Availability of data on exposure | coded data made available by the authors |
| **Methodological approach** | |
| Empirical approach | descriptive |
| Use of exposure variation | only variation over time for a single population |
| Method | comparison of populations |
| Code availability | none |
| **Effect assessment** | |
| Reporting of intervention effect | quantification of change in outcome values |
| Measure of effect | growth rate |
| Reporting of uncertainty | yes |
| Sensitivity analysis | none |
| Subgroup assessment | none |

| Coletti et al. 2020 | |
|---|---|
| **Study information** | |
| Author | Coletti P, Wambua J, Gimma A, Willem L, Vercruysse S, Vanhoutte B, Jarvis CI, Van Zandvoort K, Edmunds J, Beutels P, Hens N |
| Title | CoMix: Comparing Mixing Patterns in the Belgian Population during and after Lockdown |
| Year | 2020 |
| Journal | Scientific Reports |
| DOI | 10.1038/s41598-020-78540-7 |
| Date received | 2020-08-25 |
| Date accepted | 2020-12-14 |
| **Study setting** | |
| Number of populations included | single |
| Level of populations included | national (Belgium) |
| Geographic areas covered | Europe |
| Number of countries covered | single country (Belgium) |
| Study period | start and end date span first epidemic wave: behavioral |
| **Outcome** | |
| Raw outcome | behavioral outcome (survey: number of contacts & face mask use) |
| Time resolution of raw outcome | – not applicable – |
| Computed outcome | epidemiological parameter (reproduction number) |
| Method to obtain the computed outcome | statistical estimation of reproduction number from individual data (Diekmann et al.) |
| Data source | data collected by authors |
| Data availability | data made available by the authors |
| **Intervention** | |
| Terminology for interventions | measures |

– continued on next page –

| | |
|---|---|
| Terminology for the specific type of non-pharmaceutical interventions | social distancing |
| Exposure types | all interventions together (timeline of measures) |
| Types of single interventions | – not applicable – |
| Coding of interventions | – not applicable – |
| Source of intervention data | – not applicable – |
| Availability of data on exposure | – not applicable – |
| **Methodological approach** | |
| Empirical approach | descriptive |
| Use of exposure variation | only variation over time for a single population |
| Method | description of change over time (comparison of time periods) |
| Code availability | none |
| **Effect assessment** | |
| Reporting of intervention effect | comparison of outcome values |
| Measure of effect | number of contacts, face mask usage, reproduction number |
| Reporting of uncertainty | no |
| Sensitivity analysis | none |
| Subgroup assessment | socioeconomic indicators (age, gender, household size) |

**Collins & Duffy 2020**

| | |
|---|---|
| **Study information** | |
| Author | Collins OC, Duffy KJ |
| Title | Estimating the Impact of Lock-down, Quarantine and Sensitization in a COVID-19 Outbreak: Lessons from the COVID-19 Outbreak in China |

– continued on next page –

| | |
|---|---|
| Year | 2020 |
| Journal | PeerJ |
| DOI | 10.7717/peerj.9933 |
| Date received | 2020-05-27 |
| Date accepted | 2020-08-23 |
| **Study setting** | |
| Number of populations included | single |
| Level of populations included | national (China) |
| Geographic areas covered | Asia |
| Number of countries covered | single country (China) |
| Study period | start and end date span first epidemic wave |
| **Outcome** | |
| Raw outcome | epidemiological population-level outcome (active cases) |
| Time resolution of raw outcome | daily |
| Computed outcome | none |
| Method to obtain the computed outcome | – not applicable – |
| Data source | data from publicly available cross-country selections (Worldometer) |
| Data availability | data made available by the authors |
| **Intervention** | |
| Terminology for interventions | measures |
| Terminology for the specific type of non-pharmaceutical interventions | control |
| Exposure types | categories of interventions (sensitization, quarantine, lockdown and the combination of all three) |
| Types of single interventions | – not applicable – |

– continued on next page –

| | |
|---|---|
| Coding of interventions | – not applicable – |
| Source of intervention data | – not applicable – |
| Availability of data on exposure | – not applicable – |
| **Methodological approach** | |
| Empirical approach | parametric |
| Use of exposure variation | only variation over time for a single population |
| Method | mechanistic model (compartmental single-population transmission model) |
| Latent variable functionally linked to intervention | susceptible decay, transmission rate, quarantine rate |
| Code availability | publicly available |
| **Effect assessment** | |
| Reporting of intervention effect | quantification of change in outcome values |
| Measure of effect | susceptible decay rate, transmission rate, quarantine rate |
| Reporting of uncertainty | no |
| Sensitivity analysis | none |
| Subgroup assessment | none |

**Cooper et al. 2020**

| | |
|---|---|
| **Study information** | |
| Author | Cooper I, Mondal A, Antonopoulos CG |
| Title | A SIR Model Assumption for the Spread of COVID-19 in Different Communities |
| Year | 2020 |
| Journal | Chaos, Solitons & Fractals |

– continued on next page –

| | |
|---|---|
| DOI | 10.1016/j.chaos.2020.110057 |
| Date received | 2020-06-18 |
| Date accepted | 2020-06-23 |
| **Study setting** | |
| Number of populations included | multiple |
| Level of populations included | both national and subnational (China, South Korea, India, Australia, USA, Italy, Texas) |
| Geographic areas covered | Asia, Oceania, Europe, North America |
| Number of countries covered | multiple countries |
| Study period | same end date for several populations with diverse epidemic trajectories |
| **Outcome** | |
| Raw outcome | epidemiological population-level outcome (cases, deaths, recovered cases) |
| Time resolution of raw outcome | daily |
| Computed outcome | other (infections) |
| Method to obtain the computed outcome | compartmental transmission model (single-population) |
| Data source | data from publicly available cross-country selections (Worldometer) |
| Data availability | data access via source |
| **Intervention** | |
| Terminology for interventions | measures |
| Terminology for the specific type of non-pharmaceutical interventions | control |
| Exposure types | all interventions together |
| Types of single interventions | – not applicable – |
| Coding of interventions | – not applicable – |
| Source of intervention data | – not applicable – |

– continued on next page –

| | |
|---|---|
| Availability of data on exposure | – not applicable – |
| **Methodological approach** | |
| Empirical approach | descriptive |
| Use of exposure variation | only variation over time for multiple populations |
| Method | description of change over time (description of time course) |
| Code availability | none |
| **Effect assessment** | |
| Reporting of intervention effect | qualitative statement |
| Measure of effect | infections |
| Reporting of uncertainty | – not applicable – |
| Sensitivity analysis | none |
| Subgroup assessment | none |

**Courtemanche et al. 2020**

| | |
|---|---|
| **Study information** | |
| Author | Courtemanche C, Garuccio J, Le A, Pinkston J, Yelowitz A |
| Title | Strong Social Distancing Measures in the United States Reduced the COVID-19 Growth Rate |
| Year | 2020 |
| Journal | Health Affairs |
| DOI | 10.1377/hlthaff.2020.00608 |
| Date received | – could not be evaluated – |
| Date accepted | 2020-05-14 (published) |
| **Study setting** | |

– continued on next page –

| | |
|---|---|
| Number of populations included | multiple |
| Level of populations included | both national and subnational (US counties) |
| Geographic areas covered | North America |
| Number of countries covered | single country (US) |
| Study period | same end date for several populations with diverse epidemic trajectories |
| **Outcome** | |
| Raw outcome | epidemiological population-level outcome (cases) |
| Time resolution of raw outcome | daily |
| Computed outcome | measure of epidemic trend (growth rate) |
| Method to obtain the computed outcome | simple computation |
| Data source | data from publicly available cross-country selections (JHU) |
| Data availability | data access via source |
| **Intervention** | |
| Terminology for interventions | measures |
| Terminology for the specific type of non-pharmaceutical interventions | social distancing |
| Exposure types | multiple separate interventions (bans of large gatherings, school closure, venue closure, stay-at-home order) |
| Types of single interventions | bans of large gatherings, school closure, venue closure, stay-at-home order |
| Coding of interventions | necessary |
| Source of intervention data | use of externally coded data (same as in Killeen et al.) |
| Availability of data on exposure | coded data made available by the authors |
| **Methodological approach** | |
| Empirical approach | parametric |
| Use of exposure variation | both variation over time and between populations |

– continued on next page –

| | |
|---|---|
| Method | non-mechanistic model (generalized linear model) |
| Code availability | none |
| **Effect assessment** | |
| Reporting of intervention effect | quantification of change in outcome values |
| Measure of effect | growth rate |
| Reporting of uncertainty | yes |
| Sensitivity analysis | different coding of interventions used (gathering bans of 50 instead of 500, closure of restaurants AND gyms, not OR), same analysis with (sub)population(s) excluded, model specification varied |
| Subgroup assessment | none |

| | |
|---|---|
| | **Cowling et al. 2020 (Analysis 1)** |
| **Study information** | |
| Author | Cowling BJ, Ali ST, Ng TW, Tsang TK, Li JC, Fong MW, Liao Q, Kwan MY, Lee SL, Chiu SS, Wu JT, Wu P, Leung GM |
| Title | Impact Assessment of Non-Pharmaceutical Interventions against Coronavirus Disease 2019 and Influenza in Hong Kong: An Observational Study |
| Year | 2020 |
| Journal | The Lancet Public Health |
| DOI | 10.1016/s2468-2667(20)30090-6 |
| Date received | – could not be evaluated – |
| Date accepted | 2020-04-17 (published) |
| **Study setting** | |
| Number of populations included | single |

– continued on next page –

| | |
|---|---|
| Level of populations included | national (Hong Kong) |
| Geographic areas covered | Asia |
| Number of countries covered | single country (Hong Kong) |
| Study period | start and end date span first epidemic wave |
| **Outcome** | |
| Raw outcome | epidemiological individual-level outcome (cases, surrogate disease: cases with influenza & hospitalizations children with influenza) |
| Time resolution of raw outcome | daily |
| Computed outcome | epidemiological parameter (reproduction number, reproduction number: influenza, reproduction number: influenza of hospitalized children) |
| Method to obtain the computed outcome | statistical estimation of reproduction number (Cori et al.) |
| Data source | data from (sub)national authorities |
| Data availability | data access via source |
| **Intervention** | |
| Terminology for interventions | interventions |
| Terminology for the specific type of non-pharmaceutical interventions | non-pharmaceutical |
| Exposure types | one single intervention (school closure) |
| Types of single interventions | school closure |
| Coding of interventions | not necessary |
| Source of intervention data | – could not be evaluated – |
| Availability of data on exposure | raw data documented in the manuscript |
| **Methodological approach** | |
| Empirical approach | descriptive |
| Use of exposure variation | only variation over time for a single population |

– continued on next page –

| | |
|---|---|
| Method | description of change over time (comparison of time periods) |
| Code availability | none |
| **Effect assessment** | |
| Reporting of intervention effect | quantification of change in outcome values |
| Measure of effect | reproduction number: SARS-CoV-2, surrogate disease: influenza, surrogate disease: influenza in hospitalized children |
| Reporting of uncertainty | yes |
| Sensitivity analysis | none |
| Subgroup assessment | none |

| | **Cowling et al. 2020 (Analysis 2)** |
|---|---|
| **Study information** | |
| Author | Cowling BJ, Ali ST, Ng TW, Tsang TK, Li JC, Fong MW, Liao Q, Kwan MY, Lee SL, Chiu SS, Wu JT, Wu P, Leung GM |
| Title | Impact Assessment of Non-Pharmaceutical Interventions against Coronavirus Disease 2019 and Influenza in Hong Kong: An Observational Study |
| Year | 2020 |
| Journal | The Lancet Public Health |
| DOI | 10.1016/s2468-2667(20)30090-6 |
| Date received | – could not be evaluated – |
| Date accepted | 2020-04-17 (published) |
| **Study setting** | |
| Number of populations included | single |
| Level of populations included | national (Hong Kong) |
| Geographic areas covered | Asia |

– continued on next page –

| | |
|---|---|
| Number of countries covered | single country (Hong Kong) |
| Study period | start and end date span first epidemic wave: behavioral |
| **Outcome** | |
| Raw outcome | behavioral outcome (survey: responses to behavioral adherence to measures, e.g., use of face masks & avoidance of public places) |
| Time resolution of raw outcome | – not applicable – |
| Computed outcome | none |
| Method to obtain the computed outcome | – not applicable – |
| Data source | data collected by authors |
| Data availability | data not accessible |
| **Intervention** | |
| Terminology for interventions | interventions |
| Terminology for the specific type of non-pharmaceutical interventions | non-pharmaceutical |
| Exposure types | all interventions together (timeline of measures) |
| Types of single interventions | – not applicable – |
| Coding of interventions | – not applicable – |
| Source of intervention data | – not applicable – |
| Availability of data on exposure | – not applicable – |
| **Methodological approach** | |
| Empirical approach | descriptive |
| Use of exposure variation | only variation over time for a single population |
| Method | description of change over time (description of time course) |
| Code availability | none |
| **Effect assessment** | |

– continued on next page –

| | |
|---|---|
| Reporting of intervention effect | comparison of outcome values |
| Measure of effect | face mask usage, avoidance of public places |
| Reporting of uncertainty | yes |
| Sensitivity analysis | none |
| Subgroup assessment | none |

| **Crokidakis 2020** | |
|---|---|
| **Study information** | |
| Author | Crokidakis N |
| Title | COVID-19 Spreading in Rio de Janeiro, Brazil: Do the Policies of Social Isolation Really Work? |
| Year | 2020 |
| Journal | Chaos, Solitons & Fractals |
| DOI | 10.1016/j.chaos.2020.109930 |
| Date received | 2020-05-05 |
| Date accepted | 2020-05-20 |
| **Study setting** | |
| Number of populations included | single |
| Level of populations included | subnational (Rio de Janeiro) |
| Geographic areas covered | Central and South America |
| Number of countries covered | single country (Brazil) |
| Study period | end date in growth phase of wave |
| **Outcome** | |
| Raw outcome | epidemiological population-level outcome (cases) |

– continued on next page –

| | |
|---|---|
| Time resolution of raw outcome | daily |
| Computed outcome | none |
| Method to obtain the computed outcome | – not applicable – |
| Data source | data from (sub)national authorities |
| Data availability | data access via source |
| **Intervention** | |
| Terminology for interventions | policies |
| Terminology for the specific type of non-pharmaceutical interventions | social isolation |
| Exposure types | all interventions together |
| Types of single interventions | – not applicable – |
| Coding of interventions | – not applicable – |
| Source of intervention data | – not applicable – |
| Availability of data on exposure | – not applicable – |
| **Methodological approach** | |
| Empirical approach | parametric |
| Use of exposure variation | only variation over time for a single population |
| Method | mechanistic model (compartmental single-population transmission model) |
| Latent variable functionally linked to intervention | susceptible decay |
| Code availability | none |
| **Effect assessment** | |
| Reporting of intervention effect | quantification of change in outcome values |
| Measure of effect | cases, quarantined |
| Reporting of uncertainty | no |

– continued on next page –

| | |
|---|---|
| Sensitivity analysis | none |
| Subgroup assessment | none |

| **Czerwińska & Szenborn 2020** | |
|---|---|
| **Study information** | |
| Author | Czerwińska E, Szenborn L |
| Title | The COVID-19 Pandemic Lockdown Causes a Decrease in the Number of Cases of Other Infectious Diseases in Poland |
| Year | 2020 |
| Journal | Pediatria Polska |
| DOI | 10.5114/polp.2020.97221 |
| Date received | 2020-06-04 |
| Date accepted | 2020-06-22 |
| **Study setting** | |
| Number of populations included | single |
| Level of populations included | national (Poland) |
| Geographic areas covered | Europe |
| Number of countries covered | single country (Poland) |
| Study period | end date at peak of wave |
| **Outcome** | |
| Raw outcome | epidemiological population-level outcome (surrogate disease: cases of multiple other infectious diseases) |
| Time resolution of raw outcome | biweekly |
| Computed outcome | none |

– continued on next page –

| | |
|---|---|
| Method to obtain the computed outcome | – not applicable – |
| Data source | data from (sub)national authorities |
| Data availability | data access via source |
| **Intervention** | |
| Terminology for interventions | measures |
| Terminology for the specific type of non-pharmaceutical interventions | social distancing |
| Exposure types | all interventions together (stay-at-home order, school closure, bans of small gatherings, mask mandate) |
| Types of single interventions | – not applicable – |
| Coding of interventions | – not applicable – |
| Source of intervention data | – not applicable – |
| Availability of data on exposure | – not applicable – |
| **Methodological approach** | |
| Empirical approach | descriptive |
| Use of exposure variation | only variation over time for a single population |
| Method | description of change over time (comparison of time periods) |
| Code availability | none |
| **Effect assessment** | |
| Reporting of intervention effect | quantification of change in outcome values |
| Measure of effect | hospitalizations: acute bronchiolitis in infants |
| Reporting of uncertainty | no |
| Sensitivity analysis | none |
| Subgroup assessment | none |

| da Silva & Tsigaris 2020 | |
|---|---|
| **Study information** | |
| Author | da Silva JA, Tsigaris P |
| Title | Policy Determinants of COVID-19 Pandemic–Induced Fatality Rates across Nations |
| Year | 2020 |
| Journal | Public Health |
| DOI | 10.1016/j.puhe.2020.08.008 |
| Date received | 2020-05-09 |
| Date accepted | 2020-08-12 |
| **Study setting** | |
| Number of populations included | multiple |
| Level of populations included | national (121 countries areound the world) |
| Geographic areas covered | Europe, Middle East and Africa, Asia, Oceania, Central and South America, North America |
| Number of countries covered | multiple countries |
| Study period | same end date for several populations with diverse epidemic trajectories |
| **Outcome** | |
| Raw outcome | epidemiological population-level outcome (cases, deaths) |
| Time resolution of raw outcome | daily |
| Computed outcome | epidemiological parameter (case-fatality rate) |
| Method to obtain the computed outcome | simple computation |
| Data source | data from publicly available cross-country selections (Worldometer) |
| Data availability | data access via source |

– continued on next page –

| | |
|---|---|
| **Intervention** | |
| Terminology for interventions | measures |
| Terminology for the specific type of non-pharmaceutical interventions | containment |
| Exposure types | start of interventions (days since first policy, delay for international travel restrictions, delay for public information campain, delay for testing policy) |
| Types of single interventions | – not applicable – |
| Coding of interventions | necessary |
| Source of intervention data | use of externally coded data (OxCGRT) |
| Availability of data on exposure | access to externally coded data via source |
| **Methodological approach** | |
| Empirical approach | parametric |
| Use of exposure variation | only variation between populations |
| Method | non-mechanistic model (generalized linear model) |
| Code availability | none |
| **Effect assessment** | |
| Reporting of intervention effect | quantification of change in outcome values |
| Measure of effect | case-fatality rate |
| Reporting of uncertainty | yes |
| Sensitivity analysis | none |
| Subgroup assessment | socioeconomic indicators (income) |

**Dai et al. 2020**

| |
|---|
| **Study information** |

– continued on next page –

| | |
|---|---|
| Author | Dai C, Yang AJ, Wang K, and |
| Title | Evaluation of Prevention and Control Interventions and Its Impact on the Epidemic of Coronavirus Disease 2019 in Chongqing and Guizhou Provinces |
| Year | 2020 |
| Journal | Mathematical Biosciences and Engineering |
| DOI | 10.3934/mbe.2020152 |
| Date received | 2020-02-12 |
| Date accepted | 2020-03-11 |
| **Study setting** | |
| Number of populations included | multiple |
| Level of populations included | subnational (Chongqing and Guizhou province in China) |
| Geographic areas covered | Asia |
| Number of countries covered | single country (China) |
| Study period | start and end date span first epidemic wave |
| **Outcome** | |
| Raw outcome | epidemiological population-level outcome (cases) |
| Time resolution of raw outcome | daily |
| Computed outcome | none |
| Method to obtain the computed outcome | – not applicable – |
| Data source | data from (sub)national authorities |
| Data availability | data access via source |
| **Intervention** | |
| Terminology for interventions | measures |
| Terminology for the specific type of non-pharmaceutical interventions | prevention, control |

– continued on next page –

| | |
|---|---|
| Exposure types | one combination of interventions (Wuhan City Lockdown) |
| Types of single interventions | – not applicable – |
| Coding of interventions | not necessary |
| Source of intervention data | – could not be evaluated – |
| Availability of data on exposure | raw data documented in the manuscript |
| **Methodological approach** | |
| Empirical approach | parametric |
| Use of exposure variation | only variation over time for multiple populations |
| Method | mechanistic model (compartmental single-population transmission model) |
| Latent variable functionally linked to intervention | contact rate |
| Code availability | none |
| **Effect assessment** | |
| Reporting of intervention effect | quantification of change in outcome values |
| Measure of effect | contact rate |
| Reporting of uncertainty | no |
| Sensitivity analysis | none |
| Subgroup assessment | none |

**Dandekar et al. 2020**

| | |
|---|---|
| **Study information** | |
| Author | Dandekar R, Rackauckas C, Barbastathis G |
| Title | A Machine Learning-Aided Global Diagnostic and Comparative Tool to Assess Effect of Quarantine Control in COVID-19 Spread |

– continued on next page –

| | |
|---|---|
| Year | 2020 |
| Journal | Patterns |
| DOI | 10.1016/j.patter.2020.100145 |
| Date received | 2020-07-22 |
| Date accepted | 2020-10-21 |
| **Study setting** | |
| Number of populations included | multiple |
| Level of populations included | both national and subnational (70 countries worldwide, 20 major US states) |
| Geographic areas covered | Europe, Middle East and Africa, Asia, Oceania, Central and South America, North America |
| Number of countries covered | multiple countries |
| Study period | same end date for several populations with diverse epidemic trajectories |
| **Outcome** | |
| Raw outcome | epidemiological population-level outcome (cases, recovered cases) |
| Time resolution of raw outcome | daily |
| Computed outcome | none |
| Method to obtain the computed outcome | – not applicable – |
| Data source | data from publicly available cross-country selections (JHU) |
| Data availability | data made available by the authors |
| **Intervention** | |
| Terminology for interventions | policies |
| Terminology for the specific type of non-pharmaceutical interventions | quarantine |
| Exposure types | all interventions together |
| Types of single interventions | – not applicable – |

– continued on next page –

| | |
|---|---|
| Coding of interventions | – not applicable – |
| Source of intervention data | – not applicable – |
| Availability of data on exposure | – not applicable – |
| **Methodological approach** | |
| Empirical approach | parametric |
| Use of exposure variation | only variation over time for multiple populations |
| Method | mechanistic model (compartmental single-population transmission model) |
| Latent variable functionally linked to intervention | quarantine rate |
| Code availability | publicly available |
| **Effect assessment** | |
| Reporting of intervention effect | quantification of change in outcome values |
| Measure of effect | quarantine rate |
| Reporting of uncertainty | no |
| Sensitivity analysis | none |
| Subgroup assessment | none |

| **de Brito Cruz 2020 (Analysis 1)** | |
|---|---|
| **Study information** | |
| Author | de Brito Cruz CH |
| Title | Social Distancing in São Paulo State: Demonstrating the Reduction in Cases Using Time Series Analysis of Deaths Due to COVID-19 |
| Year | 2020 |

– continued on next page –

| | |
|---|---|
| Journal | Revista Brasileira de Epidemiologia |
| DOI | 10.1590/1980-549720200056 |
| Date received | 2020-05-12 |
| Date accepted | 2020-05-14 |
| **Study setting** | |
| Number of populations included | multiple |
| Level of populations included | both national and subnational (Brazil, Sao Paulo State, Sao Paulo City) |
| Geographic areas covered | Central and South America |
| Number of countries covered | single country (Brazil) |
| Study period | end date in growth phase of wave |
| **Outcome** | |
| Raw outcome | epidemiological population-level outcome (deaths) |
| Time resolution of raw outcome | daily |
| Computed outcome | none |
| Method to obtain the computed outcome | – not applicable – |
| Data source | data from (sub)national authorities (National Association of the Registrars for Natural Persons) |
| Data availability | data access via source |
| **Intervention** | |
| Terminology for interventions | measures |
| Terminology for the specific type of non-pharmaceutical interventions | social distancing |
| Exposure types | all interventions together |
| Types of single interventions | – not applicable – |
| Coding of interventions | – not applicable – |

– continued on next page –

| | |
|---|---|
| Source of intervention data | – not applicable – |
| Availability of data on exposure | – not applicable – |
| **Methodological approach** | |
| Empirical approach | descriptive |
| Use of exposure variation | only variation over time for multiple populations |
| Method | description of change over time (description of time course) |
| Code availability | none |
| **Effect assessment** | |
| Reporting of intervention effect | qualitative statement |
| Measure of effect | deaths |
| Reporting of uncertainty | – not applicable – |
| Sensitivity analysis | none |
| Subgroup assessment | none |

**de Brito Cruz 2020 (Analysis 2, mobility as exposure)**

| | |
|---|---|
| **Study information** | |
| Author | de Brito Cruz CH |
| Title | Social Distancing in São Paulo State: Demonstrating the Reduction in Cases Using Time Series Analysis of Deaths Due to COVID-19 |
| Year | 2020 |
| Journal | Revista Brasileira de Epidemiologia |
| DOI | 10.1590/1980-549720200056 |
| Date received | 12.05.2020 |
| Date accepted | 14.05.2020 |

– continued on next page –

| | |
|---|---|
| **Study setting** | |
| Number of populations included | multiple |
| Level of populations included | both national and subnational (Brazil, Sao Paulo State, Sao Paulo City) |
| Geographic areas covered | Central and South America |
| Number of countries covered | single country (Brazil) |
| Study period | end date in growth phase of wave |
| **Outcome** | |
| Raw outcome | epidemiological population-level outcome (deaths) |
| Time resolution of raw outcome | daily |
| Computed outcome | measure of epidemic trend (growth rate: deaths) |
| Method to obtain the computed outcome | simple computation |
| Data source | data from (sub)national authorities (National Association of the Registrars for Natural Persons) |
| Data availability | data access via source |
| **Intervention** | |
| Terminology for interventions | measures |
| Terminology for the specific type of non-pharmaceutical interventions | social distancing |
| Exposure types | mobility (index based on São Paulo Intelligent Monitoring System) |
| Types of single interventions | – not applicable – |
| Coding of interventions | – not applicable – |
| Source of intervention data | data collected by authors |
| Availability of data on exposure | data access via source |
| **Methodological approach** | |
| Empirical approach | descriptive |

– continued on next page –

| | |
|---|---|
| Use of exposure variation | only variation over time for multiple populations |
| Method | non-mechanistic model (correlation) |
| Code availability | none |
| **Effect assessment** | |
| Reporting of intervention effect | comparison of outcome values |
| Measure of effect | growth rate: deaths |
| Reporting of uncertainty | no |
| Sensitivity analysis | none |
| Subgroup assessment | none |

**Dehning et al. 2020**

| | |
|---|---|
| **Study information** | |
| Author | Dehning J, Zierenberg J, Spitzner FP, Wibral M, Neto JP, Wilczek M, Priesemann V |
| Title | Inferring Change Points in the Spread of COVID-19 Reveals the Effectiveness of Interventions |
| Year | 2020 |
| Journal | Science |
| DOI | 10.1126/science.abb9789 |
| Date received | 2020-04-03 |
| Date accepted | 2020-05-12 |
| **Study setting** | |
| Number of populations included | single |
| Level of populations included | national (Germany) |

– continued on next page –

| | |
|---|---|
| Geographic areas covered | Europe |
| Number of countries covered | single country (Germany) |
| Study period | start and end date span first epidemic wave |
| **Outcome** | |
| Raw outcome | epidemiological population-level outcome (cases) |
| Time resolution of raw outcome | daily |
| Computed outcome | none |
| Method to obtain the computed outcome | – not applicable – |
| Data source | data from publicly available cross-country selections (JHU) |
| Data availability | data made available by the authors |
| **Intervention** | |
| Terminology for interventions | interventions |
| Terminology for the specific type of non-pharmaceutical interventions | – not applicable – |
| Exposure types | one combination of interventions (stage 1: event ban, stage 2: school and venue closure, stage 3: contact ban, lockdown) |
| Types of single interventions | – not applicable – |
| Coding of interventions | not necessary |
| Source of intervention data | – could not be evaluated – |
| Availability of data on exposure | raw data documented in the manuscript |
| **Methodological approach** | |
| Empirical approach | parametric |
| Use of exposure variation | only variation over time for a single population |
| Method | mechanistic model (compartmental single-population transmission model) |

– continued on next page –

| | |
|---|---|
| Latent variable functionally linked to intervention | transmission rate |
| Code availability | publicly available |
| **Effect assessment** | |
| Reporting of intervention effect | quantification of change in outcome values |
| Measure of effect | transmission rate |
| Reporting of uncertainty | yes |
| Sensitivity analysis | epidemiological parameters varied, model specification varied |
| Subgroup assessment | none |

| **Dickson et al. 2020** | |
|---|---|
| **Study information** | |
| Author | Dickson MM, Espa G, Giuliani D, Santi F, Savadori L |
| Title | Assessing the Effect of Containment Measures on the Spatio-Temporal Dynamic of COVID-19 in Italy |
| Year | 2020 |
| Journal | Nonlinear Dynamics |
| DOI | 10.1007/s11071-020-05853-7 |
| Date received | 2020-04-27 |
| Date accepted | 2020-07-28 |
| **Study setting** | |
| Number of populations included | multiple |
| Level of populations included | both national and subnational (Italy and its 107 provinces) |
| Geographic areas covered | Europe |

– continued on next page –

| | |
|---|---|
| Number of countries covered | single country (Italy) |
| Study period | start and end date span first epidemic wave |
| **Outcome** | |
| Raw outcome | epidemiological population-level outcome (cases) |
| Time resolution of raw outcome | daily |
| Computed outcome | measure of epidemic trend (other: autoregressive parameters) |
| Method to obtain the computed outcome | other (endemic-epidemic regression model based on hhh4 framework with time-varying temporal and spatial autoregressive parameters representing disease spread within and across provinces) |
| Data source | data from (sub)national authorities |
| Data availability | data access via source |
| **Intervention** | |
| Terminology for interventions | measures |
| Terminology for the specific type of non-pharmaceutical interventions | containment |
| Exposure types | multiple combinations of interventions (national quarantine following DPCM of 11 March 2020, closure of non-essential businesses and lockdown on March 25 ) |
| Types of single interventions | – not applicable – |
| Coding of interventions | not necessary |
| Source of intervention data | – could not be evaluated – |
| Availability of data on exposure | raw data documented in the manuscript |
| **Methodological approach** | |
| Empirical approach | descriptive |
| Use of exposure variation | only variation over time for multiple populations |
| Method | description of change over time (description of time course) |

– continued on next page –

| | |
|---|---|
| Code availability | none |
| **Effect assessment** | |
| Reporting of intervention effect | qualitative statement |
| Measure of effect | autoregressive parameters |
| Reporting of uncertainty | – not applicable – |
| Sensitivity analysis | none |
| Subgroup assessment | none |

**Dikid et al. 2020**

| | |
|---|---|
| **Study information** | |
| Author | Dikid T, Patel P, Athotra A, Vaisakh TP, Jain S, Covid N |
| Title | Impact of Nonpharmacological Interventions on COVID-19 Transmission Dynamics in India |
| Year | 2020 |
| Journal | Indian Journal of Public Health |
| DOI | 10.4103/ijph.ijph__510__20 |
| Date received | 2020-05-05 |
| Date accepted | 2020-05-12 |
| **Study setting** | |
| Number of populations included | single |
| Level of populations included | national (India) |
| Geographic areas covered | Asia |
| Number of countries covered | single country (India) |
| Study period | end date in growth phase of wave |

– continued on next page –

| | |
|---|---|
| **Outcome** | |
| Raw outcome | epidemiological population-level outcome (cases) |
| Time resolution of raw outcome | daily |
| Computed outcome | measure of epidemic trend (doubling time), epidemiological parameter (reproduction number) |
| Method to obtain the computed outcome | statistical estimation of reproduction number (White et al., Wallinga and Teunis), simple computation (doubling time) |
| Data source | data from (sub)national authorities |
| Data availability | data access via source |
| **Intervention** | |
| Terminology for interventions | interventions |
| Terminology for the specific type of non-pharmaceutical interventions | non-pharmacological |
| Exposure types | one combination of interventions (Indian National Lockdown) |
| Types of single interventions | – not applicable – |
| Coding of interventions | not necessary |
| Source of intervention data | government or news websites |
| Availability of data on exposure | raw data documented in the manuscript |
| **Methodological approach** | |
| Empirical approach | descriptive |
| Use of exposure variation | only variation over time for a single population |
| Method | description of change over time (comparison of time periods) |
| Code availability | none |
| **Effect assessment** | |
| Reporting of intervention effect | comparison of outcome values |
| Measure of effect | reproduction number, doubling time |

– continued on next page –

| | |
|---|---|
| Reporting of uncertainty | yes |
| Sensitivity analysis | none |
| Subgroup assessment | none |

**Djurović 2020**

| | |
|---|---|
| **Study information** | |
| Author | Djurović I |
| Title | Epidemiological Control Measures and Predicted Number of Infections for SARS-CoV-2 Pandemic: Case Study Serbia March–April 2020 |
| Year | 2020 |
| Journal | Heliyon |
| DOI | 10.1016/j.heliyon.2020.e04238 |
| Date received | 2020-04-21 |
| Date accepted | 2020-06-15 |
| **Study setting** | |
| Number of populations included | single |
| Level of populations included | national (Serbia) |
| Geographic areas covered | Europe |
| Number of countries covered | single country (Serbia) |
| Study period | end date at peak of wave |
| **Outcome** | |
| Raw outcome | epidemiological population-level outcome (cases) |
| Time resolution of raw outcome | daily |
| Computed outcome | none |

– continued on next page –

| | |
|---|---|
| Method to obtain the computed outcome | – not applicable – |
| Data source | data from (sub)national authorities |
| Data availability | data access via source |
| **Intervention** | |
| Terminology for interventions | measures |
| Terminology for the specific type of non-pharmaceutical interventions | control |
| Exposure types | all interventions together (timeline of measures) |
| Types of single interventions | – not applicable – |
| Coding of interventions | – not applicable – |
| Source of intervention data | – not applicable – |
| Availability of data on exposure | – not applicable – |
| **Methodological approach** | |
| Empirical approach | counterfactual |
| Use of exposure variation | only variation over time for a single population |
| Method | non-mechanistic model (exponential growth model) |
| Code availability | none |
| **Effect assessment** | |
| Reporting of intervention effect | qualitative statement |
| Measure of effect | cases |
| Reporting of uncertainty | – not applicable – |
| Sensitivity analysis | different coding of interventions used (model fit on different time periods with NPIs) |
| Subgroup assessment | none |

| Domenico et al. 2020 | |
|---|---|
| **Study information** | |
| Author | Domenico LD, Pullano G, Sabbatini CE, Boëlle PY, Colizza V |
| Title | Impact of Lockdown on COVID-19 Epidemic in Île-de-France and Possible Exit Strategies |
| Year | 2020 |
| Journal | BMC Medicine |
| DOI | 10.1186/s12916-020-01698-4 |
| Date received | 2020-04-28 |
| Date accepted | 2020-07-06 |
| **Study setting** | |
| Number of populations included | single |
| Level of populations included | subnational (Ile de France) |
| Geographic areas covered | Europe |
| Number of countries covered | single country (France) |
| Study period | start and end date span first epidemic wave |
| **Outcome** | |
| Raw outcome | epidemiological population-level outcome (hospitalizations, ICU beds occupation) |
| Time resolution of raw outcome | daily |
| Computed outcome | none |
| Method to obtain the computed outcome | – not applicable – |
| Data source | data from (sub)national authorities |
| Data availability | data access via source |

– continued on next page –

| | |
|---|---|
| **Intervention** | |
| Terminology for interventions | measures |
| Terminology for the specific type of non-pharmaceutical interventions | social distancing |
| Exposure types | one combination of interventions (national lockdown per decree of March 18) |
| Types of single interventions | – not applicable – |
| Coding of interventions | not necessary |
| Source of intervention data | government or news websites |
| Availability of data on exposure | raw data documented in the manuscript |
| **Methodological approach** | |
| Empirical approach | counterfactual |
| Use of exposure variation | only variation over time for a single population |
| Method | mechanistic model (compartmental single-population transmission model) |
| Code availability | none |
| **Effect assessment** | |
| Reporting of intervention effect | qualitative statement |
| Measure of effect | ICU beds occupation |
| Reporting of uncertainty | – not applicable – |
| Sensitivity analysis | epidemiological parameters varied |
| Subgroup assessment | none |

**Donsimoni et al. 2020**

**Study information**

– continued on next page –

| | |
|---|---|
| Author | Donsimoni JR, Glawion R, Plachter B, Wälde K, Weiser C |
| Title | Should Contact Bans Have Been Lifted More in Germany? |
| Year | 2020 |
| Journal | CESifo Economic Studies |
| DOI | 10.1093/cesifo/ifaa004 |
| Date received | – could not be evaluated – |
| Date accepted | 2020-07-03(published) |
| **Study setting** | |
| Number of populations included | single |
| Level of populations included | national (Germany) |
| Geographic areas covered | Europe |
| Number of countries covered | single country (Germany) |
| Study period | start and end date span first epidemic wave |
| **Outcome** | |
| Raw outcome | epidemiological population-level outcome (cases) |
| Time resolution of raw outcome | daily |
| Computed outcome | none |
| Method to obtain the computed outcome | – not applicable – |
| Data source | data from (sub)national authorities |
| Data availability | data access via source |
| **Intervention** | |
| Terminology for interventions | measures |
| Terminology for the specific type of non-pharmaceutical interventions | contact ban |

– continued on next page –

| | |
|---|---|
| Exposure types | one combination of interventions (social contact restrictions (including major public events suspended, schools closed, domestic movements banned)) |
| Types of single interventions | – not applicable – |
| Coding of interventions | necessary |
| Source of intervention data | use of externally coded data (OxCGRT) |
| Availability of data on exposure | access to externally coded data via source |
| **Methodological approach** | |
| Empirical approach | counterfactual |
| Use of exposure variation | only variation over time for a single population |
| Method | mechanistic model (compartmental single-population transmission model) |
| Code availability | none |
| **Effect assessment** | |
| Reporting of intervention effect | qualitative statement |
| Measure of effect | cases |
| Reporting of uncertainty | – not applicable – |
| Sensitivity analysis | none |
| Subgroup assessment | none |

**Du et al. 2020 (Analysis 1)**

| | |
|---|---|
| **Study information** | |
| Author | Du Z, Xu X, Wang L, Fox SJ, Cowling BJ, Galvani AP, Meyers LA |
| Title | Effects of Proactive Social Distancing on COVID-19 Outbreaks in 58 Cities, China |

– continued on next page –

| | |
|---|---|
| Year | 2020 |
| Journal | Emerging Infectious Diseases |
| DOI | 10.3201/eid2609.201932 |
| Date received | – could not be evaluated – |
| Date accepted | 2020-06-09 (published) |
| **Study setting** | |
| Number of populations included | multiple |
| Level of populations included | subnational (58 cities in China outside of Wuhan) |
| Geographic areas covered | Asia |
| Number of countries covered | single country (China) |
| Study period | start and end date span first epidemic wave |
| **Outcome** | |
| Raw outcome | epidemiological population-level outcome (cases) |
| Time resolution of raw outcome | daily |
| Computed outcome | epidemiological parameter (reproduction number) |
| Method to obtain the computed outcome | statistical estimation of reproduction number (Cori et al.) |
| Data source | data from (sub)national authorities |
| Data availability | data access via source |
| **Intervention** | |
| Terminology for interventions | measures |
| Terminology for the specific type of non-pharmaceutical interventions | social distancing |
| Exposure types | start of interventions (first of many social distancing measures implemented (including venue closure, isolation, travel restrictions)) |
| Types of single interventions | – not applicable – |

– continued on next page –

| | |
|---|---|
| Coding of interventions | necessary |
| Source of intervention data | coding done by authors |
| Availability of data on exposure | coded data made available by the authors |
| **Methodological approach** | |
| Empirical approach | descriptive |
| Use of exposure variation | only variation over time for multiple populations |
| Method | description of change over time (comparison of time periods) |
| Code availability | none |
| **Effect assessment** | |
| Reporting of intervention effect | quantification of change in outcome values |
| Measure of effect | reproduction number |
| Reporting of uncertainty | yes |
| Sensitivity analysis | none |
| Subgroup assessment | none |

**Du et al. 2020 (Analysis 2)**

| | |
|---|---|
| **Study information** | |
| Author | Du Z, Xu X, Wang L, Fox SJ, Cowling BJ, Galvani AP, Meyers LA |
| Title | Effects of Proactive Social Distancing on COVID-19 Outbreaks in 58 Cities, China |
| Year | 2020 |
| Journal | Emerging Infectious Diseases |
| DOI | 10.3201/eid2609.201932 |
| Date received | – could not be evaluated – |

– continued on next page –

| | |
|---|---|
| Date accepted | 2020-06-09 (published) |
| **Study setting** | |
| Number of populations included | multiple |
| Level of populations included | subnational (58 cities in China outside of Wuhan) |
| Geographic areas covered | Asia |
| Number of countries covered | single country (China) |
| Study period | start and end date span first epidemic wave |
| **Outcome** | |
| Raw outcome | epidemiological population-level outcome (cases) |
| Time resolution of raw outcome | daily |
| Computed outcome | summary statistic (time until reproduction number below 1) |
| Method to obtain the computed outcome | statistical estimation of reproduction number (Cori et al.) |
| Data source | data from (sub)national authorities |
| Data availability | data access via source |
| **Intervention** | |
| Terminology for interventions | measures |
| Terminology for the specific type of non-pharmaceutical interventions | social distancing |
| Exposure types | start of interventions (first of many social distancing measures implemnted (including venue closure, isolation, travel restrictions)) |
| Types of single interventions | – not applicable – |
| Coding of interventions | necessary |
| Source of intervention data | coding done by authors |
| Availability of data on exposure | coded data made available by the authors |
| **Methodological approach** | |

– continued on next page –

| | |
|---|---|
| Empirical approach | parametric |
| Use of exposure variation | only variation between populations |
| Method | non-mechanistic model (generalized linear model) |
| Code availability | none |
| **Effect assessment** | |
| Reporting of intervention effect | quantification of change in outcome values |
| Measure of effect | time until reproduction number below 1 |
| Reporting of uncertainty | yes |
| Sensitivity analysis | none |
| Subgroup assessment | none |

**Duhon et al. 2021**

| | |
|---|---|
| **Study information** | |
| Author | Duhon J, Bragazzi N, Kong JD |
| Title | The Impact of Non-Pharmaceutical Interventions, Demographic, Social, and Climatic Factors on the Initial Growth Rate of COVID-19: A Cross-Country Study |
| Year | 2021 |
| Journal | Science of The Total Environment |
| DOI | 10.1016/j.scitotenv.2020.144325 |
| Date received | 2020-09-19 |
| Date accepted | 2020-12-05 |
| **Study setting** | |
| Number of populations included | multiple |

– continued on next page –

| | |
|---|---|
| Level of populations included | national (– could not be evaluated – (probably many countries, over 100)) |
| Geographic areas covered | Europe, Middle East and Africa, Asia, Oceania, Central and South America, North America |
| Number of countries covered | multiple countries |
| Study period | same end date for several populations with diverse epidemic trajectories |
| **Outcome** | |
| Raw outcome | epidemiological population-level outcome (cases) |
| Time resolution of raw outcome | daily |
| Computed outcome | measure of epidemic trend (growth rate) |
| Method to obtain the computed outcome | exponential growth model |
| Data source | data from publicly available cross-country selections (OurWorldInData) |
| Data availability | data access via source |
| **Intervention** | |
| Terminology for interventions | interventions |
| Terminology for the specific type of non-pharmaceutical interventions | non-pharmaceutical |
| Exposure types | multiple separate interventions (workplace closure, internal movement restrictions) |
| Types of single interventions | workplace closure, internal movement restrictions |
| Coding of interventions | necessary |
| Source of intervention data | use of externally coded data (OxCGRT) |
| Availability of data on exposure | access to externally coded data via source |
| **Methodological approach** | |
| Empirical approach | parametric |
| Use of exposure variation | both variation over time and between populations |

– continued on next page –

| | |
|---|---|
| Method | non-mechanistic model (generalized linear model) |
| Code availability | none |
| **Effect assessment** | |
| Reporting of intervention effect | quantification of change in outcome values |
| Measure of effect | growth rate |
| Reporting of uncertainty | yes |
| Sensitivity analysis | none |
| Subgroup assessment | none |

**Džiugys et al. 2020**

| | |
|---|---|
| **Study information** | |
| Author | Džiugys A, Bieliūnas M, Skarbalius G, Misiulis E, Navakas R |
| Title | Simplified Model of Covid-19 Epidemic Prognosis under Quarantine and Estimation of Quarantine Effectiveness |
| Year | 2020 |
| Journal | Chaos, Solitons & Fractals |
| DOI | 10.1016/j.chaos.2020.110162 |
| Date received | 2020-06-18 |
| Date accepted | 2020-07-25 |
| **Study setting** | |
| Number of populations included | multiple |
| Level of populations included | national (53 countries worldwide) |
| Geographic areas covered | Europe, Middle East and Africa, Asia, Oceania, Central and South America, North America |

– continued on next page –

| | |
|---|---|
| Number of countries covered | multiple countries |
| Study period | same end date for several populations with diverse epidemic trajectories |
| **Outcome** | |
| Raw outcome | epidemiological population-level outcome (cases) |
| Time resolution of raw outcome | daily |
| Computed outcome | measure of epidemic trend (growth rate) |
| Method to obtain the computed outcome | simple computation |
| Data source | data from publicly available cross-country selections (ECDC) |
| Data availability | data access via source |
| **Intervention** | |
| Terminology for interventions | measures |
| Terminology for the specific type of non-pharmaceutical interventions | quarantine |
| Exposure types | start of interventions (day of the first intervention leading to lockdown) |
| Types of single interventions | – not applicable – |
| Coding of interventions | necessary |
| Source of intervention data | use of externally coded data (Wikipedia) |
| Availability of data on exposure | access to externally coded data via source |
| **Methodological approach** | |
| Empirical approach | parametric |
| Use of exposure variation | only variation over time for multiple populations |
| Method | non-mechanistic model (generalized linear model) |
| Code availability | none |
| **Effect assessment** | |
| Reporting of intervention effect | quantification of change in outcome values |

– continued on next page –

| | |
|---|---|
| Measure of effect | growth rate |
| Reporting of uncertainty | no |
| Sensitivity analysis | none |
| Subgroup assessment | none |

**Edjoc et al. 2020**

| | |
|---|---|
| **Study information** | |
| Author | Edjoc R, Atchessi N, Lien A, Smith BA, Gabrani-Juma I, Abalos C, Heisz M |
| Title | Assessing the Progression of the COVID-19 Pandemic in Canada Using Testing Data and Time-Dependent Reproduction Numbers |
| Year | 2020 |
| Journal | Canadian Journal of Public Health |
| DOI | 10.17269/s41997-020-00428-w |
| Date received | 2020-06-23 |
| Date accepted | 2020-09-29 |
| **Study setting** | |
| Number of populations included | multiple |
| Level of populations included | both national and subnational (Canada) |
| Geographic areas covered | North America |
| Number of countries covered | single country (Canada) |
| Study period | start and end date span first epidemic wave |
| **Outcome** | |
| Raw outcome | epidemiological population-level outcome (cases) |

– continued on next page –

| | |
|---|---|
| Time resolution of raw outcome | daily |
| Computed outcome | epidemiological parameter (reproduction number) |
| Method to obtain the computed outcome | statistical estimation of reproduction number (Walinga et al.) |
| Data source | data from (sub)national authorities |
| Data availability | data access via source |
| **Intervention** | |
| Terminology for interventions | interventions |
| Terminology for the specific type of non-pharmaceutical interventions | public health |
| Exposure types | all interventions together (gathering bans, closing non-essential businesses, implementation of telework, closing public venues, closing schools and universities) |
| Types of single interventions | – not applicable – |
| Coding of interventions | – not applicable – |
| Source of intervention data | – not applicable – |
| Availability of data on exposure | – not applicable – |
| **Methodological approach** | |
| Empirical approach | descriptive |
| Use of exposure variation | only variation over time for multiple populations |
| Method | description of change over time (description of time course) |
| Code availability | none |
| **Effect assessment** | |
| Reporting of intervention effect | qualitative statement |
| Measure of effect | cases, reproduction number |
| Reporting of uncertainty | – not applicable – |

– continued on next page –

| | |
|---|---|
| Sensitivity analysis | none |
| Subgroup assessment | none |

**Eguchi et al. 2020**

| | |
|---|---|
| **Study information** | |
| Author | Eguchi A, Yoneoka D, Shi S, Tanoue Y, Kawashima T, Nomura S, Matsuura K, Makiyama K, Ejima K, Gilmour S, Nishiura H, Miyata H |
| Title | Trend Change of the Transmission Route of COVID-19–Related Symptoms in Japan |
| Year | 2020 |
| Journal | Public Health |
| DOI | 10.1016/j.puhe.2020.08.020 |
| Date received | 2020-06-16 |
| Date accepted | 2020-08-20 |
| **Study setting** | |
| Number of populations included | single |
| Level of populations included | national (Japan) |
| Geographic areas covered | Asia |
| Number of countries covered | single country (Japan) |
| Study period | start and end date span first epidemic wave: behavioral |
| **Outcome** | |
| Raw outcome | behavioral outcome (survey: contacts with infected individuals and fever symptoms) |
| Time resolution of raw outcome | daily |
| Computed outcome | none |

– continued on next page –

| | |
|---|---|
| Method to obtain the computed outcome | – not applicable – |
| Data source | data from (sub)national authorities |
| Data availability | data not accessible |
| **Intervention** | |
| Terminology for interventions | – not applicable – |
| Terminology for the specific type of non-pharmaceutical interventions | emergency declaration |
| Exposure types | one single intervention (declaration of a state of emergency on April 7 in some prefectures and nationwide on April 16) |
| Types of single interventions | declaration of a state of emergency |
| Coding of interventions | not necessary |
| Source of intervention data | government or news websites |
| Availability of data on exposure | raw data documented in the manuscript |
| **Methodological approach** | |
| Empirical approach | descriptive |
| Use of exposure variation | only variation over time for a single population |
| Method | description of change over time (description of time course) |
| Code availability | none |
| **Effect assessment** | |
| Reporting of intervention effect | qualitative statement |
| Measure of effect | contacts with infected individuals, fever symptoms |
| Reporting of uncertainty | – not applicable – |
| Sensitivity analysis | none |
| Subgroup assessment | epidemiological indicators (infection setting) |

| Fatmi 2020 | |
|---|---|
| **Study information** | |
| Author | Fatmi MR |
| Title | COVID-19 Impact on Urban Mobility |
| Year | 2020 |
| Journal | Journal of Urban Management |
| DOI | 10.1016/j.jum.2020.08.002 |
| Date received | 2020-07-22 |
| Date accepted | 2020-08-03 |
| **Study setting** | |
| Number of populations included | single |
| Level of populations included | subnational (Kelowna region of British Columbia) |
| Geographic areas covered | North America |
| Number of countries covered | single country (Canada) |
| Study period | start and end date span first epidemic wave: behavioral |
| **Outcome** | |
| Raw outcome | behavioral outcome (survey: travel activities) |
| Time resolution of raw outcome | – not applicable – |
| Computed outcome | none |
| Method to obtain the computed outcome | – not applicable – |
| Data source | data collected by authors |
| Data availability | data not accessible |
| **Intervention** | |
| Terminology for interventions | – not applicable – |

– continued on next page –

| | |
|---|---|
| Terminology for the specific type of non-pharmaceutical interventions | travel restrictions |
| Exposure types | all interventions together |
| Types of single interventions | – not applicable – |
| Coding of interventions | – not applicable – |
| Source of intervention data | – not applicable – |
| Availability of data on exposure | – not applicable – |
| **Methodological approach** | |
| Empirical approach | descriptive |
| Use of exposure variation | only variation over time for a single population |
| Method | description of change over time (comparison of time periods) |
| Code availability | none |
| **Effect assessment** | |
| Reporting of intervention effect | comparison of outcome values |
| Measure of effect | mobility: travel activities |
| Reporting of uncertainty | no |
| Sensitivity analysis | none |
| Subgroup assessment | none |

**Feng et al. 2020**

| | |
|---|---|
| **Study information** | |
| Author | Feng X, Chen AJ, Wang K, Wang L, Zhang F, Jin Z, Zou L, Wang X, and, and, And |
| Title | Phase-Adjusted Estimation of the COVID-19 Outbreak in South Korea under Multi-Source Data and Adjustment Measures: A Modelling Study |

– continued on next page –

| | |
|---|---|
| Year | 2020 |
| Journal | Mathematical Biosciences and Engineering |
| DOI | 10.3934/mbe.2020205 |
| Date received | 2020-03-30 |
| Date accepted | 2020-05-06 |
| **Study setting** | |
| Number of populations included | single |
| Level of populations included | national (South Korea) |
| Geographic areas covered | Asia |
| Number of countries covered | single country (South Korea) |
| Study period | start and end date span first epidemic wave |
| **Outcome** | |
| Raw outcome | epidemiological population-level outcome (cases, deaths, recovered cases) |
| Time resolution of raw outcome | daily |
| Computed outcome | epidemiological parameter (reproduction number) |
| Method to obtain the computed outcome | compartmental transmission model (single-population) |
| Data source | data from (sub)national authorities |
| Data availability | data access via source |
| **Intervention** | |
| Terminology for interventions | measures |
| Terminology for the specific type of non-pharmaceutical interventions | lockdown, control |
| Exposure types | multiple combinations of interventions (second phase: people are encouraged to work at home and reduce the unnecessary gathering activities and so on, third phase: free tests to people in a less crowded manner) |

– continued on next page –

| | |
|---|---|
| Types of single interventions | – not applicable – |
| Coding of interventions | not necessary |
| Source of intervention data | – could not be evaluated – |
| Availability of data on exposure | raw data documented in the manuscript |
| **Methodological approach** | |
| Empirical approach | descriptive |
| Use of exposure variation | only variation over time for a single population |
| Method | description of change over time (comparison of time periods) |
| Code availability | none |
| **Effect assessment** | |
| Reporting of intervention effect | comparison of outcome values |
| Measure of effect | reproduction number |
| Reporting of uncertainty | yes |
| Sensitivity analysis | epidemiological parameters varied |
| Subgroup assessment | none |

**Fernández-Recio 2020**

| | |
|---|---|
| **Study information** | |
| Author | Fernández-Recio J |
| Title | Modelling the Evolution of COVID-19 in High-Incidence European Countries and Regions: Estimated Number of Infections and Impact of Past and Future Intervention Measures |
| Year | 2020 |
| Journal | Journal of Clinical Medicine |

– continued on next page –

| | |
|---|---|
| DOI | 10.3390/jcm9061825 |
| Date received | 2020-05-18 |
| Date accepted | 2020-06-08 |
| **Study setting** | |
| Number of populations included | multiple |
| Level of populations included | both national and subnational (Spain, Italy, UK, Germany, France, Iceland, La Rioja) |
| Geographic areas covered | Europe |
| Number of countries covered | multiple countries |
| Study period | start and end date span first epidemic wave |
| **Outcome** | |
| Raw outcome | epidemiological population-level outcome (cases, deaths) |
| Time resolution of raw outcome | daily |
| Computed outcome | none |
| Method to obtain the computed outcome | – not applicable – |
| Data source | data from publicly available cross-country selections (ECDC) |
| Data availability | data access via source |
| **Intervention** | |
| Terminology for interventions | measures |
| Terminology for the specific type of non-pharmaceutical interventions | intervention |
| Exposure types | one combination of interventions (lockdown (dates inferred from Flaxman study)) |
| Types of single interventions | – not applicable – |
| Coding of interventions | necessary |

– continued on next page –

**Fernández-Recio 2020** – continued from previous page

| | |
|---|---|
| Source of intervention data | use of externally coded data (same as in Flaxman et al.) |
| Availability of data on exposure | coded data made available by the authors |
| **Methodological approach** | |
| Empirical approach | parametric |
| Use of exposure variation | only variation over time for multiple populations |
| Method | mechanistic model (semi-mechanistic Bayesian transmission model) |
| Latent variable functionally linked to intervention | reproduction number |
| Code availability | none |
| **Effect assessment** | |
| Reporting of intervention effect | quantification of change in outcome values |
| Measure of effect | reproduction number |
| Reporting of uncertainty | yes |
| Sensitivity analysis | none |
| Subgroup assessment | none |

**Flaxman et al. 2020**

| |
|---|
| **Study information** |

– continued on next page –

| | |
|---|---|
| Author | Flaxman S, Mishra S, Gandy A, Unwin HJ, Mellan TA, Coupland H, Whittaker C, Zhu H, Berah T, Eaton JW, Monod M, Perez-Guzman PN, Schmit N, Cilloni L, Ainslie KE, Baguelin M, Boonyasiri A, Boyd O, Cattarino L, Cooper LV, Cucunubá Z, Cuomo-Dannenburg G, Dighe A, Djaafara B, Dorigatti I, van Elsland SL, FitzJohn RG, Gaythorpe KA, Geidelberg L, Grassly NC, Green WD, Hallett T, Hamlet A, Hinsley W, Jeffrey B, Knock E, Laydon DJ, Nedjati-Gilani G, Nouvellet P, Parag KV, Siveroni I, Thompson HA, Verity R, Volz E, Walters CE, Wang H, Wang Y, Watson OJ, Winskill P, Xi X, Walker PG, Ghani AC, Donnelly CA, Riley S, Vollmer MA, Ferguson NM, Okell LC, And SB |
| Title | Estimating the Effects of Non-Pharmaceutical Interventions on COVID-19 in Europe |
| Year | 2020 |
| Journal | Nature |
| DOI | 10.1038/s41586-020-2405-7 |
| Date received | 2020-03-30 |
| Date accepted | 2020-05-22 |
| **Study setting** | |
| Number of populations included | multiple |
| Level of populations included | national (11 European countries) |
| Geographic areas covered | Europe |
| Number of countries covered | multiple countries |
| Study period | start and end date span first epidemic wave |
| **Outcome** | |
| Raw outcome | epidemiological population-level outcome (deaths) |
| Time resolution of raw outcome | daily |
| Computed outcome | none |

– continued on next page –

| | |
|---|---|
| Method to obtain the computed outcome | – not applicable – |
| Data source | data from publicly available cross-country selections (ECDC) |
| Data availability | data made available by the authors |
| **Intervention** | |
| Terminology for interventions | interventions |
| Terminology for the specific type of non-pharmaceutical interventions | non-pharmaceutical |
| Exposure types | multiple separate interventions (lockdown (gathering bans, venue closure), public events banned, primary and secondary school closure, self isolaton, social distancing encouraged) |
| Types of single interventions | – not applicable – |
| Coding of interventions | necessary |
| Source of intervention data | coding done by authors |
| Availability of data on exposure | coded data made available by the authors |
| **Methodological approach** | |
| Empirical approach | parametric |
| Use of exposure variation | both variation over time and between populations |
| Method | mechanistic model (semi-mechanistic Bayesian transmission model) |
| Latent variable functionally linked to intervention | reproduction number |
| Code availability | publicly available |
| **Effect assessment** | |
| Reporting of intervention effect | quantification of change in outcome values |
| Measure of effect | reproduction number |
| Reporting of uncertainty | yes |

– continued on next page –

| | |
|---|---|
| Sensitivity analysis | start or end date of study period varied, same analysis with (sub)population(s) excluded, model specification varied, epidemiological parameters varied |
| Subgroup assessment | none |

| **Frank 2020** | |
|---|---|
| **Study information** | |
| Author | Frank TD |
| Title | COVID-19 Interventions in Some European Countries Induced Bifurcations Stabilizing Low Death States against High Death States: An Eigenvalue Analysis Based on the Order Parameter Concept of Synergetics |
| Year | 2020 |
| Journal | Chaos, Solitons & Fractals |
| DOI | 10.1016/j.chaos.2020.110194 |
| Date received | 2020-07-15 |
| Date accepted | 2020-08-06 |
| **Study setting** | |
| Number of populations included | multiple |
| Level of populations included | national (20 European countries) |
| Geographic areas covered | Europe |
| Number of countries covered | multiple countries |
| Study period | start and end date span first epidemic wave |
| **Outcome** | |
| Raw outcome | epidemiological population-level outcome (cases) |
| Time resolution of raw outcome | daily |

– continued on next page –

| | |
|---|---|
| Computed outcome | other (order parameter eigenvalue) |
| Method to obtain the computed outcome | compartmental transmission model (single-population) |
| Data source | data from publicly available cross-country selections (JHU) |
| Data availability | data access via source |
| **Intervention** | |
| Terminology for interventions | interventions |
| Terminology for the specific type of non-pharmaceutical interventions | government |
| Exposure types | all interventions together |
| Types of single interventions | – not applicable – |
| Coding of interventions | – not applicable – |
| Source of intervention data | – not applicable – |
| Availability of data on exposure | – not applicable – |
| **Methodological approach** | |
| Empirical approach | descriptive |
| Use of exposure variation | only variation over time for multiple populations |
| Method | description of change over time (comparison of time periods) |
| Code availability | none |
| **Effect assessment** | |
| Reporting of intervention effect | qualitative statement |
| Measure of effect | order parameter eigenvalue |
| Reporting of uncertainty | – not applicable – |
| Sensitivity analysis | none |
| Subgroup assessment | none |

| Friedrich et al. 2020 | |
|---|---|
| **Study information** | |
| Author | Friedrich F, Ongaratto R, Scotta MC, Veras TN, Stein RT, Lumertz MS, Jones MH, Comaru T, Pinto LA |
| Title | Early Impact of Social Distancing in Response to Coronavirus Disease 2019 on Hospitalizations for Acute Bronchiolitis in Infants in Brazil |
| Year | 2020 |
| Journal | Clinical Infectious Diseases |
| DOI | 10.1093/cid/ciaa1458 |
| Date received | 2020-08-12 |
| Date accepted | 2020-09-22 |
| **Study setting** | |
| Number of populations included | multiple |
| Level of populations included | both national and subnational (Brazil and 5 of its macro-regions) |
| Geographic areas covered | Central and South America |
| Number of countries covered | single country (Brazil) |
| Study period | end date in growth phase of wave |
| **Outcome** | |
| Raw outcome | epidemiological population-level outcome (surrogate disease: hospitalizations with acute bronchiolitis in infants) |
| Time resolution of raw outcome | monthly |
| Computed outcome | none |
| Method to obtain the computed outcome | – not applicable – |
| Data source | data from (sub)national authorities |
| Data availability | data access via source |

– continued on next page –

| | |
|---|---|
| **Intervention** | |
| Terminology for interventions | measures |
| Terminology for the specific type of non-pharmaceutical interventions | social distancing |
| Exposure types | all interventions together |
| Types of single interventions | – not applicable – |
| Coding of interventions | – not applicable – |
| Source of intervention data | – not applicable – |
| Availability of data on exposure | – not applicable – |
| **Methodological approach** | |
| Empirical approach | descriptive |
| Use of exposure variation | only variation over time for multiple populations |
| Method | description of change over time (comparison of time periods) |
| Code availability | none |
| **Effect assessment** | |
| Reporting of intervention effect | quantification of change in outcome values |
| Measure of effect | hospitalizations: acute bronchiolitis in infants |
| Reporting of uncertainty | yes |
| Sensitivity analysis | none |
| Subgroup assessment | none |

**Friedson et al. 2020 (Analysis 1)**

**Study information**

– continued on next page –

| | |
|---|---|
| Author | Friedson AI, McNichols D, Sabia JJ, Dave D |
| Title | Shelter-in-Place Orders and Public Health: Evidence from California during the COVID-19 Pandemic |
| Year | 2020 |
| Journal | Journal of Policy Analysis and Management |
| DOI | 10.1002/pam.22267 |
| Date received | – could not be evaluated – |
| Date accepted | 2020-12-11 |
| **Study setting** | |
| Number of populations included | single |
| Level of populations included | subnational (California) |
| Geographic areas covered | North America |
| Number of countries covered | single country (California) |
| Study period | end date in growth phase of wave |
| **Outcome** | |
| Raw outcome | epidemiological population-level outcome (cases, deaths), behavioral outcome (mobility: shelter-in-place index) |
| Time resolution of raw outcome | daily |
| Computed outcome | none |
| Method to obtain the computed outcome | – not applicable – |
| Data source | data from public media outlets (New York Times: cases), mobility data from corporate organizations (SafeGraph: mobility) |
| Data availability | data access via source |
| **Intervention** | |
| Terminology for interventions | – not applicable – |

– continued on next page –

| | |
|---|---|
| Terminology for the specific type of non-pharmaceutical interventions | shelter-in-place order |
| Exposure types | one single intervention (stay-at-home order) |
| Types of single interventions | stay-at-home order |
| Coding of interventions | not necessary |
| Source of intervention data | – could not be evaluated – |
| Availability of data on exposure | raw data documented in the manuscript |
| **Methodological approach** | |
| Empirical approach | counterfactual |
| Use of exposure variation | only variation over time for a single population |
| Method | synthetic controls |
| Code availability | none |
| **Effect assessment** | |
| Reporting of intervention effect | quantification of change in outcome values |
| Measure of effect | cases, deaths, mobility: shelter-in-place index |
| Reporting of uncertainty | yes |
| Sensitivity analysis | same analysis with (sub)population(s) excluded, model specification varied |
| Subgroup assessment | none |

**Friedson et al. 2020 (Analysis 2)**

| | |
|---|---|
| **Study information** | |
| Author | Friedson AI, McNichols D, Sabia JJ, Dave D |
| Title | Shelter-in-Place Orders and Public Health: Evidence from California during the COVID-19 Pandemic |
| Year | 2020 |

– continued on next page –

| | |
|---|---|
| Journal | Journal of Policy Analysis and Management |
| DOI | 10.1002/pam.22267 |
| Date received | – could not be evaluated – |
| Date accepted | 2020-12-11 |
| **Study setting** | |
| Number of populations included | multiple |
| Level of populations included | both national and subnational (US states) |
| Geographic areas covered | North America |
| Number of countries covered | single country (US) |
| Study period | end date in growth phase of wave |
| **Outcome** | |
| Raw outcome | epidemiological population-level outcome (cases, deaths) |
| Time resolution of raw outcome | daily |
| Computed outcome | none |
| Method to obtain the computed outcome | – not applicable – |
| Data source | data from public media outlets (New York Times) |
| Data availability | data access via source |
| **Intervention** | |
| Terminology for interventions | – not applicable – |
| Terminology for the specific type of non-pharmaceutical interventions | shelter-in-place order |
| Exposure types | one single intervention (stay-at-home order) |
| Types of single interventions | stay-at-home order |
| Coding of interventions | not necessary |
| Source of intervention data | – could not be evaluated – |

– continued on next page –

| | |
|---|---|
| Availability of data on exposure | raw data documented in the manuscript |
| **Methodological approach** | |
| Empirical approach | parametric |
| Use of exposure variation | only variation over time for a single population |
| Method | non-mechanistic model (generalized linear model) |
| Code availability | none |
| **Effect assessment** | |
| Reporting of intervention effect | quantification of change in outcome values |
| Measure of effect | cases, deaths |
| Reporting of uncertainty | yes |
| Sensitivity analysis | same analysis with (sub)population(s) excluded, model specification varied |
| Subgroup assessment | none |

| **Gallaway et al. 2020** | |
|---|---|
| **Study information** | |
| Author | Gallaway MS, Rigler J, Robinson S, Herrick K, Livar E, Komatsu KK, Brady S, Cunico J, Christ CM |
| Title | Trends in COVID-19 Incidence after Implementation of Mitigation Measures —Arizona, January 22–August 7, 2020 |
| Year | 2020 |
| Journal | Morbidity and Mortality Weekly Report |
| DOI | 10.15585/mmwr.mm6940e3 |
| Date received | – could not be evaluated – |
| Date accepted | 2020-08-07 (published) |

– continued on next page –

| | |
|---|---|
| **Study setting** | |
| Number of populations included | single |
| Level of populations included | subnational (Arizona) |
| Geographic areas covered | North America |
| Number of countries covered | single country (US) |
| Study period | start and end date span first epidemic wave |
| **Outcome** | |
| Raw outcome | epidemiological population-level outcome (cases) |
| Time resolution of raw outcome | daily |
| Computed outcome | none |
| Method to obtain the computed outcome | – not applicable – |
| Data source | data from (sub)national authorities |
| Data availability | data access via source |
| **Intervention** | |
| Terminology for interventions | measures |
| Terminology for the specific type of non-pharmaceutical interventions | mitigation |
| Exposure types | all interventions together (timeline of measures) |
| Types of single interventions | – not applicable – |
| Coding of interventions | – not applicable – |
| Source of intervention data | – not applicable – |
| Availability of data on exposure | – not applicable – |
| **Methodological approach** | |
| Empirical approach | descriptive |
| Use of exposure variation | only variation over time for a single population |

– continued on next page –

| | |
|---|---|
| Method | description of change over time (comparison of time periods) |
| Code availability | none |
| **Effect assessment** | |
| Reporting of intervention effect | comparison of outcome values |
| Measure of effect | cases |
| Reporting of uncertainty | no |
| Sensitivity analysis | none |
| Subgroup assessment | none |

**Gao et al. 2020 (Analysis 1)**

| | |
|---|---|
| **Study information** | |
| Author | Gao S, Rao J, Kang Y, Liang Y, Kruse J, Dopfer D, Sethi AK, Reyes JF, Yandell BS, Patz JA |
| Title | Association of Mobile Phone Location Data Indications of Travel and Stay-at-Home Mandates with COVID-19 Infection Rates in the US |
| Year | 2020 |
| Journal | JAMA Network Open |
| DOI | 10.1001/jamanetworkopen.2020.20485 |
| Date received | – could not be evaluated – |
| Date accepted | 2020-07-31 |
| **Study setting** | |
| Number of populations included | multiple |
| Level of populations included | both national and subnational (US and all states) |

– continued on next page –

| | |
|---|---|
| Geographic areas covered | North America |
| Number of countries covered | single country (US) |
| Study period | same end date for several populations with diverse epidemic trajectories |
| **Outcome** | |
| Raw outcome | epidemiological population-level outcome (cases) |
| Time resolution of raw outcome | daily |
| Computed outcome | measure of epidemic trend (doubling time) |
| Method to obtain the computed outcome | exponential growth model |
| Data source | data from publicly available open-source projects (Corona Data Scraper: cases), data from (sub)national authorities (health authority: cases) |
| Data availability | data access via source |
| **Intervention** | |
| Terminology for interventions | measures |
| Terminology for the specific type of non-pharmaceutical interventions | non-pharmacological |
| Exposure types | one single intervention (stay-at-home order) |
| Types of single interventions | stay-at-home order |
| Coding of interventions | necessary |
| Source of intervention data | coding done by authors |
| Availability of data on exposure | coded data made available by the authors |
| **Methodological approach** | |
| Empirical approach | descriptive |
| Use of exposure variation | only variation over time for multiple populations |
| Method | description of change over time (comparison of time periods) |
| Code availability | none |

– continued on next page –

| | |
|---|---|
| **Effect assessment** | |
| Reporting of intervention effect | quantification of change in outcome values |
| Measure of effect | doubling time |
| Reporting of uncertainty | yes |
| Sensitivity analysis | model specification varied |
| Subgroup assessment | none |

| **Gao et al. 2020 (Analysis 2)** | |
|---|---|
| **Study information** | |
| Author | Gao S, Rao J, Kang Y, Liang Y, Kruse J, Dopfer D, Sethi AK, Reyes JF, Yandell BS, Patz JA |
| Title | Association of Mobile Phone Location Data Indications of Travel and Stay-at-Home Mandates with COVID-19 Infection Rates in the US |
| Year | 2020 |
| Journal | JAMA Network Open |
| DOI | 10.1001/jamanetworkopen.2020.20485 |
| Date received | – could not be evaluated – |
| Date accepted | 2020-07-31 |
| **Study setting** | |
| Number of populations included | multiple |
| Level of populations included | both national and subnational (US and all states) |
| Geographic areas covered | North America |
| Number of countries covered | single country (US) |
| Study period | start and end date span first epidemic wave: behavioral |
| **Outcome** | |

– continued on next page –

| | |
|---|---|
| Raw outcome | behavioral outcome (mobility: number of trips between and within counties relative to baseline) |
| Time resolution of raw outcome | daily |
| Computed outcome | none |
| Method to obtain the computed outcome | – not applicable – |
| Data source | mobility data from corporate organizations (Descartes Labs (mobility: travel distances), Safegraph (mobility: home dwell time)) |
| Data availability | data access via source |
| **Intervention** | |
| Terminology for interventions | measures |
| Terminology for the specific type of non-pharmaceutical interventions | non-pharmacological |
| Exposure types | one single intervention (stay-at-home order) |
| Types of single interventions | stay-at-home order |
| Coding of interventions | necessary |
| Source of intervention data | coding done by authors |
| Availability of data on exposure | coded data made available by the authors |
| **Methodological approach** | |
| Empirical approach | descriptive |
| Use of exposure variation | only variation over time for multiple populations |
| Method | description of change over time (comparison of time periods) |
| Code availability | none |
| **Effect assessment** | |
| Reporting of intervention effect | quantification of change in outcome values |
| Measure of effect | mobility: travel distance & home dwell time |

– continued on next page –

| | |
|---|---|
| Reporting of uncertainty | yes |
| Sensitivity analysis | none |
| Subgroup assessment | none |

**Gatto et al. 2020**

| | |
|---|---|
| **Study information** | |
| Author | Gatto M, Bertuzzo E, Mari L, Miccoli S, Carraro L, Casagrandi R, Rinaldo A |
| Title | Spread and Dynamics of the COVID-19 Epidemic in Italy: Effects of Emergency Containment Measures |
| Year | 2020 |
| Journal | Proceedings of the National Academy of Sciences |
| DOI | 10.1073/pnas.2004978117 |
| Date received | 2020-03-26 (sent for review) |
| Date accepted | 2020-04-06 |
| **Study setting** | |
| Number of populations included | multiple |
| Level of populations included | both national and subnational (Italy and 107 provinces) |
| Geographic areas covered | Europe |
| Number of countries covered | single country (Italy) |
| Study period | end date at peak of wave |
| **Outcome** | |
| Raw outcome | epidemiological population-level outcome (hospitalizations, deaths) |
| Time resolution of raw outcome | daily |

– continued on next page –

| | |
|---|---|
| Computed outcome | none |
| Method to obtain the computed outcome | – not applicable – |
| Data source | data from (sub)national authorities |
| Data availability | data access via source |
| **Intervention** | |
| Terminology for interventions | measures |
| Terminology for the specific type of non-pharmaceutical interventions | containment |
| Exposure types | multiple combinations of interventions (two sets of measures adopted at unspecific dates and to a varying extent in different subnational regions) |
| Types of single interventions | – not applicable – |
| Coding of interventions | not necessary |
| Source of intervention data | government or news websites |
| Availability of data on exposure | raw data documented in the manuscript |
| **Methodological approach** | |
| Empirical approach | parametric |
| Use of exposure variation | both variation over time and between populations |
| Method | mechanistic model (compartmental single-population transmission model) |
| Latent variable functionally linked to intervention | transmission rate |
| Code availability | none |
| **Effect assessment** | |
| Reporting of intervention effect | quantification of change in outcome values |
| Measure of effect | transmission rate |
| Reporting of uncertainty | yes |

– continued on next page –

| | |
|---|---|
| Sensitivity analysis | none |
| Subgroup assessment | none |

| **Ge et al. 2020** | |
|---|---|
| **Study information** | |
| Author | Ge J, He D, Lin Z, Zhu H, Zhuang Z |
| Title | Four-Tier Response System and Spatial Propagation of COVID-19 in China by a Network Model |
| Year | 2020 |
| Journal | Mathematical Biosciences |
| DOI | 10.1016/j.mbs.2020.108484 |
| Date received | 2020-08-16 |
| Date accepted | 2020-09-28 |
| **Study setting** | |
| Number of populations included | multiple |
| Level of populations included | subnational (Wuhan and 18 other cities/regions) |
| Geographic areas covered | Asia |
| Number of countries covered | single country (China) |
| Study period | start and end date span first epidemic wave |
| **Outcome** | |
| Raw outcome | epidemiological population-level outcome (cases) |
| Time resolution of raw outcome | daily |
| Computed outcome | none |

– continued on next page –

| | |
|---|---|
| Method to obtain the computed outcome | – not applicable – |
| Data source | – could not be evaluated – |
| Data availability | data not accessible |
| **Intervention** | |
| Terminology for interventions | measures |
| Terminology for the specific type of non-pharmaceutical interventions | social distancing |
| Exposure types | phases of interventions (different response levels in Chinas response plan) |
| Types of single interventions | – not applicable – |
| Coding of interventions | not necessary |
| Source of intervention data | government or news websites |
| Availability of data on exposure | raw data documented in the manuscript |
| **Methodological approach** | |
| Empirical approach | parametric |
| Use of exposure variation | both variation over time and between populations |
| Method | mechanistic model (compartmental metapopulation transmission model) |
| Latent variable functionally linked to intervention | transmission rate |
| Code availability | none |
| **Effect assessment** | |
| Reporting of intervention effect | quantification of change in outcome values |
| Measure of effect | transmission rate |
| Reporting of uncertainty | yes |
| Sensitivity analysis | none |
| Subgroup assessment | none |

| Ghosal et al. 2020 | |
|---|---|
| **Study information** | |
| Author | Ghosal S, Bhattacharyya R, Majumder M |
| Title | Impact of Complete Lockdown on Total Infection and Death Rates: A Hierarchical Cluster Analysis |
| Year | 2020 |
| Journal | Diabetes & Metabolic Syndrome: Clinical Research & Reviews |
| DOI | 10.1016/j.dsx.2020.05.026 |
| Date received | 2020-05-08 |
| Date accepted | 2020-05-11 |
| **Study setting** | |
| Number of populations included | multiple |
| Level of populations included | national (Spain, Germany, Italy, UK, France, Belgium, Austria, New Zealand, India, Hungary, Poland, Malysia) |
| Geographic areas covered | Asia, Europe, Oceania |
| Number of countries covered | multiple countries |
| Study period | end date could not be evaluated |
| **Outcome** | |
| Raw outcome | epidemiological population-level outcome (growth rate, growth rate: deaths) |
| Time resolution of raw outcome | daily |
| Computed outcome | none |
| Method to obtain the computed outcome | – not applicable – |
| Data source | data from publicly available cross-country selections (Worldometer) |
| Data availability | data access via source |

– continued on next page –

| | |
|---|---|
| **Intervention** | |
| Terminology for interventions | strategies |
| Terminology for the specific type of non-pharmaceutical interventions | preventive |
| Exposure types | one combination of interventions (lockdown) |
| Types of single interventions | – not applicable – |
| Coding of interventions | necessary |
| Source of intervention data | coding done by authors |
| Availability of data on exposure | coded data made available by the authors |
| **Methodological approach** | |
| Empirical approach | descriptive |
| Use of exposure variation | only variation over time for multiple populations |
| Method | description of change over time (description of time course) |
| Code availability | none |
| **Effect assessment** | |
| Reporting of intervention effect | qualitative statement |
| Measure of effect | growth rate, growth rate: deaths |
| Reporting of uncertainty | – not applicable – |
| Sensitivity analysis | none |
| Subgroup assessment | none |

**Gill et al. 2020 (Analysis 1)**

| |
|---|
| **Study information** |

– continued on next page –

| | |
|---|---|
| Author | Gill BS, Jayaraj VJ, Singh S, Ghazali SM, Cheong YL, Iderus NH, Sundram BM, Aris TB, Ibrahim HM, Hong BH, Labadin J |
| Title | Modelling the Effectiveness of Epidemic Control Measures in Preventing the Transmission of COVID-19 in Malaysia |
| Year | 2020 |
| Journal | International Journal of Environmental Research and Public Health |
| DOI | 10.3390/ijerph17155509 |
| Date received | 2020-04-28 |
| Date accepted | 2020-07-02 |
| **Study setting** | |
| Number of populations included | single |
| Level of populations included | national (Malaysia) |
| Geographic areas covered | Asia |
| Number of countries covered | single country (Malaysia) |
| Study period | start and end date span first epidemic wave |
| **Outcome** | |
| Raw outcome | epidemiological population-level outcome (active cases) |
| Time resolution of raw outcome | daily |
| Computed outcome | none |
| Method to obtain the computed outcome | – not applicable – |
| Data source | data from (sub)national authorities |
| Data availability | data access via source |
| **Intervention** | |
| Terminology for interventions | measures |

– continued on next page –

| | |
|---|---|
| Terminology for the specific type of non-pharmaceutical interventions | control |
| Exposure types | one combination of interventions (movement control order decreed by the Malaysian government on March 18) |
| Types of single interventions | – not applicable – |
| Coding of interventions | not necessary |
| Source of intervention data | – could not be evaluated – |
| Availability of data on exposure | raw data documented in the manuscript |
| **Methodological approach** | |
| Empirical approach | counterfactual |
| Use of exposure variation | only variation over time for a single population |
| Method | mechanistic model (compartmental single-population transmission model) |
| Code availability | none |
| **Effect assessment** | |
| Reporting of intervention effect | quantification of change in outcome values |
| Measure of effect | active cases |
| Reporting of uncertainty | no |
| Sensitivity analysis | none |
| Subgroup assessment | none |

**Gill et al. 2020 (Analysis 2)**

| | |
|---|---|
| **Study information** | |
| Author | Gill BS, Jayaraj VJ, Singh S, Ghazali SM, Cheong YL, Iderus NH, Sundram BM, Aris TB, Ibrahim HM, Hong BH, Labadin J |

– continued on next page –

| | |
|---|---|
| Title | Modelling the Effectiveness of Epidemic Control Measures in Preventing the Transmission of COVID-19 in Malaysia |
| Year | 2020 |
| Journal | International Journal of Environmental Research and Public Health |
| DOI | 10.3390/ijerph17155509 |
| Date received | 2020-04-28 |
| Date accepted | 2020-07-02 |
| **Study setting** | |
| Number of populations included | single |
| Level of populations included | national (Malaysia) |
| Geographic areas covered | Asia |
| Number of countries covered | single country (Malaysia) |
| Study period | start and end date span first epidemic wave |
| **Outcome** | |
| Raw outcome | epidemiological population-level outcome (active cases) |
| Time resolution of raw outcome | daily |
| Computed outcome | epidemiological parameter (contact rate) |
| Method to obtain the computed outcome | compartmental transmission model (single-population) |
| Data source | data from (sub)national authorities |
| Data availability | data access via source |
| **Intervention** | |
| Terminology for interventions | measures |
| Terminology for the specific type of non-pharmaceutical interventions | control |

– continued on next page –

| | |
|---|---|
| Exposure types | one combination of interventions (movement control order decreed by the Malaysian government on March 18) |
| Types of single interventions | – not applicable – |
| Coding of interventions | not necessary |
| Source of intervention data | – could not be evaluated – |
| Availability of data on exposure | raw data documented in the manuscript |
| **Methodological approach** | |
| Empirical approach | descriptive |
| Use of exposure variation | only variation over time for a single population |
| Method | description of change over time (comparison of time periods) |
| Code availability | none |
| **Effect assessment** | |
| Reporting of intervention effect | comparison of outcome values |
| Measure of effect | contact rate |
| Reporting of uncertainty | no |
| Sensitivity analysis | none |
| Subgroup assessment | none |

**Gregori et al. 2020**

| | |
|---|---|
| **Study information** | |
| Author | Gregori D, Azzolina D, Lanera C, Prosepe I, Destro N, Lorenzoni G, Berchialla P |
| Title | A First Estimation of the Impact of Public Health Actions against COVID-19 in Veneto (Italy) |
| Year | 2020 |

– continued on next page –

| | |
|---|---|
| Journal | Journal of Epidemiology and Community Health |
| DOI | 10.1136/jech-2020-214209 |
| Date received | 2020-03-28 |
| Date accepted | 2020-04-21 |
| **Study setting** | |
| Number of populations included | single |
| Level of populations included | subnational (Venice (Italy)) |
| Geographic areas covered | Europe |
| Number of countries covered | single country (Italy) |
| Study period | end date at peak of wave |
| **Outcome** | |
| Raw outcome | epidemiological population-level outcome (hospitalizations) |
| Time resolution of raw outcome | daily |
| Computed outcome | none |
| Method to obtain the computed outcome | – not applicable – |
| Data source | data from (sub)national authorities |
| Data availability | data access via source |
| **Intervention** | |
| Terminology for interventions | interventions |
| Terminology for the specific type of non-pharmaceutical interventions | public health |
| Exposure types | one combination of interventions (change point 17 days after the first decree law that established a lockdown) |
| Types of single interventions | – not applicable – |
| Coding of interventions | not necessary |

– continued on next page –

| | |
|---|---|
| Source of intervention data | government or news websites |
| Availability of data on exposure | raw data documented in the manuscript |
| **Methodological approach** | |
| Empirical approach | counterfactual |
| Use of exposure variation | only variation over time for a single population |
| Method | non-mechanistic model (other: generalized additive model) |
| Code availability | none |
| **Effect assessment** | |
| Reporting of intervention effect | quantification of change in outcome values |
| Measure of effect | hospitalizations |
| Reporting of uncertainty | yes |
| Sensitivity analysis | none |
| Subgroup assessment | none |

**Guirao 2020**

| | |
|---|---|
| **Study information** | |
| Author | Guirao A |
| Title | The COVID-19 Outbreak in Spain. A Simple Dynamics Model, Some Lessons, and a Theoretical Framework for Control Response |
| Year | 2020 |
| Journal | Infectious Disease Modelling |
| DOI | 10.1016/j.idm.2020.08.010 |
| Date received | 2020-07-02 |
| Date accepted | 2020-08-21 |

– continued on next page –

| | |
|---|---|
| **Study setting** | |
| Number of populations included | multiple |
| Level of populations included | national (Spain, Italy, Germany) |
| Geographic areas covered | Europe |
| Number of countries covered | multiple countries |
| Study period | start and end date span first epidemic wave |
| **Outcome** | |
| Raw outcome | epidemiological population-level outcome (cases) |
| Time resolution of raw outcome | daily |
| Computed outcome | epidemiological parameter (reproduction number) |
| Method to obtain the computed outcome | compartmental transmission model (single-population) |
| Data source | data from (sub)national authorities (Spain, Italy, Germany), data from publicly available cross-country selections (WHO: Italy, Germany) |
| Data availability | data access via source |
| **Intervention** | |
| Terminology for interventions | measures |
| Terminology for the specific type of non-pharmaceutical interventions | control |
| Exposure types | one combination of interventions (state decrees establishing a system of work-from-home and the mandatory closure of non-emergency businesses and educational institutions throughout the state) |
| Types of single interventions | – not applicable – |
| Coding of interventions | not necessary |
| Source of intervention data | – could not be evaluated – |
| Availability of data on exposure | raw data documented in the manuscript |

– continued on next page –

| | |
|---|---|
| **Methodological approach** | |
| Empirical approach | descriptive |
| Use of exposure variation | only variation over time for multiple populations |
| Method | description of change over time (comparison of time periods) |
| Code availability | none |
| **Effect assessment** | |
| Reporting of intervention effect | comparison of outcome values |
| Measure of effect | reproduction number |
| Reporting of uncertainty | yes |
| Sensitivity analysis | none |
| Subgroup assessment | none |

**Guo & Xiao 2020**

| | |
|---|---|
| **Study information** | |
| Author | Guo Z, Xiao D |
| Title | Analysis and Prediction of the Coronavirus Disease Epidemic in China Based on an Individual-Based Model |
| Year | 2020 |
| Journal | Scientific Reports |
| DOI | 10.1038/s41598-020-76969-4 |
| Date received | 2020-04-06 |
| Date accepted | 2020-10-30 |
| **Study setting** | |
| Number of populations included | single |

– continued on next page –

| | |
|---|---|
| Level of populations included | national (China) |
| Geographic areas covered | Asia |
| Number of countries covered | single country (China) |
| Study period | start and end date span first epidemic wave |
| **Outcome** | |
| Raw outcome | epidemiological population-level outcome (cases) |
| Time resolution of raw outcome | daily |
| Computed outcome | none |
| Method to obtain the computed outcome | – not applicable – |
| Data source | data from (sub)national authorities |
| Data availability | data made available by the authors |
| **Intervention** | |
| Terminology for interventions | measures |
| Terminology for the specific type of non-pharmaceutical interventions | prevention, control |
| Exposure types | one single intervention (traffic control restrictions to and from Hubei) |
| Types of single interventions | internal movement restrictions |
| Coding of interventions | not necessary |
| Source of intervention data | government or news websites |
| Availability of data on exposure | raw data documented in the manuscript |
| **Methodological approach** | |
| Empirical approach | counterfactual |
| Use of exposure variation | only variation over time for a single population |
| Method | mechanistic model (individual-based transmission model) |

– continued on next page –

| | |
|---|---|
| Code availability | none |
| **Effect assessment** | |
| Reporting of intervention effect | quantification of change in outcome values |
| Measure of effect | cases |
| Reporting of uncertainty | no |
| Sensitivity analysis | epidemiological parameters varied |
| Subgroup assessment | none |

**Gupta et al. 2021 (Analysis 1)**

| | |
|---|---|
| **Study information** | |
| Author | Gupta M, Mohanta SS, Rao A, Parameswaran GG, Agarwal M, Arora M, Mazumder A, Lohiya A, Behera P, Bansal A, Kumar R, Meena VP, Tiwari P, Mohan A, Bhatnagar S |
| Title | Transmission Dynamics of the COVID-19 Epidemic in India and Modeling Optimal Lockdown Exit Strategies |
| Year | 2021 |
| Journal | International Journal of Infectious Diseases |
| DOI | 10.1016/j.ijid.2020.11.206 |
| Date received | 2020-07-01 |
| Date accepted | 2020-11-28 |
| **Study setting** | |
| Number of populations included | single |
| Level of populations included | national (India) |
| Geographic areas covered | Asia |

– continued on next page –

| | |
|---|---|
| Number of countries covered | single country (India) |
| Study period | end date in growth phase of wave |
| **Outcome** | |
| Raw outcome | epidemiological population-level outcome (cases) |
| Time resolution of raw outcome | daily |
| Computed outcome | epidemiological parameter (reproduction number) |
| Method to obtain the computed outcome | back-projection (infections), statistical estimation of reproduction number (White et al., Walinga et al., Cori et al.) |
| Data source | data from publicly available open-source projects (covid19India.org) |
| Data availability | data made available by the authors |
| **Intervention** | |
| Terminology for interventions | measures |
| Terminology for the specific type of non-pharmaceutical interventions | control |
| Exposure types | one combination of interventions (Indian National Lockdown) |
| Types of single interventions | – not applicable – |
| Coding of interventions | not necessary |
| Source of intervention data | – could not be evaluated – |
| Availability of data on exposure | raw data documented in the manuscript |
| **Methodological approach** | |
| Empirical approach | descriptive |
| Use of exposure variation | only variation over time for a single population |
| Method | description of change over time (description of time course) |
| Code availability | publicly available |
| **Effect assessment** | |
| Reporting of intervention effect | comparison of outcome values |

– continued on next page –

| | |
|---|---|
| Measure of effect | reproduction number |
| Reporting of uncertainty | yes |
| Sensitivity analysis | none |
| Subgroup assessment | none |

**Gupta et al. 2021 (Analysis 2)**

| | |
|---|---|
| **Study information** | |
| Author | Gupta M, Mohanta SS, Rao A, Parameswaran GG, Agarwal M, Arora M, Mazumder A, Lohiya A, Behera P, Bansal A, Kumar R, Meena VP, Tiwari P, Mohan A, Bhatnagar S |
| Title | Transmission Dynamics of the COVID-19 Epidemic in India and Modeling Optimal Lockdown Exit Strategies |
| Year | 2021 |
| Journal | International Journal of Infectious Diseases |
| DOI | 10.1016/j.ijid.2020.11.206 |
| Date received | 2020-07-01 |
| Date accepted | 2020-11-28 |
| **Study setting** | |
| Number of populations included | single |
| Level of populations included | national (India) |
| Geographic areas covered | Asia |
| Number of countries covered | single country (India) |
| Study period | start and end date span first epidemic wave: behavioral |
| **Outcome** | |
| Raw outcome | behavioral outcome (mobility: time spent in various locations) |

– continued on next page –

| | |
|---|---|
| Time resolution of raw outcome | daily |
| Computed outcome | none |
| Method to obtain the computed outcome | – not applicable – |
| Data source | mobility data from corporate organizations (Google) |
| Data availability | data made available by the authors |
| **Intervention** | |
| Terminology for interventions | measures |
| Terminology for the specific type of non-pharmaceutical interventions | control |
| Exposure types | one combination of interventions (Indian National Lockdown) |
| Types of single interventions | – not applicable – |
| Coding of interventions | not necessary |
| Source of intervention data | – could not be evaluated – |
| Availability of data on exposure | raw data documented in the manuscript |
| **Methodological approach** | |
| Empirical approach | descriptive |
| Use of exposure variation | only variation over time for a single population |
| Method | description of change over time (description of time course) |
| Code availability | publicly available |
| **Effect assessment** | |
| Reporting of intervention effect | qualitative statement |
| Measure of effect | mobility: time spent in various locations |
| Reporting of uncertainty | – not applicable – |
| Sensitivity analysis | none |

– continued on next page –

| | |
|---|---|
| Subgroup assessment | none |

| **Hao et al. 2020** | |
|---|---|
| **Study information** | |
| Author | Hao X, Cheng S, Wu D, Wu T, Lin X, Wang C |
| Title | Reconstruction of the Full Transmission Dynamics of COVID-19 in Wuhan |
| Year | 2020 |
| Journal | Nature |
| DOI | 10.1038/s41586-020-2554-8 |
| Date received | 2020-04-14 |
| Date accepted | 2020-07-10 |
| **Study setting** | |
| Number of populations included | single |
| Level of populations included | subnational (Wuhan (China)) |
| Geographic areas covered | Asia |
| Number of countries covered | single country (China) |
| Study period | start and end date span first epidemic wave |
| **Outcome** | |
| Raw outcome | epidemiological individual-level outcome (individual cases) |
| Time resolution of raw outcome | daily |
| Computed outcome | epidemiological parameter (reproduction number) |
| Method to obtain the computed outcome | compartmental transmission model (single-population) |
| Data source | data from (sub)national authorities |

– continued on next page –

| | |
|---|---|
| Data availability | data made available by the authors |
| **Intervention** | |
| Terminology for interventions | interventions |
| Terminology for the specific type of non-pharmaceutical interventions | non-pharmaceutical |
| Exposure types | multiple combinations of interventions (Wuhan City Lockdown, centralized isolation and quarantine, community screening) |
| Types of single interventions | – not applicable – |
| Coding of interventions | not necessary |
| Source of intervention data | another study (Pan et al.) |
| Availability of data on exposure | raw data documented in the manuscript |
| **Methodological approach** | |
| Empirical approach | descriptive |
| Use of exposure variation | only variation over time for a single population |
| Method | description of change over time (comparison of time periods) |
| Code availability | publicly available |
| **Effect assessment** | |
| Reporting of intervention effect | quantification of change in outcome values |
| Measure of effect | reproduction number, infections |
| Reporting of uncertainty | yes |
| Sensitivity analysis | different or modified outcome used (smoothing of outliers), epidemiological parameters varied |
| Subgroup assessment | none |

| Haug et al. 2020 | |
|---|---|
| **Study information** | |
| Author | Haug N, Geyrhofer L, Londei A, Dervic E, Desvars-Larrive A, Loreto V, Pinior B, Thurner S, Klimek P |
| Title | Ranking the Effectiveness of Worldwide COVID-19 Government Interventions |
| Year | 2020 |
| Journal | Nature Human Behaviour |
| DOI | 10.1038/s41562-020-01009-0 |
| Date received | 2020-10-15 |
| Date accepted | 2020-10-28 |
| **Study setting** | |
| Number of populations included | multiple |
| Level of populations included | both national and subnational (79 territories around the globe) |
| Geographic areas covered | Europe, Middle East and Africa, Asia, Oceania, Central and South America, North America |
| Number of countries covered | multiple countries |
| Study period | same end date for several populations with diverse epidemic trajectories |
| **Outcome** | |
| Raw outcome | epidemiological population-level outcome (cases) |
| Time resolution of raw outcome | daily |
| Computed outcome | epidemiological parameter (reproduction number) |
| Method to obtain the computed outcome | statistical estimation of reproduction number (Cori et al.) |
| Data source | data from publicly available cross-country selections (JHU) |
| Data availability | data made available by the authors |
| **Intervention** | |

– continued on next page –

| | |
|---|---|
| Terminology for interventions | interventions |
| Terminology for the specific type of non-pharmaceutical interventions | non-pharmaceutical |
| Exposure types | multiple separate interventions (large list of hierarchically coded NPIs) |
| Types of single interventions | stay-at-home order, school closure, workplace closure, venue closure, declaration of a state of emergency, bans of large gatherings, bans of small gatherings, internal movement restrictions, public transport closure, international travel restrictions, mask mandate |
| Coding of interventions | necessary |
| Source of intervention data | use of externally coded data (Complexity Science Hub COVID-19 Control Strategies List) |
| Availability of data on exposure | coded data made available by the authors |
| **Methodological approach** | |
| Empirical approach | parametric |
| Use of exposure variation | both variation over time and between populations |
| Method | non-mechanistic model (other: several machine learning models of different complexity) |
| Code availability | publicly available |
| **Effect assessment** | |
| Reporting of intervention effect | quantification of change in outcome values |
| Measure of effect | reproduction number |
| Reporting of uncertainty | yes |
| Sensitivity analysis | different coding of interventions used (validation with other NPI datasets, implementation date varied) |
| Subgroup assessment | none |

| He et al. 2020a | |
|---|---|
| **Study information** | |
| Author | He J, Chen G, Jiang Y, Jin R, Shortridge A, Agusti S, He M, Wu J, Duarte CM, Christakos G |
| Title | Comparative Infection Modeling and Control of COVID-19 Transmission Patterns in China, South Korea, Italy and Iran |
| Year | 2020 |
| Journal | Science of The Total Environment |
| DOI | 10.1016/j.scitotenv.2020.141447 |
| Date received | 2020-07-05 |
| Date accepted | 2020-08-01 |
| **Study setting** | |
| Number of populations included | multiple |
| Level of populations included | both national and subnational (Chinese regions (Zhejiang, Guangdong, Xinjiang), South Korea, Italy, Iran) |
| Geographic areas covered | Asia, Europe, Middle East and Africa |
| Number of countries covered | multiple countries |
| Study period | start and end date span first epidemic wave |
| **Outcome** | |
| Raw outcome | epidemiological population-level outcome (cases, deaths, recovered cases) |
| Time resolution of raw outcome | daily |
| Computed outcome | none |
| Method to obtain the computed outcome | – not applicable – |
| Data source | data from (sub)national authorities |
| Data availability | data access via source |
| **Intervention** | |

– continued on next page –

| | |
|---|---|
| Terminology for interventions | measures |
| Terminology for the specific type of non-pharmaceutical interventions | control |
| Exposure types | all interventions together (timeline of measures) |
| Types of single interventions | – not applicable – |
| Coding of interventions | – not applicable – |
| Source of intervention data | – not applicable – |
| Availability of data on exposure | – not applicable – |
| **Methodological approach** | |
| Empirical approach | counterfactual |
| Use of exposure variation | only variation over time for multiple populations |
| Method | mechanistic model (compartmental single-population transmission model) |
| Code availability | none |
| **Effect assessment** | |
| Reporting of intervention effect | quantification of change in outcome values |
| Measure of effect | cases |
| Reporting of uncertainty | no |
| Sensitivity analysis | none |
| Subgroup assessment | none |

**He et al. 2020b**

| | |
|---|---|
| **Study information** | |
| Author | He S, Tang AS, And LR |

– continued on next page –

| | |
|---|---|
| Title | A Discrete Stochastic Model of the COVID-19 Outbreak: Forecast and Control |
| Year | 2020 |
| Journal | Mathematical Biosciences and Engineering |
| DOI | 10.3934/mbe.2020153 |
| Date received | 2020-02-22 |
| Date accepted | 2020-03-12 |
| **Study setting** | |
| Number of populations included | single |
| Level of populations included | national (China) |
| Geographic areas covered | Asia |
| Number of countries covered | single country (China) |
| Study period | start and end date span first epidemic wave |
| **Outcome** | |
| Raw outcome | epidemiological population-level outcome (cases, deaths, recovered cases) |
| Time resolution of raw outcome | daily |
| Computed outcome | epidemiological parameter (reproduction number, contact rate) |
| Method to obtain the computed outcome | compartmental transmission model (single-population) |
| Data source | data from (sub)national authorities |
| Data availability | data access via source |
| **Intervention** | |
| Terminology for interventions | measures |
| Terminology for the specific type of non-pharmaceutical interventions | control |
| Exposure types | all interventions together |

– continued on next page –

| | |
|---|---|
| Types of single interventions | – not applicable – |
| Coding of interventions | – not applicable – |
| Source of intervention data | – not applicable – |
| Availability of data on exposure | – not applicable – |
| **Methodological approach** | |
| Empirical approach | descriptive |
| Use of exposure variation | only variation over time for a single population |
| Method | description of change over time (description of time course) |
| Code availability | none |
| **Effect assessment** | |
| Reporting of intervention effect | qualitative statement |
| Measure of effect | reproduction number, contact rate |
| Reporting of uncertainty | – not applicable – |
| Sensitivity analysis | none |
| Subgroup assessment | none |

**Henríquez et al. 2020 (Analysis 1)**

| | |
|---|---|
| **Study information** | |
| Author | Henríquez J, Gonzalo-Almorox E, García-Goñi M, Paolucci F |
| Title | The First Months of the COVID-19 Pandemic in Spain |
| Year | 2020 |
| Journal | Health Policy and Technology |
| DOI | 10.1016/j.hlpt.2020.08.013 |

– continued on next page –

| | |
|---|---|
| Date received | – could not be evaluated – |
| Date accepted | 2020-08-27 (published) |
| **Study setting** | |
| Number of populations included | single |
| Level of populations included | national (Spain) |
| Geographic areas covered | Europe |
| Number of countries covered | single country (Spain) |
| Study period | start and end date span first epidemic wave |
| **Outcome** | |
| Raw outcome | epidemiological population-level outcome (cases) |
| Time resolution of raw outcome | daily |
| Computed outcome | none |
| Method to obtain the computed outcome | – not applicable – |
| Data source | data from (sub)national authorities |
| Data availability | data access via source |
| **Intervention** | |
| Terminology for interventions | measures |
| Terminology for the specific type of non-pharmaceutical interventions | containment, mitigation |
| Exposure types | stringency index (OxCGRT Stringency Index) |
| Types of single interventions | – not applicable – |
| Coding of interventions | necessary |
| Source of intervention data | use of externally coded data (OxCGRT) |
| Availability of data on exposure | access to externally coded data via source |
| **Methodological approach** | |

– continued on next page –

| | |
|---|---|
| Empirical approach | descriptive |
| Use of exposure variation | only variation over time for a single population |
| Method | description of change over time (description of time course) |
| Code availability | none |
| **Effect assessment** | |
| Reporting of intervention effect | qualitative statement |
| Measure of effect | cases |
| Reporting of uncertainty | – not applicable – |
| Sensitivity analysis | none |
| Subgroup assessment | none |

**Henríquez et al. 2020 (Analysis 2)**

| | |
|---|---|
| **Study information** | |
| Author | Henríquez J, Gonzalo-Almorox E, García-Goñi M, Paolucci F |
| Title | The First Months of the COVID-19 Pandemic in Spain |
| Year | 2020 |
| Journal | Health Policy and Technology |
| DOI | 10.1016/j.hlpt.2020.08.013 |
| Date received | – could not be evaluated – |
| Date accepted | 2020-08-27 (published) |
| **Study setting** | |
| Number of populations included | multiple |
| Level of populations included | both national and subnational (Spain and all its autonomous communities) |
| Geographic areas covered | Europe |

– continued on next page –

| | |
|---|---|
| Number of countries covered | single country (Spain) |
| Study period | start and end date span first epidemic wave: behavioral |
| **Outcome** | |
| Raw outcome | behavioral outcome (mobility) |
| Time resolution of raw outcome | daily |
| Computed outcome | none |
| Method to obtain the computed outcome | – not applicable – |
| Data source | – could not be evaluated – |
| Data availability | data not accessible |
| **Intervention** | |
| Terminology for interventions | measures |
| Terminology for the specific type of non-pharmaceutical interventions | containment, mitigation |
| Exposure types | all interventions together |
| Types of single interventions | – not applicable – |
| Coding of interventions | – not applicable – |
| Source of intervention data | – not applicable – |
| Availability of data on exposure | – not applicable – |
| **Methodological approach** | |
| Empirical approach | descriptive |
| Use of exposure variation | only variation over time for a single population |
| Method | description of change over time (description of time course) |
| Code availability | none |
| **Effect assessment** | |
| Reporting of intervention effect | qualitative statement |

– continued on next page –

| | |
|---|---|
| Measure of effect | mobility |
| Reporting of uncertainty | – not applicable – |
| Sensitivity analysis | none |
| Subgroup assessment | none |

**Holtz et al. 2020**

| | |
|---|---|
| **Study information** | |
| Author | Holtz D, Zhao M, Benzell SG, Cao CY, Rahimian MA, Yang J, Allen J, Collis A, Moehring A, Sowrirajan T, Ghosh D, Zhang Y, Dhillon PS, Nicolaides C, Eckles D, Aral S |
| Title | Interdependence and the Cost of Uncoordinated Responses to COVID-19 |
| Year | 2020 |
| Journal | Proceedings of the National Academy of Sciences |
| DOI | 10.1073/pnas.2009522117 |
| Date received | 2020-05-12 |
| Date accepted | 2020-07-10 |
| **Study setting** | |
| Number of populations included | multiple |
| Level of populations included | both national and subnational (2,502 US counties) |
| Geographic areas covered | North America |
| Number of countries covered | single country (US) |
| Study period | start and end date span first epidemic wave: behavioral |
| **Outcome** | |
| Raw outcome | behavioral outcome (mobility: fraction of mobile devices leaving home and mean number of locations visited per device) |

– continued on next page –

| | |
|---|---|
| Time resolution of raw outcome | daily |
| Computed outcome | none |
| Method to obtain the computed outcome | – not applicable – |
| Data source | mobility data from corporate organizations (Safegraph (fraction of mobile devices leaving home), Facebook (mean number of locations visited per device)) |
| Data availability | data made available by the authors |
| **Intervention** | |
| Terminology for interventions | measures |
| Terminology for the specific type of non-pharmaceutical interventions | social distancing |
| Exposure types | multiple separate interventions (stay-at-home order, venue closure) |
| Types of single interventions | stay-at-home order, venue closure |
| Coding of interventions | necessary |
| Source of intervention data | use of externally coded data (same as in Killeen et al.) |
| Availability of data on exposure | access to externally coded data via source |
| **Methodological approach** | |
| Empirical approach | parametric |
| Use of exposure variation | both variation over time and between populations |
| Method | non-mechanistic model (generalized linear model) |
| Code availability | publicly available |
| **Effect assessment** | |
| Reporting of intervention effect | quantification of change in outcome values |
| Measure of effect | mobility: fraction of mobile devices leaving home and mean number of locations visited per device |

– continued on next page –

| | |
|---|---|
| Reporting of uncertainty | yes |
| Sensitivity analysis | different coding of interventions used (policy effects considered with lags and leads), model specification varied, same analysis with (sub)population(s) excluded |
| Subgroup assessment | none |

| **Hsiang et al. 2020** | |
|---|---|
| **Study information** | |
| Author | Hsiang S, Allen D, Annan-Phan S, Bell K, Bolliger I, Chong T, Druckenmiller H, Huang LY, Hultgren A, Krasovich E, Lau P, Lee J, Rolf E, Tseng J, Wu T |
| Title | The Effect of Large-Scale Anti-Contagion Policies on the COVID-19 Pandemic |
| Year | 2020 |
| Journal | Nature |
| DOI | 10.1038/s41586-020-2404-8 |
| Date received | 2020-03-22 |
| Date accepted | 2020-05-26 |
| **Study setting** | |
| Number of populations included | multiple |
| Level of populations included | both national and subnational (China, South Korea, Italy, Iran, France, the US and the subnational regions of these countries) |
| Geographic areas covered | Asia, Europe, Middle East and Africa, North America |
| Number of countries covered | multiple countries |
| Study period | same end date for several populations with diverse epidemic trajectories |

– continued on next page –

| | |
|---|---|
| **Outcome** | |
| Raw outcome | epidemiological population-level outcome (cases, active cases, deaths, recovered cases) |
| Time resolution of raw outcome | daily |
| Computed outcome | measure of epidemic trend (growth rate, growth rate: active cases) |
| Method to obtain the computed outcome | simple computation |
| Data source | data from (sub)national authorities (China, South Korea, Iran, Italy, France), public (USAFacts: US) |
| Data availability | data made available by the authors |
| **Intervention** | |
| Terminology for interventions | policies |
| Terminology for the specific type of non-pharmaceutical interventions | anti-contagion |
| Exposure types | multiple separate interventions (individual and aggregated country-specific measures (includes travel restrictions school closure, business closure, religious closure, work bans, event bans gathering bans, home isolation, emergency declaration, etc)) |
| Types of single interventions | international travel restrictions, workplace closure, venue closure, bans of large gatherings, stay-at-home order, declaration of a state of emergency |
| Coding of interventions | necessary |
| Source of intervention data | coding done by authors |
| Availability of data on exposure | coded data made available by the authors |
| **Methodological approach** | |
| Empirical approach | parametric |
| Use of exposure variation | both variation over time and between populations |
| Method | non-mechanistic model (generalized linear model) |

– continued on next page –

| | |
|---|---|
| Code availability | publicly available |
| **Effect assessment** | |
| Reporting of intervention effect | quantification of change in outcome values |
| Measure of effect | growth rate, growth rate: active cases |
| Reporting of uncertainty | yes |
| Sensitivity analysis | different or modified outcome used (bias related to underreporting of cases estimated), model specification varied, epidemiological parameters varied |
| Subgroup assessment | none |

**Huang et al. 2020**

| | |
|---|---|
| **Study information** | |
| Author | Huang X, Li Z, Jiang Y, Li X, Porter D |
| Title | Twitter Reveals Human Mobility Dynamics during the COVID-19 Pandemic |
| Year | 2020 |
| Journal | PLOS ONE |
| DOI | 10.1371/journal.pone.0241957 |
| Date received | 2020-06-24 |
| Date accepted | 2020-10-23 |
| **Study setting** | |
| Number of populations included | multiple |
| Level of populations included | both national and subnational (20 selected countries around the world, US states) |
| Geographic areas covered | Europe, Middle East and Africa, Asia, Oceania, Central and South America, North America |
| Number of countries covered | multiple countries |

– continued on next page –

| | |
|---|---|
| Study period | start and end date span first epidemic wave: behavioral |
| **Outcome** | |
| Raw outcome | behavioral outcome (mobility: index based on distances) |
| Time resolution of raw outcome | daily |
| Computed outcome | none |
| Method to obtain the computed outcome | – not applicable – |
| Data source | mobility data from corporate organizations (Twitter) |
| Data availability | data made available by the authors |
| **Intervention** | |
| Terminology for interventions | measures |
| Terminology for the specific type of non-pharmaceutical interventions | mitigation |
| Exposure types | all interventions together |
| Types of single interventions | – not applicable – |
| Coding of interventions | – not applicable – |
| Source of intervention data | – not applicable – |
| Availability of data on exposure | – not applicable – |
| **Methodological approach** | |
| Empirical approach | descriptive |
| Use of exposure variation | only variation over time for multiple populations |
| Method | description of change over time (description of time course) |
| Code availability | none |
| **Effect assessment** | |
| Reporting of intervention effect | qualitative statement |
| Measure of effect | mobility: index based on distances |

– continued on next page –

| | |
|---|---|
| Reporting of uncertainty | – not applicable – |
| Sensitivity analysis | none |
| Subgroup assessment | none |

| **Huber & Langen 2020 (Analysis 1)** | |
|---|---|
| **Study information** | |
| Author | Huber M, Langen H |
| Title | Timing Matters: The Impact of Response Measures on COVID-19-related Hospitalization and Death Rates in Germany and Switzerland |
| Year | 2020 |
| Journal | Swiss Journal of Economics and Statistics |
| DOI | 10.1186/s41937-020-00054-w |
| Date received | 2020-06-18 |
| Date accepted | 2020-07-27 |
| **Study setting** | |
| Number of populations included | multiple |
| Level of populations included | both national and subnational (Germany, Switzerland and its states/counties/cantons) |
| Geographic areas covered | Europe |
| Number of countries covered | multiple countries |
| Study period | start and end date span first epidemic wave |
| **Outcome** | |
| Raw outcome | epidemiological population-level outcome (hospitalizations, deaths) |

– continued on next page –

| | |
|---|---|
| Time resolution of raw outcome | daily |
| Computed outcome | none |
| Method to obtain the computed outcome | – not applicable – |
| Data source | data from (sub)national authorities |
| Data availability | data access via source |
| **Intervention** | |
| Terminology for interventions | measures |
| Terminology for the specific type of non-pharmaceutical interventions | response |
| Exposure types | start of interventions (start date of lockdown measures (including the closure of non-essential shops, schools, childcare centers, cafes, bars, and restaurants, bans on gatherings)) |
| Types of single interventions | – not applicable – |
| Coding of interventions | not necessary |
| Source of intervention data | government or news websites |
| Availability of data on exposure | raw data documented in the manuscript |
| **Methodological approach** | |
| Empirical approach | parametric |
| Use of exposure variation | only variation between populations |
| Method | non-mechanistic model (generalized linear model) |
| Code availability | none |
| **Effect assessment** | |
| Reporting of intervention effect | quantification of change in outcome values |
| Measure of effect | hospitalizations, deaths |
| Reporting of uncertainty | yes |

– continued on next page –

| | |
|---|---|
| Sensitivity analysis | model specification varied |
| Subgroup assessment | none |

**Huber & Langen 2020 (Analysis 2)**

| | |
|---|---|
| **Study information** | |
| Author | Huber M, Langen H |
| Title | Timing Matters: The Impact of Response Measures on COVID-19-related Hospitalization and Death Rates in Germany and Switzerland |
| Year | 2020 |
| Journal | Swiss Journal of Economics and Statistics |
| DOI | 10.1186/s41937-020-00054-w |
| Date received | 2020-06-18 |
| Date accepted | 2020-07-27 |
| **Study setting** | |
| Number of populations included | multiple |
| Level of populations included | subnational (2 cantons in Switzerland) |
| Geographic areas covered | Europe |
| Number of countries covered | single country (Switzerland) |
| Study period | start and end date span first epidemic wave |
| **Outcome** | |
| Raw outcome | epidemiological population-level outcome (hospitalizations, deaths) |
| Time resolution of raw outcome | daily |
| Computed outcome | none |
| Method to obtain the computed outcome | – not applicable – |

– continued on next page –

| | |
|---|---|
| Data source | data from (sub)national authorities |
| Data availability | data access via source |
| **Intervention** | |
| Terminology for interventions | measures |
| Terminology for the specific type of non-pharmaceutical interventions | response |
| Exposure types | start of interventions (start date of lockdown measures (including the closure of non-essential shops, schools, childcare centers, cafes, bars, and restaurants, bans on gatherings)) |
| Types of single interventions | – not applicable – |
| Coding of interventions | not necessary |
| Source of intervention data | government or news websites |
| Availability of data on exposure | raw data documented in the manuscript |
| **Methodological approach** | |
| Empirical approach | counterfactual |
| Use of exposure variation | only variation over time for multiple populations |
| Method | synthetic controls |
| Code availability | none |
| **Effect assessment** | |
| Reporting of intervention effect | quantification of change in outcome values |
| Measure of effect | hospitalizations, deaths |
| Reporting of uncertainty | no |
| Sensitivity analysis | none |
| Subgroup assessment | none |

| Huber & Langen 2020 (Analysis 3) | |
|---|---|
| **Study information** | |
| Author | Huber M, Langen H |
| Title | Timing Matters: The Impact of Response Measures on COVID-19-related Hospitalization and Death Rates in Germany and Switzerland |
| Year | 2020 |
| Journal | Swiss Journal of Economics and Statistics |
| DOI | 10.1186/s41937-020-00054-w |
| Date received | 2020-06-18 |
| Date accepted | 2020-07-27 |
| **Study setting** | |
| Number of populations included | multiple |
| Level of populations included | both national and subnational (Germany and its states) |
| Geographic areas covered | Europe |
| Number of countries covered | single country (Germany) |
| Study period | start and end date span first epidemic wave |
| **Outcome** | |
| Raw outcome | epidemiological population-level outcome (hospitalizations, deaths) |
| Time resolution of raw outcome | daily |
| Computed outcome | none |
| Method to obtain the computed outcome | – not applicable – |
| Data source | data from (sub)national authorities |
| Data availability | data access via source |
| **Intervention** | |
| Terminology for interventions | measures |

– continued on next page –

| **Huber & Langen 2020 (Analysis 3)** – continued from previous page | |
|---|---|
| Terminology for the specific type of non-pharmaceutical interventions | response |
| Exposure types | one single intervention (curfew) |
| Types of single interventions | stay-at-home order |
| Coding of interventions | not necessary |
| Source of intervention data | government or news websites |
| Availability of data on exposure | raw data documented in the manuscript |
| **Methodological approach** | |
| Empirical approach | parametric |
| Use of exposure variation | only variation between populations |
| Method | non-mechanistic model (generalized linear model) |
| Code availability | none |
| **Effect assessment** | |
| Reporting of intervention effect | quantification of change in outcome values |
| Measure of effect | hospitalizations, deaths |
| Reporting of uncertainty | yes |
| Sensitivity analysis | none |
| Subgroup assessment | none |

| **Hyafil & Moriña 2021** | |
|---|---|
| **Study information** | |
| Author | Hyafil A, Moriña D |
| Title | Analysis of the Impact of Lockdown on the Reproduction Number of the SARS-Cov-2 in Spain |

– continued on next page –

| | |
|---|---|
| Year | 2021 |
| Journal | Gaceta Sanitaria |
| DOI | 10.1016/j.gaceta.2020.05.003 |
| Date received | 2020-04-18 |
| Date accepted | 2020-05-06 |
| **Study setting** | |
| Number of populations included | single |
| Level of populations included | national (Spain) |
| Geographic areas covered | Europe |
| Number of countries covered | single country (Spain) |
| Study period | start and end date span first epidemic wave |
| **Outcome** | |
| Raw outcome | epidemiological population-level outcome (hospitalizations) |
| Time resolution of raw outcome | daily |
| Computed outcome | epidemiological parameter (reproduction number) |
| Method to obtain the computed outcome | compartmental transmission model (single-population) |
| Data source | data from (sub)national authorities |
| Data availability | data access via source |
| **Intervention** | |
| Terminology for interventions | interventions |
| Terminology for the specific type of non-pharmaceutical interventions | non-pharmaceutical |
| Exposure types | multiple separate interventions (emergency declaration, stay-at-home order) |
| Types of single interventions | declaration of a state of emergency, stay-at-home order |
| Coding of interventions | not necessary |

– continued on next page –

| | |
|---|---|
| Source of intervention data | – could not be evaluated – |
| Availability of data on exposure | raw data documented in the manuscript |
| **Methodological approach** | |
| Empirical approach | descriptive |
| Use of exposure variation | only variation over time for a single population |
| Method | description of change over time (comparison of time periods) |
| Code availability | none |
| **Effect assessment** | |
| Reporting of intervention effect | comparison of outcome values |
| Measure of effect | reproduction number |
| Reporting of uncertainty | yes |
| Sensitivity analysis | different or modified outcome used (use cases and deaths instead of hospitalizations as raw different or modified outcome used), epidemiological parameters varied |
| Subgroup assessment | none |

**Islam et al. 2020**

| | |
|---|---|
| **Study information** | |
| Author | Islam N, Sharp SJ, Chowell G, Shabnam S, Kawachi I, Lacey B, Massaro JM, D'Agostino RB, White M |
| Title | Physical Distancing Interventions and Incidence of Coronavirus Disease 2019: Natural Experiment in 149 Countries |
| Year | 2020 |
| Journal | BMJ |
| DOI | 10.1136/bmj.m2743 |

– continued on next page –

| | |
|---|---|
| Date received | – could not be evaluated – |
| Date accepted | 2020-07-08 |
| **Study setting** | |
| Number of populations included | multiple |
| Level of populations included | national (149 countries and regions) |
| Geographic areas covered | Europe, Middle East and Africa, Asia, Oceania, Central and South America, North America |
| Number of countries covered | multiple countries |
| Study period | end date could not be evaluated |
| **Outcome** | |
| Raw outcome | epidemiological population-level outcome (cases) |
| Time resolution of raw outcome | daily |
| Computed outcome | none |
| Method to obtain the computed outcome | – not applicable – |
| Data source | data from publicly available cross-country selections (ECDC) |
| Data availability | data made available by the authors |
| **Intervention** | |
| Terminology for interventions | measures |
| Terminology for the specific type of non-pharmaceutical interventions | physical distancing |
| Exposure types | multiple separate interventions (school closure, bans of large gatherings, workplace closure, stay-at-home order, public transport closure) |
| Types of single interventions | school closure, bans of large gatherings, workplace closure, stay-at-home order, public transport closure |
| Coding of interventions | necessary |

– continued on next page –

| | |
|---|---|
| Source of intervention data | use of externally coded data (OxCGRT) |
| Availability of data on exposure | coded data made available by the authors |
| **Methodological approach** | |
| Empirical approach | parametric |
| Use of exposure variation | only variation over time for multiple populations |
| Method | non-mechanistic model (generalized linear model) |
| Code availability | none |
| **Effect assessment** | |
| Reporting of intervention effect | quantification of change in outcome values |
| Measure of effect | incidence rate ratio |
| Reporting of uncertainty | yes |
| Sensitivity analysis | epidemiological parameters varied, model specification varied, same analysis with (sub)population(s) excluded |
| Subgroup assessment | none |

**Jardine et al. 2020**

| | |
|---|---|
| **Study information** | |
| Author | Jardine R, Wright J, Samad Z, Bhutta ZA |
| Title | Analysis of COVID-19 Burden, Epidemiology and Mitigation Strategies in Muslim Majority Countries |
| Year | 2020 |
| Journal | Eastern Mediterranean Health Journal |
| DOI | 10.26719/emhj.20.120 |
| Date received | 2020-06-26 |

– continued on next page –

| | |
|---|---|
| Date accepted | 2020-09-14 |
| **Study setting** | |
| Number of populations included | multiple |
| Level of populations included | national (44 countries with Muslim majority) |
| Geographic areas covered | Middle East and Africa |
| Number of countries covered | multiple countries |
| Study period | same end date for several populations with diverse epidemic trajectories |
| **Outcome** | |
| Raw outcome | epidemiological population-level outcome (cases) |
| Time resolution of raw outcome | daily |
| Computed outcome | measure of epidemic trend (doubling time) |
| Method to obtain the computed outcome | simple computation |
| Data source | data from publicly available cross-country selections (JHU) |
| Data availability | data access via source |
| **Intervention** | |
| Terminology for interventions | interventions |
| Terminology for the specific type of non-pharmaceutical interventions | non-pharmaceutical |
| Exposure types | one combination of interventions (strict movement restrictions (visa restrictions, domestic travel restrictions and curfew) ) |
| Types of single interventions | – not applicable – |
| Coding of interventions | necessary |
| Source of intervention data | use of externally coded data (ACAPS Government Measures Dataset) |
| Availability of data on exposure | access to externally coded data via source |
| **Methodological approach** | |

– continued on next page –

| | |
|---|---|
| Empirical approach | descriptive |
| Use of exposure variation | only variation between populations |
| Method | comparison of populations |
| Code availability | none |
| **Effect assessment** | |
| Reporting of intervention effect | quantification of change in outcome values |
| Measure of effect | cases, doubling time |
| Reporting of uncertainty | yes |
| Sensitivity analysis | none |
| Subgroup assessment | none |

| **Jarvis et al. 2020** | |
|---|---|
| **Study information** | |
| Author | Jarvis CI, Van Zandvoort AK, Gimma A, Prem K, Klepac P, Rubin GJ, Edmunds WJ |
| Title | Quantifying the Impact of Physical Distance Measures on the Transmission of COVID-19 in the UK |
| Year | 2020 |
| Journal | BMC Medicine |
| DOI | 10.1186/s12916-020-01597-8 |
| Date received | 2020-04-03 |
| Date accepted | 2020-04-17 |
| **Study setting** | |
| Number of populations included | single |

– continued on next page –

| | |
|---|---|
| Level of populations included | national (UK) |
| Geographic areas covered | Europe |
| Number of countries covered | single country (UK) |
| Study period | late start |
| **Outcome** | |
| Raw outcome | behavioral outcome (survey: number of contacts & adherence to NPIs) |
| Time resolution of raw outcome | – not applicable – |
| Computed outcome | epidemiological parameter (reproduction number) |
| Method to obtain the computed outcome | statistical estimation of reproduction number from individual data (Diekmann et al.) |
| Data source | data collected by authors |
| Data availability | data made available by the authors |
| **Intervention** | |
| Terminology for interventions | measures |
| Terminology for the specific type of non-pharmaceutical interventions | physical distancing |
| Exposure types | one combination of interventions (lockdown on March 23) |
| Types of single interventions | – not applicable – |
| Coding of interventions | not necessary |
| Source of intervention data | – could not be evaluated – |
| Availability of data on exposure | raw data documented in the manuscript |
| **Methodological approach** | |
| Empirical approach | descriptive |
| Use of exposure variation | only variation over time for a single population |
| Method | description of change over time (comparison of time periods) |

– continued on next page –

| | |
|---|---|
| Code availability | publicly available |
| **Effect assessment** | |
| Reporting of intervention effect | quantification of change in outcome values |
| Measure of effect | number of contacts, adherence to NPIs, reproduction number |
| Reporting of uncertainty | yes |
| Sensitivity analysis | epidemiological parameters varied |
| Subgroup assessment | socioeconomic indicators (age, gender, household size) |

**Jefferies et al. 2020**

| | |
|---|---|
| **Study information** | |
| Author | Jefferies S, French N, Gilkison C, Graham G, Hope V, Marshall J, McElnay C, McNeill A, Muellner P, Paine S, Prasad N, Scott J, Sherwood J, Yang L, Priest P |
| Title | COVID-19 in New Zealand and the Impact of the National Response: A Descriptive Epidemiological Study |
| Year | 2020 |
| Journal | The Lancet Public Health |
| DOI | 10.1016/s2468-2667(20)30225-5 |
| Date received | – could not be evaluated – |
| Date accepted | 2020-10-13 (published) |
| **Study setting** | |
| Number of populations included | single |
| Level of populations included | national (New Zealand) |
| Geographic areas covered | Oceania |
| Number of countries covered | single country (New Zealand) |

– continued on next page –

| | |
|---|---|
| Study period | start and end date span first epidemic wave |
| **Outcome** | |
| Raw outcome | epidemiological individual-level outcome (individual cases) |
| Time resolution of raw outcome | daily |
| Computed outcome | epidemiological parameter (time from symptom onset to isolation, isolation interval) |
| Method to obtain the computed outcome | simple computation |
| Data source | data from (sub)national authorities |
| Data availability | data not accessible |
| **Intervention** | |
| Terminology for interventions | responses |
| Terminology for the specific type of non-pharmaceutical interventions | government |
| Exposure types | phases of interventions (five phases of different nonpharmaceutical interventions which ronly oughly correspond to different alert levels) |
| Types of single interventions | – not applicable – |
| Coding of interventions | not necessary |
| Source of intervention data | – could not be evaluated – |
| Availability of data on exposure | raw data documented in the manuscript |
| **Methodological approach** | |
| Empirical approach | descriptive |
| Use of exposure variation | only variation over time for a single population |
| Method | description of change over time (comparison of time periods) |
| Code availability | none |
| **Effect assessment** | |

– continued on next page –

| | |
|---|---|
| Reporting of intervention effect | comparison of outcome values |
| Measure of effect | cases, hospitalizations, ICU hospitalizations, deaths, time from symptom onset to isolation, isolation interval |
| Reporting of uncertainty | yes |
| Sensitivity analysis | same analysis with (sub)population(s) excluded |
| Subgroup assessment | socioeconomic indicators (age, gender, income, health care occupation, ethnicity), epidemiological indicators (source of infection) |

**Jeffrey et al. 2020 (Analysis 1)**

| | |
|---|---|
| **Study information** | |
| Author | Jeffrey B, Walters CE, Ainslie KE, Eales O, Ciavarella C, Bhatia S, Hayes S, Baguelin M, Boonyasiri A, Brazeau NF, Cuomo-Dannenburg G, FitzJohn RG, Gaythorpe K, Green W, Imai N, Mellan TA, Mishra S, Nouvellet P, Unwin HJ, Verity R, Vollmer M, Whittaker C, Ferguson NM, Donnelly CA, Riley S |
| Title | Anonymised and Aggregated Crowd Level Mobility Data from Mobile Phones Suggests That Initial Compliance with COVID-19 Social Distancing Interventions Was High and Geographically Consistent across the UK |
| Year | 2020 |
| Journal | Wellcome Open Research |
| DOI | 10.12688/wellcomeopenres.15997.1 |
| Date received | – could not be evaluated – |
| Date accepted | 2020-07-17 (published) |
| **Study setting** | |
| Number of populations included | multiple |

– continued on next page –

| | |
|---|---|
| Level of populations included | both national and subnational (UK countries, local authority districts, cities) |
| Geographic areas covered | Europe |
| Number of countries covered | single country (UK) |
| Study period | start and end date span first epidemic wave: behavioral |
| **Outcome** | |
| Raw outcome | epidemiological population-level outcome (cases), behavioral outcome (mobility: transit station mobility index & subway validations) |
| Time resolution of raw outcome | daily |
| Computed outcome | none |
| Method to obtain the computed outcome | – not applicable – |
| Data source | mobility data from corporate organizations (Facebook (number of trips and trip lengths), O2 (number of trips and trip lengths)) |
| Data availability | data made available by the authors |
| **Intervention** | |
| Terminology for interventions | interventions |
| Terminology for the specific type of non-pharmaceutical interventions | social distancing |
| Exposure types | multiple separate interventions (school closure, venue closure, stay-at-home order) |
| Types of single interventions | school closure, venue closure, stay-at-home order |
| Coding of interventions | not necessary |
| Source of intervention data | government or news websites |
| Availability of data on exposure | raw data documented in the manuscript |
| **Methodological approach** | |
| Empirical approach | descriptive |

– continued on next page –

| | |
|---|---|
| Use of exposure variation | only variation over time for multiple populations |
| Method | description of change over time (comparison of time periods) |
| Code availability | publicly available |
| **Effect assessment** | |
| Reporting of intervention effect | comparison of outcome values |
| Measure of effect | mobility: percentage difference from baseline number of trips and trip lengths |
| Reporting of uncertainty | no |
| Sensitivity analysis | different or modified outcome used (different mobility data used) |
| Subgroup assessment | socioeconomic indicators (population density) |

**Jeffrey et al. 2020 (Analysis 2, mobility as exposure)**

| | |
|---|---|
| **Study information** | |
| Author | Jeffrey B, Walters CE, Ainslie KE, Eales O, Ciavarella C, Bhatia S, Hayes S, Baguelin M, Boonyasiri A, Brazeau NF, Cuomo-Dannenburg G, FitzJohn RG, Gaythorpe K, Green W, Imai N, Mellan TA, Mishra S, Nouvellet P, Unwin HJ, Verity R, Vollmer M, Whittaker C, Ferguson NM, Donnelly CA, Riley S |
| Title | Anonymised and Aggregated Crowd Level Mobility Data from Mobile Phones Suggests That Initial Compliance with COVID-19 Social Distancing Interventions Was High and Geographically Consistent across the UK |
| Year | 2020 |
| Journal | Wellcome Open Research |
| DOI | 10.12688/wellcomeopenres.15997.1 |
| Date received | – could not be evaluated – |
| Date accepted | 17.07.2020 (published) |

– continued on next page –

| | |
|---|---|
| **Study setting** | |
| Number of populations included | multiple |
| Level of populations included | subnational (local authority districts in UK) |
| Geographic areas covered | Europe |
| Number of countries covered | single country (UK) |
| Study period | start and end date span first epidemic wave |
| **Outcome** | |
| Raw outcome | behavioral outcome (mobility: number of trips) |
| Time resolution of raw outcome | daily |
| Computed outcome | none |
| Method to obtain the computed outcome | – not applicable – |
| Data source | mobility data by corporate organizations (Facebook) |
| Data availability | data made available by the authors |
| **Intervention** | |
| Terminology for interventions | interventions |
| Terminology for the specific type of non-pharmaceutical interventions | social distancing |
| Exposure types | change points (mobility: number of trips) |
| Types of single interventions | – not applicable – |
| Coding of interventions | – not applicable – |
| Source of intervention data | data collected by authors |
| Availability of data on exposure | data made available by the authors |
| **Methodological approach** | |
| Empirical approach | descriptive |
| Use of exposure variation | only variation over time for multiple populations |

– continued on next page –

**Jeffrey et al. 2020 (Analysis 2, mobility as exposure)** – continued from previous page

| | |
|---|---|
| Method | description of change over time (comparison of time periods) |
| Code availability | publicly available |
| **Effect assessment** | |
| Reporting of intervention effect | quantification of change in outcome values |
| Measure of effect | mobility: percentage difference from baseline number of trips |
| Reporting of uncertainty | yes |
| Sensitivity analysis | model specification varied |
| Subgroup assessment | none |

**Ji et al. 2020**

| | |
|---|---|
| **Study information** | |
| Author | Ji T, Chen HL, Xu J, Wu LN, Li JJ, Chen K, Qin G |
| Title | Lockdown Contained the Spread of 2019 Novel Coronavirus Disease in Huangshi City, China: Early Epidemiological Findings |
| Year | 2020 |
| Journal | Clinical Infectious Diseases |
| DOI | 10.1093/cid/ciaa390 |
| Date received | 2020-02-24 |
| Date accepted | 2020-04-03 |
| **Study setting** | |
| Number of populations included | single |
| Level of populations included | subnational (Huangshi (China)) |
| Geographic areas covered | Asia |
| Number of countries covered | single country (China) |

– continued on next page –

| | |
|---|---|
| Study period | start and end date span first epidemic wave |
| **Outcome** | |
| Raw outcome | epidemiological population-level outcome (cases) |
| Time resolution of raw outcome | daily |
| Computed outcome | none |
| Method to obtain the computed outcome | – not applicable – |
| Data source | data from (sub)national authorities |
| Data availability | data access via source |
| **Intervention** | |
| Terminology for interventions | interventions |
| Terminology for the specific type of non-pharmaceutical interventions | non-pharmaceutical |
| Exposure types | one combination of interventions (Wuhan City Lockdown) |
| Types of single interventions | – not applicable – |
| Coding of interventions | not necessary |
| Source of intervention data | another study (Rubin et al.) |
| Availability of data on exposure | raw data documented in the manuscript |
| **Methodological approach** | |
| Empirical approach | descriptive |
| Use of exposure variation | only variation over time for a single population |
| Method | description of change over time (description of time course) |
| Code availability | none |
| **Effect assessment** | |
| Reporting of intervention effect | qualitative statement |
| Measure of effect | cases |

– continued on next page –

| | |
|---|---|
| Reporting of uncertainty | – not applicable – |
| Sensitivity analysis | none |
| Subgroup assessment | none |

| **Jia et al. 2020** | |
|---|---|
| **Study information** | |
| Author | Jia JS, Lu X, Yuan Y, Xu G, Jia J, Christakis NA |
| Title | Population Flow Drives Spatio-Temporal Distribution of COVID-19 in China |
| Year | 2020 |
| Journal | Nature |
| DOI | 10.1038/s41586-020-2284-y |
| Date received | 2020-02-18 |
| Date accepted | 2020-04-21 |
| **Study setting** | |
| Number of populations included | multiple |
| Level of populations included | both national and subnational (China and its provinces) |
| Geographic areas covered | Asia |
| Number of countries covered | single country (China) |
| Study period | start and end date span first epidemic wave: behavioral |
| **Outcome** | |
| Raw outcome | behavioral outcome (mobility: migration from Wuhan) |
| Time resolution of raw outcome | daily |
| Computed outcome | none |

– continued on next page –

| | |
|---|---|
| Method to obtain the computed outcome | – not applicable – |
| Data source | mobility data from corporate organizations (major national carrier) |
| Data availability | data made available by the authors |
| **Intervention** | |
| Terminology for interventions | interventions |
| Terminology for the specific type of non-pharmaceutical interventions | public health |
| Exposure types | one combination of interventions (Wuhan City Lockdown) |
| Types of single interventions | – not applicable – |
| Coding of interventions | not necessary |
| Source of intervention data | – could not be evaluated – |
| Availability of data on exposure | raw data documented in the manuscript |
| **Methodological approach** | |
| Empirical approach | descriptive |
| Use of exposure variation | only variation over time for multiple populations |
| Method | description of change over time (description of time course) |
| Code availability | publicly available |
| **Effect assessment** | |
| Reporting of intervention effect | quantification of change in outcome values |
| Measure of effect | mobility: migration from Wuhan |
| Reporting of uncertainty | no |
| Sensitivity analysis | none |
| Subgroup assessment | none |

| Jüni et al. 2020 | |
| --- | --- |
| **Study information** | |
| Author | Jüni P, Rothenbühler M, Bobos P, Thorpe KE, da Costa BR, Fisman DN, Slutsky AS, Gesink D |
| Title | Impact of Climate and Public Health Interventions on the COVID-19 Pandemic: A Prospective Cohort Study |
| Year | 2020 |
| Journal | Canadian Medical Association Journal |
| DOI | 10.1503/cmaj.200920 |
| Date received | – could not be evaluated – |
| Date accepted | 2020-05-05 |
| **Study setting** | |
| Number of populations included | multiple |
| Level of populations included | both national and subnational (144 "geopolitical areas") |
| Geographic areas covered | Europe, Middle East and Africa, Asia, Oceania, Central and South America, North America |
| Number of countries covered | multiple countries |
| Study period | late start |
| **Outcome** | |
| Raw outcome | epidemiological population-level outcome (cases) |
| Time resolution of raw outcome | – not applicable – |
| Computed outcome | measure of epidemic trend (growth rate) |
| Method to obtain the computed outcome | simple computation |
| Data source | data from publicly available cross-country selections (JHU) |
| Data availability | data access via source |
| **Intervention** | |

– continued on next page –

| | |
|---|---|
| Terminology for interventions | interventions |
| Terminology for the specific type of non-pharmaceutical interventions | public health |
| Exposure types | multiple combinations of interventions (bans of small gatherings, school closure, other social distancing measures (there was variation in measures of social distancing reported by different geopolitical areas, including recommendations or requirements regarding social distancing, closure of sit-in restaurants and bars, or closure of nongrocery stores)) |
| Types of single interventions | – not applicable – |
| Coding of interventions | necessary |
| Source of intervention data | coding done by authors |
| Availability of data on exposure | coded data not available |
| **Methodological approach** | |
| Empirical approach | parametric |
| Use of exposure variation | only variation between populations |
| Method | non-mechanistic model (generalized linear model) |
| Code availability | none |
| **Effect assessment** | |
| Reporting of intervention effect | quantification of change in outcome values |
| Measure of effect | growth rate |
| Reporting of uncertainty | yes |
| Sensitivity analysis | model specification varied |
| Subgroup assessment | none |

| Kang & Kim 2020 | |
|---|---|
| **Study information** | |
| Author | Kang N, Kim B |
| Title | The Effects of Border Shutdowns on the Spread of COVID-19 |
| Year | 2020 |
| Journal | Journal of Preventive Medicine and Public Health |
| DOI | 10.3961/jpmph.20.332 |
| Date received | 2020-07-17 |
| Date accepted | 2020-07-20 |
| **Study setting** | |
| Number of populations included | multiple |
| Level of populations included | national (Australia, Singapore, USA, Vietnam, Taiwan, Hong Kong) |
| Geographic areas covered | Europe, Middle East and Africa, Asia, Oceania, Central and South America, North America |
| Number of countries covered | multiple countries |
| Study period | end date in growth phase of wave |
| **Outcome** | |
| Raw outcome | epidemiological population-level outcome (cases) |
| Time resolution of raw outcome | daily |
| Computed outcome | none |
| Method to obtain the computed outcome | – not applicable – |
| Data source | data from publicly available cross-country selections (WHO) |
| Data availability | data access via source |
| **Intervention** | |
| Terminology for interventions | – not applicable – |

– continued on next page –

| | |
|---|---|
| Terminology for the specific type of non-pharmaceutical interventions | border shutdown |
| Exposure types | one single intervention (travel restrictions (entry bans against China)) |
| Types of single interventions | international travel restrictions |
| Coding of interventions | not necessary |
| Source of intervention data | government or news websites |
| Availability of data on exposure | raw data documented in the manuscript |
| **Methodological approach** | |
| Empirical approach | counterfactual |
| Use of exposure variation | only variation over time for multiple populations |
| Method | synthetic controls |
| Code availability | none |
| **Effect assessment** | |
| Reporting of intervention effect | quantification of change in outcome values |
| Measure of effect | cases |
| Reporting of uncertainty | no |
| Sensitivity analysis | different or modified outcome used (new cases used instead of cumulative cases), model specification varied |
| Subgroup assessment | none |

**Karnakov et al. 2020 (Analysis 1)**

| | |
|---|---|
| **Study information** | |
| Author | Karnakov P, Arampatzis G, Kii I, Wermelinger F, Wlchli D, Papadimitriou C, Koumoutsakos P |

– continued on next page –

| | |
|---|---|
| Title | Data-Driven Inference of the Reproduction Number for COVID-19 before and after Interventions for 51 European Countries |
| Year | 2020 |
| Journal | Swiss Medical Weekly |
| DOI | 10.4414/smw.2020.20313 |
| Date received | – could not be evaluated – |
| Date accepted | 2020-07-17 (published) |
| **Study setting** | |
| Number of populations included | multiple |
| Level of populations included | national (51 countries) |
| Geographic areas covered | Europe |
| Number of countries covered | multiple countries |
| Study period | start and end date span first epidemic wave |
| **Outcome** | |
| Raw outcome | epidemiological population-level outcome (cases) |
| Time resolution of raw outcome | daily |
| Computed outcome | none |
| Method to obtain the computed outcome | – not applicable – |
| Data source | data from publicly available open-source projects (data from publicly available open-source projects by the Humanistic GIS Lab at University of Washington) |
| Data availability | data made available by the authors |
| **Intervention** | |
| Terminology for interventions | interventions |

– continued on next page –

| | |
|---|---|
| Terminology for the specific type of non-pharmaceutical interventions | non-pharmaceutical |
| Exposure types | start of interventions (estimated change points when interventions change R (roughly corresponding to the day of the first intervention leading to lockdown)) |
| Types of single interventions | – not applicable – |
| Coding of interventions | – not applicable – |
| Source of intervention data | – not applicable – |
| Availability of data on exposure | – not applicable – |
| **Methodological approach** | |
| Empirical approach | parametric |
| Use of exposure variation | only variation over time for multiple populations |
| Method | mechanistic model (compartmental single-population transmission model) |
| Latent variable functionally linked to intervention | reproduction number |
| Code availability | publicly available |
| **Effect assessment** | |
| Reporting of intervention effect | comparison of outcome values |
| Measure of effect | reproduction number |
| Reporting of uncertainty | yes |
| Sensitivity analysis | model specification varied |
| Subgroup assessment | none |

**Karnakov et al. 2020 (Analysis 2)**

**Study information**

– continued on next page –

| | |
|---|---|
| Author | Karnakov P, Arampatzis G, Kii I, Wermelinger F, Wlchli D, Papadimitriou C, Koumoutsakos P |
| Title | Data-Driven Inference of the Reproduction Number for COVID-19 before and after Interventions for 51 European Countries |
| Year | 2020 |
| Journal | Swiss Medical Weekly |
| DOI | 10.4414/smw.2020.20313 |
| Date received | – could not be evaluated – |
| Date accepted | 2020-07-17 (published) |
| **Study setting** | |
| Number of populations included | multiple |
| Level of populations included | national (51 countries) |
| Geographic areas covered | Europe |
| Number of countries covered | multiple countries |
| Study period | start and end date span first epidemic wave |
| **Outcome** | |
| Raw outcome | epidemiological population-level outcome (cases) |
| Time resolution of raw outcome | daily |
| Computed outcome | change points (change points: reproduction number) |
| Method to obtain the computed outcome | compartmental transmission model (compartmental single-population transmission model, with change points) |
| Data source | data from publicly available open-source projects (data from publicly available open-source projects by the Humanistic GIS Lab at University of Washington) |
| Data availability | data made available by the authors |
| **Intervention** | |

– continued on next page –

| | |
|---|---|
| Terminology for interventions | interventions |
| Terminology for the specific type of non-pharmaceutical interventions | non-pharmaceutical |
| Exposure types | start of interventions (start date of non-pharmaceutical interventions (various)) |
| Types of single interventions | – not applicable – |
| Coding of interventions | necessary |
| Source of intervention data | use of externally coded data (Wikipedia) |
| Availability of data on exposure | access to externally coded data via source |
| **Methodological approach** | |
| Empirical approach | descriptive |
| Use of exposure variation | only variation over time for multiple populations |
| Method | comparison of change points with intervention dates |
| Code availability | publicly available |
| **Effect assessment** | |
| Reporting of intervention effect | quantification of change in outcome values |
| Measure of effect | change points: reproduction number |
| Reporting of uncertainty | yes |
| Sensitivity analysis | none |
| Subgroup assessment | none |

**Kaufman et al. 2020**

| | |
|---|---|
| **Study information** | |
| Author | Kaufman BG, Whitaker R, Mahendraratnam N, Smith VA, McClellan MB |

– continued on next page –

| | |
|---|---|
| Title | Comparing Associations of State Reopening Strategies with COVID-19 Burden |
| Year | 2020 |
| Journal | Journal of General Internal Medicine |
| DOI | 10.1007/s11606-020-06277-0 |
| Date received | 2020-09-12 |
| Date accepted | 2020-09-28 |
| **Study setting** | |
| Number of populations included | multiple |
| Level of populations included | both national and subnational (50 US states and Washington D.C.) |
| Geographic areas covered | North America |
| Number of countries covered | single country (US) |
| Study period | same end date for several populations with diverse epidemic trajectories |
| **Outcome** | |
| Raw outcome | epidemiological population-level outcome (cases, deaths) |
| Time resolution of raw outcome | daily |
| Computed outcome | none |
| Method to obtain the computed outcome | – not applicable – |
| Data source | data from public media outlets (New York Times) |
| Data availability | data access via source |
| **Intervention** | |
| Terminology for interventions | measures |
| Terminology for the specific type of non-pharmaceutical interventions | mitigation |
| Exposure types | one single intervention (mask mandate (statewide)) |

– continued on next page –

| | |
|---|---|
| Types of single interventions | mask mandate |
| Coding of interventions | necessary |
| Source of intervention data | use of externally coded data (Boston University COVID-19 US State Policy Database) |
| Availability of data on exposure | coded data made available by the authors |
| **Methodological approach** | |
| Empirical approach | parametric |
| Use of exposure variation | both variation over time and between populations |
| Method | non-mechanistic model (generalized linear model) |
| Code availability | none |
| **Effect assessment** | |
| Reporting of intervention effect | quantification of change in outcome values |
| Measure of effect | cases, deaths |
| Reporting of uncertainty | yes |
| Sensitivity analysis | model specification varied, same analysis with (sub)population(s) excluded |
| Subgroup assessment | none |

**Kendall et al. 2020 (Analysis 1)**

| | |
|---|---|
| **Study information** | |
| Author | Kendall M, Milsom L, Abeler-Dörner L, Wymant C, Ferretti L, Briers M, Holmes C, Bonsall D, Abeler J, Fraser C |
| Title | Epidemiological Changes on the Isle of Wight after the Launch of the NHS Test and Trace Programme: A Preliminary Analysis |
| Year | 2020 |

– continued on next page –

| | |
|---|---|
| Journal | The Lancet Digital Health |
| DOI | 10.1016/s2589-7500(20)30241-7 |
| Date received | – could not be evaluated – |
| Date accepted | 2020-10-14 (published) |
| **Study setting** | |
| Number of populations included | single |
| Level of populations included | national (Isle of Wight (UK)) |
| Geographic areas covered | Europe |
| Number of countries covered | single country (UK) |
| Study period | start and end date span first epidemic wave |
| **Outcome** | |
| Raw outcome | epidemiological population-level outcome (cases) |
| Time resolution of raw outcome | daily |
| Computed outcome | epidemiological parameter (reproduction number) |
| Method to obtain the computed outcome | back projection, statistical estimation of reproduction number (Cori et al.) |
| Data source | data collected by authors (hospitals: cases), data from (sub)national authorities (cases) |
| Data availability | data made available by the authors |
| **Intervention** | |
| Terminology for interventions | interventions |
| Terminology for the specific type of non-pharmaceutical interventions | non-pharmaceutical |
| Exposure types | one combination of interventions (NHS Test and Trace programme) |
| Types of single interventions | – not applicable – |
| Coding of interventions | not necessary |

– continued on next page –

**Kendall et al. 2020 (Analysis 1)** – continued from previous page

| | |
|---|---|
| Source of intervention data | – could not be evaluated – |
| Availability of data on exposure | raw data documented in the manuscript |
| **Methodological approach** | |
| Empirical approach | descriptive |
| Use of exposure variation | only variation over time for a single population |
| Method | description of change over time (comparison of time periods) |
| Code availability | publicly available |
| **Effect assessment** | |
| Reporting of intervention effect | comparison of outcome values |
| Measure of effect | reproduction number, infections |
| Reporting of uncertainty | no |
| Sensitivity analysis | none |
| Subgroup assessment | none |

| **Kendall et al. 2020 (Analysis 2)** | |
|---|---|
| **Study information** | |
| Author | Kendall M, Milsom L, Abeler-Dörner L, Wymant C, Ferretti L, Briers M, Holmes C, Bonsall D, Abeler J, Fraser C |
| Title | Epidemiological Changes on the Isle of Wight after the Launch of the NHS Test and Trace Programme: A Preliminary Analysis |
| Year | 2020 |
| Journal | The Lancet Digital Health |
| DOI | 10.1016/s2589-7500(20)30241-7 |
| Date received | – could not be evaluated – |
| Date accepted | 2020-10-14 (published) |

– continued on next page –

| | |
|---|---|
| **Study setting** | |
| Number of populations included | single |
| Level of populations included | national (Isle of Wight (UK)) |
| Geographic areas covered | Europe |
| Number of countries covered | single country (UK) |
| Study period | start and end date span first epidemic wave |
| **Outcome** | |
| Raw outcome | epidemiological population-level outcome (cases) |
| Time resolution of raw outcome | daily |
| Computed outcome | epidemiological parameter (reproduction number) |
| Method to obtain the computed outcome | back projection, statistical estimation of reproduction number (Cori et al.) |
| Data source | data collected by authors (hospitals: cases), data from (sub)national authorities (cases) |
| Data availability | data made available by the authors |
| **Intervention** | |
| Terminology for interventions | interventions |
| Terminology for the specific type of non-pharmaceutical interventions | non-pharmaceutical |
| Exposure types | one combination of interventions (NHS Test and Trace programme) |
| Types of single interventions | – not applicable – |
| Coding of interventions | not necessary |
| Source of intervention data | – could not be evaluated – |
| Availability of data on exposure | raw data documented in the manuscript |
| **Methodological approach** | |
| Empirical approach | counterfactual |

– continued on next page –

**Kendall et al. 2020 (Analysis 2)** – continued from previous page

| | |
|---|---|
| Use of exposure variation | only variation over time for a single population |
| Method | synthetic controls |
| Code availability | publicly available |
| **Effect assessment** | |
| Reporting of intervention effect | quantification of change in outcome values |
| Measure of effect | reproduction number |
| Reporting of uncertainty | yes |
| Sensitivity analysis | none |
| Subgroup assessment | none |

**Khan et al. 2020**

| | |
|---|---|
| **Study information** | |
| Author | Khan NM, Soobhug AD, Khan MH |
| Title | Studying the Trend of the Novel Coronavirus Series in Mauritius and Its Implications |
| Year | 2020 |
| Journal | PLOS ONE |
| DOI | 10.1371/journal.pone.0235730 |
| Date received | 2020-05-04 |
| Date accepted | 2020-06-10 |
| **Study setting** | |
| Number of populations included | single |
| Level of populations included | national (Mauritius) |
| Geographic areas covered | Middle East and Africa |

– continued on next page –

| | |
|---|---|
| Number of countries covered | single country (Mauritius) |
| Study period | start and end date span first epidemic wave |
| **Outcome** | |
| Raw outcome | epidemiological population-level outcome (active cases) |
| Time resolution of raw outcome | daily |
| Computed outcome | none |
| Method to obtain the computed outcome | – not applicable – |
| Data source | data from publicly available cross-country selections (ECDC) |
| Data availability | data made available by the authors |
| **Intervention** | |
| Terminology for interventions | measures |
| Terminology for the specific type of non-pharmaceutical interventions | confinement |
| Exposure types | one combination of interventions (sanitary curfew on March 20 (work bans, non-essential business restrictions, social distancing in public places)) |
| Types of single interventions | – not applicable – |
| Coding of interventions | not necessary |
| Source of intervention data | – could not be evaluated – |
| Availability of data on exposure | raw data documented in the manuscript |
| **Methodological approach** | |
| Empirical approach | parametric |
| Use of exposure variation | only variation over time for a single population |
| Method | non-mechanistic model (generalized linear model) |
| Code availability | none |
| **Effect assessment** | |

– continued on next page –

| | |
|---|---|
| Reporting of intervention effect | quantification of change in outcome values |
| Measure of effect | active cases |
| Reporting of uncertainty | yes |
| Sensitivity analysis | model specification varied |
| Subgroup assessment | none |

| **Kharroubi & Saleh 2020** | |
|---|---|
| **Study information** | |
| Author | Kharroubi S, Saleh F |
| Title | Are Lockdown Measures Effective against COVID-19? |
| Year | 2020 |
| Journal | Frontiers in Public Health |
| DOI | 10.3389/fpubh.2020.549692 |
| Date received | 2020-04-09 |
| Date accepted | 2020-09-09 |
| **Study setting** | |
| Number of populations included | single |
| Level of populations included | national (Lebanon) |
| Geographic areas covered | Middle East and Africa |
| Number of countries covered | single country (Lebanon) |
| Study period | start and end date span first epidemic wave |
| **Outcome** | |
| Raw outcome | epidemiological population-level outcome (cases) |
| Time resolution of raw outcome | daily |

– continued on next page –

| | |
|---|---|
| Computed outcome | none |
| Method to obtain the computed outcome | – not applicable – |
| Data source | data from (sub)national authorities |
| Data availability | data access via source |
| **Intervention** | |
| Terminology for interventions | measures |
| Terminology for the specific type of non-pharmaceutical interventions | containment |
| Exposure types | one combination of interventions (lockdown decreed on March 15 (full lockdown, shutting down airport, imposing travel restrictions and completely sealing the borders)) |
| Types of single interventions | – not applicable – |
| Coding of interventions | not necessary |
| Source of intervention data | – could not be evaluated – |
| Availability of data on exposure | raw data documented in the manuscript |
| **Methodological approach** | |
| Empirical approach | parametric |
| Use of exposure variation | only variation over time for a single population |
| Method | non-mechanistic model (generalized linear model) |
| Code availability | none |
| **Effect assessment** | |
| Reporting of intervention effect | quantification of change in outcome values |
| Measure of effect | cases |
| Reporting of uncertainty | no |
| Sensitivity analysis | none |

– continued on next page –

| | |
|---|---|
| Subgroup assessment | none |

| **Kim et al. 2020a** | |
|---|---|
| **Study information** | |
| Author | Kim YJ, Seo MH, Yeom HE |
| Title | Estimating a Breakpoint in the Pattern of Spread of COVID-19 in South Korea |
| Year | 2020 |
| Journal | International Journal of Infectious Diseases |
| DOI | 10.1016/j.ijid.2020.06.055 |
| Date received | 2020-05-05 |
| Date accepted | 2020-06-16 |
| **Study setting** | |
| Number of populations included | single |
| Level of populations included | national (South Korea) |
| Geographic areas covered | Asia |
| Number of countries covered | single country (South Korea) |
| Study period | start and end date span first epidemic wave |
| **Outcome** | |
| Raw outcome | epidemiological population-level outcome (cases) |
| Time resolution of raw outcome | daily |
| Computed outcome | none |
| Method to obtain the computed outcome | – not applicable – |

– continued on next page –

| | |
|---|---|
| Data source | data from (sub)national authorities |
| Data availability | data access via source |
| **Intervention** | |
| Terminology for interventions | interventions |
| Terminology for the specific type of non-pharmaceutical interventions | public health |
| Exposure types | all interventions together |
| Types of single interventions | – not applicable – |
| Coding of interventions | – not applicable – |
| Source of intervention data | – not applicable – |
| Availability of data on exposure | – not applicable – |
| **Methodological approach** | |
| Empirical approach | parametric |
| Use of exposure variation | only variation over time for a single population |
| Method | mechanistic model (compartmental single-population transmission model) |
| Latent variable functionally linked to intervention | transmission rate |
| Code availability | none |
| **Effect assessment** | |
| Reporting of intervention effect | quantification of change in outcome values |
| Measure of effect | transmission rate |
| Reporting of uncertainty | no |
| Sensitivity analysis | epidemiological parameters varied |
| Subgroup assessment | none |

| Kim et al. 2020b | |
|---|---|
| **Study information** | |
| Author | Kim S, Ko Y, Kim YJ, Jung E |
| Title | The Impact of Social Distancing and Public Behavior Changes on COVID-19 Transmission Dynamics in the Republic of Korea |
| Year | 2020 |
| Journal | PLOS ONE |
| DOI | 10.1371/journal.pone.0238684 |
| Date received | 2020-05-01 |
| Date accepted | 2020-08-22 |
| **Study setting** | |
| Number of populations included | single |
| Level of populations included | national (Korea) |
| Geographic areas covered | Asia |
| Number of countries covered | single country (Korea) |
| Study period | start and end date span first epidemic wave |
| **Outcome** | |
| Raw outcome | epidemiological population-level outcome (cases) |
| Time resolution of raw outcome | daily |
| Computed outcome | none |
| Method to obtain the computed outcome | – not applicable – |
| Data source | data from (sub)national authorities |
| Data availability | data access via source |
| **Intervention** | |
| Terminology for interventions | interventions |

– continued on next page –

| | |
|---|---|
| Terminology for the specific type of non-pharmaceutical interventions | non-pharmaceutical |
| Exposure types | all interventions together |
| Types of single interventions | – not applicable – |
| Coding of interventions | – not applicable – |
| Source of intervention data | – not applicable – |
| Availability of data on exposure | – not applicable – |
| **Methodological approach** | |
| Empirical approach | parametric |
| Use of exposure variation | only variation over time for a single population |
| Method | mechanistic model (compartmental single-population transmission model) |
| Latent variable functionally linked to intervention | other (behavior change rate: transition of susceptibles to compartment with lower transmission rate) |
| Code availability | none |
| **Effect assessment** | |
| Reporting of intervention effect | quantification of change in outcome values |
| Measure of effect | behavior change rate |
| Reporting of uncertainty | no |
| Sensitivity analysis | none |
| Subgroup assessment | none |

**Klimek-Tulwin & Tulwin 2020**

| | |
|---|---|
| **Study information** | |
| Author | Klimek-Tulwin M, Tulwin T |

– continued on next page –

| | |
|---|---|
| Title | Early School Closures Can Reduce the First-Wave of the COVID-19 Pandemic Development |
| Year | 2020 |
| Journal | Journal of Public Health |
| DOI | 10.1007/s10389-020-01391-z |
| Date received | 2020-05-30 |
| Date accepted | 2020-10-01 |
| **Study setting** | |
| Number of populations included | multiple |
| Level of populations included | national (15 countries) |
| Geographic areas covered | Asia, Central and South America, Europe |
| Number of countries covered | multiple countries |
| Study period | end date could not be evaluated |
| **Outcome** | |
| Raw outcome | epidemiological population-level outcome (cases) |
| Time resolution of raw outcome | daily |
| Computed outcome | none |
| Method to obtain the computed outcome | – not applicable – |
| Data source | data from publicly available cross-country selections (Worldometer) |
| Data availability | data access via source |
| **Intervention** | |
| Terminology for interventions | interventions |
| Terminology for the specific type of non-pharmaceutical interventions | government |

– continued on next page –

| | |
|---|---|
| Exposure types | one single intervention (school closure (relative timing of school closure, i.e. incidence at date of closure)) |
| Types of single interventions | school closure |
| Coding of interventions | necessary |
| Source of intervention data | use of externally coded data (UNESCO Institute for Statistics Database) |
| Availability of data on exposure | access to externally coded data via source |
| **Methodological approach** | |
| Empirical approach | parametric |
| Use of exposure variation | only variation between populations |
| Method | non-mechanistic model (correlation) |
| Code availability | none |
| **Effect assessment** | |
| Reporting of intervention effect | quantification of change in outcome values |
| Measure of effect | cases |
| Reporting of uncertainty | yes |
| Sensitivity analysis | none |
| Subgroup assessment | none |

**Koh et al. 2020 (Analysis 1)**

| | |
|---|---|
| **Study information** | |
| Author | Koh WC, Naing L, Wong J |
| Title | Estimating the Impact of Physical Distancing Measures in Containing COVID-19: An Empirical Analysis |

– continued on next page –

| | |
|---|---|
| Year | 2020 |
| Journal | International Journal of Infectious Diseases |
| DOI | 10.1016/j.ijid.2020.08.026 |
| Date received | 2020-06-16 |
| Date accepted | 2020-08-08 |
| **Study setting** | |
| Number of populations included | multiple |
| Level of populations included | national (142 countries worldwide) |
| Geographic areas covered | Europe, Middle East and Africa, Asia, Oceania, Central and South America, North America |
| Number of countries covered | multiple countries |
| Study period | end date could not be evaluated, other (start at date of 100th case) |
| **Outcome** | |
| Raw outcome | epidemiological population-level outcome (cases) |
| Time resolution of raw outcome | daily |
| Computed outcome | measure of epidemic trend (growth rate), epidemiological parameter (reproduction number) |
| Method to obtain the computed outcome | statistical estimation of reproduction number (Cori et al.), simple computation (growth rate) |
| Data source | data from publicly available cross-country selections (ECDC (cases), JHU (cases)) |
| Data availability | data access via source |
| **Intervention** | |
| Terminology for interventions | measures |
| Terminology for the specific type of non-pharmaceutical interventions | social distancing |

– continued on next page –

| | |
|---|---|
| Exposure types | multiple separate interventions (school closure, bans of large gatherings, bans of small gatherings, workplace closure, stay-at-home order, international travel restrictions, internal movement restrictions, public transport closure) |
| Types of single interventions | school closure, bans of large gatherings, bans of small gatherings, workplace closure, stay-at-home order, international travel restrictions, internal movement restrictions, public transport closure |
| Coding of interventions | necessary |
| Source of intervention data | use of externally coded data (OxCGRT) |
| Availability of data on exposure | access to externally coded data via source |
| **Methodological approach** | |
| Empirical approach | parametric |
| Use of exposure variation | only variation between populations |
| Method | non-mechanistic model (generalized linear model) |
| Code availability | none |
| **Effect assessment** | |
| Reporting of intervention effect | quantification of change in outcome values |
| Measure of effect | reproduction number, growth rate |
| Reporting of uncertainty | yes |
| Sensitivity analysis | model specification varied |
| Subgroup assessment | none |

**Koh et al. 2020 (Analysis 2)**

| | |
|---|---|
| **Study information** | |
| Author | Koh WC, Naing L, Wong J |

– continued on next page –

| | |
|---|---|
| Title | Estimating the Impact of Physical Distancing Measures in Containing COVID-19: An Empirical Analysis |
| Year | 2020 |
| Journal | International Journal of Infectious Diseases |
| DOI | 10.1016/j.ijid.2020.08.026 |
| Date received | 2020-06-16 |
| Date accepted | 2020-08-08 |
| **Study setting** | |
| Number of populations included | multiple |
| Level of populations included | national (142 countries worldwide) |
| Geographic areas covered | Europe, Middle East and Africa, Asia, Oceania, Central and South America, North America |
| Number of countries covered | multiple countries |
| Study period | end date could not be evaluated, other (start at date of 100th case) |
| **Outcome** | |
| Raw outcome | epidemiological population-level outcome (cases) |
| Time resolution of raw outcome | daily |
| Computed outcome | measure of epidemic trend (growth rate), epidemiological parameter (reproduction number) |
| Method to obtain the computed outcome | statistical estimation of reproduction number (Cori et al.), simple computation (growth rate) |
| Data source | data from publicly available cross-country selections (ECDC (cases), JHU (cases)) |
| Data availability | data access via source |
| **Intervention** | |
| Terminology for interventions | measures |

– continued on next page –

| | |
|---|---|
| Terminology for the specific type of non-pharmaceutical interventions | social distancing |
| Exposure types | stringency index (OxCGRT Stringency Index) |
| Types of single interventions | – not applicable – |
| Coding of interventions | necessary |
| Source of intervention data | use of externally coded data (OxCGRT) |
| Availability of data on exposure | access to externally coded data via source |
| **Methodological approach** | |
| Empirical approach | parametric |
| Use of exposure variation | only variation between populations |
| Method | non-mechanistic model (generalized linear model) |
| Code availability | none |
| **Effect assessment** | |
| Reporting of intervention effect | quantification of change in outcome values |
| Measure of effect | reproduction number, growth rate |
| Reporting of uncertainty | yes |
| Sensitivity analysis | model specification varied |
| Subgroup assessment | none |

**Koh et al. 2020 (Analysis 3, mobility as exposure)**

| | |
|---|---|
| **Study information** | |
| Author | Koh WC, Naing L, Wong J |
| Title | Estimating the Impact of Physical Distancing Measures in Containing COVID-19: An Empirical Analysis |
| Year | 2020 |

– continued on next page –

| | |
|---|---|
| Journal | International Journal of Infectious Diseases |
| DOI | 10.1016/j.ijid.2020.08.026 |
| Date received | 16.06.2020 |
| Date accepted | 08.08.2020 |
| **Study setting** | |
| Number of populations included | multiple |
| Level of populations included | national (142 countries worldwide) |
| Geographic areas covered | Europe, Middle East and Africa, Asia, Oceania, Central and South America, North America |
| Number of countries covered | multiple countries |
| Study period | end date could not be evaluated, other (start at date of 100th case) |
| **Outcome** | |
| Raw outcome | epidemiological population-level outcome (cases) |
| Time resolution of raw outcome | daily |
| Computed outcome | epidemiological parameter (reproduction number), measure of epidemic trend (growth rate) |
| Method to obtain the computed outcome | statistical estimation of reproduction number (Cori et al.), simple computation (growth rate) |
| Data source | data from publicly available cross-country selections (ECDC, JHU) |
| Data availability | data access via source |
| **Intervention** | |
| Terminology for interventions | measures |
| Terminology for the specific type of non-pharmaceutical interventions | social distancing |
| Exposure types | mobility (time spent in various locations) |
| Types of single interventions | – not applicable – |

– continued on next page –

| | |
|---|---|
| Coding of interventions | – not applicable – |
| Source of intervention data | mobility data by corporate organizations (Google) |
| Availability of data on exposure | data access via source |
| **Methodological approach** | |
| Empirical approach | parametric |
| Use of exposure variation | only variation between populations |
| Method | non-mechanistic model (generalized linear model) |
| Code availability | none |
| **Effect assessment** | |
| Reporting of intervention effect | quantification of change in outcome values |
| Measure of effect | reproduction number, growth rate |
| Reporting of uncertainty | yes |
| Sensitivity analysis | none |
| Subgroup assessment | none |

**Kraemer et al. 2020 (Analysis 1)**

| | |
|---|---|
| **Study information** | |
| Author | Kraemer MU, Yang CH, Gutierrez B, Wu CH, Klein B, Pigott DM, du Plessis L, Faria NR, Li R, Hanage WP, Brownstein JS, Layan M, Vespignani A, Tian H, Dye C, Pybus OG, And SV |
| Title | The Effect of Human Mobility and Control Measures on the COVID-19 Epidemic in China |
| Year | 2020 |
| Journal | Science |

– continued on next page –

| | |
|---|---|
| DOI | 10.1126/science.abb4218 |
| Date received | 2020-03-03 |
| Date accepted | 2020-03-24 |
| **Study setting** | |
| Number of populations included | multiple |
| Level of populations included | both national and subnational (China and its provinces) |
| Geographic areas covered | Asia |
| Number of countries covered | single country (China) |
| Study period | start and end date span first epidemic wave: behavioral |
| **Outcome** | |
| Raw outcome | behavioral outcome (mobility: migration from Wuhan) |
| Time resolution of raw outcome | daily |
| Computed outcome | none |
| Method to obtain the computed outcome | – not applicable – |
| Data source | mobility data from corporate organizations (Baidu) |
| Data availability | data made available by the authors |
| **Intervention** | |
| Terminology for interventions | measures |
| Terminology for the specific type of non-pharmaceutical interventions | control |
| Exposure types | one combination of interventions (Wuhan City Lockdown) |
| Types of single interventions | – not applicable – |
| Coding of interventions | not necessary |
| Source of intervention data | – could not be evaluated – |

– continued on next page –

| | |
|---|---|
| Availability of data on exposure | raw data documented in the manuscript |
| **Methodological approach** | |
| Empirical approach | descriptive |
| Use of exposure variation | only variation over time for multiple populations |
| Method | description of change over time (description of time course) |
| Code availability | publicly available |
| **Effect assessment** | |
| Reporting of intervention effect | qualitative statement |
| Measure of effect | mobility: migration from Wuhan |
| Reporting of uncertainty | – not applicable – |
| Sensitivity analysis | none |
| Subgroup assessment | none |

**Kraemer et al. 2020 (Analysis 2)**

| | |
|---|---|
| **Study information** | |
| Author | Kraemer MU, Yang CH, Gutierrez B, Wu CH, Klein B, Pigott DM, du Plessis L, Faria NR, Li R, Hanage WP, Brownstein JS, Layan M, Vespignani A, Tian H, Dye C, Pybus OG, And SV |
| Title | The Effect of Human Mobility and Control Measures on the COVID-19 Epidemic in China |
| Year | 2020 |
| Journal | Science |
| DOI | 10.1126/science.abb4218 |
| Date received | 2020-03-03 |
| Date accepted | 2020-03-24 |

– continued on next page –

| | |
|---|---|
| **Study setting** | |
| Number of populations included | multiple |
| Level of populations included | both national and subnational (China and its provinces) |
| Geographic areas covered | Asia |
| Number of countries covered | single country (China) |
| Study period | start and end date span first epidemic wave |
| **Outcome** | |
| Raw outcome | epidemiological population-level outcome (cases) |
| Time resolution of raw outcome | daily |
| Computed outcome | measure of epidemic trend (growth rate) |
| Method to obtain the computed outcome | other (time series model) |
| Data source | data from (sub)national authorities |
| Data availability | data made available by the authors |
| **Intervention** | |
| Terminology for interventions | measures |
| Terminology for the specific type of non-pharmaceutical interventions | control |
| Exposure types | one combination of interventions (Wuhan City Lockdown) |
| Types of single interventions | – not applicable – |
| Coding of interventions | not necessary |
| Source of intervention data | – could not be evaluated – |
| Availability of data on exposure | raw data documented in the manuscript |
| **Methodological approach** | |
| Empirical approach | descriptive |
| Use of exposure variation | only variation over time for multiple populations |

– continued on next page –

**Kraemer et al. 2020 (Analysis 2)** – continued from previous page

| | |
|---|---|
| Method | description of change over time (comparison of time periods) |
| Code availability | publicly available |
| **Effect assessment** | |
| Reporting of intervention effect | comparison of outcome values |
| Measure of effect | growth rate |
| Reporting of uncertainty | yes |
| Sensitivity analysis | none |
| Subgroup assessment | none |

**Kraemer et al. 2020 (Analysis 3, mobility as exposure)**

| | |
|---|---|
| **Study information** | |
| Author | Kraemer MU, Yang CH, Gutierrez B, Wu CH, Klein B, Pigott DM, du Plessis L, Faria NR, Li R, Hanage WP, Brownstein JS, Layan M, Vespignani A, Tian H, Dye C, Pybus OG, And SV |
| Title | The Effect of Human Mobility and Control Measures on the COVID-19 Epidemic in China |
| Year | 2020 |
| Journal | Science |
| DOI | 10.1126/science.abb4218 |
| Date received | 03.03.2020 |
| Date accepted | 24.03.2020 |
| **Study setting** | |
| Number of populations included | multiple |
| Level of populations included | both national and subnational (China and its provinces) |
| Geographic areas covered | Asia |

– continued on next page –

| | |
|---|---|
| Number of countries covered | single country (China) |
| Study period | start and end date span first epidemic wave |
| **Outcome** | |
| Raw outcome | epidemiological population-level outcome (cases) |
| Time resolution of raw outcome | daily |
| Computed outcome | none |
| Method to obtain the computed outcome | – not applicable – |
| Data source | data from (sub)national authorities |
| Data availability | data made available by the authors |
| **Intervention** | |
| Terminology for interventions | measures |
| Terminology for the specific type of non-pharmaceutical interventions | control |
| Exposure types | mobility (migration from Wuhan) |
| Types of single interventions | – not applicable – |
| Coding of interventions | – not applicable – |
| Source of intervention data | mobility data by corporate organizations (Baidu) |
| Availability of data on exposure | data access via source |
| **Methodological approach** | |
| Empirical approach | descriptive |
| Use of exposure variation | only variation over time for multiple populations |
| Method | non-mechanistic model (correlation) |
| Code availability | publicly available |
| **Effect assessment** | |
| Reporting of intervention effect | quantification of change in outcome values |

– continued on next page –

| | |
|---|---|
| Measure of effect | cases |
| Reporting of uncertainty | no |
| Sensitivity analysis | none |
| Subgroup assessment | none |

**Krishna 2020**

| | |
|---|---|
| **Study information** | |
| Author | Krishna MV |
| Title | Mathematical Modelling on Diffusion and Control of COVID–19 |
| Year | 2020 |
| Journal | Infectious Disease Modelling |
| DOI | 10.1016/j.idm.2020.08.009 |
| Date received | 2020-06-26 |
| Date accepted | 2020-08-14 |
| **Study setting** | |
| Number of populations included | single |
| Level of populations included | subnational (Wuhan) |
| Geographic areas covered | Asia |
| Number of countries covered | single country (China) |
| Study period | end date in growth phase of wave |
| **Outcome** | |
| Raw outcome | epidemiological population-level outcome (cases) |
| Time resolution of raw outcome | daily |
| Computed outcome | epidemiological parameter (reproduction number) |

– continued on next page –

| | |
|---|---|
| Method to obtain the computed outcome | compartmental transmission model (single-population) |
| Data source | – could not be evaluated – |
| Data availability | data not accessible |
| **Intervention** | |
| Terminology for interventions | measures |
| Terminology for the specific type of non-pharmaceutical interventions | control |
| Exposure types | one combination of interventions (Wuhan City Lockdown) |
| Types of single interventions | – not applicable – |
| Coding of interventions | not necessary |
| Source of intervention data | – could not be evaluated – |
| Availability of data on exposure | raw data documented in the manuscript |
| **Methodological approach** | |
| Empirical approach | descriptive |
| Use of exposure variation | only variation over time for a single population |
| Method | description of change over time (comparison of time periods) |
| Code availability | none |
| **Effect assessment** | |
| Reporting of intervention effect | comparison of outcome values |
| Measure of effect | reproduction number |
| Reporting of uncertainty | yes |
| Sensitivity analysis | none |
| Subgroup assessment | none |

| Kucharski et al. 2020 | |
|---|---|
| **Study information** | |
| Author | Kucharski AJ, Russell TW, Diamond C, Liu Y, Edmunds J, Funk S, Eggo RM, Sun F, Jit M, Munday JD, Davies N, Gimma A, van Zandvoort K, Gibbs H, Hellewell J, Jarvis CI, Clifford S, Quilty BJ, Bosse NI, Abbott S, Klepac P, Flasche S |
| Title | Early Dynamics of Transmission and Control of COVID-19: A Mathematical Modelling Study |
| Year | 2020 |
| Journal | The Lancet Infectious Diseases |
| DOI | 10.1016/s1473-3099(20)30144-4 |
| Date received | – could not be evaluated – |
| Date accepted | 2020-03-11 (published) |
| **Study setting** | |
| Number of populations included | single |
| Level of populations included | subnational (Wuhan) |
| Geographic areas covered | Asia |
| Number of countries covered | single country (China) |
| Study period | start and end date span first epidemic wave |
| **Outcome** | |
| Raw outcome | epidemiological population-level outcome (cases) |
| Time resolution of raw outcome | daily |
| Computed outcome | epidemiological parameter (reproduction number) |
| Method to obtain the computed outcome | compartmental transmission model (metapopulation) |

– continued on next page –

| | |
|---|---|
| Data source | data from publicly available cross-country selections (WHO: exported cases), data from other research projects (study by Huang et al. 2020: cases in Wuhan, study by Liu et al. 2020: cases in China), data from public media outlets (varous data from public media outletss: prevalence of infection on evacuation flights) |
| Data availability | data made available by the authors |
| **Intervention** | |
| Terminology for interventions | measures |
| Terminology for the specific type of non-pharmaceutical interventions | travel control |
| Exposure types | one single intervention (travel restrictions (introduced on Jan 23)) |
| Types of single interventions | international travel restrictions |
| Coding of interventions | not necessary |
| Source of intervention data | – could not be evaluated – |
| Availability of data on exposure | raw data documented in the manuscript |
| **Methodological approach** | |
| Empirical approach | descriptive |
| Use of exposure variation | only variation over time for a single population |
| Method | description of change over time (comparison of time periods) |
| Code availability | publicly available |
| **Effect assessment** | |
| Reporting of intervention effect | comparison of outcome values |
| Measure of effect | reproduction number |
| Reporting of uncertainty | yes |
| Sensitivity analysis | different or modified outcome used (different confirmed case data), epidemiological parameters varied |

– continued on next page –

| | |
|---|---|
| Subgroup assessment | none |

| **Kumar et al. 2020** | |
|---|---|
| **Study information** | |
| Author | Kumar V, Sood A, Gupta S, Sood N |
| Title | Prevention- versus Promotion-Focus Regulatory Efforts on the Disease Incidence and Mortality of COVID-19: A Multinational Diffusion Study Using Functional Data Analysis |
| Year | 2020 |
| Journal | Journal of International Marketing |
| DOI | 10.1177/1069031x20966563 |
| Date received | – could not be evaluated – |
| Date accepted | 2020-12-14 (published) |
| **Study setting** | |
| Number of populations included | multiple |
| Level of populations included | national (70 countries around the globe) |
| Geographic areas covered | Europe, Middle East and Africa, Asia, Oceania, Central and South America, North America |
| Number of countries covered | multiple countries |
| Study period | start and end date span first epidemic wave |
| **Outcome** | |
| Raw outcome | epidemiological population-level outcome (cases) |
| Time resolution of raw outcome | daily |
| Computed outcome | none |

– continued on next page –

| | |
|---|---|
| Method to obtain the computed outcome | – not applicable – |
| Data source | data from publicly available cross-country selections (ECDC) |
| Data availability | data access via source |
| **Intervention** | |
| Terminology for interventions | interventions |
| Terminology for the specific type of non-pharmaceutical interventions | non-pharmaceutical |
| Exposure types | categories of interventions (containment and closure policies aggregated via principal component analysis based on individual measures (shelter-in-place orders, workplace closure, mobility restrictions, gathering bans, school closure, mask-mandates, time-based curfews, partial lockdown)) |
| Types of single interventions | – not applicable – |
| Coding of interventions | necessary |
| Source of intervention data | use of externally coded data (OxCGRT) |
| Availability of data on exposure | access to externally coded data via source |
| **Methodological approach** | |
| Empirical approach | parametric |
| Use of exposure variation | both variation over time and between populations |
| Method | non-mechanistic model (generalized linear model) |
| Code availability | none |
| **Effect assessment** | |
| Reporting of intervention effect | quantification of change in outcome values |
| Measure of effect | cases |
| Reporting of uncertainty | yes |
| Sensitivity analysis | none |

– continued on next page –

| | |
|---|---|
| Subgroup assessment | epidemiological indicators (pre-intervention epidemic trend), socioeconomic indicators (population density, age, health care occupation), public health response (subject to other measures) |

| **Kuo & Fu 2021** | |
|---|---|
| **Study information** | |
| Author | Kuo CP, Fu JS |
| Title | Evaluating the Impact of Mobility on COVID-19 Pandemic with Machine Learning Hybrid Predictions |
| Year | 2021 |
| Journal | Science of The Total Environment |
| DOI | 10.1016/j.scitotenv.2020.144151 |
| Date received | 2020-09-07 |
| Date accepted | 2020-11-24 |
| **Study setting** | |
| Number of populations included | multiple |
| Level of populations included | both national and subnational (172 US counties) |
| Geographic areas covered | North America |
| Number of countries covered | single country (US) |
| Study period | start and end date span first epidemic wave: behavioral |
| **Outcome** | |
| Raw outcome | behavioral outcome (mobility: mobility and engagement index) |
| Time resolution of raw outcome | daily |
| Computed outcome | none |

– continued on next page –

| | |
|---|---|
| Method to obtain the computed outcome | – not applicable – |
| Data source | data from (sub)national authorities (Federal Reserve Bank of Dallas) |
| Data availability | data access via source |
| **Intervention** | |
| Terminology for interventions | measures |
| Terminology for the specific type of non-pharmaceutical interventions | lockdown, control |
| Exposure types | one single intervention (stay-at-home order) |
| Types of single interventions | stay-at-home order |
| Coding of interventions | necessary |
| Source of intervention data | use of externally coded data (New York Times) |
| Availability of data on exposure | access to externally coded data via source |
| **Methodological approach** | |
| Empirical approach | descriptive |
| Use of exposure variation | only variation over time for multiple populations |
| Method | description of change over time (comparison of time periods) |
| Code availability | none |
| **Effect assessment** | |
| Reporting of intervention effect | quantification of change in outcome values |
| Measure of effect | mobility: mobility and engagement index |
| Reporting of uncertainty | no |
| Sensitivity analysis | none |
| Subgroup assessment | none |

| Lau et al. 2020 (Analysis 1) | |
|---|---|
| **Study information** | |
| Author | Lau H, Khosrawipour V, Kocbach P, Mikolajczyk A, Schubert J, Bania J, Khosrawipour T |
| Title | The Positive Impact of Lockdown in Wuhan on Containing the COVID-19 Outbreak in China |
| Year | 2020 |
| Journal | Journal of Travel Medicine |
| DOI | 10.1093/jtm/taaa037 |
| Date received | 2020-02-25 |
| Date accepted | 2020-03-12 |
| **Study setting** | |
| Number of populations included | single |
| Level of populations included | national (China) |
| Geographic areas covered | Asia |
| Number of countries covered | single country (China) |
| Study period | end date at peak of wave |
| **Outcome** | |
| Raw outcome | epidemiological population-level outcome (cases) |
| Time resolution of raw outcome | daily |
| Computed outcome | measure of epidemic trend (doubling time) |
| Method to obtain the computed outcome | exponential growth model |
| Data source | data from publicly available cross-country selections (WHO) |
| Data availability | data access via source |
| **Intervention** | |

– continued on next page –

| | |
|---|---|
| Terminology for interventions | measures |
| Terminology for the specific type of non-pharmaceutical interventions | lockdown |
| Exposure types | one combination of interventions (Wuhan City Lockdown) |
| Types of single interventions | – not applicable – |
| Coding of interventions | not necessary |
| Source of intervention data | – could not be evaluated – |
| Availability of data on exposure | raw data documented in the manuscript |
| **Methodological approach** | |
| Empirical approach | descriptive |
| Use of exposure variation | only variation over time for a single population |
| Method | description of change over time (comparison of time periods) |
| Code availability | none |
| **Effect assessment** | |
| Reporting of intervention effect | comparison of outcome values |
| Measure of effect | doubling time |
| Reporting of uncertainty | yes |
| Sensitivity analysis | none |
| Subgroup assessment | none |

**Lau et al. 2020 (Analysis 2)**

| | |
|---|---|
| **Study information** | |
| Author | Lau H, Khosrawipour V, Kocbach P, Mikolajczyk A, Schubert J, Bania J, Khosrawipour T |

– continued on next page –

| | |
|---|---|
| Title | The Positive Impact of Lockdown in Wuhan on Containing the COVID-19 Outbreak in China |
| Year | 2020 |
| Journal | Journal of Travel Medicine |
| DOI | 10.1093/jtm/taaa037 |
| Date received | 2020-02-25 |
| Date accepted | 2020-03-12 |
| **Study setting** | |
| Number of populations included | single |
| Level of populations included | national (China) |
| Geographic areas covered | Asia |
| Number of countries covered | single country (China) |
| Study period | end date at peak of wave |
| **Outcome** | |
| Raw outcome | epidemiological population-level outcome (cases), behavioral outcome (mobility: domestic air traffic) |
| Time resolution of raw outcome | daily |
| Computed outcome | other (correlation between cases and mobility) |
| Method to obtain the computed outcome | other (correlation analysis) |
| Data source | data from publicly available cross-country selections (WHO: cases), data from (sub)national authorities (Civil Aviation Administration of China: flight data) |
| Data availability | data access via source |
| **Intervention** | |
| Terminology for interventions | measures |

– continued on next page –

| | |
|---|---|
| Terminology for the specific type of non-pharmaceutical interventions | lockdown |
| Exposure types | one combination of interventions (Wuhan City Lockdown) |
| Types of single interventions | – not applicable – |
| Coding of interventions | not necessary |
| Source of intervention data | – could not be evaluated – |
| Availability of data on exposure | raw data documented in the manuscript |
| **Methodological approach** | |
| Empirical approach | descriptive |
| Use of exposure variation | only variation over time for a single population |
| Method | description of change over time (comparison of time periods) |
| Code availability | none |
| **Effect assessment** | |
| Reporting of intervention effect | comparison of outcome values |
| Measure of effect | correlation between cases and mobility |
| Reporting of uncertainty | yes |
| Sensitivity analysis | none |
| Subgroup assessment | none |

**Lee et al. 2020**

| | |
|---|---|
| **Study information** | |
| Author | Lee M, Zhao J, Sun Q, Pan Y, Zhou W, Xiong C, Zhang L |
| Title | Human Mobility Trends during the Early Stage of the COVID-19 Pandemic in the United States |

– continued on next page –

| | |
|---|---|
| Year | 2020 |
| Journal | PLOS ONE |
| DOI | 10.1371/journal.pone.0241468 |
| Date received | 2020-05-29 |
| Date accepted | 2020-10-15 |
| **Study setting** | |
| Number of populations included | multiple |
| Level of populations included | both national and subnational (50 US states) |
| Geographic areas covered | North America |
| Number of countries covered | single country (US) |
| Study period | start and end date span first epidemic wave: behavioral |
| **Outcome** | |
| Raw outcome | behavioral outcome (mobility: percentage staying home & distance traveled & percentage out-of-county trips) |
| Time resolution of raw outcome | daily |
| Computed outcome | none |
| Method to obtain the computed outcome | – not applicable – |
| Data source | data from other research projects (University of Maryland COVID-19 Impact Analysis Platform) |
| Data availability | data access via source |
| **Intervention** | |
| Terminology for interventions | interventions |
| Terminology for the specific type of non-pharmaceutical interventions | non-pharmaceutical |

– continued on next page –

| | |
|---|---|
| Exposure types | one single intervention (national analysis: national emergency declaration, state analysis: stay-at-home order) |
| Types of single interventions | declaration of a state of emergency, stay-at-home order |
| Coding of interventions | necessary |
| Source of intervention data | – could not be evaluated – |
| Availability of data on exposure | coded data not available |
| **Methodological approach** | |
| Empirical approach | descriptive |
| Use of exposure variation | only variation over time for multiple populations |
| Method | description of change over time (comparison of time periods) |
| Code availability | none |
| **Effect assessment** | |
| Reporting of intervention effect | quantification of change in outcome values |
| Measure of effect | mobility: percentage staying home & distance traveled & percentage out-of-county trips |
| Reporting of uncertainty | no |
| Sensitivity analysis | none |
| Subgroup assessment | socioeconomic indicators (income) |

**Lei et al. 2020**

| | |
|---|---|
| **Study information** | |
| Author | Lei H, Wu X, Wang X, Xu M, Xie Y, Du X, Cowling BJ, Li Y, Shu Y |
| Title | Different Transmission Dynamics of Coronavirus Disease 2019 (COVID-19) and Influenza Suggest the Relative Efficiency of Isolation/Quarantine and Social Distancing against COVID-19 in China |

– continued on next page –

| | |
|---|---|
| Year | 2020 |
| Journal | Clinical Infectious Diseases |
| DOI | 10.1093/cid/ciaa1584 |
| Date received | 2020-09-04 |
| Date accepted | 2020-10-12 |
| **Study setting** | |
| Number of populations included | single |
| Level of populations included | national (China) |
| Geographic areas covered | Asia |
| Number of countries covered | single country (China) |
| Study period | start and end date span first epidemic wave |
| **Outcome** | |
| Raw outcome | epidemiological population-level outcome (cases, surrogate disease: cases influenza) |
| Time resolution of raw outcome | daily |
| Computed outcome | epidemiological parameter (reproduction number, reproduction number: influenza) |
| Method to obtain the computed outcome | statistical estimation of reproduction number (Cori et al.) |
| Data source | data from publicly available cross-country selections (WHO (provided by Chinese National Influenza Center)) |
| Data availability | data access via source |
| **Intervention** | |
| Terminology for interventions | interventions |
| Terminology for the specific type of non-pharmaceutical interventions | non-pharmaceutical |

– continued on next page –

| | |
|---|---|
| Exposure types | one combination of interventions (Wuhan City Lockdown) |
| Types of single interventions | – not applicable – |
| Coding of interventions | not necessary |
| Source of intervention data | – could not be evaluated – |
| Availability of data on exposure | raw data documented in the manuscript |
| **Methodological approach** | |
| Empirical approach | parametric |
| Use of exposure variation | only variation over time for a single population |
| Method | non-mechanistic model (generalized linear model) |
| Code availability | none |
| **Effect assessment** | |
| Reporting of intervention effect | quantification of change in outcome values |
| Measure of effect | reproduction number, reproduction number: influenza |
| Reporting of uncertainty | yes |
| Sensitivity analysis | none |
| Subgroup assessment | none |

**Lemaitre et al. 2020 (Analysis 1)**

| | |
|---|---|
| **Study information** | |
| Author | Lemaitre JC, Perez-Saez J, Azman AS, Rinaldo A, Fellay J |
| Title | Assessing the Impact of Non-Pharmaceutical Interventions on SARS-CoV-2 Transmission in Switzerland |
| Year | 2020 |

– continued on next page –

| | |
|---|---|
| Journal | Swiss Medical Weekly |
| DOI | 10.4414/smw.2020.20295 |
| Date received | – could not be evaluated – |
| Date accepted | 2020-05-30 (published) |
| **Study setting** | |
| Number of populations included | multiple |
| Level of populations included | both national and subnational (Swiss cantons) |
| Geographic areas covered | Europe |
| Number of countries covered | single country (Switzerland) |
| Study period | start and end date span first epidemic wave |
| **Outcome** | |
| Raw outcome | epidemiological population-level outcome (hospitalizations, deaths), behavioral outcome (mobility: time spent in various locations) |
| Time resolution of raw outcome | daily |
| Computed outcome | epidemiological parameter (reproduction number), change points (change points: reproduction number, change points: mobility) |
| Method to obtain the computed outcome | compartmental transmission model (single-population), change point analysis |
| Data source | mobility data from corporate organizations (Google) |
| Data availability | data made available by the authors |
| **Intervention** | |
| Terminology for interventions | interventions |
| Terminology for the specific type of non-pharmaceutical interventions | non-pharmaceutical |
| Exposure types | one single intervention (school closure) |
| Types of single interventions | school closure |

– continued on next page –

| | |
|---|---|
| Coding of interventions | not necessary |
| Source of intervention data | – could not be evaluated – |
| Availability of data on exposure | raw data documented in the manuscript |
| **Methodological approach** | |
| Empirical approach | descriptive |
| Use of exposure variation | only variation over time for multiple populations |
| Method | comparison of change points with intervention dates |
| Code availability | publicly available |
| **Effect assessment** | |
| Reporting of intervention effect | quantification of change in outcome values |
| Measure of effect | change points: reproduction number, change points: mobility |
| Reporting of uncertainty | no |
| Sensitivity analysis | none |
| Subgroup assessment | none |

**Lemaitre et al. 2020 (Analysis 2)**

| | |
|---|---|
| **Study information** | |
| Author | Lemaitre JC, Perez-Saez J, Azman AS, Rinaldo A, Fellay J |
| Title | Assessing the Impact of Non-Pharmaceutical Interventions on SARS-CoV-2 Transmission in Switzerland |
| Year | 2020 |
| Journal | Swiss Medical Weekly |
| DOI | 10.4414/smw.2020.20295 |
| Date received | – could not be evaluated – |

– continued on next page –

| | |
|---|---|
| Date accepted | 2020-05-30 (published) |
| **Study setting** | |
| Number of populations included | multiple |
| Level of populations included | both national and subnational (Swiss cantons) |
| Geographic areas covered | Europe |
| Number of countries covered | single country (Switzerland) |
| Study period | start and end date span first epidemic wave |
| **Outcome** | |
| Raw outcome | epidemiological population-level outcome (hospitalizations, deaths) |
| Time resolution of raw outcome | daily |
| Computed outcome | epidemiological parameter (reproduction number), change points (change points: reproduction number) |
| Method to obtain the computed outcome | compartmental transmission model (single-population), change point analysis |
| Data source | data from publicly available open-source projects (openZH) |
| Data availability | data made available by the authors |
| **Intervention** | |
| Terminology for interventions | interventions |
| Terminology for the specific type of non-pharmaceutical interventions | non-pharmaceutical |
| Exposure types | multiple separate interventions (bans of small gatherings, school closure) |
| Types of single interventions | bans of small gatherings, school closure |
| Coding of interventions | not necessary |
| Source of intervention data | – could not be evaluated – |
| Availability of data on exposure | raw data documented in the manuscript |
| **Methodological approach** | |

– continued on next page –

| | |
|---|---|
| Empirical approach | descriptive |
| Use of exposure variation | only variation over time for multiple populations |
| Method | comparison of change points with intervention dates |
| Code availability | publicly available |
| **Effect assessment** | |
| Reporting of intervention effect | comparison of outcome values |
| Measure of effect | reproduction number |
| Reporting of uncertainty | yes |
| Sensitivity analysis | none |
| Subgroup assessment | none |

**Lemaitre et al. 2020 (Analysis 3, mobility as exposure)**

| | |
|---|---|
| **Study information** | |
| Author | Lemaitre JC, Perez-Saez J, Azman AS, Rinaldo A, Fellay J |
| Title | Assessing the Impact of Non-Pharmaceutical Interventions on SARS-CoV-2 Transmission in Switzerland |
| Year | 2020 |
| Journal | Swiss Medical Weekly |
| DOI | 10.4414/smw.2020.20295 |
| Date received | – could not be evaluated – |
| Date accepted | 30.05.2020 (published) |
| **Study setting** | |
| Number of populations included | multiple |
| Level of populations included | both national and subnational (Swiss cantons) |
| Geographic areas covered | Europe |

– continued on next page –

| | |
|---|---|
| Number of countries covered | single country (Switzerland) |
| Study period | start and end date span first epidemic wave |
| **Outcome** | |
| Raw outcome | epidemiological population-level outcome (hospitalized, deaths) |
| Time resolution of raw outcome | daily |
| Computed outcome | epidemiological parameter (reproduction number) |
| Method to obtain the computed outcome | compartmental transmission model (single-population) |
| Data source | data from publicly available open-source projects (openZH) |
| Data availability | data made available by the authors |
| **Intervention** | |
| Terminology for interventions | interventions |
| Terminology for the specific type of non-pharmaceutical interventions | non-pharmaceutical |
| Exposure types | mobility (time spent in various locations) |
| Types of single interventions | – not applicable – |
| Coding of interventions | – not applicable – |
| Source of intervention data | mobility data by corporate organizations (Google) |
| Availability of data on exposure | data access via source |
| **Methodological approach** | |
| Empirical approach | descriptive |
| Use of exposure variation | only variation over time for multiple populations |
| Method | non-mechanistic model (correlation) |
| Code availability | publicly available |
| **Effect assessment** | |
| Reporting of intervention effect | quantification of change in outcome values |

– continued on next page –

| | |
|---|---|
| Measure of effect | reproduction number |
| Reporting of uncertainty | no |
| Sensitivity analysis | none |
| Subgroup assessment | none |

**Li et al. 2020a**

| | |
|---|---|
| **Study information** | |
| Author | Li Y, Zhang R, Zhao J, Molina MJ |
| Title | Understanding Transmission and Intervention for the COVID-19 Pandemic in the United States |
| Year | 2020 |
| Journal | Science of The Total Environment |
| DOI | 10.1016/j.scitotenv.2020.141560 |
| Date received | 2020-07-18 |
| Date accepted | 2020-08-15 |
| **Study setting** | |
| Number of populations included | multiple |
| Level of populations included | subnational (15 US states) |
| Geographic areas covered | North America |
| Number of countries covered | single country (US) |
| Study period | same end date for several populations with diverse epidemic trajectories |
| **Outcome** | |
| Raw outcome | epidemiological population-level outcome (cases) |
| Time resolution of raw outcome | daily |

– continued on next page –

| | |
|---|---|
| Computed outcome | none |
| Method to obtain the computed outcome | – not applicable – |
| Data source | data from (sub)national authorities |
| Data availability | data access via source |
| **Intervention** | |
| Terminology for interventions | measures |
| Terminology for the specific type of non-pharmaceutical interventions | mitigation |
| Exposure types | one single intervention (mask mandate) |
| Types of single interventions | mask mandate |
| Coding of interventions | necessary |
| Source of intervention data | coding done by authors |
| Availability of data on exposure | coded data made available by the authors |
| **Methodological approach** | |
| Empirical approach | counterfactual |
| Use of exposure variation | only variation over time for multiple populations |
| Method | non-mechanistic model (exponential growth model) |
| Code availability | none |
| **Effect assessment** | |
| Reporting of intervention effect | quantification of change in outcome values |
| Measure of effect | cases |
| Reporting of uncertainty | no |
| Sensitivity analysis | none |
| Subgroup assessment | none |

| Li et al. 2020b | |
|---|---|
| **Study information** | |
| Author | Li BZ, Cao NW, Zhou HY, Chu XJ, Ye DQ |
| Title | Strong Policies Control the Spread of COVID-19 in China |
| Year | 2020 |
| Journal | Journal of Medical Virology |
| DOI | 10.1002/jmv.25934 |
| Date received | 2020-03-26 |
| Date accepted | 2020-04-21 |
| **Study setting** | |
| Number of populations included | multiple |
| Level of populations included | subnational (Hubei, Guangdong, henan, Zhejiang, Hunan, Anhui) |
| Geographic areas covered | Asia |
| Number of countries covered | single country (China) |
| Study period | start and end date span first epidemic wave |
| **Outcome** | |
| Raw outcome | epidemiological population-level outcome (cases) |
| Time resolution of raw outcome | daily |
| Computed outcome | none |
| Method to obtain the computed outcome | – not applicable – |
| Data source | data from (sub)national authorities |
| Data availability | data access via source |
| **Intervention** | |
| Terminology for interventions | policies |

– continued on next page –

| | |
|---|---|
| Terminology for the specific type of non-pharmaceutical interventions | intervention |
| Exposure types | all interventions together |
| Types of single interventions | – not applicable – |
| Coding of interventions | – not applicable – |
| Source of intervention data | – not applicable – |
| Availability of data on exposure | – not applicable – |
| **Methodological approach** | |
| Empirical approach | counterfactual |
| Use of exposure variation | only variation over time for multiple populations |
| Method | mechanistic model (compartmental single-population transmission model) |
| Code availability | none |
| **Effect assessment** | |
| Reporting of intervention effect | comparison of outcome values |
| Measure of effect | cases |
| Reporting of uncertainty | no |
| Sensitivity analysis | none |
| Subgroup assessment | none |

**Li et al. 2020c (Analysis 1)**

| | |
|---|---|
| **Study information** | |
| Author | Li Y, Wang LW, Peng ZH, Shen HB |

– continued on next page –

| | |
|---|---|
| Title | Basic Reproduction Number and Predicted Trends of Coronavirus Disease 2019 Epidemic in the Mainland of China |
| Year | 2020 |
| Journal | Infectious Diseases of Poverty |
| DOI | 10.1186/s40249-020-00704-4 |
| Date received | 2020-03-06 |
| Date accepted | 2020-06-18 |
| **Study setting** | |
| Number of populations included | multiple |
| Level of populations included | both national and subnational (China Hubei and outside Hubei) |
| Geographic areas covered | Asia |
| Number of countries covered | single country (China) |
| Study period | start and end date span first epidemic wave |
| **Outcome** | |
| Raw outcome | epidemiological population-level outcome (cases) |
| Time resolution of raw outcome | daily |
| Computed outcome | epidemiological parameter (reproduction number, transmission rate, migration rate) |
| Method to obtain the computed outcome | compartmental transmission model (single-population) |
| Data source | data from (sub)national authorities |
| Data availability | data access via source |
| **Intervention** | |
| Terminology for interventions | measures |
| Terminology for the specific type of non-pharmaceutical interventions | control |

– continued on next page –

| | |
|---|---|
| Exposure types | multiple combinations of interventions (Wuhan lockdown, Hubei lockdown, large-scale case screening) |
| Types of single interventions | – not applicable – |
| Coding of interventions | not necessary |
| Source of intervention data | – could not be evaluated – |
| Availability of data on exposure | raw data documented in the manuscript |
| **Methodological approach** | |
| Empirical approach | descriptive |
| Use of exposure variation | both variation over time and between populations |
| Method | description of change over time (comparison of time periods) |
| Code availability | none |
| **Effect assessment** | |
| Reporting of intervention effect | comparison of outcome values |
| Measure of effect | reproduction number, transmission rate, migration rate |
| Reporting of uncertainty | no |
| Sensitivity analysis | none |
| Subgroup assessment | none |

**Li et al. 2020c (Analysis 2)**

| | |
|---|---|
| **Study information** | |
| Author | Li Y, Wang LW, Peng ZH, Shen HB |
| Title | Basic Reproduction Number and Predicted Trends of Coronavirus Disease 2019 Epidemic in the Mainland of China |
| Year | 2020 |
| Journal | Infectious Diseases of Poverty |

– continued on next page –

| | |
|---|---|
| DOI | 10.1186/s40249-020-00704-4 |
| Date received | 2020-03-06 |
| Date accepted | 2020-06-18 |
| **Study setting** | |
| Number of populations included | multiple |
| Level of populations included | both national and subnational (China Hubei and outside Hubei) |
| Geographic areas covered | Asia |
| Number of countries covered | single country (China) |
| Study period | start and end date span first epidemic wave |
| **Outcome** | |
| Raw outcome | epidemiological population-level outcome (cases) |
| Time resolution of raw outcome | daily |
| Computed outcome | none |
| Method to obtain the computed outcome | – not applicable – |
| Data source | data from (sub)national authorities |
| Data availability | data access via source |
| **Intervention** | |
| Terminology for interventions | measures |
| Terminology for the specific type of non-pharmaceutical interventions | control |
| Exposure types | multiple combinations of interventions (Wuhan lockdown, Hubei lockdown, large-scale case screening) |
| Types of single interventions | – not applicable – |
| Coding of interventions | not necessary |
| Source of intervention data | – could not be evaluated – |

– continued on next page –

| | |
|---|---|
| Availability of data on exposure | raw data documented in the manuscript |
| **Methodological approach** | |
| Empirical approach | counterfactual |
| Use of exposure variation | both variation over time and between populations |
| Method | mechanistic model (compartmental metapopulation transmission model) |
| Code availability | none |
| **Effect assessment** | |
| Reporting of intervention effect | comparison of outcome values |
| Measure of effect | cases |
| Reporting of uncertainty | no |
| Sensitivity analysis | none |
| Subgroup assessment | none |

**Li et al. 2021**

| | |
|---|---|
| **Study information** | |
| Author | Li Y, Campbell H, Kulkarni D, Harpur A, Nundy M, Wang X, Nair H |
| Title | The Temporal Association of Introducing and Lifting Non-Pharmaceutical Interventions with the Time-Varying Reproduction Number (R) of SARS-CoV-2: A Modelling Study across 131 Countries |
| Year | 2021 |
| Journal | The Lancet Infectious Diseases |
| DOI | 10.1016/s1473-3099(20)30785-4 |
| Date received | – could not be evaluated – |
| Date accepted | 2020-10-22 (published) |

– continued on next page –

| | |
|---|---|
| **Study setting** | |
| Number of populations included | multiple |
| Level of populations included | national (133 countries) |
| Geographic areas covered | Europe, Middle East and Africa, Asia, Oceania, Central and South America, North America |
| Number of countries covered | multiple countries |
| Study period | start and end date span first epidemic wave |
| **Outcome** | |
| Raw outcome | epidemiological population-level outcome (effective reproduction number) |
| Time resolution of raw outcome | daily |
| Computed outcome | epidemiological parameter (reproduction number ratio) |
| Method to obtain the computed outcome | simple computation |
| Data source | data from other research projects (EpiForecasts project by the London School of Hygiene & Tropical Medicine or study by Abbott et al.) |
| Data availability | data made available by the authors |
| **Intervention** | |
| Terminology for interventions | interventions |
| Terminology for the specific type of non-pharmaceutical interventions | non-pharmaceutical |
| Exposure types | multiple separate interventions (school closure, bans of large gatherings, bans of small gatherings, workplace closure, stay-at-home order, international travel restrictions, internal movement restrictions, public transport closure) |
| Types of single interventions | school closure, bans of large gatherings, bans of small gatherings, workplace closure, stay-at-home order, international travel restrictions, internal movement restrictions, public transport closure |

– continued on next page –

| | |
|---|---|
| Coding of interventions | necessary |
| Source of intervention data | use of externally coded data (OxCGRT) |
| Availability of data on exposure | coded data made available by the authors |
| **Methodological approach** | |
| Empirical approach | parametric |
| Use of exposure variation | both variation over time and between populations |
| Method | non-mechanistic model (generalized linear model) |
| Code availability | publicly available |
| **Effect assessment** | |
| Reporting of intervention effect | quantification of change in outcome values |
| Measure of effect | reproduction number ratio |
| Reporting of uncertainty | yes |
| Sensitivity analysis | different coding of interventions used (mean effective reproduction number of the last day considered instead of effective reproduction number of the last day), same analysis with (sub)population(s) excluded |
| Subgroup assessment | none |

| **Lim et al. 2020a (Analysis 1)** | |
|---|---|
| **Study information** | |
| Author | Lim JT, Dickens BS, Choo EL, Chew LZ, Koo JR, Tam C, Park M, Cook AR |
| Title | Revealing Regional Disparities in the Transmission Potential of SARS-CoV-2 from Interventions in Southeast Asia |
| Year | 2020 |

– continued on next page –

| | |
|---|---|
| Journal | Proceedings of the Royal Society B: Biological Sciences |
| DOI | 10.1098/rspb.2020.1173 |
| Date received | 2020-05-20 |
| Date accepted | 2020-08-03 |
| **Study setting** | |
| Number of populations included | multiple |
| Level of populations included | national (9 South-east Asian countries) |
| Geographic areas covered | Asia |
| Number of countries covered | multiple countries |
| Study period | same end date for several populations with diverse epidemic trajectories |
| **Outcome** | |
| Raw outcome | epidemiological population-level outcome (cases), behavioral outcome (mobility: time spent in various locations) |
| Time resolution of raw outcome | daily |
| Computed outcome | epidemiological parameter (reproduction number) |
| Method to obtain the computed outcome | statistical estimation of reproduction number (Cori et al., extended with regression on mobility) |
| Data source | data from publicly available cross-country selections (JHU: cases), mobility data from corporate organizations (Google: mobility) |
| Data availability | data access via source |
| **Intervention** | |
| Terminology for interventions | measures |
| Terminology for the specific type of non-pharmaceutical interventions | social distancing |
| Exposure types | all interventions together (timeline of measures) |
| Types of single interventions | – not applicable – |

– continued on next page –

| | |
|---|---|
| Coding of interventions | – not applicable – |
| Source of intervention data | – not applicable – |
| Availability of data on exposure | – not applicable – |
| **Methodological approach** | |
| Empirical approach | descriptive |
| Use of exposure variation | only variation over time for multiple populations |
| Method | description of change over time (comparison of time periods) |
| Code availability | none |
| **Effect assessment** | |
| Reporting of intervention effect | comparison of outcome values |
| Measure of effect | reproduction number, cases, mobility: time spent in various locations |
| Reporting of uncertainty | yes |
| Sensitivity analysis | none |
| Subgroup assessment | none |

**Lim et al. 2020a (Analysis 2, mobility as exposure)**

| | |
|---|---|
| **Study information** | |
| Author | Lim JT, Dickens BS, Choo EL, Chew LZ, Koo JR, Tam C, Park M, Cook AR |
| Title | Revealing Regional Disparities in the Transmission Potential of SARS-CoV-2 from Interventions in Southeast Asia |
| Year | 2020 |
| Journal | Proceedings of the Royal Society B: Biological Sciences |
| DOI | 10.1098/rspb.2020.1173 |
| Date received | 20.05.2020 |

– continued on next page –

| | |
|---|---|
| Date accepted | 03.08.2020 |
| **Study setting** | |
| Number of populations included | multiple |
| Level of populations included | national (9 South-east Asian countries) |
| Geographic areas covered | Asia |
| Number of countries covered | multiple countries |
| Study period | same end date for several populations with diverse epidemic trajectories |
| **Outcome** | |
| Raw outcome | epidemiological population-level outcome (cases) |
| Time resolution of raw outcome | daily |
| Computed outcome | none |
| Method to obtain the computed outcome | – not applicable – |
| Data source | data from publicly available cross-country selections (JHU) |
| Data availability | data access via source |
| **Intervention** | |
| Terminology for interventions | measures |
| Terminology for the specific type of non-pharmaceutical interventions | social distancing |
| Exposure types | mobility (time spent in various locations) |
| Types of single interventions | – not applicable – |
| Coding of interventions | – not applicable – |
| Source of intervention data | mobility data by corporate organizations (Google) |
| Availability of data on exposure | data access via source |
| **Methodological approach** | |
| Empirical approach | parametric |

– continued on next page –

| | |
|---|---|
| Use of exposure variation | only variation over time for multiple populations |
| Method | mechanistic model (semi-mechanistic Bayesian transmission model) |
| Code availability | none |
| **Effect assessment** | |
| Reporting of intervention effect | quantification of change in outcome values |
| Measure of effect | reproduction number |
| Reporting of uncertainty | yes |
| Sensitivity analysis | epidemiological parameters varied |
| Subgroup assessment | none |

**Lim et al. 2020b**

| | |
|---|---|
| **Study information** | |
| Author | Lim JT, Dickens BS, Chew LZ, Choo EL, Koo JR, Aik J, Ng LC, Cook AR |
| Title | Impact of Sars-Cov-2 Interventions on Dengue Transmission |
| Year | 2020 |
| Journal | PLOS Neglected Tropical Diseases |
| DOI | 10.1371/journal.pntd.0008719 |
| Date received | 2020-06-09 |
| Date accepted | 2020-08-16 |
| **Study setting** | |
| Number of populations included | multiple |
| Level of populations included | national (Thailand, Malaysia, Singapore) |
| Geographic areas covered | Asia |
| Number of countries covered | multiple countries |

– continued on next page –

| | |
|---|---|
| Study period | end date could not be evaluated |
| **Outcome** | |
| Raw outcome | epidemiological population-level outcome (surrogate disease: cases dengue) |
| Time resolution of raw outcome | weekly, monthly |
| Computed outcome | none |
| Method to obtain the computed outcome | – not applicable – |
| Data source | data from (sub)national authorities (health authorities: Thailand and Singapore, Administrative Modernisation and Management Planning Unit: Malaysia), data from publicly available cross-country selections (WHO: all) |
| Data availability | data made available by the authors |
| **Intervention** | |
| Terminology for interventions | measures |
| Terminology for the specific type of non-pharmaceutical interventions | social distancing |
| Exposure types | all interventions together (social distancing policy implemented or not, timeline of measures provided) |
| Types of single interventions | – not applicable – |
| Coding of interventions | – not applicable – |
| Source of intervention data | – not applicable – |
| Availability of data on exposure | – not applicable – |
| **Methodological approach** | |
| Empirical approach | parametric |
| Use of exposure variation | only variation over time for multiple populations |
| Method | non-mechanistic model (generalized linear model) |
| Code availability | publicly available |

– continued on next page –

| | |
|---|---|
| **Effect assessment** | |
| Reporting of intervention effect | quantification of change in outcome values |
| Measure of effect | cases: dengue |
| Reporting of uncertainty | yes |
| Sensitivity analysis | model specification varied |
| Subgroup assessment | none |

| **Linka et al. 2020 (Analysis 1)** | |
|---|---|
| **Study information** | |
| Author | Linka K, Peirlinck M, Kuhl E |
| Title | The Reproduction Number of COVID-19 and Its Correlation with Public Health Interventions |
| Year | 2020 |
| Journal | Computational Mechanics |
| DOI | 10.1007/s00466-020-01880-8 |
| Date received | 2020-05-04 |
| Date accepted | 2020-07-06 |
| **Study setting** | |
| Number of populations included | multiple |
| Level of populations included | national (all 27 EU countries) |
| Geographic areas covered | Europe |
| Number of countries covered | multiple countries |
| Study period | start and end date span first epidemic wave |

– continued on next page –

| | |
|---|---|
| **Outcome** | |
| Raw outcome | epidemiological population-level outcome (cases) |
| Time resolution of raw outcome | daily |
| Computed outcome | epidemiological parameter (reproduction number) |
| Method to obtain the computed outcome | compartmental transmission model (single-population) |
| Data source | data from publicly available cross-country selections (ECDC) |
| Data availability | data access via source |
| **Intervention** | |
| Terminology for interventions | interventions |
| Terminology for the specific type of non-pharmaceutical interventions | public health |
| Exposure types | all interventions together |
| Types of single interventions | – not applicable – |
| Coding of interventions | – not applicable – |
| Source of intervention data | – not applicable – |
| Availability of data on exposure | – not applicable – |
| **Methodological approach** | |
| Empirical approach | descriptive |
| Use of exposure variation | only variation over time for multiple populations |
| Method | description of change over time (comparison of time periods) |
| Code availability | none |
| **Effect assessment** | |
| Reporting of intervention effect | comparison of outcome values |
| Measure of effect | reproduction number |
| Reporting of uncertainty | yes |

– continued on next page –

| | |
|---|---|
| Sensitivity analysis | model specification varied |
| Subgroup assessment | none |

| **Linka et al. 2020 (Analysis 2, mobility as exposure)** | |
|---|---|
| **Study information** | |
| Author | Linka K, Peirlinck M, Kuhl E |
| Title | The Reproduction Number of COVID-19 and Its Correlation with Public Health Interventions |
| Year | 2020 |
| Journal | Computational Mechanics |
| DOI | 10.1007/s00466-020-01880-8 |
| Date received | 04.05.2020 |
| Date accepted | 06.07.2020 |
| **Study setting** | |
| Number of populations included | multiple |
| Level of populations included | national (all 27 EU countries) |
| Geographic areas covered | Europe |
| Number of countries covered | multiple countries |
| Study period | start and end date span first epidemic wave |
| **Outcome** | |
| Raw outcome | epidemiological population-level outcome (cases) |
| Time resolution of raw outcome | daily |
| Computed outcome | epidemiological parameter (reproduction number) |
| Method to obtain the computed outcome | compartmental transmission model (single-population) |

– continued on next page –

| | |
|---|---|
| Data source | data from publicly available cross-country selections (ECDC) |
| Data availability | data access via source |
| **Intervention** | |
| Terminology for interventions | interventions |
| Terminology for the specific type of non-pharmaceutical interventions | public health |
| Exposure types | mobility (air travel data, number of requests for directions in Maps) |
| Types of single interventions | – not applicable – |
| Coding of interventions | – not applicable – |
| Source of intervention data | mobility data by corporate organizations (Eurocontrol: air travel data, Apple mobility: number of trips by mode) |
| Availability of data on exposure | data access via source |
| **Methodological approach** | |
| Empirical approach | descriptive |
| Use of exposure variation | only variation over time for multiple populations |
| Method | non-mechanistic model (correlation) |
| Code availability | none |
| **Effect assessment** | |
| Reporting of intervention effect | quantification of change in outcome values |
| Measure of effect | reproduction number |
| Reporting of uncertainty | no |
| Sensitivity analysis | none |
| Subgroup assessment | none |

| Liu et al. 2020a | |
|---|---|
| **Study information** | |
| Author | Liu PY, He S, Rong LB, Tang SY |
| Title | The Effect of Control Measures on COVID-19 Transmission in Italy: Comparison with Guangdong Province in China |
| Year | 2020 |
| Journal | Infectious Diseases of Poverty |
| DOI | 10.1186/s40249-020-00730-2 |
| Date received | 2020-04-01 |
| Date accepted | 2020-07-22 |
| **Study setting** | |
| Number of populations included | single |
| Level of populations included | national (Italy) |
| Geographic areas covered | Europe |
| Number of countries covered | single country (Italy) |
| Study period | end date in growth phase of wave |
| **Outcome** | |
| Raw outcome | epidemiological population-level outcome (cases, deaths, recovered cases) |
| Time resolution of raw outcome | daily |
| Computed outcome | summary statistic (initial contact rate, prevalence ratio: cases, cure ratio: recovered, time until 1000 cases) |
| Method to obtain the computed outcome | simple computation (ratios), compartmental transmission model (compartmental single-population transmission model: initial contact rate) |
| Data source | data from (sub)national authorities (Italy and Guangdong) |
| Data availability | data access via source |
| **Intervention** | |
| Terminology for interventions | measures |

– continued on next page –

| | |
|---|---|
| Terminology for the specific type of non-pharmaceutical interventions | control |
| Exposure types | all interventions together (timeline of measures) |
| Types of single interventions | – not applicable – |
| Coding of interventions | – not applicable – |
| Source of intervention data | – not applicable – |
| Availability of data on exposure | – not applicable – |
| **Methodological approach** | |
| Empirical approach | descriptive |
| Use of exposure variation | only variation over time for a single population |
| Method | comparison of populations |
| Code availability | none |
| **Effect assessment** | |
| Reporting of intervention effect | comparison of outcome values |
| Measure of effect | contact rate: initial contact rate, cases, recovered, time until 1000 cases |
| Reporting of uncertainty | no |
| Sensitivity analysis | none |
| Subgroup assessment | none |

**Liu et al. 2020b (Analysis 1)**

| | |
|---|---|
| **Study information** | |
| Author | Liu K, Ai S, Song S, Zhu G, Tian F, Li H, Gao Y, Wu Y, Zhang S, Shao Z, Liu Q, Lin H |

– continued on next page –

| | |
|---|---|
| Title | Population Movement, City Closure in Wuhan, and Geographical Expansion of the COVID-19 Infection in China in January 2020 |
| Year | 2020 |
| Journal | Clinical Infectious Diseases |
| DOI | 10.1093/cid/ciaa422 |
| Date received | 2020-02-11 |
| Date accepted | 2020-04-17 |
| **Study setting** | |
| Number of populations included | multiple |
| Level of populations included | both national and subnational (Chinese provinces) |
| Geographic areas covered | Asia |
| Number of countries covered | single country (China) |
| Study period | start and end date span first epidemic wave: behavioral |
| **Outcome** | |
| Raw outcome | behavioral outcome (mobility: population outflow from Wuhan) |
| Time resolution of raw outcome | daily |
| Computed outcome | none |
| Method to obtain the computed outcome | – not applicable – |
| Data source | mobility data from corporate organizations (Baidu) |
| Data availability | data access via source |
| **Intervention** | |
| Terminology for interventions | measures |
| Terminology for the specific type of non-pharmaceutical interventions | prevention, control |
| Exposure types | one combination of interventions (Wuhan City Lockdown) |

– continued on next page –

| | |
|---|---|
| Types of single interventions | – not applicable – |
| Coding of interventions | not necessary |
| Source of intervention data | – could not be evaluated – |
| Availability of data on exposure | raw data documented in the manuscript |
| **Methodological approach** | |
| Empirical approach | descriptive |
| Use of exposure variation | only variation over time for multiple populations |
| Method | description of change over time (comparison of time periods) |
| Code availability | none |
| **Effect assessment** | |
| Reporting of intervention effect | comparison of outcome values |
| Measure of effect | mobility: population outflow from Wuhan |
| Reporting of uncertainty | no |
| Sensitivity analysis | none |
| Subgroup assessment | none |

**Liu et al. 2020b (Analysis 2, mobility as exposure)**

| | |
|---|---|
| **Study information** | |
| Author | Liu K, Ai S, Song S, Zhu G, Tian F, Li H, Gao Y, Wu Y, Zhang S, Shao Z, Liu Q, Lin H |
| Title | Population Movement, City Closure in Wuhan, and Geographical Expansion of the COVID-19 Infection in China in January 2020 |
| Year | 2020 |
| Journal | Clinical Infectious Diseases |
| DOI | 10.1093/cid/ciaa422 |

– continued on next page –

| | |
|---|---|
| Date received | 11.02.2020 |
| Date accepted | 17.04.2020 |
| **Study setting** | |
| Number of populations included | multiple |
| Level of populations included | both national and subnational (Chinese provinces) |
| Geographic areas covered | Asia |
| Number of countries covered | single country (China) |
| Study period | end date in growth phase of wave |
| **Outcome** | |
| Raw outcome | epidemiological population-level outcome (cases) |
| Time resolution of raw outcome | daily |
| Computed outcome | none |
| Method to obtain the computed outcome | – not applicable – |
| Data source | data from (sub)national authorities |
| Data availability | data access via source |
| **Intervention** | |
| Terminology for interventions | measures |
| Terminology for the specific type of non-pharmaceutical interventions | prevention, control |
| Exposure types | mobility (population outflow from Wuhan) |
| Types of single interventions | – not applicable – |
| Coding of interventions | – not applicable – |
| Source of intervention data | mobility data by corporate organizations (Baidu) |
| Availability of data on exposure | data access via source |
| **Methodological approach** | |

– continued on next page –

| | |
|---|---|
| Empirical approach | counterfactual |
| Use of exposure variation | only variation between populations |
| Method | other |
| Code availability | none |
| **Effect assessment** | |
| Reporting of intervention effect | quantification of change in outcome values |
| Measure of effect | cases |
| Reporting of uncertainty | yes |
| Sensitivity analysis | none |
| Subgroup assessment | none |

**Loeffler-Wirth et al. 2020**

| | |
|---|---|
| **Study information** | |
| Author | Loeffler-Wirth H, Schmidt M, Binder H |
| Title | Covid-19 Transmission Trajectories–Monitoring the Pandemic in the World-wide Context |
| Year | 2020 |
| Journal | Viruses |
| DOI | 10.3390/v12070777 |
| Date received | 2020-06-30 |
| Date accepted | 2020-07-14 |
| **Study setting** | |
| Number of populations included | multiple |
| Level of populations included | both national and subnational (187 countries, 17 German states) |

– continued on next page –

| | |
|---|---|
| Geographic areas covered | Europe, Middle East and Africa, Asia, Oceania, Central and South America, North America |
| Number of countries covered | multiple countries |
| Study period | same end date for several populations with diverse epidemic trajectories |
| **Outcome** | |
| Raw outcome | epidemiological population-level outcome (cases) |
| Time resolution of raw outcome | daily |
| Computed outcome | epidemiological parameter (reproduction number) |
| Method to obtain the computed outcome | compartmental transmission model (single-population) |
| Data source | data from publicly available cross-country selections (JHU: all other countries), data from (sub)national authorities (Germany) |
| Data availability | data access via source |
| **Intervention** | |
| Terminology for interventions | interventions |
| Terminology for the specific type of non-pharmaceutical interventions | non-pharmaceutical |
| Exposure types | all interventions together |
| Types of single interventions | – not applicable – |
| Coding of interventions | – not applicable – |
| Source of intervention data | – not applicable – |
| Availability of data on exposure | – not applicable – |
| **Methodological approach** | |
| Empirical approach | descriptive |
| Use of exposure variation | only variation over time for multiple populations |
| Method | description of change over time (description of time course) |

– continued on next page –

| | |
|---|---|
| Code availability | none |
| **Effect assessment** | |
| Reporting of intervention effect | qualitative statement |
| Measure of effect | cases, reproduction number |
| Reporting of uncertainty | – not applicable – |
| Sensitivity analysis | none |
| Subgroup assessment | none |

| **Lurie et al. 2020 (Analysis 1)** | |
|---|---|
| **Study information** | |
| Author | Lurie MN, Silva J, Yorlets RR, Tao J, Chan PA |
| Title | Coronavirus Disease 2019 Epidemic Doubling Time in the United States before and during Stay-at-Home Restrictions |
| Year | 2020 |
| Journal | The Journal of Infectious Diseases |
| DOI | 10.1093/infdis/jiaa491 |
| Date received | 2020-06-15 |
| Date accepted | 2020-07-30 |
| **Study setting** | |
| Number of populations included | multiple |
| Level of populations included | both national and subnational (US and its states) |
| Geographic areas covered | North America |
| Number of countries covered | single country (US) |

– continued on next page –

| | |
|---|---|
| Study period | same end date for several populations with diverse epidemic trajectories |
| **Outcome** | |
| Raw outcome | epidemiological population-level outcome (cases) |
| Time resolution of raw outcome | daily |
| Computed outcome | measure of epidemic trend (doubling time) |
| Method to obtain the computed outcome | simple computation |
| Data source | data from publicly available cross-country selections (JHU) |
| Data availability | data access via source |
| **Intervention** | |
| Terminology for interventions | strategies |
| Terminology for the specific type of non-pharmaceutical interventions | mitigation |
| Exposure types | one single intervention (stay-at-home order) |
| Types of single interventions | stay-at-home order |
| Coding of interventions | necessary |
| Source of intervention data | use of externally coded data (New York Times, OxCGRT (for national level)) |
| Availability of data on exposure | access to externally coded data via source |
| **Methodological approach** | |
| Empirical approach | descriptive |
| Use of exposure variation | only variation over time for multiple populations |
| Method | description of change over time (comparison of time periods) |
| Code availability | none |
| **Effect assessment** | |
| Reporting of intervention effect | quantification of change in outcome values |

– continued on next page –

| | |
|---|---|
| Measure of effect | doubling time |
| Reporting of uncertainty | yes |
| Sensitivity analysis | none |
| Subgroup assessment | none |

| **Lurie et al. 2020 (Analysis 2)** | |
|---|---|
| **Study information** | |
| Author | Lurie MN, Silva J, Yorlets RR, Tao J, Chan PA |
| Title | Coronavirus Disease 2019 Epidemic Doubling Time in the United States before and during Stay-at-Home Restrictions |
| Year | 2020 |
| Journal | The Journal of Infectious Diseases |
| DOI | 10.1093/infdis/jiaa491 |
| Date received | 2020-06-15 |
| Date accepted | 2020-07-30 |
| **Study setting** | |
| Number of populations included | single |
| Level of populations included | national (US) |
| Geographic areas covered | North America |
| Number of countries covered | single country (US) |
| Study period | start and end date span first epidemic wave |
| **Outcome** | |
| Raw outcome | epidemiological population-level outcome (cases) |
| Time resolution of raw outcome | daily |
| Computed outcome | measure of epidemic trend (doubling time) |

– continued on next page –

| | |
|---|---|
| Method to obtain the computed outcome | simple computation |
| Data source | data from publicly available cross-country selections (JHU) |
| Data availability | data access via source |
| **Intervention** | |
| Terminology for interventions | strategies |
| Terminology for the specific type of non-pharmaceutical interventions | mitigation |
| Exposure types | stringency index (OxCGRT Stringency Index) |
| Types of single interventions | – not applicable – |
| Coding of interventions | necessary |
| Source of intervention data | use of externally coded data (OxCGRT) |
| Availability of data on exposure | access to externally coded data via source |
| **Methodological approach** | |
| Empirical approach | descriptive |
| Use of exposure variation | only variation over time for a single population |
| Method | description of change over time (comparison of time periods) |
| Code availability | none |
| **Effect assessment** | |
| Reporting of intervention effect | quantification of change in outcome values |
| Measure of effect | doubling time |
| Reporting of uncertainty | yes |
| Sensitivity analysis | none |
| Subgroup assessment | none |

| Maier & Brockmann 2020 | |
|---|---|
| **Study information** | |
| Author | Maier BF, Brockmann D |
| Title | Effective Containment Explains Subexponential Growth in Recent Confirmed COVID-19 Cases in China |
| Year | 2020 |
| Journal | Science |
| DOI | 10.1126/science.abb4557 |
| Date received | 2020-02-25 |
| Date accepted | 2020-04-04 |
| **Study setting** | |
| Number of populations included | multiple |
| Level of populations included | both national and subnational (Hubei + 28 other provinces (China)) |
| Geographic areas covered | Asia |
| Number of countries covered | single country (China) |
| Study period | end date at peak of wave |
| **Outcome** | |
| Raw outcome | epidemiological population-level outcome (cases) |
| Time resolution of raw outcome | daily |
| Computed outcome | none |
| Method to obtain the computed outcome | – not applicable – |
| Data source | data from publicly available cross-country selections (JHU) |
| Data availability | data made available by the authors |
| **Intervention** | |
| Terminology for interventions | measures |

– continued on next page –

| | |
|---|---|
| Terminology for the specific type of non-pharmaceutical interventions | containment |
| Exposure types | categories of interventions (containment and quarantining) |
| Types of single interventions | – not applicable – |
| Coding of interventions | – not applicable – |
| Source of intervention data | – not applicable – |
| Availability of data on exposure | – not applicable – |
| **Methodological approach** | |
| Empirical approach | parametric |
| Use of exposure variation | only variation over time for multiple populations |
| Method | mechanistic model (compartmental single-population transmission model) |
| Latent variable functionally linked to intervention | susceptible decay |
| Code availability | publicly available |
| **Effect assessment** | |
| Reporting of intervention effect | quantification of change in outcome values |
| Measure of effect | reproduction number |
| Reporting of uncertainty | no |
| Sensitivity analysis | epidemiological parameters varied |
| Subgroup assessment | none |

**Malheiro et al. 2020**

| | |
|---|---|
| **Study information** | |
| Author | Malheiro R, Figueiredo AL, Magalhães JP, Teixeira P, Moita I, Moutinho MC, Mansilha RB, Gonçalves LM, Ferreira E |

– continued on next page –

| | |
|---|---|
| Title | Effectiveness of Contact Tracing and Quarantine on Reducing COVID-19 Transmission: A Retrospective Cohort Study |
| Year | 2020 |
| Journal | Public Health |
| DOI | 10.1016/j.puhe.2020.09.012 |
| Date received | 2020-07-03 |
| Date accepted | 2020-09-16 |
| **Study setting** | |
| Number of populations included | single |
| Level of populations included | subnational (Eastern Porto (Portugal)) |
| Geographic areas covered | Europe |
| Number of countries covered | single country (Portugal) |
| Study period | start and end date span first epidemic wave |
| **Outcome** | |
| Raw outcome | epidemiological individual-level outcome (individual cases and transmission chains) |
| Time resolution of raw outcome | daily |
| Computed outcome | epidemiological parameter (secondary cases per index case, proportion of index cases with secondary cases, time between symptom onset and testing, number of close contacts) |
| Method to obtain the computed outcome | simple computation |
| Data source | data from (sub)national authorities (National Epidemiological Surveillance System and protected databases used for the daily monitoring and registration of close contacts for follow-up) |
| Data availability | data not accessible |
| **Intervention** | |

– continued on next page –

| | |
|---|---|
| Terminology for interventions | measures |
| Terminology for the specific type of non-pharmaceutical interventions | lockdown |
| Exposure types | one combination of interventions (state of emergency as decreed by the Portugese government on March 18 (lockdown measures, including constraints in public circulation, closure of commercial activities, and resort to homeworking)) |
| Types of single interventions | – not applicable – |
| Coding of interventions | not necessary |
| Source of intervention data | government or news websites |
| Availability of data on exposure | raw data documented in the manuscript |
| **Methodological approach** | |
| Empirical approach | descriptive |
| Use of exposure variation | only variation over time for a single population |
| Method | description of change over time (comparison of time periods) |
| Code availability | none |
| **Effect assessment** | |
| Reporting of intervention effect | quantification of change in outcome values |
| Measure of effect | secondary cases per index case, proportion of index cases with secondary cases, time between symptom onset and testing, number of contacts |
| Reporting of uncertainty | yes |
| Sensitivity analysis | none |
| Subgroup assessment | public health response (subject to other measures: contact tracing and quarantine measures) |

| Manevski et al. 2020 | |
|---|---|
| **Study information** | |
| Author | Manevski D, Gorenjec NR, Kejžar N, Blagus R |
| Title | Modeling COVID-19 Pandemic Using Bayesian Analysis with Application to Slovene Data |
| Year | 2020 |
| Journal | Mathematical Biosciences |
| DOI | 10.1016/j.mbs.2020.108466 |
| Date received | 2020-07-08 |
| Date accepted | 2020-09-05 |
| **Study setting** | |
| Number of populations included | single |
| Level of populations included | national (Slovenia) |
| Geographic areas covered | Europe |
| Number of countries covered | single country (Slovenia) |
| Study period | start and end date span first epidemic wave |
| **Outcome** | |
| Raw outcome | epidemiological population-level outcome (cases, deaths, hospitalizations, ICU hospitalizations) |
| Time resolution of raw outcome | daily |
| Computed outcome | none |
| Method to obtain the computed outcome | – not applicable – |
| Data source | data from publicly available open-source projects (COVID-19 Tracker Slovenia) |
| Data availability | data made available by the authors |
| **Intervention** | |

– continued on next page –

| | |
|---|---|
| Terminology for interventions | interventions |
| Terminology for the specific type of non-pharmaceutical interventions | non-pharmaceutical |
| Exposure types | all interventions together (event bans, lockdown, prohibition of movement outside municipality of residence) |
| Types of single interventions | – not applicable – |
| Coding of interventions | – not applicable – |
| Source of intervention data | – not applicable – |
| Availability of data on exposure | – not applicable – |
| **Methodological approach** | |
| Empirical approach | parametric |
| Use of exposure variation | only variation over time for a single population |
| Method | mechanistic model (semi-mechanistic Bayesian transmission model) |
| Latent variable functionally linked to intervention | reproduction number |
| Code availability | publicly available |
| **Effect assessment** | |
| Reporting of intervention effect | comparison of outcome values |
| Measure of effect | reproduction number |
| Reporting of uncertainty | yes |
| Sensitivity analysis | different or modified outcome used (only deaths used), model specification varied, epidemiological parameters varied |
| Subgroup assessment | none |

| Marschner 2020 | |
|---|---|
| **Study information** | |
| Author | Marschner IC |
| Title | Back-Projection of COVID-19 Diagnosis Counts to Assess Infection Incidence and Control Measures: Analysis of Australian Data |
| Year | 2020 |
| Journal | Epidemiology and Infection |
| DOI | 10.1017/s0950268820001065 |
| Date received | 2020-04-26 |
| Date accepted | 2020-05-11 |
| **Study setting** | |
| Number of populations included | single |
| Level of populations included | national (Australia) |
| Geographic areas covered | Oceania |
| Number of countries covered | single country (Australia) |
| Study period | start and end date span first epidemic wave |
| **Outcome** | |
| Raw outcome | epidemiological population-level outcome (cases) |
| Time resolution of raw outcome | daily |
| Computed outcome | other (infections) |
| Method to obtain the computed outcome | back projection |
| Data source | data from (sub)national authorities |
| Data availability | data made available by the authors |
| **Intervention** | |
| Terminology for interventions | measures |

– continued on next page –

| | |
|---|---|
| Terminology for the specific type of non-pharmaceutical interventions | control |
| Exposure types | phases of interventions (border closure and three stages of social distancing measures) |
| Types of single interventions | – not applicable – |
| Coding of interventions | not necessary |
| Source of intervention data | government or news websites |
| Availability of data on exposure | raw data documented in the manuscript |
| **Methodological approach** | |
| Empirical approach | descriptive |
| Use of exposure variation | only variation over time for a single population |
| Method | description of change over time (description of time course) |
| Code availability | publicly available |
| **Effect assessment** | |
| Reporting of intervention effect | qualitative statement |
| Measure of effect | infections |
| Reporting of uncertainty | – not applicable – |
| Sensitivity analysis | different or modified outcome used (undetected cases adjusted for), epidemiological parameters varied |
| Subgroup assessment | none |

**McCarthy et al. 2020**

| | |
|---|---|
| **Study information** | |
| Author | McCarthy Z, Xiao Y, Scarabel F, Tang B, Bragazzi NL, Nah K, Heffernan JM, Asgary A, Murty VK, Ogden NH, Wu J |

– continued on next page –

| | |
|---|---|
| Title | Quantifying the Shift in Social Contact Patterns in Response to Non-Pharmaceutical Interventions |
| Year | 2020 |
| Journal | Journal of Mathematics in Industry |
| DOI | 10.1186/s13362-020-00096-y |
| Date received | 2020-10-15 |
| Date accepted | 2020-11-25 |
| **Study setting** | |
| Number of populations included | single |
| Level of populations included | subnational (Ontario (Canada)) |
| Geographic areas covered | North America |
| Number of countries covered | single country (Canada) |
| Study period | start and end date span first epidemic wave |
| **Outcome** | |
| Raw outcome | epidemiological population-level outcome (cases) |
| Time resolution of raw outcome | daily |
| Computed outcome | none |
| Method to obtain the computed outcome | – not applicable – |
| Data source | data from other research projects (publicly funded research project: contact matrices), data from (sub)national authorities (cases) |
| Data availability | data access via source |
| **Intervention** | |
| Terminology for interventions | measures |
| Terminology for the specific type of non-pharmaceutical interventions | physical distancing |

– continued on next page –

| | |
|---|---|
| Exposure types | multiple combinations of interventions (public school closure (from March 14), declaration of state of emergency, with closure of public venues and events and physical distancing advisories (from March 18), and closure of non-essential establishments (from March 24)) |
| Types of single interventions | – not applicable – |
| Coding of interventions | not necessary |
| Source of intervention data | government or news websites |
| Availability of data on exposure | raw data documented in the manuscript |
| **Methodological approach** | |
| Empirical approach | parametric |
| Use of exposure variation | only variation over time for a single population |
| Method | mechanistic model (compartmental single-population transmission model) |
| Latent variable functionally linked to intervention | contact matrix |
| Code availability | none |
| **Effect assessment** | |
| Reporting of intervention effect | quantification of change in outcome values |
| Measure of effect | reproduction number, contact rate, contact matrix |
| Reporting of uncertainty | no |
| Sensitivity analysis | epidemiological parameters varied |
| Subgroup assessment | socioeconomic indicators (age), epidemiological indicators (infection setting) |

**McGrail et al. 2020**

| | |
|---|---|
| **Study information** | |
| Author | McGrail DJ, Dai J, McAndrews KM, Kalluri R |

– continued on next page –

| | |
|---|---|
| Title | Enacting National Social Distancing Policies Corresponds with Dramatic Reduction in COVID19 Infection Rates |
| Year | 2020 |
| Journal | PLOS ONE |
| DOI | 10.1371/journal.pone.0236619 |
| Date received | 2020-05-01 |
| Date accepted | 2020-07-10 |
| **Study setting** | |
| Number of populations included | multiple |
| Level of populations included | both national and subnational (50 US states, 120 countries) |
| Geographic areas covered | Europe, Middle East and Africa, Asia, Oceania, Central and South America, North America |
| Number of countries covered | multiple countries |
| Study period | same end date for several populations with diverse epidemic trajectories |
| **Outcome** | |
| Raw outcome | epidemiological population-level outcome (cases), behavioral outcome (mobility: average reduction in time spent in various locations) |
| Time resolution of raw outcome | daily |
| Computed outcome | measure of epidemic trend (growth rate) |
| Method to obtain the computed outcome | exponential growth model |
| Data source | data from publicly available cross-country selections (JHU) |
| Data availability | data access via source |
| **Intervention** | |
| Terminology for interventions | measures |

– continued on next page –

| | |
|---|---|
| Terminology for the specific type of non-pharmaceutical interventions | social distancing |
| Exposure types | one single intervention (workplace closure) |
| Types of single interventions | workplace closure |
| Coding of interventions | necessary |
| Source of intervention data | use of externally coded data (Aura Vision Global Covid-19 Lockdown Tracker) |
| Availability of data on exposure | coded data made available by the authors |
| **Methodological approach** | |
| Empirical approach | descriptive |
| Use of exposure variation | both variation over time and between populations |
| Method | description of change over time (comparison of time periods) |
| Code availability | none |
| **Effect assessment** | |
| Reporting of intervention effect | quantification of change in outcome values |
| Measure of effect | growth rate, mobility: average reduction in time spent in various locations |
| Reporting of uncertainty | yes |
| Sensitivity analysis | start or end date of study period varied |
| Subgroup assessment | none |

**McKee et al. 2020**

| | |
|---|---|
| **Study information** | |
| Author | McKee KL, Crandell IC, Hanlon AL |
| Title | County-Level Social Distancing and Policy Impact in the United States: A Dynamical Systems Model |

– continued on next page –

| | |
|---|---|
| Year | 2020 |
| Journal | JMIR Public Health and Surveillance |
| DOI | 10.2196/23902 |
| Date received | 2020-09-10 |
| Date accepted | 2020-11-27 |
| **Study setting** | |
| Number of populations included | multiple |
| Level of populations included | both national and subnational (3050 counties in the US) |
| Geographic areas covered | North America |
| Number of countries covered | single country (US) |
| Study period | start and end date span first epidemic wave: behavioral |
| **Outcome** | |
| Raw outcome | behavioral outcome (mobility: distance traveled & visitation to key sites & log number of interpersonal encounters) |
| Time resolution of raw outcome | daily |
| Computed outcome | none |
| Method to obtain the computed outcome | – not applicable – |
| Data source | mobility data from corporate organizations (Unacast) |
| Data availability | data access via source |
| **Intervention** | |
| Terminology for interventions | measures |
| Terminology for the specific type of non-pharmaceutical interventions | social distancing |
| Exposure types | multiple separate interventions (stay-at-home order, bans of small gatherings, school closure, workplace closure) |

– continued on next page –

| | |
|---|---|
| Types of single interventions | stay-at-home order, bans of small gatherings, school closure, workplace closure |
| Coding of interventions | necessary |
| Source of intervention data | use of externally coded data (Kaiser Fung Family) |
| Availability of data on exposure | access to externally coded data via source |
| **Methodological approach** | |
| Empirical approach | parametric |
| Use of exposure variation | both variation over time and between populations |
| Method | other (differential equation-based state-space model to estimate effects of interventions on mobility) |
| Code availability | none |
| **Effect assessment** | |
| Reporting of intervention effect | quantification of change in outcome values |
| Measure of effect | mobility: distance traveled & visitation to key sites & log number of interpersonal encounters |
| Reporting of uncertainty | yes |
| Sensitivity analysis | none |
| Subgroup assessment | none |

**McKenzie & Adams 2020**

| | |
|---|---|
| **Study information** | |
| Author | McKenzie G, Adams B |
| Title | A Country Comparison of Place-Based Activity Response to COVID-19 Policies |
| Year | 2020 |

– continued on next page –

| | |
|---|---|
| Journal | Applied Geography |
| DOI | 10.1016/j.apgeog.2020.102363 |
| Date received | 2020-05-18 |
| Date accepted | 2020-10-26 |
| **Study setting** | |
| Number of populations included | multiple |
| Level of populations included | both national and subnational (108 countries around the globe, 46 with subnational regions) |
| Geographic areas covered | Europe, Middle East and Africa, Asia, Oceania, Central and South America, North America |
| Number of countries covered | multiple countries |
| Study period | start and end date span first epidemic wave: behavioral |
| **Outcome** | |
| Raw outcome | behavioral outcome (mobility: time spent in various locations) |
| Time resolution of raw outcome | daily |
| Computed outcome | none |
| Method to obtain the computed outcome | – not applicable – |
| Data source | mobility data from corporate organizations (Google) |
| Data availability | data made available by the authors |
| **Intervention** | |
| Terminology for interventions | responses |
| Terminology for the specific type of non-pharmaceutical interventions | government |
| Exposure types | stringency index (OxCGRT Stringency Index) |
| Types of single interventions | – not applicable – |

– continued on next page –

| | |
|---|---|
| Coding of interventions | necessary |
| Source of intervention data | use of externally coded data (OxCGRT) |
| Availability of data on exposure | coded data made available by the authors |
| **Methodological approach** | |
| Empirical approach | parametric |
| Use of exposure variation | only variation over time for multiple populations |
| Method | non-mechanistic model (correlation) |
| Code availability | publicly available |
| **Effect assessment** | |
| Reporting of intervention effect | quantification of change in outcome values |
| Measure of effect | mobility: time spent in various locations |
| Reporting of uncertainty | yes |
| Sensitivity analysis | none |
| Subgroup assessment | geographic areas (regions) |

| **Medline et al. 2020** | |
|---|---|
| **Study information** | |
| Author | Medline A, Hayes L, Valdez K, Hayashi A, Vahedi F, Capell W, Sonnenberg J, Glick Z, Klausner JD |
| Title | Evaluating the Impact of Stay-at-Home Orders on the Time to Reach the Peak Burden of COVID-19 Cases and Deaths: Does Timing Matter? |
| Year | 2020 |
| Journal | BMC Public Health |
| DOI | 10.1186/s12889-020-09817-9 |

– continued on next page –

| | |
|---|---|
| Date received | 2020-06-20 |
| Date accepted | 2020-11-02 |
| **Study setting** | |
| Number of populations included | multiple |
| Level of populations included | both national and subnational (41 countries, US and its states) |
| Geographic areas covered | Europe, Middle East and Africa, Asia, Oceania, Central and South America, North America |
| Number of countries covered | multiple countries |
| Study period | end date could not be evaluated |
| **Outcome** | |
| Raw outcome | epidemiological population-level outcome (cases, deaths) |
| Time resolution of raw outcome | daily |
| Computed outcome | summary statistic (time from first case to peak of cases/deaths) |
| Method to obtain the computed outcome | simple computation |
| Data source | data from publicly available cross-country selections (WHO: all countries, Worldometer: all countries), data from (sub)national authorities (US) |
| Data availability | data access via source |
| **Intervention** | |
| Terminology for interventions | interventions |
| Terminology for the specific type of non-pharmaceutical interventions | non-pharmaceutical |
| Exposure types | one single intervention (stay-at-home order) |
| Types of single interventions | stay-at-home order |
| Coding of interventions | necessary |
| Source of intervention data | government or news websites (countries), New York Times (US) |

– continued on next page –

| | |
|---|---|
| Availability of data on exposure | access to externally coded data via source |
| **Methodological approach** | |
| Empirical approach | parametric |
| Use of exposure variation | only variation between populations |
| Method | non-mechanistic model (generalized linear model) |
| Code availability | none |
| **Effect assessment** | |
| Reporting of intervention effect | quantification of change in outcome values |
| Measure of effect | time to peak: cases/deaths |
| Reporting of uncertainty | yes |
| Sensitivity analysis | none |
| Subgroup assessment | none |

**Miller et al. 2020**

| | |
|---|---|
| **Study information** | |
| Author | Miller D, Martin MA, Harel N, Tirosh O, Kustin T, Meir M, Sorek N, Gefen-Halevi S, Amit S, Vorontsov O, Shaag A, Wolf D, Peretz A, Shemer-Avni Y, Roif-Kaminsky D, Kopelman NM, Huppert A, Koelle K, Stern A |
| Title | Full Genome Viral Sequences Inform Patterns of SARS-CoV-2 Spread into and within Israel |
| Year | 2020 |
| Journal | Nature Communications |
| DOI | 10.1038/s41467-020-19248-0 |
| Date received | 2020-06-12 |

– continued on next page –

| | |
|---|---|
| Date accepted | 2020-10-02 |
| **Study setting** | |
| Number of populations included | single |
| Level of populations included | national (Israel) |
| Geographic areas covered | Asia |
| Number of countries covered | single country (Israel) |
| Study period | start and end date span first epidemic wave |
| **Outcome** | |
| Raw outcome | epidemiological individual-level outcome (genome sequence data) |
| Time resolution of raw outcome | daily |
| Computed outcome | none |
| Method to obtain the computed outcome | – not applicable – |
| Data source | data collected by authors, data from publicly available cross-country selections (GISAID) |
| Data availability | data made available by the authors |
| **Intervention** | |
| Terminology for interventions | measures |
| Terminology for the specific type of non-pharmaceutical interventions | social distancing |
| Exposure types | one combination of interventions (interventions implemented on March 19) |
| Types of single interventions | – not applicable – |
| Coding of interventions | not necessary |
| Source of intervention data | – could not be evaluated – |
| Availability of data on exposure | raw data documented in the manuscript |
| **Methodological approach** | |

– continued on next page –

| | |
|---|---|
| Empirical approach | parametric |
| Use of exposure variation | only variation over time for a single population |
| Method | mechanistic model (phylodynamic model) |
| Latent variable functionally linked to intervention | reproduction number |
| Code availability | publicly available |
| **Effect assessment** | |
| Reporting of intervention effect | quantification of change in outcome values |
| Measure of effect | reproduction number |
| Reporting of uncertainty | yes |
| Sensitivity analysis | model specification varied, epidemiological parameters varied |
| Subgroup assessment | none |

| **Mitra et al. 2020** | |
|---|---|
| **Study information** | |
| Author | Mitra A, Pakhare AP, Roy A, Joshi A |
| Title | Impact of COVID-19 Epidemic Curtailment Strategies in Selected Indian States: An Analysis by Reproduction Number and Doubling Time with Incidence Modelling |
| Year | 2020 |
| Journal | PLOS ONE |
| DOI | 10.1371/journal.pone.0239026 |
| Date received | 2020-05-18 |
| Date accepted | 2020-08-28 |
| **Study setting** | |

– continued on next page –

| | |
|---|---|
| Number of populations included | multiple |
| Level of populations included | subnational (Top 10 Indian states with highest incidence) |
| Geographic areas covered | Asia |
| Number of countries covered | single country (India) |
| Study period | end date in growth phase of wave |
| **Outcome** | |
| Raw outcome | epidemiological population-level outcome (cases) |
| Time resolution of raw outcome | daily |
| Computed outcome | measure of epidemic trend (growth rate, doubling time), epidemiological parameter (reproduction number) |
| Method to obtain the computed outcome | exponential growth model (growth rate), exponential growth model (doubling time), statistical estimation of reproduction number (White et al.) |
| Data source | data from publicly available open-source projects (covid19India.org) |
| Data availability | data made available by the authors |
| **Intervention** | |
| Terminology for interventions | strategies |
| Terminology for the specific type of non-pharmaceutical interventions | curtailment |
| Exposure types | one combination of interventions (Indian National Lockdown) |
| Types of single interventions | – not applicable – |
| Coding of interventions | not necessary |
| Source of intervention data | – could not be evaluated – |
| Availability of data on exposure | raw data documented in the manuscript |
| **Methodological approach** | |
| Empirical approach | descriptive |
| Use of exposure variation | only variation over time for multiple populations |

– continued on next page –

**Mitra et al. 2020** – continued from previous page

| | |
|---|---|
| Method | description of change over time (comparison of time periods) |
| Code availability | publicly available |
| **Effect assessment** | |
| Reporting of intervention effect | comparison of outcome values |
| Measure of effect | growth rate, doubling time, reproduction number |
| Reporting of uncertainty | yes |
| Sensitivity analysis | none |
| Subgroup assessment | none |

**Moon et al. 2020**

| | |
|---|---|
| **Study information** | |
| Author | Moon SG, Kim YK, Son WS, Kim JH, Choi J, Na BJ, Park B |
| Title | Time Variant Reproductive Number of COVID-19 in Seoul, Korea |
| Year | 2020 |
| Journal | Epidemiology and Health |
| DOI | 10.4178/epih.e2020047 |
| Date received | 2020-04-01 |
| Date accepted | 2020-06-20 |
| **Study setting** | |
| Number of populations included | single |
| Level of populations included | subnational (Seoul (Korea)) |
| Geographic areas covered | Asia |
| Number of countries covered | single country (South Korea) |
| Study period | start and end date span first epidemic wave |

– continued on next page –

| | |
|---|---|
| **Outcome** | |
| Raw outcome | epidemiological population-level outcome (cases) |
| Time resolution of raw outcome | daily |
| Computed outcome | epidemiological parameter (reproduction number) |
| Method to obtain the computed outcome | statistical estimation of reproduction number (Cori et al.) |
| Data source | data from (sub)national authorities |
| Data availability | data access via source |
| **Intervention** | |
| Terminology for interventions | policies |
| Terminology for the specific type of non-pharmaceutical interventions | quarantine |
| Exposure types | all interventions together |
| Types of single interventions | – not applicable – |
| Coding of interventions | – not applicable – |
| Source of intervention data | – not applicable – |
| Availability of data on exposure | – not applicable – |
| **Methodological approach** | |
| Empirical approach | descriptive |
| Use of exposure variation | only variation over time for a single population |
| Method | description of change over time (description of time course) |
| Code availability | none |
| **Effect assessment** | |
| Reporting of intervention effect | qualitative statement |
| Measure of effect | reproduction number |
| Reporting of uncertainty | – not applicable – |

– continued on next page –

| | |
|---|---|
| Sensitivity analysis | different or modified outcome used (use effective reproduction number based on cases by date of confirmation and by date of onset), epidemiological parameters varied |
| Subgroup assessment | none |

**Moreno et al. 2020**

| | |
|---|---|
| **Study information** | |
| Author | Moreno GK, Braun KM, Riemersma KK, Martin MA, Halfmann PJ, Crooks CM, Prall T, Baker D, Baczenas JJ, Heffron AS, Ramuta M, Khubbar M, Weiler AM, Accola MA, Rehrauer WM, O'Connor SL, Safdar N, Pepperell CS, Dasu T, Bhattacharyya S, Kawaoka Y, Koelle K, O'Connor DH, Friedrich TC |
| Title | Revealing Fine-Scale Spatiotemporal Differences in SARS-CoV-2 Introduction and Spread |
| Year | 2020 |
| Journal | Nature Communications |
| DOI | 10.1038/s41467-020-19346-z |
| Date received | 2020-06-30 |
| Date accepted | 2020-10-06 |
| **Study setting** | |
| Number of populations included | multiple |
| Level of populations included | subnational (2 US counties (Dane and Milwaukee)) |
| Geographic areas covered | North America |
| Number of countries covered | single country (US) |
| Study period | end date in growth phase of wave |

– continued on next page –

| | |
|---|---|
| **Outcome** | |
| Raw outcome | epidemiological individual-level outcome (genome sequence data) |
| Time resolution of raw outcome | daily |
| Computed outcome | none |
| Method to obtain the computed outcome | – not applicable – |
| Data source | data collected by authors (hospitals and laboratories in the counties) |
| Data availability | data made available by the authors |
| **Intervention** | |
| Terminology for interventions | interventions |
| Terminology for the specific type of non-pharmaceutical interventions | public health |
| Exposure types | one single intervention (stay-at-home order (business closure, stay-at-home order, gathering bans)) |
| Types of single interventions | stay-at-home order |
| Coding of interventions | not necessary |
| Source of intervention data | government or news websites |
| Availability of data on exposure | raw data documented in the manuscript |
| **Methodological approach** | |
| Empirical approach | parametric |
| Use of exposure variation | only variation over time for multiple populations |
| Method | mechanistic model (phylodynamic model) |
| Latent variable functionally linked to intervention | reproduction number |
| Code availability | publicly available |
| **Effect assessment** | |

– continued on next page –

| | |
|---|---|
| Reporting of intervention effect | quantification of change in outcome values |
| Measure of effect | reproduction number |
| Reporting of uncertainty | yes |
| Sensitivity analysis | epidemiological parameters varied |
| Subgroup assessment | none |

| **Moshammer et al. 2020** | |
|---|---|
| **Study information** | |
| Author | Moshammer H, Poteser M, Lemmerer K, Wallner P, Hutter HP |
| Title | Time Course of COVID-19 Cases in Austria |
| Year | 2020 |
| Journal | International Journal of Environmental Research and Public Health |
| DOI | 10.3390/ijerph17093270 |
| Date received | 2020-04-15 |
| Date accepted | 2020-05-06 |
| **Study setting** | |
| Number of populations included | single |
| Level of populations included | national (Austria) |
| Geographic areas covered | Europe |
| Number of countries covered | single country (Austria) |
| Study period | start and end date span first epidemic wave |
| **Outcome** | |
| Raw outcome | epidemiological population-level outcome (cases) |
| Time resolution of raw outcome | daily |

– continued on next page –

| | |
|---|---|
| Computed outcome | epidemiological parameter (reproduction number) |
| Method to obtain the computed outcome | back projection, statistical estimation of reproduction number (other, very simplistic approach) |
| Data source | data from publicly available cross-country selections (JHU) |
| Data availability | data access via source |
| **Intervention** | |
| Terminology for interventions | measures |
| Terminology for the specific type of non-pharmaceutical interventions | public health |
| Exposure types | all interventions together (closure of schools, restraurants, shops, cancellation of conferences etc) |
| Types of single interventions | – not applicable – |
| Coding of interventions | – not applicable – |
| Source of intervention data | – not applicable – |
| Availability of data on exposure | – not applicable – |
| **Methodological approach** | |
| Empirical approach | descriptive |
| Use of exposure variation | only variation over time for a single population |
| Method | description of change over time (description of time course) |
| Code availability | none |
| **Effect assessment** | |
| Reporting of intervention effect | qualitative statement |
| Measure of effect | cases, reproduction number |
| Reporting of uncertainty | – not applicable – |
| Sensitivity analysis | none |
| Subgroup assessment | none |

| Mukandavire et al. 2020 | |
|---|---|
| **Study information** | |
| Author | Mukandavire Z, Nyabadza F, Malunguza NJ, Cuadros DF, Shiri T, Musuka G |
| Title | Quantifying Early COVID-19 Outbreak Transmission in South Africa and Exploring Vaccine Efficacy Scenarios |
| Year | 2020 |
| Journal | PLOS ONE |
| DOI | 10.1371/journal.pone.0236003 |
| Date received | 2020-04-23 |
| Date accepted | 2020-06-27 |
| **Study setting** | |
| Number of populations included | single |
| Level of populations included | national (South Africa) |
| Geographic areas covered | Central and South America |
| Number of countries covered | single country (South Africa) |
| Study period | end date in growth phase of wave |
| **Outcome** | |
| Raw outcome | epidemiological population-level outcome (cases) |
| Time resolution of raw outcome | daily |
| Computed outcome | epidemiological parameter (transmission rate) |
| Method to obtain the computed outcome | compartmental transmission model (single-population) |
| Data source | data from (sub)national authorities |
| Data availability | data made available by the authors |

– continued on next page –

| | |
|---|---|
| **Intervention** | |
| Terminology for interventions | measures |
| Terminology for the specific type of non-pharmaceutical interventions | social distancing |
| Exposure types | one combination of interventions (lockdown issued on March 26) |
| Types of single interventions | – not applicable – |
| Coding of interventions | not necessary |
| Source of intervention data | – could not be evaluated – |
| Availability of data on exposure | raw data documented in the manuscript |
| **Methodological approach** | |
| Empirical approach | descriptive |
| Use of exposure variation | only variation over time for a single population |
| Method | description of change over time (comparison of time periods) |
| Code availability | none |
| **Effect assessment** | |
| Reporting of intervention effect | quantification of change in outcome values |
| Measure of effect | transmission rate |
| Reporting of uncertainty | yes |
| Sensitivity analysis | none |
| Subgroup assessment | none |

**Munayco et al. 2020**

**Study information**

– continued on next page –

| | |
|---|---|
| Author | Munayco CV, Tariq A, Rothenberg R, Soto-Cabezas GG, Reyes MF, Valle A, Rojas-Mezarina L, Cabezas C, Loayza M, Chowell G, Garro DC, Vasquez KM, Castro ES, Ordinola IS, Mimbela JM, Cornejo KM, Quijano FC, Rosillo LT, Ibarguen LO, Dominguez MV, Seminario RV, Silva MC, Dreyfus MS, Pineda ML, Durand M, Janampa N, Chuquihuaccha J, Lizarbe SM, Cusi DE, Pilco IM, Jaramillo A, Vargas K, Cabanillas O, Arrasco J, Vargas M, Ramos W |
| Title | Early Transmission Dynamics of COVID-19 in a Southern Hemisphere Setting: Lima-Peru: February 29th–March 30th, 2020 |
| Year | 2020 |
| Journal | Infectious Disease Modelling |
| DOI | 10.1016/j.idm.2020.05.001 |
| Date received | 2020-04-24 |
| Date accepted | 2020-05-07 |
| **Study setting** | |
| Number of populations included | single |
| Level of populations included | subnational (Lima (Peru)) |
| Geographic areas covered | Central and South America |
| Number of countries covered | single country (Peru) |
| Study period | start and end date span first epidemic wave |
| **Outcome** | |
| Raw outcome | epidemiological population-level outcome (cases) |
| Time resolution of raw outcome | daily |
| Computed outcome | none |
| Method to obtain the computed outcome | – not applicable – |
| Data source | data from (sub)national authorities |

– continued on next page –

| | |
|---|---|
| Data availability | data access via source |
| **Intervention** | |
| Terminology for interventions | measures |
| Terminology for the specific type of non-pharmaceutical interventions | social distancing |
| Exposure types | all interventions together (school closure, emergency declaration (national), other social distancing interventions) |
| Types of single interventions | – not applicable – |
| Coding of interventions | – not applicable – |
| Source of intervention data | – not applicable – |
| Availability of data on exposure | – not applicable – |
| **Methodological approach** | |
| Empirical approach | counterfactual |
| Use of exposure variation | only variation over time for a single population |
| Method | non-mechanistic model (exponential growth model) |
| Code availability | none |
| **Effect assessment** | |
| Reporting of intervention effect | qualitative statement |
| Measure of effect | cases |
| Reporting of uncertainty | – not applicable – |
| Sensitivity analysis | none |
| Subgroup assessment | none |

| Murillo-Zamora et al. 2020 | |
|---|---|
| **Study information** | |
| Author | Murillo-Zamora E, Guzmán-Esquivel J, Sánchez-Piña RA, Cedeño-Laurent G, Delgado-Enciso I, Mendoza-Cano O |
| Title | Physical Distancing Reduced the Incidence of Influenza and Supports a Favorable Impact on SARS-CoV-2 Spread in Mexico |
| Year | 2020 |
| Journal | The Journal of Infection in Developing Countries |
| DOI | 10.3855/jidc.13250 |
| Date received | 2020-06-12 |
| Date accepted | 2020-08-11 |
| **Study setting** | |
| Number of populations included | multiple |
| Level of populations included | subnational (Maxican states) |
| Geographic areas covered | Central and South America |
| Number of countries covered | single country (Mexico) |
| Study period | end date in growth phase of wave |
| **Outcome** | |
| Raw outcome | epidemiological population-level outcome (cases) |
| Time resolution of raw outcome | daily |
| Computed outcome | measure of epidemic trend (growth rate) |
| Method to obtain the computed outcome | simple computation |
| Data source | data from (sub)national authorities |
| Data availability | data access via source |
| **Intervention** | |
| Terminology for interventions | measures |

– continued on next page –

| | |
|---|---|
| Terminology for the specific type of non-pharmaceutical interventions | physical distancing |
| Exposure types | one single intervention (school closure) |
| Types of single interventions | school closure |
| Coding of interventions | not necessary |
| Source of intervention data | government or news websites |
| Availability of data on exposure | raw data documented in the manuscript |
| **Methodological approach** | |
| Empirical approach | descriptive |
| Use of exposure variation | both variation over time and between populations |
| Method | comparison of populations |
| Code availability | none |
| **Effect assessment** | |
| Reporting of intervention effect | quantification of change in outcome values |
| Measure of effect | growth rate |
| Reporting of uncertainty | yes |
| Sensitivity analysis | none |
| Subgroup assessment | socioeconomic indicators (age), public health response (response time) |

**Ng et al. 2020**

| | |
|---|---|
| **Study information** | |
| Author | Ng Y, Li Z, Chua YX, Chaw WL, Zhao Z, Er B, Pung R, Chiew CJ, Lye DC, Heng D, Lee VJ |

– continued on next page –

| | |
|---|---|
| Title | Evaluation of the Effectiveness of Surveillance and Containment Measures for the First 100 Patients with COVID-19 in Singapore —January 2–February 29, 2020 |
| Year | 2020 |
| Journal | Morbidity and Mortality Weekly Report |
| DOI | 10.15585/mmwr.mm6911e1 |
| Date received | – could not be evaluated – |
| Date accepted | 2020-03-20 (published) |
| **Study setting** | |
| Number of populations included | single |
| Level of populations included | national (Singapore) |
| Geographic areas covered | Asia |
| Number of countries covered | single country (Singapore) |
| Study period | end date in growth phase of wave |
| **Outcome** | |
| Raw outcome | epidemiological individual-level outcome (individual cases) |
| Time resolution of raw outcome | daily |
| Computed outcome | epidemiological parameter (time from symptom onset to isolation) |
| Method to obtain the computed outcome | simple computation |
| Data source | data from (sub)national authorities |
| Data availability | data not accessible |
| **Intervention** | |
| Terminology for interventions | measures |
| Terminology for the specific type of non-pharmaceutical interventions | surveillance, containment |

– continued on next page –

| | |
|---|---|
| Exposure types | all interventions together |
| Types of single interventions | – not applicable – |
| Coding of interventions | – not applicable – |
| Source of intervention data | – not applicable – |
| Availability of data on exposure | – not applicable – |
| **Methodological approach** | |
| Empirical approach | descriptive |
| Use of exposure variation | only variation over time for a single population |
| Method | description of change over time (comparison of time periods) |
| Code availability | none |
| **Effect assessment** | |
| Reporting of intervention effect | quantification of change in outcome values |
| Measure of effect | time from symptom onset to isolation |
| Reporting of uncertainty | no |
| Sensitivity analysis | none |
| Subgroup assessment | epidemiological indicators (source of infection) |

**Niazkar et al. 2020 (Analysis 1)**

| | |
|---|---|
| **Study information** | |
| Author | Niazkar M, Türkkan GE, Niazkar HR, Türkkan YA |
| Title | Assessment of Three Mathematical Prediction Models for Forecasting the COVID-19 Outbreak in Iran and Turkey |
| Year | 2020 |

– continued on next page –

| | |
|---|---|
| Journal | Computational and Mathematical Methods in Medicine |
| DOI | 10.1155/2020/7056285 |
| Date received | 2020-07-27 |
| Date accepted | 2020-11-03 |
| **Study setting** | |
| Number of populations included | single |
| Level of populations included | national (Iran) |
| Geographic areas covered | Middle East and Africa |
| Number of countries covered | single country (Iran) |
| Study period | start and end date span first epidemic wave |
| **Outcome** | |
| Raw outcome | epidemiological population-level outcome (cases) |
| Time resolution of raw outcome | daily |
| Computed outcome | none |
| Method to obtain the computed outcome | – not applicable – |
| Data source | data from publicly available cross-country selections (WHO) |
| Data availability | data access via source |
| **Intervention** | |
| Terminology for interventions | measures |
| Terminology for the specific type of non-pharmaceutical interventions | control |
| Exposure types | one single intervention (workplace closure (reopening of low-risk businesses)) |
| Types of single interventions | workplace closure |
| Coding of interventions | not necessary |
| Source of intervention data | government or news websites |

– continued on next page –

| | |
|---|---|
| Availability of data on exposure | raw data documented in the manuscript |
| **Methodological approach** | |
| Empirical approach | counterfactual |
| Use of exposure variation | only variation over time for a single population |
| Method | non-mechanistic model (exponential growth model) |
| Code availability | none |
| **Effect assessment** | |
| Reporting of intervention effect | quantification of change in outcome values |
| Measure of effect | cases |
| Reporting of uncertainty | no |
| Sensitivity analysis | none |
| Subgroup assessment | none |

| | Niazkar et al. 2020 (Analysis 2) |
|---|---|
| **Study information** | |
| Author | Niazkar M, Türkkan GE, Niazkar HR, Türkkan YA |
| Title | Assessment of Three Mathematical Prediction Models for Forecasting the COVID-19 Outbreak in Iran and Turkey |
| Year | 2020 |
| Journal | Computational and Mathematical Methods in Medicine |
| DOI | 10.1155/2020/7056285 |
| Date received | 2020-07-27 |
| Date accepted | 2020-11-03 |
| **Study setting** | |

– continued on next page –

| | |
|---|---|
| Number of populations included | single |
| Level of populations included | national (Turkey) |
| Geographic areas covered | Middle East and Africa |
| Number of countries covered | single country (Turkey) |
| Study period | end date in growth phase of wave |
| **Outcome** | |
| Raw outcome | epidemiological population-level outcome (cases) |
| Time resolution of raw outcome | daily |
| Computed outcome | none |
| Method to obtain the computed outcome | – not applicable – |
| Data source | data from (sub)national authorities |
| Data availability | data access via source |
| **Intervention** | |
| Terminology for interventions | measures |
| Terminology for the specific type of non-pharmaceutical interventions | control |
| Exposure types | one combination of interventions (curfew, bans of large gatherings (canelling of sport events), venue closure (closure of mosques)) |
| Types of single interventions | – not applicable – |
| Coding of interventions | not necessary |
| Source of intervention data | government or news websites |
| Availability of data on exposure | raw data documented in the manuscript |
| **Methodological approach** | |
| Empirical approach | counterfactual |
| Use of exposure variation | only variation over time for a single population |

– continued on next page –

| | |
|---|---|
| Method | non-mechanistic model (exponential growth model) |
| Code availability | none |
| **Effect assessment** | |
| Reporting of intervention effect | quantification of change in outcome values |
| Measure of effect | cases |
| Reporting of uncertainty | no |
| Sensitivity analysis | none |
| Subgroup assessment | none |

**Nie et al. 2020**

| | |
|---|---|
| **Study information** | |
| Author | Nie L, Guo AX, Yi C, And RW |
| Title | Analyzing the Effects of Public Interventions on Reducing Public Gatherings in China during the COVID-19 Epidemic via Mobile Terminals Positioning Data |
| Year | 2020 |
| Journal | Mathematical Biosciences and Engineering |
| DOI | 10.3934/mbe.2020265 |
| Date received | 2020-05-07 |
| Date accepted | 2020-07-09 |
| **Study setting** | |
| Number of populations included | multiple |
| Level of populations included | both national and subnational (China and its 31 provinces) |
| Geographic areas covered | Asia |

– continued on next page –

| | |
|---|---|
| Number of countries covered | single country (China) |
| Study period | start and end date span first epidemic wave: behavioral |
| **Outcome** | |
| Raw outcome | behavioral outcome (mobility: number of mobile terminals with specific points of interest) |
| Time resolution of raw outcome | daily |
| Computed outcome | none |
| Method to obtain the computed outcome | – not applicable – |
| Data source | – could not be evaluated – |
| Data availability | data not accessible |
| **Intervention** | |
| Terminology for interventions | interventions |
| Terminology for the specific type of non-pharmaceutical interventions | public health |
| Exposure types | multiple combinations of interventions (five strategies involving work-frome-home orders and venue closure as well as their partial resumption) |
| Types of single interventions | – not applicable – |
| Coding of interventions | not necessary |
| Source of intervention data | – could not be evaluated – |
| Availability of data on exposure | raw data documented in the manuscript |
| **Methodological approach** | |
| Empirical approach | parametric |
| Use of exposure variation | only variation over time for a single population |
| Method | non-mechanistic model (generalized linear model) |
| Code availability | none |

– continued on next page –

| | |
|---|---|
| **Effect assessment** | |
| Reporting of intervention effect | quantification of change in outcome values |
| Measure of effect | mobility: number of people in workplace or consumption areas |
| Reporting of uncertainty | yes |
| Sensitivity analysis | none |
| Subgroup assessment | none |

**Niwa et al. 2020**

| | |
|---|---|
| **Study information** | |
| Author | Niwa M, Hara Y, Sengoku S, Kodama K |
| Title | Effectiveness of Social Measures against COVID-19 Outbreaks in Selected Japanese Regions Analyzed by System Dynamic Modeling |
| Year | 2020 |
| Journal | International Journal of Environmental Research and Public Health |
| DOI | 10.3390/ijerph17176238 |
| Date received | 2020-07-17 |
| Date accepted | 2020-08-19 |
| **Study setting** | |
| Number of populations included | multiple |
| Level of populations included | subnational (3 regions in Japan (Tokyo metropolitan area, Osaka prefecture, Hokkaido prefecture)) |
| Geographic areas covered | Asia |
| Number of countries covered | single country (Japan) |
| Study period | start and end date span first epidemic wave |

– continued on next page –

| | |
|---|---|
| **Outcome** | |
| Raw outcome | epidemiological population-level outcome (cases) |
| Time resolution of raw outcome | daily |
| Computed outcome | epidemiological parameter (other: transmission efficiency) |
| Method to obtain the computed outcome | other (stock-and-flow model based on system dynamics) |
| Data source | data from (sub)national authorities |
| Data availability | data made available by the authors |
| **Intervention** | |
| Terminology for interventions | measures |
| Terminology for the specific type of non-pharmaceutical interventions | control |
| Exposure types | all interventions together |
| Types of single interventions | – not applicable – |
| Coding of interventions | – not applicable – |
| Source of intervention data | – not applicable – |
| Availability of data on exposure | – not applicable – |
| **Methodological approach** | |
| Empirical approach | descriptive |
| Use of exposure variation | only variation over time for multiple populations |
| Method | description of change over time (comparison of time periods) |
| Code availability | none |
| **Effect assessment** | |
| Reporting of intervention effect | quantification of change in outcome values |
| Measure of effect | transmission efficiency |
| Reporting of uncertainty | no |

– continued on next page –

| | |
|---|---|
| Sensitivity analysis | none |
| Subgroup assessment | none |

| **Odagaki 2020** | |
|---|---|
| **Study information** | |
| Author | Odagaki T |
| Title | Analysis of the Outbreak of COVID-19 in Japan by SIQR Model |
| Year | 2020 |
| Journal | Infectious Disease Modelling |
| DOI | 10.1016/j.idm.2020.08.013 |
| Date received | 2020-06-11 |
| Date accepted | 2020-08-28 |
| **Study setting** | |
| Number of populations included | single |
| Level of populations included | national (Japan) |
| Geographic areas covered | Asia |
| Number of countries covered | single country (Japan) |
| Study period | start and end date span first epidemic wave |
| **Outcome** | |
| Raw outcome | epidemiological population-level outcome (cases) |
| Time resolution of raw outcome | daily |
| Computed outcome | epidemiological parameter (transmission rate) |
| Method to obtain the computed outcome | compartmental transmission model (single-population) |

– continued on next page –

| | |
|---|---|
| Data source | data from (sub)national authorities |
| Data availability | data access via source |
| **Intervention** | |
| Terminology for interventions | measures |
| Terminology for the specific type of non-pharmaceutical interventions | lockdown, quarantine |
| Exposure types | one single intervention (emergency declaration) |
| Types of single interventions | declaration of a state of emergency |
| Coding of interventions | not necessary |
| Source of intervention data | – could not be evaluated – |
| Availability of data on exposure | raw data documented in the manuscript |
| **Methodological approach** | |
| Empirical approach | descriptive |
| Use of exposure variation | only variation over time for a single population |
| Method | description of change over time (comparison of time periods) |
| Code availability | none |
| **Effect assessment** | |
| Reporting of intervention effect | quantification of change in outcome values |
| Measure of effect | transmission rate, reproduction number |
| Reporting of uncertainty | no |
| Sensitivity analysis | none |
| Subgroup assessment | none |

| **Ouchetto et al. 2020** | |
|---|---|
| **Study information** | |
| Author | Ouchetto O, Bourhanbour AD, Boumhamdi M |
| Title | Effectiveness of Containment Measures to Control the Spread of COVID-19 in North Africa |
| Year | 2020 |
| Journal | Disaster Medicine and Public Health Preparedness |
| DOI | 10.1017/dmp.2020.314 |
| Date received | – could not be evaluated – |
| Date accepted | 2020-09-03 (published) |
| **Study setting** | |
| Number of populations included | multiple |
| Level of populations included | national (Morocco, Algeria, Tunisia) |
| Geographic areas covered | Middle East and Africa |
| Number of countries covered | multiple countries |
| Study period | end date at peak of wave |
| **Outcome** | |
| Raw outcome | epidemiological population-level outcome (cases) |
| Time resolution of raw outcome | daily |
| Computed outcome | epidemiological parameter (reproduction number) |
| Method to obtain the computed outcome | statistical estimation of reproduction number (Wallinga and Teunis) |
| Data source | data from publicly available cross-country selections (ECDC) |
| Data availability | data access via source |
| **Intervention** | |
| Terminology for interventions | measures |

– continued on next page –

| | |
|---|---|
| Terminology for the specific type of non-pharmaceutical interventions | containment |
| Exposure types | one single intervention (stay-at-home order) |
| Types of single interventions | stay-at-home order |
| Coding of interventions | not necessary |
| Source of intervention data | – could not be evaluated – |
| Availability of data on exposure | raw data documented in the manuscript |
| **Methodological approach** | |
| Empirical approach | descriptive |
| Use of exposure variation | only variation over time for multiple populations |
| Method | description of change over time (comparison of time periods) |
| Code availability | none |
| **Effect assessment** | |
| Reporting of intervention effect | comparison of outcome values |
| Measure of effect | reproduction number |
| Reporting of uncertainty | no |
| Sensitivity analysis | none |
| Subgroup assessment | none |

**Pan et al. 2020a**

| | |
|---|---|
| **Study information** | |
| Author | Pan Y, Darzi A, Kabiri A, Zhao G, Luo W, Xiong C, Zhang L |
| Title | Quantifying Human Mobility Behaviour Changes during the COVID-19 Outbreak in the United States |

– continued on next page –

| | |
|---|---|
| Year | 2020 |
| Journal | Scientific Reports |
| DOI | 10.1038/s41598-020-77751-2 |
| Date received | 2020-06-18 |
| Date accepted | 2020-11-03 |
| **Study setting** | |
| Number of populations included | multiple |
| Level of populations included | both national and subnational (US and all states) |
| Geographic areas covered | North America |
| Number of countries covered | single country (US) |
| Study period | start and end date span first epidemic wave: behavioral |
| **Outcome** | |
| Raw outcome | behavioral outcome (mobility: percentage staying at home & daily work trips & non-work trips & distance travelled & out-of-county trips) |
| Time resolution of raw outcome | daily |
| Computed outcome | none |
| Method to obtain the computed outcome | – not applicable – |
| Data source | public (Maryland Transportation Institute led by the University of Maryland) |
| Data availability | data made available by the authors |
| **Intervention** | |
| Terminology for interventions | orders |
| Terminology for the specific type of non-pharmaceutical interventions | government |

– continued on next page –

| | |
|---|---|
| Exposure types | multiple separate interventions (stay-at-home order, emergency declaration (national)) |
| Types of single interventions | stay-at-home order, declaration of a state of emergency |
| Coding of interventions | necessary |
| Source of intervention data | – could not be evaluated – |
| Availability of data on exposure | coded data not available |
| **Methodological approach** | |
| Empirical approach | descriptive |
| Use of exposure variation | only variation over time for multiple populations |
| Method | description of change over time (description of time course) |
| Code availability | publicly available |
| **Effect assessment** | |
| Reporting of intervention effect | qualitative statement |
| Measure of effect | mobility: social distancing index |
| Reporting of uncertainty | – not applicable – |
| Sensitivity analysis | none |
| Subgroup assessment | none |

**Pan et al. 2020b**

| | |
|---|---|
| **Study information** | |
| Author | Pan A, Liu L, Wang C, Guo H, Hao X, Wang Q, Huang J, He N, Yu H, Lin X, Wei S, Wu T |
| Title | Association of Public Health Interventions with the Epidemiology of the COVID-19 Outbreak in Wuhan, China |
| Year | 2020 |

– continued on next page –

| | |
|---|---|
| Journal | JAMA |
| DOI | 10.1001/jama.2020.6130 |
| Date received | – could not be evaluated – |
| Date accepted | 2020-04-10 (published) |
| **Study setting** | |
| Number of populations included | single |
| Level of populations included | subnational (Wuhan) |
| Geographic areas covered | Asia |
| Number of countries covered | single country (China) |
| Study period | start and end date span first epidemic wave |
| **Outcome** | |
| Raw outcome | epidemiological individual-level outcome (individual cases) |
| Time resolution of raw outcome | daily |
| Computed outcome | epidemiological parameter (reproduction number) |
| Method to obtain the computed outcome | statistical estimation of reproduction number (Cori et al.) |
| Data source | data from (sub)national authorities |
| Data availability | data not accessible |
| **Intervention** | |
| Terminology for interventions | interventions |
| Terminology for the specific type of non-pharmaceutical interventions | public health |
| Exposure types | one combination of interventions (Wuhan City Lockdown, centralized quarantine and treatment, universal symptom survey) |
| Types of single interventions | – not applicable – |
| Coding of interventions | not necessary |

– continued on next page –

| | |
|---|---|
| Source of intervention data | – could not be evaluated – |
| Availability of data on exposure | raw data documented in the manuscript |
| **Methodological approach** | |
| Empirical approach | descriptive |
| Use of exposure variation | only variation over time for a single population |
| Method | description of change over time (comparison of time periods) |
| Code availability | none |
| **Effect assessment** | |
| Reporting of intervention effect | comparison of outcome values |
| Measure of effect | cases, reproduction number |
| Reporting of uncertainty | yes |
| Sensitivity analysis | none |
| Subgroup assessment | socioeconomic indicators (age, gender, health care occupation), epidemiological indicators (disease severity) |

**Pang et al. 2020**

| | |
|---|---|
| **Study information** | |
| Author | Pang L, Liu S, Zhang X, Tian T, Zhao Z |
| Title | Transmission Dynamics and Control Strategies of COVID-19 in Wuhan, China |
| Year | 2020 |
| Journal | Journal of Biological Systems |
| DOI | 10.1142/s0218339020500096 |
| Date received | 2020-02-23 |

– continued on next page –

| | |
|---|---|
| Date accepted | 2020-04-14 |
| **Study setting** | |
| Number of populations included | single |
| Level of populations included | subnational (Wuhan) |
| Geographic areas covered | Asia |
| Number of countries covered | single country (China) |
| Study period | start and end date span first epidemic wave |
| **Outcome** | |
| Raw outcome | epidemiological population-level outcome (cases) |
| Time resolution of raw outcome | daily |
| Computed outcome | epidemiological parameter (reproduction number, transmission rate) |
| Method to obtain the computed outcome | compartmental transmission model (single-population) |
| Data source | data from (sub)national authorities |
| Data availability | data access via source |
| **Intervention** | |
| Terminology for interventions | measures |
| Terminology for the specific type of non-pharmaceutical interventions | control |
| Exposure types | one combination of interventions (Wuhan City Lockdown and strict quarantine measures) |
| Types of single interventions | – not applicable – |
| Coding of interventions | not necessary |
| Source of intervention data | – could not be evaluated – |
| Availability of data on exposure | raw data documented in the manuscript |
| **Methodological approach** | |

– continued on next page –

| | |
|---|---|
| Empirical approach | descriptive |
| Use of exposure variation | only variation over time for a single population |
| Method | description of change over time (comparison of time periods) |
| Code availability | none |
| **Effect assessment** | |
| Reporting of intervention effect | comparison of outcome values |
| Measure of effect | reproduction number, transmission rate |
| Reporting of uncertainty | yes |
| Sensitivity analysis | none |
| Subgroup assessment | none |

| **Pei et al. 2020** | |
|---|---|
| **Study information** | |
| Author | Pei S, Kandula S, Shaman J |
| Title | Differential Effects of Intervention Timing on COVID-19 Spread in the United States |
| Year | 2020 |
| Journal | Science Advances |
| DOI | 10.1126/sciadv.abd6370 |
| Date received | 2020-07-02 |
| Date accepted | 2020-10-20 |
| **Study setting** | |
| Number of populations included | multiple |
| Level of populations included | both national and subnational (US counties) |

– continued on next page –

| | |
|---|---|
| Geographic areas covered | North America |
| Number of countries covered | single country (US) |
| Study period | same end date for several populations with diverse epidemic trajectories |
| **Outcome** | |
| Raw outcome | epidemiological population-level outcome (cases, deaths) |
| Time resolution of raw outcome | daily |
| Computed outcome | epidemiological parameter (reproduction number) |
| Method to obtain the computed outcome | compartmental transmission model (single-population) |
| Data source | public (USAFacts) |
| Data availability | data made available by the authors |
| **Intervention** | |
| Terminology for interventions | measures |
| Terminology for the specific type of non-pharmaceutical interventions | social distancing, control |
| Exposure types | all interventions together |
| Types of single interventions | – not applicable – |
| Coding of interventions | – not applicable – |
| Source of intervention data | – not applicable – |
| Availability of data on exposure | – not applicable – |
| **Methodological approach** | |
| Empirical approach | descriptive |
| Use of exposure variation | only variation over time for multiple populations |
| Method | description of change over time (description of time course) |
| Code availability | publicly available |
| **Effect assessment** | |

– continued on next page –

| | |
|---|---|
| Reporting of intervention effect | comparison of outcome values |
| Measure of effect | reproduction number |
| Reporting of uncertainty | yes |
| Sensitivity analysis | model specification varied, epidemiological parameters varied |
| Subgroup assessment | none |

| **Peixoto et al. 2020** | |
|---|---|
| **Study information** | |
| Author | Peixoto VR, Vieira A, Aguiar P, Carvalho C, Thomas DR, Abrantes A |
| Title | Initial Assessment of the Impact of the Emergency State Lockdown Measures on the 1st Wave of the COVID-19 Epidemic in Portugal |
| Year | 2020 |
| Journal | Acta Médica Portuguesa |
| DOI | 10.20344/amp.14129 |
| Date received | 2020-05-13 |
| Date accepted | 2020-07-31 |
| **Study setting** | |
| Number of populations included | single |
| Level of populations included | national (Portugal) |
| Geographic areas covered | Europe |
| Number of countries covered | single country (Portugal) |
| Study period | end date at peak of wave |
| **Outcome** | |

– continued on next page –

| | |
|---|---|
| Raw outcome | epidemiological population-level outcome (cases, deaths, hospitalizations, ICU hospitalizations) |
| Time resolution of raw outcome | daily |
| Computed outcome | none |
| Method to obtain the computed outcome | – not applicable – |
| Data source | data from (sub)national authorities |
| Data availability | data access via source |
| **Intervention** | |
| Terminology for interventions | measures |
| Terminology for the specific type of non-pharmaceutical interventions | containment |
| Exposure types | one combination of interventions (lockdown measures decreed on March 16) |
| Types of single interventions | – not applicable – |
| Coding of interventions | not necessary |
| Source of intervention data | – could not be evaluated – |
| Availability of data on exposure | raw data documented in the manuscript |
| **Methodological approach** | |
| Empirical approach | counterfactual |
| Use of exposure variation | only variation over time for a single population |
| Method | non-mechanistic model (time series model) |
| Code availability | none |
| **Effect assessment** | |
| Reporting of intervention effect | quantification of change in outcome values |
| Measure of effect | cases, deaths, hospitalizations, ICU hospitalizations |
| Reporting of uncertainty | no |

– continued on next page –

| | |
|---|---|
| Sensitivity analysis | none |
| Subgroup assessment | none |

| **Peng et al. 2020** | |
|---|---|
| **Study information** | |
| Author | Peng Z, Song W, Ding Z, Guan Q, Yang X, Xu Q, Wang X, Xia Y |
| Title | Linking Key Intervention Timings to Rapid Declining Effective Reproduction Number to Quantify Lessons against COVID-19 |
| Year | 2020 |
| Journal | Frontiers of Medicine |
| DOI | 10.1007/s11684-020-0788-3 |
| Date received | 2020-03-20 |
| Date accepted | 2020-04-08 |
| **Study setting** | |
| Number of populations included | multiple |
| Level of populations included | national (China, Italy, Iran, South Korea, Japan) |
| Geographic areas covered | Asia, Europe, Middle East and Africa |
| Number of countries covered | multiple countries |
| Study period | end date in growth phase of wave |
| **Outcome** | |
| Raw outcome | epidemiological population-level outcome (cases) |
| Time resolution of raw outcome | daily |
| Computed outcome | epidemiological parameter (reproduction number) |

– continued on next page –

| | |
|---|---|
| Method to obtain the computed outcome | statistical estimation of reproduction number (White et al., Walinga et al., Cori et al.) |
| Data source | data from (sub)national authorities (China), data from publicly available cross-country selections (WHO: other countries) |
| Data availability | data access via source |
| **Intervention** | |
| Terminology for interventions | measures |
| Terminology for the specific type of non-pharmaceutical interventions | prevention, control |
| Exposure types | all interventions together |
| Types of single interventions | – not applicable – |
| Coding of interventions | – not applicable – |
| Source of intervention data | – not applicable – |
| Availability of data on exposure | – not applicable – |
| **Methodological approach** | |
| Empirical approach | descriptive |
| Use of exposure variation | only variation over time for multiple populations |
| Method | description of change over time (description of time course) |
| Code availability | none |
| **Effect assessment** | |
| Reporting of intervention effect | qualitative statement |
| Measure of effect | reproduction number |
| Reporting of uncertainty | – not applicable – |
| Sensitivity analysis | none |
| Subgroup assessment | none |

| Pépin et al. 2020 | |
|---|---|
| **Study information** | |
| Author | Pépin JL, Bruno RM, Yang RY, Vercamer V, Jouhaud P, Escourrou P, Boutouyrie P |
| Title | Wearable Activity Trackers for Monitoring Adherence to Home Confinement During the COVID-19 Pandemic Worldwide: Data Aggregation and Analysis |
| Year | 2020 |
| Journal | Journal of Medical Internet Research |
| DOI | 10.2196/19787 |
| Date received | 2020-05-01 |
| Date accepted | 2020-06-04 |
| **Study setting** | |
| Number of populations included | multiple |
| Level of populations included | both national and subnational (Multiple countries around the world including some subnational regions) |
| Geographic areas covered | Asia, Oceania, Europe, North America |
| Number of countries covered | multiple countries |
| Study period | start and end date span first epidemic wave: behavioral |
| **Outcome** | |
| Raw outcome | behavioral outcome (mobility: number of steps made) |
| Time resolution of raw outcome | daily |
| Computed outcome | none |
| Method to obtain the computed outcome | – not applicable – |
| Data source | mobility data from corporate organizations (Withings) |

– continued on next page –

| | |
|---|---|
| Data availability | data not accessible |
| **Intervention** | |
| Terminology for interventions | policies |
| Terminology for the specific type of non-pharmaceutical interventions | confinement |
| Exposure types | multiple combinations of interventions (lockdown: stay-at-home order, partial lockdown: e.g. school closures, venue closures, bans of gatherings) |
| Types of single interventions | – not applicable – |
| Coding of interventions | necessary |
| Source of intervention data | – could not be evaluated – |
| Availability of data on exposure | coded data made available by the authors |
| **Methodological approach** | |
| Empirical approach | descriptive |
| Use of exposure variation | only variation over time for multiple populations |
| Method | description of change over time (comparison of time periods) |
| Code availability | none |
| **Effect assessment** | |
| Reporting of intervention effect | quantification of change in outcome values |
| Measure of effect | mobility: number of steps made |
| Reporting of uncertainty | yes |
| Sensitivity analysis | none |
| Subgroup assessment | none |

| Piovani et al. 2021 | |
|---|---|
| **Study information** | |
| Author | Piovani D, Christodoulou MN, Hadjidemetriou A, Pantavou K, Zaza P, Bagos PG, Bonovas S, Nikolopoulos GK |
| Title | Effect of Early Application of Social Distancing Interventions on COVID-19 Mortality over the First Pandemic Wave: An Analysis of Longitudinal Data from 37 Countries |
| Year | 2021 |
| Journal | Journal of Infection |
| DOI | 10.1016/j.jinf.2020.11.033 |
| Date received | – could not be evaluated – |
| Date accepted | 2020-11-28 |
| **Study setting** | |
| Number of populations included | multiple |
| Level of populations included | national (37 OECD countries) |
| Geographic areas covered | Europe, Middle East and Africa, Asia, Oceania, Central and South America, North America |
| Number of countries covered | multiple countries |
| Study period | same end date for several populations with diverse epidemic trajectories |
| **Outcome** | |
| Raw outcome | epidemiological population-level outcome (deaths) |
| Time resolution of raw outcome | daily |
| Computed outcome | none |
| Method to obtain the computed outcome | – not applicable – |
| Data source | data from publicly available cross-country selections (WHO) |
| Data availability | data access via source |

– continued on next page –

| | |
|---|---|
| **Intervention** | |
| Terminology for interventions | measures |
| Terminology for the specific type of non-pharmaceutical interventions | social distancing |
| Exposure types | one single intervention (bans of large gatherings, school closure) |
| Types of single interventions | bans of large gatherings, school closure |
| Coding of interventions | necessary |
| Source of intervention data | use of externally coded data (OxCGRT) |
| Availability of data on exposure | access to externally coded data via source |
| **Methodological approach** | |
| Empirical approach | parametric |
| Use of exposure variation | both variation over time and between populations |
| Method | non-mechanistic model (generalized linear model) |
| Code availability | none |
| **Effect assessment** | |
| Reporting of intervention effect | quantification of change in outcome values |
| Measure of effect | deaths |
| Reporting of uncertainty | yes |
| Sensitivity analysis | epidemiological parameters varied |
| Subgroup assessment | socioeconomic indicators (age, population density), epidemiological indicators (pre-intervention epidemic trend) |

**Price et al. 2020**

| |
|---|
| **Study information** |

– continued on next page –

| | |
|---|---|
| Author | Price DJ, Shearer FM, Meehan MT, McBryde E, Moss R, Golding N, Conway EJ, Dawson P, Cromer D, Wood J, Abbott S, McVernon J, McCaw JM |
| Title | Early Analysis of the Australian COVID-19 Epidemic |
| Year | 2020 |
| Journal | eLife |
| DOI | 10.7554/elife.58785 |
| Date received | 2020-05-11 |
| Date accepted | 2020-08-12 |
| **Study setting** | |
| Number of populations included | multiple |
| Level of populations included | both national and subnational (Australia and its states) |
| Geographic areas covered | Oceania |
| Number of countries covered | single country (Australia) |
| Study period | start and end date span first epidemic wave |
| **Outcome** | |
| Raw outcome | epidemiological population-level outcome (cases, deaths) |
| Time resolution of raw outcome | daily |
| Computed outcome | epidemiological parameter (reproduction number) |
| Method to obtain the computed outcome | statistical estimation of reproduction number (Cori et al.) |
| Data source | data from (sub)national authorities |
| Data availability | data made available by the authors |
| **Intervention** | |
| Terminology for interventions | interventions |

– continued on next page –

| | |
|---|---|
| Terminology for the specific type of non-pharmaceutical interventions | public health |
| Exposure types | all interventions together (timeline of measures) |
| Types of single interventions | – not applicable – |
| Coding of interventions | – not applicable – |
| Source of intervention data | – not applicable – |
| Availability of data on exposure | – not applicable – |
| **Methodological approach** | |
| Empirical approach | descriptive |
| Use of exposure variation | only variation over time for multiple populations |
| Method | description of change over time (description of time course) |
| Code availability | publicly available |
| **Effect assessment** | |
| Reporting of intervention effect | qualitative statement |
| Measure of effect | cases, reproduction number |
| Reporting of uncertainty | – not applicable – |
| Sensitivity analysis | different or modified outcome used (percentage of imported cases varied) |
| Subgroup assessment | none |

**Pullano et al. 2020 (Analysis 1)**

| | |
|---|---|
| **Study information** | |
| Author | Pullano G, Valdano E, Scarpa N, Rubrichi S, Colizza V |

– continued on next page –

| | |
|---|---|
| Title | Evaluating the Effect of Demographic Factors, Socioeconomic Factors, and Risk Aversion on Mobility during the COVID-19 Epidemic in France under Lockdown: A Population-Based Study |
| Year | 2020 |
| Journal | The Lancet Digital Health |
| DOI | 10.1016/s2589-7500(20)30243-0 |
| Date received | – could not be evaluated – |
| Date accepted | 2020-10-28 (published) |
| **Study setting** | |
| Number of populations included | multiple |
| Level of populations included | both national and subnational (France, regions) |
| Geographic areas covered | Europe |
| Number of countries covered | single country (France) |
| Study period | start and end date span first epidemic wave: behavioral |
| **Outcome** | |
| Raw outcome | behavioral outcome (mobility: number of trips) |
| Time resolution of raw outcome | daily |
| Computed outcome | none |
| Method to obtain the computed outcome | – not applicable – |
| Data source | mobility data from corporate organizations (Orange Business Service Flux Vision) |
| Data availability | data not accessible |
| **Intervention** | |
| Terminology for interventions | – not applicable – |

– continued on next page –

| | |
|---|---|
| Terminology for the specific type of non-pharmaceutical interventions | lockdown |
| Exposure types | phases of interventions (phase 2: school closure, gathering bans, public transport bans, phase 3: nationwide restrictions culminating in lockdown) |
| Types of single interventions | – not applicable – |
| Coding of interventions | not necessary |
| Source of intervention data | – could not be evaluated – |
| Availability of data on exposure | raw data documented in the manuscript |
| **Methodological approach** | |
| Empirical approach | descriptive |
| Use of exposure variation | only variation over time for multiple populations |
| Method | description of change over time (comparison of time periods) |
| Code availability | none |
| **Effect assessment** | |
| Reporting of intervention effect | quantification of change in outcome values |
| Measure of effect | mobility: number of trips |
| Reporting of uncertainty | no |
| Sensitivity analysis | none |
| Subgroup assessment | socioeconomic indicators (age) |

**Pullano et al. 2020 (Analysis 2)**

| | |
|---|---|
| **Study information** | |
| Author | Pullano G, Valdano E, Scarpa N, Rubrichi S, Colizza V |

– continued on next page –

| | |
|---|---|
| Title | Evaluating the Effect of Demographic Factors, Socioeconomic Factors, and Risk Aversion on Mobility during the COVID-19 Epidemic in France under Lockdown: A Population-Based Study |
| Year | 2020 |
| Journal | The Lancet Digital Health |
| DOI | 10.1016/s2589-7500(20)30243-0 |
| Date received | – could not be evaluated – |
| Date accepted | 2020-10-28 (published) |
| **Study setting** | |
| Number of populations included | multiple |
| Level of populations included | both national and subnational (France, regions) |
| Geographic areas covered | Europe |
| Number of countries covered | single country (France) |
| Study period | start and end date span first epidemic wave: behavioral |
| **Outcome** | |
| Raw outcome | behavioral outcome (mobility: number of trips) |
| Time resolution of raw outcome | daily |
| Computed outcome | none |
| Method to obtain the computed outcome | – not applicable – |
| Data source | mobility data from corporate organizations (Orange Business Service Flux Vision) |
| Data availability | data not accessible |
| **Intervention** | |
| Terminology for interventions | – not applicable – |

– continued on next page –

| | |
|---|---|
| Terminology for the specific type of non-pharmaceutical interventions | lockdown |
| Exposure types | phases of interventions (Combining phase 2: school closure, gathering bans, public transport bans, and phase 3: nationwide restrictions culminating in lockdown) |
| Types of single interventions | – not applicable – |
| Coding of interventions | not necessary |
| Source of intervention data | – could not be evaluated – |
| Availability of data on exposure | raw data documented in the manuscript |
| **Methodological approach** | |
| Empirical approach | counterfactual |
| Use of exposure variation | only variation over time for multiple populations |
| Method | non-mechanistic model (time series model) |
| Code availability | none |
| **Effect assessment** | |
| Reporting of intervention effect | quantification of change in outcome values |
| Measure of effect | mobility: number of trips |
| Reporting of uncertainty | yes |
| Sensitivity analysis | none |
| Subgroup assessment | socioeconomic indicators (age) |

**Pullano et al. 2020 (Analysis 3)**

| | |
|---|---|
| **Study information** | |
| Author | Pullano G, Valdano E, Scarpa N, Rubrichi S, Colizza V |

– continued on next page –

| | |
|---|---|
| Title | Evaluating the Effect of Demographic Factors, Socioeconomic Factors, and Risk Aversion on Mobility during the COVID-19 Epidemic in France under Lockdown: A Population-Based Study |
| Year | 2020 |
| Journal | The Lancet Digital Health |
| DOI | 10.1016/s2589-7500(20)30243-0 |
| Date received | – could not be evaluated – |
| Date accepted | 2020-10-28 (published) |
| **Study setting** | |
| Number of populations included | multiple |
| Level of populations included | both national and subnational (France, regions) |
| Geographic areas covered | Europe |
| Number of countries covered | single country (France) |
| Study period | start and end date span first epidemic wave: behavioral |
| **Outcome** | |
| Raw outcome | behavioral outcome (mobility: number of mobility connections between locations) |
| Time resolution of raw outcome | daily |
| Computed outcome | none |
| Method to obtain the computed outcome | – not applicable – |
| Data source | mobility data from corporate organizations (Orange Business Service Flux Vision) |
| Data availability | data not accessible |
| **Intervention** | |
| Terminology for interventions | – not applicable – |

– continued on next page –

| | |
|---|---|
| Terminology for the specific type of non-pharmaceutical interventions | lockdown |
| Exposure types | phases of interventions (phase 2: school closure, gathering bans, public transport bans, phase 3: nationwide restrictions culminating in lockdown) |
| Types of single interventions | – not applicable – |
| Coding of interventions | not necessary |
| Source of intervention data | – could not be evaluated – |
| Availability of data on exposure | raw data documented in the manuscript |
| **Methodological approach** | |
| Empirical approach | descriptive |
| Use of exposure variation | only variation over time for multiple populations |
| Method | description of change over time (comparison of time periods) |
| Code availability | none |
| **Effect assessment** | |
| Reporting of intervention effect | comparison of outcome values |
| Measure of effect | mobility: number of mobility connections between locations |
| Reporting of uncertainty | no |
| Sensitivity analysis | none |
| Subgroup assessment | none |

**Pung et al. 2020**

| | |
|---|---|
| **Study information** | |
| Author | Pung R, Cook AR, Chiew CJ, Clapham HE, Sun Y, Li Z, Dickens BL, Ma S, Mak K, Tan CC, Heng D, Chen MI, Lee VJ |
| Title | Effectiveness of Containment Measures against COVID-19 in Singapore |

– continued on next page –

| | |
|---|---|
| Year | 2020 |
| Journal | Epidemiology |
| DOI | 10.1097/ede.0000000000001257 |
| Date received | 2020-05-09 |
| Date accepted | 2020-09-03 |
| **Study setting** | |
| Number of populations included | single |
| Level of populations included | national (Singapore) |
| Geographic areas covered | Asia |
| Number of countries covered | single country (Singapore) |
| Study period | start and end date span first epidemic wave |
| **Outcome** | |
| Raw outcome | epidemiological individual-level outcome (individual cases and transmission chains) |
| Time resolution of raw outcome | daily |
| Computed outcome | epidemiological parameter (reproduction number) |
| Method to obtain the computed outcome | statistical estimation of reproduction number from individual data (Wallinga and Teunis, extended version with Bayesian data augmentation of transmission chains, incorporating individual data) |
| Data source | data from (sub)national authorities |
| Data availability | data not accessible |
| **Intervention** | |
| Terminology for interventions | measures |
| Terminology for the specific type of non-pharmaceutical interventions | containment |
| Exposure types | all interventions together |

– continued on next page –

| | |
|---|---|
| Types of single interventions | – not applicable – |
| Coding of interventions | – not applicable – |
| Source of intervention data | – not applicable – |
| Availability of data on exposure | – not applicable – |
| **Methodological approach** | |
| Empirical approach | descriptive |
| Use of exposure variation | only variation over time for a single population |
| Method | description of change over time (description of time course) |
| Code availability | none |
| **Effect assessment** | |
| Reporting of intervention effect | qualitative statement |
| Measure of effect | reproduction number |
| Reporting of uncertainty | – not applicable – |
| Sensitivity analysis | none |
| Subgroup assessment | none |

**Qiu et al. 2020 (Analysis 1)**

| | |
|---|---|
| **Study information** | |
| Author | Qiu Y, Chen X, Shi W |
| Title | Impacts of Social and Economic Factors on the Transmission of Coronavirus Disease 2019 (COVID-19) in China |
| Year | 2020 |
| Journal | Journal of Population Economics |

– continued on next page –

| | |
|---|---|
| DOI | 10.1007/s00148-020-00778-2 |
| Date received | – could not be evaluated – |
| Date accepted | 2020-05-09 (published) |
| **Study setting** | |
| Number of populations included | multiple |
| Level of populations included | both national and subnational (304 prefecture-level cities (China), excluding Wuhan) |
| Geographic areas covered | Asia |
| Number of countries covered | single country (China) |
| Study period | start and end date span first epidemic wave |
| **Outcome** | |
| Raw outcome | epidemiological population-level outcome (cases) |
| Time resolution of raw outcome | daily |
| Computed outcome | epidemiological parameter (reproduction number) |
| Method to obtain the computed outcome | other (fit regression model with lagged cases as predictor and interpret coefficient as transmission rate/reproduction number) |
| Data source | data from (sub)national authorities |
| Data availability | data access via source |
| **Intervention** | |
| Terminology for interventions | measures |
| Terminology for the specific type of non-pharmaceutical interventions | public health |
| Exposure types | one combination of interventions (Wuhan City Lockdown) |
| Types of single interventions | – not applicable – |
| Coding of interventions | not necessary |
| Source of intervention data | – could not be evaluated – |

– continued on next page –

| | |
|---|---|
| Availability of data on exposure | raw data documented in the manuscript |
| **Methodological approach** | |
| Empirical approach | descriptive |
| Use of exposure variation | only variation over time for multiple populations |
| Method | description of change over time (comparison of time periods) |
| Code availability | none |
| **Effect assessment** | |
| Reporting of intervention effect | comparison of outcome values |
| Measure of effect | reproduction number |
| Reporting of uncertainty | no |
| Sensitivity analysis | model specification varied |
| Subgroup assessment | none |

**Qiu et al. 2020 (Analysis 2)**

| | |
|---|---|
| **Study information** | |
| Author | Qiu Y, Chen X, Shi W |
| Title | Impacts of Social and Economic Factors on the Transmission of Coronavirus Disease 2019 (COVID-19) in China |
| Year | 2020 |
| Journal | Journal of Population Economics |
| DOI | 10.1007/s00148-020-00778-2 |
| Date received | – could not be evaluated – |
| Date accepted | 2020-05-09 (published) |
| **Study setting** | |

– continued on next page –

| | |
|---|---|
| Number of populations included | multiple |
| Level of populations included | both national and subnational (<300 prefecture-level cities (China), excluded cities in Hubei) |
| Geographic areas covered | Asia |
| Number of countries covered | single country (China) |
| Study period | other (starts with start of NPIs) |
| **Outcome** | |
| Raw outcome | epidemiological population-level outcome (cases) |
| Time resolution of raw outcome | daily |
| Computed outcome | none |
| Method to obtain the computed outcome | – not applicable – |
| Data source | data from (sub)national authorities |
| Data availability | data access via source |
| **Intervention** | |
| Terminology for interventions | measures |
| Terminology for the specific type of non-pharmaceutical interventions | public health |
| Exposure types | multiple separate interventions (bans of small gatherings (closed management of communities), stay-at-home order) |
| Types of single interventions | bans of small gatherings, stay-at-home order |
| Coding of interventions | not necessary |
| Source of intervention data | – could not be evaluated – |
| Availability of data on exposure | raw data documented in the manuscript |
| **Methodological approach** | |
| Empirical approach | parametric |

– continued on next page –

| | |
|---|---|
| Use of exposure variation | both variation over time and between populations |
| Method | non-mechanistic model (generalized linear model) |
| Code availability | none |
| **Effect assessment** | |
| Reporting of intervention effect | quantification of change in outcome values |
| Measure of effect | cases |
| Reporting of uncertainty | yes |
| Sensitivity analysis | model specification varied |
| Subgroup assessment | none |

**Quaife et al. 2020**

| | |
|---|---|
| **Study information** | |
| Author | Quaife M, van Zandvoort K, Gimma A, Shah K, McCreesh N, Prem K, Barasa E, Mwanga D, Kangwana B, Pinchoff J, Bosse NI, Medley G, O'Reilly K, Leclerc QJ, Jit M, Lowe R, Davies NG, Deol AK, Knight GM, Auzenbergs M, Edmunds WJ, Atkins KE, Jombart T, Eggo RM, Foss AM, Flasche S, Pearson CA, Nightingale ES, Munday JD, Abbott S, Quilty BJ, Gibbs HP, Simons D, Emery JC, Hué S, Liu Y, Tully DC, Rosello A, Procter SR, Endo A, Russell TW, Rees EM, Diamond C, Meakin SR, Kucharski AJ, Gore-Langton GR, Klepac P, Hellewell J, Sun FY, Houben RM, Villabona-Arenas CJ, Funk S, Clifford S, Brady O, Edmunds WJ, Jarvis CI, And KA |
| Title | The Impact of COVID-19 Control Measures on Social Contacts and Transmission in Kenyan Informal Settlements |
| Year | 2020 |
| Journal | BMC Medicine |

– continued on next page –

| | |
|---|---|
| DOI | 10.1186/s12916-020-01779-4 |
| Date received | 2020-06-09 |
| Date accepted | 2020-09-09 |
| **Study setting** | |
| Number of populations included | single |
| Level of populations included | subnational (informal settlements around Nairobi, Kenia) |
| Geographic areas covered | Middle East and Africa |
| Number of countries covered | single country (Kenia) |
| Study period | other (surveys conducted only in May) |
| **Outcome** | |
| Raw outcome | behavioral outcome (survey: number of physical and non-physical contacts) |
| Time resolution of raw outcome | – not applicable – |
| Computed outcome | epidemiological parameter (reproduction number) |
| Method to obtain the computed outcome | statistical estimation of reproduction number from individual data (Diekmann et al., contact matrices estimated from survey responses) |
| Data source | data collected by authors |
| Data availability | data made available by the authors |
| **Intervention** | |
| Terminology for interventions | measures |
| Terminology for the specific type of non-pharmaceutical interventions | control |
| Exposure types | all interventions together |
| Types of single interventions | – not applicable – |
| Coding of interventions | – not applicable – |
| Source of intervention data | – not applicable – |

– continued on next page –

| | |
|---|---|
| Availability of data on exposure | – not applicable – |
| **Methodological approach** | |
| Empirical approach | descriptive |
| Use of exposure variation | only variation over time for a single population |
| Method | description of change over time (comparison of time periods) |
| Code availability | publicly available |
| **Effect assessment** | |
| Reporting of intervention effect | comparison of outcome values |
| Measure of effect | reproduction number |
| Reporting of uncertainty | yes |
| Sensitivity analysis | model specification varied |
| Subgroup assessment | none |

**Rahman et al. 2020 (Analysis 1)**

| | |
|---|---|
| **Study information** | |
| Author | Rahman MM, Thill JC, Paul KC |
| Title | COVID-19 Pandemic Severity, Lockdown Regimes, and People's Mobility: Early Evidence from 88 Countries |
| Year | 2020 |
| Journal | Sustainability |
| DOI | 10.3390/su12219101 |
| Date received | 2020-10-02 |
| Date accepted | 2020-10-30 |

– continued on next page –

| | |
|---|---|
| **Study setting** | |
| Number of populations included | multiple |
| Level of populations included | national (88 countries around the globe) |
| Geographic areas covered | Europe, Middle East and Africa, Asia, Oceania, Central and South America, North America |
| Number of countries covered | multiple countries |
| Study period | start and end date span first epidemic wave: behavioral |
| **Outcome** | |
| Raw outcome | behavioral outcome (mobility: time spent in various locations) |
| Time resolution of raw outcome | daily |
| Computed outcome | none |
| Method to obtain the computed outcome | – not applicable – |
| Data source | mobility data from corporate organizations (Google) |
| Data availability | data access via source |
| **Intervention** | |
| Terminology for interventions | – not applicable – |
| Terminology for the specific type of non-pharmaceutical interventions | lockdown |
| Exposure types | one combination of interventions (lockdown measures (e.g., restriction on public gathering, workplace closing, and stay-at-home order) with specific dates) |
| Types of single interventions | – not applicable – |
| Coding of interventions | necessary |
| Source of intervention data | use of externally coded data (OxCGRT, and same as in Dunford et al.) |
| Availability of data on exposure | access to externally coded data via source |

– continued on next page –

| | |
|---|---|
| **Methodological approach** | |
| Empirical approach | parametric |
| Use of exposure variation | both variation over time and between populations |
| Method | non-mechanistic model (other: structural equation model) |
| Code availability | none |
| **Effect assessment** | |
| Reporting of intervention effect | quantification of change in outcome values |
| Measure of effect | mobility: time spent in various locations |
| Reporting of uncertainty | yes |
| Sensitivity analysis | none |
| Subgroup assessment | none |

**Rahman et al. 2020 (Analysis 2, mobility as exposure)**

| | |
|---|---|
| **Study information** | |
| Author | Rahman MM, Thill JC, Paul KC |
| Title | COVID-19 Pandemic Severity, Lockdown Regimes, and People's Mobility: Early Evidence from 88 Countries |
| Year | 2020 |
| Journal | Sustainability |
| DOI | 10.3390/su12219101 |
| Date received | 02.10.2020 |
| Date accepted | 30.10.2020 |
| **Study setting** | |
| Number of populations included | multiple |
| Level of populations included | national (88 countries around the globe) |

– continued on next page –

| | |
|---|---|
| Geographic areas covered | Europe, Middle East and Africa, Asia, Oceania, Central and South America, North America |
| Number of countries covered | multiple countries |
| Study period | same end date for several populations with diverse epidemic trajectories |
| **Outcome** | |
| Raw outcome | epidemiological population-level outcome (cases, deaths) |
| Time resolution of raw outcome | daily |
| Computed outcome | none |
| Method to obtain the computed outcome | – not applicable – |
| Data source | data from publicly available cross-country selections (Worldometer) |
| Data availability | data access via source |
| **Intervention** | |
| Terminology for interventions | – not applicable – |
| Terminology for the specific type of non-pharmaceutical interventions | lockdown |
| Exposure types | mobility (time spent in various locations) |
| Types of single interventions | – not applicable – |
| Coding of interventions | – not applicable – |
| Source of intervention data | mobility data by corporate organizations (Google) |
| Availability of data on exposure | data access via source |
| **Methodological approach** | |
| Empirical approach | parametric |
| Use of exposure variation | both variation over time and between populations |
| Method | non-mechanistic model (generalized linear model) |
| Code availability | none |

– continued on next page –

| | |
|---|---|
| **Effect assessment** | |
| Reporting of intervention effect | quantification of change in outcome values |
| Measure of effect | cases, deaths |
| Reporting of uncertainty | yes |
| Sensitivity analysis | none |
| Subgroup assessment | none |

**Reis et al. 2020**

| | |
|---|---|
| **Study information** | |
| Author | Reis RF, de Melo Quintela B, de Oliveira Campos J, Gomes JM, Rocha BM, Lobosco M, dos Santos RW |
| Title | Characterization of the COVID-19 Pandemic and the Impact of Uncertainties, Mitigation Strategies, and Underreporting of Cases in South Korea, Italy, and Brazil |
| Year | 2020 |
| Journal | Chaos, Solitons & Fractals |
| DOI | 10.1016/j.chaos.2020.109888 |
| Date received | 2020-04-11 |
| Date accepted | 2020-05-10 |
| **Study setting** | |
| Number of populations included | multiple |
| Level of populations included | national (Brazil, Italy, South Korea) |
| Geographic areas covered | Asia, Central and South America, Europe |
| Number of countries covered | multiple countries |

– continued on next page –

| | |
|---|---|
| Study period | end date in growth phase of wave |
| **Outcome** | |
| Raw outcome | epidemiological population-level outcome (cases, deaths) |
| Time resolution of raw outcome | daily |
| Computed outcome | none |
| Method to obtain the computed outcome | – not applicable – |
| Data source | data from publicly available cross-country selections (JHU) |
| Data availability | data made available by the authors |
| **Intervention** | |
| Terminology for interventions | measures |
| Terminology for the specific type of non-pharmaceutical interventions | mitigation |
| Exposure types | all interventions together |
| Types of single interventions | – not applicable – |
| Coding of interventions | – not applicable – |
| Source of intervention data | – not applicable – |
| Availability of data on exposure | – not applicable – |
| **Methodological approach** | |
| Empirical approach | parametric |
| Use of exposure variation | only variation over time for multiple populations |
| Method | mechanistic model (compartmental single-population transmission model) |
| Latent variable functionally linked to intervention | transmission rate |
| Code availability | publicly available |
| **Effect assessment** | |

– continued on next page –

| | |
|---|---|
| Reporting of intervention effect | quantification of change in outcome values |
| Measure of effect | transmission rate |
| Reporting of uncertainty | yes |
| Sensitivity analysis | none |
| Subgroup assessment | none |

**Riccardo et al. 2020**

| | |
|---|---|
| **Study information** | |
| Author | Riccardo F, Ajelli M, Andrianou XD, Bella A, Manso M, Fabiani M, Bellino S, Boros S, Urdiales AM, Marziano V, Rota MC, Filia A, DAncona F, Siddu A, Punzo O, Trentini F, Guzzetta G, Poletti P, Stefanelli P, Castrucci MR, Ciervo A, Benedetto CD, Tallon M, Piccioli A, Brusaferro S, Rezza G, Merler S, And PP |
| Title | Epidemiological Characteristics of COVID-19 Cases and Estimates of the Reproductive Numbers 1 Month into the Epidemic, Italy, 28 January to 31 March 2020 |
| Year | 2020 |
| Journal | Eurosurveillance |
| DOI | 10.2807/1560-7917.es.2020.25.49.2000790 |
| Date received | 2020-05-03 |
| Date accepted | 2020-08-14 |
| **Study setting** | |
| Number of populations included | multiple |
| Level of populations included | both national and subnational (Italy and its regions) |
| Geographic areas covered | Europe |
| Number of countries covered | single country (Italy) |

– continued on next page –

| | |
|---|---|
| Study period | start and end date span first epidemic wave |
| **Outcome** | |
| Raw outcome | epidemiological population-level outcome (cases) |
| Time resolution of raw outcome | daily |
| Computed outcome | epidemiological parameter (reproduction number) |
| Method to obtain the computed outcome | statistical estimation of reproduction number (Cori et al.) |
| Data source | data from (sub)national authorities |
| Data availability | data access via source |
| **Intervention** | |
| Terminology for interventions | measures |
| Terminology for the specific type of non-pharmaceutical interventions | non-pharmacological |
| Exposure types | all interventions together (timeline of measures) |
| Types of single interventions | – not applicable – |
| Coding of interventions | – not applicable – |
| Source of intervention data | – not applicable – |
| Availability of data on exposure | – not applicable – |
| **Methodological approach** | |
| Empirical approach | descriptive |
| Use of exposure variation | only variation over time for multiple populations |
| Method | description of change over time (description of time course) |
| Code availability | none |
| **Effect assessment** | |
| Reporting of intervention effect | qualitative statement |
| Measure of effect | reproduction number |

– continued on next page –

| | |
|---|---|
| Reporting of uncertainty | – not applicable – |
| Sensitivity analysis | none |
| Subgroup assessment | none |

| **Rieger & Wang 2020** | |
|---|---|
| **Study information** | |
| Author | Rieger MO, Wang M |
| Title | Secret Erosion of the "Lockdown"? Patterns in Daily Activities during the SARS-Cov2 Pandemics around the World |
| Year | 2020 |
| Journal | Review of Behavioral Economics |
| DOI | 10.1561/105.00000124 |
| Date received | – could not be evaluated – |
| Date accepted | 2020-08-10 (published) |
| **Study setting** | |
| Number of populations included | multiple |
| Level of populations included | national (Germany, France, UK, US) |
| Geographic areas covered | Europe, North America |
| Number of countries covered | multiple countries |
| Study period | start and end date span first epidemic wave: behavioral |
| **Outcome** | |
| Raw outcome | behavioral outcome (mobility: number of trips by mode & time spent in various locations & intention) |
| Time resolution of raw outcome | daily |

– continued on next page –

| | |
|---|---|
| Computed outcome | none |
| Method to obtain the computed outcome | – not applicable – |
| Data source | mobility data from corporate organizations (Apple: number of trips by mode, Google: time spent in various locations), data from other research projects (study by Fetzer et al.: intention) |
| Data availability | data access via source |
| **Intervention** | |
| Terminology for interventions | – not applicable – |
| Terminology for the specific type of non-pharmaceutical interventions | lockdown |
| Exposure types | one combination of interventions (lockdown with specific date for each country) |
| Types of single interventions | – not applicable – |
| Coding of interventions | not necessary |
| Source of intervention data | – could not be evaluated – |
| Availability of data on exposure | raw data documented in the manuscript |
| **Methodological approach** | |
| Empirical approach | parametric |
| Use of exposure variation | only variation over time for multiple populations |
| Method | non-mechanistic model (correlation) |
| Code availability | none |
| **Effect assessment** | |
| Reporting of intervention effect | quantification of change in outcome values |
| Measure of effect | mobility: number of trips by mode & time spent in various locations & intention |
| Reporting of uncertainty | yes |

– continued on next page –

| | |
|---|---|
| Sensitivity analysis | none |
| Subgroup assessment | none |

| **Ryu et al. 2020** | |
|---|---|
| **Study information** | |
| Author | Ryu S, Ali ST, Jang C, Kim B, Cowling BJ |
| Title | Effect of Nonpharmaceutical Interventions on Transmission of Severe Acute Respiratory Syndrome Coronavirus 2, South Korea, 2020 |
| Year | 2020 |
| Journal | Emerging Infectious Diseases |
| DOI | 10.3201/eid2610.201886 |
| Date received | – could not be evaluated – |
| Date accepted | 2020-10-15 (published) |
| **Study setting** | |
| Number of populations included | single |
| Level of populations included | national (South Korea) |
| Geographic areas covered | Asia |
| Number of countries covered | single country (South Korea) |
| Study period | start and end date span first epidemic wave |
| **Outcome** | |
| Raw outcome | epidemiological individual-level outcome (individual cases and transmission chains) |
| Time resolution of raw outcome | daily |
| Computed outcome | epidemiological parameter (reproduction number) |

– continued on next page –

| | |
|---|---|
| Method to obtain the computed outcome | statistical estimation of reproduction number (Cori et al.) |
| Data source | data from (sub)national authorities |
| Data availability | data not accessible |
| **Intervention** | |
| Terminology for interventions | interventions |
| Terminology for the specific type of non-pharmaceutical interventions | non-pharmaceutical |
| Exposure types | one single intervention (emergency declaration (declaration of highest public alert)) |
| Types of single interventions | declaration of a state of emergency |
| Coding of interventions | not necessary |
| Source of intervention data | another study (Kim et al.) |
| Availability of data on exposure | raw data documented in the manuscript |
| **Methodological approach** | |
| Empirical approach | descriptive |
| Use of exposure variation | only variation over time for a single population |
| Method | description of change over time (comparison of time periods) |
| Code availability | none |
| **Effect assessment** | |
| Reporting of intervention effect | quantification of change in outcome values |
| Measure of effect | reproduction number |
| Reporting of uncertainty | yes |
| Sensitivity analysis | none |
| Subgroup assessment | none |

| Saez et al. 2020 | |
|---|---|
| **Study information** | |
| Author | Saez M, Tobias A, Varga D, Barceló MA |
| Title | Effectiveness of the Measures to Flatten the Epidemic Curve of COVID-19. The Case of Spain |
| Year | 2020 |
| Journal | Science of The Total Environment |
| DOI | 10.1016/j.scitotenv.2020.138761 |
| Date received | 2020-04-06 |
| Date accepted | 2020-04-15 |
| **Study setting** | |
| Number of populations included | single |
| Level of populations included | national (Spain) |
| Geographic areas covered | Europe |
| Number of countries covered | single country (Spain) |
| Study period | start and end date span first epidemic wave |
| **Outcome** | |
| Raw outcome | epidemiological population-level outcome (cases) |
| Time resolution of raw outcome | daily |
| Computed outcome | measure of epidemic trend (growth rate) |
| Method to obtain the computed outcome | simple computation |
| Data source | data from (sub)national authorities |
| Data availability | data access via source |
| **Intervention** | |

– continued on next page –

| | |
|---|---|
| Terminology for interventions | measures |
| Terminology for the specific type of non-pharmaceutical interventions | physical distancing |
| Exposure types | one combination of interventions (confinement of the population by decree of the Spanish Government on March 14) |
| Types of single interventions | – not applicable – |
| Coding of interventions | not necessary |
| Source of intervention data | – could not be evaluated – |
| Availability of data on exposure | raw data documented in the manuscript |
| **Methodological approach** | |
| Empirical approach | parametric |
| Use of exposure variation | only variation over time for a single population |
| Method | non-mechanistic model (generalized linear model) |
| Code availability | none |
| **Effect assessment** | |
| Reporting of intervention effect | quantification of change in outcome values |
| Measure of effect | growth rate |
| Reporting of uncertainty | yes |
| Sensitivity analysis | none |
| Subgroup assessment | none |

**Salje et al. 2020**

**Study information**

– continued on next page –

| | |
|---|---|
| Author | Salje H, Kiem CT, Lefrancq N, Courtejoie N, Bosetti P, Paireau J, Andronico A, Hozé N, Richet J, Dubost CL, Strat YL, Lessler J, Levy-Bruhl D, Fontanet A, Opatowski L, Boelle PY, Cauchemez S |
| Title | Estimating the Burden of SARS-CoV-2 in France |
| Year | 2020 |
| Journal | Science |
| DOI | 10.1126/science.abc3517 |
| Date received | 2020-04-20 |
| Date accepted | 2020-05-11 |
| **Study setting** | |
| Number of populations included | single |
| Level of populations included | national (France) |
| Geographic areas covered | Europe |
| Number of countries covered | single country (France) |
| Study period | start and end date span first epidemic wave |
| **Outcome** | |
| Raw outcome | epidemiological population-level outcome (hospitalizations, deaths) |
| Time resolution of raw outcome | daily |
| Computed outcome | none |
| Method to obtain the computed outcome | – not applicable – |
| Data source | data from (sub)national authorities |
| Data availability | data made available by the authors |
| **Intervention** | |
| Terminology for interventions | – not applicable – |

– continued on next page –

| | |
|---|---|
| Terminology for the specific type of non-pharmaceutical interventions | lockdown |
| Exposure types | one single intervention (stay-at-home order) |
| Types of single interventions | stay-at-home order |
| Coding of interventions | not necessary |
| Source of intervention data | – could not be evaluated – |
| Availability of data on exposure | raw data documented in the manuscript |
| **Methodological approach** | |
| Empirical approach | parametric |
| Use of exposure variation | only variation over time for a single population |
| Method | mechanistic model (compartmental single-population transmission model) |
| Latent variable functionally linked to intervention | transmission rate |
| Code availability | publicly available |
| **Effect assessment** | |
| Reporting of intervention effect | quantification of change in outcome values |
| Measure of effect | transmission rate, reproduction number |
| Reporting of uncertainty | yes |
| Sensitivity analysis | epidemiological parameters varied |
| Subgroup assessment | none |

**Salvatore et al. 2020**

| | |
|---|---|
| **Study information** | |
| Author | Salvatore M, Basu D, Ray D, Kleinsasser M, Purkayastha S, Bhattacharyya R, Mukherjee B |

– continued on next page –

| | |
|---|---|
| Title | Comprehensive Public Health Evaluation of Lockdown as a Non-Pharmaceutical Intervention on COVID-19 Spread in India: National Trends Masking State-Level Variations |
| Year | 2020 |
| Journal | BMJ Open |
| DOI | 10.1136/bmjopen-2020-041778 |
| Date received | 2020-06-17 |
| Date accepted | 2020-11-11 |
| **Study setting** | |
| Number of populations included | multiple |
| Level of populations included | both national and subnational (India and 20 states of India) |
| Geographic areas covered | Asia |
| Number of countries covered | single country (India) |
| Study period | end date in growth phase of wave |
| **Outcome** | |
| Raw outcome | epidemiological population-level outcome (cases) |
| Time resolution of raw outcome | daily |
| Computed outcome | measure of epidemic trend (doubling time), epidemiological parameter (reproduction number) |
| Method to obtain the computed outcome | statistical estimation of reproduction number (Cori et al.), exponential growth model (doubling time) |
| Data source | data from publicly available open-source projects (covid19india.org: cases), data from publicly available cross-country selections (OurWorldInData: cases) |
| Data availability | data made available by the authors |
| **Intervention** | |
| Terminology for interventions | interventions |

– continued on next page –

| | |
|---|---|
| Terminology for the specific type of non-pharmaceutical interventions | non-pharmaceutical |
| Exposure types | one combination of interventions (Indian National Lockdown) |
| Types of single interventions | – not applicable – |
| Coding of interventions | not necessary |
| Source of intervention data | government or news websites |
| Availability of data on exposure | raw data documented in the manuscript |
| **Methodological approach** | |
| Empirical approach | descriptive |
| Use of exposure variation | only variation over time for multiple populations |
| Method | description of change over time (comparison of time periods) |
| Code availability | publicly available |
| **Effect assessment** | |
| Reporting of intervention effect | comparison of outcome values |
| Measure of effect | reproduction number, doubling time |
| Reporting of uncertainty | yes |
| Sensitivity analysis | none |
| Subgroup assessment | none |

**Santamaria et al. 2020**

| | |
|---|---|
| **Study information** | |
| Author | Santamaria C, Sermi F, Spyratos S, Iacus SM, Annunziato A, Tarchi D, Vespe M |
| Title | Measuring the Impact of COVID-19 Confinement Measures on Human Mobility Using Mobile Positioning Data. A European Regional Analysis |

– continued on next page –

| | |
|---|---|
| Year | 2020 |
| Journal | Safety Science |
| DOI | 10.1016/j.ssci.2020.104925 |
| Date received | 2020-07-09 |
| Date accepted | 2020-07-17 |
| **Study setting** | |
| Number of populations included | multiple |
| Level of populations included | national (14 European countries) |
| Geographic areas covered | Europe |
| Number of countries covered | multiple countries |
| Study period | start and end date span first epidemic wave: behavioral |
| **Outcome** | |
| Raw outcome | behavioral outcome (mobility: incoming and outgoing travels in a region) |
| Time resolution of raw outcome | daily |
| Computed outcome | none |
| Method to obtain the computed outcome | – not applicable – |
| Data source | mobility data from corporate organizations (European Mobile Network Operators) |
| Data availability | data not accessible |
| **Intervention** | |
| Terminology for interventions | measures |
| Terminology for the specific type of non-pharmaceutical interventions | confinement |

– continued on next page –

| | |
|---|---|
| Exposure types | multiple separate interventions (school closure, bans of large gatherings, bans of small gatherings, workplace closure, stay-at-home order, international travel restrictions, internal movement restrictions, public transport closure) |
| Types of single interventions | school closure, bans of large gatherings, bans of small gatherings, workplace closure, stay-at-home order, international travel restrictions, internal movement restrictions, public transport closure |
| Coding of interventions | necessary |
| Source of intervention data | use of externally coded data (OxCGRT) |
| Availability of data on exposure | access to externally coded data via source |
| **Methodological approach** | |
| Empirical approach | parametric |
| Use of exposure variation | both variation over time and between populations |
| Method | non-mechanistic model (generalized linear model) |
| Code availability | none |
| **Effect assessment** | |
| Reporting of intervention effect | quantification of change in outcome values |
| Measure of effect | mobility: mobility index of number of movements |
| Reporting of uncertainty | yes |
| Sensitivity analysis | none |
| Subgroup assessment | none |

**Saurabh et al. 2020**

**Study information**

– continued on next page –

| | |
|---|---|
| Author | Saurabh S, Verma MK, Gautam V, Kumar N, Goel AD, Gupta MK, Bhardwaj P, Misra S |
| Title | Transmission Dynamics of the COVID-19 Epidemic at the District Level in India: Prospective Observational Study |
| Year | 2020 |
| Journal | JMIR Public Health and Surveillance |
| DOI | 10.2196/22678 |
| Date received | 2020-07-21 |
| Date accepted | 2020-09-09 |
| **Study setting** | |
| Number of populations included | single |
| Level of populations included | subnational (Jodhpur district (India)) |
| Geographic areas covered | Asia |
| Number of countries covered | single country (India) |
| Study period | end date in growth phase of wave |
| **Outcome** | |
| Raw outcome | epidemiological individual-level outcome (individual cases and transmission chains, cases) |
| Time resolution of raw outcome | daily |
| Computed outcome | epidemiological parameter (reproduction number) |
| Method to obtain the computed outcome | statistical estimation of reproduction number from individual data (Simplified version of Cori et al., Wallinga and Teunis) |
| Data source | data from publicly available open-source projects (covid19india.org) |
| Data availability | data access via source |
| **Intervention** | |
| Terminology for interventions | measures |

– continued on next page –

| | |
|---|---|
| Terminology for the specific type of non-pharmaceutical interventions | control |
| Exposure types | all interventions together |
| Types of single interventions | – not applicable – |
| Coding of interventions | – not applicable – |
| Source of intervention data | – not applicable – |
| Availability of data on exposure | – not applicable – |
| **Methodological approach** | |
| Empirical approach | descriptive |
| Use of exposure variation | only variation over time for a single population |
| Method | description of change over time (description of time course) |
| Code availability | none |
| **Effect assessment** | |
| Reporting of intervention effect | qualitative statement |
| Measure of effect | reproduction number |
| Reporting of uncertainty | – not applicable – |
| Sensitivity analysis | none |
| Subgroup assessment | none |

**Scarabel et al. 2020**

| | |
|---|---|
| **Study information** | |
| Author | Scarabel F, Pellis L, Bragazzi NL, Wu J |
| Title | Canada Needs to Rapidly Escalate Public Health Interventions for Its COVID-19 Mitigation Strategies |

– continued on next page –

| | |
|---|---|
| Year | 2020 |
| Journal | Infectious Disease Modelling |
| DOI | 10.1016/j.idm.2020.03.004 |
| Date received | 2020-03-25 |
| Date accepted | 2020-03-30 |
| **Study setting** | |
| Number of populations included | single |
| Level of populations included | national (Italy) |
| Geographic areas covered | Europe |
| Number of countries covered | single country (Italy) |
| Study period | end date in growth phase of wave |
| **Outcome** | |
| Raw outcome | epidemiological population-level outcome (cases) |
| Time resolution of raw outcome | daily |
| Computed outcome | measure of epidemic trend (growth rate) |
| Method to obtain the computed outcome | exponential growth model |
| Data source | data from publicly available cross-country selections (ECDC) |
| Data availability | data access via source |
| **Intervention** | |
| Terminology for interventions | interventions |
| Terminology for the specific type of non-pharmaceutical interventions | public health |
| Exposure types | multiple combinations of interventions (lockdown (movement restrictions), closure of non-essential businesses) |
| Types of single interventions | – not applicable – |

– continued on next page –

| | |
|---|---|
| Coding of interventions | not necessary |
| Source of intervention data | – could not be evaluated – |
| Availability of data on exposure | raw data documented in the manuscript |
| **Methodological approach** | |
| Empirical approach | descriptive |
| Use of exposure variation | only variation over time for a single population |
| Method | description of change over time (comparison of time periods) |
| Code availability | none |
| **Effect assessment** | |
| Reporting of intervention effect | comparison of outcome values |
| Measure of effect | growth rate |
| Reporting of uncertainty | no |
| Sensitivity analysis | none |
| Subgroup assessment | none |

**Sebastian et al. 2020 (Analysis 1)**

| | |
|---|---|
| **Study information** | |
| Author | Sebastian S, Paul A, Joby J, Saijan S, Joseph J, Kunjumon J |
| Title | Impact of Lockdown in India: A Case Study on Karnataka with International Model |
| Year | 2020 |
| Journal | Kesmas: National Public Health Journal |
| DOI | 10.21109/kesmas.v15i2.3978 |

– continued on next page –

| | |
|---|---|
| Date received | 2020-07-02 |
| Date accepted | 2020-07-03 |
| **Study setting** | |
| Number of populations included | multiple |
| Level of populations included | both national and subnational (Karnataka and Pubjab (India), Australia, UK) |
| Geographic areas covered | Asia, Europe, Oceania |
| Number of countries covered | multiple countries |
| Study period | end date could not be evaluated |
| **Outcome** | |
| Raw outcome | epidemiological population-level outcome (cases, recovered cases, deaths) |
| Time resolution of raw outcome | daily |
| Computed outcome | measure of epidemic trend (growth rate, growth rate: recovered, growth rate: deaths, doubling time) |
| Method to obtain the computed outcome | simple computation (growth rate), exponential growth model (doubling time) |
| Data source | data from publicly available cross-country selections (WHO) |
| Data availability | data access via source |
| **Intervention** | |
| Terminology for interventions | measures |
| Terminology for the specific type of non-pharmaceutical interventions | social distancing |
| Exposure types | one combination of interventions (Indian National Lockdown) |
| Types of single interventions | – not applicable – |
| Coding of interventions | not necessary |
| Source of intervention data | – could not be evaluated – |

– continued on next page –

| | |
|---|---|
| Availability of data on exposure | raw data documented in the manuscript |
| **Methodological approach** | |
| Empirical approach | descriptive |
| Use of exposure variation | only variation over time for multiple populations |
| Method | description of change over time (comparison of time periods) |
| Code availability | none |
| **Effect assessment** | |
| Reporting of intervention effect | quantification of change in outcome values |
| Measure of effect | growth rate, growth rate: recovered, growth rate: deaths, doubling time |
| Reporting of uncertainty | no |
| Sensitivity analysis | none |
| Subgroup assessment | none |

| **Sebastian et al. 2020 (Analysis 2)** | |
|---|---|
| **Study information** | |
| Author | Sebastian S, Paul A, Joby J, Saijan S, Joseph J, Kunjumon J |
| Title | Impact of Lockdown in India: A Case Study on Karnataka with International Model |
| Year | 2020 |
| Journal | Kesmas: National Public Health Journal |
| DOI | 10.21109/kesmas.v15i2.3978 |
| Date received | 2020-07-02 |
| Date accepted | 2020-07-03 |
| **Study setting** | |

– continued on next page –

| | |
|---|---|
| Number of populations included | multiple |
| Level of populations included | both national and subnational (Karnataka and Pubjab (India), Australia, UK) |
| Geographic areas covered | Asia, Europe, Oceania |
| Number of countries covered | multiple countries |
| Study period | end date could not be evaluated |
| **Outcome** | |
| Raw outcome | epidemiological population-level outcome (cases) |
| Time resolution of raw outcome | daily |
| Computed outcome | none |
| Method to obtain the computed outcome | – not applicable – |
| Data source | data from publicly available cross-country selections (WHO) |
| Data availability | data access via source |
| **Intervention** | |
| Terminology for interventions | measures |
| Terminology for the specific type of non-pharmaceutical interventions | social distancing |
| Exposure types | one combination of interventions (Indian National Lockdown) |
| Types of single interventions | – not applicable – |
| Coding of interventions | not necessary |
| Source of intervention data | – could not be evaluated – |
| Availability of data on exposure | raw data documented in the manuscript |
| **Methodological approach** | |
| Empirical approach | counterfactual |
| Use of exposure variation | only variation over time for multiple populations |

– continued on next page –

**Sebastian et al. 2020 (Analysis 2)** – continued from previous page

| | |
|---|---|
| Method | non-mechanistic model (exponential growth model) |
| Code availability | none |
| **Effect assessment** | |
| Reporting of intervention effect | quantification of change in outcome values |
| Measure of effect | cases |
| Reporting of uncertainty | no |
| Sensitivity analysis | none |
| Subgroup assessment | none |

**Sebastiani et al. 2020**

| | |
|---|---|
| **Study information** | |
| Author | Sebastiani G, Massa M, Riboli E |
| Title | COVID-19 Epidemic in Italy: Evolution, Projections and Impact of Government Measures |
| Year | 2020 |
| Journal | European Journal of Epidemiology |
| DOI | 10.1007/s10654-020-00631-6 |
| Date received | 2020-04-06 |
| Date accepted | 2020-04-08 |
| **Study setting** | |
| Number of populations included | multiple |
| Level of populations included | both national and subnational (Italy and 107 provinces) |
| Geographic areas covered | Europe |
| Number of countries covered | single country (Italy) |

– continued on next page –

| | |
|---|---|
| Study period | end date at peak of wave |
| **Outcome** | |
| Raw outcome | epidemiological population-level outcome (cases) |
| Time resolution of raw outcome | daily |
| Computed outcome | none |
| Method to obtain the computed outcome | – not applicable – |
| Data source | data from (sub)national authorities |
| Data availability | data access via source |
| **Intervention** | |
| Terminology for interventions | measures |
| Terminology for the specific type of non-pharmaceutical interventions | government |
| Exposure types | all interventions together (lockdown (quarantining, movement restrictions, school and venue closure etc.)) |
| Types of single interventions | – not applicable – |
| Coding of interventions | – not applicable – |
| Source of intervention data | – not applicable – |
| Availability of data on exposure | – not applicable – |
| **Methodological approach** | |
| Empirical approach | counterfactual |
| Use of exposure variation | only variation over time for multiple populations |
| Method | non-mechanistic model (exponential growth model) |
| Code availability | none |
| **Effect assessment** | |
| Reporting of intervention effect | qualitative statement |

– continued on next page –

| | |
|---|---|
| Measure of effect | cases |
| Reporting of uncertainty | – not applicable – |
| Sensitivity analysis | none |
| Subgroup assessment | none |

| **Seemann et al. 2020** | |
|---|---|
| **Study information** | |
| Author | Seemann T, Lane CR, Sherry NL, Duchene S, Gonçalves da Silva A, Caly L, Sait M, Ballard SA, Horan K, Schultz MB, Hoang T, Easton M, Dougall S, Stinear TP, Druce J, Catton M, Sutton B, van Diemen A, Alpren C, Williamson DA, Howden BP |
| Title | Tracking the COVID-19 Pandemic in Australia Using Genomics |
| Year | 2020 |
| Journal | Nature Communications |
| DOI | 10.1038/s41467-020-18314-x |
| Date received | 2020-07-06 |
| Date accepted | 2020-08-17 |
| **Study setting** | |
| Number of populations included | single |
| Level of populations included | subnational (Victoria (Australia)) |
| Geographic areas covered | Oceania |
| Number of countries covered | single country (Australia) |
| Study period | start and end date span first epidemic wave |
| **Outcome** | |
| Raw outcome | epidemiological individual-level outcome (genome sequence data) |

– continued on next page –

| | |
|---|---|
| Time resolution of raw outcome | daily |
| Computed outcome | epidemiological parameter (reproduction number) |
| Method to obtain the computed outcome | phylodynamic model |
| Data source | data from (sub)national authorities |
| Data availability | data made available by the authors |
| **Intervention** | |
| Terminology for interventions | – not applicable – |
| Terminology for the specific type of non-pharmaceutical interventions | travel restrictions, physical distancing |
| Exposure types | one combination of interventions (estimated change point around March 27 roughly corresponding to the implementation of social distancing restrictions) |
| Types of single interventions | – not applicable – |
| Coding of interventions | – not applicable – |
| Source of intervention data | – not applicable – |
| Availability of data on exposure | – not applicable – |
| **Methodological approach** | |
| Empirical approach | descriptive |
| Use of exposure variation | only variation over time for a single population |
| Method | description of change over time (comparison of time periods) |
| Code availability | none |
| **Effect assessment** | |
| Reporting of intervention effect | comparison of outcome values |
| Measure of effect | reproduction number |
| Reporting of uncertainty | yes |

– continued on next page –

| | |
|---|---|
| Sensitivity analysis | none |
| Subgroup assessment | none |

**Sehra et al. 2020 (Analysis 1)**

| | |
|---|---|
| **Study information** | |
| Author | Sehra ST, George M, Wiebe DJ, Fundin S, Baker JF |
| Title | Cell Phone Activity in Categories of Places and Associations with Growth in Cases of COVID-19 in the US |
| Year | 2020 |
| Journal | JAMA Internal Medicine |
| DOI | 10.1001/jamainternmed.2020.4288 |
| Date received | – could not be evaluated – |
| Date accepted | 2020-08-31 (published) |
| **Study setting** | |
| Number of populations included | multiple |
| Level of populations included | both national and subnational (US counties) |
| Geographic areas covered | North America |
| Number of countries covered | single country (US) |
| Study period | start and end date span first epidemic wave: behavioral |
| **Outcome** | |
| Raw outcome | behavioral outcome (mobility: time spent in various locations) |
| Time resolution of raw outcome | daily |
| Computed outcome | none |

– continued on next page –

| | |
|---|---|
| Method to obtain the computed outcome | – not applicable – |
| Data source | mobility data from corporate organizations (Google) |
| Data availability | data access via source |
| **Intervention** | |
| Terminology for interventions | measures |
| Terminology for the specific type of non-pharmaceutical interventions | social distancing |
| Exposure types | one single intervention (stay-at-home order) |
| Types of single interventions | stay-at-home order |
| Coding of interventions | necessary |
| Source of intervention data | coding done by authors |
| Availability of data on exposure | coded data not available |
| **Methodological approach** | |
| Empirical approach | descriptive |
| Use of exposure variation | only variation over time for multiple populations |
| Method | description of change over time (comparison of time periods) |
| Code availability | none |
| **Effect assessment** | |
| Reporting of intervention effect | quantification of change in outcome values |
| Measure of effect | mobility: time spent in various locations |
| Reporting of uncertainty | yes |
| Sensitivity analysis | none |
| Subgroup assessment | none |

| Sehra et al. 2020 (Analysis 2, mobility as exposure) | |
|---|---|
| **Study information** | |
| Author | Sehra ST, George M, Wiebe DJ, Fundin S, Baker JF |
| Title | Cell Phone Activity in Categories of Places and Associations with Growth in Cases of COVID-19 in the US |
| Year | 2020 |
| Journal | JAMA Internal Medicine |
| DOI | 10.1001/jamainternmed.2020.4288 |
| Date received | – could not be evaluated – |
| Date accepted | 31.08.2020 (published) |
| **Study setting** | |
| Number of populations included | multiple |
| Level of populations included | both national and subnational (US counties) |
| Geographic areas covered | North America |
| Number of countries covered | single country (US) |
| Study period | same end date for several populations with diverse epidemic trajectories |
| **Outcome** | |
| Raw outcome | epidemiological population-level outcome (cases) |
| Time resolution of raw outcome | daily |
| Computed outcome | measure of epidemic trend (growth rate) |
| Method to obtain the computed outcome | simple computation |
| Data source | data from publicly available cross-country selections (JHU) |
| Data availability | data access via source |
| **Intervention** | |
| Terminology for interventions | measures |

– continued on next page –

**Sehra et al. 2020 (Analysis 2, mobility as exposure)** – continued from previous page

| | |
|---|---|
| Terminology for the specific type of non-pharmaceutical interventions | social distancing |
| Exposure types | mobility (time spent in various locations) |
| Types of single interventions | – not applicable – |
| Coding of interventions | – not applicable – |
| Source of intervention data | mobility data by corporate organizations (Google) |
| Availability of data on exposure | data access via source |
| **Methodological approach** | |
| Empirical approach | parametric |
| Use of exposure variation | only variation over time for multiple populations |
| Method | non-mechanistic model (generalized linear model) |
| Code availability | none |
| **Effect assessment** | |
| Reporting of intervention effect | quantification of change in outcome values |
| Measure of effect | growth rate |
| Reporting of uncertainty | yes |
| Sensitivity analysis | model specification varied, epidemiological parameters varied |
| Subgroup assessment | none |

**Sekine et al. 2020**

| | |
|---|---|
| **Study information** | |
| Author | Sekine I, Uojima H, Koyama H, Kamio T, Sato M, Yamamoto T, Fukaguchi K, Fukui H, Yamagami H |

– continued on next page –

| | |
|---|---|
| Title | Impact of Non-Pharmaceutical Interventions for the COVID-19 Pandemic on Emergency Department Patient Trends in Japan: A Retrospective Analysis |
| Year | 2020 |
| Journal | Acute Medicine & Surgery |
| DOI | 10.1002/ams2.603 |
| Date received | 2020-08-11 |
| Date accepted | 2020-10-26 |
| **Study setting** | |
| Number of populations included | single |
| Level of populations included | subnational (Shonan Kamakura General Hospital in Kanagawa, Japan) |
| Geographic areas covered | Asia |
| Number of countries covered | single country (Japan) |
| Study period | start and end date span first epidemic wave |
| **Outcome** | |
| Raw outcome | epidemiological population-level outcome (emergency room visits) |
| Time resolution of raw outcome | daily |
| Computed outcome | none |
| Method to obtain the computed outcome | – not applicable – |
| Data source | data collected by authors (Shonan Kamakura General Hospital in Kanagawa) |
| Data availability | data access via source |
| **Intervention** | |
| Terminology for interventions | interventions |

– continued on next page –

| | |
|---|---|
| Terminology for the specific type of non-pharmaceutical interventions | non-pharmaceutical |
| Exposure types | multiple separate interventions (school closure, emergency declaration) |
| Types of single interventions | school closure, declaration of a state of emergency |
| Coding of interventions | not necessary |
| Source of intervention data | – could not be evaluated – |
| Availability of data on exposure | raw data documented in the manuscript |
| **Methodological approach** | |
| Empirical approach | parametric |
| Use of exposure variation | only variation over time for a single population |
| Method | non-mechanistic model (generalized linear model) |
| Code availability | none |
| **Effect assessment** | |
| Reporting of intervention effect | quantification of change in outcome values |
| Measure of effect | emergency room visits |
| Reporting of uncertainty | yes |
| Sensitivity analysis | none |
| Subgroup assessment | socioeconomic indicators (age, gender) |

**Sewell & And 2020 (Analysis 1)**

| | |
|---|---|
| **Study information** | |
| Author | Sewell DK, And AM |

– continued on next page –

| | |
|---|---|
| Title | Simulation-Free Estimation of an Individual-Based SEIR Model for Evaluating Nonpharmaceutical Interventions with an Application to COVID-19 in the District of Columbia |
| Year | 2020 |
| Journal | PLOS ONE |
| DOI | 10.1371/journal.pone.0241949 |
| Date received | 2020-05-28 |
| Date accepted | 2020-09-26 |
| **Study setting** | |
| Number of populations included | single |
| Level of populations included | subnational (Washingston D.C.) |
| Geographic areas covered | North America |
| Number of countries covered | single country (US) |
| Study period | start and end date span first epidemic wave: behavioral |
| **Outcome** | |
| Raw outcome | behavioral outcome (mobility: time spent in locations for retail and recreation) |
| Time resolution of raw outcome | daily |
| Computed outcome | none |
| Method to obtain the computed outcome | – not applicable – |
| Data source | mobility data from corporate organizations (Google) |
| Data availability | data made available by the authors |
| **Intervention** | |
| Terminology for interventions | interventions |

– continued on next page –

| **Sewell & And 2020 (Analysis 1)** – continued from previous page | |
|---|---|
| Terminology for the specific type of non-pharmaceutical interventions | non-pharmaceutical |
| Exposure types | one single intervention (emergency declaration) |
| Types of single interventions | declaration of a state of emergency |
| Coding of interventions | not necessary |
| Source of intervention data | use of externally coded data (Boston University COVID-19 US State Policy Database) |
| Availability of data on exposure | raw data documented in the manuscript |
| **Methodological approach** | |
| Empirical approach | descriptive |
| Use of exposure variation | only variation over time for a single population |
| Method | description of change over time (description of time course) |
| Code availability | publicly available |
| **Effect assessment** | |
| Reporting of intervention effect | qualitative statement |
| Measure of effect | mobility: time spent in locations for retail and recreation |
| Reporting of uncertainty | – not applicable – |
| Sensitivity analysis | none |
| Subgroup assessment | none |

| **Sewell & And 2020 (Analysis 2)** | |
|---|---|
| **Study information** | |
| Author | Sewell DK, And AM |

– continued on next page –

| | |
|---|---|
| Title | Simulation-Free Estimation of an Individual-Based SEIR Model for Evaluating Nonpharmaceutical Interventions with an Application to COVID-19 in the District of Columbia |
| Year | 2020 |
| Journal | PLOS ONE |
| DOI | 10.1371/journal.pone.0241949 |
| Date received | 2020-05-28 |
| Date accepted | 2020-09-26 |
| **Study setting** | |
| Number of populations included | single |
| Level of populations included | subnational (Washingston D.C.) |
| Geographic areas covered | North America |
| Number of countries covered | single country (US) |
| Study period | start and end date span first epidemic wave |
| **Outcome** | |
| Raw outcome | epidemiological population-level outcome (cases) |
| Time resolution of raw outcome | daily |
| Computed outcome | none |
| Method to obtain the computed outcome | – not applicable – |
| Data source | data from public media outlets (COVID Tracking Project by the Atlantic) |
| Data availability | data made available by the authors |
| **Intervention** | |
| Terminology for interventions | interventions |
| Terminology for the specific type of non-pharmaceutical interventions | non-pharmaceutical |

– continued on next page –

| | |
|---|---|
| Exposure types | one single intervention (mask mandate) |
| Types of single interventions | mask mandate |
| Coding of interventions | not necessary |
| Source of intervention data | use of externally coded data (Boston University COVID-19 US State Policy Database) |
| Availability of data on exposure | raw data documented in the manuscript |
| **Methodological approach** | |
| Empirical approach | parametric |
| Use of exposure variation | only variation over time for a single population |
| Method | mechanistic model (individual-based transmission model) |
| Latent variable functionally linked to intervention | probability of transmission upon contact |
| Code availability | publicly available |
| **Effect assessment** | |
| Reporting of intervention effect | quantification of change in outcome values |
| Measure of effect | probability of transmission upon contact |
| Reporting of uncertainty | no |
| Sensitivity analysis | none |
| Subgroup assessment | none |

**Shanmugam 2020**

| | |
|---|---|
| **Study information** | |
| Author | Shanmugam R |
| Title | Restricted Prevalence Rates of COVID-19's Infectivity, Hospitalization, Recovery, Mortality in the USA and Their Implications |

– continued on next page –

| | |
|---|---|
| Year | 2020 |
| Journal | Journal of Healthcare Informatics Research |
| DOI | 10.1007/s41666-020-00078-0 |
| Date received | 2020-05-28 |
| Date accepted | 2020-09-25 |
| **Study setting** | |
| Number of populations included | multiple |
| Level of populations included | both national and subnational (4 regions in the US: eastern, central, mountain, and pacific time zone states) |
| Geographic areas covered | North America |
| Number of countries covered | single country (US) |
| Study period | end date could not be evaluated |
| **Outcome** | |
| Raw outcome | epidemiological population-level outcome (cases, deaths, hospitalizations, recovered cases) |
| Time resolution of raw outcome | – not applicable – |
| Computed outcome | none |
| Method to obtain the computed outcome | – not applicable – |
| Data source | public (Kaiser Family Foundation) |
| Data availability | data access via source |
| **Intervention** | |
| Terminology for interventions | measures |
| Terminology for the specific type of non-pharmaceutical interventions | social distancing |
| Exposure types | complete set of interventions |

– continued on next page –

| | |
|---|---|
| Types of single interventions | – not applicable – |
| Coding of interventions | – not applicable – |
| Source of intervention data | – not applicable – |
| Availability of data on exposure | – not applicable – |
| **Methodological approach** | |
| Empirical approach | parametric |
| Use of exposure variation | only variation over time for multiple populations |
| Method | non-mechanistic model (generalized linear model) |
| Code availability | none |
| **Effect assessment** | |
| Reporting of intervention effect | quantification of change in outcome values |
| Measure of effect | cases, deaths, hospitalizations, recovered |
| Reporting of uncertainty | no |
| Sensitivity analysis | none |
| Subgroup assessment | geographic areas (regions) |

**Shi et al. 2020a (Analysis 1)**

| | |
|---|---|
| **Study information** | |
| Author | Shi Q, Hu Y, Peng B, Tang XJ, Wang W, Su K, Luo C, Wu B, Zhang F, Zhang Y, Anderson B, Zhong XN, Qiu JF, Yang CY, Huang AL |
| Title | Effective Control of SARS-CoV-2 Transmission in Wanzhou, China |
| Year | 2020 |
| Journal | Nature Medicine |

– continued on next page –

| | |
|---|---|
| DOI | 10.1038/s41591-020-01178-5 |
| Date received | 2020-06-22 |
| Date accepted | 2020-11-13 |
| **Study setting** | |
| Number of populations included | single |
| Level of populations included | subnational (Wanzhou (China)) |
| Geographic areas covered | Asia |
| Number of countries covered | single country (China) |
| Study period | start and end date span first epidemic wave |
| **Outcome** | |
| Raw outcome | epidemiological individual-level outcome (individual cases and transmission chains) |
| Time resolution of raw outcome | daily |
| Computed outcome | epidemiological parameter (reproduction number) |
| Method to obtain the computed outcome | statistical estimation of reproduction number from individual data (Lloyd and Smith, between generations) |
| Data source | data from (sub)national authorities |
| Data availability | data not accessible |
| **Intervention** | |
| Terminology for interventions | measures |
| Terminology for the specific type of non-pharmaceutical interventions | control |
| Exposure types | one combination of interventions (Wuhan City Lockdown) |
| Types of single interventions | – not applicable – |
| Coding of interventions | not necessary |
| Source of intervention data | – could not be evaluated – |

– continued on next page –

| | |
|---|---|
| Availability of data on exposure | raw data documented in the manuscript |
| **Methodological approach** | |
| Empirical approach | descriptive |
| Use of exposure variation | only variation over time for a single population |
| Method | description of change over time (comparison of time periods) |
| Code availability | publicly available |
| **Effect assessment** | |
| Reporting of intervention effect | comparison of outcome values |
| Measure of effect | reproduction number |
| Reporting of uncertainty | yes |
| Sensitivity analysis | none |
| Subgroup assessment | none |

**Shi et al. 2020a (Analysis 2)**

| | |
|---|---|
| **Study information** | |
| Author | Shi Q, Hu Y, Peng B, Tang XJ, Wang W, Su K, Luo C, Wu B, Zhang F, Zhang Y, Anderson B, Zhong XN, Qiu JF, Yang CY, Huang AL |
| Title | Effective Control of SARS-CoV-2 Transmission in Wanzhou, China |
| Year | 2020 |
| Journal | Nature Medicine |
| DOI | 10.1038/s41591-020-01178-5 |
| Date received | 2020-06-22 |
| Date accepted | 2020-11-13 |
| **Study setting** | |

– continued on next page –

| | |
|---|---|
| Number of populations included | single |
| Level of populations included | subnational (Wanzhou (China)) |
| Geographic areas covered | Asia |
| Number of countries covered | single country (China) |
| Study period | start and end date span first epidemic wave |
| **Outcome** | |
| Raw outcome | epidemiological population-level outcome (cases) |
| Time resolution of raw outcome | daily |
| Computed outcome | none |
| Method to obtain the computed outcome | – not applicable – |
| Data source | data from (sub)national authorities |
| Data availability | data access via source |
| **Intervention** | |
| Terminology for interventions | measures |
| Terminology for the specific type of non-pharmaceutical interventions | control |
| Exposure types | one combination of interventions (Wuhan City Lockdown) |
| Types of single interventions | – not applicable – |
| Coding of interventions | not necessary |
| Source of intervention data | – could not be evaluated – |
| Availability of data on exposure | raw data documented in the manuscript |
| **Methodological approach** | |
| Empirical approach | counterfactual |
| Use of exposure variation | only variation over time for a single population |
| Method | mechanistic model (individual-based transmission model) |

– continued on next page –

| | |
|---|---|
| Code availability | publicly available |
| **Effect assessment** | |
| Reporting of intervention effect | comparison of outcome values |
| Measure of effect | cases |
| Reporting of uncertainty | no |
| Sensitivity analysis | none |
| Subgroup assessment | none |

**Shi et al. 2020b**

| | |
|---|---|
| **Study information** | |
| Author | Shi S, Tanaka S, Ueno R, Gilmour S, Tanoue Y, Kawashima T, Nomura S, Eguchi A, Miyata H, Yoneoka D |
| Title | Travel Restrictions and SARS-CoV-2 Transmission: An Effective Distance Approach to Estimate Impact |
| Year | 2020 |
| Journal | Bulletin of the World Health Organization |
| DOI | 10.2471/blt.20.255679 |
| Date received | 2020-03-18 |
| Date accepted | 2020-05-12 |
| **Study setting** | |
| Number of populations included | multiple |
| Level of populations included | both national and subnational (80 countries around the globe with their airports) |
| Geographic areas covered | Europe, Middle East and Africa, Asia, Oceania, Central and South America, North America |

– continued on next page –

| | |
|---|---|
| Number of countries covered | multiple countries |
| Study period | same end date for several populations with diverse epidemic trajectories |
| **Outcome** | |
| Raw outcome | epidemiological population-level outcome (dates of first reported case in country), behavioral outcome (mobility: transportation network data) |
| Time resolution of raw outcome | daily |
| Computed outcome | none |
| Method to obtain the computed outcome | – not applicable – |
| Data source | public (Automatic Dependent Surveillance-Broadcast exchange data: mobility), – could not be evaluated – (first case date) |
| Data availability | data access via source |
| **Intervention** | |
| Terminology for interventions | – not applicable – |
| Terminology for the specific type of non-pharmaceutical interventions | travel restrictions |
| Exposure types | one single intervention (travel restrictions (day of introduction of travel restrictions: entry bans, visa restrictions, flight suspensions)) |
| Types of single interventions | international travel restrictions |
| Coding of interventions | necessary |
| Source of intervention data | use of externally coded data (WHO, Think Global Health) |
| Availability of data on exposure | coded data made available by the authors |
| **Methodological approach** | |
| Empirical approach | counterfactual |
| Use of exposure variation | both variation over time and between populations |
| Method | other (hazard-based survival model for countries' risk of case importation) |

– continued on next page –

| | |
|---|---|
| Code availability | publicly available |
| **Effect assessment** | |
| Reporting of intervention effect | quantification of change in outcome values |
| Measure of effect | countries' risk of case importation |
| Reporting of uncertainty | no |
| Sensitivity analysis | none |
| Subgroup assessment | none |

**Shim et al. 2021**

| | |
|---|---|
| **Study information** | |
| Author | Shim E, Tariq A, Chowell G |
| Title | Spatial Variability in Reproduction Number and Doubling Time across Two Waves of the COVID-19 Pandemic in South Korea, February to July, 2020 |
| Year | 2021 |
| Journal | International Journal of Infectious Diseases |
| DOI | 10.1016/j.ijid.2020.10.007 |
| Date received | 2020-08-30 |
| Date accepted | 2020-10-03 |
| **Study setting** | |
| Number of populations included | multiple |
| Level of populations included | subnational (Seoul, Gyeonggi Province, Gyeongbuk Province, and Daegu (South Korea)) |
| Geographic areas covered | Asia |
| Number of countries covered | single country (South Korea) |

– continued on next page –

| | |
|---|---|
| Study period | includes second wave (beginning of second wave for some regions) |
| **Outcome** | |
| Raw outcome | epidemiological population-level outcome (cases) |
| Time resolution of raw outcome | daily |
| Computed outcome | measure of epidemic trend (doubling time), epidemiological parameter (reproduction number) |
| Method to obtain the computed outcome | statistical estimation of reproduction number, simple computation (doubling time) |
| Data source | data from (sub)national authorities |
| Data availability | data access via source |
| **Intervention** | |
| Terminology for interventions | measures |
| Terminology for the specific type of non-pharmaceutical interventions | control |
| Exposure types | all interventions together |
| Types of single interventions | – not applicable – |
| Coding of interventions | – not applicable – |
| Source of intervention data | – not applicable – |
| Availability of data on exposure | – not applicable – |
| **Methodological approach** | |
| Empirical approach | descriptive |
| Use of exposure variation | only variation over time for multiple populations |
| Method | description of change over time (description of time course) |
| Code availability | none |
| **Effect assessment** | |
| Reporting of intervention effect | qualitative statement |

– continued on next page –

| | |
|---|---|
| Measure of effect | reproduction number, doubling time |
| Reporting of uncertainty | – not applicable – |
| Sensitivity analysis | none |
| Subgroup assessment | none |

**Siedner et al. 2020**

| | |
|---|---|
| **Study information** | |
| Author | Siedner MJ, Harling G, Reynolds Z, Gilbert RF, Haneuse S, Venkataramani AS, Tsai AC |
| Title | Social Distancing to Slow the US COVID-19 Epidemic: Longitudinal Pretest–Posttest Comparison Group Study |
| Year | 2020 |
| Journal | PLOS Medicine |
| DOI | 10.1371/journal.pmed.1003244 |
| Date received | 2020-04-27 |
| Date accepted | 2020-07-02 |
| **Study setting** | |
| Number of populations included | multiple |
| Level of populations included | both national and subnational (US states) |
| Geographic areas covered | North America |
| Number of countries covered | single country (US) |
| Study period | same end date for several populations with diverse epidemic trajectories |
| **Outcome** | |
| Raw outcome | epidemiological population-level outcome (cases, deaths) |

– continued on next page –

| | |
|---|---|
| Time resolution of raw outcome | daily |
| Computed outcome | measure of epidemic trend (growth rate, growth rate: deaths) |
| Method to obtain the computed outcome | simple computation |
| Data source | data from public media outlets (New York Times) |
| Data availability | data access via source |
| **Intervention** | |
| Terminology for interventions | measures |
| Terminology for the specific type of non-pharmaceutical interventions | social distancing |
| Exposure types | start of interventions (days since first social distancing measure (closure of schools, closure of workplaces, cancellations of public events, stay-at-home order, closure of state borders)) |
| Types of single interventions | – not applicable – |
| Coding of interventions | necessary |
| Source of intervention data | coding done by authors |
| Availability of data on exposure | coded data made available by the authors |
| **Methodological approach** | |
| Empirical approach | parametric |
| Use of exposure variation | both variation over time and between populations |
| Method | non-mechanistic model (generalized linear model) |
| Code availability | none |
| **Effect assessment** | |
| Reporting of intervention effect | quantification of change in outcome values |
| Measure of effect | growth rate, growth rate: deaths |
| Reporting of uncertainty | yes |

– continued on next page –

| | |
|---|---|
| Sensitivity analysis | model specification varied, epidemiological parameters varied |
| Subgroup assessment | none |

| **Signorelli et al. 2020** | |
|---|---|
| **Study information** | |
| Author | Signorelli C, Scognamiglio T, Odone A |
| Title | COVID-19 in Italy: Impact of Containment Measures and Prevalence Estimates of Infection in the General Population |
| Year | 2020 |
| Journal | Acta Bio Medica Atenei Parmensis |
| DOI | 10.23750/abm.v91i3-S.9511 |
| Date received | 2020-04-07 |
| Date accepted | 2020-04-08 |
| **Study setting** | |
| Number of populations included | multiple |
| Level of populations included | both national and subnational (Italy and regions) |
| Geographic areas covered | Europe |
| Number of countries covered | single country (Italy) |
| Study period | start and end date span first epidemic wave |
| **Outcome** | |
| Raw outcome | epidemiological population-level outcome (cases) |
| Time resolution of raw outcome | daily |
| Computed outcome | none |

– continued on next page –

| | |
|---|---|
| Method to obtain the computed outcome | – not applicable – |
| Data source | data from (sub)national authorities |
| Data availability | data access via source |
| **Intervention** | |
| Terminology for interventions | measures |
| Terminology for the specific type of non-pharmaceutical interventions | containment |
| Exposure types | all interventions together (timeline of measures) |
| Types of single interventions | – not applicable – |
| Coding of interventions | – not applicable – |
| Source of intervention data | – not applicable – |
| Availability of data on exposure | – not applicable – |
| **Methodological approach** | |
| Empirical approach | descriptive |
| Use of exposure variation | only variation over time for multiple populations |
| Method | description of change over time (description of time course) |
| Code availability | none |
| **Effect assessment** | |
| Reporting of intervention effect | qualitative statement |
| Measure of effect | cases |
| Reporting of uncertainty | – not applicable – |
| Sensitivity analysis | none |
| Subgroup assessment | none |

| Silva et al. 2020 | |
|---|---|
| **Study information** | |
| Author | Silva L, Filho DF, Fernandes A |
| Title | The Effect of Lockdown on the COVID-19 Epidemic in Brazil: Evidence from an Interrupted Time Series Design |
| Year | 2020 |
| Journal | Cadernos de Saúde Pública |
| DOI | 10.1590/0102-311x00213920 |
| Date received | 2020-07-21 |
| Date accepted | 2020-08-31 |
| **Study setting** | |
| Number of populations included | multiple |
| Level of populations included | subnational (4 cities (Brazil)) |
| Geographic areas covered | Central and South America |
| Number of countries covered | single country (Brazil) |
| Study period | end date in growth phase of wave |
| **Outcome** | |
| Raw outcome | epidemiological population-level outcome (cases, deaths) |
| Time resolution of raw outcome | daily |
| Computed outcome | none |
| Method to obtain the computed outcome | – not applicable – |
| Data source | data from (sub)national authorities |
| Data availability | data made available by the authors |
| **Intervention** | |
| Terminology for interventions | – not applicable – |

– continued on next page –

| | |
|---|---|
| Terminology for the specific type of non-pharmaceutical interventions | lockdown |
| Exposure types | one combination of interventions (lockdown) |
| Types of single interventions | – not applicable – |
| Coding of interventions | not necessary |
| Source of intervention data | – could not be evaluated – |
| Availability of data on exposure | raw data documented in the manuscript |
| **Methodological approach** | |
| Empirical approach | parametric |
| Use of exposure variation | only variation over time for multiple populations |
| Method | non-mechanistic model (generalized linear model) |
| Code availability | publicly available |
| **Effect assessment** | |
| Reporting of intervention effect | quantification of change in outcome values |
| Measure of effect | cases, deaths |
| Reporting of uncertainty | yes |
| Sensitivity analysis | none |
| Subgroup assessment | none |

**Singh & Singh 2020**

| | |
|---|---|
| **Study information** | |
| Author | Singh BP, Singh G |
| Title | Modeling Tempo ofCOVID-19 Pandemic in India and Significance of Lockdown |

– continued on next page –

| | |
|---|---|
| Year | 2020 |
| Journal | Journal of Public Affairs |
| DOI | 10.1002/pa.2257 |
| Date received | 2020-06-24 |
| Date accepted | 2020-06-27 |
| **Study setting** | |
| Number of populations included | multiple |
| Level of populations included | both national and subnational (India and its states) |
| Geographic areas covered | Asia |
| Number of countries covered | single country (India) |
| Study period | end date in growth phase of wave |
| **Outcome** | |
| Raw outcome | epidemiological population-level outcome (cases) |
| Time resolution of raw outcome | daily |
| Computed outcome | measure of epidemic trend (growth rate) |
| Method to obtain the computed outcome | exponential growth model |
| Data source | data from publicly available open-source projects (covid19india.org) |
| Data availability | data access via source |
| **Intervention** | |
| Terminology for interventions | strategies |
| Terminology for the specific type of non-pharmaceutical interventions | physical distancing |
| Exposure types | one combination of interventions (Indian National Lockdown) |
| Types of single interventions | – not applicable – |
| Coding of interventions | not necessary |

– continued on next page –

| | |
|---|---|
| Source of intervention data | – could not be evaluated – |
| Availability of data on exposure | raw data documented in the manuscript |
| **Methodological approach** | |
| Empirical approach | descriptive |
| Use of exposure variation | only variation over time for a single population |
| Method | description of change over time (comparison of time periods) |
| Code availability | none |
| **Effect assessment** | |
| Reporting of intervention effect | quantification of change in outcome values |
| Measure of effect | growth rate |
| Reporting of uncertainty | yes |
| Sensitivity analysis | none |
| Subgroup assessment | none |

**Singh et al. 2020**

| | |
|---|---|
| **Study information** | |
| Author | Singh BB, Lowerison M, Lewinson RT, Vallerand IA, Deardon R, Gill JP, Singh B, Barkema HW |
| Title | Public Health Interventions Slowed but Did Not Halt the Spread of COVID-19 in India |
| Year | 2020 |
| Journal | Transboundary and Emerging Diseases |
| DOI | 10.1111/tbed.13868 |
| Date received | 2020-06-20 |

– continued on next page –

| | |
|---|---|
| Date accepted | 2020-09-26 |
| **Study setting** | |
| Number of populations included | multiple |
| Level of populations included | both national and subnational (India and 12 states) |
| Geographic areas covered | Asia |
| Number of countries covered | single country (India) |
| Study period | end date in growth phase of wave |
| **Outcome** | |
| Raw outcome | epidemiological population-level outcome (cases), behavioral outcome (mobility: time spent in various locations) |
| Time resolution of raw outcome | daily |
| Computed outcome | epidemiological parameter (reproduction number) |
| Method to obtain the computed outcome | statistical estimation of reproduction number (Cori et al.) |
| Data source | data from publicly available cross-country selections (WHO: cases), mobility data from corporate organizations (Google: mobility) |
| Data availability | data access via source |
| **Intervention** | |
| Terminology for interventions | interventions |
| Terminology for the specific type of non-pharmaceutical interventions | public health |
| Exposure types | one combination of interventions (Indian National Lockdown) |
| Types of single interventions | – not applicable – |
| Coding of interventions | not necessary |
| Source of intervention data | government or news websites |
| Availability of data on exposure | raw data documented in the manuscript |

– continued on next page –

| | |
|---|---|
| **Methodological approach** | |
| Empirical approach | descriptive |
| Use of exposure variation | only variation over time for multiple populations |
| Method | description of change over time (comparison of time periods) |
| Code availability | none |
| **Effect assessment** | |
| Reporting of intervention effect | comparison of outcome values |
| Measure of effect | cases, reproduction number, mobility: time spent in various locations |
| Reporting of uncertainty | yes |
| Sensitivity analysis | none |
| Subgroup assessment | none |

**Sinha 2020**

| | |
|---|---|
| **Study information** | |
| Author | Sinha S |
| Title | Epidemiological Dynamics of the COVID-19 Pandemic in India: An Interim Assessment |
| Year | 2020 |
| Journal | Statistics and Applications |
| DOI | – not applicable – |
| Date received | 2020-07-20 |
| Date accepted | 2020-07-30 |
| **Study setting** | |
| Number of populations included | single |

– continued on next page –

| | |
|---|---|
| Level of populations included | national (India) |
| Geographic areas covered | Asia |
| Number of countries covered | single country (India) |
| Study period | end date in growth phase of wave |
| **Outcome** | |
| Raw outcome | epidemiological population-level outcome (cases, deaths, recovered cases) |
| Time resolution of raw outcome | daily |
| Computed outcome | epidemiological parameter (reproduction number) |
| Method to obtain the computed outcome | exponential growth model |
| Data source | data from publicly available cross-country selections (JHU) |
| Data availability | data access via source |
| **Intervention** | |
| Terminology for interventions | interventions |
| Terminology for the specific type of non-pharmaceutical interventions | non-pharmaceutical |
| Exposure types | one combination of interventions (Indian National Lockdown) |
| Types of single interventions | – not applicable – |
| Coding of interventions | not necessary |
| Source of intervention data | – could not be evaluated – |
| Availability of data on exposure | raw data documented in the manuscript |
| **Methodological approach** | |
| Empirical approach | descriptive |
| Use of exposure variation | only variation over time for a single population |
| Method | description of change over time (comparison of time periods) |

– continued on next page –

| | |
|---|---|
| Code availability | none |
| **Effect assessment** | |
| Reporting of intervention effect | comparison of outcome values |
| Measure of effect | reproduction number |
| Reporting of uncertainty | yes |
| Sensitivity analysis | none |
| Subgroup assessment | none |

**Spaccaferri et al. 2020**

| | |
|---|---|
| **Study information** | |
| Author | Spaccaferri G, Larrieu S, Pouey J, Calba C, Benet T, Sommen C, Lévy-Bruhl D, Smaili S, Che D, Filleul L, Caserio-Schönemann C, Ait-El-Belghiti F, Haeghebaert S, Desenclos JC, Huiart L, Laporte A, Rolland P |
| Title | Early Assessment of the Impact of Mitigation Measures to Control COVID-19 in 22 French Metropolitan Areas, October to November 2020 |
| Year | 2020 |
| Journal | Eurosurveillance |
| DOI | 10.2807/1560-7917.es.2020.25.50.2001974 |
| Date received | 2020-11-21 |
| Date accepted | 2020-12-17 |
| **Study setting** | |
| Number of populations included | multiple |
| Level of populations included | both national and subnational (22 French metropolitan areas) |
| Geographic areas covered | Europe |
| Number of countries covered | single country (France) |

– continued on next page –

| | |
|---|---|
| Study period | includes second wave (focus on second wave: start date October 28, 2020, end date November 15, 2020) |
| **Outcome** | |
| Raw outcome | epidemiological population-level outcome (cases, hospitalizations, test positivity rate) |
| Time resolution of raw outcome | daily |
| Computed outcome | measure of epidemic trend (growth rate, growth rate: hospitalizations, growth rate: test positivity rate) |
| Method to obtain the computed outcome | simple computation |
| Data source | data from (sub)national authorities |
| Data availability | data access via source |
| **Intervention** | |
| Terminology for interventions | measures |
| Terminology for the specific type of non-pharmaceutical interventions | mitigation |
| Exposure types | one combination of interventions (Wuhan City Lockdown) |
| Types of single interventions | – not applicable – |
| Coding of interventions | not necessary |
| Source of intervention data | – could not be evaluated – |
| Availability of data on exposure | raw data documented in the manuscript |
| **Methodological approach** | |
| Empirical approach | descriptive |
| Use of exposure variation | only variation over time for multiple populations |
| Method | description of change over time (comparison of time periods) |
| Code availability | none |

– continued on next page –

| | |
|---|---|
| **Effect assessment** | |
| Reporting of intervention effect | comparison of outcome values |
| Measure of effect | growth rate, growth rate: hospitalizations, growth rate: test positivity rate |
| Reporting of uncertainty | no |
| Sensitivity analysis | none |
| Subgroup assessment | public health response (response time) |

**Sun et al. 2020**

| | |
|---|---|
| **Study information** | |
| Author | Sun S, Folarin AA, Ranjan Y, Rashid Z, Conde P, Stewart C, Cummins N, Matcham F, Dalla Costa G, Simblett S, Leocani L, Lamers F, Sørensen PS, Buron M, Zabalza A, Guerrero Pérez AI, Penninx BW, Siddi S, Haro JM, Myin-Germeys I, Rintala A, Wykes T, Narayan VA, Comi G, Hotopf M, Dobson RJ, RADAR-CNS Consortium |
| Title | Using Smartphones and Wearable Devices to Monitor Behavioral Changes During COVID-19 |
| Year | 2020 |
| Journal | Journal of Medical Internet Research |
| DOI | 10.2196/19992 |
| Date received | 2020-05-08 |
| Date accepted | 2020-07-26 |
| **Study setting** | |
| Number of populations included | multiple |
| Level of populations included | national (Italy, Spain, Denmark, UK, Netherlands) |
| Geographic areas covered | Europe |

– continued on next page –

| | |
|---|---|
| Number of countries covered | multiple countries |
| Study period | start and end date span first epidemic wave: behavioral |
| **Outcome** | |
| Raw outcome | behavioral outcome (mobility: time spent at home, maximum distance from place of residence, steps, maximum number of nearby devices) |
| Time resolution of raw outcome | daily |
| Computed outcome | none |
| Method to obtain the computed outcome | – not applicable – |
| Data source | data collected by authors (RADAR CNS study using wearable devices and smartphones) |
| Data availability | data not accessible |
| **Intervention** | |
| Terminology for interventions | interventions |
| Terminology for the specific type of non-pharmaceutical interventions | non-pharmaceutical |
| Exposure types | start of interventions (date of first restrictive measure introducing national lockdown) |
| Types of single interventions | – not applicable – |
| Coding of interventions | not necessary |
| Source of intervention data | use of externally coded data (OxCGRT) |
| Availability of data on exposure | access to externally coded data via source |
| **Methodological approach** | |
| Empirical approach | descriptive |
| Use of exposure variation | only variation over time for multiple populations |
| Method | description of change over time (comparison of time periods) |

– continued on next page –

| | |
|---|---|
| Code availability | none |
| **Effect assessment** | |
| Reporting of intervention effect | quantification of change in outcome values |
| Measure of effect | mobility: time spent at home & maximum distance from place of residence & steps & maximum number of nearby devices |
| Reporting of uncertainty | yes |
| Sensitivity analysis | none |
| Subgroup assessment | none |

**Supino et al. 2020**

| | |
|---|---|
| **Study information** | |
| Author | Supino M, d'Onofrio A, Luongo F, Occhipinti G, Co AD |
| Title | The Effects of Containment Measures in the Italian Outbreak of COVID-19 |
| Year | 2020 |
| Journal | BMC Public Health |
| DOI | 10.1186/s12889-020-09913-w |
| Date received | 2020-04-03 |
| Date accepted | 2020-11-18 |
| **Study setting** | |
| Number of populations included | multiple |
| Level of populations included | both national and subnational (Italy (excluding regions with saturated ICU capacity), Lombardy) |
| Geographic areas covered | Europe |
| Number of countries covered | single country (Italy) |

– continued on next page –

| | |
|---|---|
| Study period | start and end date span first epidemic wave |
| **Outcome** | |
| Raw outcome | epidemiological population-level outcome (ICU hospitalizations, deaths) |
| Time resolution of raw outcome | daily |
| Computed outcome | none |
| Method to obtain the computed outcome | – not applicable – |
| Data source | data from (sub)national authorities |
| Data availability | data access via source |
| **Intervention** | |
| Terminology for interventions | measures |
| Terminology for the specific type of non-pharmaceutical interventions | containment |
| Exposure types | multiple combinations of interventions (lockdown on March 9, full lockdown on March 21 involving work ban) |
| Types of single interventions | – not applicable – |
| Coding of interventions | not necessary |
| Source of intervention data | – could not be evaluated – |
| Availability of data on exposure | raw data documented in the manuscript |
| **Methodological approach** | |
| Empirical approach | counterfactual |
| Use of exposure variation | only variation over time for a single population |
| Method | non-mechanistic model (exponential growth model) |
| Code availability | none |
| **Effect assessment** | |
| Reporting of intervention effect | qualitative statement |

– continued on next page –

| | |
|---|---|
| Measure of effect | ICU hospitalizations, deaths |
| Reporting of uncertainty | – not applicable – |
| Sensitivity analysis | none |
| Subgroup assessment | none |

| **Suraya et al. 2020 (Analysis 1)** | |
|---|---|
| **Study information** | |
| Author | Suraya I, Nurmansyah MI, Rachmawati E, Aufa BA, Koire II |
| Title | The Impact of Large-Scale Social Restriction on COVID-19 Incidence: A Case Study of Four Regions in Indonesia |
| Year | 2020 |
| Journal | Kesmas: National Public Health Journal |
| DOI | 10.21109/kesmas.v15i2.3990 |
| Date received | 2020-06-19 |
| Date accepted | 2020-06-20 |
| **Study setting** | |
| Number of populations included | multiple |
| Level of populations included | subnational (4 provinces in Indonesia) |
| Geographic areas covered | Asia |
| Number of countries covered | single country (Indonesia) |
| Study period | end date in growth phase of wave |
| **Outcome** | |
| Raw outcome | epidemiological population-level outcome (cases) |

– continued on next page –

| | |
|---|---|
| Time resolution of raw outcome | daily |
| Computed outcome | none |
| Method to obtain the computed outcome | – not applicable – |
| Data source | – could not be evaluated – |
| Data availability | data not accessible |
| **Intervention** | |
| Terminology for interventions | – not applicable – |
| Terminology for the specific type of non-pharmaceutical interventions | social restrictions |
| Exposure types | one combination of interventions (large scale social restrictions (school closure, non-essential business closures, restrictions of religious activities, limitations on activities in public places or facilities, limitations on mode of transportations, restrictions on activities related to defense and security)) |
| Types of single interventions | – not applicable – |
| Coding of interventions | not necessary |
| Source of intervention data | – could not be evaluated – |
| Availability of data on exposure | raw data documented in the manuscript |
| **Methodological approach** | |
| Empirical approach | descriptive |
| Use of exposure variation | only variation over time for multiple populations |
| Method | description of change over time (comparison of time periods) |
| Code availability | none |
| **Effect assessment** | |
| Reporting of intervention effect | comparison of outcome values |
| Measure of effect | cases |

– continued on next page –

| | |
|---|---|
| Reporting of uncertainty | yes |
| Sensitivity analysis | none |
| Subgroup assessment | none |

**Suraya et al. 2020 (Analysis 2)**

| | |
|---|---|
| **Study information** | |
| Author | Suraya I, Nurmansyah MI, Rachmawati E, Aufa BA, Koire II |
| Title | The Impact of Large-Scale Social Restriction on COVID-19 Incidence: A Case Study of Four Regions in Indonesia |
| Year | 2020 |
| Journal | Kesmas: National Public Health Journal |
| DOI | 10.21109/kesmas.v15i2.3990 |
| Date received | 2020-06-19 |
| Date accepted | 2020-06-20 |
| **Study setting** | |
| Number of populations included | multiple |
| Level of populations included | subnational (4 provinces in Indonesia) |
| Geographic areas covered | Asia |
| Number of countries covered | single country (Indonesia) |
| Study period | end date in growth phase of wave |
| **Outcome** | |
| Raw outcome | epidemiological population-level outcome (cases) |
| Time resolution of raw outcome | daily |
| Computed outcome | none |

– continued on next page –

| | |
|---|---|
| Method to obtain the computed outcome | – not applicable – |
| Data source | – could not be evaluated – |
| Data availability | data not accessible |
| **Intervention** | |
| Terminology for interventions | – not applicable – |
| Terminology for the specific type of non-pharmaceutical interventions | social restrictions |
| Exposure types | one combination of interventions (large scale social restrictions (school closure, non-essential business closures, restrictions of religious activities, limitations on activities in public places or facilities, limitations on mode of transportations, restrictions on activities related to defense and security)) |
| Types of single interventions | – not applicable – |
| Coding of interventions | not necessary |
| Source of intervention data | – could not be evaluated – |
| Availability of data on exposure | raw data documented in the manuscript |
| **Methodological approach** | |
| Empirical approach | parametric |
| Use of exposure variation | only variation over time for multiple populations |
| Method | non-mechanistic model (generalized linear model) |
| Code availability | none |
| **Effect assessment** | |
| Reporting of intervention effect | quantification of change in outcome values |
| Measure of effect | incidence rate ratio |
| Reporting of uncertainty | yes |
| Sensitivity analysis | none |

– continued on next page –

| | |
|---|---|
| Subgroup assessment | none |

| **Talmoudi et al. 2020** | |
|---|---|
| **Study information** | |
| Author | Talmoudi K, Safer M, Letaief H, Hchaichi A, Harizi C, Dhaouadi S, Derouiche S, Bouaziz I, Gharbi D, Najar N, Osman M, Cherif I, Mlallekh R, Ben-Ayed O, Ayedi Y, Bouabid L, Bougatef S, ép Ben-Alaya NB, Chahed MK |
| Title | Estimating Transmission Dynamics and Serial Interval of the First Wave of COVID-19 Infections under Different Control Measures: A Statistical Analysis in Tunisia from February 29 to May 5, 2020 |
| Year | 2020 |
| Journal | BMC Infectious Diseases |
| DOI | 10.1186/s12879-020-05577-4 |
| Date received | 2020-05-23 |
| Date accepted | 2020-11-03 |
| **Study setting** | |
| Number of populations included | single |
| Level of populations included | national (Tunisia) |
| Geographic areas covered | Middle East and Africa |
| Number of countries covered | single country (Tunisia) |
| Study period | start and end date span first epidemic wave |
| **Outcome** | |
| Raw outcome | epidemiological individual-level outcome (cases, individual cases) |
| Time resolution of raw outcome | daily |

– continued on next page –

| | |
|---|---|
| Computed outcome | epidemiological parameter (reproduction number) |
| Method to obtain the computed outcome | statistical estimation of reproduction number (Cori et al.) |
| Data source | data from (sub)national authorities (National Notifiable Diseases Reporting System: influenza, National Sentinel Influenza Surveillance Network: influenza-like illnesses) |
| Data availability | data access via source |
| **Intervention** | |
| Terminology for interventions | measures |
| Terminology for the specific type of non-pharmaceutical interventions | control |
| Exposure types | multiple combinations of interventions (curfew, lockdown) |
| Types of single interventions | – not applicable – |
| Coding of interventions | not necessary |
| Source of intervention data | – could not be evaluated – |
| Availability of data on exposure | raw data documented in the manuscript |
| **Methodological approach** | |
| Empirical approach | descriptive |
| Use of exposure variation | only variation over time for a single population |
| Method | description of change over time (comparison of time periods) |
| Code availability | none |
| **Effect assessment** | |
| Reporting of intervention effect | comparison of outcome values |
| Measure of effect | reproduction number |
| Reporting of uncertainty | yes |
| Sensitivity analysis | none |

– continued on next page –

| | |
|---|---|
| Subgroup assessment | none |

| **Tamagusko & Ferreira 2020 (Analysis 1)** | |
|---|---|
| **Study information** | |
| Author | Tamagusko T, Ferreira A |
| Title | Data-Driven Approach to Understand the Mobility Patterns of the Portuguese Population during the COVID-19 Pandemic |
| Year | 2020 |
| Journal | Sustainability |
| DOI | 10.3390/su12229775 |
| Date received | 2020-10-14 |
| Date accepted | 2020-11-20 |
| **Study setting** | |
| Number of populations included | single |
| Level of populations included | national (Portugal) |
| Geographic areas covered | Europe |
| Number of countries covered | single country (Portugal) |
| Study period | start and end date span first epidemic wave: behavioral |
| **Outcome** | |
| Raw outcome | behavioral outcome (mobility: time spent in various locations) |
| Time resolution of raw outcome | daily |
| Computed outcome | change points (change points: mobility) |

– continued on next page –

| | |
|---|---|
| Method to obtain the computed outcome | change point analysis |
| Data source | mobility data from corporate organizations (Google) |
| Data availability | data made available by the authors |
| **Intervention** | |
| Terminology for interventions | measures, policies |
| Terminology for the specific type of non-pharmaceutical interventions | social distancing |
| Exposure types | all interventions together (timeline of measures) |
| Types of single interventions | – not applicable – |
| Coding of interventions | – not applicable – |
| Source of intervention data | – not applicable – |
| Availability of data on exposure | – not applicable – |
| **Methodological approach** | |
| Empirical approach | descriptive |
| Use of exposure variation | only variation over time for a single population |
| Method | comparison of change points with intervention dates |
| Code availability | publicly available |
| **Effect assessment** | |
| Reporting of intervention effect | comparison of outcome values |
| Measure of effect | change points: mobility |
| Reporting of uncertainty | no |
| Sensitivity analysis | none |
| Subgroup assessment | none |

| Tamagusko & Ferreira 2020 (Analysis 2) | |
|---|---|
| **Study information** | |
| Author | Tamagusko T, Ferreira A |
| Title | Data-Driven Approach to Understand the Mobility Patterns of the Portuguese Population during the COVID-19 Pandemic |
| Year | 2020 |
| Journal | Sustainability |
| DOI | 10.3390/su12229775 |
| Date received | 2020-10-14 |
| Date accepted | 2020-11-20 |
| **Study setting** | |
| Number of populations included | single |
| Level of populations included | national (Portugal) |
| Geographic areas covered | Europe |
| Number of countries covered | single country (Portugal) |
| Study period | start and end date span first epidemic wave |
| **Outcome** | |
| Raw outcome | epidemiological population-level outcome (cases) |
| Time resolution of raw outcome | daily |
| Computed outcome | epidemiological parameter (reproduction number) |
| Method to obtain the computed outcome | statistical estimation of reproduction number (Cori et al.) |
| Data source | data from publicly available cross-country selections (OurWorldInData) |
| Data availability | data made available by the authors |
| **Intervention** | |
| Terminology for interventions | measures, policies |

– continued on next page –

| | |
|---|---|
| Terminology for the specific type of non-pharmaceutical interventions | social distancing |
| Exposure types | all interventions together (timeline of measures) |
| Types of single interventions | – not applicable – |
| Coding of interventions | – not applicable – |
| Source of intervention data | – not applicable – |
| Availability of data on exposure | – not applicable – |
| **Methodological approach** | |
| Empirical approach | descriptive |
| Use of exposure variation | only variation over time for a single population |
| Method | description of change over time (description of time course) |
| Code availability | publicly available |
| **Effect assessment** | |
| Reporting of intervention effect | qualitative statement |
| Measure of effect | reproduction number |
| Reporting of uncertainty | – not applicable – |
| Sensitivity analysis | none |
| Subgroup assessment | none |

**Tchole et al. 2020**

| | |
|---|---|
| **Study information** | |
| Author | Tchole AI, Li ZW, Wei JT, Ye RZ, Wang WJ, Du WY, Wang HT, Yin CN, Ji XK, Xue FZ, Bachir AM, Zhao L, Cao WC |
| Title | Epidemic and Control of COVID-19 in Niger: Quantitative Analyses in a Least Developed Country |

– continued on next page –

| | |
|---|---|
| Year | 2020 |
| Journal | Journal of Global Health |
| DOI | 10.7189/jogh.10.020513 |
| Date received | – could not be evaluated – |
| Date accepted | 2020-12-06 |
| **Study setting** | |
| Number of populations included | single |
| Level of populations included | national (Niger) |
| Geographic areas covered | Middle East and Africa |
| Number of countries covered | single country (Niger) |
| Study period | start and end date span first epidemic wave |
| **Outcome** | |
| Raw outcome | epidemiological individual-level outcome (individual cases) |
| Time resolution of raw outcome | daily |
| Computed outcome | epidemiological parameter (reproduction number) |
| Method to obtain the computed outcome | statistical estimation of reproduction number (Cori et al.) |
| Data source | data from (sub)national authorities |
| Data availability | data not accessible |
| **Intervention** | |
| Terminology for interventions | measures |
| Terminology for the specific type of non-pharmaceutical interventions | public health |
| Exposure types | all interventions together (timeline of measures) |
| Types of single interventions | – not applicable – |
| Coding of interventions | – not applicable – |

– continued on next page –

| | |
|---|---|
| Source of intervention data | – not applicable – |
| Availability of data on exposure | – not applicable – |
| **Methodological approach** | |
| Empirical approach | descriptive |
| Use of exposure variation | only variation over time for a single population |
| Method | description of change over time (description of time course) |
| Code availability | none |
| **Effect assessment** | |
| Reporting of intervention effect | qualitative statement |
| Measure of effect | reproduction number |
| Reporting of uncertainty | – not applicable – |
| Sensitivity analysis | none |
| Subgroup assessment | none |

**Thai et al. 2020**

**Study information**

– continued on next page –

| | |
|---|---|
| Author | Thai PQ, Rabaa MA, Luong DH, Tan DQ, Quang TD, Quach HL, Thi NA, Dinh PC, Nghia ND, Tu TA, Quang N, Phuc TM, Chau V, Khanh NC, Anh DD, Duong TN, Thwaites G, van Doorn HR, Choisy M, Chambers M, Choisy M, Day J, Trinh DH, Tam DT, Donovan J, Duc DH, Geskus RB, Chanh HQ, Van HH, Thao HD, Huy Hle, Ha HN, Trieu HT, Yen HX, Kestelyn E, Kesteman T, Nguyet LA, Yen LM, Lawson K, Thanh LK, Nhu LN, Nhat LT, Lan LT, Van TL, Lewycka SO, Tran NB, Nguyet NM, Quyen NT, Ngoc NT, Ny NT, Thuong NT, Trang NT, Tuyen NT, Diep NT, Dung NT, Tam NT, Hong NT, Trang NT, Van VC, Truong NX, Van NT, Khanh PN, Lam PK, Yen PL, Nhat PT, Rabaa M, Thuong TN, Thwaites G, Thwaites L, Phuc TM, Thanh TT, Ngoc TT, Hien TT, Van DH, Van NJ, Chau V, Bich VT, Hang VT, And SY |
| Title | The First 100 Days of Severe Acute Respiratory Syndrome Coronavirus 2 (SARS-CoV-2) Control in Vietnam |
| Year | 2020 |
| Journal | Clinical Infectious Diseases |
| DOI | 10.1093/cid/ciaa1130 |
| Date received | 2020-06-02 |
| Date accepted | 2020-07-28 |
| **Study setting** | |
| Number of populations included | single |
| Level of populations included | national (Vietnam) |
| Geographic areas covered | Asia |
| Number of countries covered | single country (Vietnam) |
| Study period | start and end date span first epidemic wave |
| **Outcome** | |
| Raw outcome | epidemiological population-level outcome (cases), behavioral outcome (mobility: number of trips by mode and time spent in various locations) |
| Time resolution of raw outcome | daily |

– continued on next page –

| | |
|---|---|
| Computed outcome | epidemiological parameter (reproduction number) |
| Method to obtain the computed outcome | statistical estimation of reproduction number (Cori et al.) |
| Data source | data from (sub)national authorities (cases), mobility data from corporate organizations (Apple: number of trips by mode, Google: time spent in various locations) |
| Data availability | data access via source |
| **Intervention** | |
| Terminology for interventions | measures |
| Terminology for the specific type of non-pharmaceutical interventions | control |
| Exposure types | all interventions together (timeline of measures) |
| Types of single interventions | – not applicable – |
| Coding of interventions | – not applicable – |
| Source of intervention data | – not applicable – |
| Availability of data on exposure | – not applicable – |
| **Methodological approach** | |
| Empirical approach | descriptive |
| Use of exposure variation | only variation over time for a single population |
| Method | description of change over time (description of time course) |
| Code availability | none |
| **Effect assessment** | |
| Reporting of intervention effect | qualitative statement |
| Measure of effect | cases, reproduction number, mobility: number of trips by mode and and time spent in various locations |
| Reporting of uncertainty | – not applicable – |

– continued on next page –

| | |
|---|---|
| Sensitivity analysis | none |
| Subgroup assessment | none |

| **Tian et al. 2020 (Analysis 1)** | |
|---|---|
| **Study information** | |
| Author | Tian J, Wu AJ, Bao Y, Weng X, Shi L, Liu B, Yu X, Qi L, And ZL |
| Title | Modeling Analysis of COVID-19 Based on Morbidity Data in Anhui, China |
| Year | 2020 |
| Journal | Mathematical Biosciences and Engineering |
| DOI | 10.3934/mbe.2020158 |
| Date received | 2020-02-22 |
| Date accepted | 2020-03-20 |
| **Study setting** | |
| Number of populations included | single |
| Level of populations included | subnational (Anhui province (China)) |
| Geographic areas covered | Asia |
| Number of countries covered | single country (China) |
| Study period | start and end date span first epidemic wave |
| **Outcome** | |
| Raw outcome | epidemiological population-level outcome (deaths) |
| Time resolution of raw outcome | daily |
| Computed outcome | none |

– continued on next page –

| | |
|---|---|
| Method to obtain the computed outcome | – not applicable – |
| Data source | data from (sub)national authorities |
| Data availability | data access via source |
| **Intervention** | |
| Terminology for interventions | measures |
| Terminology for the specific type of non-pharmaceutical interventions | intervention |
| Exposure types | one combination of interventions (Wuhan City Lockdown) |
| Types of single interventions | – not applicable – |
| Coding of interventions | not necessary |
| Source of intervention data | – could not be evaluated – |
| Availability of data on exposure | raw data documented in the manuscript |
| **Methodological approach** | |
| Empirical approach | parametric |
| Use of exposure variation | only variation over time for a single population |
| Method | non-mechanistic model (generalized linear model) |
| Code availability | none |
| **Effect assessment** | |
| Reporting of intervention effect | quantification of change in outcome values |
| Measure of effect | deaths |
| Reporting of uncertainty | yes |
| Sensitivity analysis | none |
| Subgroup assessment | none |

| Tian et al. 2020 (Analysis 2) | |
|---|---|
| **Study information** | |
| Author | Tian J, Wu AJ, Bao Y, Weng X, Shi L, Liu B, Yu X, Qi L, And ZL |
| Title | Modeling Analysis of COVID-19 Based on Morbidity Data in Anhui, China |
| Year | 2020 |
| Journal | Mathematical Biosciences and Engineering |
| DOI | 10.3934/mbe.2020158 |
| Date received | 2020-02-22 |
| Date accepted | 2020-03-20 |
| **Study setting** | |
| Number of populations included | single |
| Level of populations included | subnational (Anhui province (China)) |
| Geographic areas covered | Asia |
| Number of countries covered | single country (China) |
| Study period | start and end date span first epidemic wave |
| **Outcome** | |
| Raw outcome | epidemiological population-level outcome (deaths) |
| Time resolution of raw outcome | daily |
| Computed outcome | epidemiological parameter (reproduction number) |
| Method to obtain the computed outcome | compartmental transmission model (single-population) |
| Data source | data from (sub)national authorities |
| Data availability | data access via source |
| **Intervention** | |
| Terminology for interventions | measures |
| Terminology for the specific type of non-pharmaceutical interventions | intervention |

– continued on next page –

| | |
|---|---|
| Exposure types | one combination of interventions (Wuhan City Lockdown) |
| Types of single interventions | – not applicable – |
| Coding of interventions | not necessary |
| Source of intervention data | – could not be evaluated – |
| Availability of data on exposure | raw data documented in the manuscript |
| **Methodological approach** | |
| Empirical approach | descriptive |
| Use of exposure variation | only variation over time for a single population |
| Method | description of change over time (comparison of time periods) |
| Code availability | none |
| **Effect assessment** | |
| Reporting of intervention effect | comparison of outcome values |
| Measure of effect | reproduction number |
| Reporting of uncertainty | no |
| Sensitivity analysis | none |
| Subgroup assessment | none |

**Tian et al. 2021**

| | |
|---|---|
| **Study information** | |
| Author | Tian T, Luo W, Tan J, Jiang Y, Chen M, Pan W, Yang S, Zhao J, Wang X, Zhang H |
| Title | The Timing and Effectiveness of Implementing Mild Interventions of COVID-19 in Large Industrial Regions via a Synthetic Control Method |
| Year | 2021 |

– continued on next page –

| | |
|---|---|
| Journal | Statistics and Its Interface |
| DOI | 10.4310/20-sii634 |
| Date received | 2020-07-16 |
| Date accepted | 2020-08-30 |
| **Study setting** | |
| Number of populations included | single |
| Level of populations included | subnational (Shenzhen) |
| Geographic areas covered | Asia |
| Number of countries covered | single country (China) |
| Study period | end date in growth phase of wave |
| **Outcome** | |
| Raw outcome | epidemiological population-level outcome (cases) |
| Time resolution of raw outcome | daily |
| Computed outcome | none |
| Method to obtain the computed outcome | – not applicable – |
| Data source | data from other research projects (study by Wu et al.) |
| Data availability | data made available by the authors |
| **Intervention** | |
| Terminology for interventions | interventions |
| Terminology for the specific type of non-pharmaceutical interventions | – not applicable – |
| Exposure types | one combination of interventions (Wuhan City Lockdown) |
| Types of single interventions | – not applicable – |
| Coding of interventions | not necessary |
| Source of intervention data | – could not be evaluated – |

– continued on next page –

| | |
|---|---|
| Availability of data on exposure | raw data documented in the manuscript |
| **Methodological approach** | |
| Empirical approach | counterfactual |
| Use of exposure variation | only variation over time for a single population |
| Method | synthetic controls |
| Code availability | publicly available |
| **Effect assessment** | |
| Reporting of intervention effect | quantification of change in outcome values |
| Measure of effect | cases |
| Reporting of uncertainty | yes |
| Sensitivity analysis | none |
| Subgroup assessment | none |

**Tiwari et al. 2020**

| | |
|---|---|
| **Study information** | |
| Author | Tiwari V, Deyal N, Bisht NS |
| Title | Mathematical Modeling Based Study and Prediction of COVID-19 Epidemic Dissemination under the Impact of Lockdown in India |
| Year | 2020 |
| Journal | Frontiers in Physics |
| DOI | 10.3389/fphy.2020.586899 |
| Date received | 2020-07-24 |
| Date accepted | 2020-09-07 |
| **Study setting** | |

– continued on next page –

| | |
|---|---|
| Number of populations included | single |
| Level of populations included | national (India) |
| Geographic areas covered | Asia |
| Number of countries covered | single country (India) |
| Study period | end date in growth phase of wave |
| **Outcome** | |
| Raw outcome | epidemiological population-level outcome (cases, deaths, recovered cases) |
| Time resolution of raw outcome | daily |
| Computed outcome | epidemiological parameter (reproduction number) |
| Method to obtain the computed outcome | compartmental transmission model (single-population) |
| Data source | data from (sub)national authorities |
| Data availability | data access via source |
| **Intervention** | |
| Terminology for interventions | – not applicable – |
| Terminology for the specific type of non-pharmaceutical interventions | lockdown |
| Exposure types | one combination of interventions (Indian National Lockdown) |
| Types of single interventions | – not applicable – |
| Coding of interventions | not necessary |
| Source of intervention data | government or news websites |
| Availability of data on exposure | raw data documented in the manuscript |
| **Methodological approach** | |
| Empirical approach | descriptive |
| Use of exposure variation | only variation over time for a single population |
| Method | description of change over time (comparison of time periods) |

– continued on next page –

| | |
|---|---|
| Code availability | none |
| **Effect assessment** | |
| Reporting of intervention effect | qualitative statement |
| Measure of effect | cases, reproduction number |
| Reporting of uncertainty | – not applicable – |
| Sensitivity analysis | none |
| Subgroup assessment | none |

| **Tsai et al. 2020** | |
|---|---|
| **Study information** | |
| Author | Tsai AC, Harling G, Reynolds Z, Gilbert RF, Siedner MJ |
| Title | Coronavirus Disease 2019 (COVID-19) Transmission in the United States before versus after Relaxation of Statewide Social Distancing Measures |
| Year | 2020 |
| Journal | Clinical Infectious Diseases |
| DOI | 10.1093/cid/ciaa1502 |
| Date received | 2020-08-04 |
| Date accepted | 2020-09-28 |
| **Study setting** | |
| Number of populations included | multiple |
| Level of populations included | both national and subnational (US and its states) |
| Geographic areas covered | North America |
| Number of countries covered | single country (US) |
| Study period | same end date for several populations with diverse epidemic trajectories |

– continued on next page –

| | |
|---|---|
| **Outcome** | |
| Raw outcome | epidemiological population-level outcome (effective reproduction number) |
| Time resolution of raw outcome | daily |
| Computed outcome | none |
| Method to obtain the computed outcome | – not applicable – |
| Data source | data from other research projects (study by Unwin et al.) |
| Data availability | data access via source |
| **Intervention** | |
| Terminology for interventions | measures |
| Terminology for the specific type of non-pharmaceutical interventions | social distancing |
| Exposure types | multiple separate interventions (school closure (reopening of schools), workplace closure (easing of working restrictions), venue closure (reopening of businesses, reopening of outdoor recreational facilities), stay-at-home order (rescission of state-wide restrictions on movement: shelter-in-place order or lockdown), travel restrictions (rescission of mandatory quarantines for interstate travel)) |
| Types of single interventions | school closure, stay-at-home order, international travel restrictions, workplace closure, venue closure |
| Coding of interventions | necessary |
| Source of intervention data | coding done by authors |
| Availability of data on exposure | coded data made available by the authors |
| **Methodological approach** | |
| Empirical approach | parametric |
| Use of exposure variation | both variation over time and between populations |
| Method | non-mechanistic model (generalized linear model) |

– continued on next page –

| | |
|---|---|
| Code availability | none |
| **Effect assessment** | |
| Reporting of intervention effect | quantification of change in outcome values |
| Measure of effect | reproduction number |
| Reporting of uncertainty | yes |
| Sensitivity analysis | different or modified outcome used (cases and deaths used as raw different or modified outcome used, estimates of effective reproduction number used from other paper), different coding of interventions used (days since relaxation of SIPO used as primary explanatory variable), model specification varied |
| Subgroup assessment | none |

**Turk et al. 2020**

| | |
|---|---|
| **Study information** | |
| Author | Turk PJ, Chou SH, Kowalkowski MA, Palmer PP, Priem JS, Spencer MD, Taylor YJ, McWilliams AD |
| Title | Modeling COVID-19 Latent Prevalence to Assess a Public Health Intervention at a State and Regional Scale: Retrospective Cohort Study |
| Year | 2020 |
| Journal | JMIR Public Health and Surveillance |
| DOI | 10.2196/19353 |
| Date received | 2020-04-14 |
| Date accepted | 2020-05-18 |
| **Study setting** | |
| Number of populations included | multiple |

– continued on next page –

| | |
|---|---|
| Level of populations included | subnational (North Carolina and Charlotte region) |
| Geographic areas covered | North America |
| Number of countries covered | single country (US) |
| Study period | end date in growth phase of wave |
| **Outcome** | |
| Raw outcome | epidemiological population-level outcome (cases, deaths) |
| Time resolution of raw outcome | daily |
| Computed outcome | measure of epidemic trend (doubling time), epidemiological parameter (reproduction number) |
| Method to obtain the computed outcome | compartmental transmission model (single-population) |
| Data source | data from (sub)national authorities |
| Data availability | data access via source |
| **Intervention** | |
| Terminology for interventions | interventions |
| Terminology for the specific type of non-pharmaceutical interventions | public health |
| Exposure types | one single intervention (stay-at-home order (in Mecklenburg & Co)) |
| Types of single interventions | stay-at-home order |
| Coding of interventions | not necessary |
| Source of intervention data | – could not be evaluated – |
| Availability of data on exposure | raw data documented in the manuscript |
| **Methodological approach** | |
| Empirical approach | descriptive |
| Use of exposure variation | only variation over time for multiple populations |
| Method | description of change over time (comparison of time periods) |

– continued on next page –

| | |
|---|---|
| Code availability | none |
| **Effect assessment** | |
| Reporting of intervention effect | comparison of outcome values |
| Measure of effect | reproduction number, doubling time |
| Reporting of uncertainty | yes |
| Sensitivity analysis | none |
| Subgroup assessment | none |

**Tutsoy et al. 2020**

| | |
|---|---|
| **Study information** | |
| Author | Tutsoy O, Polat A, Colak S, Balikci K |
| Title | Development of a Multi-Dimensional Parametric Model with Non-Pharmacological Policies for Predicting the COVID-19 Pandemic Casualties |
| Year | 2020 |
| Journal | IEEE Access |
| DOI | 10.1109/access.2020.3044929 |
| Date received | 2020-12-09 |
| Date accepted | 2020-12-11 |
| **Study setting** | |
| Number of populations included | single |
| Level of populations included | national (Turkey) |
| Geographic areas covered | Middle East and Africa |
| Number of countries covered | single country (Turkey) |
| Study period | start and end date span first epidemic wave |

– continued on next page –

| | |
|---|---|
| **Outcome** | |
| Raw outcome | epidemiological population-level outcome (tests, cases, deaths) |
| Time resolution of raw outcome | daily |
| Computed outcome | none |
| Method to obtain the computed outcome | – not applicable – |
| Data source | data from (sub)national authorities |
| Data availability | data access via source |
| **Intervention** | |
| Terminology for interventions | policies |
| Terminology for the specific type of non-pharmaceutical interventions | non-pharmacological |
| Exposure types | multiple separate interventions (stay-at-home order (curfews for people with chronic diseases, curfews for people over 65, curfews for people under 20, curfews for everyone), school closure) |
| Types of single interventions | stay-at-home order, school closure |
| Coding of interventions | not necessary |
| Source of intervention data | – could not be evaluated – |
| Availability of data on exposure | raw data documented in the manuscript |
| **Methodological approach** | |
| Empirical approach | parametric |
| Use of exposure variation | only variation over time for a single population |
| Method | other (non-standard, semi-mechanistic transmission model) |
| Code availability | none |
| **Effect assessment** | |
| Reporting of intervention effect | quantification of change in outcome values |

– continued on next page –

| | |
|---|---|
| Measure of effect | tests, cases, deaths |
| Reporting of uncertainty | no |
| Sensitivity analysis | none |
| Subgroup assessment | none |

**Utamura et al. 2020**

| | |
|---|---|
| **Study information** | |
| Author | Utamura M, Koizumi M, Kirikami S |
| Title | An Epidemiological Model Considering Isolation to Predict COVID-19 Trends in Tokyo, Japan: Numerical Analysis |
| Year | 2020 |
| Journal | JMIR Public Health and Surveillance |
| DOI | 10.2196/23624 |
| Date received | 2020-08-18 |
| Date accepted | 2020-11-30 |
| **Study setting** | |
| Number of populations included | single |
| Level of populations included | subnational (Tokyo (Japan)) |
| Geographic areas covered | Asia |
| Number of countries covered | single country (Japan) |
| Study period | start and end date span first epidemic wave |
| **Outcome** | |
| Raw outcome | epidemiological population-level outcome (cases) |
| Time resolution of raw outcome | daily |

– continued on next page –

| | |
|---|---|
| Computed outcome | none |
| Method to obtain the computed outcome | – not applicable – |
| Data source | data from (sub)national authorities |
| Data availability | data access via source |
| **Intervention** | |
| Terminology for interventions | interventions |
| Terminology for the specific type of non-pharmaceutical interventions | public health |
| Exposure types | one single intervention (stay-at-home order) |
| Types of single interventions | stay-at-home order |
| Coding of interventions | not necessary |
| Source of intervention data | – could not be evaluated – |
| Availability of data on exposure | raw data documented in the manuscript |
| **Methodological approach** | |
| Empirical approach | parametric |
| Use of exposure variation | only variation over time for a single population |
| Method | mechanistic model (compartmental single-population transmission model) |
| Latent variable functionally linked to intervention | transmission rate |
| Code availability | none |
| **Effect assessment** | |
| Reporting of intervention effect | quantification of change in outcome values |
| Measure of effect | transmission rate |
| Reporting of uncertainty | no |

– continued on next page –

| | |
|---|---|
| Sensitivity analysis | none |
| Subgroup assessment | none |

**Valcarcel et al. 2020**

| | |
|---|---|
| **Study information** | |
| Author | Valcarcel B, Avilez JL, Torres-Roman JS, Poterico JA, Bazalar-Palacios J, Vecchia C |
| Title | The Effect of Early-Stage Public Health Policies in the Transmission of COVID-19 for South American Countries |
| Year | 2020 |
| Journal | Revista Panamericana de Salud Pública |
| DOI | 10.26633/rpsp.2020.148 |
| Date received | – could not be evaluated – |
| Date accepted | 2020-11-10 (published) |
| **Study setting** | |
| Number of populations included | multiple |
| Level of populations included | national (10 South American countries) |
| Geographic areas covered | Central and South America |
| Number of countries covered | multiple countries |
| Study period | start and end date span first epidemic wave |
| **Outcome** | |
| Raw outcome | epidemiological population-level outcome (cases) |
| Time resolution of raw outcome | daily |
| Computed outcome | epidemiological parameter (reproduction number) |

– continued on next page –

| | |
|---|---|
| Method to obtain the computed outcome | statistical estimation of reproduction number (Thompson et al.) |
| Data source | data from publicly available cross-country selections (ECDC) |
| Data availability | data made available by the authors |
| **Intervention** | |
| Terminology for interventions | measures |
| Terminology for the specific type of non-pharmaceutical interventions | public health |
| Exposure types | multiple combinations of interventions (group 1: isolation, quarantine, and social distancing, group 2: community-wide containment) |
| Types of single interventions | – not applicable – |
| Coding of interventions | necessary |
| Source of intervention data | – could not be evaluated – |
| Availability of data on exposure | coded data not available |
| **Methodological approach** | |
| Empirical approach | descriptive |
| Use of exposure variation | only variation over time for multiple populations |
| Method | comparison of points in time (description of time course) |
| Code availability | publicly available |
| **Effect assessment** | |
| Reporting of intervention effect | comparison of outcome values |
| Measure of effect | reproduction number |
| Reporting of uncertainty | yes |
| Sensitivity analysis | model specification varied |
| Subgroup assessment | public health response (response time) |

| Valencia et al. 2020 (Analysis 1) | |
|---|---|
| **Study information** | |
| Author | Valencia M, Becerra JE, Reyes JC, Castro KG |
| Title | Assessment of Early Mitigation Measures against COVID-19 in Puerto Rico: March 15-May 15, 2020 |
| Year | 2020 |
| Journal | PLOS ONE |
| DOI | 10.1371/journal.pone.0240013 |
| Date received | 2020-06-05 |
| Date accepted | 2020-09-17 |
| **Study setting** | |
| Number of populations included | single |
| Level of populations included | subnational (Puerto Rico) |
| Geographic areas covered | North America |
| Number of countries covered | single country (US) |
| Study period | start and end date span first epidemic wave |
| **Outcome** | |
| Raw outcome | epidemiological population-level outcome (cases, deaths) |
| Time resolution of raw outcome | daily |
| Computed outcome | epidemiological parameter (reproduction number) |
| Method to obtain the computed outcome | exponential growth model |
| Data source | data from (sub)national authorities |
| Data availability | data made available by the authors |

– continued on next page –

| | |
|---|---|
| **Intervention** | |
| Terminology for interventions | interventions |
| Terminology for the specific type of non-pharmaceutical interventions | non-pharmaceutical |
| Exposure types | one combination of interventions (curfew (as part of a state of emergency declaration closing public and private schools and universities, all nonessential businesses and public agencies, and implementing travel restrictions)) |
| Types of single interventions | – not applicable – |
| Coding of interventions | not necessary |
| Source of intervention data | government or news websites |
| Availability of data on exposure | raw data documented in the manuscript |
| **Methodological approach** | |
| Empirical approach | descriptive |
| Use of exposure variation | only variation over time for a single population |
| Method | description of change over time (comparison of time periods) |
| Code availability | publicly available |
| **Effect assessment** | |
| Reporting of intervention effect | comparison of outcome values |
| Measure of effect | reproduction number |
| Reporting of uncertainty | yes |
| Sensitivity analysis | none |
| Subgroup assessment | none |

| Valencia et al. 2020 (Analysis 2) | |
|---|---|
| **Study information** | |
| Author | Valencia M, Becerra JE, Reyes JC, Castro KG |
| Title | Assessment of Early Mitigation Measures against COVID-19 in Puerto Rico: March 15-May 15, 2020 |
| Year | 2020 |
| Journal | PLOS ONE |
| DOI | 10.1371/journal.pone.0240013 |
| Date received | 2020-06-05 |
| Date accepted | 2020-09-17 |
| **Study setting** | |
| Number of populations included | single |
| Level of populations included | subnational (Puerto Rico) |
| Geographic areas covered | North America |
| Number of countries covered | single country (US) |
| Study period | start and end date span first epidemic wave |
| **Outcome** | |
| Raw outcome | epidemiological population-level outcome (cases, deaths) |
| Time resolution of raw outcome | daily |
| Computed outcome | none |
| Method to obtain the computed outcome | – not applicable – |
| Data source | data from (sub)national authorities |
| Data availability | data made available by the authors |
| **Intervention** | |
| Terminology for interventions | interventions |

– continued on next page –

| | |
|---|---|
| Terminology for the specific type of non-pharmaceutical interventions | non-pharmaceutical |
| Exposure types | one combination of interventions (curfew (as part of a state of emergency declaration closing public and private schools and universities, all nonessential businesses and public agencies, and implementing travel restrictions)) |
| Types of single interventions | – not applicable – |
| Coding of interventions | not necessary |
| Source of intervention data | government or news websites |
| Availability of data on exposure | raw data documented in the manuscript |
| **Methodological approach** | |
| Empirical approach | counterfactual |
| Use of exposure variation | only variation over time for a single population |
| Method | non-mechanistic model (exponential growth model) |
| Code availability | publicly available |
| **Effect assessment** | |
| Reporting of intervention effect | comparison of outcome values |
| Measure of effect | cases, deaths |
| Reporting of uncertainty | no |
| Sensitivity analysis | none |
| Subgroup assessment | none |

**Van Dyke et al. 2020**

| | |
|---|---|
| **Study information** | |
| Author | Van Dyke ME, Rogers TM, Pevzner E, Satterwhite CL, Shah HB, Beckman WJ, Ahmed F, Hunt DC, Rule J |

– continued on next page –

| | |
|---|---|
| Title | Trends in County-Level COVID-19 Incidence in Counties with and without a Mask Mandate —Kansas, June 1–August 23, 2020 |
| Year | 2020 |
| Journal | Morbidity and Mortality Weekly Report |
| DOI | 10.15585/mmwr.mm6947e2 |
| Date received | – could not be evaluated – |
| Date accepted | 2020-11-20 |
| **Study setting** | |
| Number of populations included | multiple |
| Level of populations included | subnational (Counties in Kansas) |
| Geographic areas covered | North America |
| Number of countries covered | single country (US) |
| Study period | includes second wave (two waves) |
| **Outcome** | |
| Raw outcome | epidemiological population-level outcome (cases) |
| Time resolution of raw outcome | daily |
| Computed outcome | none |
| Method to obtain the computed outcome | – not applicable – |
| Data source | public (USAFacts) |
| Data availability | data access via source |
| **Intervention** | |
| Terminology for interventions | strategies |
| Terminology for the specific type of non-pharmaceutical interventions | mitigation |
| Exposure types | one single intervention (mask mandate) |

– continued on next page –

| | |
|---|---|
| Types of single interventions | mask mandate |
| Coding of interventions | necessary |
| Source of intervention data | use of externally coded data (Kansas Health Institute) |
| Availability of data on exposure | access to externally coded data via source |
| **Methodological approach** | |
| Empirical approach | parametric |
| Use of exposure variation | both variation over time and between populations |
| Method | non-mechanistic model (generalized linear model) |
| Code availability | none |
| **Effect assessment** | |
| Reporting of intervention effect | quantification of change in outcome values |
| Measure of effect | cases |
| Reporting of uncertainty | yes |
| Sensitivity analysis | different coding of interventions used (different NPI coding, i.e., nonmandated counties that included cities mandating masks coded as mandated counties) |
| Subgroup assessment | public health response (subject to other measures: with and without other NPIs) |

**Vannoni et al. 2020**

| | |
|---|---|
| **Study information** | |
| Author | Vannoni M, McKee M, Semenza JC, Bonell C, Stuckler D |
| Title | Using Volunteered Geographic Information to Assess Mobility in the Early Phases of the COVID-19 Pandemic: A Cross-City Time Series Analysis of 41 Cities in 22 Countries from March 2nd to 26th 2020 |

– continued on next page –

| | |
|---|---|
| Year | 2020 |
| Journal | Globalization and Health |
| DOI | 10.1186/s12992-020-00598-9 |
| Date received | 2020-04-21 |
| Date accepted | 2020-07-13 |
| **Study setting** | |
| Number of populations included | multiple |
| Level of populations included | subnational (41 cities in 22 countries) |
| Geographic areas covered | Europe, Middle East and Africa, Asia, Oceania, Central and South America, North America |
| Number of countries covered | multiple countries |
| Study period | start and end date span first epidemic wave: behavioral |
| **Outcome** | |
| Raw outcome | behavioral outcome (mobility: mobility index based on number of trips planned compared to a reference period) |
| Time resolution of raw outcome | daily |
| Computed outcome | none |
| Method to obtain the computed outcome | – not applicable – |
| Data source | mobility data from corporate organizations (Citymapper) |
| Data availability | data access via source |
| **Intervention** | |
| Terminology for interventions | interventions |
| Terminology for the specific type of non-pharmaceutical interventions | non-pharmaceutical |

– continued on next page –

| | |
|---|---|
| Exposure types | multiple separate interventions (school closure, bans of large gatherings, workplace closure, international travel restrictions, internal movement restrictions, public transport closure) |
| Types of single interventions | school closure, bans of large gatherings, workplace closure, international travel restrictions, internal movement restrictions, public transport closure |
| Coding of interventions | necessary |
| Source of intervention data | use of externally coded data (OxCGRT) |
| Availability of data on exposure | access to externally coded data via source |
| **Methodological approach** | |
| Empirical approach | parametric |
| Use of exposure variation | both variation over time and between populations |
| Method | non-mechanistic model (generalized linear model) |
| Code availability | none |
| **Effect assessment** | |
| Reporting of intervention effect | quantification of change in outcome values |
| Measure of effect | mobility: mobility index based on number of trips planned compared to a reference period |
| Reporting of uncertainty | yes |
| Sensitivity analysis | different or modified outcome used (different mobility data used) |
| Subgroup assessment | none |

**Verma et al. 2020**

| | |
|---|---|
| **Study information** | |
| Author | Verma BK, Verma M, Verma VK, Abdullah RB, Nath DC, Khan HT, Verma A, Vishwakarma RK, Verma V |

– continued on next page –

| | |
|---|---|
| Title | Global Lockdown: An Effective Safeguard in Responding to the Threat of COVID-19 |
| Year | 2020 |
| Journal | Journal of Evaluation in Clinical Practice |
| DOI | 10.1111/jep.13483 |
| Date received | 2020-06-22 |
| Date accepted | 2020-09-01 |
| **Study setting** | |
| Number of populations included | multiple |
| Level of populations included | national (United States, Italy, United Kingdom, France, India, Russia) |
| Geographic areas covered | Asia, Europe, North America |
| Number of countries covered | multiple countries |
| Study period | same end date for several populations with diverse epidemic trajectories |
| **Outcome** | |
| Raw outcome | epidemiological population-level outcome (cases) |
| Time resolution of raw outcome | daily |
| Computed outcome | change points (change points: cases) |
| Method to obtain the computed outcome | change point analysis |
| Data source | data from publicly available cross-country selections (OurWorldInData) |
| Data availability | data access via source |
| **Intervention** | |
| Terminology for interventions | – not applicable – |
| Terminology for the specific type of non-pharmaceutical interventions | lockdown |

– continued on next page –

| | |
|---|---|
| Exposure types | one combination of interventions (lockdown with specific date for each country) |
| Types of single interventions | – not applicable – |
| Coding of interventions | not necessary |
| Source of intervention data | – could not be evaluated – |
| Availability of data on exposure | raw data documented in the manuscript |
| **Methodological approach** | |
| Empirical approach | descriptive |
| Use of exposure variation | only variation over time for multiple populations |
| Method | comparison of change points with intervention dates |
| Code availability | none |
| **Effect assessment** | |
| Reporting of intervention effect | quantification of change in outcome values |
| Measure of effect | change points: cases |
| Reporting of uncertainty | yes |
| Sensitivity analysis | none |
| Subgroup assessment | none |

**Vicentini et al. 2020**

| | |
|---|---|
| **Study information** | |
| Author | Vicentini C, Bordino V, Gardois P, Zotti CM |
| Title | Early Assessment of the Impact of Mitigation Measures on the COVID-19 Outbreak in Italy |
| Year | 2020 |

– continued on next page –

| | |
|---|---|
| Journal | Public Health |
| DOI | 10.1016/j.puhe.2020.06.028 |
| Date received | 2020-04-28 |
| Date accepted | 2020-06-10 |
| **Study setting** | |
| Number of populations included | single |
| Level of populations included | national (Italy) |
| Geographic areas covered | Europe |
| Number of countries covered | single country (Italy) |
| Study period | start and end date span first epidemic wave |
| **Outcome** | |
| Raw outcome | epidemiological population-level outcome (ICU hospitalizations) |
| Time resolution of raw outcome | daily |
| Computed outcome | none |
| Method to obtain the computed outcome | – not applicable – |
| Data source | data from (sub)national authorities |
| Data availability | data access via source |
| **Intervention** | |
| Terminology for interventions | measures |
| Terminology for the specific type of non-pharmaceutical interventions | mitigation |
| Exposure types | multiple combinations of interventions (lockdown of the epicenter of the outbreak, school closure and nationwide lockdown) |
| Types of single interventions | – not applicable – |
| Coding of interventions | not necessary |

– continued on next page –

| | |
|---|---|
| Source of intervention data | – could not be evaluated – |
| Availability of data on exposure | raw data documented in the manuscript |
| **Methodological approach** | |
| Empirical approach | counterfactual |
| Use of exposure variation | only variation over time for a single population |
| Method | non-mechanistic model (exponential growth model) |
| Code availability | none |
| **Effect assessment** | |
| Reporting of intervention effect | comparison of outcome values |
| Measure of effect | ICU hospitalizations |
| Reporting of uncertainty | no |
| Sensitivity analysis | none |
| Subgroup assessment | none |

**Wagner et al. 2020**

| | |
|---|---|
| **Study information** | |
| Author | Wagner AB, Hill EL, Ryan SE, Sun Z, Deng G, Bhadane S, Martinez VH, Wu P, Li D, Anand A, Acharya J, Matteson DS |
| Title | Social Distancing Merely Stabilized COVID-19 in the United States |
| Year | 2020 |
| Journal | Stat |
| DOI | 10.1002/sta4.302 |
| Date received | 2020-05-11 |
| Date accepted | 2020-06-28 |

– continued on next page –

| | |
|---|---|
| **Study setting** | |
| Number of populations included | multiple |
| Level of populations included | both national and subnational (US and all states) |
| Geographic areas covered | North America |
| Number of countries covered | single country (US) |
| Study period | same end date for several populations with diverse epidemic trajectories |
| **Outcome** | |
| Raw outcome | epidemiological population-level outcome (cases) |
| Time resolution of raw outcome | daily |
| Computed outcome | none |
| Method to obtain the computed outcome | – not applicable – |
| Data source | data from public media outlets (New York Times) |
| Data availability | data made available by the authors |
| **Intervention** | |
| Terminology for interventions | measures |
| Terminology for the specific type of non-pharmaceutical interventions | social distancing |
| Exposure types | start of interventions (first NPI out of either school closure or venue closure (restaurant closures)) |
| Types of single interventions | – not applicable – |
| Coding of interventions | necessary |
| Source of intervention data | use of externally coded data (Education Week, Wida) |
| Availability of data on exposure | coded data made available by the authors |
| **Methodological approach** | |
| Empirical approach | parametric |

– continued on next page –

| | |
|---|---|
| Use of exposure variation | only variation over time for multiple populations |
| Method | non-mechanistic model (generalized linear model) |
| Code availability | publicly available |
| **Effect assessment** | |
| Reporting of intervention effect | quantification of change in outcome values |
| Measure of effect | doubling time |
| Reporting of uncertainty | yes |
| Sensitivity analysis | different or modified outcome used (deaths used instead of confirmed cases), model specification varied |
| Subgroup assessment | none |

**Wahaibi et al. 2020**

| | |
|---|---|
| **Study information** | |
| Author | Wahaibi AA, Manji AA, Maani AA, Rawahi BA, Harthy KA, Alyaquobi F, Al-Jardani A, Petersen E, Abri SA |
| Title | COVID-19 Epidemic Monitoring after Non-Pharmaceutical Interventions: The Use of Time-Varying Reproduction Number in a Country with a Large Migrant Population |
| Year | 2020 |
| Journal | International Journal of Infectious Diseases |
| DOI | 10.1016/j.ijid.2020.08.039 |
| Date received | 2020-07-27 |
| Date accepted | 2020-08-16 |
| **Study setting** | |
| Number of populations included | single |

– continued on next page –

| | |
|---|---|
| Level of populations included | national (Oman) |
| Geographic areas covered | Middle East and Africa |
| Number of countries covered | single country (Oman) |
| Study period | end date in growth phase of wave |
| **Outcome** | |
| Raw outcome | epidemiological individual-level outcome (individual cases and transmission chains) |
| Time resolution of raw outcome | daily |
| Computed outcome | epidemiological parameter (reproduction number) |
| Method to obtain the computed outcome | statistical estimation of reproduction number (Cori et al.) |
| Data source | data from (sub)national authorities |
| Data availability | data not accessible |
| **Intervention** | |
| Terminology for interventions | interventions |
| Terminology for the specific type of non-pharmaceutical interventions | non-pharmaceutical |
| Exposure types | phases of interventions (various measures separated into three phases) |
| Types of single interventions | – not applicable – |
| Coding of interventions | not necessary |
| Source of intervention data | – could not be evaluated – |
| Availability of data on exposure | raw data documented in the manuscript |
| **Methodological approach** | |
| Empirical approach | descriptive |
| Use of exposure variation | only variation over time for a single population |
| Method | description of change over time (comparison of time periods) |

– continued on next page –

| | |
|---|---|
| Code availability | none |
| **Effect assessment** | |
| Reporting of intervention effect | comparison of outcome values |
| Measure of effect | reproduction number |
| Reporting of uncertainty | yes |
| Sensitivity analysis | none |
| Subgroup assessment | socioeconomic indicators (nationality) |

| **Wan et al. 2020** | |
|---|---|
| **Study information** | |
| Author | Wan H, Cui JA, Yang GJ |
| Title | Risk Estimation and Prediction of the Transmission of Coronavirus Disease-2019 (COVID-19) in the Mainland of China Excluding Hubei Province |
| Year | 2020 |
| Journal | Infectious Diseases of Poverty |
| DOI | 10.1186/s40249-020-00683-6 |
| Date received | 2020-03-02 |
| Date accepted | 2020-05-28 |
| **Study setting** | |
| Number of populations included | single |
| Level of populations included | national (China exluding Hubei) |
| Geographic areas covered | Asia |
| Number of countries covered | single country (China) |
| Study period | start and end date span first epidemic wave |

– continued on next page –

| | |
|---|---|
| **Outcome** | |
| Raw outcome | epidemiological population-level outcome (cases, deaths, recovered cases) |
| Time resolution of raw outcome | daily |
| Computed outcome | epidemiological parameter (reproduction number) |
| Method to obtain the computed outcome | compartmental transmission model (single-population) |
| Data source | data from (sub)national authorities |
| Data availability | data access via source |
| **Intervention** | |
| Terminology for interventions | strategies, measures |
| Terminology for the specific type of non-pharmaceutical interventions | containment, control, self-protection |
| Exposure types | all interventions together |
| Types of single interventions | – not applicable – |
| Coding of interventions | – not applicable – |
| Source of intervention data | – not applicable – |
| Availability of data on exposure | – not applicable – |
| **Methodological approach** | |
| Empirical approach | descriptive |
| Use of exposure variation | only variation over time for a single population |
| Method | description of change over time (description of time course) |
| Code availability | none |
| **Effect assessment** | |
| Reporting of intervention effect | comparison of outcome values |
| Measure of effect | reproduction number |
| Reporting of uncertainty | no |

– continued on next page –

| | |
|---|---|
| Sensitivity analysis | none |
| Subgroup assessment | none |

**Wang et al. 2020a**

| | |
|---|---|
| **Study information** | |
| Author | Wang Q, Xie S, Wang Y, Zeng D |
| Title | Survival-Convolution Models for Predicting COVID-19 Cases and Assessing Effects of Mitigation Strategies |
| Year | 2020 |
| Journal | Frontiers in Public Health |
| DOI | 10.3389/fpubh.2020.00325 |
| Date received | 2020-05-11 |
| Date accepted | 2020-06-15 |
| **Study setting** | |
| Number of populations included | multiple |
| Level of populations included | national (Italy and US) |
| Geographic areas covered | Europe, North America |
| Number of countries covered | multiple countries |
| Study period | start and end date span first epidemic wave |
| **Outcome** | |
| Raw outcome | epidemiological population-level outcome (cases) |
| Time resolution of raw outcome | daily |
| Computed outcome | none |

– continued on next page –

| | |
|---|---|
| Method to obtain the computed outcome | – not applicable – |
| Data source | data from publicly available cross-country selections (Worldometer) |
| Data availability | data made available by the authors |
| **Intervention** | |
| Terminology for interventions | interventions |
| Terminology for the specific type of non-pharmaceutical interventions | public health |
| Exposure types | one combination of interventions (nationwide lockdown of March 11 (Italy), emergency declaration (national US)) |
| Types of single interventions | – not applicable – |
| Coding of interventions | not necessary |
| Source of intervention data | – could not be evaluated – |
| Availability of data on exposure | raw data documented in the manuscript |
| **Methodological approach** | |
| Empirical approach | parametric |
| Use of exposure variation | only variation over time for multiple populations |
| Method | other (discrete, deterministic, non-compartmental transmission model fit via gradient-based optimization) |
| Latent variable functionally linked to intervention | transmission rate |
| Code availability | publicly available |
| **Effect assessment** | |
| Reporting of intervention effect | quantification of change in outcome values |
| Measure of effect | reproduction number, transmission rate |
| Reporting of uncertainty | yes |

– continued on next page –

| | |
|---|---|
| Sensitivity analysis | none |
| Subgroup assessment | none |

| **Wang et al. 2020b** | |
|---|---|
| **Study information** | |
| Author | Wang T, Wu Y, Lau JY, Yu Y, Liu L, Li J, Zhang K, Tong W, Jiang B |
| Title | A Four-Compartment Model for the COVID-19 Infection —Implications on Infection Kinetics, Control Measures, and Lockdown Exit Strategies |
| Year | 2020 |
| Journal | Precision Clinical Medicine |
| DOI | 10.1093/pcmedi/pbaa018 |
| Date received | 2020-05-21 |
| Date accepted | 2020-05-22 |
| **Study setting** | |
| Number of populations included | multiple |
| Level of populations included | both national and subnational (Italy, UK, US, Wuhan) |
| Geographic areas covered | Asia, Europe, North America |
| Number of countries covered | multiple countries |
| Study period | end date at peak of wave |
| **Outcome** | |
| Raw outcome | epidemiological population-level outcome (cases) |
| Time resolution of raw outcome | daily |
| Computed outcome | none |

– continued on next page –

| | |
|---|---|
| Method to obtain the computed outcome | – not applicable – |
| Data source | – could not be evaluated – |
| Data availability | data not accessible |
| **Intervention** | |
| Terminology for interventions | measures |
| Terminology for the specific type of non-pharmaceutical interventions | control |
| Exposure types | one combination of interventions (Wuhan City Lockdown (China), lockdown decreed on March 10 (Italy), lockdown decreed March 23 and strengthened on March 30 (UK), shelter-in-place order (US)) |
| Types of single interventions | – not applicable – |
| Coding of interventions | not necessary |
| Source of intervention data | – could not be evaluated – |
| Availability of data on exposure | raw data documented in the manuscript |
| **Methodological approach** | |
| Empirical approach | parametric |
| Use of exposure variation | only variation over time for multiple populations |
| Method | mechanistic model (compartmental single-population transmission model) |
| Latent variable functionally linked to intervention | contact rate, probability of transmission upon contact |
| Code availability | none |
| **Effect assessment** | |
| Reporting of intervention effect | quantification of change in outcome values |
| Measure of effect | contact rate, probability of transmission upon contact |
| Reporting of uncertainty | no |

– continued on next page –

| | |
|---|---|
| Sensitivity analysis | none |
| Subgroup assessment | none |

| **Wang et al. 2020c (Analysis 1)** | |
|---|---|
| **Study information** | |
| Author | Wang S, Yang X, Li L, Nadler P, Arcucci R, Huang Y, Teng Z, Guo Y |
| Title | A Bayesian Updating Scheme for Pandemics: Estimating the Infection Dynamics of COVID-19 |
| Year | 2020 |
| Journal | IEEE Computational Intelligence Magazine |
| DOI | 10.1109/mci.2020.3019874 |
| Date received | – could not be evaluated – |
| Date accepted | 2020-10-15 |
| **Study setting** | |
| Number of populations included | multiple |
| Level of populations included | national (14 European countries) |
| Geographic areas covered | Europe |
| Number of countries covered | multiple countries |
| Study period | start and end date span first epidemic wave |
| **Outcome** | |
| Raw outcome | epidemiological population-level outcome (cases) |
| Time resolution of raw outcome | daily |

– continued on next page –

| | |
|---|---|
| Computed outcome | epidemiological parameter (reproduction number, infectious period length, overall infectiousness) |
| Method to obtain the computed outcome | semi-mechanistic Bayesian transmission model |
| Data source | data from publicly available cross-country selections (JHU) |
| Data availability | data access via source |
| **Intervention** | |
| Terminology for interventions | measures |
| Terminology for the specific type of non-pharmaceutical interventions | intervention |
| Exposure types | stringency index (OxCGRT Stringency Index (categorized into five levels)) |
| Types of single interventions | – not applicable – |
| Coding of interventions | necessary |
| Source of intervention data | use of externally coded data (OxCGRT) |
| Availability of data on exposure | access to externally coded data via source |
| **Methodological approach** | |
| Empirical approach | parametric |
| Use of exposure variation | both variation over time and between populations |
| Method | non-mechanistic model (generalized linear model) |
| Code availability | none |
| **Effect assessment** | |
| Reporting of intervention effect | quantification of change in outcome values |
| Measure of effect | reproduction number, infectious period length, overall infectiousness |
| Reporting of uncertainty | yes |
| Sensitivity analysis | none |
| Subgroup assessment | none |

| Wang et al. 2020c (Analysis 2) | |
|---|---|
| **Study information** | |
| Author | Wang S, Yang X, Li L, Nadler P, Arcucci R, Huang Y, Teng Z, Guo Y |
| Title | A Bayesian Updating Scheme for Pandemics: Estimating the Infection Dynamics of COVID-19 |
| Year | 2020 |
| Journal | IEEE Computational Intelligence Magazine |
| DOI | 10.1109/mci.2020.3019874 |
| Date received | – could not be evaluated – |
| Date accepted | 2020-10-15 |
| **Study setting** | |
| Number of populations included | single |
| Level of populations included | subnational (Wuhan) |
| Geographic areas covered | Asia |
| Number of countries covered | single country (China) |
| Study period | start and end date span first epidemic wave |
| **Outcome** | |
| Raw outcome | epidemiological population-level outcome (cases) |
| Time resolution of raw outcome | daily |
| Computed outcome | epidemiological parameter (reproduction number, infectious period length, overall infectiousness) |
| Method to obtain the computed outcome | semi-mechanistic Bayesian transmission model |
| Data source | data from (sub)national authorities |
| Data availability | data access via source |
| **Intervention** | |

– continued on next page –

| | |
|---|---|
| Terminology for interventions | measures |
| Terminology for the specific type of non-pharmaceutical interventions | intervention |
| Exposure types | one combination of interventions (Wuhan City Lockdown) |
| Types of single interventions | – not applicable – |
| Coding of interventions | not necessary |
| Source of intervention data | – could not be evaluated – |
| Availability of data on exposure | raw data documented in the manuscript |
| **Methodological approach** | |
| Empirical approach | descriptive |
| Use of exposure variation | only variation over time for a single population |
| Method | description of change over time (comparison of time periods) |
| Code availability | none |
| **Effect assessment** | |
| Reporting of intervention effect | quantification of change in outcome values |
| Measure of effect | reproduction number, infectious period length, overall infectiousness |
| Reporting of uncertainty | no |
| Sensitivity analysis | none |
| Subgroup assessment | none |

**Wang et al. 2020d**

| | |
|---|---|
| **Study information** | |
| Author | Wang K, Zhao S, Li H, Song Y, Wang L, Wang MH, Peng Z, Li H, He D |

– continued on next page –

| | |
|---|---|
| Title | Real-Time Estimation of the Reproduction Number of the Novel Coronavirus Disease (COVID-19) in China in 2020 Based on Incidence Data |
| Year | 2020 |
| Journal | Annals of Translational Medicine |
| DOI | 10.21037/atm-20-1944 |
| Date received | 2020-02-26 |
| Date accepted | 2020-04-21 |
| **Study setting** | |
| Number of populations included | multiple |
| Level of populations included | both national and subnational (China, Hubei province and Wuhan) |
| Geographic areas covered | Asia |
| Number of countries covered | single country (China) |
| Study period | start and end date span first epidemic wave |
| **Outcome** | |
| Raw outcome | epidemiological individual-level outcome (individual cases) |
| Time resolution of raw outcome | daily |
| Computed outcome | epidemiological parameter (reproduction number) |
| Method to obtain the computed outcome | statistical estimation of reproduction number (Wallinga and Teunis) |
| Data source | data from (sub)national authorities |
| Data availability | data not accessible |
| **Intervention** | |
| Terminology for interventions | – not applicable – |
| Terminology for the specific type of non-pharmaceutical interventions | lockdown |
| Exposure types | one combination of interventions (Wuhan City Lockdown) |

– continued on next page –

| | |
|---|---|
| Types of single interventions | – not applicable – |
| Coding of interventions | not necessary |
| Source of intervention data | – could not be evaluated – |
| Availability of data on exposure | raw data documented in the manuscript |
| **Methodological approach** | |
| Empirical approach | descriptive |
| Use of exposure variation | only variation over time for multiple populations |
| Method | description of change over time (comparison of time periods) |
| Code availability | none |
| **Effect assessment** | |
| Reporting of intervention effect | comparison of outcome values |
| Measure of effect | reproduction number |
| Reporting of uncertainty | yes |
| Sensitivity analysis | different or modified outcome used (varying levels of underreporting accounted for when computing effective reproduction number) |
| Subgroup assessment | none |

**Wang et al. 2020e**

| | |
|---|---|
| **Study information** | |
| Author | Wang X, Ren R, Kattan MW, Jehi L, Cheng Z, Fang K |
| Title | Public Health Interventions' Effect on Hospital Use in Patients with COVID-19: Comparative Study |
| Year | 2020 |
| Journal | JMIR Public Health and Surveillance |

– continued on next page –

| | |
|---|---|
| DOI | 10.2196/25174 |
| Date received | 2020-11-12 |
| Date accepted | 2020-12-06 |
| **Study setting** | |
| Number of populations included | multiple |
| Level of populations included | subnational (New York, Ohio, Hubei) |
| Geographic areas covered | Asia, North America |
| Number of countries covered | multiple countries |
| Study period | start and end date span first epidemic wave |
| **Outcome** | |
| Raw outcome | epidemiological population-level outcome (cases, deaths, recovered cases) |
| Time resolution of raw outcome | daily |
| Computed outcome | none |
| Method to obtain the computed outcome | – not applicable – |
| Data source | data from (sub)national authorities (British Columbia and New Zealand), data from public media outlets (COVID Tracking Project by The Atlantic: US) |
| Data availability | data access via source |
| **Intervention** | |
| Terminology for interventions | interventions |
| Terminology for the specific type of non-pharmaceutical interventions | public health |
| Exposure types | one combination of interventions (Wuhan City Lockdown) |
| Types of single interventions | – not applicable – |
| Coding of interventions | not necessary |

– continued on next page –

| | |
|---|---|
| Source of intervention data | – could not be evaluated – |
| Availability of data on exposure | raw data documented in the manuscript |
| **Methodological approach** | |
| Empirical approach | parametric |
| Use of exposure variation | only variation over time for multiple populations |
| Method | mechanistic model (compartmental single-population transmission model) |
| Latent variable functionally linked to intervention | transmission rate |
| Code availability | none |
| **Effect assessment** | |
| Reporting of intervention effect | quantification of change in outcome values |
| Measure of effect | transmission rate, reproduction number |
| Reporting of uncertainty | yes |
| Sensitivity analysis | none |
| Subgroup assessment | none |

**Wells et al. 2020**

| | |
|---|---|
| **Study information** | |
| Author | Wells CR, Sah P, Moghadas SM, Pandey A, Shoukat A, Wang Y, Wang Z, Meyers LA, Singer BH, Galvani AP |
| Title | Impact of International Travel and Border Control Measures on the Global Spread of the Novel 2019 Coronavirus Outbreak |
| Year | 2020 |
| Journal | Proceedings of the National Academy of Sciences |
| DOI | 10.1073/pnas.2002616117 |

– continued on next page –

| | |
|---|---|
| Date received | 2020-02-12 (sent for review) |
| Date accepted | 2020-03-13 |
| **Study setting** | |
| Number of populations included | multiple |
| Level of populations included | national (China and 21 countries with imported cases) |
| Geographic areas covered | Asia |
| Number of countries covered | single country (China) |
| Study period | other (start date in December 2019 to estimate early exportation risk) |
| **Outcome** | |
| Raw outcome | epidemiological population-level outcome (cases, exported cases) |
| Time resolution of raw outcome | daily |
| Computed outcome | none |
| Method to obtain the computed outcome | – not applicable – |
| Data source | data collected by authors (from various sources) |
| Data availability | data made available by the authors |
| **Intervention** | |
| Terminology for interventions | measures |
| Terminology for the specific type of non-pharmaceutical interventions | travel and border control measures |
| Exposure types | one single intervention (travel restrictions) |
| Types of single interventions | international travel restrictions |
| Coding of interventions | not necessary |
| Source of intervention data | – could not be evaluated – |
| Availability of data on exposure | raw data documented in the manuscript |
| **Methodological approach** | |

– continued on next page –

| | |
|---|---|
| Empirical approach | counterfactual |
| Use of exposure variation | only variation over time for a single population |
| Method | other (extrapolate probability and risk of travels outside of China to predict exported cases in absence of intervention) |
| Code availability | publicly available |
| **Effect assessment** | |
| Reporting of intervention effect | quantification of change in outcome values |
| Measure of effect | exported cases |
| Reporting of uncertainty | yes |
| Sensitivity analysis | none |
| Subgroup assessment | none |

**Wibbens et al. 2020**

| | |
|---|---|
| **Study information** | |
| Author | Wibbens PD, Koo WW, McGahan AM |
| Title | Which COVID Policies Are Most Effective? A Bayesian Analysis of COVID-19 by Jurisdiction |
| Year | 2020 |
| Journal | PLOS ONE |
| DOI | 10.1371/journal.pone.0244177 |
| Date received | 2020-05-19 |
| Date accepted | 2020-12-04 |
| **Study setting** | |
| Number of populations included | multiple |

– continued on next page –

| | |
|---|---|
| Level of populations included | both national and subnational (40 countries around the globe and US states) |
| Geographic areas covered | Europe, Middle East and Africa, Asia, Oceania, Central and South America, North America |
| Number of countries covered | multiple countries |
| Study period | start and end date span first epidemic wave |
| **Outcome** | |
| Raw outcome | epidemiological population-level outcome (cases, deaths, excess deaths) |
| Time resolution of raw outcome | daily |
| Computed outcome | none |
| Method to obtain the computed outcome | – not applicable – |
| Data source | data from publicly available cross-country selections (JHU) |
| Data availability | data made available by the authors |
| **Intervention** | |
| Terminology for interventions | measures |
| Terminology for the specific type of non-pharmaceutical interventions | control |
| Exposure types | multiple separate interventions (school closure, bans of large gatherings, bans of small gatherings, workplace closure, stay-at-home order, international travel restrictions, internal movement restrictions, public transport closure) |
| Types of single interventions | school closure, bans of large gatherings, bans of small gatherings, workplace closure, stay-at-home order, international travel restrictions, internal movement restrictions, public transport closure |
| Coding of interventions | necessary |
| Source of intervention data | use of externally coded data (OxCGRT) |

– continued on next page –

| | |
|---|---|
| Availability of data on exposure | coded data made available by the authors |
| **Methodological approach** | |
| Empirical approach | parametric |
| Use of exposure variation | both variation over time and between populations |
| Method | mechanistic model (semi-mechanistic Bayesian transmission model) |
| Latent variable functionally linked to intervention | other (growth rate of new cases) |
| Code availability | publicly available |
| **Effect assessment** | |
| Reporting of intervention effect | quantification of change in outcome values |
| Measure of effect | growth rate: cases |
| Reporting of uncertainty | yes |
| Sensitivity analysis | same analysis with (sub)population(s) excluded, model specification varied |
| Subgroup assessment | epidemiological indicators (pre-intervention epidemic trend) |

**Wieland 2020 (Analysis 1)**

| | |
|---|---|
| **Study information** | |
| Author | Wieland T |
| Title | A Phenomenological Approach to Assessing the Effectiveness of COVID-19 Related Nonpharmaceutical Interventions in Germany |
| Year | 2020 |
| Journal | Safety Science |
| DOI | 10.1016/j.ssci.2020.104924 |

– continued on next page –

| | |
|---|---|
| Date received | – could not be evaluated – |
| Date accepted | 2020-07-21 (published) |
| **Study setting** | |
| Number of populations included | single |
| Level of populations included | national (Germany) |
| Geographic areas covered | Europe |
| Number of countries covered | single country (Germany) |
| Study period | start and end date span first epidemic wave |
| **Outcome** | |
| Raw outcome | epidemiological individual-level outcome (individual cases) |
| Time resolution of raw outcome | daily |
| Computed outcome | measure of epidemic trend (growth rate: infections), epidemiological parameter (reproduction number), change points (change points: infections) |
| Method to obtain the computed outcome | back projection, change point analysis, statistical estimation of reproduction number (AD Heiden and Hamouda) |
| Data source | data from (sub)national authorities |
| Data availability | data not accessible |
| **Intervention** | |
| Terminology for interventions | interventions |
| Terminology for the specific type of non-pharmaceutical interventions | non-pharmaceutical |
| Exposure types | multiple combinations of interventions (1. cancellation of mass events after March 8, 2. closure of schools and child day care facilities between March 16 and 18, and 3. a contact ban, bans of gatherings and closures of "nonessential" services from March 23) |
| Types of single interventions | – not applicable – |
| Coding of interventions | not necessary |

– continued on next page –

| | |
|---|---|
| Source of intervention data | – could not be evaluated – |
| Availability of data on exposure | raw data documented in the manuscript |
| **Methodological approach** | |
| Empirical approach | descriptive |
| Use of exposure variation | only variation over time for a single population |
| Method | comparison of change points with intervention dates |
| Code availability | none |
| **Effect assessment** | |
| Reporting of intervention effect | comparison of outcome values |
| Measure of effect | change points: infections, growth rate: infections, reproduction number |
| Reporting of uncertainty | yes |
| Sensitivity analysis | none |
| Subgroup assessment | none |

**Wieland 2020 (Analysis 2)**

| | |
|---|---|
| **Study information** | |
| Author | Wieland T |
| Title | A Phenomenological Approach to Assessing the Effectiveness of COVID-19 Related Nonpharmaceutical Interventions in Germany |
| Year | 2020 |
| Journal | Safety Science |
| DOI | 10.1016/j.ssci.2020.104924 |
| Date received | – could not be evaluated – |
| Date accepted | 2020-07-21 (published) |

– continued on next page –

| | |
|---|---|
| **Study setting** | |
| Number of populations included | single |
| Level of populations included | national (Germany) |
| Geographic areas covered | Europe |
| Number of countries covered | single country (Germany) |
| Study period | start and end date span first epidemic wave |
| **Outcome** | |
| Raw outcome | epidemiological individual-level outcome (individual cases) |
| Time resolution of raw outcome | daily |
| Computed outcome | measure of epidemic trend (growth rate: infections), epidemiological parameter (reproduction number) |
| Method to obtain the computed outcome | back projection, change point analysis, statistical estimation of reproduction number (AD Heiden and Hamouda) |
| Data source | data from (sub)national authorities |
| Data availability | data not accessible |
| **Intervention** | |
| Terminology for interventions | interventions |
| Terminology for the specific type of non-pharmaceutical interventions | non-pharmaceutical |
| Exposure types | multiple combinations of interventions (estimated change points from time series that correspond with the implementation dates of NPIs to some extent of schools and child day care facilities between March 16 and 18, and 3. a contact ban, bans of gatherings and closures of “nonessential” services from March 23) |
| Types of single interventions | – not applicable – |
| Coding of interventions | not necessary |
| Source of intervention data | – could not be evaluated – |

– continued on next page –

**Wieland 2020 (Analysis 2)** – continued from previous page

| | |
|---|---|
| Availability of data on exposure | raw data documented in the manuscript |
| **Methodological approach** | |
| Empirical approach | descriptive |
| Use of exposure variation | only variation over time for a single population |
| Method | description of change over time (comparison of time periods) |
| Code availability | none |
| **Effect assessment** | |
| Reporting of intervention effect | comparison of outcome values |
| Measure of effect | infections, growth rate: infections, reproduction number |
| Reporting of uncertainty | yes |
| Sensitivity analysis | none |
| Subgroup assessment | none |

**Wielechowski et al. 2020 (Analysis 1)**

| | |
|---|---|
| **Study information** | |
| Author | Wielechowski M, Czech K, Grzeda L |
| Title | Decline in Mobility: Public Transport in Poland in the Time of the COVID-19 Pandemic |
| Year | 2020 |
| Journal | Economies |
| DOI | 10.3390/economies8040078 |
| Date received | 2020-08-13 |
| Date accepted | 2020-09-15 |

– continued on next page –

| | |
|---|---|
| **Study setting** | |
| Number of populations included | multiple |
| Level of populations included | both national and subnational (Poland and its 16 vovoidships) |
| Geographic areas covered | Europe |
| Number of countries covered | single country (Poland) |
| Study period | start and end date span first epidemic wave: behavioral |
| **Outcome** | |
| Raw outcome | behavioral outcome (mobility: time spent in public transport stations) |
| Time resolution of raw outcome | daily |
| Computed outcome | none |
| Method to obtain the computed outcome | – not applicable – |
| Data source | mobility data from corporate organizations (Google) |
| Data availability | data access via source |
| **Intervention** | |
| Terminology for interventions | policies |
| Terminology for the specific type of non-pharmaceutical interventions | government |
| Exposure types | stringency index (OxCGRT Stringency Index) |
| Types of single interventions | – not applicable – |
| Coding of interventions | necessary |
| Source of intervention data | use of externally coded data (OxCGRT) |
| Availability of data on exposure | access to externally coded data via source |
| **Methodological approach** | |
| Empirical approach | parametric |
| Use of exposure variation | only variation over time for multiple populations |

– continued on next page –

| | |
|---|---|
| Method | non-mechanistic model (correlation) |
| Code availability | none |
| **Effect assessment** | |
| Reporting of intervention effect | quantification of change in outcome values |
| Measure of effect | mobility: time spent in public transport stations |
| Reporting of uncertainty | yes |
| Sensitivity analysis | none |
| Subgroup assessment | none |

**Wielechowski et al. 2020 (Analysis 2, mobility as exposure)**

| | |
|---|---|
| **Study information** | |
| Author | Wielechowski M, Czech K, Grzeda L |
| Title | Decline in Mobility: Public Transport in Poland in the Time of the COVID-19 Pandemic |
| Year | 2020 |
| Journal | Economies |
| DOI | 10.3390/economies8040078 |
| Date received | 13.08.2020 |
| Date accepted | 15.09.2020 |
| **Study setting** | |
| Number of populations included | multiple |
| Level of populations included | both national and subnational (Poland and its 16 vovoidships) |
| Geographic areas covered | Europe |
| Number of countries covered | single country (Poland) |

– continued on next page –

| | |
|---|---|
| Study period | start and end date span first epidemic wave |
| **Outcome** | |
| Raw outcome | epidemiological population-level outcome (cases) |
| Time resolution of raw outcome | daily |
| Computed outcome | none |
| Method to obtain the computed outcome | – not applicable – |
| Data source | data from (sub)national authorities |
| Data availability | data access via source |
| **Intervention** | |
| Terminology for interventions | policies |
| Terminology for the specific type of non-pharmaceutical interventions | government |
| Exposure types | mobility (time spent in public transport stations) |
| Types of single interventions | – not applicable – |
| Coding of interventions | – not applicable – |
| Source of intervention data | mobility data by corporate organizations (Google) |
| Availability of data on exposure | data access via source |
| **Methodological approach** | |
| Empirical approach | descriptive |
| Use of exposure variation | only variation over time for multiple populations |
| Method | non-mechanistic model (correlation) |
| Code availability | none |
| **Effect assessment** | |
| Reporting of intervention effect | quantification of change in outcome values |
| Measure of effect | cases |

– continued on next page –

| | |
|---|---|
| Reporting of uncertainty | yes |
| Sensitivity analysis | none |
| Subgroup assessment | none |

| **Wong et al. 2020a** | |
|---|---|
| **Study information** | |
| Author | Wong CK, Wong JY, Tang EH, Au CH, Lau KT, Wai AK |
| Title | Impact of National Containment Measures on Decelerating the Increase in Daily New Cases of COVID-19 in 54 Countries and 4 Epicenters of the Pandemic: Comparative Observational Study |
| Year | 2020 |
| Journal | Journal of Medical Internet Research |
| DOI | 10.2196/19904 |
| Date received | 2020-05-06 |
| Date accepted | 2020-07-13 |
| **Study setting** | |
| Number of populations included | multiple |
| Level of populations included | both national and subnational (54 countries, Wuhan, New York State, Lombardy, Madrid) |
| Geographic areas covered | Europe, Middle East and Africa, Asia, Oceania, Central and South America, North America |
| Number of countries covered | multiple countries |
| Study period | same end date for several populations with diverse epidemic trajectories |
| **Outcome** | |
| Raw outcome | epidemiological population-level outcome (cases) |

– continued on next page –

| | |
|---|---|
| Time resolution of raw outcome | daily |
| Computed outcome | none |
| Method to obtain the computed outcome | – not applicable – |
| Data source | data from publicly available cross-country selections (OurWorldInData) |
| Data availability | data access via source |
| **Intervention** | |
| Terminology for interventions | measures |
| Terminology for the specific type of non-pharmaceutical interventions | containment |
| Exposure types | multiple separate interventions (stay-at-home order (limited outdoor movement), curfew (stay-at-home order during specific time periods), lockdown (restriction of population mobility)) |
| Types of single interventions | stay-at-home order, internal movement restrictions |
| Coding of interventions | necessary |
| Source of intervention data | coding done by authors |
| Availability of data on exposure | coded data made available by the authors |
| **Methodological approach** | |
| Empirical approach | descriptive |
| Use of exposure variation | only variation over time for multiple populations |
| Method | description of change over time (description of time course) |
| Code availability | none |
| **Effect assessment** | |
| Reporting of intervention effect | comparison of outcome values |
| Measure of effect | cases |
| Reporting of uncertainty | yes |

– continued on next page –

| | |
|---|---|
| Sensitivity analysis | none |
| Subgroup assessment | geographic areas (regions), socioeconomic indicators (income), epidemiological indicators (pre-intervention epidemic trend) |

| **Wong et al. 2020b (Analysis 1)** | |
|---|---|
| **Study information** | |
| Author | Wong GN, Weiner ZJ, Tkachenko AV, Elbanna A, Maslov S, Goldenfeld N |
| Title | Modeling COVID-19 Dynamics in Illinois under Nonpharmaceutical Interventions |
| Year | 2020 |
| Journal | Physical Review X |
| DOI | 10.1103/physrevx.10.041033 |
| Date received | 2020-06-15 |
| Date accepted | 2020-10-22 |
| **Study setting** | |
| Number of populations included | single |
| Level of populations included | subnational (Illinois) |
| Geographic areas covered | North America |
| Number of countries covered | single country (US) |
| Study period | end date at peak of wave |
| **Outcome** | |
| Raw outcome | epidemiological population-level outcome (hospitalizations, ICU hospitalizations, deaths) |
| Time resolution of raw outcome | daily |

– continued on next page –

| | |
|---|---|
| Computed outcome | none |
| Method to obtain the computed outcome | – not applicable – |
| Data source | data from (sub)national authorities |
| Data availability | data made available by the authors |
| **Intervention** | |
| Terminology for interventions | interventions |
| Terminology for the specific type of non-pharmaceutical interventions | non-pharmaceutical |
| Exposure types | all interventions together |
| Types of single interventions | – not applicable – |
| Coding of interventions | – not applicable – |
| Source of intervention data | – not applicable – |
| Availability of data on exposure | – not applicable – |
| **Methodological approach** | |
| Empirical approach | parametric |
| Use of exposure variation | only variation over time for a single population |
| Method | mechanistic model (compartmental single-population transmission model) |
| Latent variable functionally linked to intervention | reproduction number |
| Code availability | publicly available |
| **Effect assessment** | |
| Reporting of intervention effect | quantification of change in outcome values |
| Measure of effect | reproduction number |
| Reporting of uncertainty | yes |

– continued on next page –

| | |
|---|---|
| Sensitivity analysis | none |
| Subgroup assessment | geographic areas (regions) |

| **Wong et al. 2020b (Analysis 2, mobility as exposure)** | |
|---|---|
| **Study information** | |
| Author | Wong GN, Weiner ZJ, Tkachenko AV, Elbanna A, Maslov S, Goldenfeld N |
| Title | Modeling COVID-19 Dynamics in Illinois under Nonpharmaceutical Interventions |
| Year | 2020 |
| Journal | Physical Review X |
| DOI | 10.1103/physrevx.10.041033 |
| Date received | 15.06.2020 |
| Date accepted | 22.10.2020 |
| **Study setting** | |
| Number of populations included | single |
| Level of populations included | subnational (Illinois) |
| Geographic areas covered | North America |
| Number of countries covered | single country (US) |
| Study period | end date at peak of wave |
| **Outcome** | |
| Raw outcome | epidemiological population-level outcome (hospitalizations, ICU hospitalizations, deaths) |
| Time resolution of raw outcome | daily |
| Computed outcome | other (mitigation factor) |

– continued on next page –

| | |
|---|---|
| Method to obtain the computed outcome | compartmental transmission model (single-population) |
| Data source | data from (sub)national authorities |
| Data availability | data made available by the authors |
| **Intervention** | |
| Terminology for interventions | interventions |
| Terminology for the specific type of non-pharmaceutical interventions | non-pharmaceutical |
| Exposure types | mobility (activity, distance, non-essential visits) |
| Types of single interventions | – not applicable – |
| Coding of interventions | – not applicable – |
| Source of intervention data | mobility data by corporate organizations (Google, Unacast) |
| Availability of data on exposure | data access via source |
| **Methodological approach** | |
| Empirical approach | descriptive |
| Use of exposure variation | only variation over time for a single population |
| Method | description of change over time (description of time course) |
| Code availability | publicly available |
| **Effect assessment** | |
| Reporting of intervention effect | qualitative statement |
| Measure of effect | mitigation factor |
| Reporting of uncertainty | – not applicable – |
| Sensitivity analysis | none |
| Subgroup assessment | none |

| Wong et al. 2020c (Analysis 1) | |
|---|---|
| **Study information** | |
| Author | Wong MC, Ng RW, Chong KC, Lai CK, Huang J, Chen Z, Boon SS, Chan PK |
| Title | Stringent Containment Measures without Complete City Lockdown to Achieve Low Incidence and Mortality across Two Waves of COVID-19 in Hong Kong |
| Year | 2020 |
| Journal | BMJ Global Health |
| DOI | 10.1136/bmjgh-2020-003573 |
| Date received | 2020-07-29 |
| Date accepted | 2020-09-07 |
| **Study setting** | |
| Number of populations included | single |
| Level of populations included | national (Hong Kong) |
| Geographic areas covered | Asia |
| Number of countries covered | single country (Hong Kong) |
| Study period | includes second wave (two waves) |
| **Outcome** | |
| Raw outcome | epidemiological individual-level outcome (individual cases) |
| Time resolution of raw outcome | daily |
| Computed outcome | epidemiological parameter (reproduction number) |
| Method to obtain the computed outcome | statistical estimation of reproduction number (Thompson et al.) |
| Data source | data from (sub)national authorities |
| Data availability | data not accessible |

– continued on next page –

| | |
|---|---|
| **Intervention** | |
| Terminology for interventions | measures |
| Terminology for the specific type of non-pharmaceutical interventions | containment |
| Exposure types | all interventions together (timeline of measures) |
| Types of single interventions | – not applicable – |
| Coding of interventions | – not applicable – |
| Source of intervention data | – not applicable – |
| Availability of data on exposure | – not applicable – |
| **Methodological approach** | |
| Empirical approach | descriptive |
| Use of exposure variation | only variation over time for a single population |
| Method | description of change over time (description of time course) |
| Code availability | none |
| **Effect assessment** | |
| Reporting of intervention effect | comparison of outcome values |
| Measure of effect | reproduction number |
| Reporting of uncertainty | yes |
| Sensitivity analysis | none |
| Subgroup assessment | none |

**Wong et al. 2020c (Analysis 2)**

| | |
|---|---|
| **Study information** | |
| Author | Wong MC, Ng RW, Chong KC, Lai CK, Huang J, Chen Z, Boon SS, Chan PK |

– continued on next page –

| | |
|---|---|
| Title | Stringent Containment Measures without Complete City Lockdown to Achieve Low Incidence and Mortality across Two Waves of COVID-19 in Hong Kong |
| Year | 2020 |
| Journal | BMJ Global Health |
| DOI | 10.1136/bmjgh-2020-003573 |
| Date received | 2020-07-29 |
| Date accepted | 2020-09-07 |
| **Study setting** | |
| Number of populations included | single |
| Level of populations included | national (Hong Kong) |
| Geographic areas covered | Asia |
| Number of countries covered | single country (Hong Kong) |
| Study period | includes second wave (two waves) |
| **Outcome** | |
| Raw outcome | epidemiological population-level outcome (cases) |
| Time resolution of raw outcome | daily |
| Computed outcome | none |
| Method to obtain the computed outcome | – not applicable – |
| Data source | data from (sub)national authorities |
| Data availability | data access via source |
| **Intervention** | |
| Terminology for interventions | measures |
| Terminology for the specific type of non-pharmaceutical interventions | containment |

– continued on next page –

**Wong et al. 2020c (Analysis 2)** – continued from previous page

| | |
|---|---|
| Exposure types | all interventions together (timeline of measures) |
| Types of single interventions | – not applicable – |
| Coding of interventions | – not applicable – |
| Source of intervention data | – not applicable – |
| Availability of data on exposure | – not applicable – |
| **Methodological approach** | |
| Empirical approach | counterfactual |
| Use of exposure variation | only variation over time for a single population |
| Method | other (extrapolation of cases by forward application of renewal equation used by Thompson et al.) |
| Code availability | none |
| **Effect assessment** | |
| Reporting of intervention effect | qualitative statement |
| Measure of effect | cases |
| Reporting of uncertainty | – not applicable – |
| Sensitivity analysis | none |
| Subgroup assessment | none |

**Wu et al. 2020 (Analysis 1)**

| | |
|---|---|
| **Study information** | |
| Author | Wu J, Tang B, Bragazzi NL, Nah K, McCarthy Z |
| Title | Quantifying the Role of Social Distancing, Personal Protection and Case Detection in Mitigating COVID-19 Outbreak in Ontario, Canada |

– continued on next page –

| | |
|---|---|
| Year | 2020 |
| Journal | Journal of Mathematics in Industry |
| DOI | 10.1186/s13362-020-00083-3 |
| Date received | 2020-04-24 |
| Date accepted | 2020-05-12 |
| **Study setting** | |
| Number of populations included | single |
| Level of populations included | subnational (Ontario) |
| Geographic areas covered | North America |
| Number of countries covered | single country (Canada) |
| Study period | end date in growth phase of wave |
| **Outcome** | |
| Raw outcome | epidemiological population-level outcome (cases) |
| Time resolution of raw outcome | daily |
| Computed outcome | epidemiological parameter (reproduction number) |
| Method to obtain the computed outcome | compartmental transmission model (single-population) |
| Data source | data from (sub)national authorities |
| Data availability | data access via source |
| **Intervention** | |
| Terminology for interventions | interventions |
| Terminology for the specific type of non-pharmaceutical interventions | public health |
| Exposure types | all interventions together |
| Types of single interventions | – not applicable – |
| Coding of interventions | – not applicable – |

– continued on next page –

| | |
|---|---|
| Source of intervention data | – not applicable – |
| Availability of data on exposure | – not applicable – |
| **Methodological approach** | |
| Empirical approach | descriptive |
| Use of exposure variation | only variation over time for a single population |
| Method | description of change over time (description of time course) |
| Code availability | none |
| **Effect assessment** | |
| Reporting of intervention effect | qualitative statement |
| Measure of effect | reproduction number |
| Reporting of uncertainty | – not applicable – |
| Sensitivity analysis | none |
| Subgroup assessment | none |

**Wu et al. 2020 (Analysis 2)**

| | |
|---|---|
| **Study information** | |
| Author | Wu J, Tang B, Bragazzi NL, Nah K, McCarthy Z |
| Title | Quantifying the Role of Social Distancing, Personal Protection and Case Detection in Mitigating COVID-19 Outbreak in Ontario, Canada |
| Year | 2020 |
| Journal | Journal of Mathematics in Industry |
| DOI | 10.1186/s13362-020-00083-3 |
| Date received | 2020-04-24 |
| Date accepted | 2020-05-12 |

– continued on next page –

| | |
|---|---|
| **Study setting** | |
| Number of populations included | single |
| Level of populations included | subnational (Ontario) |
| Geographic areas covered | North America |
| Number of countries covered | single country (Canada) |
| Study period | end date in growth phase of wave |
| **Outcome** | |
| Raw outcome | epidemiological population-level outcome (cases) |
| Time resolution of raw outcome | daily |
| Computed outcome | none |
| Method to obtain the computed outcome | – not applicable – |
| Data source | data from (sub)national authorities |
| Data availability | data access via source |
| **Intervention** | |
| Terminology for interventions | interventions |
| Terminology for the specific type of non-pharmaceutical interventions | public health |
| Exposure types | one single intervention (workplace closure on March 24) |
| Types of single interventions | workplace closure |
| Coding of interventions | not necessary |
| Source of intervention data | – could not be evaluated – |
| Availability of data on exposure | raw data documented in the manuscript |
| **Methodological approach** | |
| Empirical approach | parametric |
| Use of exposure variation | only variation over time for a single population |

– continued on next page –

| | |
|---|---|
| Method | mechanistic model (compartmental single-population transmission model) |
| Latent variable functionally linked to intervention | contact rate, quarantine rate |
| Code availability | none |
| **Effect assessment** | |
| Reporting of intervention effect | quantification of change in outcome values |
| Measure of effect | contact rate, quarantine rate, reproduction number |
| Reporting of uncertainty | no |
| Sensitivity analysis | none |
| Subgroup assessment | none |

**Xia et al. 2020**

| | |
|---|---|
| **Study information** | |
| Author | Xia F, Xiao AY, Liu P, Cheke RA, Li X, And |
| Title | Differences in How Interventions Coupled with Effective Reproduction Numbers Account for Marked Variations in COVID-19 Epidemic Outcomes |
| Year | 2020 |
| Journal | Mathematical Biosciences and Engineering |
| DOI | 10.3934/mbe.2020274 |
| Date received | 2020-06-05 |
| Date accepted | 2020-07-21 |
| **Study setting** | |
| Number of populations included | multiple |
| Level of populations included | national (China, South Korea, Japan, Spain) |

– continued on next page –

| | |
|---|---|
| Geographic areas covered | Asia, Europe |
| Number of countries covered | multiple countries |
| Study period | start and end date span first epidemic wave |
| **Outcome** | |
| Raw outcome | epidemiological population-level outcome (cases) |
| Time resolution of raw outcome | daily |
| Computed outcome | epidemiological parameter (reproduction number) |
| Method to obtain the computed outcome | compartmental transmission model (single-population) |
| Data source | data from (sub)national authorities (health authority in each country), data from publicly available cross-country selections (WHO: all countries) |
| Data availability | data access via source |
| **Intervention** | |
| Terminology for interventions | strategies |
| Terminology for the specific type of non-pharmaceutical interventions | containment |
| Exposure types | all interventions together |
| Types of single interventions | – not applicable – |
| Coding of interventions | – not applicable – |
| Source of intervention data | – not applicable – |
| Availability of data on exposure | – not applicable – |
| **Methodological approach** | |
| Empirical approach | descriptive |
| Use of exposure variation | only variation over time for multiple populations |
| Method | description of change over time (description of time course) |
| Code availability | none |

– continued on next page –

| | |
|---|---|
| **Effect assessment** | |
| Reporting of intervention effect | qualitative statement |
| Measure of effect | reproduction number |
| Reporting of uncertainty | – not applicable – |
| Sensitivity analysis | none |
| Subgroup assessment | none |

**Xiao et al. 2020**

| | |
|---|---|
| **Study information** | |
| Author | Xiao Y, Tang B, Wu J, Cheke RA, Tang S |
| Title | Linking Key Intervention Timing to Rapid Decline of the COVID-19 Effective Reproductive Number to Quantify Lessons from Mainland China |
| Year | 2020 |
| Journal | International Journal of Infectious Diseases |
| DOI | 10.1016/j.ijid.2020.06.030 |
| Date received | 2020-05-08 |
| Date accepted | 2020-06-04 |
| **Study setting** | |
| Number of populations included | single |
| Level of populations included | national (China) |
| Geographic areas covered | Asia |
| Number of countries covered | single country (China) |
| Study period | start and end date span first epidemic wave |
| **Outcome** | |

– continued on next page –

| | |
|---|---|
| Raw outcome | epidemiological population-level outcome (cases) |
| Time resolution of raw outcome | daily |
| Computed outcome | epidemiological parameter (reproduction number) |
| Method to obtain the computed outcome | statistical estimation of reproduction number (– could not be evaluated –) |
| Data source | – could not be evaluated – |
| Data availability | data not accessible |
| **Intervention** | |
| Terminology for interventions | measures |
| Terminology for the specific type of non-pharmaceutical interventions | intervention |
| Exposure types | all interventions together (timeline of measures) |
| Types of single interventions | – not applicable – |
| Coding of interventions | – not applicable – |
| Source of intervention data | – not applicable – |
| Availability of data on exposure | – not applicable – |
| **Methodological approach** | |
| Empirical approach | descriptive |
| Use of exposure variation | only variation over time for a single population |
| Method | description of change over time (description of time course) |
| Code availability | none |
| **Effect assessment** | |
| Reporting of intervention effect | qualitative statement |
| Measure of effect | reproduction number |
| Reporting of uncertainty | – not applicable – |
| Sensitivity analysis | none |

– continued on next page –

| | |
|---|---|
| Subgroup assessment | none |

**Xu et al. 2020 (Analysis 1)**

| | |
|---|---|
| **Study information** | |
| Author | Xu P, Dredze M, Broniatowski DA |
| Title | The Twitter Social Mobility Index: Measuring Social Distancing Practices with Geolocated Tweets |
| Year | 2020 |
| Journal | Journal of Medical Internet Research |
| DOI | 10.2196/21499 |
| Date received | 2020-06-16 |
| Date accepted | 2020-10-11 |
| **Study setting** | |
| Number of populations included | multiple |
| Level of populations included | both national and subnational (US) |
| Geographic areas covered | North America |
| Number of countries covered | single country (US) |
| Study period | start and end date span first epidemic wave: behavioral |
| **Outcome** | |
| Raw outcome | behavioral outcome (mobility: area and frequency of travel) |
| Time resolution of raw outcome | daily |
| Computed outcome | none |

– continued on next page –

| | |
|---|---|
| Method to obtain the computed outcome | – not applicable – |
| Data source | mobility data from corporate organizations (Twitter) |
| Data availability | data not accessible |
| **Intervention** | |
| Terminology for interventions | policies |
| Terminology for the specific type of non-pharmaceutical interventions | social distancing |
| Exposure types | one combination of interventions (social distancing measures, "slow the spread guidelines" by White house) |
| Types of single interventions | – not applicable – |
| Coding of interventions | not necessary |
| Source of intervention data | government or news websites |
| Availability of data on exposure | raw data documented in the manuscript |
| **Methodological approach** | |
| Empirical approach | descriptive |
| Use of exposure variation | only variation over time for multiple populations |
| Method | description of change over time (comparison of time periods) |
| Code availability | none |
| **Effect assessment** | |
| Reporting of intervention effect | quantification of change in outcome values |
| Measure of effect | mobility: area and frequency of travel |
| Reporting of uncertainty | no |
| Sensitivity analysis | none |
| Subgroup assessment | none |

| **Xu et al. 2020 (Analysis 2)** | |
|---|---|
| **Study information** | |
| Author | Xu P, Dredze M, Broniatowski DA |
| Title | The Twitter Social Mobility Index: Measuring Social Distancing Practices with Geolocated Tweets |
| Year | 2020 |
| Journal | Journal of Medical Internet Research |
| DOI | 10.2196/21499 |
| Date received | 2020-06-16 |
| Date accepted | 2020-10-11 |
| **Study setting** | |
| Number of populations included | multiple |
| Level of populations included | subnational (US) |
| Geographic areas covered | North America |
| Number of countries covered | single country (US) |
| Study period | same end date for several populations with diverse epidemic trajectories |
| **Outcome** | |
| Raw outcome | epidemiological population-level outcome (cases) |
| Time resolution of raw outcome | daily |
| Computed outcome | none |
| Method to obtain the computed outcome | – not applicable – |
| Data source | data from publicly available cross-country selections (JHU) |
| Data availability | data made available by the authors |
| **Intervention** | |
| Terminology for interventions | policies |

– continued on next page –

| | |
|---|---|
| Terminology for the specific type of non-pharmaceutical interventions | social distancing |
| Exposure types | multiple separate interventions (state of emergency declaration, stay-at-home order, business closure, and venue closure (in various forms), other (date banned visitors to nursing homes, evictions frozen, day care closed)) |
| Types of single interventions | declaration of a state of emergency, stay-at-home order, workplace closure, venue closure, other |
| Coding of interventions | necessary |
| Source of intervention data | use of externally coded data (COVID-19 US State Policy Database openICPSR) |
| Availability of data on exposure | coded data made available by the authors |
| **Methodological approach** | |
| Empirical approach | parametric |
| Use of exposure variation | only variation between populations |
| Method | non-mechanistic model (correlation) |
| Code availability | none |
| **Effect assessment** | |
| Reporting of intervention effect | quantification of change in outcome values |
| Measure of effect | cases |
| Reporting of uncertainty | yes |
| Sensitivity analysis | none |
| Subgroup assessment | none |

**Xu et al. 2020 (Analysis 3, mobility as exposure)**

| | |
|---|---|
| **Study information** | |
| Author | Xu P, Dredze M, Broniatowski DA |

– continued on next page –

| | |
|---|---|
| Title | The Twitter Social Mobility Index: Measuring Social Distancing Practices with Geolocated Tweets |
| Year | 2020 |
| Journal | Journal of Medical Internet Research |
| DOI | 10.2196/21499 |
| Date received | 16.06.2020 |
| Date accepted | 11.10.2020 |
| **Study setting** | |
| Number of populations included | multiple |
| Level of populations included | both national and subnational (US) |
| Geographic areas covered | North America |
| Number of countries covered | single country (US) |
| Study period | same end date for several populations with diverse epidemic trajectories |
| **Outcome** | |
| Raw outcome | epidemiological population-level outcome (cases) |
| Time resolution of raw outcome | daily |
| Computed outcome | none |
| Method to obtain the computed outcome | – not applicable – |
| Data source | data from publicly available cross-country selections (JHU) |
| Data availability | data made available by the authors |
| **Intervention** | |
| Terminology for interventions | policies |
| Terminology for the specific type of non-pharmaceutical interventions | social distancing |
| Exposure types | mobility (area and frequency of travel) |

– continued on next page –

| **Xu et al. 2020 (Analysis 3, mobility as exposure)** – continued from previous page | |
|---|---|
| Types of single interventions | – not applicable – |
| Coding of interventions | – not applicable – |
| Source of intervention data | mobility data by corporate organizations (Twitter) |
| Availability of data on exposure | data not accessible |
| **Methodological approach** | |
| Empirical approach | descriptive |
| Use of exposure variation | both variation over time and between populations |
| Method | non-mechanistic model (correlation) |
| Code availability | none |
| **Effect assessment** | |
| Reporting of intervention effect | quantification of change in outcome values |
| Measure of effect | cases |
| Reporting of uncertainty | no |
| Sensitivity analysis | none |
| Subgroup assessment | none |

| **Yang et al. 2020** | |
|---|---|
| **Study information** | |
| Author | Yang Q, Yi C, Vajdi A, Cohnstaedt LW, Wu H, Guo X, Scoglio CM |
| Title | Short-Term Forecasts and Long-Term Mitigation Evaluations for the COVID-19 Epidemic in Hubei Province, China |
| Year | 2020 |
| Journal | Infectious Disease Modelling |
| DOI | 10.1016/j.idm.2020.08.001 |

– continued on next page –

| | |
|---|---|
| Date received | 2020-03-27 |
| Date accepted | 2020-07-05 |
| **Study setting** | |
| Number of populations included | single |
| Level of populations included | subnational (Hubei province) |
| Geographic areas covered | Asia |
| Number of countries covered | single country (China) |
| Study period | start and end date span first epidemic wave |
| **Outcome** | |
| Raw outcome | epidemiological population-level outcome (cases) |
| Time resolution of raw outcome | daily |
| Computed outcome | epidemiological parameter (reproduction number, transmission rate) |
| Method to obtain the computed outcome | compartmental transmission model (metapopulation) |
| Data source | data from (sub)national authorities |
| Data availability | data access via source |
| **Intervention** | |
| Terminology for interventions | interventions |
| Terminology for the specific type of non-pharmaceutical interventions | non-pharmaceutical |
| Exposure types | one combination of interventions (Wuhan City Lockdown) |
| Types of single interventions | – not applicable – |
| Coding of interventions | not necessary |
| Source of intervention data | another study (Du et al., Wu et al.) |
| Availability of data on exposure | raw data documented in the manuscript |
| **Methodological approach** | |

– continued on next page –

| | |
|---|---|
| Empirical approach | descriptive |
| Use of exposure variation | only variation over time for a single population |
| Method | description of change over time (comparison of time periods) |
| Code availability | none |
| **Effect assessment** | |
| Reporting of intervention effect | comparison of outcome values |
| Measure of effect | transmission rate, reproduction number |
| Reporting of uncertainty | no |
| Sensitivity analysis | none |
| Subgroup assessment | none |

| **Ye & Hu 2020** | |
|---|---|
| **Study information** | |
| Author | Ye L, Hu L |
| Title | Spatiotemporal Distribution and Trend of COVID-19 in the Yangtze River Delta Region of the People's Republic of China |
| Year | 2020 |
| Journal | Geospatial Health |
| DOI | 10.4081/gh.2020.889 |
| Date received | 2020-04-11 |
| Date accepted | 2020-04-30 |
| **Study setting** | |
| Number of populations included | multiple |
| Level of populations included | subnational (Yangtze river delta region (China) + 3 subregions) |

– continued on next page –

| | |
|---|---|
| Geographic areas covered | Asia |
| Number of countries covered | single country (China) |
| Study period | start and end date span first epidemic wave |
| **Outcome** | |
| Raw outcome | epidemiological population-level outcome (cases) |
| Time resolution of raw outcome | daily |
| Computed outcome | none |
| Method to obtain the computed outcome | – not applicable – |
| Data source | data from (sub)national authorities |
| Data availability | data access via source |
| **Intervention** | |
| Terminology for interventions | measures |
| Terminology for the specific type of non-pharmaceutical interventions | control |
| Exposure types | one combination of interventions (Wuhan City Lockdown) |
| Types of single interventions | – not applicable – |
| Coding of interventions | not necessary |
| Source of intervention data | – could not be evaluated – |
| Availability of data on exposure | raw data documented in the manuscript |
| **Methodological approach** | |
| Empirical approach | descriptive |
| Use of exposure variation | only variation over time for multiple populations |
| Method | description of change over time (description of time course) |
| Code availability | none |
| **Effect assessment** | |

– continued on next page –

| | |
|---|---|
| Reporting of intervention effect | qualitative statement |
| Measure of effect | cases |
| Reporting of uncertainty | – not applicable – |
| Sensitivity analysis | none |
| Subgroup assessment | none |

| **Yehya et al. 2020** | |
|---|---|
| **Study information** | |
| Author | Yehya N, Venkataramani A, Harhay MO |
| Title | Statewide Interventions and Coronavirus Disease 2019 Mortality in the United States: An Observational Study |
| Year | 2020 |
| Journal | Clinical Infectious Diseases |
| DOI | 10.1093/cid/ciaa923 |
| Date received | 2020-05-22 |
| Date accepted | 2020-06-26 |
| **Study setting** | |
| Number of populations included | multiple |
| Level of populations included | both national and subnational (37 US states) |
| Geographic areas covered | North America |
| Number of countries covered | single country (US) |
| Study period | same end date for several populations with diverse epidemic trajectories |
| **Outcome** | |
| Raw outcome | epidemiological population-level outcome (deaths) |

– continued on next page –

| | |
|---|---|
| Time resolution of raw outcome | daily |
| Computed outcome | summary statistic (mortality rate ratio: mortality rate relative to mortality rate on day 1 in a country) |
| Method to obtain the computed outcome | simple computation |
| Data source | data from publicly available cross-country selections (JHU) |
| Data availability | data access via source |
| **Intervention** | |
| Terminology for interventions | measures |
| Terminology for the specific type of non-pharmaceutical interventions | social distancing |
| Exposure types | multiple separate interventions (emergency declaration, school closure) |
| Types of single interventions | declaration of a state of emergency, school closure |
| Coding of interventions | necessary |
| Source of intervention data | coding done by authors |
| Availability of data on exposure | coded data not available |
| **Methodological approach** | |
| Empirical approach | parametric |
| Use of exposure variation | both variation over time and between populations |
| Method | non-mechanistic model (generalized linear model) |
| Code availability | none |
| **Effect assessment** | |
| Reporting of intervention effect | quantification of change in outcome values |
| Measure of effect | mortality rate ratio: mortality rate relative to mortality rate on day 1 in a country |
| Reporting of uncertainty | yes |

– continued on next page –

| | |
|---|---|
| Sensitivity analysis | different or modified outcome used (per capita deaths used rather than absolute deaths), start or end date of study period varied, same analysis with (sub)population(s) excluded |
| Subgroup assessment | none |

| **You 2020** | |
|---|---|
| **Study information** | |
| Author | You JH |
| Title | Impact of COVID-19 Infection Control Measures on Influenza-Related Outcomes in Hong Kong |
| Year | 2020 |
| Journal | Pathogens and Global Health |
| DOI | 10.1080/20477724.2020.1857492 |
| Date received | – could not be evaluated – |
| Date accepted | 2020-12-15 (published) |
| **Study setting** | |
| Number of populations included | single |
| Level of populations included | national (Hong Kong) |
| Geographic areas covered | Asia |
| Number of countries covered | single country (Hong Kong) |
| Study period | end date could not be evaluated |
| **Outcome** | |
| Raw outcome | epidemiological population-level outcome (surrogate disease: hospitalizations influenza) |
| Time resolution of raw outcome | weekly |

– continued on next page –

| | |
|---|---|
| Computed outcome | other (cases: influenza, deaths: influenza, clinical visits: influenza) |
| Method to obtain the computed outcome | other (multiply observed cases with ratios for clinical visits, deaths, hospitalizations, perform Monte Carlo simulation) |
| Data source | data from (sub)national authorities |
| Data availability | data access via source |
| **Intervention** | |
| Terminology for interventions | measures |
| Terminology for the specific type of non-pharmaceutical interventions | infection control |
| Exposure types | all interventions together |
| Types of single interventions | – not applicable – |
| Coding of interventions | – not applicable – |
| Source of intervention data | – not applicable – |
| Availability of data on exposure | – not applicable – |
| **Methodological approach** | |
| Empirical approach | descriptive |
| Use of exposure variation | only variation over time for a single population |
| Method | description of change over time (comparison of time periods) |
| Code availability | none |
| **Effect assessment** | |
| Reporting of intervention effect | quantification of change in outcome values |
| Measure of effect | hospitalizations: influenza |
| Reporting of uncertainty | yes |
| Sensitivity analysis | none |
| Subgroup assessment | none |

| You et al. 2020 | |
|---|---|
| **Study information** | |
| Author | You C, Deng Y, Hu W, Sun J, Lin Q, Zhou F, Pang CH, Zhang Y, Chen Z, Zhou XH |
| Title | Estimation of the Time-Varying Reproduction Number of COVID-19 Outbreak in China |
| Year | 2020 |
| Journal | International Journal of Hygiene and Environmental Health |
| DOI | 10.1016/j.ijheh.2020.113555 |
| Date received | 2020-03-10 |
| Date accepted | 2020-05-07 |
| **Study setting** | |
| Number of populations included | multiple |
| Level of populations included | both national and subnational (Hubei, Beijing, Shanghai, Guangdong, Zhejiang, and Henan (cities/provinces)) |
| Geographic areas covered | Asia |
| Number of countries covered | single country (China) |
| Study period | start and end date span first epidemic wave |
| **Outcome** | |
| Raw outcome | epidemiological individual-level outcome (individual cases and transmission chains) |
| Time resolution of raw outcome | daily |
| Computed outcome | epidemiological parameter (reproduction number) |
| Method to obtain the computed outcome | statistical estimation of reproduction number (White et al., Walinga et al.), compartmental transmission model (single-population) |
| Data source | data from (sub)national authorities |

– continued on next page –

| | |
|---|---|
| Data availability | data not accessible |
| **Intervention** | |
| Terminology for interventions | measures |
| Terminology for the specific type of non-pharmaceutical interventions | containment |
| Exposure types | one combination of interventions (Wuhan City Lockdown) |
| Types of single interventions | – not applicable – |
| Coding of interventions | not necessary |
| Source of intervention data | – could not be evaluated – |
| Availability of data on exposure | raw data documented in the manuscript |
| **Methodological approach** | |
| Empirical approach | descriptive |
| Use of exposure variation | only variation over time for multiple populations |
| Method | description of change over time (comparison of time periods) |
| Code availability | none |
| **Effect assessment** | |
| Reporting of intervention effect | comparison of outcome values |
| Measure of effect | reproduction number |
| Reporting of uncertainty | yes |
| Sensitivity analysis | none |
| Subgroup assessment | none |

| Yuan et al. 2020 | |
|---|---|
| **Study information** | |
| Author | Yuan Z, Xiao Y, Dai Z, Huang J, Zhang Z, Chen Y |
| Title | Modelling the Effects of Wuhan's Lockdown during COVID-19, China |
| Year | 2020 |
| Journal | Bulletin of the World Health Organization |
| DOI | 10.2471/blt.20.254045 |
| Date received | 2020-02-29 |
| Date accepted | 2020-05-01 |
| **Study setting** | |
| Number of populations included | multiple |
| Level of populations included | both national and subnational (China + 44 non-Wuhan provinces, cities) |
| Geographic areas covered | Asia |
| Number of countries covered | single country (China) |
| Study period | start and end date span first epidemic wave |
| **Outcome** | |
| Raw outcome | epidemiological population-level outcome (cases) |
| Time resolution of raw outcome | – not applicable – |
| Computed outcome | none |
| Method to obtain the computed outcome | – not applicable – |
| Data source | data from (sub)national authorities |
| Data availability | data made available by the authors |
| **Intervention** | |
| Terminology for interventions | measures |
| Terminology for the specific type of non-pharmaceutical interventions | control |

– continued on next page –

| | |
|---|---|
| Exposure types | all interventions together |
| Types of single interventions | – not applicable – |
| Coding of interventions | – not applicable – |
| Source of intervention data | – not applicable – |
| Availability of data on exposure | – not applicable – |
| **Methodological approach** | |
| Empirical approach | counterfactual |
| Use of exposure variation | both variation over time and between populations |
| Method | non-mechanistic model (generalized linear model) |
| Code availability | none |
| **Effect assessment** | |
| Reporting of intervention effect | comparison of outcome values |
| Measure of effect | cases |
| Reporting of uncertainty | no |
| Sensitivity analysis | none |
| Subgroup assessment | none |

**Zhang et al. 2020a**

| | |
|---|---|
| **Study information** | |
| Author | Zhang X, Ji Z, Zheng Y, Ye X, Li D |
| Title | Evaluating the Effect of City Lockdown on Controlling COVID-19 Propagation through Deep Learning and Network Science Models |
| Year | 2020 |
| Journal | Cities |

– continued on next page –

| | |
|---|---|
| DOI | 10.1016/j.cities.2020.102869 |
| Date received | 2020-05-08 |
| Date accepted | 2020-07-07 |
| **Study setting** | |
| Number of populations included | multiple |
| Level of populations included | both national and subnational (China and its provinces) |
| Geographic areas covered | Asia |
| Number of countries covered | single country (China) |
| Study period | start and end date span first epidemic wave |
| **Outcome** | |
| Raw outcome | epidemiological population-level outcome (cases), behavioral outcome (mobility: population flow) |
| Time resolution of raw outcome | daily |
| Computed outcome | other (correlation between mobility and infection rate between provinces) |
| Method to obtain the computed outcome | compartmental transmission model (metapopulation) |
| Data source | data from publicly available cross-country selections (WHO: cases), data from (sub)national authorities (Civil Aviation Administration of China: flight data) |
| Data availability | data access via source |
| **Intervention** | |
| Terminology for interventions | – not applicable – |
| Terminology for the specific type of non-pharmaceutical interventions | lockdown |
| Exposure types | one combination of interventions (Wuhan City Lockdown) |
| Types of single interventions | – not applicable – |

– continued on next page –

| | |
|---|---|
| Coding of interventions | not necessary |
| Source of intervention data | – could not be evaluated – |
| Availability of data on exposure | raw data documented in the manuscript |
| **Methodological approach** | |
| Empirical approach | descriptive |
| Use of exposure variation | both variation over time and between populations |
| Method | description of change over time (comparison of time periods) |
| Code availability | none |
| **Effect assessment** | |
| Reporting of intervention effect | qualitative statement |
| Measure of effect | correlation between mobility and infection rate between provinces |
| Reporting of uncertainty | – not applicable – |
| Sensitivity analysis | none |
| Subgroup assessment | none |

**Zhang et al. 2020b**

| | |
|---|---|
| **Study information** | |
| Author | Zhang L, Yang H, Wang K, Zhan Y, Bian L |
| Title | Measuring Imported Case Risk of COVID-19 from Inbound International Flights —A Case Study on China |
| Year | 2020 |
| Journal | Journal of Air Transport Management |
| DOI | 10.1016/j.jairtraman.2020.101918 |
| Date received | 2020-05-29 |

– continued on next page –

| | |
|---|---|
| Date accepted | 2020-08-23 |
| **Study setting** | |
| Number of populations included | multiple |
| Level of populations included | both national and subnational (China and its provinces) |
| Geographic areas covered | Asia |
| Number of countries covered | single country (China) |
| Study period | start and end date span first epidemic wave |
| **Outcome** | |
| Raw outcome | epidemiological population-level outcome (cases, mobility: international flight data) |
| Time resolution of raw outcome | daily |
| Computed outcome | other (imported case risk index) |
| Method to obtain the computed outcome | other (ODE model) |
| Data source | data from publicly available cross-country selections (JHU: cases), mobility data from corporate organizations (UMETRIP: mobility) |
| Data availability | data access via source |
| **Intervention** | |
| Terminology for interventions | – not applicable – |
| Terminology for the specific type of non-pharmaceutical interventions | travel ban |
| Exposure types | one single intervention (international travel restrictions) |
| Types of single interventions | international travel restrictions |
| Coding of interventions | not necessary |
| Source of intervention data | – could not be evaluated – |
| Availability of data on exposure | raw data documented in the manuscript |

– continued on next page –

| | |
|---|---|
| **Methodological approach** | |
| Empirical approach | counterfactual |
| Use of exposure variation | only variation over time for multiple populations |
| Method | other (simulation of risk index using mobility before intervention) |
| Code availability | none |
| **Effect assessment** | |
| Reporting of intervention effect | comparison of outcome values |
| Measure of effect | imported case risk index |
| Reporting of uncertainty | no |
| Sensitivity analysis | none |
| Subgroup assessment | none |

**Zhang et al. 2020c**

| | |
|---|---|
| **Study information** | |
| Author | Zhang J, Dong L, Zhang Y, Chen X, Yao G, Han Z |
| Title | Investigating Time, Strength, and Duration of Measures in Controlling the Spread of COVID-19 Using a Networked Meta-Population Model |
| Year | 2020 |
| Journal | Nonlinear Dynamics |
| DOI | 10.1007/s11071-020-05769-2 |
| Date received | 2020-04-16 |
| Date accepted | 2020-06-14 |
| **Study setting** | |
| Number of populations included | multiple |

– continued on next page –

| | |
|---|---|
| Level of populations included | both national and subnational (China and some of its large cities) |
| Geographic areas covered | Asia |
| Number of countries covered | single country (China) |
| Study period | start and end date span first epidemic wave |
| **Outcome** | |
| Raw outcome | epidemiological population-level outcome (cases) |
| Time resolution of raw outcome | daily |
| Computed outcome | none |
| Method to obtain the computed outcome | – not applicable – |
| Data source | data from (sub)national authorities |
| Data availability | data made available by the authors |
| **Intervention** | |
| Terminology for interventions | measures |
| Terminology for the specific type of non-pharmaceutical interventions | intervention |
| Exposure types | one combination of interventions (Wuhan City Lockdown) |
| Types of single interventions | – not applicable – |
| Coding of interventions | not necessary |
| Source of intervention data | government or news websites |
| Availability of data on exposure | raw data documented in the manuscript |
| **Methodological approach** | |
| Empirical approach | counterfactual |
| Use of exposure variation | both variation over time and between populations |
| Method | mechanistic model (compartmental metapopulation transmission model) |

– continued on next page –

| | |
|---|---|
| Code availability | publicly available |
| **Effect assessment** | |
| Reporting of intervention effect | comparison of outcome values |
| Measure of effect | infections |
| Reporting of uncertainty | yes |
| Sensitivity analysis | none |
| Subgroup assessment | none |

| **Zhang et al. 2020d** | |
|---|---|
| **Study information** | |
| Author | Zhang B, Zhou H, Zhou F |
| Title | Study on SARS-CoV-2 Transmission and the Effects of Control Measures in China |
| Year | 2020 |
| Journal | PLOS ONE |
| DOI | 10.1371/journal.pone.0242649 |
| Date received | 2020-02-18 |
| Date accepted | 2020-10-16 |
| **Study setting** | |
| Number of populations included | multiple |
| Level of populations included | both national and subnational (China, Wuhan, Hubei) |
| Geographic areas covered | Asia |
| Number of countries covered | single country (China) |
| Study period | start and end date span first epidemic wave |

– continued on next page –

| | |
|---|---|
| **Outcome** | |
| Raw outcome | epidemiological population-level outcome (cases, deaths) |
| Time resolution of raw outcome | daily |
| Computed outcome | epidemiological parameter (reproduction number) |
| Method to obtain the computed outcome | compartmental transmission model (single-population) |
| Data source | data from (sub)national authorities |
| Data availability | data made available by the authors |
| **Intervention** | |
| Terminology for interventions | measures |
| Terminology for the specific type of non-pharmaceutical interventions | control |
| Exposure types | one combination of interventions (Wuhan City Lockdown) |
| Types of single interventions | – not applicable – |
| Coding of interventions | not necessary |
| Source of intervention data | – could not be evaluated – |
| Availability of data on exposure | raw data documented in the manuscript |
| **Methodological approach** | |
| Empirical approach | descriptive |
| Use of exposure variation | only variation over time for multiple populations |
| Method | description of change over time (comparison of time periods) |
| Code availability | none |
| **Effect assessment** | |
| Reporting of intervention effect | comparison of outcome values |
| Measure of effect | reproduction number |
| Reporting of uncertainty | no |

– continued on next page –

| | |
|---|---|
| Sensitivity analysis | none |
| Subgroup assessment | none |

| **Zhao & Chen 2020** | |
|---|---|
| **Study information** | |
| Author | Zhao S, Chen H |
| Title | Modeling the Epidemic Dynamics and Control of COVID-19 Outbreak in China |
| Year | 2020 |
| Journal | Quantitative Biology |
| DOI | 10.1007/s40484-020-0199-0 |
| Date received | 2020-02-29 |
| Date accepted | 2020-03-04 |
| **Study setting** | |
| Number of populations included | multiple |
| Level of populations included | both national and subnational (Wuhan, rest of Hubei, rest of China, four Chinese cities) |
| Geographic areas covered | Asia |
| Number of countries covered | single country (China) |
| Study period | start and end date span first epidemic wave |
| **Outcome** | |
| Raw outcome | epidemiological population-level outcome (cases) |
| Time resolution of raw outcome | daily |
| Computed outcome | none |

– continued on next page –

| | |
|---|---|
| Method to obtain the computed outcome | – not applicable – |
| Data source | data from (sub)national authorities |
| Data availability | data access via source |
| **Intervention** | |
| Terminology for interventions | measures |
| Terminology for the specific type of non-pharmaceutical interventions | control |
| Exposure types | phases of interventions (three stages of measures in Wuhan/rest of China) |
| Types of single interventions | – not applicable – |
| Coding of interventions | not necessary |
| Source of intervention data | – could not be evaluated – |
| Availability of data on exposure | raw data documented in the manuscript |
| **Methodological approach** | |
| Empirical approach | counterfactual |
| Use of exposure variation | only variation over time for multiple populations |
| Method | mechanistic model (compartmental single-population transmission model) |
| Code availability | none |
| **Effect assessment** | |
| Reporting of intervention effect | comparison of outcome values |
| Measure of effect | cases |
| Reporting of uncertainty | no |
| Sensitivity analysis | model specification varied |
| Subgroup assessment | none |

| Zhao et al. 2020 | |
|---|---|
| **Study information** | |
| Author | Zhao L, Feng D, Ye RZ, Wang HT, Zhou YH, Wei JT, J. de Vlas S, Cui XM, Jia N, Yin CN, Li SX, Wang ZQ, Cao WC |
| Title | Outbreak of COVID-19 and SARS in Mainland China: A Comparative Study Based on National Surveillance Data |
| Year | 2020 |
| Journal | BMJ Open |
| DOI | 10.1136/bmjopen-2020-043411 |
| Date received | 2020-08-04 |
| Date accepted | 2020-10-02 |
| **Study setting** | |
| Number of populations included | single |
| Level of populations included | subnational (Hubei (China)) |
| Geographic areas covered | Asia |
| Number of countries covered | single country (China) |
| Study period | start and end date span first epidemic wave |
| **Outcome** | |
| Raw outcome | epidemiological individual-level outcome (individual cases) |
| Time resolution of raw outcome | daily |
| Computed outcome | epidemiological parameter (reproduction number) |
| Method to obtain the computed outcome | statistical estimation of reproduction number (Cori et al.) |
| Data source | data from (sub)national authorities |
| Data availability | data not accessible |
| **Intervention** | |
| Terminology for interventions | measures |

– continued on next page –

| | |
|---|---|
| Terminology for the specific type of non-pharmaceutical interventions | control |
| Exposure types | one combination of interventions (Wuhan City Lockdown) |
| Types of single interventions | – not applicable – |
| Coding of interventions | not necessary |
| Source of intervention data | – could not be evaluated – |
| Availability of data on exposure | raw data documented in the manuscript |
| **Methodological approach** | |
| Empirical approach | descriptive |
| Use of exposure variation | only variation over time for a single population |
| Method | description of change over time (description of time course) |
| Code availability | none |
| **Effect assessment** | |
| Reporting of intervention effect | qualitative statement |
| Measure of effect | reproduction number |
| Reporting of uncertainty | no |
| Sensitivity analysis | none |
| Subgroup assessment | none |

### F Excluded studies

Through title and abstract screening, 411 studies were identified as potentially relevant and their full texts evaluated. Of these, 163 studies did not meet the eligibility criteria and were subsequently excluded. In the following, these excluded studies are listed together with the reason for exclusion.

### Only simulation of NPI effects in hypothetical scenarios

| Only simulation of NPI effects in hypothetical scenarios | |
|---|---|
| Author | Aleta A, Moreno Y |
| Title | Evaluation of the potential incidence of COVID-19 and effectiveness of containment measures in Spain: A data-driven approach |
| Year | 2020 |
| Journal | BMC Medicine |
| DOI | 10.1186/s12916-020-01619-5 |
| Author | Alrashed S, Min-Allah N, Saxena A, Ali I, Mehmood R |
| Title | Impact of lockdowns on the spread of COVID-19 in Saudi Arabia |
| Year | 2020 |
| Journal | Informatics in Medicine Unlocked |
| DOI | 10.1016/j.imu.2020.100420 |
| Author | Alrasheed H, Althnian A, Kurdi H, Al-Mgren H, Alharbi S |
| Title | COVID-19 spread in Saudi Arabia: Modeling, simulation and analysis |
| Year | 2020 |
| Journal | International Journal of Environmental Research and Public Health |
| DOI | 10.3390/ijerph17217744 |
| Author | Arino J, Bajeux N, Portet S, Watmough J |
| Title | Quarantine and the risk of COVID-19 importation |
| Year | 2020 |
| Journal | Epidemiology and Infection |
| DOI | 10.1017/S0950268820002988 |
| Author | Awaidy SA, Mahomed O |
| Title | Impact of non-pharmaceutical interventions on the COVID-19 epidemic: A modelling study |

– continued on next page –

| | |
|---|---|
| Year | 2020 |
| Journal | SAGE Open Medicine |
| DOI | 10.1177/2050312120979462 |
| Author | Azanza Ricardo CL, Hernandez-Vargas EA |
| Title | Epidemiological Characteristics of COVID-19 in Mexico and the Potential Impact of Lifting Confinement Across Regions |
| Year | 2020 |
| Journal | Frontiers in Physics |
| DOI | 10.3389/fphy.2020.573322 |
| Author | Baek YJ, Lee T, Cho Y, Hyun JH, Kim MH, Sohn Y, Kim JH, Ahn JY, Jeong SJ, Ku NS, Yeom JS, Lee J, Choi JY |
| Title | A mathematical model of COVID-19 transmission in a tertiary hospital and assessment of the effects of different intervention strategies |
| Year | 2020 |
| Journal | PLOS ONE |
| DOI | 10.1371/journal.pone.0241169 |
| Author | Bemanian A, Ahn KW, O'Brien M, Rausch DJ, Weston B, Beyer KMM |
| Title | Investigating the Trajectory of the COVID-19 Outbreak in Milwaukee County and Projected Effects of Relaxed Distancing |
| Year | 2020 |
| Journal | WMJ |
| DOI | not available |
| Author | Bentout S, Tridane A, Djilali S, Touaoula TM |
| Title | Age-Structured Modeling of COVID-19 Epidemic in the USA, UAE and Algeria |
| Year | 2020 |

– continued on next page –

| | |
|---|---|
| Journal | Alexandria Engineering Journal |
| DOI | 10.1016/j.aej.2020.08.053 |
| Author | Bouchnita A, Jebrane A |
| Title | A hybrid multi-scale model of COVID-19 transmission dynamics to assess the potential of non-pharmaceutical interventions |
| Year | 2020 |
| Journal | Chaos, Solitons and Fractals |
| DOI | 10.1016/j.chaos.2020.109941 |
| Author | Buonomo, B., Della Marca, R. |
| Title | Effects of information-induced behavioural changes during the COVID-19 lockdowns: The case of Italy: COVID-19 lockdowns and behavioral change |
| Year | 2020 |
| Journal | Royal Society Open Science |
| DOI | 10.1098/rsos.201635rsos201635 |
| Author | Cabore JW, Karamagi HC, Kipruto H, Asamani JA, Droti B, Seydi ABW, Titi-Ofei R, Impouma B, Yao M, Yoti Z, Zawaira F, Tumusiime P, Talisuna A, Kasolo FC, Moeti MR |
| Title | The potential effects of widespread community transmission of SARS-CoV-2 infection in the World Health Organization African Region: A predictive model |
| Year | 2020 |
| Journal | BMJ Global Health |
| DOI | 10.1136/bmjgh-2020-002647 |
| Author | Cano OB, Morales SC, Bendtsen C |
| Title | Covid-19 modelling: The effects of social distancing |
| Year | 2020 |

– continued on next page –

| | |
|---|---|
| Journal | Interdisciplinary Perspectives on Infectious Diseases |
| DOI | 10.1155/2020/2041743 |
| Author | Casares M, Khan H |
| Title | The timing and intensity of social distancing to flatten the COVID-19 curve: The case of Spain |
| Year | 2020 |
| Journal | International Journal of Environmental Research and Public Health |
| DOI | 10.3390/ijerph17197283 |
| Author | Castilho C, Gondim JAM, Marchesin M, Sabeti M |
| Title | Assessing the efficiency of different control strategies for the covid-19 epidemic |
| Year | 2020 |
| Journal | Electronic Journal of Differential Equations |
| DOI | not available |
| Author | Chang SL, Harding N, Zachreson C, Cliff OM, Prokopenko M |
| Title | Modelling transmission and control of the COVID-19 pandemic in Australia |
| Year | 2020 |
| Journal | Nature Communications |
| DOI | 10.1038/s41467-020-19393-6 |
| Author | Chen S, Li Q, Gao S, Kang Y, Shi X |
| Title | State-specific projection of COVID-19 infection in the United States and evaluation of three major control measures |
| Year | 2020 |
| Journal | Scientific Reports |
| DOI | 10.1038/s41598-020-80044-3 |

– continued on next page –

| | |
|---|---|
| Author | Choi S, Ki M |
| Title | Estimating the reproductive number and the outbreak size of COVID-19 in Korea |
| Year | 2020 |
| Journal | Epidemiology and health |
| DOI | 10.4178/epih.e2020011 |
| Author | Costantino V, Heslop DJ, MacIntyre CR |
| Title | The effectiveness of full and partial travel bans against COVID-19 spread in Australia for travellers from China during and after the epidemic peak in China |
| Year | 2020 |
| Journal | Journal of travel medicine |
| DOI | 10.1093/jtm/taaa081 |
| Author | DeFries R, Agarwala M, Baquie S, Choksi P, Dogra N, Preetha GS, Khanwilkar S, Mondal P, Nagendra H, Urpelainen J |
| Title | Post-lockdown spread of COVID-19 from cities to vulnerable forest-fringe villages in Central India |
| Year | 2020 |
| Journal | Current Science |
| DOI | 10.18520/cs/v119/i1/52-58 |
| Author | Faranda D, Alberti T |
| Title | Modeling the second wave of COVID-19 infections in France and Italy via a stochastic SEIR model |
| Year | 2020 |
| Journal | Chaos |
| DOI | 10.1063/5.0015943 |

– continued on next page –

| | |
|---|---|
| Author | Feng LX, Jing SL, Hu SK, Wang DF, Huo HF |
| Title | Modelling the effects of media coverage and quarantine on the COVID-19 infections in the UK |
| Year | 2020 |
| Journal | Mathematical Biosciences and Engineering |
| DOI | 10.3934/MBE.2020204 |
| Author | Giordano G, Blanchini F, Bruno R, Colaneri P, Di Filippo A, Di Matteo A, Colaneri M |
| Title | Modelling the COVID-19 epidemic and implementation of population-wide interventions in Italy |
| Year | 2020 |
| Journal | Nature Medicine |
| DOI | 10.1038/s41591-020-0883-7 |
| Author | Girona T |
| Title | Confinement Time Required to Avoid a Quick Rebound of COVID-19: Predictions From a Monte Carlo Stochastic Model |
| Year | 2020 |
| Journal | Frontiers in Physics |
| DOI | 10.3389/fphy.2020.00186 |
| Author | Godin A, Xia Y, Buckeridge DL, Mishra S, Douwes-Schultz D, Shen Y, Lavigne M, Drolet M, Schmidt AM, Brisson M, Maheu-Giroux M |
| Title | The role of case importation in explaining differences in early SARS-CoV-2 transmission dynamics in Canada—A mathematical modeling study of surveillance data |
| Year | 2020 |
| Journal | International Journal of Infectious Diseases |

– continued on next page –

| | |
|---|---|
| DOI | 10.1016/j.ijid.2020.10.046 |
| Author | Gountas I, Hillas G, Souliotis K |
| Title | Act early, save lives: managing COVID-19 in Greece |
| Year | 2020 |
| Journal | Public Health |
| DOI | 10.1016/j.puhe.2020.08.016 |
| Author | Gul S, Tuncay K, Binici B, Aydin BB |
| Title | Transmission dynamics of Covid-19 in Italy, Germany and Turkey considering social distancing, testing and quarantine |
| Year | 2020 |
| Journal | Journal of Infection in Developing Countries |
| DOI | 10.3855/jidc.12844 |
| Author | Gupta SD, Jain R, Bhatnagar S |
| Title | COVID-19 Pandemic in Rajasthan: Mathematical Modelling and Social Distancing |
| Year | 2020 |
| Journal | Journal of Health Management |
| DOI | 10.1177/0972063420935537 |
| Author | Guzzetta G, Poletti P, Ajelli M, Trentini F, Marziano V, Cereda D, Tirani M, Diurno G, Bodina A, Barone A, Crottogini L, Gramegna M, Melegaro A, Merler S |
| Title | Potential short-term outcome of an uncontrolled COVID-19 epidemic in Lombardy, Italy, February to March 2020 |
| Year | 2020 |
| Journal | Eurosurveillance |
| DOI | 10.2807/1560-7917.ES.2020.25.12.2000293 |

– continued on next page –

| | |
|---|---|
| Author | Hammoumi A, Qesmi R |
| Title | Impact assessment of containment measure against COVID-19 spread in Morocco |
| Year | 2020 |
| Journal | Chaos, Solitons and Fractals |
| DOI | 10.1016/j.chaos.2020.110231 |
| Author | Hogan AB, Jewell BL, Sherrard-Smith E, Vesga JF, Watson OJ, Whittaker C, Hamlet A, Smith JA, Winskill P, Verity R, Baguelin M, Lees JA, Whittles LK, Ainslie KEC, Bhatt S, Boonyasiri A, Brazeau NF, Cattarino L, Cooper LV, Coupland H, Cuomo-Dannenburg G, Dighe A, Djaafara BA, Donnelly CA, Eaton JW, van Elsland SL, FitzJohn RG, Fu H, Gaythorpe KAM, Green W, Haw DJ, Hayes S, Hinsley W, Imai N, Laydon DJ, Mangal TD, Mellan TA, Mishra S, Nedjati-Gilani G, Parag KV, Thompson HA, Unwin HJT, Vollmer MAC, Walters CE, Wang HW, Wang YR, Xi XY, Ferguson NM, Okell LC, Churcher TS, Arinaminpathy N, Ghani AC, Walker PGT, Hallett TB |
| Title | Potential impact of the COVID-19 pandemic on HIV, tuberculosis, and malaria in low-income and middle-income countries: a modelling study |
| Year | 2020 |
| Journal | Lancet Global Health |
| DOI | 10.1016/S2214-109X(20)30288-6 |
| Author | Hossain MP, Junus A, Zhu X, Jia P, Wen TH, Pfeiffer D, Yuan HY |
| Title | The effects of border control and quarantine measures on the spread of COVID-19 |
| Year | 2020 |
| Journal | Epidemics |
| DOI | 10.1016/j.epidem.2020.100397 |

– continued on next page –

| | |
|---|---|
| Author | Hou C, Chen JX, Zhou YQ, Hua L, Yuan JX, He S, Guo Y, Zhang S, Jia QW, Zhao CH, Zhang J, Xu GX, Jia EZ |
| Title | The effectiveness of quarantine of Wuhan city against the Corona Virus Disease 2019 (COVID-19): A well-mixed SEIR model analysis |
| Year | 2020 |
| Journal | Journal of Medical Virology |
| DOI | 10.1002/jmv.25827 |
| Author | Hu L, Nie LF |
| Title | Dynamic modeling and analysis of COVID-19 in different transmission process and control strategies |
| Year | 2020 |
| Journal | Mathematical Methods in the Applied Sciences |
| DOI | 10.1002/mma.6839 |
| Author | Huang YB, Wu Y, Zhang WD |
| Title | Comprehensive identification and isolation policies have effectively suppressed the spread of COVID-19 |
| Year | 2020 |
| Journal | Chaos, Solitons and Fractals |
| DOI | 10.1016/j.chaos.2020.110041 |
| Author | Iboi EA, Sharomi O, Ngonghala CN, Gumel AB |
| Title | Mathematical modeling and analysis of COVID-19 pandemic in Nigeria |
| Year | 2020 |
| Journal | Mathematical Biosciences and Engineering |
| DOI | 10.3934/MBE.2020369 |
| Author | Jackson ML |

– continued on next page –

| | |
|---|---|
| Title | Low-impact social distancing interventions to mitigate local epidemics of SARS-CoV-2 |
| Year | 2020 |
| Journal | Microbes and Infection |
| DOI | 10.1016/j.micinf.2020.09.006 |
| Author | Jia JW, Ding J, Liu SY, Liao GD, Li JZ, Duan B, Wang GQ, Zhang R |
| Title | Modeling the control of Covid-19: Impact of policy interventions and meteorological factors |
| Year | 2020 |
| Journal | Electronic Journal Of Differential Equations |
| DOI | not available |
| Author | Jiang S, Li Q, Li C, Liu S, He X, Wang T, Li H, Corpe C, Zhang X, Xu J, Wang J |
| Title | Mathematical models for devising the optimal SARS-CoV-2 strategy for eradication in China, South Korea, and Italy |
| Year | 2020 |
| Journal | Journal of Translational Medicine |
| DOI | 10.1186/s12967-020-02513-7 |
| Author | Kane GQ, Amba MCO, Mbratana T |
| Title | Early view of the potential impact of government policy responses on reproductive number of COVID-19 in African countries: Evidence from Cameroon |
| Year | 2020 |
| Journal | Economics Bulletin |
| DOI | not available |
| Author | Kassa SM, Njagarah JBH, Terefe YA |

– continued on next page –

| | |
|---|---|
| Title | Analysis of the mitigation strategies for COVID-19: From mathematical modelling perspective |
| Year | 2020 |
| Journal | Chaos, Solitons and Fractals |
| DOI | 10.1016/j.chaos.2020.109968 |
| Author | Khajji B, Kada D, Balatif O, Rachik M |
| Title | A multi-region discrete time mathematical modeling of the dynamics of Covid-19 virus propagation using optimal control |
| Year | 2020 |
| Journal | Journal of Applied Mathematics and Computing |
| DOI | 10.1007/s12190-020-01354-3 |
| Author | Kheirallah KA, Alsinglawi B, Alzoubi A, Saidan MN, Mubin O, Alorjani MS, Mzayek F |
| Title | The effect of strict state measures on the epidemiologic curve of covid-19 infection in the context of a developing country: A simulation from Jordan |
| Year | 2020 |
| Journal | International Journal of Environmental Research and Public Health |
| DOI | 10.3390/ijerph17186530 |
| Author | Kim BN, Kim E, Lee S, Oh C |
| Title | Mathematical model of covid-19 transmission dynamics in South Korea: The impacts of travel restrictions, social distancing, and early detection |
| Year | 2020 |
| Journal | Processes |
| DOI | 10.3390/pr8101304 |
| Author | Kim S, Kim YJ, Peck KR, Jung E |

– continued on next page –

| | |
|---|---|
| Title | School Opening Delay Effect on Transmission Dynamics of Coronavirus Disease 2019 in Korea: Based on Mathematical Modeling and Simulation Study |
| Year | 2020 |
| Journal | Journal of Korean medical science |
| DOI | 10.3346/jkms.2020.35.e143 |
| Author | Kimathi M, Mwalili S, Ojiambo V, Gathungu DK |
| Title | Age-structured model for COVID-19: Effectiveness of social distancing and contact reduction in Kenya |
| Year | 2020 |
| Journal | Infectious Disease Modelling |
| DOI | 10.1016/j.idm.2020.10.012 |
| Author | Kucharski AJ, Klepac P, Conlan AJK, Kissler SM, Tang ML, Fry H, Gog JR, Edmunds WJ, Emery JC, Medley G, Munday JD, Russell TW, Leclerc QJ, Diamond C, Procter SR, Gimma A, Sun FY, Gibbs HP, Rosello A, van Zandvoort K, Hué S, Meakin SR, Deol AK, Knight G, Jombart T, Foss AM, Bosse NI, Atkins KE, Quilty BJ, Lowe R, Prem K, Flasche S, Pearson CAB, Houben RMGJ, Nightingale ES, Endo A, Tully DC, Liu Y, Villabona-Arenas J, O'Reilly K, Funk S, Eggo RM, Jit M, Rees EM, Hellewell J, Clifford S, Jarvis CI, Abbott S, Auzenbergs M, Davies NG, Simons D |
| Title | Effectiveness of isolation, testing, contact tracing, and physical distancing on reducing transmission of SARS-CoV-2 in different settings: a mathematical modelling study |
| Year | 2020 |
| Journal | The Lancet Infectious Diseases |
| DOI | 10.1016/S1473-3099(20)30457-6 |
| Author | Kuniya T, Inaba H |

– continued on next page –

| | |
|---|---|
| Title | Possible effects of mixed prevention strategy for COVID-19 epidemic: massive testing, quarantine and social distancing |
| Year | 2020 |
| Journal | AIMS Public Health |
| DOI | 10.3934/publichealth.2020040 |
| Author | Leung K, Wu JT, Liu D, Leung GM |
| Title | First-wave COVID-19 transmissibility and severity in China outside Hubei after control measures, and second-wave scenario planning: a modelling impact assessment |
| Year | 2020 |
| Journal | The Lancet |
| DOI | 10.1016/S0140-6736(20)30746-7 |
| Author | Lhous M, Zakary O, Rachik M, Magri EM, Tridane A |
| Title | Optimal containment control strategy of the second phase of the covid-19 lockdown in morocco |
| Year | 2020 |
| Journal | Applied Sciences (Switzerland) |
| DOI | 10.3390/app10217559 |
| Author | Linka K, Rahman P, Goriely A, Kuhl E |
| Title | Is it safe to lift COVID-19 travel bans? The Newfoundland story |
| Year | 2020 |
| Journal | Computational Mechanics |
| DOI | 10.1007/s00466-020-01899-x |
| Author | Loewenthal G, Abadi S, Avram O, Halabi K, Ecker N, Nagar N, Mayrose I, Pupko T |
| Title | An evaluation of COVID-19 in Italy: A data-driven modeling analysis |

– continued on next page –

| | |
|---|---|
| Year | 2020 |
| Journal | Infectious Disease Modelling |
| DOI | 10.1016/j.idm.2020.06.007 |
| Author | Mahdizadeh Gharakhanlou N, Hooshangi N |
| Title | Spatio-temporal simulation of the novel coronavirus (COVID-19) outbreak using the agent-based modeling approach (case study: Urmia, Iran) |
| Year | 2020 |
| Journal | Informatics in Medicine Unlocked |
| DOI | 10.1016/j.imu.2020.100403 |
| Author | Marimuthu Y, Nagappa B, Sharma N, Basu S, Chopra KK |
| Title | COVID-19 and tuberculosis: A mathematical model based forecasting in Delhi, India |
| Year | 2020 |
| Journal | Indian Journal of Tuberculosis |
| DOI | 10.1016/j.ijtb.2020.05.006 |
| Author | Matrajt L, Leung T |
| Title | Evaluating the effectiveness of social distancing interventions to delay or flatten the epidemic curve of Coronavirus disease |
| Year | 2020 |
| Journal | Emerging Infectious Diseases |
| DOI | 10.3201/eid2608.201093 |
| Author | McCombs A, Kadelka C |
| Title | A model-based evaluation of the efficacy of COVID-19 social distancing, testing and hospital triage policies |
| Year | 2020 |
| Journal | PLoS Computational Biology |

– continued on next page –

| | |
|---|---|
| DOI | 10.1371/journal.pcbi.1008388 |
| Author | Milhinhos A, Costa PM |
| Title | On the Progression of COVID-19 in Portugal: A Comparative Analysis of Active Cases Using Non-linear Regression |
| Year | 2020 |
| Journal | Frontiers in Public Health |
| DOI | 10.3389/fpubh.2020.00495 |
| Author | Min KD, Kang H, Lee JY, Jeon S, Cho SI |
| Title | Estimating the effectiveness of non-pharmaceutical interventions on COVID-19 control in Korea |
| Year | 2020 |
| Journal | Journal of Korean Medical Science |
| DOI | 10.3346/JKMS.2020.35.E321 |
| Author | Murillo MFM, Murillo WAM, Pandey D |
| Title | Efficient Prediction and Analysis of the possible evolution of the SARS-CoV-2 in COVID-19 applying the SEIR model in Santo Domingo de los Tsachilas, Ecuador |
| Year | 2020 |
| Journal | Enfoque UTE |
| DOI | 10.29019/enfoqueute.v11n4.678 |
| Author | Nawaz A, Su X, Iqbal S, Zahoor H, Asad A, Asghar S, Basit F, Barkat MQ, Souhail A, Raheel Shah SA |
| Title | Validating a Phenomenological Mathematical Model for Public Health and Safety Interventions Influencing the Evolutionary Stages of Recent Outbreak for Long-Term and Short-Term Domains in Pakistan |
| Year | 2020 |
| Journal | Complexity |

– continued on next page –

| | |
|---|---|
| DOI | 10.1155/2020/8866071 |
| Author | Ng WL |
| Title | To lockdown? When to peak? Will there be an end? A macroeconomic analysis on COVID-19 epidemic in the United States |
| Year | 2020 |
| Journal | Journal of Macroeconomics |
| DOI | 10.1016/j.jmacro.2020.103230 |
| Author | Ngonghala CN, Iboi E, Eikenberry S, Scotch M, MacIntyre CR, Bonds MH, Gumel AB |
| Title | Mathematical assessment of the impact of non-pharmaceutical interventions on curtailing the 2019 novel Coronavirus |
| Year | 2020 |
| Journal | Mathematical Biosciences |
| DOI | 10.1016/j.mbs.2020.108364 |
| Author | Nkwayep CH, Bowong S, Tewa JJ, Kurths J |
| Title | Short-term forecasts of the COVID-19 pandemic: a study case of Cameroon |
| Year | 2020 |
| Journal | Chaos, Solitons and Fractals |
| DOI | 10.1016/j.chaos.2020.110106 |
| Author | Nuzzo A, Tan CO, Raskar R, DeSimone DC, Kapa S, Gupta R |
| Title | Universal Shelter-in-Place Versus Advanced Automated Contact Tracing and Targeted Isolation: A Case for 21st-Century Technologies for SARS-CoV-2 and Future Pandemics |
| Year | 2020 |
| Journal | Mayo Clinic Proceedings |
| DOI | 10.1016/j.mayocp.2020.06.027 |

– continued on next page –

| | |
|---|---|
| Author | Olaniyi S, Obabiyi OS, Okosun KO, Oladipo AT, Adewale SO |
| Title | Mathematical modelling and optimal cost-effective control of COVID-19 transmission dynamics |
| Year | 2020 |
| Journal | European Physical Journal Plus |
| DOI | 10.1140/epjp/s13360-020-00954-z |
| Author | Olfatifar M, Houri H, Shojaee S, Pourhoseingholi MA, Alali WQ, Busani L, Ashtari S, Shahrokh S, Vahedian A, Aghdaei HA |
| Title | The Required Confronting Approaches Efficacy and Time to Control COVID-19 Outbreak in Iran |
| Year | 2020 |
| Journal | Archives ff Clinical Infectious Diseases |
| DOI | 10.5812/archcid.102633 |
| Author | Overton CE, Stage HB, Ahmad S, Curran-Sebastian J, Dark P, Das R, Fearon E, Felton T, Fyles M, Gent N, Hall I, House T, Lewkowicz H, Pang X, Pellis L, Sawko R, Ustianowski A, Vekaria B, Webb L |
| Title | Using statistics and mathematical modelling to understand infectious disease outbreaks: COVID-19 as an example |
| Year | 2020 |
| Journal | Infectious Disease Modelling |
| DOI | 10.1016/j.idm.2020.06.008 |
| Author | Ozair M, Hussain T, Hussain M, Awan AU, Baleanu D, Abro KA |
| Title | A Mathematical and Statistical Estimation of Potential Transmission and Severity of COVID-19: A Combined Study of Romania and Pakistan |
| Year | 2020 |
| Journal | BioMed Research International |

– continued on next page –

| | |
|---|---|
| DOI | 10.1155/2020/5607236 |
| Author | Pais RJ, Taveira N |
| Title | Predicting the evolution and control of the COVID-19 pandemic in Portugal |
| Year | 2020 |
| Journal | F1000Research |
| DOI | 10.12688/f1000research.23401.2 |
| Author | Pan J, Tian J, Xiong H, Liu Z, Yao Y, Wang Y, Zhu W, Wang Y, Wang W |
| Title | Risk assessment and evaluation of China's policy to prevent COVID-19 cases imported by plane |
| Year | 2020 |
| Journal | PLoS neglected tropical diseases |
| DOI | 10.1371/journal.pntd.0008908 |
| Author | Peirlinck M, Linka K, Sahli Costabal F, Kuhl E |
| Title | Outbreak dynamics of COVID-19 in China and the United States |
| Year | 2020 |
| Journal | Biomechanics and Modeling in Mechanobiology |
| DOI | 10.1007/s10237-020-01332-5 |
| Author | Pernice S, Castagno P, Marcotulli L, Maule MM, Richiardi L, Moirano G, Sereno M, Cordero F, Beccuti M |
| Title | Impacts of reopening strategies for COVID-19 epidemic: a modeling study in Piedmont region |
| Year | 2020 |
| Journal | BMC Infectious Diseases |
| DOI | 10.1186/s12879-020-05490-w |
| Author | Rădulescu A, Williams C, Cavanagh K |

– continued on next page –

| | |
|---|---|
| Title | Management strategies in a SEIR-type model of COVID 19 community spread |
| Year | 2020 |
| Journal | Scientific Reports |
| DOI | 10.1038/s41598-020-77628-4 |
| Author | Rainisch G, Undurraga EA, Chowell G |
| Title | A dynamic modeling tool for estimating healthcare demand from the COVID19 epidemic and evaluating population-wide interventions |
| Year | 2020 |
| Journal | International Journal of Infectious Diseases |
| DOI | 10.1016/j.ijid.2020.05.043 |
| Author | Razzaq OA, Rehman DU, Khan NA, Ahmadian A, Ferrara M |
| Title | Optimal surveillance mitigation of COVID'19 disease outbreak: Fractional order optimal control of compartment model |
| Year | 2020 |
| Journal | Results in Physics |
| DOI | 10.1016/j.rinp.2020.103715 |
| Author | Rivera-Rodriguez C, Urdinola BP |
| Title | Predicting Hospital Demand During the COVID-19 Outbreak in Bogota, Colombia |
| Year | 2020 |
| Journal | Frontiers in Public Health |
| DOI | 10.3389/fpubh.2020.582706 |
| Author | Ruktanonchai NW, Floyd JR, Lai S, Ruktanonchai CW, Sadilek A, Rente-Lourenco P, Ben X, Carioli A, Gwinn J, Steele JE, Prosper O, Schneider A, Oplinger A, Eastham P, Tatem AJ |

– continued on next page –

| | |
|---|---|
| Title | Assessing the impact of coordinated COVID-19 exit strategies across Europe |
| Year | 2020 |
| Journal | Science |
| DOI | 10.1126/science.abc5096 |
| Author | Saldaña F, Flores-Arguedas H, Camacho-Gutiérrez JA, Barradas I |
| Title | Modeling the transmission dynamics and the impact of the control interventions for the COVID-19 epidemic outbreak |
| Year | 2020 |
| Journal | Mathematical biosciences and engineering |
| DOI | 10.3934/mbe.2020231 |
| Author | Schneider KA, Ngwa GA, Schwehm M, Eichner L, Eichner M |
| Title | The COVID-19 pandemic preparedness simulation tool: CovidSIM |
| Year | 2020 |
| Journal | BMC Infectious Diseases |
| DOI | 10.1186/s12879-020-05566-7 |
| Author | Singh A, Chandra SK, Bajpai MK |
| Title | Study of non-pharmacological interventions on covid-19 spread |
| Year | 2020 |
| Journal | CMES - Computer Modeling in Engineering and Sciences |
| DOI | 10.32604/cmes.2020.011601 |
| Author | Sjödin H, Johansson AF, Brännström Å, Farooq Z, Kriit HK, Wilder-Smith A, Åström C, Thunberg J, Söderquist M, Rocklöv J |
| Title | COVID-19 healthcare demand and mortality in Sweden in response to non-pharmaceutical mitigation and suppression scenarios |
| Year | 2020 |
| Journal | International Journal of Epidemiology |

– continued on next page –

| | |
|---|---|
| DOI | 10.1093/ije/dyaa121 |
| Author | Su L, Hong N, Zhou X, He J, Ma Y, Jiang H, Han L, Chang F, Shan G, Zhu W, Long Y |
| Title | Evaluation of the Secondary Transmission Pattern and Epidemic Prediction of COVID-19 in the Four Metropolitan Areas of China |
| Year | 2020 |
| Journal | Frontiers in Medicine |
| DOI | 10.3389/fmed.2020.00171 |
| Author | Sy C, Bernardo E, Miguel A, San Juan JL, Mayol AP, Ching PM, Culaba A, Ubando A, Mutuc JE |
| Title | Policy Development for Pandemic Response Using System Dynamics: a Case Study on COVID-19 |
| Year | 2020 |
| Journal | Process Integration and Optimization for Sustainability |
| DOI | 10.1007/s41660-020-00130-x |
| Author | Taboe HB, Salako KV, Tison JM, Ngonghala CN, Glèlè Kakaï R |
| Title | Predicting COVID-19 spread in the face of control measures in West Africa |
| Year | 2020 |
| Journal | Mathematical Biosciences |
| DOI | 10.1016/j.mbs.2020.108431 |
| Author | Tang B, Scarabel F, Bragazzi NL, McCarthy Z, Glazer M, Xiao Y, Heffernan JM, Asgary A, Ogden NH, Wu J |
| Title | De-escalation by reversing the escalation with a stronger synergistic package of contact tracing, quarantine, isolation and personal protection: Feasibility of preventing a covid-19 rebound in Ontario, Canada, as a case study |
| Year | 2020 |
| Journal | Biology |

– continued on next page –

| | |
|---|---|
| DOI | 10.3390/biology9050100 |
| Author | Tantrakarnapa K, Bhopdhornangkul B |
| Title | Challenging the spread of COVID-19 in Thailand |
| Year | 2020 |
| Journal | One Health |
| DOI | 10.1016/j.onehlt.2020.100173 |
| Author | Tatapudi H, Das R, Das TK |
| Title | Impact assessment of full and partial stay-at-home orders, face mask usage, and contact tracing: An agent-based simulation study of COVID-19 for an urban region |
| Year | 2020 |
| Journal | Global Epidemiology |
| DOI | 10.1016/j.gloepi.2020.100036 |
| Author | Tocto-Erazo MR, Espíndola-Zepeda JA, Montoya-Laos JA, Acuña-Zegarra MA, Olmos-Liceaga D, Reyes-Castro PA, FigueroaPreciado G |
| Title | Lockdown, relaxation, and acme period in COVID-19: A study of disease dynamics in Hermosillo, Sonora, Mexico |
| Year | 2020 |
| Journal | PLOS ONE |
| DOI | 10.1371/journal.pone.0242957 |
| Author | Traini MC, Caponi C, Ferrari R, De Socio GV |
| Title | A study of SARS-CoV-2 epidemiology in Italy: from early days to secondary effects after social distancing |
| Year | 2020 |
| Journal | Infectious Diseases |
| DOI | 10.1080/23744235.2020.1797157 |

– continued on next page –

| | |
|---|---|
| Author | Tsay C, Lejarza F, Stadtherr MA, Baldea M |
| Title | Modeling, state estimation, and optimal control for the US COVID-19 outbreak |
| Year | 2020 |
| Journal | Scientific reports |
| DOI | 10.1038/s41598-020-67459-8 |
| Author | Valenti VE, de Lemos Menezes P, de Abreu ACG, Vieira GNA, Garner DM |
| Title | Social distancing measures could have reduced estimated deaths related to COVID-19 in Brazil |
| Year | 2020 |
| Journal | Journal of Human Growth and Development |
| DOI | 10.7322/JHGD.V30.10360 |
| Author | Varotsos CA, Krapivin VF |
| Title | A new model for the spread of COVID-19 and the improvement of safety |
| Year | 2020 |
| Journal | Safety Science |
| DOI | 10.1016/j.ssci.2020.104962 |
| Author | Volpert V, Banerjee M, Petrovskii S |
| Title | On a quarantine model of coronavirus infection and data analysis |
| Year | 2020 |
| Journal | Mathematical Modelling of Natural Phenomena |
| DOI | 10.1051/mmnp/2020006 |
| Author | Wang K, Lu ZZ, Wang XM, Li H, Li HL, Lin DD, Cai YL, Feng X, Song YT, Feng ZW, Ji WD, Wang XY, Yin Y, Wang L, Peng ZH |
| Title | Current trends and future prediction of novel coronavirus disease (COVID-19) epidemic in China: a dynamical modeling analysis |

– continued on next page –

| | |
|---|---|
| Year | 2020 |
| Journal | Mathematical biosciences and engineering |
| DOI | 10.3934/mbe.2020173 |
| Author | Wang X, Du Z, Huang G, Pasco RF, Fox SJ, Galvani AP, Pignone M, Claiborne Johnston S, Meyers LA |
| Title | Effects of cocooning on coronavirus disease rates after relaxing social distancing |
| Year | 2020 |
| Journal | Emerging Infectious Diseases |
| DOI | 10.3201/EID2612.201930 |
| Author | Wang X, Pasco RF, Du Z, Petty M, Fox SJ, Galvani AP, Pignone M, Claiborne Johnston S, Meyers LA |
| Title | Impact of social distancing measures on coronavirus disease healthcare demand, central Texas, USA |
| Year | 2020 |
| Journal | Emerging Infectious Diseases |
| DOI | 10.3201/eid2610.201702 |
| Author | Wei Y, Wang J, Song W, Xiu C, Ma L, Pei T |
| Title | Spread of COVID-19 in China: analysis from a city-based epidemic and mobility model |
| Year | 2020 |
| Journal | Cities |
| DOI | 10.1016/j.cities.2020.103010 |
| Author | Yang JY, Wang GQ, Zhang S |
| Title | Impact of household quarantine on SARS-Cov-2 infection in mainland China: A mean-field modelling approach |

– continued on next page –

| | |
|---|---|
| Year | 2020 |
| Journal | Mathematical biosciences and engineering |
| DOI | 10.3934/mbe.2020248 |
| Author | Yoo S, Managi S |
| Title | Global mortality benefits of COVID-19 action |
| Year | 2020 |
| Journal | Technological Forecasting And Social Change |
| DOI | 10.1016/j.techfore.2020.120231 |
| Author | Zhang K, Vilches TN, Tariq M, Galvani AP, Moghadas SM |
| Title | The impact of mask-wearing and shelter-in-place on COVID-19 outbreaks in the United States |
| Year | 2020 |
| Journal | International Journal of Infectious Diseases |
| DOI | 10.1016/j.ijid.2020.10.002 |
| Author | Zhang Y, Yu X, Sun H, Tick GR, Wei W, Jin B |
| Title | Applicability of time fractional derivative models for simulating the dynamics and mitigation scenarios of COVID-19 |
| Year | 2020 |
| Journal | Chaos, Solitons and Fractals |
| DOI | 10.1016/j.chaos.2020.109959 |
| Author | Zhou Y, Xu RZ, Hu DS, Yue Y, Li QQ, Xia JZ |
| Title | Effects of human mobility restrictions on the spread of COVID-19 in Shenzhen, China: a modelling study using mobile phone data |
| Year | 2020 |
| Journal | Lancet Digital Health |

– continued on next page –

| | |
|---|---|
| DOI | 10.1016/S2589-7500(20)30165-5 |
| Author | Zhu HJ, Li Y, Jin XL, Huang JP, Liu X, Qian Y, Tan JD |
| Title | Transmission dynamics and control methodology of COVID-19: A modeling study |
| Year | 2020 |
| Journal | Applied Mathematical Modelling |
| DOI | 10.1016/j.apm.2020.08.056 |
| Author | Zongo P, Zorom M, Mophou G, Dorville R, Beaumont C |
| Title | A model of covid-19 transmission to understand the effectiveness of the containment measures: Application to data from France |
| Year | 2020 |
| Journal | Epidemiology and Infection |
| DOI | 10.1017/S0950268820002162 |
| Author | Zu J, Li ML, Li ZF, Shen MW, Xiao YN, Ji FP |
| Title | Transmission patterns of COVID-19 in the mainland of China and the efficacy of different control strategies: a data- And model-driven study |
| Year | 2020 |
| Journal | Infectious Diseases of Poverty |
| DOI | 10.1186/s40249-020-00709-z |

### Different research objective than assessment of NPI effects

**Different research objective than assessment of NPI effects**

| | |
|---|---|
| Author | Bajiya VP, Bugalia S, Tripathi JP |
| Title | Mathematical modeling of COVID-19: Impact of non-pharmaceutical interventions in India |
| Year | 2020 |

– continued on next page –

| | |
|---|---|
| Journal | Chaos |
| DOI | 10.1063/5.0021353 |
| Author | Bianconi A, Marcelli A, Campi G, Perali A |
| Title | Ostwald Growth Rate in Controlled Covid-19 Epidemic Spreading as in Arrested Growth in Quantum Complex Matter |
| Year | 2020 |
| Journal | Condensed Matter |
| DOI | 10.3390/condmat5020023 |
| Author | Biswas SK, Ghosh JK, Sarkar S, Ghosh U |
| Title | COVID-19 pandemic in India: a mathematical model study |
| Year | 2020 |
| Journal | Nonlinear Dynamics |
| DOI | 10.1007/s11071-020-05958-z |
| Author | Bright A, Glynn-Robinson AJ, Kane S, Wright R, Saul N |
| Title | The effect of COVID-19 public health measures on nationally notifiable diseases in Australia: preliminary analysis |
| Year | 2020 |
| Journal | Communicable diseases intelligence |
| DOI | 10.33321/cdi.2020.44.85 |
| Author | De Visscher A |
| Title | The COVID-19 pandemic: model-based evaluation of non-pharmaceutical interventions and prognoses |
| Year | 2020 |
| Journal | Nonlinear Dynamics |
| DOI | 10.1007/s11071-020-05861-7 |

– continued on next page –

| | |
|---|---|
| Author | Debecker A, Modis T |
| Title | Poorly known aspects of flattening the curve of COVID-19 |
| Year | 2020 |
| Journal | Technological Forecasting and Social Change |
| DOI | 10.1016/j.techfore.2020.120432 |
| Author | Grossman G., Kim S., Rexer J.M., Thirumurthy H. |
| Title | Political partisanship influences behavioral responses to governors' recommendations for COVID-19 prevention in the United States |
| Year | 2020 |
| Journal | Proceedings of the National Academy of Sciences of the United States of America |
| DOI | 10.1073/pnas.2007835117 |
| Author | Hong HG, Li Y |
| Title | Estimation of time-varying reproduction numbers underlying epidemiological processes: A new statistical tool for the COVID-19 pandemic |
| Year | 2020 |
| Journal | PLOS ONE |
| DOI | 10.1371/journal.pone.0236464 |
| Author | Kaxiras E, Neofotistos G |
| Title | Multiple epidemic wave model of the COVID-19 pandemic: Modeling study |
| Year | 2020 |
| Journal | Journal of Medical Internet Research |
| DOI | 10.2196/20912 |
| Author | Kosmidis K, Macheras P |
| Title | A fractal kinetics SI model can explain the dynamics of COVID-19 epidemics |
| Year | 2020 |

– continued on next page –

| | |
|---|---|
| Journal | PLOS ONE |
| DOI | 10.1371/journal.pone.0237304 |
| Author | Lee W, Hwang SS, Song I, Park C, Kim H, Song IK, Choi HM, Prifti K, Kwon Y, Kim J, Oh S, Yang J, Cha M, Kim Y, Bell ML, Kim H |
| Title | COVID-19 in South Korea: Epidemiological and spatiotemporal patterns of the spread and the role of aggressive diagnostic tests in the early phase |
| Year | 2020 |
| Journal | International Journal of Epidemiology |
| DOI | 10.1093/ije/dyaa119 |
| Author | Lupu D, Maha LG, Viorică ED |
| Title | Covid-19 incidence in europe: Drivers and government interventions |
| Year | 2020 |
| Journal | Transylvanian Review of Administrative Sciences |
| DOI | 10.24193/tras.SI2020.5 |
| Author | Mahajan P, Kaushal J |
| Title | Epidemic Trend of COVID-19 Transmission in India During Lockdown-1 Phase |
| Year | 2020 |
| Journal | Journal of community health |
| DOI | 10.1007/s10900-020-00863-3 |
| Author | Maziarz M, Zach M |
| Title | Agent-based modelling for SARS-CoV-2 epidemic prediction and intervention assessment: A methodological appraisal |
| Year | 2020 |
| Journal | Journal of Evaluation in Clinical Practice |
| DOI | 10.1111/jep.13459 |

– continued on next page –

| | |
|---|---|
| Author | Monllor P, Su Z, Gabrielli L, Taltavull de La Paz P |
| Title | COVID-19 Infection Process in Italy and Spain: Are Data Talking? Evidence From ARMA and Vector Autoregression Models |
| Year | 2020 |
| Journal | Frontiers in Public Health |
| DOI | 10.3389/fpubh.2020.550602 |
| Author | Ponjavić M, Karabegović A, Ferhatbegović E, Tahirović E, Uzunović S, Travar M, Pilav A, Mulić M, Karakaš S, Avdić N, Mulabdić Z, Pavić G, Bičo M, Vasilj I, Mamić D, Hukić M |
| Title | Spatio-temporal data visualization for monitoring of control measures in the prevention of the spread of COVID-19 in Bosnia and Herzegovina |
| Year | 2020 |
| Journal | Medicinski Glasnik |
| DOI | 10.17392/1215-20 |
| Author | Pratomo H |
| Title | From Social Distancing to Physical Distancing: A Challenge for Evaluating Public Health Intervention against COVID-19 |
| Year | 2020 |
| Journal | Kesmas: National Public Health Journal |
| DOI | 10.21109/kesmas.v15i2.4010 |
| Author | Rajendrakumar AL, Nair ATN, Nangia C, Chourasia PK, Chourasia MK, Syed MG, Nair AS, Nair AB, Koya MSF |
| Title | Epidemic Landscape and Forecasting of SARS-CoV-2 in India |
| Year | 2020 |
| Journal | Journal of Epidemiology and Global Health |
| DOI | 10.2991/jegh.k.200823.001 |

– continued on next page –

| | |
|---|---|
| Author | Ramchandani A, Fan C, Mostafavi A |
| Title | DeepCOVIDNet: An Interpretable Deep Learning Model for Predictive Surveillance of COVID-19 Using Heterogeneous Features and Their Interactions |
| Year | 2020 |
| Journal | IEEE Access |
| DOI | 10.1109/ACCESS.2020.3019989 |
| Author | Rotejanaprasert C, Lawpoolsri S, Pan-Ngum W, Maude RJ |
| Title | Preliminary estimation of temporal and spatiotemporal dynamic measures of COVID- 19 transmission in Thailand |
| Year | 2020 |
| Journal | PLOS ONE |
| DOI | 10.1371/journal.pone.0239645 |
| Author | Sun J, Chen X, Zhang Z, Lai S, Zhao B, Liu H, Wang S, Huan W, Zhao R, Ng MTA, Zheng Y |
| Title | Forecasting the long-term trend of COVID-19 epidemic using a dynamic model |
| Year | 2020 |
| Journal | Scientific Reports |
| DOI | 10.1038/s41598-020-78084-w |
| Author | Sun ZB, Zhang H, Yang YF, Wan H, Wang YX |
| Title | Impacts of geographic factors and population density on the COVID-19 spreading under the lockdown policies of China |
| Year | 2020 |
| Journal | Science ff The Total Environment |
| DOI | 10.1016/j.scitotenv.2020.141347 |

– continued on next page –

| | |
|---|---|
| Author | Tariq A, Lee Y, Roosa K, Blumberg S, Yan P, Ma S, Chowell G |
| Title | Real-time monitoring the transmission potential of COVID-19 in Singapore, March 2020 |
| Year | 2020 |
| Journal | BMC Medicine |
| DOI | 10.1186/s12916-020-01615-9 |
| Author | Wang KW, Gao J, Wang H, Wu XL, Yuan QF, Guo FY, Zhang ZJ, Cheng Y |
| Title | Epidemiology of 2019 novel coronavirus in Jiangsu Province, China after wartime control measures: A population-level retrospective study |
| Year | 2020 |
| Journal | Travel Medicine and Infectious Disease |
| DOI | 10.1016/j.tmaid.2020.101654 |
| Author | Wang PP, Zheng XQ, Ai G, Liu DY, Zhu BR |
| Title | Time series prediction for the epidemic trends of COVID-19 using the improved LSTM deep learning method: Case studies in Russia, Peru and Iran |
| Year | 2020 |
| Journal | Chaos Solitons & Fractals |
| DOI | 10.1016/j.chaos.2020.110214 |
| Author | Ylli A, Wu YY, Burazeri G, Pirkle C, Sentell T |
| Title | The lower COVID-19 related mortality and incidence rates in Eastern European countries are associated with delayed start of community circulation |
| Year | 2020 |
| Journal | PLOS ONE |
| DOI | 10.1371/journal.pone.0243411 |

– continued on next page –

| | |
|---|---|
| Author | Zhang N, Shi T, Zhong H, Guo Y |
| Title | COVID-19 Prevention and Control Public Health Strategies in Shanghai, China |
| Year | 2020 |
| Journal | Journal of public health management and practice : JPHMP |
| DOI | 10.1097/PHH.0000000000001202 |
| Author | Zhou L, Liu JM, Dong XP, McGoogan JM, Wu ZY |
| Title | COVID-19 seeding time and doubling time model: An early epidemic risk assessment tool |
| Year | 2020 |
| Journal | Infectious Diseases of Poverty |
| DOI | 10.1186/s40249-020-00685-4 |
| Author | Zhou WK, Wang AL, Xia F, Xiao YN, Tang SY |
| Title | Effects of media reporting on mitigating spread of COVID-19 in the early phase of the outbreak |
| Year | 2020 |
| Journal | Mathematical biosciences and engineering |
| DOI | 10.3934/mbe.2020147 |
| Author | Zhou YM, Li J, Ye LJ, Chen ZG, Luo QS, Wu XD, Ni HY |
| Title | Random Network Transmission and Countermeasures in Containing Global Spread of COVID-19-Alike Pandemic: A Hybrid Modelling Approach |
| Year | 2020 |
| Journal | Complexity |
| DOI | 10.1155/2020/6703703 |

### Only assessment of effects of mobility or behavior, not of NPIs

| Only assessment of effects of mobility or behavior, not of NPIs | |
|---|---|
| Author | Borjas GJ |
| Title | Business closures, stay-at-home restrictions, and COVID-19 testing outcomes in New York City |
| Year | 2020 |
| Journal | Preventing Chronic Disease |
| DOI | 10.5888/PCD17.200264 |
| Author | Bryant P, Elofsson A |
| Title | Estimating the impact of mobility patterns on COVID-19 infection rates in 11 European countries |
| Year | 2020 |
| Journal | PeerJ |
| DOI | 10.7717/peerj.9879 |
| Author | Delen D, Eryarsoy E, Davazdahemami B |
| Title | No place like home: Cross-national data analysis of the efficacy of social distancing during the COVID-19 pandemic |
| Year | 2020 |
| Journal | JMIR Public Health and Surveillance |
| DOI | 10.2196/19862 |
| Author | Durmus H, Gökler ME, Metintas S |
| Title | The effectiveness of community-based social distancing for mitigating the spread of the COVID-19 pandemic in Turkey |
| Year | 2020 |
| Journal | Journal of Preventive Medicine and Public Health |
| DOI | 10.3961/JPMPH.20.381 |

– continued on next page –

| | |
|---|---|
| Author | Ferguson NM, Ainslie KEC, Walters CE, Fu H, Bhatia S, Wang H, Xi X, Baguelin M, Bhatt S, Boonyasiri A, Boyd O, Cattarino L, Ciavarella C, Cucunuba Z, Cuomo-Dannenburg G, Dighe A, Dorigatti I, van Elsland SL, FitzJohn R, Gaythorpe K, Ghani AC, Green W, Hamlet A, Hinsley W, Imai N, Jorgensen D, Knock E, Laydon D, Nedjati-Gilani G, Okell LC, Siveroni I, Thompson HA, Unwin HJT, Verity R, Vollmer M, Walker PGT, Wang Y, Watson OJ, Whittaker C, Winskill P, Donnelly CA, Riley S |
| Title | Evidence of initial success for China exiting COVID-19 social distancing policy after achieving containment |
| Year | 2020 |
| Journal | Wellcome Open Research |
| DOI | 10.12688/wellcomeopenres.15843.1 |
| Author | Iacus SM, Santamaria C, Sermi F, Spyratos S, Tarchi D, Vespe M |
| Title | Human mobility and COVID-19 initial dynamics |
| Year | 2020 |
| Journal | Nonlinear Dynamics |
| DOI | 10.1007/s11071-020-05854-6 |
| Author | Jang WM, Jang DH, Lee JY |
| Title | Social Distancing and Transmission-reducing Practices during the 2019 Coronavirus Disease and 2015 Middle East Respiratory Syndrome Coronavirus Outbreaks in Korea |
| Year | 2020 |
| Journal | Journal of Korean Medical Science |
| DOI | 10.3346/jkms.2020.35.e220 |
| Author | Kain MP, Childs ML, Becker AD, Mordecai EA |
| Title | Chopping the tail: How preventing superspreading can help to maintain COVID-19 control |

– continued on next page –

| | |
|---|---|
| Year | 2020 |
| Journal | Epidemics |
| DOI | 10.1016/j.epidem.2020.100430 |
| Author | Lai S, Ruktanonchai NW, Zhou L, Prosper O, Luo W, Floyd JR, Wesolowski A, Santillana M, Zhang C, Du X, Yu H, Tatem AJ |
| Title | Effect of non-pharmaceutical interventions to contain COVID-19 in China |
| Year | 2020 |
| Journal | Nature |
| DOI | 10.1038/s41586-020-2293-x |
| Author | Loewenthal G, Abadi S, Avram O, Halabi K, Ecker N, Nagar N, Mayrose I, Pupko T |
| Title | COVID-19 pandemic-related lockdown: response time is more important than its strictness |
| Year | 2020 |
| Journal | Embo Molecular Medicine |
| DOI | 10.15252/emmm.202013171 |
| Author | Morley CP, Anderson KB, Shaw J, Stewart T, Thomas SJ, Wang D |
| Title | Social Distancing Metrics and Estimates of SARS-CoV-2 Transmission Rates: Associations Between Mobile Telephone Data Tracking and R |
| Year | 2020 |
| Journal | Journal of Public Health Management and Practice |
| DOI | 10.1097/PHH.0000000000001240 |
| Author | Ogundokun RO, Lukman AF, Kibria GBM, Awotunde JB, Aladeitan BB |
| Title | Predictive modelling of COVID-19 confirmed cases in Nigeria |
| Year | 2020 |
| Journal | Infectious Disease Modelling |

– continued on next page –

| | |
|---|---|
| DOI | 10.1016/j.idm.2020.08.003 |
| Author | Rubin D, Huang J, Fisher BT, Gasparrini A, Tam V, Song L, Wang X, Kaufman J, Fitzpatrick K, Jain A, Griffis H, Crammer K, Morris J, Tasian G |
| Title | Association of Social Distancing, Population Density, and Temperature with the Instantaneous Reproduction Number of SARS-CoV-2 in Counties across the United States |
| Year | 2020 |
| Journal | JAMA Network Open |
| DOI | 10.1001/jamanetworkopen.2020.16099 |
| Author | Unwin HJT, Mishra S, Bradley VC, Gandy A, Mellan TA, Coupland H, Ish-Horowicz J, Vollmer MAC, Whittaker C, Filippi SL, Xi X, Monod M, Ratmann O, Hutchinson M, Valka F, Zhu H, Hawryluk I, Milton P, Ainslie KEC, Baguelin M, Boonyasiri A, Brazeau NF, Cattarino L, Cucunuba Z, Cuomo-Dannenburg G, Dorigatti I, Eales OD, Eaton JW, van Elsland SL, FitzJohn RG, Gaythorpe KAM, Green W, Hinsley W, Jeffrey B, Knock E, Laydon DJ, Lees J, Nedjati-Gilani G, Nouvellet P, Okell L, Parag KV, Siveroni I, Thompson HA, Walker P, Walters CE, Watson OJ, Whittles LK, Ghani AC, Ferguson NM, Riley S, Donnelly CA, Bhatt S, Flaxman S |
| Title | State-level tracking of COVID-19 in the United States |
| Year | 2020 |
| Journal | Nature Communications |
| DOI | 10.1038/s41467-020-19652-6 |
| Author | Vokó Z, Pitter JG |
| Title | The effect of social distance measures on COVID-19 epidemics in Europe: an interrupted time series analysis |
| Year | 2020 |
| Journal | GeroScience |

– continued on next page –

**Only assessment of effects of mobility or behavior, not of NPIs** – continued from previous page

| | |
|---|---|
| DOI | 10.1007/s11357-020-00205-0 |
| Author | Yuan X, Hu K, Xu J, Zhang X, Bao W, Lynch CF, Zhang L |
| Title | State heterogeneity in the associations of human mobility with COVID-19 epidemics in the European Union |
| Year | 2020 |
| Journal | American Journal of Translational Research |
| DOI | not available |
| Author | Zhao Y, Wang R, Li J, Zhang Y, Yang H |
| Title | Analysis of the Transmissibility Change of 2019-Novel Coronavirus Pneumonia and Its Potential Factors in China from 2019 to 2020 |
| Year | 2020 |
| Journal | BioMed Research International |
| DOI | 10.1155/2020/3842470 |

### Use of results from other study

| Use of results from other study | |
|---|---|
| Author | Dighe A, Cattarino L, Cuomo-Dannenburg G, Skarp J, Imai N, Bhatia S, Gaythorpe KAM, Ainslie KEC, Baguelin M, Bhatt S, Boonyasiri A, Brazeau NF, Cooper LV, Coupland H, Cucunuba Z, Dorigatti I, Eales OD, Van Elsland SL, Fitzjohn RG, Green WD, Haw DJ, Hinsley W, Knock E, Laydon DJ, Mellan T, Mishra S, Nedjati-Gilani G, Nouvellet P, Pons-Salort M, Thompson HA, Unwin HJT, Verity R, Vollmer MAC, Walters CE, Watson OJ, Whittaker C, Whittles LK, Ghani AC, Donnelly CA, Ferguson NM, Riley S |
| Title | Response to COVID-19 in South Korea and implications for lifting stringent interventions |
| Year | 2020 |

– continued on next page –

| | |
|---|---|
| Journal | BMC Medicine |
| DOI | 10.1186/s12916-020-01791-8 |
| Author | Galbán-García E, Más-Bermejo P |
| Title | COVID-19 in Cuba: Assessing the national response |
| Year | 2020 |
| Journal | MEDICC Review |
| DOI | 10.37757/MR2020.V22.N4.5 |
| Author | Giachino M, Valera CBG, Velásquez SR, Dohrendorf-Wyss MA, Rozanova L, Flahault A |
| Title | Understanding the dynamics of the covid-19 pandemic: A real-time analysis of switzerland's first wave |
| Year | 2020 |
| Journal | International Journal of Environmental Research and Public Health |
| DOI | 10.3390/ijerph17238825 |
| Author | Hassan R, Dosar AS, Mondol JK, Khan TH, Noman AA, Sayem MS, Hasan M, Juyena NS |
| Title | Prediction of Epidemics Trend of COVID-19 in Bangladesh |
| Year | 2020 |
| Journal | Frontiers in Public Health |
| DOI | 10.3389/fpubh.2020.559437 |
| Author | Imai N, Gaythorpe KAM, Abbott S, Bhatia S, van Elsland S, Prem K, Liu Y, Ferguson NM |
| Title | Adoption and impact of non-pharmaceutical interventions for COVID-19 |
| Year | 2020 |
| Journal | Wellcome Open Research |
| DOI | 10.12688/wellcomeopenres.15808.1 |

– continued on next page –

**Use of results from other study** – continued from previous page

| | |
|---|---|
| Author | Medina-Ortiz D, Contreras S, Barrera-Saavedra Y, Cabas-Mora G, Olivera-Nappa Á |
| Title | Country-Wise Forecast Model for the Effective Reproduction Number R<inf>t</inf> of Coronavirus Disease |
| Year | 2020 |
| Journal | Frontiers in Physics |
| DOI | 10.3389/fphy.2020.00304 |

### Extraction not feasible due to quality issues

**Extraction not feasible due to quality issues**

| | |
|---|---|
| Author | Adly HM, AlJahdali IA, Garout MA, Khafagy AA, Saati AA, Saleh SAK |
| Title | Correlation of COVID-19 Pandemic with Healthcare System Response and Prevention Measures in Saudi Arabia |
| Year | 2020 |
| Journal | International Journal of Environmental Research and Public Health |
| DOI | 10.3390/ijerph17186666 |
| Author | Al Zobbi M, Alsinglawi B, Mubin O, Alnajjar F |
| Title | Measurement Method for Evaluating the Lockdown Policies during the COVID-19 Pandemic |
| Year | 2020 |
| Journal | International Journal of Environmental Research and Public Health |
| DOI | 10.3390/ijerph17155574 |
| Author | Rajendran S, Jayagopal P |
| Title | Accessing Covid19 epidemic outbreak in Tamilnadu and the impact of lockdown through epidemiological models and dynamic systems |
| Year | 2020 |

– continued on next page –

**Extraction not feasible due to quality issues** – continued from previous page

| | |
|---|---|
| Journal | Measurement |
| DOI | 10.1016/j.measurement.2020.108432 |
| Author | Rui R, Tian M |
| Title | Joint estimation of case fatality rate of COVID-19 and power of quarantine strategy performed in Wuhan, China |
| Year | 2020 |
| Journal | Biometrical Journal |
| DOI | 10.1002/bimj.202000116 |

### Not available in English language

| **Not available in English language** | |
|---|---|
| Author | Cao S, Feng P, Shi P |
| Title | Study on the epidemic development of COVID-19 in Hubei province by a modified SEIR model |
| Year | 2020 |
| Journal | Journal of Zhejiang University |
| DOI | not available |

### G PRISMA checklist

| Section and Topic | Item # | Checklist Item | Location where item is reported |
|---|---|---|---|
| **TITLE** | | | |
| Title | 1 | Identify the report as a systematic review. | p. 1 |
| **ABSTRACT** | | | |
| Abstract | 2 | See the PRISMA 2020 for Abstracts checklist | p. 1 |
| **INTRODUCTION** | | | |
| Rationale | 3 | Describe the rationale for the review in the context of existing knowledge. | p. 2 |
| Objectives | 4 | Provide an explicit statement of the objective(s) or question(s) the review addresses. | p. 3 |
| **METHODS** | | | |
| Eligibility criteria | 5 | Specify the inclusion and exclusion criteria for the review and how studies were grouped for the syntheses. | pp. 29–30 |
| Information sources | 6 | Specify all databases, registers, websites, organisations, reference lists and other sources searched or consulted to identify studies. Specify the date when each source was last searched or consulted. | p. 31 |
| Search strategy | 7 | Present the full search strategies for all databases, registers and websites, including any filters and limits used. | p. 31, SI Appendix B pp. 3–7 |
| Selection process | 8 | Specify the methods used to decide whether a study met the inclusion criteria of the review, including how many reviewers screened each record and each report retrieved, whether they worked independently, and if applicable, details of automation tools used in the process. | p. 32 |
| Data collection process | 9 | Specify the methods used to collect data from reports, including how many reviewers collected data from each report, whether they worked independently, any processes for obtaining or confirming data from study investigators, and if applicable, details of automation tools used in the process. | pp. 32–33 |

– continued on next page –

| | | | |
|---|---|---|---|
| Data items | 10a | List and define all outcomes for which data were sought. Specify whether all results that were compatible with each outcome domain in each study were sought (e.g. for all measures, time points, analyses), and if not, the methods used to decide which results to collect. | SI Appendix D pp. 9–24 |
| | 10b | List and define all other variables for which data were sought (e.g. participant and intervention characteristics, funding sources). Describe any assumptions made about any missing or unclear information. | SI Appendix D pp. 9–24 |
| Study risk of bias assessment | 11 | Specify the methods used to assess risk of bias in the included studies, including details of the tool(s) used, how many reviewers assessed each study and whether they worked independently, and if applicable, details of automation tools used in the process. | p. 33 |
| Effect measures | 12 | Specify for each outcome the effect measure(s) (e.g. risk ratio, mean difference) used in the synthesis or presentation of results. | – not applicable – |
| Synthesis methods | 13a | Describe the processes used to decide which studies were eligible for each synthesis (e.g. tabulating the study intervention characteristics and comparing against the planned groups for each synthesis (item 5)). | pp. 29–30 |
| | 13b | Describe any methods required to prepare the data for presentation or synthesis, such as handling of missing summary statistics, or data conversions. | – not applicable – |
| | 13c | Describe any methods used to tabulate or visually display results of individual studies and syntheses. | pp. 33-34 |
| | 13d | Describe any methods used to synthesize results and provide a rationale for the choice(s). If meta-analysis was performed, describe the model(s), method(s) to identify the presence and extent of statistical heterogeneity, and software package(s) used. | pp. 33-34 |
| | 13e | Describe any methods used to explore possible causes of heterogeneity among study results (e.g. subgroup analysis, meta-regression). | – not applicable – |
| | 13f | Describe any sensitivity analyses conducted to assess robustness of the synthesized results. | – not applicable – |

– continued on next page –

| | | | |
|---|---|---|---|
| Reporting bias assessment | 14 | Describe any methods used to assess risk of bias due to missing results in a synthesis (arising from reporting biases). | – not applicable – |
| Certainty assessment | 15 | Describe any methods used to assess certainty (or confidence) in the body of evidence for an outcome. | – not applicable – |
| **RESULTS** | | | |
| Study selection | 16a | Describe the results of the search and selection process, from the number of records identified in the search to the number of studies included in the review, ideally using a flow diagram. | pp. 3–4 |
| | 16b | Cite studies that might appear to meet the inclusion criteria, but which were excluded, and explain why they were excluded. | p. 3, SI Appendix F pp. 618–659 |
| Study characteristics | 17 | Cite each included study and present its characteristics. | SI Appendix E pp. 25–617 |
| Risk of bias in studies | 18 | Present assessments of risk of bias for each included study. | – not applicable – |
| Results of individual studies | 19 | For all outcomes, present, for each study: (a) summary statistics for each group (where appropriate) and (b) an effect estimate and its precision (e.g. confidence/credible interval), ideally using structured tables or plots. | – not applicable – |
| Results of syntheses | 20a | For each synthesis, briefly summarise the characteristics and risk of bias among contributing studies. | – not applicable – |
| | 20b | Present results of all statistical syntheses conducted. If meta-analysis was done, present for each the summary estimate and its precision (e.g. confidence/credible interval) and measures of statistical heterogeneity. If comparing groups, describe the direction of the effect. | – not applicable – |
| | 20c | Present results of all investigations of possible causes of heterogeneity among study results. | – not applicable – |
| | 20d | Present results of all sensitivity analyses conducted to assess the robustness of the synthesized results. | – not applicable – |
| Reporting biases | 21 | Present assessments of risk of bias due to missing results (arising from reporting biases) for each synthesis assessed. | – not applicable – |

– continued on next page –

| | | | |
|---|---|---|---|
| Certainty of evidence | 22 | Present assessments of certainty (or confidence) in the body of evidence for each outcome assessed. | – not applicable – |
| **DISCUSSION** | | | |
| Discussion | 23a | Provide a general interpretation of the results in the context of other evidence. | pp. 21–27 |
| | 23b | Discuss any limitations of the evidence included in the review. | p. 28 |
| | 23c | Discuss any limitations of the review processes used. | pp. 29-31 |
| | 23d | Discuss implications of the results for practice, policy, and future research. | pp. 27-28 |
| **OTHER INFORMATION** | | | |
| Registration and protocol | 24a | Provide registration information for the review, including register name and registration number, or state that the review was not registered. | p. 29 |
| | 24b | Indicate where the review protocol can be accessed, or state that a protocol was not prepared. | p. 29 |
| | 24c | Describe and explain any amendments to information provided at registration or in the protocol. | p. 31 |
| Support | 25 | Describe sources of financial or non-financial support for the review, and the role of the funders or sponsors in the review. | p. 46 |
| Competing interests | 26 | Declare any competing interests of review authors. | p. 46 |
| Availability of data, code and other materials | 27 | Report which of the following are publicly available and where they can be found: template data collection forms; data extracted from included studies; data used for all analyses; analytic code; any other materials used in the review. | p. 46 |
